# Societal costs for the closest family relatives of patients with brain disorders in Denmark: a population-based cohort study

**DOI:** 10.1101/2025.06.23.25329206

**Authors:** Mette Kielsholm Thomsen, Thomas Bøjer Rasmussen, Jens Olsen, Niels Skipper, Sandra B Stallknecht, Oleguer Plana-Ripoll, Christian Fynbo Christiansen

## Abstract

**Background:** Brain disorders, including neurological conditions and mental disorders, pose a considerable economic burden in Denmark. However, the costs of illness are not limited to the patients themselves, as the consequences of living with a brain disorder may also impact close relatives.

**Aim:** We aimed to assess excess societal costs related to healthcare utilization and income loss for the closest family relatives of patients with brain disorders.

**Methods:** This population-based cohort study included the closest family relatives of patients with prevalent (by Jan 1, 2021) or incident (during 2016-2021) brain disorders. They were compared with corresponding relatives of matched population comparisons. Patients were categorized into three age strata: children and young people (0-24 years), adults (25-64 years), and older adults (65+ years). We specified criteria for identifying the closest family relative for each age stratum. Using data from national registries, we estimated attributable healthcare costs, and for working-age relatives (18-65 years), we also estimated income loss.

**Results:** In 2021, close relatives of patients with brain disorders included 125,495 fathers and 96,154 mothers of children and young people, 880,661 relatives of adults, and 378,826 relatives of older adults. The pooled attributable costs of brain disorders incurred by closest family relatives were 2,407 million EUR for prevalent disease in 2021 and 794 million EUR for incident disease the first year following incidence. The dominating cost component was income loss for working-age relatives.

**Conclusion:** The higher healthcare costs and especially the lower income for relatives of patients with brain disorders adds to the societal economic burden of brain disorders in Denmark. Informal care provided by relatives may contribute to this, underscoring the need to consider their caregiving burden in questions of healthcare prioritization.

## Introduction

Brain disorders, including neurological conditions and mental disorders, are a considerable health burden which incurred more than 500 million disability-adjusted life-years (DALYs) worldwide in 2021. The high overall disability burden is driven both by disease severity, and a high prevalence (1). The prevalence of brain disorders in Europe is considerable, with the three most common types being 152.8 million persons affected by headaches, 69.1 million affected by anxiety disorders, and 44.9 million affected by sleep disorders, in the total population of 514 million inhabitants of 30 European countries (2). In Denmark, brain disorders affected one third of the population in 2021 (3).

Brain disorders have previously been found to pose a considerable economic burden across countries (2–4). Costs of brain disorders in Europe were estimated at 798 billion EUR, or 5,550 EUR per inhabitant, in 2010 (2). In the Danish population of ∼6 million people, the total healthcare costs attributable to brain disorders is €7.5 billion yearly, in addition to an average income loss of 17,587 EUR per person with a brain disorder in Denmark (3). However, the costs of an illness are not limited to the patients themselves, as the consequences of suffering from a brain disorder impact close relatives as well, not least from the patient’s need of support and informal care in daily life and managing disease (5).

Informal caregiving is most often provided by a close family member, and more seldom by a neighbor or friend (6). Family roles, and thus the type of family relative most likely to provide informal care, change over the life course. A Danish survey among informal caregivers of patients treated in out-patient psychiatric services, found that for adult patients, 35% of their caregivers were partners and 49% were parents (7). For patients in later life, a German study of primary informal caregivers of patients with dementia, found that the primary caregiver was most often the partner or an adult child, accounting for 93% of patients (8).

Keating et al. has established a taxonomy for economic costs of informal caregiving, identifying categories of economic costs of caregiving, namely loss of income, out-of-pocket expenses and the value of time spent undertaking informal care, in addition to health and social consequences of care labor (9). Family relatives undertaking informal care may be subject to consequences for their own health, as well as their ability to undertake paid work (10), thus adding to the sum cost of brain disorders.

A survey-based study from Sweden found that even in a relatively expansive and universal welfare system, significant resources went toward providing informal care across the population, with costs of 14.5 billion EUR per year, equaling 3% of the Swedish GDP (11). Likewise, replacement costs of caregiving (costs, if all time spent on informal care were to be replaced by paid care) was estimated in Australia in 2020 at 4% of GDP (12).

Previously, the economic burden incurred by relatives of patients with brain disorders in Denmark has only been evaluated in selected mental disorders and stroke, and generally focusing on one type of relative (e.g. parents or partners) (13–19).

We aimed to extend the current evidence on societal costs of all brain disorders in Denmark, by assessing the costs related to health and income consequences of caregiving for the closest family relatives of patients with brain disorders across stages of life.

## Methods

### Setting

In Denmark, healthcare is primarily tax-funded and provided free of charge at the point of use. All residents are assigned a unique and universal civil person registration (CPR) number at birth or immigration, which enables data linkage across national registries (20).

According to Danish law, non-interventional registry-based studies do not require informed consent or approval by a research ethics committee. The study was registered at Aarhus University (j. no. 2016-051-000001/603).

### Study design

In this registry-based cohort study, we assessed societal costs among closest family relatives of persons with brain disorders in Denmark. They were compared with corresponding relatives of matched population comparisons. We estimated the excess costs in family relatives of patients with incident and prevalent brain disorders, respectively, and considered costs related to healthcare and loss of income.

### Study cohorts

#### Brain disorder cohorts

We considered 25 types of brain disorders (Supplementary Table 1) as previously defined (3).

We established an incident and a prevalent cohort for any brain disorder and for each subtype of brain disorder. The prevalent cohorts included persons with the brain disorder diagnosed within the past 20 years, among people alive and residing in Denmark on January 1, 2021. The incident cohorts included persons with a first-time diagnosis of the brain disorder of interest during January 1, 2016, to December 31, 2021, among those with no diagnosis of the specific disorder in the previous 15 years.

#### Definition of closest family relatives by age strata

We used age as a stratifying factor throughout, assessed on diagnosis date for incident cohorts and on January 1, 2021, for the prevalent cohorts. Age was categorized into three categories by “life phases”, according to World Health Organization (WHO) and National Health Services (NHS) definitions: 1) children and young people 0-24 years (21), 2) adults 25-64 years, and 3) older adults 65+ years (22).

For each patient, we identified the one or two closest family relatives among relatives that had not previously been diagnosed with the brain disorder of interest themselves. As criteria to identify who would typically be the primary relative(s) of a patient, we considered both age strata, type of family relation, geographical proximity and age of family relatives (Table 1). For children and young people (0-24 years), we included both parents if possible. For adults (25–64) we included one relative, prioritized as 1) partners, 2) parents, 3) children and 4) adult siblings as closest relatives. For older adults (65+), we also included one relative, prioritized as 1) partners, 2) adult children and 3) adult siblings as closest relatives.

**Table 1.** Identification of relatives of patients with brain disorders by age strata.

|  | **Closest family relative(s) of index patients** |
| --- | --- |
| **Children and young people 0-24 years<sup>1</sup>** | Up to two: Mother and/or father |
| **Adults 25-64 years** | One, according to priority:<br>1. Married and/or cohabiting partner<br>2. Parent living closest, by same: 1) municipality, or 2) region, or youngest parent (if living together or both same municipality/region)<br>3. Adult<sup>3</sup> child living closest, by same 1) municipality, or 2) region, or oldest child (if more than one in same municipality/region)<br>4. Adult<sup>3</sup> sibling living closest, by same 1) municipality or 2) region, or oldest sibling (if more than one in same municipality/region) |
| **Older adults 65+ years<sup>2</sup>** | One, according to priority:<br>1. Married and/or cohabiting partner<br>2. Adult<sup>3</sup> child living closest, by same 1) municipality, or 2) region, or oldest child (if more than one in same municipality/region)<br>3. Adult<sup>3</sup> sibling living closest, by same 1) municipality or 2) region, or youngest sibling (if more than one in same municipality/region) |

<sup>1</sup>According to WHO definition (21). <sup>2</sup>According to NHS (22). <sup>3</sup>Age>18 years.

#### Population comparisons and comparison relatives

We matched each set of patient and patient relative(s) with one set of comparison person and comparison relative(s).

Due to computational restrictions, this was done in two steps. First, to allow for a sufficient number of individuals to identify family relatives of same type as the patient relative, we matched each patient with up to 100 persons from the general population, among those who had not previously been diagnosed with the brain disorder of interest. Matching was done by sex and birth year using sampling with replacement, i.e. a person can be a control for multiple cases, but only once for each case. Second, for each person in the comparison cohort, we identified their family relatives without preexisting the brain disorder of interest and restricted to the closest according to the same criteria as for patient relatives. Then we restricted the comparison relatives to those who were of the same relation-type, year of birth (within five years), sex and education level as the patient relative. If this resulted in more than one eligible set of comparison person and comparison relative(s), we drew one set at random (Figure 1).

**Figure 1.**
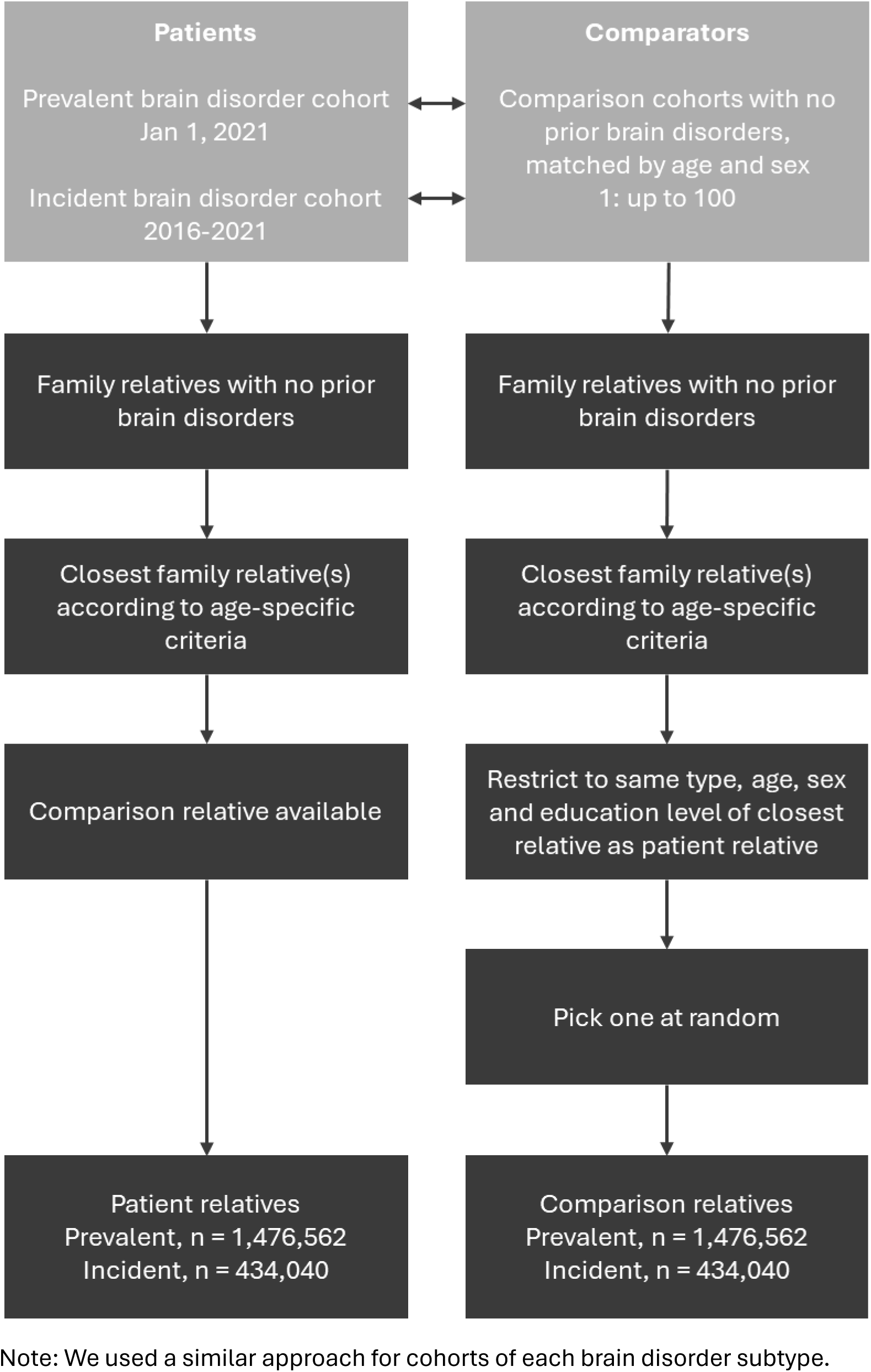
Flow diagram of identifying relatives of patients with any brain disorder, and comparison relatives Note: We used a similar approach for cohorts of each brain disorder subtype.

**Figure 2.**
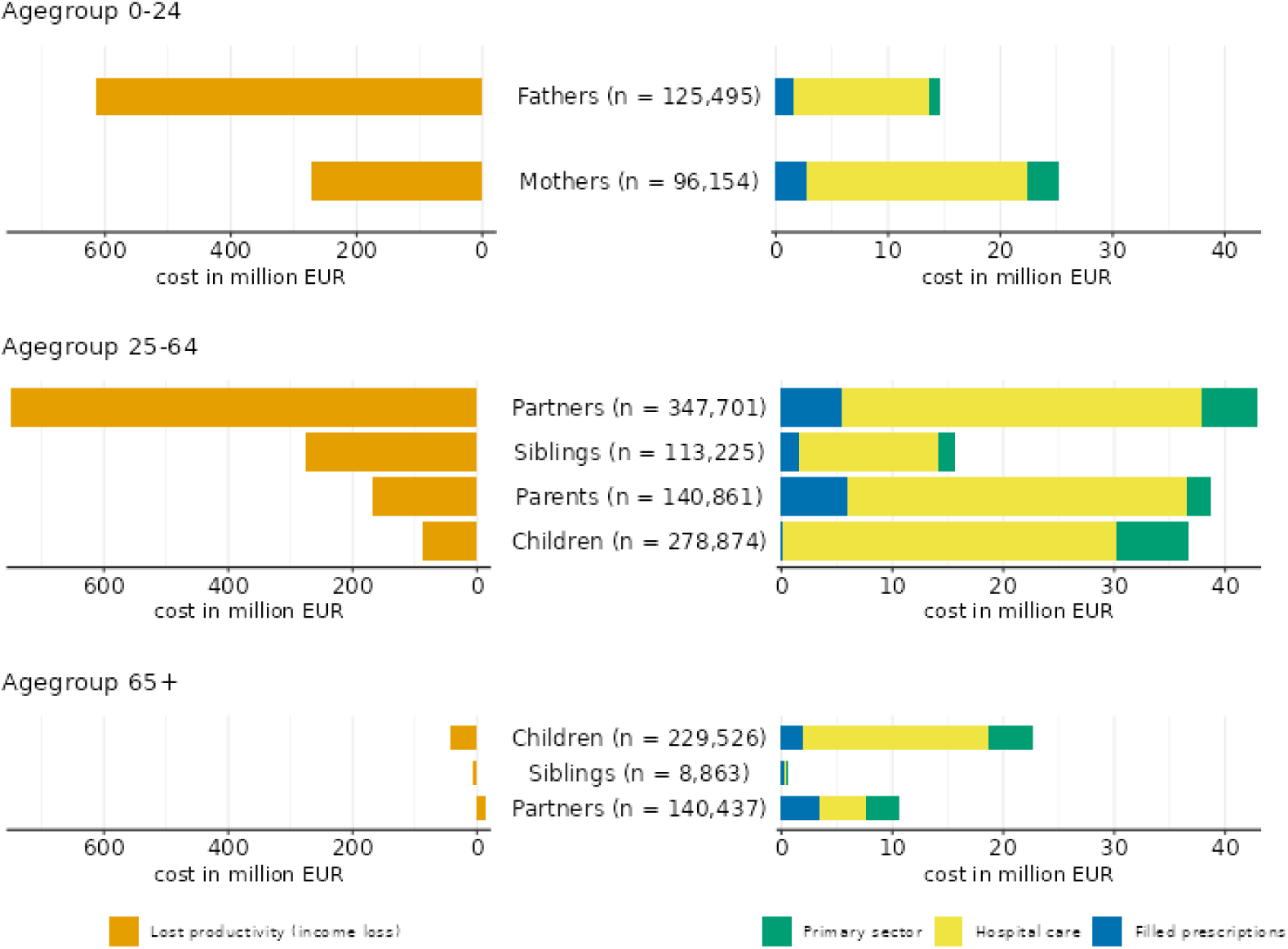
Total attributable costs of income loss (left) and healthcare (right) for relatives of patients with any prevalent brain disorder in 2021, by age strata and type of relative

### Data sources and variables

We retrieved data from the Danish National Patient Registry (DNPR), including diagnoses from both somatic and psychiatric hospital contacts (23), and the prescriptions registry (24) to identify diagnoses of brain disorders (Supplementary Table 1). From the DNPR, we also retrieved information on comorbidities in the past 10 years and calculated the Charlson Comorbidity Index (CCI) Score based on these data (25).

The specific diagnosis included in the CCI is listed in Supplementary Table 2.

To assess healthcare costs, we included primary sector healthcare, hospital care, and filled prescriptions. For primary sector healthcare, we used data from the Danish Health Service Registry (26), to identify subsidized services provided by general practitioners, private practicing specialists and dentists, and the cost of each type of services was retrieved from payment-agreements between Danish Regions and provider unions.

Hospital-based healthcare was assessed as admissions, outpatient contacts and emergency room visits from the DNPR, and linked to Diagnosis-Related Group (DRG) and Danish Ambulatory Grouping System (DAGS) tariffs. Tariffs for psychiatric hospital care was only available until 2019, and therefore we imputed their values for the period 2020-2022 based on data from the years prior, following the same method as described in a previous study (3).

Filled prescriptions were identified using data from the Prescription Registry, which contains information on all prescriptions filled at community pharmacies, including the price of each specific drug (24).

Data on income was retrieved from the Income Registry (27), and defined as individual earnings (not including social benefits) before taxes. An overview of data tables and variables used to calculate costs are available in Supplementary Table 3.

To adjust for inflation, all costs except prescription medication costs due to its fluctuating price-structure (elaborated in Supplementary Table 3), were adjusted to the 2023 level using the Danish consumer price index (28). Original cost data was in Danish currency (DKK), and we used a fixed exchange rate of 1 EUR = 7.45 DKK for conversion to EUR.

### Statistical analysis

We used descriptive statistics to characterize patient and comparison cohorts, patient relatives and comparison relatives according to sex, age, education level, and CCI score. In the main analysis we focused on any brain disorder and secondarily reported corresponding results for specific brain disorders.

We assessed the societal costs of brain disorders in family relatives including costs both related to healthcare services and income loss. For the prevalent cohorts, costs were assessed for the calendar year 2021, and for the incident cohorts, costs were assessed in the year following diagnosis.

We computed both the actual and attributable costs of healthcare services. The attributable healthcare costs were calculated as the difference in healthcare service costs between the patient relatives and their matched comparison relatives.

We estimated attributable income loss in family relatives of working age, i.e. 18 to 65 years. The attributable income loss was calculated as the yearly income (earnings) before taxes for patient relatives, minus the yearly income (earnings) of comparison relatives. Secondarily, we estimated the relative income loss as the mean attributable income loss proportional to the mean income of comparison relatives.

## Results

### Prevalent brain disorder cohort

We identified 263,026 children and young people (0-24 years), 1,103,732 adults (25-64 years), and 566,848 older adults (65+ years) with any prevalent brain disorder on January 1, 2021. Among these, 34%, 10% and 28% in each age strata, respectively, had no available closest relative alive, living in Denmark and without prior brain disorders (Supplementary Table 4a). The proportions of patients without an eligible closest family relative were lower when considering specific prevalent brain disorders in the two youngest age strata (ranging between 1% for adults with eating disorders to 19% for children and young people with depression) but was up to 36% and 68% for older adults with cerebral palsy or intellectual disabilities, respectively (Supplementary Table 60 and 168).

Sub-cohorts of patient relatives each consisted of 125,495 fathers, and 96,154 mothers of children and young people, 880,661 relatives of adults, and 378,826 relatives of older adults with any type of brain disorders (Table 2).

**Table 2.**
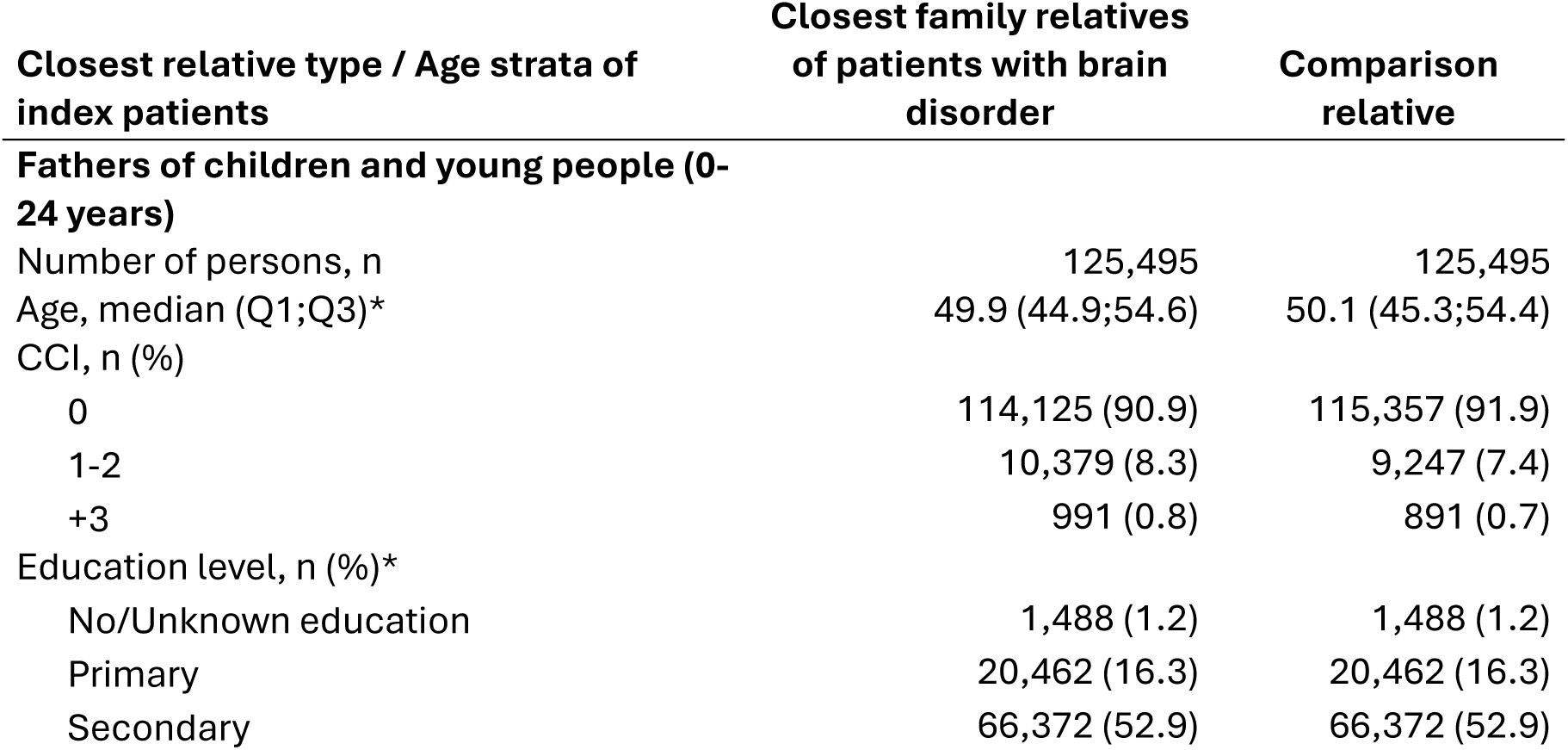
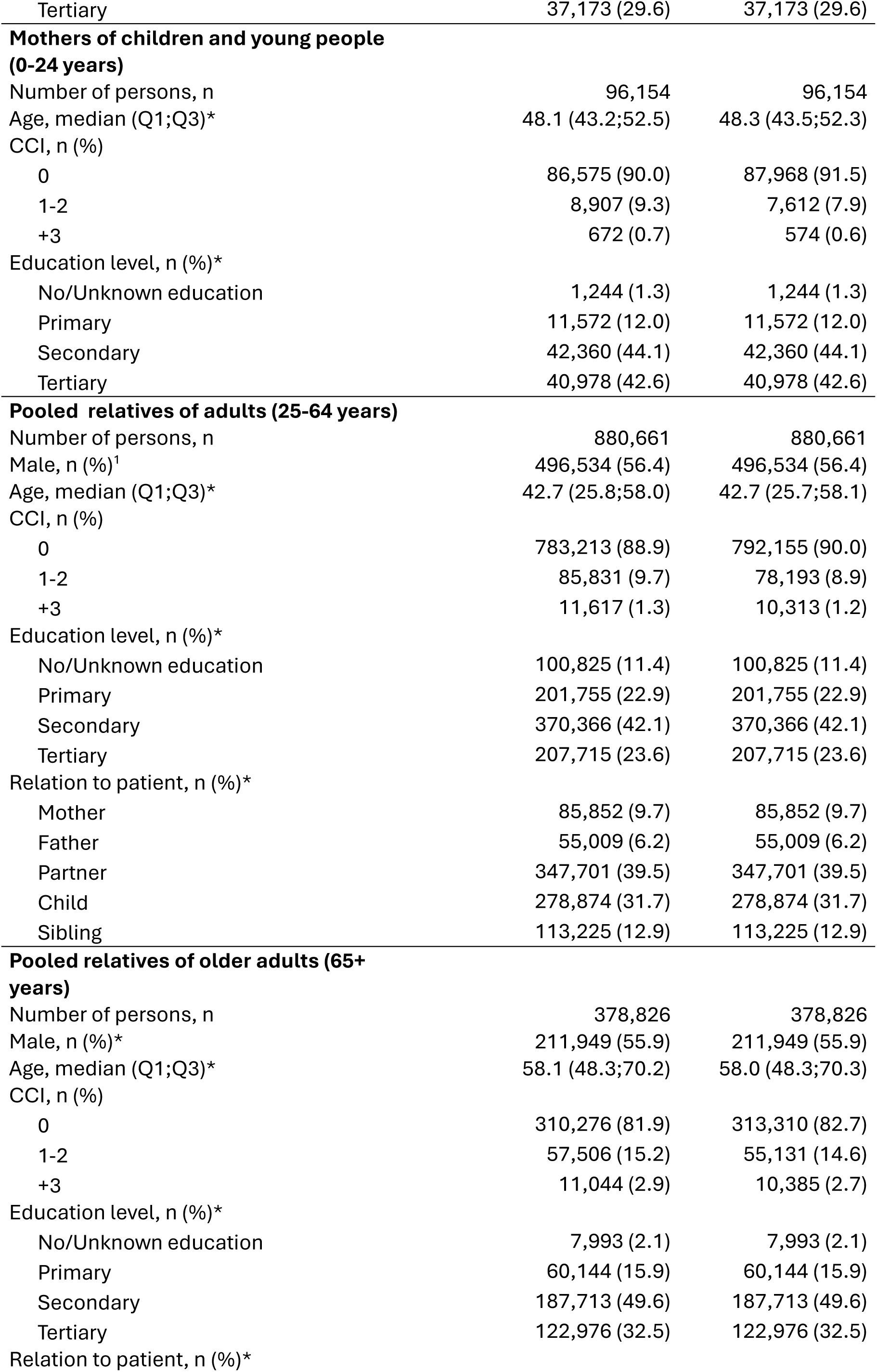
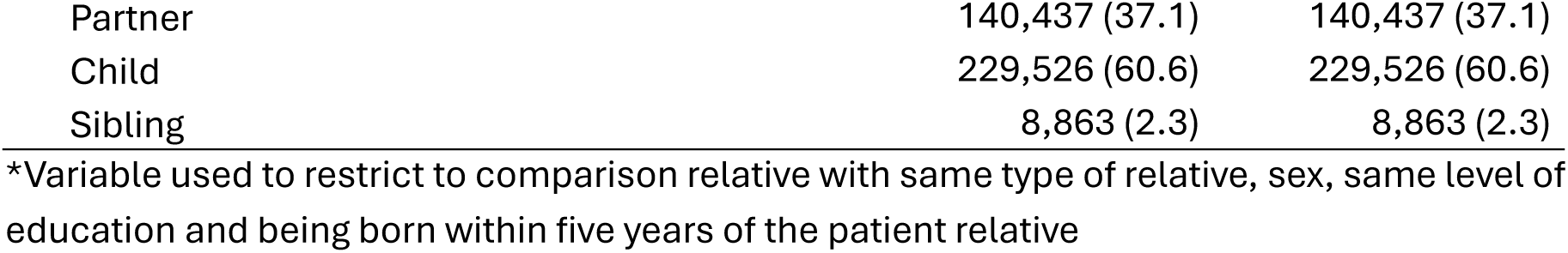
Characteristics of patient and comparison relatives of patients with any prevalent brain disorder, by patient age strata.

| **Closest relative type / Age strata of index patients** | **Closest family relatives of patients with brain disorder** | **Comparison relative** |
| --- | --- | --- |
| **Fathers of children and young people (0-24 years)** |  |  |
| Number of persons, n | 125,495 | 125,495 |
| Age, median (Q1;Q3)* | 49.9 (44.9;54.6) | 50.1 (45.3;54.4) |
| CCI, n (%) |  |  |
| 0 | 114,125 (90.9) | 115,357 (91.9) |
| 1-2 | 10,379 (8.3) | 9,247 (7.4) |
| +3 | 991 (0.8) | 891 (0.7) |
| Education level, n (%)* |  |  |
| No/Unknown education | 1,488 (1.2) | 1,488 (1.2) |
| Primary | 20,462 (16.3) | 20,462 (16.3) |
| Secondary | 66,372 (52.9) | 66,372 (52.9) |
| Tertiary | 37,173 (29.6) | 37,173 (29.6) |
| **Mothers of children and young people (0-24 years)** |  |  |
| Number of persons, n | 96,154 | 96,154 |
| Age, median (Q1;Q3)* | 48.1 (43.2;52.5) | 48.3 (43.5;52.3) |
| CCI, n (%) |  |  |
| 0 | 86,575 (90.0) | 87,968 (91.5) |
| 1-2 | 8,907 (9.3) | 7,612 (7.9) |
| +3 | 672 (0.7) | 574 (0.6) |
| Education level, n (%)* |  |  |
| No/Unknown education | 1,244 (1.3) | 1,244 (1.3) |
| Primary | 11,572 (12.0) | 11,572 (12.0) |
| Secondary | 42,360 (44.1) | 42,360 (44.1) |
| Tertiary | 40,978 (42.6) | 40,978 (42.6) |
| **Pooled relatives of adults (25-64 years)** |  |  |
| Number of persons, n | 880,661 | 880,661 |
| Male, n (%)<sup>1</sup> | 496,534 (56.4) | 496,534 (56.4) |
| Age, median (Q1;Q3)* | 42.7 (25.8;58.0) | 42.7 (25.7;58.1) |
| CCI, n (%) |  |  |
| 0 | 783,213 (88.9) | 792,155 (90.0) |
| 1-2 | 85,831 (9.7) | 78,193 (8.9) |
| +3 | 11,617 (1.3) | 10,313 (1.2) |
| Education level, n (%)* |  |  |
| No/Unknown education | 100,825 (11.4) | 100,825 (11.4) |
| Primary | 201,755 (22.9) | 201,755 (22.9) |
| Secondary | 370,366 (42.1) | 370,366 (42.1) |
| Tertiary | 207,715 (23.6) | 207,715 (23.6) |
| Relation to patient, n (%)* |  |  |
| Mother | 85,852 (9.7) | 85,852 (9.7) |
| Father | 55,009 (6.2) | 55,009 (6.2) |
| Partner | 347,701 (39.5) | 347,701 (39.5) |
| Child | 278,874 (31.7) | 278,874 (31.7) |
| Sibling | 113,225 (12.9) | 113,225 (12.9) |
| **Pooled relatives of older adults (65+ years)** |  |  |
| Number of persons, n | 378,826 | 378,826 |
| Male, n (%)* | 211,949 (55.9) | 211,949 (55.9) |
| Age, median (Q1;Q3)* | 58.1 (48.3;70.2) | 58.0 (48.3;70.3) |
| CCI, n (%) |  |  |
| 0 | 310,276 (81.9) | 313,310 (82.7) |
| 1-2 | 57,506 (15.2) | 55,131 (14.6) |
| +3 | 11,044 (2.9) | 10,385 (2.7) |
| Education level, n (%)* |  |  |
| No/Unknown education | 7,993 (2.1) | 7,993 (2.1) |
| Primary | 60,144 (15.9) | 60,144 (15.9) |
| Secondary | 187,713 (49.6) | 187,713 (49.6) |
| Tertiary | 122,976 (32.5) | 122,976 (32.5) |
| Relation to patient, n (%)* |  |  |
| Partner | 140,437 (37.1) | 140,437 (37.1) |
| Child | 229,526 (60.6) | 229,526 (60.6) |
| Sibling | 8,863 (2.3) | 8,863 (2.3) |

\*Variable used to restrict to comparison relative with same type of relative, sex, same level of education and being born within five years of the patient relative

The median age of fathers and mothers of children and young people with any brain disorder was 49.9 years (inter quartile range, IQR: 44.9;54.6), and 48.1 years (IQR: 43.2;52.5), respectively. The majority had a low comorbidity burden, with 90.9% and 90.0% having a CCI score of 0 (Table 2).

The closest relatives of adults with any brain disorder, were mostly partners (39.5%) and adult children (31.7%), and 15.9% and 12.9% were parents and siblings, respectively.

Overall, 56.4% of them were men, median age was 42.7 years (IQR: 25.8;58.0) and 88.9% had a CCI score of 0. The closest relatives of older adults were 37.1% partners, 60.6% adult children and 2.3% siblings, with overall 55.9% men, and median age 58.1 years (IQR: 48.3;70.2). More relatives of older adults had comorbidities compared to both relatives of children and young people and relatives of adults with only 81.9% having a CCI score of 0. Comparison relatives had similar characteristics as patient relatives (Table 2).

### Costs of prevalent brain disorders

In sum, the total attributable costs of brain disorders incurred by closest relatives of patients with any brain disorder was 2,407 million EUR in 2021.

Mothers and fathers of children and young people with any brain disorder had total attributable healthcare costs of 14.7 and 25.2 million EUR in 2021, which corresponds to 116 and 262 EUR per person, respectively. The attributable healthcare costs in 2021 were 134.1 million EUR in total and 152 EUR per person for closest relatives of adults, and 34.0 million EUR in total and 90 EUR per person for closest relatives of older adults. The costliest component of healthcare was consistently hospital care (Table 3).

**Table 3.**
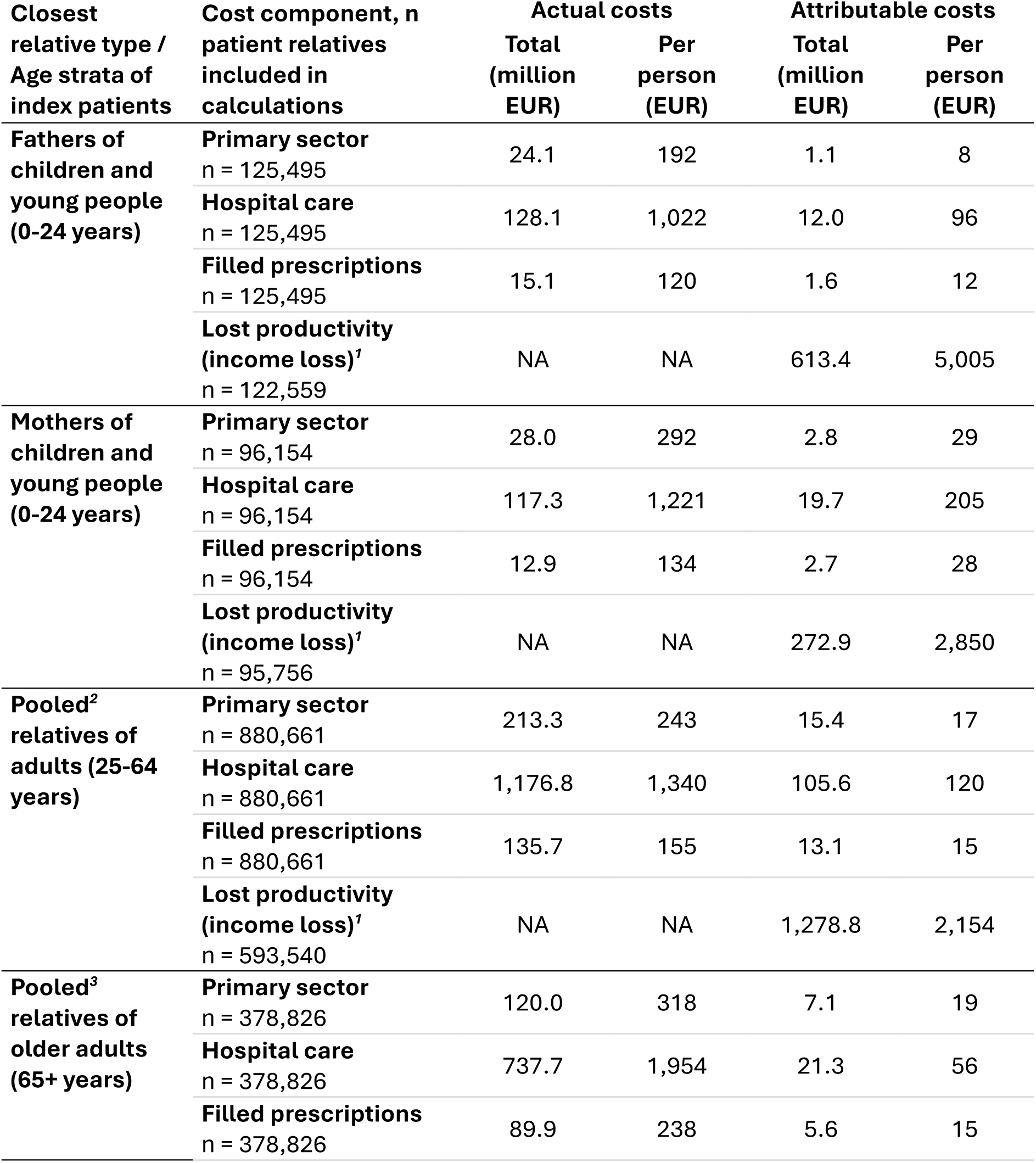
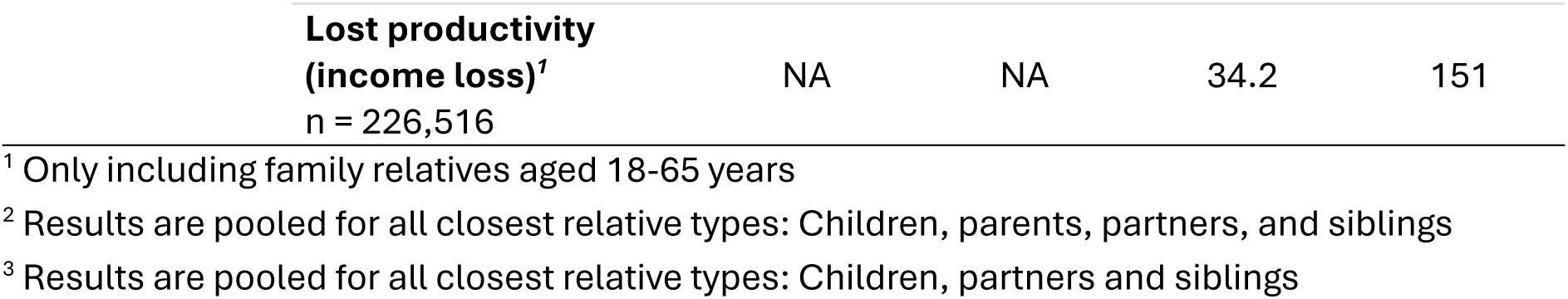
Societal costs in 2021 of prevalent brain disorder in family relatives of patients with any prevalent brain disorder, by patient age strata.

Income loss led to total attributable costs of 613.4 million EUR for fathers and 272.9 million EUR for mothers of children and young people, 1,278.8 million EUR for relatives of adults, and 34.2 million EUR for relatives of older adults, or between 151 and 5,005 EUR per person depending on type of relative in 2021 (Table 3).

Income loss on the relative scale was more similar for mothers and fathers of children and young people with 5.4% for fathers and 4.5% for mothers, respectively. The proportional income loss was 3.3% for relatives of adults and 0.2% for relatives of older adults.

For mothers and fathers of children and young people, the total attributable costs and income loss was highest when patients suffered from developmental and behavioral disorders, whereas the costliest disorder for relatives of adults and older adults was depression (Supplementary Table 88 and 100).

### Incident brain disorder cohort

The proportions of patients with incident brain disorders, who did not have a closest family relative, were similar to those for patients with prevalent brain disorders (Supplementary Table 4b). The incident cohorts of patient relatives each consisted of 79,433 fathers and 63,931 mothers of children and young people (0-24 years), 188,475 relatives (53.4% men) of adults (25-64 years) and 102,748 relatives (50.7% men) of older adults (65+ years) with incident brain disorders of any type (Table 4).

**Table 4.** Characteristics of patient and comparison relatives of patients with any incident brain disorder, by patient age strata.

| **Closest relative type / Age strata of index patients** | **Closest family relatives of patients with brain disorder** | **Comparison relative** |
| --- | --- | --- |
| **Fathers of children and young people (0-24 years)** |  |  |
| Number of persons, n | 79,433 | 79,433 |
| Age, median (Q1;Q3)* | 47.7 (41.9;52.8) | 47.8 (42.1;52.6) |
| CCI, n (%) |  |  |
| 0 | 73,070 (92.0) | 73,557 (92.6) |
| 1-2 | 5,858 (7.4) | 5,387 (6.8) |
| +3 | 505 (0.6) | 489 (0.6) |
| Education level, n (%)* |  |  |
| No/Unknown education | 1,332 (1.7) | 1,332 (1.7) |
| Primary | 13,155 (16.6) | 13,155 (16.6) |
| Secondary | 41,212 (51.9) | 41,212 (51.9) |
| Tertiary | 23,734 (29.9) | 23,734 (29.9) |
| **Mothers of children and young people (0-24 years)** |  |  |
| Number of persons, n | 63,931 | 63,931 |
| Age, median (Q1;Q3)* | 45.8 (39.9;50.7) | 45.9 (40.2;50.5) |
| CCI, n (%) |  |  |
| 0 | 58,137 (90.9) | 58,929 (92.2) |
| 1-2 | 5,369 (8.4) | 4,624 (7.2) |
| +3 | 425 (0.7) | 378 (0.6) |
| Education level, n (%)* |  |  |
| No/Unknown education | 1,319 (2.1) | 1,319 (2.1) |
| Primary | 8,285 (13.0) | 8,285 (13.0) |
| Secondary | 27,433 (42.9) | 27,433 (42.9) |
| Tertiary | 26,894 (42.1) | 26,894 (42.1) |
| **Pooled relatives of adults (25-64 years)** |  |  |
| Number of persons, n | 188,475 | 188,475 |
| Male, n (%)* | 100,623 (53.4) | 100,623 (53.4) |
| Age, median (Q1;Q3)* | 41.4 (25.9;56.1) | 41.4 (25.8;56.1) |
| CCI, n (%) |  |  |
| 0 | 169,354 (89.9) | 170,729 (90.6) |
| 1-2 | 16,859 (8.9) | 15,776 (8.4) |
| +3 | 2,262 (1.2) | 1,970 (1.0) |
| Education level, n (%)* |  |  |
| No/Unknown education | 25,157 (13.3) | 25,157 (13.3) |
| Primary | 41,204 (21.9) | 41,204 (21.9) |
| Secondary | 75,802 (40.2) | 75,802 (40.2) |
| Tertiary | 46,312 (24.6) | 46,312 (24.6) |
| Relation to patient, n (%)<sup>*</sup> |  |  |
| Mother | 17,685 (9.4) | 17,685 (9.4) |
| Father | 10,654 (5.7) | 10,654 (5.7) |
| Partner | 85,677 (45.5) | 85,677 (45.5) |
| Child | 53,740 (28.5) | 53,740 (28.5) |
| Sibling | 20,719 (11.0) | 20,719 (11.0) |
| **Pooled relatives of older adults (65+ years)** |  |  |
| Number of persons, n | 102,748 | 102,748 |
| Male, n (%)<sup>*</sup> | 52,125 (50.7) | 52,125 (50.7) |
| Age, median (Q1;Q3)<sup>*</sup> | 58.6 (49.6;70.9) | 58.4 (49.6;70.9) |
| CCI, n (%) |  |  |
| 0 | 83,799 (81.6) | 84,861 (82.6) |
| 1-2 | 15,646 (15.2) | 14,970 (14.6) |
| +3 | 3,303 (3.2) | 2,917 (2.8) |
| Education level, n (%)<sup>*</sup> |  |  |
| No/Unknown education | 2,163 (2.1) | 2,163 (2.1) |
| Primary | 18,860 (18.4) | 18,860 (18.4) |
| Secondary | 51,579 (50.2) | 51,579 (50.2) |
| Tertiary | 30,146 (29.3) | 30,146 (29.3) |
| Relation to patient, n (%)<sup>*</sup> |  |  |
| Partner | 39,382 (38.3) | 39,382 (38.3) |
| Child | 62,495 (60.8) | 62,495 (60.8) |
| Sibling | 871 (0.8) | 871 (0.8) |

<sup>\*</sup>Variable used to restrict to comparison relative with same type of relative, sex, same level of education and being born within five years of the patient relative

### Costs of incident brain disorders

In sum, the total attributable costs of brain disorders incurred by closest relatives in the first year after incidence of any brain disorder was 794 million EUR.

By patient age strata, total attributable healthcare costs for relatives amounted to 8.7 million EUR and 11.4 million EUR for fathers and mothers of children and young people, 31.6 million EUR for relatives of adults and 19.5 million EUR for relatives of older adults in the first year (Table 5 and Figure 4). Per person, attributable healthcare costs in the first year after incidence varied between 111 EUR for fathers of children and young people and 191 EUR for relatives of older adults. Income loss posed the highest costs with 328.2 million EUR for fathers and 137.6 million EUR for mothers of children and young people, 209 million EUR for relatives of adults and 47.6 million EUR for relatives of older adults in the first year after incidence. Per person, income loss varied between 792 EUR for relatives of older adults and 4,219 EUR for fathers of children and young people. The proportional income loss was 5.0% for fathers and 3.8% for mothers of children and young people, 2.6% for relatives of adults, and 1.1% for relatives of older adults in the first year.

**Figure 4.**
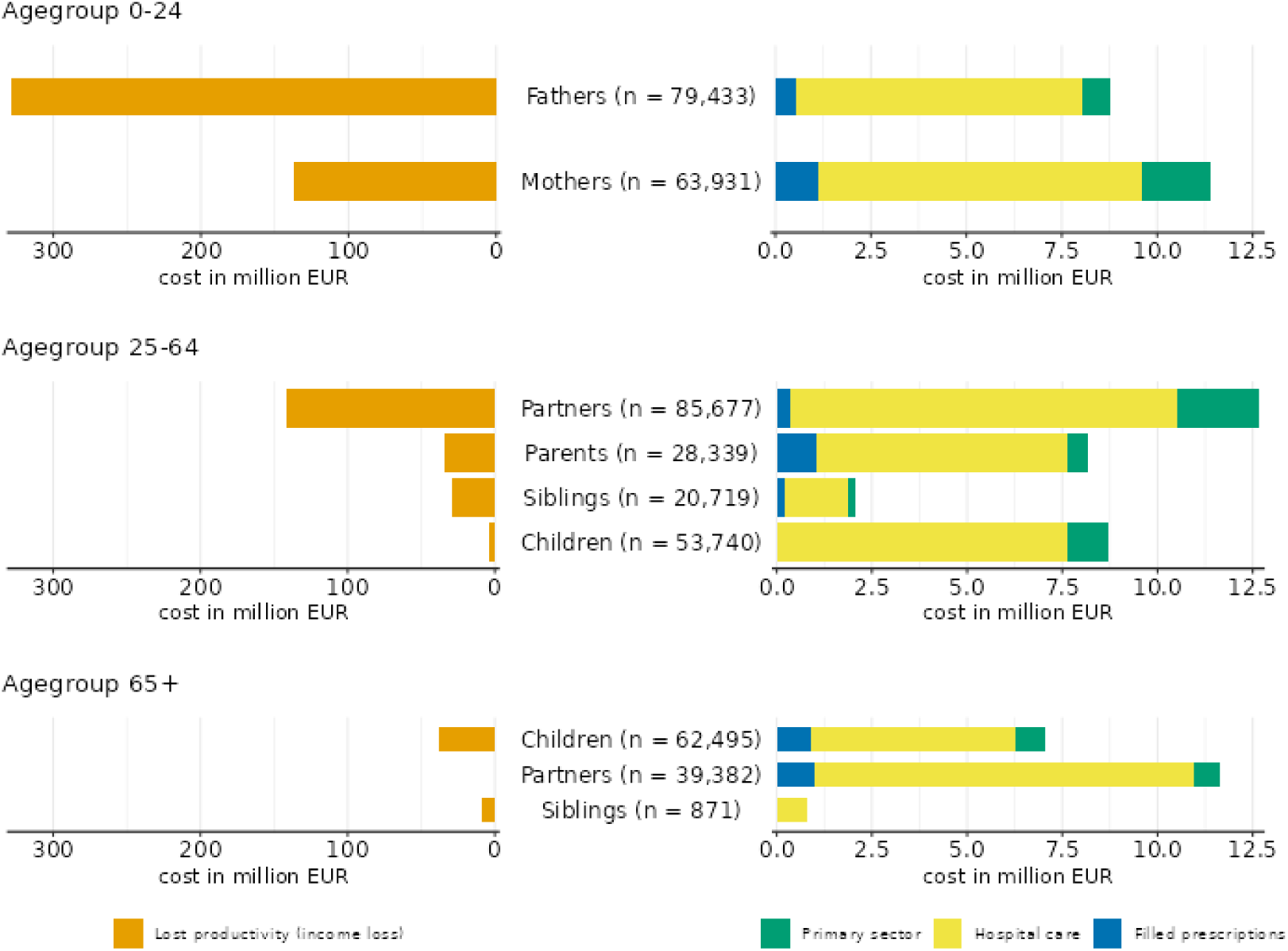
Total attributable costs of income loss (left) and healthcare (right) for relatives of patients with any incident brain disorder in 2016-2021 the first year following incidence, by age strata and type of relative

**Table 5.**
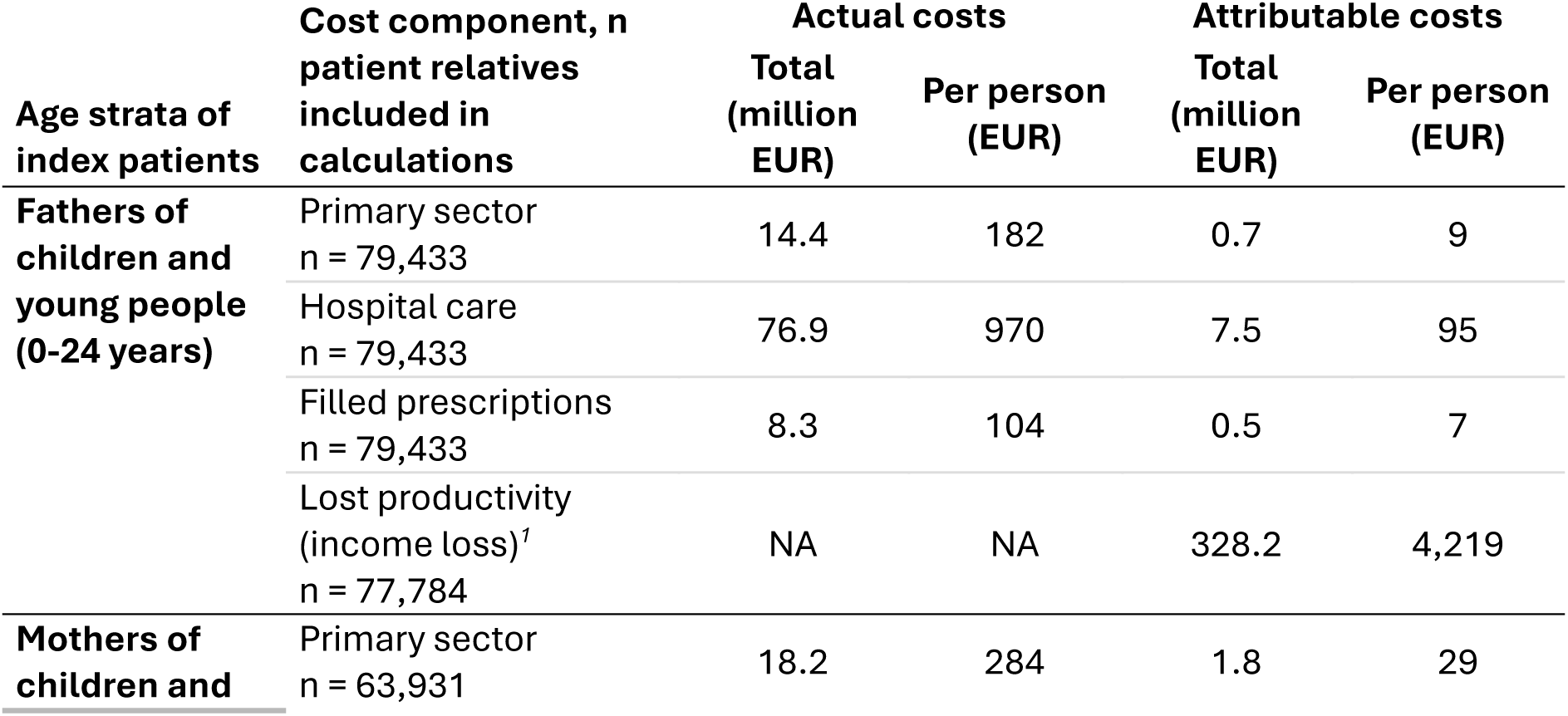
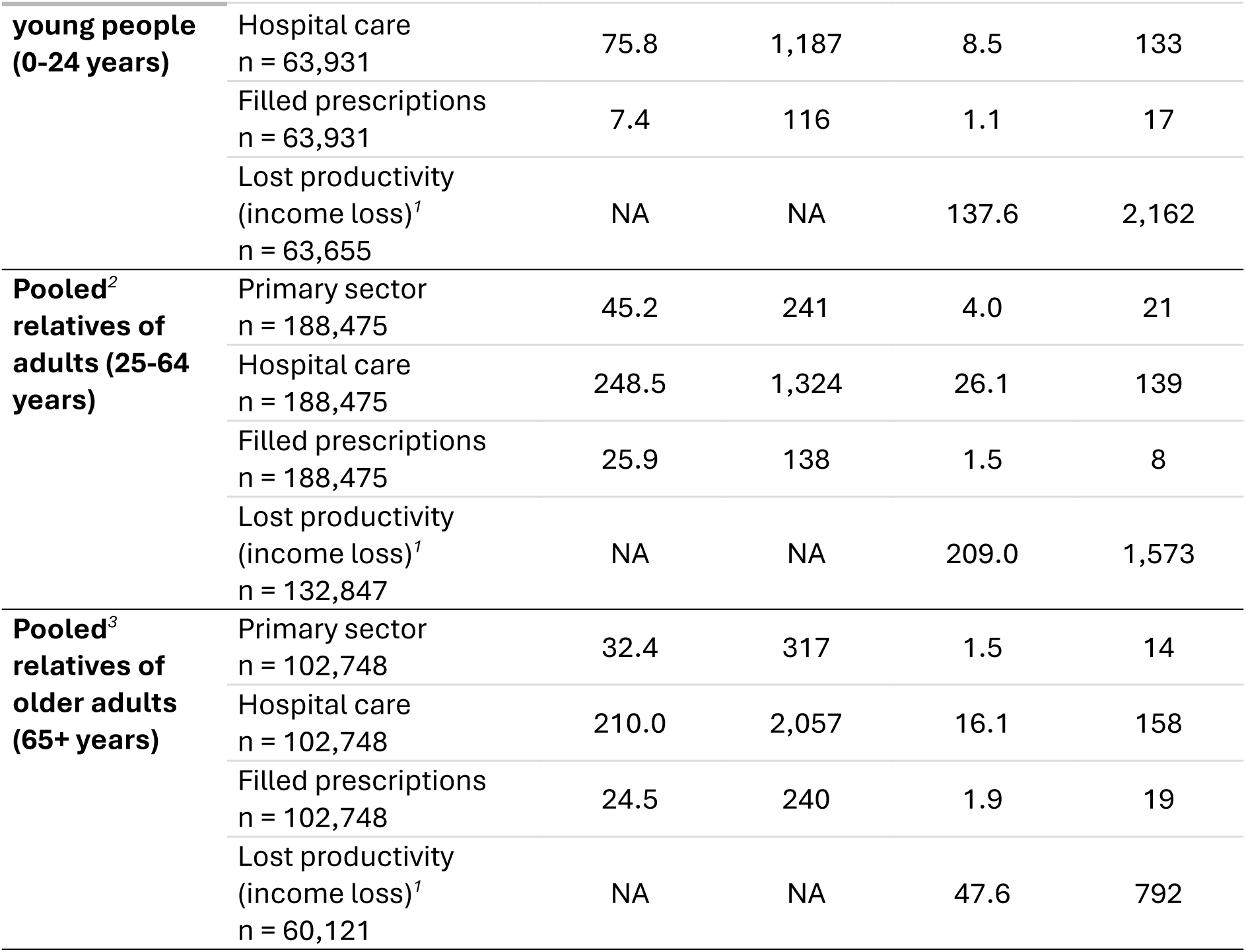
Societal costs of incident brain disorder in family relatives of patients with any incident brain disorder the first year following incidence, by patient age strata.

For mothers and fathers of children and young people, the healthcare costs and income loss in the first year was highest when their children suffered from developmental and behavioral disorders, stress-related disorders or sleep disorders, whereas the costliest disorders for relatives of adults were depression, stress-related disorders and sleep disorders. For relatives of older adults, healthcare costs were highest when the patient suffered from sleep disorders or depression, and income loss were highest when the patient suffered from stroke or dementia (Supplementary Table 70, 82, 262, and 286).

## Discussion

In 2021, close relatives of patients with brain disorders included 125,495 fathers and 96,154 mothers of children and young people, 880,661 relatives of adults, and 378,826 relatives of older adults. The pooled societal costs of brain disorders incurred by closest family relatives was 2,407 million EUR for prevalent disease in 2021 and 794 million EUR for incident disease the first year. The dominating cost component was income loss, which was proportionally highest for fathers of children and young people with brain disorders.

Previously, Fuglsang et al. estimated the patient-incurred costs of brain disorders which were measured in billions rather than millions of EUR (3), and yet, our cost estimates from the family-relative perspective add an important component to the overall economic burden of brain disorders in Denmark. At the same time, the costs illustrate both the importance of informal care provided by family relatives, and the personal health and income consequences of caregiving for a family member with a brain disorder.

The previous study by Fuglsang et al. demonstrated a high degree of excess costs for patients with brain disorders (3), and therefore including relatives who themselves had brain disorders would overestimate our results. When considering any brain disorder, a fair proportion of patients had no close relative alive and with no previous brain disorder of their own; this was overall less of an issue when considering specific brain disorders.

At the cost of representativeness of patients with brain disorders in general, our inclusion criteria led to relatives of patients with brain disorders being similar to the identified comparators on age, sex, education level and morbidity burden. However, there may be some residual confounding that we were not able to account for, as e.g. Komulainen et al previously found that parents of children with mental disorders had lower income compared with population comparators, even five years before the child was diagnosed (14). This could be due to chronic disease persisting years before diagnosis, or due to an excess risk of mental disorders in lower income families.

Using registry data to identify close family members, i.e. the assumed informal caregivers in our study, enables a population perspective, and also identified all family members of persons with brain disorders, and not only caregivers of patients able to refer a relative to e.g. participate in a survey, directly ascertaining the primary informal caregiver. As such, we may have underestimated the societal costs of brain disorders incurred by caregivers, as presumably not all who were defined as closest family members in our study, provided informal care. Such care may not have been received at all or may have been provided by another person in the family or social network, whom we were not able to account for.

We considered societal costs of the health and income consequences of informal caregiving, whereas other approaches include estimating the value of time spent caregiving, as e.g. a multinational European study, which estimated the cost of care overall per dementia patient at €7,820 (95% CI: €7,194–€8,446), of which 54% was the value of informal care, and only 16% was direct medical costs (13).

An Australian caregiver study found that the longer a person is a caregiver, the greater the consequences it has for paid work, and thereby income (29), which is in line with our finding that the income loss was higher when looking at prevalent brain disorders compared with incident brain disorders, for relatives of children and young people and of adults.

Denmark traditionally had a higher degree of institutionalized care, as opposed to family-based informal care playing a larger role in Southern Europe (13), however care is increasingly moving out of hospitals, and a higher degree of family involvement is expected (30). The costs of brain disorders incurred from patient relatives may therefore increase in the future in Demark.

Due to the high prevalence of brain disorders in the population, it is likely that many families suffer from more than one case of brain disorders, increasing the family burden. Accumulation of brain disorders in families, or couples, as well as the burden of caregiving in “sandwich” generations caring for both offspring and parents with brain disorders at the same time, pose important topics for future research on the caregiver burden of brain disorders.

## Conclusions

We found that costs incurring from the consequences experienced by relatives of patients with brain disorders contribute to the overall societal burden of brain disorders in Denmark. Informal care provided by relatives may contribute to this finding and underscores the need to acknowledge and address their caregiving burden, especially in light of future healthcare prioritizations that may shift more care responsibilities onto patient relatives.

## Data Availability

The data underlying the results are from the specified Danish registries, and not publicly available. Researchers who fulfil the requirements set by the data providers (Statistics Denmark and the Danish Health Data Authority) can obtain similar data.

