## Supplementary materials for "Societal costs for the closest family relatives of patients with brain disorders in Denmark: a population-based cohort study"

Contents

Supplementary Table 1 Definitions of brain disorders
Supplementary Table 2 Diagnosis codes Charlson Comorbidity Index
Supplementary Table 3 Data and variables to assess costs
Supplementary Table 4a Patients with prevalent any brain disorder, closest relatives identified and with at least one eligible comparator
Supplementary Table 4b Patients with incident any brain disorder, closest relatives identified and with at least one eligible comparator
Supplementary Table 5 Characteristics of included patients with any prevalent or incident brain disorder, by age patient strata
Supplementary Tables 6-17 Alcohol use disorder
Supplementary Tables 18-29 Anxiety
Supplementary Tables 30-41 Bipolar disorder
Supplementary Tables 42-53 Brain tumors
Supplementary Tables 54-65 Cerebral palsy
Supplementary Tables 66-77 Dementia
Supplementary Tables 78-89 Depression
Supplementary Tables 90-101 Developmental and behavioural disorders
Supplementary Tables 102-113 Drug use disorder
Supplementary Tables 114-125 Eating disorders
Supplementary Tables 126-137 Epilepsy
Supplementary Tables 138-149 Headache
Supplementary Tables 150-161 Infections of the CNS
Supplementary Tables 162-173 Intellectual disability
Supplementary Tables 174-185 Multiple sclerosis
Supplementary Tables 186-197 Neuromuscular disorders
Supplementary Tables 198-209 Other neurodegenerative disorders
Supplementary Tables 210-221 Parkinson’s disease
Supplementary Tables 222-233 Personality disorders
Supplementary Tables 234-245 Polyneuropathy
Supplementary Tables 246-257 Schizophrenia spectrum disorders
Supplementary Tables 258-269 Sleep disorders
Supplementary Tables 270-281 Stress-related disorders
Supplementary Tables 282-293 Stroke
Supplementary Tables 294-305 Traumatic brain injury

#### Supplementary Table 1 Definitions of brain disorders

| Brain disorder | Code type (and/or) | Codes | Subcodes excluded |
|---|---|---|---|
| Alcohol abuse | ICD-10 | F10, E244, G312, G621, G721, I426, K292, K70, K852, K860, Q860, Z502, or Z714 | F100 |
| | ATC | V03AA or N07BB | |
| Anxiety disorders | ICD-10 | F40, F41, or F42 | |
| | ATC | N06A with indication 371, 830, or 163 | |
| Bipolar disorder | ICD-10 | F30 or F31 | |
| | ATC | N05AN01 | |
| Brain tumors | ICD-10 | C70, C71, C72, D32, D33, D42, or D43 | |
| Cerebral palsy | ICD-10 | G80 | |
| Dementia | ICD-10 | B220A, F00, F01, F02, F03, F1073, F1173, F1273, F1373, F1473, F1573, F1673, F1873, F1973, G30, G310B, G311, G318, or G319 | |
| | ATC | N06D | |
| Depression | ICD-10 | F32 or F33 | |
| | ATC | N06A with indication 168 or 270 | |
| Developmental and behavioural disorders | ICD-10 | F84 or F9 | F99 |
| | ATC | N06BA02, N06BA04, N06BA09, NA06BA12, or C02AC02 with indication 375, 376, or 391 | |
| Drug abuse | ICD-10 | F1, Z722 or Z864 | F10, F110, F130, F17 or F180 |
| | ATC | N07BC01 or N07BC51 | |
| Eating disorders | ICD-10 | F500, F501, F502, or F503 | |
| Epilepsy | ICD-10 | G40 | |
| Headache | ICD-10 | G43 or G44 | |
| | ATC | N02C | |
| Infections of the CNS | ICD-10 | G0, A17, A321, A390, A521, A522, A523, A83, A84, A85, A87, A89, B003, B004, B010, B011, B020, B021, B582, B451, or B375 | G08 or G09 |
| Intellectual disability | ICD-10 | F7, Q90, or Q992 | |
| Multiple sclerosis | ICD-10 | G35 | |
| Neuromuscular disorders | ICD-10 | G70, G71, G72, or G73 | |
| Other neurodegenerative disorders | ICD-10 | G10, G11, G12, G13, or G14 | G130 or G139 |
| Parkinson's disease | ICD-10 | G20 or G23 | |
| | ATC | N04BA, N04BB, N04BD, or N04BX | |
| Personality disorders | ICD-10 | F60 | |
| Polyneuropathy | ICD-10 | G60, G61, G62, G63, G64, or G130 | |
| Schizophrenia spectrum disorders | ICD-10 | F2 | |
| Sleep disorders | ICD-10 | F51, G470, G471, G473, or G474 | |
| | ATC | N05CF01, N05CH01, N06BA07, N06BA14, N07XX04, or N07XX11 | |
| Stress related disorders | ICD-10 | F43 | |
| Stroke | ICD-10 | I60, I61, I63, or I64 | I608 |
| Traumatic brain injury | ICD-10 | F072, S020, S021, S027, S029, or S06 | |

**Supplementary Table 2 Diagnosis codes Charlson Comorbidity Index**

| CCI disease | ICD-10 codes |
|---|---|
| Myocardial infarction | I21, I22, or I23 |
| Congestive heart failure | I50, I110, I130, or I132 |
| Peripheral vascular disease | I70, I71, I72, I73, I74, or I77 |
| Cerebrovascular disease | I6, G45, or G46 |
| Dementia | F00, F01, F02, F03, F051, or G30 |
| Chronic pulmonary disease | J40, J41, J42, J43, J44, J45, J46, J47, J60, J61, J62, J63, J64, J65, J66, J67, J684, J701, J703, J841, J920, J961, J982, or J983 |
| Connective tissue disease | M05, M06, M08, M09, M30, M31, M32, M33, M34, M35, M36, or D86 |
| Ulcer disease | K221, K25, K26, K27, or K28 |
| Mild liver disease | B18, K700, K701, K702, K703, K709, K71, K73, K74, or K760 |
| Diabetes without end-organ damage | E100, E101, E109, E110, E111, or E119 |
| Hemiplegia | G81 or G82 |
| Moderate to severe renal disease | I12, I13, N00, N01, N02, N03, N04, N05, N07, N11, N14, N17, N18, N19, or Q61 |
| Diabetes with end-organ damage | E102, E103, E104, E105, E106, E107, E108, E112, E113, E114, E115, E116, E117, or E118 |
| Solid cancer tumour | C0, C1, C2, C3, C4, C5, C6, C70, C71, C72, C73, C74, or C75 |
| Leukaemia | C91, C92, C93, C94, or C95 |
| Lymphoma | C81, C82, C83, C84, C85, C88, C90, or C96 |
| Moderate to severe liver disease | B150, B160, B162, B190, K704, K72, K766, or I85 |
| Metastatic solid tumour | C76, C77, C78, C79, or C80 |
| AIDS | B21, B22, B23, or B24 |

#### Supplementary Table 3 Data and variables to assess costs

| Type of information | Registry or information source | Variable(s) | Price level |
|---|---|---|---|
| **Primary health care sector** | | | |
| n of contacts | Health service register (1) | SPECIALE | |
| Cost | Payment-agreements between Danish Regions and provider unions | - | Adjusted to 2023 level according to consumer price index (2). |
| **Hospital care** | | | |
| n and cost | DRG registries:<br>Drgshel2015-2017<br>drgphel2015-2017<br>drgpsyk_hel2018<br>drg_kontakter2021-2023<br>drsamb2015-2017<br>drgpamb2015-2017<br>drpsyk_amb2018 | v_totpris<br>totalpris_drg<br>v_pris pris_drg | Adjusted to 2023 level according to consumer price index (2). |
| **Filled prescriptions** | | | Not adjusted due to price irregularities distinct from typical price development over time. The irregular development in prices is primarily caused by patent expirations and bi-weekly price regulations related to government reimbursement. |
| n and cost | Prescriptions Registry (3) | AUP | |
| **Lost productivity, i.e. loss of earned income** | | | Adjusted to 2023 level according to consumer price index (2). |
| Income | Income registry (4) | erhvervsindk_13 | |

Abbreviations. n=number

References: 1. Sahl Andersen J, De Fine Olivarius N, Krasnik A. The Danish National Health Service Register. Scand J Public Health. 2011 Jul 20;39(7_suppl):34–7.
2. Statistics Denmark. PRIS8: Consumer price index, annual average (1900=100) by type [Internet, cited 2025 Apr 4]. Available from: https://www.statbank.dk/pris8.
3. Wallach Kildemoes H, Toft Sørensen H, Hallas J. The Danish national prescription registry. Scand J Public Health. 2011 Jul;39(7):38–41.
4. Baadsgaard M, Quitzau J. Danish registers on personal income and transfer payments. Scand J Public Health. 2011 Jul;39(7):103–5.

**Supplementary Table 4a Patients with prevalent any brain disorder, closest relatives identified and with at least one eligible comparator**

| Agegroup of index patients in matched population | Closest relative type identified | Available control with same type of closest relative |
|---|---|---|
| Children and young people (0-24 years)<br>n = 263,026 | Fathers<br>n = 131,149 (50%) | 95% |
| | Mothers<br>n = 100,583 (38%) | 95% |
| | No available closest relative<br>n = 89,843 (34%) | n/a |
| Adults (25-64 years)<br>n = 1,103,732 | Children<br>n = 290,975 (26%) | 95% |
| | Parents<br>n = 166,991 (15%) | 84% |
| | Partners<br>n = 365,434 (33%) | 95% |
| | Siblings<br>n = 172,504 (16%) | 65% |
| | Pooled<br>n = 995,904 (90%) | 88% |
| | No available closest relative<br>n = 107,828 (10%) | n/a |
| Older people (65+ years)<br>n = 566,848 | Children<br>n = 241,536 (43%) | 95% |
| | Partners<br>n = 148,676 (26%) | 94% |
| | Siblings<br>n = 18,066 (3%) | 49% |
| | Pooled<br>n = 408,278 (72%) | 92% |
| | No available closest relative<br>n = 171,819 (28%) | n/a |

**Supplementary Table 4b Patients with incident any brain disorder, closest relatives identified and with at least one eligible comparator**

| Agegroup of index patients in matched population | Closest relative type identified | Available control with same type of closest relative |
|---|---|---|
| Children and young people (0-24 years)<br>n = 163,194 | Fathers<br>n = 83,187 (51%) | 95% |
| | Mothers<br>n = 67,078 (41%) | 95% |
| | No available closest relative<br>n = 53,605 (33%) | n/a |
| Adults (25-64 years)<br>n = 233,850 | Children<br>n = 55,721 (24%) | 96% |
| | Parents<br>n = 32,805 (14%) | 86% |
| | Partners<br>n = 90,205 (39%) | 94% |
| | Siblings<br>n = 30,537 (13%) | 67% |
| | Pooled<br>n = 209,268 (89%) | 90% |
| | No available closest relative<br>n = 24,582 (11%) | n/a |
| Older people (65+ years)<br>n = 146,680 | Children<br>n = 65,680 (45%) | 95% |
| | Partners<br>n = 41,696 (28%) | 94% |
| | Siblings<br>n = 2,377 (2%) | 36% |
| | Pooled<br>n = 109,753 (75%) | 94% |
| | No available closest relative<br>n = 36,927 (25%) | n/a |

**Supplementary Table 5 Characteristics of included patients with any prevalent or incident brain disorder, by age patient strata**

| Characteristics by patient age strata | Patients with prevalent brain disorders | Patients with incident brain disorders |
|---|---|---|
| **Children and young people 0-24 years** | | |
| Number of persons, n | 221,649 | 143,364 |
| Male, n (%) | 118,396 (53.4) | 67,309 (46.9) |
| Age, median (Q1;Q3) | 18.3 (13.5;21.8) | 15.8 (10.0;20.2) |
| **Adults 25-64 years** | | |
| Number of persons, n | 880,661 | 188,475 |
| Male, n (%) | 371,473 (42.2) | 89,814 (47.7) |
| Age, median (Q1;Q3) | 47.6 (36.5;56.1) | 45.1 (34.3;54.3) |
| CCI, n (%) | | |
| 0 | 727,360 (82.6) | 161,677 (85.8) |
| 1-2 | 132,232 (15.0) | 22,787 (12.1) |
| +3 | 21,069 (2.4) | 4,011 (2.1) |
| Education level, n (%) | | |
| No/Unknown education | 19,866 (2.3) | 8,584 (4.6) |
| Primary | 194,916 (22.1) | 34,565 (18.3) |
| Secondary | 396,550 (45.0) | 84,171 (44.7) |
| Tertiary | 269,329 (30.6) | 61,155 (32.4) |
| **Older adults 65- years** | | |
| Number of persons, n | 378,826 | 102,748 |
| Male, n (%) | 156,364 (41.3) | 53,103 (51.7) |
| Age, median (Q1;Q3) | 74.7 (69.8;80.4) | 76.2 (71.3;82.2) |
| CCI, n (%) | | |
| 0 | 181,023 (47.8) | 54,130 (52.7) |
| 1-2 | 141,189 (37.3) | 33,708 (32.8) |
| +3 | 56,614 (14.9) | 14,910 (14.5) |
| Education level, n (%) | | |
| No/Unknown education | 20,881 (5.5) | 6,195 (6.0) |
| Primary | 111,561 (29.4) | 34,121 (33.2) |
| Secondary | 171,453 (45.3) | 45,543 (44.3) |
| Tertiary | 74,931 (19.8) | 16,889 (16.4) |

#### Supplementary Table 6
#### Alcohol use disorder - Incident cohort 2016-2021
#### Identification of populations of relatives

| ***Agegroup of index patients in matched population*** | | ***Closest relative type identified*** | | ***Available control with same type of closest relative*** |
|---|---|---|---|---|
| Children and young people (0-24 years) | n = 3,309 | Fathers | n = 2,264 (68%) | 95% |
| | | Mothers | n = 2,767 (84%) | 96% |
| | | No available closest relative | n = 308 (9%) | n/a |
| Young adults and adults (25-64 years) | n = 30,453 | Children | n = 5,471 (18%) | 92% |
| | | Parents | n = 5,194 (17%) | 88% |
| | | Partners | n = 13,934 (46%) | 95% |
| | | Siblings | n = 4,336 (14%) | 48% |
| | | Pooled | n = 28,935 (95%) | 86% |
| | | No available closest relative | n = 1,518 (5%) | n/a |
| Older people (65+ years) | n = 12,154 | Children | n = 4,908 (40%) | 92% |
| | | Partners | n = 5,175 (43%) | 95% |
| | | Siblings | n = 343 (3%) | 40% |
| | | Pooled | n = 10,426 (86%) | 92% |
| | | No available closest relative | n = 1,728 (14%) | n/a |

**Supplementary Table 7**
**Alcohol use disorder - Incident cohort 2016-2021**
**Patient characteristics - cases**

| *Age strata of index patients* | *Characteristic* | |
|---|---|---|
| Children and young people (0-24 years) | Number of persons, n | 2,882 (100.0) |
| | Male, n (%) | 1,821 (63.2) |
| | Age, median (Q1;Q3) | 21.5 (19.6;23.4) |
| | CCI, n (%) | |
| | 0 | 2,653 (92.1) |
| | 1-2 | 215 (7.5) |
| | +3 | 14 (0.5) |
| | Education level, n (%) | |
| | No/Unknown education | 35 (1.2) |
| | Primary | 2,120 (73.6) |
| | Secondary | 683 (23.7) |
| | Tertiary | 44 (1.5) |
| Young adults and adults (25-64 years) | Number of persons, n | 25,119 (100.0) |
| | Male, n (%) | 13,992 (55.7) |
| | Age, median (Q1;Q3) | 49.5 (39.7;56.9) |
| | CCI, n (%) | |
| | 0 | 19,766 (78.7) |
| | 1-2 | 4,523 (18.0) |
| | +3 | 830 (3.3) |
| | Education level, n (%) | |
| | No/Unknown education | 744 (3.0) |
| | Primary | 7,172 (28.6) |
| | Secondary | 12,183 (48.5) |
| | Tertiary | 5,020 (20.0) |
| Older people (65+ years) | Number of persons, n | 9,615 (100.0) |
| | Male, n (%) | 5,777 (60.1) |
| | Age, median (Q1;Q3) | 72.2 (68.4;76.6) |
| | CCI, n (%) | |
| | 0 | 4,582 (47.7) |
| | 1-2 | 3,454 (35.9) |
| | +3 | 1,579 (16.4) |
| | Education level, n (%) | |
| | No/Unknown education | 699 (7.3) |
| | Primary | 2,306 (24.0) |
| | Secondary | 4,770 (49.6) |
| | Tertiary | 1,840 (19.1) |

**Supplementary Table 8**
**Alcohol use disorder - Incident cohort 2016-2021**
**Patient characteristics - relatives - pooled**

| *Age strata of index patients* | *Closest relative type* | *Characteristic of family relatives by agestrata of index patients* | *Closest family relatives of brain disorder patients* | *Comparison relative* |
|---|---|---|---|---|
| Children and young people (0-24 years) | Fathers | Number of persons, n | 2,168 (100.0) | 2,168 (100.0) |
| | | Male, n (%) | 2,168 (100.0) | 2,168 (100.0) |
| | | Age, median (Q1;Q3) | 52.4 (48.4;56.1) | 52.4 (49.1;56.0) |
| | | CCI, n (%) | | |
| | | 0 | 1,826 (84.2) | 1,875 (86.5) |
| | | 1-2 | 300 (13.8) | 263 (12.1) |
| | | +3 | 42 (1.9) | 30 (1.4) |
| | | Education level, n (%) | | |
| | | No/Unknown education | 34 (1.6) | 34 (1.6) |
| | | Primary | 586 (27.0) | 586 (27.0) |
| | | Secondary | 1,111 (51.2) | 1,111 (51.2) |
| | | Tertiary | 437 (20.2) | 437 (20.2) |
| | | Relation to patient, n (%) | | |
| | | Father | 2,168 (100.0) | 2,168 (100.0) |
| Children and young people (0-24 years) | Mothers | Number of persons, n | 2,658 (100.0) | 2,658 (100.0) |
| | | Male, n (%) | 0 (0.0) | 0 (0.0) |
| | | Age, median (Q1;Q3) | 50.0 (46.2;53.6) | 50.2 (46.6;53.4) |
| | | CCI, n (%) | | |
| | | 0 | 2,217 (83.4) | 2,292 (86.2) |
| | | 1-2 | 402 (15.1) | 331 (12.5) |
| | | +3 | 39 (1.5) | 35 (1.3) |
| | | Education level, n (%) | | |
| | | No/Unknown education | 16 (0.6) | 16 (0.6) |
| | | Primary | 668 (25.1) | 668 (25.1) |
| | | Secondary | 1,282 (48.2) | 1,282 (48.2) |
| | | Tertiary | 692 (26.0) | 692 (26.0) |
| | | Relation to patient, n (%) | | |
| | | Mother | 2,658 (100.0) | 2,658 (100.0) |
| Young adults and adults (25-64 years) | Pooled | Number of persons, n | 25,119 (100.0) | 25,119 (100.0) |
| | | Male, n (%) | 11,235 (44.7) | 11,235 (44.7) |
| | | Age, median (Q1;Q3) | 50.0 (34.1;59.8) | 50.1 (34.0;59.7) |
| | | CCI, n (%) | | |
| | | 0 | 20,801 (82.8) | 21,153 (84.2) |
| | | 1-2 | 3,626 (14.4) | 3,387 (13.5) |
| | | +3 | 692 (2.8) | 579 (2.3) |
| | | Education level, n (%) | | |
| | | No/Unknown education | 1,522 (6.1) | 1,522 (6.1) |
| | | Primary | 7,094 (28.2) | 7,094 (28.2) |
| | | Secondary | 11,450 (45.6) | 11,450 (45.6) |
| | | Tertiary | 5,053 (20.1) | 5,053 (20.1) |
| | | Relation to patient, n (%) | | |
| | | Mother | 3,575 (14.2) | 3,575 (14.2) |
| | | Father | 1,025 (4.1) | 1,025 (4.1) |
| | | Partner | 13,339 (53.1) | 13,339 (53.1) |
| | | Child | 5,061 (20.1) | 5,061 (20.1) |

**Supplementary Table 8**
**Alcohol use disorder - Incident cohort 2016-2021**
**Patient characteristics - relatives - pooled**

| *Age strata of index patients* | *Closest relative type* | *Characteristic of family relatives by agestrata of index patients* | *Closest family relatives of brain disorder patients* | *Comparison relative* |
|---|---|---|---|---|
| | | Sibling | 2,119 (8.4) | 2,119 (8.4) |
| Older people (65+ years) | Pooled | Number of persons, n | 9,615 (100.0) | 9,615 (100.0) |
| | | Male, n (%) | 4,137 (43.0) | 4,137 (43.0) |
| | | Age, median (Q1;Q3) | 61.4 (48.3;71.2) | 61.8 (48.1;71.2) |
| | | CCI, n (%) | | |
| | | 0 | 7,179 (74.7) | 7,366 (76.6) |
| | | 1-2 | 1,917 (19.9) | 1,798 (18.7) |
| | | +3 | 519 (5.4) | 451 (4.7) |
| | | Education level, n (%) | | |
| | | No/Unknown education | 284 (3.0) | 284 (3.0) |
| | | Primary | 1,840 (19.1) | 1,840 (19.1) |
| | | Secondary | 4,665 (48.5) | 4,665 (48.5) |
| | | Tertiary | 2,826 (29.4) | 2,826 (29.4) |
| | | Relation to patient, n (%) | | |
| | | Partner | 4,948 (51.5) | 4,948 (51.5) |
| | | Child | 4,529 (47.1) | 4,529 (47.1) |
| | | Sibling | 138 (1.4) | 138 (1.4) |

**Supplementary Table 9**
**Alcohol use disorder - Incident cohort 2016-2021**
**Patient characteristics - relatives - not pooled**

| *Age strata of index patients* | *Closest relative type* | *Characteristic of family relatives by agestrata of index patients* | *Closest family relatives of brain disorder patients* | *Comparison relative* |
|---|---|---|---|---|
| Children and young people (0-24 years) | Fathers | Number of persons, n | 2,168 (100.0) | 2,168 (100.0) |
| | | Male, n (%) | 2,168 (100.0) | 2,168 (100.0) |
| | | Age, median (Q1;Q3) | 52.4 (48.4;56.1) | 52.4 (49.1;56.0) |
| | | CCI, n (%) | | |
| | | 0 | 1,826 (84.2) | 1,875 (86.5) |
| | | 1-2 | 300 (13.8) | 263 (12.1) |
| | | +3 | 42 (1.9) | 30 (1.4) |
| | | Education level, n (%) | | |
| | | No/Unknown education | 34 (1.6) | 34 (1.6) |
| | | Primary | 586 (27.0) | 586 (27.0) |
| | | Secondary | 1,111 (51.2) | 1,111 (51.2) |
| | | Tertiary | 437 (20.2) | 437 (20.2) |
| | | Relation to patient, n (%) | | |
| | | Father | 2,168 (100.0) | 2,168 (100.0) |
| Children and young people (0-24 years) | Mothers | Number of persons, n | 2,658 (100.0) | 2,658 (100.0) |
| | | Male, n (%) | 0 (0.0) | 0 (0.0) |
| | | Age, median (Q1;Q3) | 50.0 (46.2;53.6) | 50.2 (46.6;53.4) |
| | | CCI, n (%) | | |
| | | 0 | 2,217 (83.4) | 2,292 (86.2) |
| | | 1-2 | 402 (15.1) | 331 (12.5) |
| | | +3 | 39 (1.5) | 35 (1.3) |
| | | Education level, n (%) | | |
| | | No/Unknown education | 16 (0.6) | 16 (0.6) |
| | | Primary | 668 (25.1) | 668 (25.1) |
| | | Secondary | 1,282 (48.2) | 1,282 (48.2) |
| | | Tertiary | 692 (26.0) | 692 (26.0) |
| | | Relation to patient, n (%) | | |
| | | Mother | 2,658 (100.0) | 2,658 (100.0) |
| Young adults and adults (25-64 years) | Children | Number of persons, n | 5,061 (100.0) | 5,061 (100.0) |
| | | Male, n (%) | 2,617 (51.7) | 2,617 (51.7) |
| | | Age, median (Q1;Q3) | 21.9 (14.7;30.6) | 21.4 (14.7;30.3) |
| | | CCI, n (%) | | |
| | | 0 | 4,742 (93.7) | 4,739 (93.6) |
| | | 1-2 | 302 (6.0) | 315 (6.2) |
| | | +3 | 17 (0.3) | 7 (0.1) |
| | | Education level, n (%) | | |
| | | No/Unknown education | 1,051 (20.8) | 1,051 (20.8) |
| | | Primary | 2,098 (41.5) | 2,098 (41.5) |
| | | Secondary | 1,310 (25.9) | 1,310 (25.9) |
| | | Tertiary | 602 (11.9) | 602 (11.9) |
| | | Relation to patient, n (%) | | |
| | | Child | 5,061 (100.0) | 5,061 (100.0) |
| Young adults and adults (25-64 years) | Parents | Number of persons, n | 4,600 (100.0) | 4,600 (100.0) |
| | | Male, n (%) | 1,025 (22.3) | 1,025 (22.3) |
| | | Age, median (Q1;Q3) | 67.7 (59.5;75.4) | 67.8 (59.6;75.5) |

### Supplementary Table 9
### Alcohol use disorder - Incident cohort 2016-2021
### Patient characteristics - relatives - not pooled

| *Age strata of index patients* | *Closest relative type* | *Characteristic of family relatives by agestrata of index patients* | *Closest family relatives of brain disorder patients* | *Comparison relative* |
|---|---|---|---|---|
| | | CCI, n (%) | | |
| | | 0 | 3,009 (65.4) | 3,080 (67.0) |
| | | 1-2 | 1,216 (26.4) | 1,200 (26.1) |
| | | +3 | 375 (8.2) | 320 (7.0) |
| | | Education level, n (%) | | |
| | | No/Unknown education | 75 (1.6) | 75 (1.6) |
| | | Primary | 1,648 (35.8) | 1,648 (35.8) |
| | | Secondary | 2,142 (46.6) | 2,142 (46.6) |
| | | Tertiary | 735 (16.0) | 735 (16.0) |
| | | Relation to patient, n (%) | | |
| | | Mother | 3,575 (77.7) | 3,575 (77.7) |
| | | Father | 1,025 (22.3) | 1,025 (22.3) |
| Young adults and adults (25-64 years) | Partners | Number of persons, n | 13,339 (100.0) | 13,339 (100.0) |
| | | Male, n (%) | 6,275 (47.0) | 6,275 (47.0) |
| | | Age, median (Q1;Q3) | 50.2 (41.1;57.5) | 50.3 (41.3;57.5) |
| | | CCI, n (%) | | |
| | | 0 | 11,274 (84.5) | 11,530 (86.4) |
| | | 1-2 | 1,815 (13.6) | 1,597 (12.0) |
| | | +3 | 250 (1.9) | 212 (1.6) |
| | | Education level, n (%) | | |
| | | No/Unknown education | 384 (2.9) | 384 (2.9) |
| | | Primary | 2,769 (20.8) | 2,769 (20.8) |
| | | Secondary | 6,851 (51.4) | 6,851 (51.4) |
| | | Tertiary | 3,335 (25.0) | 3,335 (25.0) |
| | | Relation to patient, n (%) | | |
| | | Partner | 13,339 (100.0) | 13,339 (100.0) |
| Young adults and adults (25-64 years) | Siblings | Number of persons, n | 2,119 (100.0) | 2,119 (100.0) |
| | | Male, n (%) | 1,318 (62.2) | 1,318 (62.2) |
| | | Age, median (Q1;Q3) | 53.6 (40.6;59.3) | 53.3 (40.5;59.4) |
| | | CCI, n (%) | | |
| | | 0 | 1,776 (83.8) | 1,804 (85.1) |
| | | 1-2 | 293 (13.8) | 275 (13.0) |
| | | +3 | 50 (2.4) | 40 (1.9) |
| | | Education level, n (%) | | |
| | | No/Unknown education | 12 (0.6) | 12 (0.6) |
| | | Primary | 579 (27.3) | 579 (27.3) |
| | | Secondary | 1,147 (54.1) | 1,147 (54.1) |
| | | Tertiary | 381 (18.0) | 381 (18.0) |
| | | Relation to patient, n (%) | | |
| | | Sibling | 2,119 (100.0) | 2,119 (100.0) |
| Older people (65+ years) | Children | Number of persons, n | 4,529 (100.0) | 4,529 (100.0) |
| | | Male, n (%) | 2,305 (50.9) | 2,305 (50.9) |
| | | Age, median (Q1;Q3) | 47.6 (42.5;53.0) | 47.5 (42.4;52.8) |
| | | CCI, n (%) | | |
| | | 0 | 3,976 (87.8) | 4,020 (88.8) |

**Supplementary Table 9**
**Alcohol use disorder - Incident cohort 2016-2021**
**Patient characteristics - relatives - not pooled**

| *Age strata of index patients* | *Closest relative type* | *Characteristic of family relatives by agestrata of index patients* | *Closest family relatives of brain disorder patients* | *Comparison relative* |
|---|---|---|---|---|
| | | 1-2 | 494 (10.9) | 455 (10.0) |
| | | +3 | 59 (1.3) | 54 (1.2) |
| | | Education level, n (%) | | |
| | | No/Unknown education | 7 (0.2) | 7 (0.2) |
| | | Primary | 760 (16.8) | 760 (16.8) |
| | | Secondary | 2,175 (48.0) | 2,175 (48.0) |
| | | Tertiary | 1,587 (35.0) | 1,587 (35.0) |
| | | Relation to patient, n (%) | | |
| | | Child | 4,529 (100.0) | 4,529 (100.0) |
| Older people (65+ years) | Partners | Number of persons, n | 4,948 (100.0) | 4,948 (100.0) |
| | | Male, n (%) | 1,766 (35.7) | 1,766 (35.7) |
| | | Age, median (Q1;Q3) | 70.9 (66.4;75.2) | 71.0 (66.7;75.0) |
| | | CCI, n (%) | | |
| | | 0 | 3,099 (62.6) | 3,242 (65.5) |
| | | 1-2 | 1,390 (28.1) | 1,316 (26.6) |
| | | +3 | 459 (9.3) | 390 (7.9) |
| | | Education level, n (%) | | |
| | | No/Unknown education | 276 (5.6) | 276 (5.6) |
| | | Primary | 1,061 (21.4) | 1,061 (21.4) |
| | | Secondary | 2,411 (48.7) | 2,411 (48.7) |
| | | Tertiary | 1,200 (24.3) | 1,200 (24.3) |
| | | Relation to patient, n (%) | | |
| | | Partner | 4,948 (100.0) | 4,948 (100.0) |
| Older people (65+ years) | Siblings | Number of persons, n | 138 (100.0) | 138 (100.0) |
| | | Male, n (%) | 66 (47.8) | 66 (47.8) |
| | | Age, median (Q1;Q3) | 61.8 (59.0;64.3) | 62.4 (59.5;64.4) |
| | | CCI, n (%) | | |
| | | 0 | 104 (75.4) | 104 (75.4) |
| | | 1-2 | * | 27 (19.6) |
| | | +3 | * | 7 (5.1) |
| | | Education level, n (%) | | |
| | | No/Unknown education | * | * |
| | | Primary | * | * |
| | | Secondary | 79 (57.2) | 79 (57.2) |
| | | Tertiary | 39 (28.3) | 39 (28.3) |
| | | Relation to patient, n (%) | | |
| | | Sibling | 138 (100.0) | 138 (100.0) |

#### Supplementary Table 10
#### Alcohol use disorder - Incident cohort 2016-2021
#### Cost analysis - pooled

| | | | | *Actual costs* | | *Attributable costs* | |
|---|---|---|---|---|---|---|---|
| ***Age strata of index patients*** | ***Closest relative type*** | ***Number of index patients included in cost component calculations*** | ***Cost component*** | ***Total (million EUR)*** | ***Per person (EUR)*** | ***Total (million EUR)*** | ***Per person (EUR)*** |
| Children and young people (0-24 years) | Fathers | 2,168 | Primary sector | 0.5 | 245 | 0.0 | 9 |
| | | 2,168 | Hospital care | 4.4 | 2,015 | 1.2 | 532 |
| | | 2,168 | Filled prescriptions | 0.5 | 244 | -0.0 | -9 |
| | | 2,050 | Lost productivity (income loss) | . | . | 14.2 | 6,923 |
| Children and young people (0-24 years) | Mothers | 2,658 | Primary sector | 1.0 | 370 | 0.1 | 35 |
| | | 2,658 | Hospital care | 5.6 | 2,097 | 0.6 | 208 |
| | | 2,658 | Filled prescriptions | 0.7 | 280 | 0.1 | 44 |
| | | 2,628 | Lost productivity (income loss) | . | . | 21.6 | 8,213 |
| Young adults and adults (25-64 years) | Pooled | 25,119 | Primary sector | 7.9 | 317 | 0.5 | 18 |
| | | 25,119 | Hospital care | 56.0 | 2,240 | 8.9 | 354 |
| | | 25,119 | Filled prescriptions | 7.0 | 280 | 1.1 | 45 |
| | | 18,331 | Lost productivity (income loss) | . | . | 99.0 | 5,398 |
| Older people (65+ years) | Pooled | 9,615 | Primary sector | 3.8 | 400 | 0.1 | 14 |
| | | 9,615 | Hospital care | 27.7 | 2,906 | 2.6 | 272 |
| | | 9,615 | Filled prescriptions | 3.5 | 367 | 0.3 | 34 |
| | | 4,882 | Lost productivity (income loss) | . | . | 7.7 | 1,585 |

#### Supplementary Table 11
#### Alcohol use disorder - Incident cohort 2016-2021
#### Cost analysis - not pooled

| *Age strata of index patients* | *Closest relative type* | *Number of index patients included in cost component calculations* | *Cost component* | *Actual costs* | | *Attributable costs* | |
|---|---|---|---|---|---|---|---|
| | | | | *Total (million EUR)* | *Per person (EUR)* | *Total (million EUR)* | *Per person (EUR)* |
| Children and young people (0-24 years) | Fathers | 2,168 | Primary sector | 0.5 | 245 | 0.0 | 9 |
| | | 2,168 | Hospital care | 4.4 | 2,015 | 1.2 | 532 |
| | | 2,168 | Filled prescriptions | 0.5 | 244 | -0.0 | -9 |
| | | 2,050 | Lost productivity (income loss) | . | . | 14.2 | 6,923 |
| Children and young people (0-24 years) | Mothers | 2,658 | Primary sector | 1.0 | 370 | 0.1 | 35 |
| | | 2,658 | Hospital care | 5.6 | 2,097 | 0.6 | 208 |
| | | 2,658 | Filled prescriptions | 0.7 | 280 | 0.1 | 44 |
| | | 2,628 | Lost productivity (income loss) | . | . | 21.6 | 8,213 |
| Young adults and adults (25-64 years) | Children | 5,061 | Primary sector | 1.1 | 223 | 0.1 | 21 |
| | | 5,061 | Hospital care | 6.6 | 1,318 | 1.5 | 304 |
| | | 5,061 | Filled prescriptions | 0.6 | 118 | 0.1 | 18 |
| | | 3,084 | Lost productivity (income loss) | . | . | 4.9 | 1,586 |
| Young adults and adults (25-64 years) | Parents | 4,600 | Primary sector | 2.1 | 462 | 0.1 | 16 |
| | | 4,600 | Hospital care | 17.2 | 3,773 | 1.7 | 385 |
| | | 4,600 | Filled prescriptions | 2.6 | 563 | 0.4 | 78 |
| | | 1,569 | Lost productivity (income loss) | . | . | 10.4 | 6,648 |
| Young adults and adults (25-64 years) | Partners | 13,339 | Primary sector | 4.1 | 306 | 0.3 | 20 |
| | | 13,339 | Hospital care | 27.9 | 2,096 | 5.3 | 397 |
| | | 13,339 | Filled prescriptions | 3.3 | 249 | 0.6 | 49 |
| | | 11,843 | Lost productivity (income loss) | . | . | 74.4 | 6,283 |
| Young adults and adults (25-64 years) | Siblings | 2,119 | Primary sector | 0.6 | 295 | 0.0 | 11 |
| | | 2,119 | Hospital care | 4.3 | 2,043 | 0.3 | 141 |
| | | 2,119 | Filled prescriptions | 0.5 | 252 | 0.0 | 17 |
| | | 1,835 | Lost productivity (income loss) | . | . | 9.2 | 5,027 |
| Older people (65+ years) | Children | 4,529 | Primary sector | 1.3 | 289 | 0.0 | 9 |
| | | 4,529 | Hospital care | 7.1 | 1,576 | 0.2 | 34 |
| | | 4,529 | Filled prescriptions | 0.9 | 198 | 0.0 | 7 |
| | | 4,435 | Lost productivity (income loss) | . | . | 9.2 | 2,071 |
| Older people (65+ years) | Partners | 4,948 | Primary sector | 2.5 | 506 | 0.1 | 21 |
| | | 4,948 | Hospital care | 20.4 | 4,181 | 2.6 | 525 |
| | | 4,948 | Filled prescriptions | 2.6 | 526 | 0.3 | 62 |
| | | 374 | Lost productivity (income loss) | . | . | -0.9 | -2,338 |
| Older people (65+ years) | Siblings | 138 | Primary sector | 0.0 | 321 | -0.0 | -44 |
| | | 138 | Hospital care | 0.2 | 1,449 | -0.1 | -846 |
| | | 138 | Filled prescriptions | 0.0 | 304 | -0.0 | -63 |
| | | 73 | Lost productivity (income loss) | . | . | -0.6 | -7,813 |

#### Supplementary Table 12
#### Alcohol use disorder - Prevalent cohort 2021
#### Identification of populations of relatives

| *Agegroup of index patients in matched population* | | *Closest relative type identified* | | *Available control with same type of closest relative* |
|---|---|---|---|---|
| Children and young people (0-24 years) | n = 2,319 | Fathers | n = 1,527 (66%) | 95% |
| | | Mothers | n = 1,807 (78%) | 96% |
| | | No available closest relative | n = 297 (13%) | n/a |
| Young adults and adults (25-64 years) | n = 100,970 | Children | n = 21,886 (22%) | 91% |
| | | Parents | n = 18,898 (19%) | 87% |
| | | Partners | n = 36,747 (36%) | 94% |
| | | Siblings | n = 19,395 (19%) | 52% |
| | | Pooled | n = 96,926 (96%) | 84% |
| | | No available closest relative | n = 4,044 (4%) | n/a |
| Older people (65+ years) | n = 48,369 | Children | n = 20,237 (42%) | 91% |
| | | Partners | n = 18,528 (38%) | 95% |
| | | Siblings | n = 2,708 (6%) | 49% |
| | | Pooled | n = 41,473 (86%) | 90% |
| | | No available closest relative | n = 6,896 (14%) | n/a |

**Supplementary Table 13**
**Alcohol use disorder - Prevalent cohort 2021**
**Patient characteristics - cases**

| ***Age strata of index patients*** | ***Characteristic*** | |
|---|---|---|
| Children and young people (0-24 years) | Number of persons, n | 1,938 (100.0) |
| | Male, n (%) | 1,178 (60.8) |
| | Age, median (Q1;Q3) | 22.6 (20.8;24.0) |
| | CCI, n (%) | |
| | 0 | 1,760 (90.8) |
| | 1-2 | 164 (8.5) |
| | +3 | 14 (0.7) |
| | Education level, n (%) | |
| | No/Unknown education | 58 (3.0) |
| | Primary | 1,362 (70.3) |
| | Secondary | 486 (25.1) |
| | Tertiary | 32 (1.7) |
| Young adults and adults (25-64 years) | Number of persons, n | 81,444 (100.0) |
| | Male, n (%) | 54,855 (67.4) |
| | Age, median (Q1;Q3) | 52.6 (43.0;58.8) |
| | CCI, n (%) | |
| | 0 | 59,000 (72.4) |
| | 1-2 | 17,713 (21.7) |
| | +3 | 4,731 (5.8) |
| | Education level, n (%) | |
| | No/Unknown education | 2,070 (2.5) |
| | Primary | 29,126 (35.8) |
| | Secondary | 37,711 (46.3) |
| | Tertiary | 12,537 (15.4) |
| Older people (65+ years) | Number of persons, n | 37,612 (100.0) |
| | Male, n (%) | 23,024 (61.2) |
| | Age, median (Q1;Q3) | 72.1 (68.2;76.5) |
| | CCI, n (%) | |
| | 0 | 15,333 (40.8) |
| | 1-2 | 14,390 (38.3) |
| | +3 | 7,889 (21.0) |
| | Education level, n (%) | |
| | No/Unknown education | 2,475 (6.6) |
| | Primary | 9,507 (25.3) |
| | Secondary | 18,423 (49.0) |
| | Tertiary | 7,207 (19.2) |

**Supplementary Table 14**
**Alcohol use disorder - Prevalent cohort 2021**
**Patient characteristics - relatives - pooled**

| *Age strata of index patients* | *Closest relative type* | *Characteristic of family relatives by agestrata of index patients* | *Closest family relatives of brain disorder patients* | *Comparison relative* |
|---|---|---|---|---|
| Children and young people (0-24 years) | Fathers | Number of persons, n | 1,459 (100.0) | 1,459 (100.0) |
| | | Male, n (%) | 1,459 (100.0) | 1,459 (100.0) |
| | | Age, median (Q1;Q3) | 53.3 (49.3;57.2) | 53.6 (50.1;56.9) |
| | | CCI, n (%) | | |
| | | 0 | 1,193 (81.8) | 1,229 (84.2) |
| | | 1-2 | 227 (15.6) | 193 (13.2) |
| | | +3 | 39 (2.7) | 37 (2.5) |
| | | Education level, n (%) | | |
| | | No/Unknown education | 24 (1.6) | 24 (1.6) |
| | | Primary | 405 (27.8) | 405 (27.8) |
| | | Secondary | 739 (50.7) | 739 (50.7) |
| | | Tertiary | 291 (19.9) | 291 (19.9) |
| | | Relation to patient, n (%) | | |
| | | Father | 1,459 (100.0) | 1,459 (100.0) |
| Children and young people (0-24 years) | Mothers | Number of persons, n | 1,736 (100.0) | 1,736 (100.0) |
| | | Male, n (%) | 0 (0.0) | 0 (0.0) |
| | | Age, median (Q1;Q3) | 51.0 (46.7;55.0) | 51.1 (47.7;54.5) |
| | | CCI, n (%) | | |
| | | 0 | 1,414 (81.5) | 1,487 (85.7) |
| | | 1-2 | 303 (17.5) | 220 (12.7) |
| | | +3 | 19 (1.1) | 29 (1.7) |
| | | Education level, n (%) | | |
| | | No/Unknown education | 17 (1.0) | 17 (1.0) |
| | | Primary | 444 (25.6) | 444 (25.6) |
| | | Secondary | 801 (46.1) | 801 (46.1) |
| | | Tertiary | 474 (27.3) | 474 (27.3) |
| | | Relation to patient, n (%) | | |
| | | Mother | 1,736 (100.0) | 1,736 (100.0) |
| Young adults and adults (25-64 years) | Pooled | Number of persons, n | 81,444 (100.0) | 81,444 (100.0) |
| | | Male, n (%) | 30,905 (37.9) | 30,905 (37.9) |
| | | Age, median (Q1;Q3) | 51.5 (33.0;61.7) | 51.5 (32.9;61.6) |
| | | CCI, n (%) | | |
| | | 0 | 65,589 (80.5) | 67,496 (82.9) |
| | | 1-2 | 13,107 (16.1) | 11,622 (14.3) |
| | | +3 | 2,748 (3.4) | 2,326 (2.9) |
| | | Education level, n (%) | | |
| | | No/Unknown education | 3,864 (4.7) | 3,864 (4.7) |
| | | Primary | 25,609 (31.4) | 25,609 (31.4) |
| | | Secondary | 36,711 (45.1) | 36,711 (45.1) |
| | | Tertiary | 15,260 (18.7) | 15,260 (18.7) |
| | | Relation to patient, n (%) | | |
| | | Mother | 12,924 (15.9) | 12,924 (15.9) |
| | | Father | 3,637 (4.5) | 3,637 (4.5) |
| | | Partner | 34,731 (42.6) | 34,731 (42.6) |
| | | Child | 20,060 (24.6) | 20,060 (24.6) |

**Supplementary Table 14**
**Alcohol use disorder - Prevalent cohort 2021**
**Patient characteristics - relatives - pooled**

| *Age strata of index patients* | *Closest relative type* | *Characteristic of family relatives by agestrata of index patients* | *Closest family relatives of brain disorder patients* | *Comparison relative* |
|---|---|---|---|---|
| | | Sibling | 10,092 (12.4) | 10,092 (12.4) |
| Older people (65+ years) | Pooled | Number of persons, n | 37,612 (100.0) | 37,612 (100.0) |
| | | Male, n (%) | 14,939 (39.7) | 14,939 (39.7) |
| | | Age, median (Q1;Q3) | 60.0 (47.4;70.4) | 60.2 (47.2;70.5) |
| | | CCI, n (%) | | |
| | | 0 | 28,607 (76.1) | 29,185 (77.6) |
| | | 1-2 | 7,201 (19.1) | 6,818 (18.1) |
| | | +3 | 1,804 (4.8) | 1,609 (4.3) |
| | | Education level, n (%) | | |
| | | No/Unknown education | 964 (2.6) | 964 (2.6) |
| | | Primary | 7,266 (19.3) | 7,266 (19.3) |
| | | Secondary | 17,785 (47.3) | 17,785 (47.3) |
| | | Tertiary | 11,597 (30.8) | 11,597 (30.8) |
| | | Relation to patient, n (%) | | |
| | | Partner | 17,745 (47.2) | 17,745 (47.2) |
| | | Child | 18,530 (49.3) | 18,530 (49.3) |
| | | Sibling | 1,337 (3.6) | 1,337 (3.6) |

**Supplementary Table 15**
**Alcohol use disorder - Prevalent cohort 2021**
**Patient characteristics - relatives - not pooled**

| *Age strata of index patients* | *Closest relative type* | *Characteristic of family relatives by agestrata of index patients* | *Closest family relatives of brain disorder patients* | *Comparison relative* |
|---|---|---|---|---|
| Children and young people (0-24 years) | Fathers | Number of persons, n | 1,459 (100.0) | 1,459 (100.0) |
| | | Male, n (%) | 1,459 (100.0) | 1,459 (100.0) |
| | | Age, median (Q1;Q3) | 53.3 (49.3;57.2) | 53.6 (50.1;56.9) |
| | | CCI, n (%) | | |
| | | 0 | 1,193 (81.8) | 1,229 (84.2) |
| | | 1-2 | 227 (15.6) | 193 (13.2) |
| | | +3 | 39 (2.7) | 37 (2.5) |
| | | Education level, n (%) | | |
| | | No/Unknown education | 24 (1.6) | 24 (1.6) |
| | | Primary | 405 (27.8) | 405 (27.8) |
| | | Secondary | 739 (50.7) | 739 (50.7) |
| | | Tertiary | 291 (19.9) | 291 (19.9) |
| | | Relation to patient, n (%) | | |
| | | Father | 1,459 (100.0) | 1,459 (100.0) |
| Children and young people (0-24 years) | Mothers | Number of persons, n | 1,736 (100.0) | 1,736 (100.0) |
| | | Male, n (%) | 0 (0.0) | 0 (0.0) |
| | | Age, median (Q1;Q3) | 51.0 (46.7;55.0) | 51.1 (47.7;54.5) |
| | | CCI, n (%) | | |
| | | 0 | 1,414 (81.5) | 1,487 (85.7) |
| | | 1-2 | 303 (17.5) | 220 (12.7) |
| | | +3 | 19 (1.1) | 29 (1.7) |
| | | Education level, n (%) | | |
| | | No/Unknown education | 17 (1.0) | 17 (1.0) |
| | | Primary | 444 (25.6) | 444 (25.6) |
| | | Secondary | 801 (46.1) | 801 (46.1) |
| | | Tertiary | 474 (27.3) | 474 (27.3) |
| | | Relation to patient, n (%) | | |
| | | Mother | 1,736 (100.0) | 1,736 (100.0) |
| Young adults and adults (25-64 years) | Children | Number of persons, n | 20,060 (100.0) | 20,060 (100.0) |
| | | Male, n (%) | 10,480 (52.2) | 10,480 (52.2) |
| | | Age, median (Q1;Q3) | 25.1 (17.5;32.1) | 24.7 (17.1;31.6) |
| | | CCI, n (%) | | |
| | | 0 | 18,759 (93.5) | 18,869 (94.1) |
| | | 1-2 | 1,250 (6.2) | 1,141 (5.7) |
| | | +3 | 51 (0.3) | 50 (0.2) |
| | | Education level, n (%) | | |
| | | No/Unknown education | 2,920 (14.6) | 2,920 (14.6) |
| | | Primary | 8,295 (41.4) | 8,295 (41.4) |
| | | Secondary | 6,128 (30.5) | 6,128 (30.5) |
| | | Tertiary | 2,717 (13.5) | 2,717 (13.5) |
| | | Relation to patient, n (%) | | |
| | | Child | 20,060 (100.0) | 20,060 (100.0) |
| Young adults and adults (25-64 years) | Parents | Number of persons, n | 16,561 (100.0) | 16,561 (100.0) |
| | | Male, n (%) | 3,637 (22.0) | 3,637 (22.0) |
| | | Age, median (Q1;Q3) | 71.8 (63.4;78.1) | 72.0 (63.7;78.3) |

**Supplementary Table 15**
**Alcohol use disorder - Prevalent cohort 2021**
**Patient characteristics - relatives - not pooled**

| *Age strata of index patients* | *Closest relative type* | *Characteristic of family relatives by agestrata of index patients* | *Closest family relatives of brain disorder patients* | *Comparison relative* |
|---|---|---|---|---|
| | | CCI, n (%) | | |
| | | 0 | 9,930 (60.0) | 10,584 (63.9) |
| | | 1-2 | 5,019 (30.3) | 4,615 (27.9) |
| | | +3 | 1,612 (9.7) | 1,362 (8.2) |
| | | Education level, n (%) | | |
| | | No/Unknown education | 220 (1.3) | 220 (1.3) |
| | | Primary | 6,512 (39.3) | 6,512 (39.3) |
| | | Secondary | 7,465 (45.1) | 7,465 (45.1) |
| | | Tertiary | 2,364 (14.3) | 2,364 (14.3) |
| | | Relation to patient, n (%) | | |
| | | Mother | 12,924 (78.0) | 12,924 (78.0) |
| | | Father | 3,637 (22.0) | 3,637 (22.0) |
| Young adults and adults (25-64 years) | Partners | Number of persons, n | 34,731 (100.0) | 34,731 (100.0) |
| | | Male, n (%) | 10,846 (31.2) | 10,846 (31.2) |
| | | Age, median (Q1;Q3) | 51.6 (41.1;58.4) | 51.5 (41.3;58.3) |
| | | CCI, n (%) | | |
| | | 0 | 28,751 (82.8) | 29,789 (85.8) |
| | | 1-2 | 5,218 (15.0) | 4,324 (12.4) |
| | | +3 | 762 (2.2) | 618 (1.8) |
| | | Education level, n (%) | | |
| | | No/Unknown education | 699 (2.0) | 699 (2.0) |
| | | Primary | 8,186 (23.6) | 8,186 (23.6) |
| | | Secondary | 17,449 (50.2) | 17,449 (50.2) |
| | | Tertiary | 8,397 (24.2) | 8,397 (24.2) |
| | | Relation to patient, n (%) | | |
| | | Partner | 34,731 (100.0) | 34,731 (100.0) |
| Young adults and adults (25-64 years) | Siblings | Number of persons, n | 10,092 (100.0) | 10,092 (100.0) |
| | | Male, n (%) | 5,942 (58.9) | 5,942 (58.9) |
| | | Age, median (Q1;Q3) | 56.3 (49.0;61.3) | 56.5 (49.0;61.2) |
| | | CCI, n (%) | | |
| | | 0 | 8,149 (80.7) | 8,254 (81.8) |
| | | 1-2 | 1,620 (16.1) | 1,542 (15.3) |
| | | +3 | 323 (3.2) | 296 (2.9) |
| | | Education level, n (%) | | |
| | | No/Unknown education | 25 (0.2) | 25 (0.2) |
| | | Primary | 2,616 (25.9) | 2,616 (25.9) |
| | | Secondary | 5,669 (56.2) | 5,669 (56.2) |
| | | Tertiary | 1,782 (17.7) | 1,782 (17.7) |
| | | Relation to patient, n (%) | | |
| | | Sibling | 10,092 (100.0) | 10,092 (100.0) |
| Older people (65+ years) | Children | Number of persons, n | 18,530 (100.0) | 18,530 (100.0) |
| | | Male, n (%) | 9,036 (48.8) | 9,036 (48.8) |
| | | Age, median (Q1;Q3) | 47.2 (41.8;53.0) | 47.1 (41.7;52.7) |
| | | CCI, n (%) | | |
| | | 0 | 16,328 (88.1) | 16,359 (88.3) |

**Supplementary Table 15**
**Alcohol use disorder - Prevalent cohort 2021**
**Patient characteristics - relatives - not pooled**

| *Age strata of index patients* | *Closest relative type* | *Characteristic of family relatives by agestrata of index patients* | *Closest family relatives of brain disorder patients* | *Comparison relative* |
|---|---|---|---|---|
| | | 1-2 | 1,971 (10.6) | 1,917 (10.3) |
| | | +3 | 231 (1.2) | 254 (1.4) |
| | | Education level, n (%) | | |
| | | No/Unknown education | 24 (0.1) | 24 (0.1) |
| | | Primary | 3,087 (16.7) | 3,087 (16.7) |
| | | Secondary | 8,544 (46.1) | 8,544 (46.1) |
| | | Tertiary | 6,875 (37.1) | 6,875 (37.1) |
| | | Relation to patient, n (%) | | |
| | | Child | 18,530 (100.0) | 18,530 (100.0) |
| Older people (65+ years) | Partners | Number of persons, n | 17,745 (100.0) | 17,745 (100.0) |
| | | Male, n (%) | 5,190 (29.2) | 5,190 (29.2) |
| | | Age, median (Q1;Q3) | 70.9 (66.4;75.3) | 70.9 (66.6;75.1) |
| | | CCI, n (%) | | |
| | | 0 | 11,260 (63.5) | 11,796 (66.5) |
| | | 1-2 | 4,968 (28.0) | 4,648 (26.2) |
| | | +3 | 1,517 (8.5) | 1,301 (7.3) |
| | | Education level, n (%) | | |
| | | No/Unknown education | 920 (5.2) | 920 (5.2) |
| | | Primary | 3,932 (22.2) | 3,932 (22.2) |
| | | Secondary | 8,427 (47.5) | 8,427 (47.5) |
| | | Tertiary | 4,466 (25.2) | 4,466 (25.2) |
| | | Relation to patient, n (%) | | |
| | | Partner | 17,745 (100.0) | 17,745 (100.0) |
| Older people (65+ years) | Siblings | Number of persons, n | 1,337 (100.0) | 1,337 (100.0) |
| | | Male, n (%) | 713 (53.3) | 713 (53.3) |
| | | Age, median (Q1;Q3) | 62.2 (59.1;64.8) | 62.4 (59.4;64.8) |
| | | CCI, n (%) | | |
| | | 0 | 1,019 (76.2) | 1,030 (77.0) |
| | | 1-2 | 262 (19.6) | 253 (18.9) |
| | | +3 | 56 (4.2) | 54 (4.0) |
| | | Education level, n (%) | | |
| | | No/Unknown education | 20 (1.5) | 20 (1.5) |
| | | Primary | 247 (18.5) | 247 (18.5) |
| | | Secondary | 814 (60.9) | 814 (60.9) |
| | | Tertiary | 256 (19.1) | 256 (19.1) |
| | | Relation to patient, n (%) | | |
| | | Sibling | 1,337 (100.0) | 1,337 (100.0) |

#### Supplementary Table 16
#### Alcohol use disorder - Prevalent cohort 2021
#### Cost analysis - pooled

| Age strata of index patients | Closest relative type | Number of index patients included in cost component calculations | Cost component | Actual costs: Total (million EUR) | Actual costs: Per person (EUR) | Attributable costs: Total (million EUR) | Attributable costs: Per person (EUR) |
|---|---|---|---|---|---|---|---|
| Children and young people (0-24 years) | Fathers | 1,459 | Primary sector | 0.4 | 249 | -0.0 | -12 |
| | | 1,459 | Hospital care | 2.6 | 1,792 | 0.4 | 254 |
| | | 1,459 | Filled prescriptions | 0.4 | 259 | 0.0 | 5 |
| | | 1,362 | Lost productivity (income loss) | . | . | 9.3 | 6,860 |
| Children and young people (0-24 years) | Mothers | 1,736 | Primary sector | 0.7 | 391 | 0.1 | 46 |
| | | 1,736 | Hospital care | 4.3 | 2,475 | 0.8 | 466 |
| | | 1,736 | Filled prescriptions | 0.5 | 295 | 0.1 | 69 |
| | | 1,709 | Lost productivity (income loss) | . | . | 17.8 | 10,442 |
| Young adults and adults (25-64 years) | Pooled | 81,444 | Primary sector | 27.5 | 339 | 1.4 | 17 |
| | | 81,444 | Hospital care | 194.4 | 2,400 | 26.6 | 328 |
| | | 81,444 | Filled prescriptions | 25.6 | 316 | 3.8 | 47 |
| | | 56,929 | Lost productivity (income loss) | . | . | 315.2 | 5,537 |
| Older people (65+ years) | Pooled | 37,612 | Primary sector | 14.9 | 397 | 0.8 | 21 |
| | | 37,612 | Hospital care | 100.4 | 2,684 | 4.8 | 128 |
| | | 37,612 | Filled prescriptions | 13.5 | 361 | 1.3 | 34 |
| | | 20,476 | Lost productivity (income loss) | . | . | 47.9 | 2,341 |

**Supplementary Table 17**
**Alcohol use disorder - Prevalent cohort 2021**
**Cost analysis - not pooled**

| *Age strata of index patients* | *Closest relative type* | *Number of index patients included in cost component calculations* | *Cost component* | *Actual costs* | | *Attributable costs* | |
|---|---|---|---|---|---|---|---|
| | | | | *Total (million EUR)* | *Per person (EUR)* | *Total (million EUR)* | *Per person (EUR)* |
| Children and young people (0-24 years) | Fathers | 1,459 | Primary sector | 0.4 | 249 | -0.0 | -12 |
| | | 1,459 | Hospital care | 2.6 | 1,792 | 0.4 | 254 |
| | | 1,459 | Filled prescriptions | 0.4 | 259 | 0.0 | 5 |
| | | 1,362 | Lost productivity (income loss) | . | . | 9.3 | 6,860 |
| Children and young people (0-24 years) | Mothers | 1,736 | Primary sector | 0.7 | 391 | 0.1 | 46 |
| | | 1,736 | Hospital care | 4.3 | 2,475 | 0.8 | 466 |
| | | 1,736 | Filled prescriptions | 0.5 | 295 | 0.1 | 69 |
| | | 1,709 | Lost productivity (income loss) | . | . | 17.8 | 10,442 |
| Young adults and adults (25-64 years) | Children | 20,060 | Primary sector | 4.9 | 243 | 0.5 | 27 |
| | | 20,060 | Hospital care | 31.9 | 1,592 | 6.4 | 318 |
| | | 20,060 | Filled prescriptions | 2.6 | 131 | 0.3 | 15 |
| | | 13,784 | Lost productivity (income loss) | . | . | 28.8 | 2,086 |
| Young adults and adults (25-64 years) | Parents | 16,561 | Primary sector | 8.0 | 494 | 0.4 | 23 |
| | | 16,561 | Hospital care | 65.6 | 4,033 | 6.0 | 368 |
| | | 16,561 | Filled prescriptions | 9.8 | 601 | 1.0 | 60 |
| | | 3,808 | Lost productivity (income loss) | . | . | 23.9 | 6,272 |
| Young adults and adults (25-64 years) | Partners | 34,731 | Primary sector | 11.4 | 330 | 0.4 | 11 |
| | | 34,731 | Hospital care | 74.2 | 2,139 | 13.1 | 377 |
| | | 34,731 | Filled prescriptions | 10.3 | 297 | 2.6 | 75 |
| | | 31,108 | Lost productivity (income loss) | . | . | 226.5 | 7,280 |
| Young adults and adults (25-64 years) | Siblings | 10,092 | Primary sector | 3.1 | 313 | 0.1 | 9 |
| | | 10,092 | Hospital care | 22.8 | 2,265 | 1.1 | 112 |
| | | 10,092 | Filled prescriptions | 2.9 | 289 | -0.1 | -9 |
| | | 8,229 | Lost productivity (income loss) | . | . | 36.1 | 4,389 |
| Older people (65+ years) | Children | 18,530 | Primary sector | 5.5 | 298 | 0.2 | 13 |
| | | 18,530 | Hospital care | 30.9 | 1,669 | -1.3 | -71 |
| | | 18,530 | Filled prescriptions | 3.9 | 209 | -0.0 | -0 |
| | | 18,099 | Lost productivity (income loss) | . | . | 47.3 | 2,613 |
| Older people (65+ years) | Partners | 17,745 | Primary sector | 8.8 | 502 | 0.5 | 28 |
| | | 17,745 | Hospital care | 65.9 | 3,754 | 5.3 | 302 |
| | | 17,745 | Filled prescriptions | 9.2 | 522 | 1.2 | 69 |
| | | 1,607 | Lost productivity (income loss) | . | . | -3.4 | -2,089 |
| Older people (65+ years) | Siblings | 1,337 | Primary sector | 0.5 | 397 | 0.1 | 45 |
| | | 1,337 | Hospital care | 3.6 | 2,678 | 0.8 | 581 |
| | | 1,337 | Filled prescriptions | 0.5 | 345 | 0.1 | 42 |
| | | 770 | Lost productivity (income loss) | . | . | 4.0 | 5,186 |

**Supplementary Table 18**
**Anxiety disorders - Incident cohort 2016-2021**
**Identification of populations of relatives**

| ***Agegroup of index patients in matched population*** | | ***Closest relative type identified*** | | ***Available control with same type of closest relative*** |
|---|---|---|---|---|
| Children and young people (0-24 years) | n = 31,219 | Fathers | n = 24,599 (79%) | 96% |
| | | Mothers | n = 23,853 (76%) | 96% |
| | | No available closest relative | n = 2,585 (8%) | n/a |
| Young adults and adults (25-64 years) | n = 59,259 | Children | n = 9,358 (16%) | 94% |
| | | Parents | n = 11,161 (19%) | 91% |
| | | Partners | n = 28,223 (48%) | 95% |
| | | Siblings | n = 7,604 (13%) | 47% |
| | | Pooled | n = 56,346 (95%) | 88% |
| | | No available closest relative | n = 2,913 (5%) | n/a |
| Older people (65+ years) | n = 22,199 | Children | n = 10,352 (47%) | 95% |
| | | Partners | n = 8,631 (39%) | 96% |
| | | Siblings | n = 360 (2%) | 31% |
| | | Pooled | n = 19,343 (87%) | 94% |
| | | No available closest relative | n = 2,856 (13%) | n/a |

**Supplementary Table 19**
**Anxiety disorders - Incident cohort 2016-2021**
**Patient characteristics - cases**

| *Age strata of index patients* | *Characteristic* | |
|---|---|---|
| Children and young people (0-24 years) | Number of persons, n | 27,688 (100.0) |
| | Male, n (%) | 9,621 (34.7) |
| | Age, median (Q1;Q3) | 19.2 (16.0;22.1) |
| | CCI, n (%) | |
| | 0 | 25,694 (92.8) |
| | 1-2 | 1,939 (7.0) |
| | +3 | 55 (0.2) |
| | Education level, n (%) | |
| | No/Unknown education | 2,831 (10.2) |
| | Primary | 18,601 (67.2) |
| | Secondary | 5,793 (20.9) |
| | Tertiary | 463 (1.7) |
| Young adults and adults (25-64 years) | Number of persons, n | 49,620 (100.0) |
| | Male, n (%) | 18,548 (37.4) |
| | Age, median (Q1;Q3) | 41.6 (31.8;52.2) |
| | CCI, n (%) | |
| | 0 | 40,957 (82.5) |
| | 1-2 | 7,095 (14.3) |
| | +3 | 1,568 (3.2) |
| | Education level, n (%) | |
| | No/Unknown education | 1,949 (3.9) |
| | Primary | 13,160 (26.5) |
| | Secondary | 20,653 (41.6) |
| | Tertiary | 13,858 (27.9) |
| Older people (65+ years) | Number of persons, n | 18,309 (100.0) |
| | Male, n (%) | 6,036 (33.0) |
| | Age, median (Q1;Q3) | 75.4 (70.5;81.5) |
| | CCI, n (%) | |
| | 0 | 7,072 (38.6) |
| | 1-2 | 6,945 (37.9) |
| | +3 | 4,292 (23.4) |
| | Education level, n (%) | |
| | No/Unknown education | 881 (4.8) |
| | Primary | 6,750 (36.9) |
| | Secondary | 7,855 (42.9) |
| | Tertiary | 2,823 (15.4) |

**Supplementary Table 20**
**Anxiety disorders - Incident cohort 2016-2021**
**Patient characteristics - relatives - pooled**

| *Age strata of index patients* | *Closest relative type* | *Characteristic of family relatives by agestrata of index patients* | *Closest family relatives of brain disorder patients* | *Comparison relative* |
|---|---|---|---|---|
| Children and young people (0-24 years) | Fathers | Number of persons, n | 23,756 (100.0) | 23,756 (100.0) |
| | | Male, n (%) | 23,756 (100.0) | 23,756 (100.0) |
| | | Age, median (Q1;Q3) | 51.0 (46.7;55.2) | 51.0 (47.1;54.8) |
| | | CCI, n (%) | | |
| | | 0 | 20,527 (86.4) | 20,913 (88.0) |
| | | 1-2 | 2,830 (11.9) | 2,522 (10.6) |
| | | +3 | 399 (1.7) | 321 (1.4) |
| | | Education level, n (%) | | |
| | | No/Unknown education | 286 (1.2) | 286 (1.2) |
| | | Primary | 4,855 (20.4) | 4,855 (20.4) |
| | | Secondary | 12,403 (52.2) | 12,403 (52.2) |
| | | Tertiary | 6,212 (26.1) | 6,212 (26.1) |
| | | Relation to patient, n (%) | | |
| | | Father | 23,756 (100.0) | 23,756 (100.0) |
| Children and young people (0-24 years) | Mothers | Number of persons, n | 23,067 (100.0) | 23,067 (100.0) |
| | | Male, n (%) | 0 (0.0) | 0 (0.0) |
| | | Age, median (Q1;Q3) | 48.9 (44.9;52.8) | 48.9 (45.2;52.5) |
| | | CCI, n (%) | | |
| | | 0 | 19,723 (85.5) | 20,176 (87.5) |
| | | 1-2 | 3,049 (13.2) | 2,645 (11.5) |
| | | +3 | 295 (1.3) | 246 (1.1) |
| | | Education level, n (%) | | |
| | | No/Unknown education | 169 (0.7) | 169 (0.7) |
| | | Primary | 3,777 (16.4) | 3,777 (16.4) |
| | | Secondary | 10,324 (44.8) | 10,324 (44.8) |
| | | Tertiary | 8,797 (38.1) | 8,797 (38.1) |
| | | Relation to patient, n (%) | | |
| | | Mother | 23,067 (100.0) | 23,067 (100.0) |
| Young adults and adults (25-64 years) | Pooled | Number of persons, n | 49,620 (100.0) | 49,620 (100.0) |
| | | Male, n (%) | 27,654 (55.7) | 27,654 (55.7) |
| | | Age, median (Q1;Q3) | 44.4 (30.6;57.4) | 44.5 (30.7;57.5) |
| | | CCI, n (%) | | |
| | | 0 | 42,136 (84.9) | 43,012 (86.7) |
| | | 1-2 | 6,387 (12.9) | 5,632 (11.4) |
| | | +3 | 1,097 (2.2) | 976 (2.0) |
| | | Education level, n (%) | | |
| | | No/Unknown education | 4,272 (8.6) | 4,272 (8.6) |
| | | Primary | 12,494 (25.2) | 12,494 (25.2) |
| | | Secondary | 21,272 (42.9) | 21,272 (42.9) |
| | | Tertiary | 11,582 (23.3) | 11,582 (23.3) |
| | | Relation to patient, n (%) | | |
| | | Mother | 7,545 (15.2) | 7,545 (15.2) |
| | | Father | 2,683 (5.4) | 2,683 (5.4) |
| | | Partner | 26,980 (54.4) | 26,980 (54.4) |
| | | Child | 8,818 (17.8) | 8,818 (17.8) |

#### Supplementary Table 20
#### Anxiety disorders - Incident cohort 2016-2021
#### Patient characteristics - relatives - pooled

| *Age strata of index patients* | *Closest relative type* | *Characteristic of family relatives by agestrata of index patients* | *Closest family relatives of brain disorder patients* | *Comparison relative* |
|---|---|---|---|---|
| | | Sibling | 3,594 (7.2) | 3,594 (7.2) |
| Older people (65+ years) | Pooled | Number of persons, n | 18,309 (100.0) | 18,309 (100.0) |
| | | Male, n (%) | 10,176 (55.6) | 10,176 (55.6) |
| | | Age, median (Q1;Q3) | 61.8 (51.8;72.7) | 61.6 (51.7;72.6) |
| | | CCI, n (%) | | |
| | | 0 | 13,249 (72.4) | 13,478 (73.6) |
| | | 1-2 | 3,922 (21.4) | 3,781 (20.7) |
| | | +3 | 1,138 (6.2) | 1,050 (5.7) |
| | | Education level, n (%) | | |
| | | No/Unknown education | 564 (3.1) | 564 (3.1) |
| | | Primary | 3,863 (21.1) | 3,863 (21.1) |
| | | Secondary | 9,282 (50.7) | 9,282 (50.7) |
| | | Tertiary | 4,600 (25.1) | 4,600 (25.1) |
| | | Relation to patient, n (%) | | |
| | | Partner | 8,313 (45.4) | 8,313 (45.4) |
| | | Child | 9,881 (54.0) | 9,881 (54.0) |
| | | Sibling | 115 (0.6) | 115 (0.6) |

**Supplementary Table 21**
**Anxiety disorders - Incident cohort 2016-2021**
**Patient characteristics - relatives - not pooled**

| *Age strata of index patients* | *Closest relative type* | *Characteristic of family relatives by agestrata of index patients* | *Closest family relatives of brain disorder patients* | *Comparison relative* |
|---|---|---|---|---|
| Children and young people (0-24 years) | Fathers | Number of persons, n | 23,756 (100.0) | 23,756 (100.0) |
| | | Male, n (%) | 23,756 (100.0) | 23,756 (100.0) |
| | | Age, median (Q1;Q3) | 51.0 (46.7;55.2) | 51.0 (47.1;54.8) |
| | | CCI, n (%) | | |
| | | 0 | 20,527 (86.4) | 20,913 (88.0) |
| | | 1-2 | 2,830 (11.9) | 2,522 (10.6) |
| | | +3 | 399 (1.7) | 321 (1.4) |
| | | Education level, n (%) | | |
| | | No/Unknown education | 286 (1.2) | 286 (1.2) |
| | | Primary | 4,855 (20.4) | 4,855 (20.4) |
| | | Secondary | 12,403 (52.2) | 12,403 (52.2) |
| | | Tertiary | 6,212 (26.1) | 6,212 (26.1) |
| | | Relation to patient, n (%) | | |
| | | Father | 23,756 (100.0) | 23,756 (100.0) |
| Children and young people (0-24 years) | Mothers | Number of persons, n | 23,067 (100.0) | 23,067 (100.0) |
| | | Male, n (%) | 0 (0.0) | 0 (0.0) |
| | | Age, median (Q1;Q3) | 48.9 (44.9;52.8) | 48.9 (45.2;52.5) |
| | | CCI, n (%) | | |
| | | 0 | 19,723 (85.5) | 20,176 (87.5) |
| | | 1-2 | 3,049 (13.2) | 2,645 (11.5) |
| | | +3 | 295 (1.3) | 246 (1.1) |
| | | Education level, n (%) | | |
| | | No/Unknown education | 169 (0.7) | 169 (0.7) |
| | | Primary | 3,777 (16.4) | 3,777 (16.4) |
| | | Secondary | 10,324 (44.8) | 10,324 (44.8) |
| | | Tertiary | 8,797 (38.1) | 8,797 (38.1) |
| | | Relation to patient, n (%) | | |
| | | Mother | 23,067 (100.0) | 23,067 (100.0) |
| Young adults and adults (25-64 years) | Children | Number of persons, n | 8,818 (100.0) | 8,818 (100.0) |
| | | Male, n (%) | 4,669 (52.9) | 4,669 (52.9) |
| | | Age, median (Q1;Q3) | 18.1 (10.4;27.1) | 17.7 (9.8;26.4) |
| | | CCI, n (%) | | |
| | | 0 | 8,221 (93.2) | 8,333 (94.5) |
| | | 1-2 | 572 (6.5) | 464 (5.3) |
| | | +3 | 25 (0.3) | 21 (0.2) |
| | | Education level, n (%) | | |
| | | No/Unknown education | 2,990 (33.9) | 2,990 (33.9) |
| | | Primary | 3,331 (37.8) | 3,331 (37.8) |
| | | Secondary | 1,716 (19.5) | 1,716 (19.5) |
| | | Tertiary | 781 (8.9) | 781 (8.9) |
| | | Relation to patient, n (%) | | |
| | | Child | 8,818 (100.0) | 8,818 (100.0) |
| Young adults and adults (25-64 years) | Parents | Number of persons, n | 10,228 (100.0) | 10,228 (100.0) |
| | | Male, n (%) | 2,683 (26.2) | 2,683 (26.2) |
| | | Age, median (Q1;Q3) | 61.9 (56.2;69.9) | 61.8 (56.5;69.9) |

**Supplementary Table 21**
**Anxiety disorders - Incident cohort 2016-2021**
**Patient characteristics - relatives - not pooled**

| *Age strata of index patients* | *Closest relative type* | *Characteristic of family relatives by agestrata of index patients* | *Closest family relatives of brain disorder patients* | *Comparison relative* |
|---|---|---|---|---|
| | | CCI, n (%) | | |
| | | 0 | 7,230 (70.7) | 7,450 (72.8) |
| | | 1-2 | 2,396 (23.4) | 2,243 (21.9) |
| | | +3 | 602 (5.9) | 535 (5.2) |
| | | Education level, n (%) | | |
| | | No/Unknown education | 204 (2.0) | 204 (2.0) |
| | | Primary | 2,843 (27.8) | 2,843 (27.8) |
| | | Secondary | 4,900 (47.9) | 4,900 (47.9) |
| | | Tertiary | 2,281 (22.3) | 2,281 (22.3) |
| | | Relation to patient, n (%) | | |
| | | Mother | 7,545 (73.8) | 7,545 (73.8) |
| | | Father | 2,683 (26.2) | 2,683 (26.2) |
| Young adults and adults (25-64 years) | Partners | Number of persons, n | 26,980 (100.0) | 26,980 (100.0) |
| | | Male, n (%) | 18,271 (67.7) | 18,271 (67.7) |
| | | Age, median (Q1;Q3) | 43.3 (34.1;53.6) | 43.4 (34.1;53.7) |
| | | CCI, n (%) | | |
| | | 0 | 23,590 (87.4) | 24,088 (89.3) |
| | | 1-2 | 2,984 (11.1) | 2,541 (9.4) |
| | | +3 | 406 (1.5) | 351 (1.3) |
| | | Education level, n (%) | | |
| | | No/Unknown education | 1,066 (4.0) | 1,066 (4.0) |
| | | Primary | 5,368 (19.9) | 5,368 (19.9) |
| | | Secondary | 12,776 (47.4) | 12,776 (47.4) |
| | | Tertiary | 7,770 (28.8) | 7,770 (28.8) |
| | | Relation to patient, n (%) | | |
| | | Partner | 26,980 (100.0) | 26,980 (100.0) |
| Young adults and adults (25-64 years) | Siblings | Number of persons, n | 3,594 (100.0) | 3,594 (100.0) |
| | | Male, n (%) | 2,031 (56.5) | 2,031 (56.5) |
| | | Age, median (Q1;Q3) | 45.0 (29.7;56.5) | 45.1 (29.6;56.5) |
| | | CCI, n (%) | | |
| | | 0 | 3,095 (86.1) | 3,141 (87.4) |
| | | 1-2 | 435 (12.1) | 384 (10.7) |
| | | +3 | 64 (1.8) | 69 (1.9) |
| | | Education level, n (%) | | |
| | | No/Unknown education | 12 (0.3) | 12 (0.3) |
| | | Primary | 952 (26.5) | 952 (26.5) |
| | | Secondary | 1,880 (52.3) | 1,880 (52.3) |
| | | Tertiary | 750 (20.9) | 750 (20.9) |
| | | Relation to patient, n (%) | | |
| | | Sibling | 3,594 (100.0) | 3,594 (100.0) |
| Older people (65+ years) | Children | Number of persons, n | 9,881 (100.0) | 9,881 (100.0) |
| | | Male, n (%) | 5,135 (52.0) | 5,135 (52.0) |
| | | Age, median (Q1;Q3) | 52.6 (46.7;58.1) | 52.5 (46.7;57.8) |
| | | CCI, n (%) | | |
| | | 0 | 8,368 (84.7) | 8,472 (85.7) |

**Supplementary Table 21**
**Anxiety disorders - Incident cohort 2016-2021**
**Patient characteristics - relatives - not pooled**

| *Age strata of index patients* | *Closest relative type* | *Characteristic of family relatives by agestrata of index patients* | *Closest family relatives of brain disorder patients* | *Comparison relative* |
|---|---|---|---|---|
| | | 1-2 | 1,305 (13.2) | 1,223 (12.4) |
| | | +3 | 208 (2.1) | 186 (1.9) |
| | | Education level, n (%) | | |
| | | No/Unknown education | 38 (0.4) | 38 (0.4) |
| | | Primary | 1,624 (16.4) | 1,624 (16.4) |
| | | Secondary | 5,165 (52.3) | 5,165 (52.3) |
| | | Tertiary | 3,054 (30.9) | 3,054 (30.9) |
| | | Relation to patient, n (%) | | |
| | | Child | 9,881 (100.0) | 9,881 (100.0) |
| Older people (65+ years) | Partners | Number of persons, n | 8,313 (100.0) | 8,313 (100.0) |
| | | Male, n (%) | 4,973 (59.8) | 4,973 (59.8) |
| | | Age, median (Q1;Q3) | 73.6 (69.1;78.5) | 73.4 (69.2;78.5) |
| | | CCI, n (%) | | |
| | | 0 | 4,802 (57.8) | 4,918 (59.2) |
| | | 1-2 | 2,590 (31.2) | 2,536 (30.5) |
| | | +3 | 921 (11.1) | 859 (10.3) |
| | | Education level, n (%) | | |
| | | No/Unknown education | 526 (6.3) | 526 (6.3) |
| | | Primary | 2,224 (26.8) | 2,224 (26.8) |
| | | Secondary | 4,041 (48.6) | 4,041 (48.6) |
| | | Tertiary | 1,522 (18.3) | 1,522 (18.3) |
| | | Relation to patient, n (%) | | |
| | | Partner | 8,313 (100.0) | 8,313 (100.0) |
| Older people (65+ years) | Siblings | Number of persons, n | 115 (100.0) | 115 (100.0) |
| | | Male, n (%) | 68 (59.1) | 68 (59.1) |
| | | Age, median (Q1;Q3) | 62.6 (59.2;65.3) | 62.2 (59.2;64.6) |
| | | CCI, n (%) | | |
| | | 0 | 79 (68.7) | 88 (76.5) |
| | | 1-2 | 27 (23.5) | 22 (19.1) |
| | | +3 | 9 (7.8) | 5 (4.3) |
| | | Education level, n (%) | | |
| | | No/Unknown education | 0 (0.0) | 0 (0.0) |
| | | Primary | 15 (13.0) | 15 (13.0) |
| | | Secondary | 76 (66.1) | 76 (66.1) |
| | | Tertiary | 24 (20.9) | 24 (20.9) |
| | | Relation to patient, n (%) | | |
| | | Sibling | 115 (100.0) | 115 (100.0) |

#### Supplementary Table 22
#### Anxiety disorders - Incident cohort 2016-2021
#### Cost analysis - pooled

| *Age strata of index patients* | *Closest relative type* | *Number of index patients included in cost component calculations* | *Cost component* | *Actual costs* | | *Attributable costs* | |
|---|---|---|---|---|---|---|---|
| | | | | *Total (million EUR)* | *Per person (EUR)* | *Total (million EUR)* | *Per person (EUR)* |
| Children and young people (0-24 years) | Fathers | 23,756 | Primary sector | 5.6 | 237 | 0.4 | 16 |
| | | 23,756 | Hospital care | 37.8 | 1,595 | 3.2 | 135 |
| | | 23,756 | Filled prescriptions | 5.0 | 213 | 1.0 | 41 |
| | | 22,758 | Lost productivity (income loss) | . | . | 159.3 | 6,999 |
| Children and young people (0-24 years) | Mothers | 23,067 | Primary sector | 8.5 | 368 | 1.1 | 50 |
| | | 23,067 | Hospital care | 42.2 | 1,829 | 6.4 | 278 |
| | | 23,067 | Filled prescriptions | 5.5 | 238 | 1.4 | 60 |
| | | 22,861 | Lost productivity (income loss) | . | . | 106.4 | 4,652 |
| Young adults and adults (25-64 years) | Pooled | 49,620 | Primary sector | 14.7 | 298 | 1.5 | 31 |
| | | 49,620 | Hospital care | 96.2 | 1,947 | 12.4 | 252 |
| | | 49,620 | Filled prescriptions | 11.4 | 230 | 1.5 | 31 |
| | | 37,451 | Lost productivity (income loss) | . | . | 187.7 | 5,012 |
| Older people (65+ years) | Pooled | 18,309 | Primary sector | 7.3 | 401 | 0.4 | 25 |
| | | 18,309 | Hospital care | 54.4 | 3,001 | 2.9 | 162 |
| | | 18,309 | Filled prescriptions | 7.0 | 386 | 0.6 | 35 |
| | | 9,311 | Lost productivity (income loss) | . | . | 40.6 | 4,365 |

**Supplementary Table 23**
**Anxiety disorders - Incident cohort 2016-2021**
**Cost analysis - not pooled**

| *Age strata of index patients* | *Closest relative type* | *Number of index patients included in cost component calculations* | *Cost component* | *Actual costs* | | *Attributable costs* | |
|---|---|---|---|---|---|---|---|
| | | | | *Total (million EUR)* | *Per person (EUR)* | *Total (million EUR)* | *Per person (EUR)* |
| Children and young people (0-24 years) | Fathers | 23,756 | Primary sector | 5.6 | 237 | 0.4 | 16 |
| | | 23,756 | Hospital care | 37.8 | 1,595 | 3.2 | 135 |
| | | 23,756 | Filled prescriptions | 5.0 | 213 | 1.0 | 41 |
| | | 22,758 | Lost productivity (income loss) | . | . | 159.3 | 6,999 |
| Children and young people (0-24 years) | Mothers | 23,067 | Primary sector | 8.5 | 368 | 1.1 | 50 |
| | | 23,067 | Hospital care | 42.2 | 1,829 | 6.4 | 278 |
| | | 23,067 | Filled prescriptions | 5.5 | 238 | 1.4 | 60 |
| | | 22,861 | Lost productivity (income loss) | . | . | 106.4 | 4,652 |
| Young adults and adults (25-64 years) | Children | 8,818 | Primary sector | 1.8 | 208 | 0.2 | 24 |
| | | 8,818 | Hospital care | 10.7 | 1,216 | 1.9 | 217 |
| | | 8,818 | Filled prescriptions | 0.9 | 106 | 0.1 | 16 |
| | | 4,148 | Lost productivity (income loss) | . | . | 13.4 | 3,222 |
| Young adults and adults (25-64 years) | Parents | 10,228 | Primary sector | 4.3 | 426 | 0.3 | 26 |
| | | 10,228 | Hospital care | 31.5 | 3,100 | 3.3 | 322 |
| | | 10,228 | Filled prescriptions | 4.4 | 436 | 0.4 | 44 |
| | | 5,248 | Lost productivity (income loss) | . | . | 26.5 | 5,047 |
| Young adults and adults (25-64 years) | Partners | 26,980 | Primary sector | 7.6 | 282 | 1.0 | 39 |
| | | 26,980 | Hospital care | 46.7 | 1,735 | 6.4 | 238 |
| | | 26,980 | Filled prescriptions | 5.2 | 194 | 0.9 | 35 |
| | | 24,839 | Lost productivity (income loss) | . | . | 140.4 | 5,653 |
| Young adults and adults (25-64 years) | Siblings | 3,594 | Primary sector | 1.0 | 283 | 0.0 | 12 |
| | | 3,594 | Hospital care | 7.4 | 2,067 | 0.9 | 242 |
| | | 3,594 | Filled prescriptions | 0.8 | 218 | 0.0 | 2 |
| | | 3,216 | Lost productivity (income loss) | . | . | 7.4 | 2,310 |
| Older people (65+ years) | Children | 9,881 | Primary sector | 3.0 | 305 | 0.2 | 19 |
| | | 9,881 | Hospital care | 18.1 | 1,831 | 0.7 | 76 |
| | | 9,881 | Filled prescriptions | 2.5 | 253 | 0.4 | 39 |
| | | 8,943 | Lost productivity (income loss) | . | . | 41.6 | 4,652 |
| Older people (65+ years) | Partners | 8,313 | Primary sector | 4.2 | 519 | 0.3 | 32 |
| | | 8,313 | Hospital care | 36.0 | 4,411 | 2.1 | 260 |
| | | 8,313 | Filled prescriptions | 4.5 | 548 | 0.2 | 29 |
| | | 310 | Lost productivity (income loss) | . | . | -0.9 | -2,810 |
| Older people (65+ years) | Siblings | 115 | Primary sector | 0.0 | 314 | -0.0 | -36 |
| | | 115 | Hospital care | 0.4 | 3,159 | 0.1 | 627 |
| | | 115 | Filled prescriptions | 0.0 | 374 | 0.0 | 101 |
| | | 58 | Lost productivity (income loss) | . | . | -0.1 | -1,493 |

**Supplementary Table 24**
**Anxiety disorders - Prevalent cohort 2021**
**Identification of populations of relatives**

| *Agegroup of index patients in matched population* | | *Closest relative type identified* | | *Available control with same type of closest relative* |
|---|---|---|---|---|
| Children and young people (0-24 years) | n = 29,299 | Fathers | n = 23,186 (79%) | 96% |
| | | Mothers | n = 22,222 (76%) | 96% |
| | | No available closest relative | n = 2,210 (8%) | n/a |
| Young adults and adults (25-64 years) | n = 209,296 | Children | n = 36,538 (17%) | 94% |
| | | Parents | n = 41,419 (20%) | 90% |
| | | Partners | n = 96,630 (46%) | 95% |
| | | Siblings | n = 28,693 (14%) | 48% |
| | | Pooled | n = 203,280 (97%) | 87% |
| | | No available closest relative | n = 6,016 (3%) | n/a |
| Older people (65+ years) | n = 67,137 | Children | n = 27,926 (42%) | 94% |
| | | Partners | n = 28,232 (42%) | 96% |
| | | Siblings | n = 2,420 (4%) | 49% |
| | | Pooled | n = 58,578 (87%) | 93% |
| | | No available closest relative | n = 8,559 (13%) | n/a |

**Supplementary Table 25**
**Anxiety disorders - Prevalent cohort 2021**
**Patient characteristics - cases**

| ***Age strata of index patients*** | ***Characteristic*** | |
|---|---|---|
| Children and young people (0-24 years) | Number of persons, n | 26,236 (100.0) |
| | Male, n (%) | 9,888 (37.7) |
| | Age, median (Q1;Q3) | 20.9 (18.1;23.1) |
| | CCI, n (%) | |
| | 0 | 24,238 (92.4) |
| | 1-2 | 1,931 (7.4) |
| | +3 | 67 (0.3) |
| | Education level, n (%) | |
| | No/Unknown education | 1,173 (4.5) |
| | Primary | 18,071 (68.9) |
| | Secondary | 6,444 (24.6) |
| | Tertiary | 548 (2.1) |
| Young adults and adults (25-64 years) | Number of persons, n | 178,115 (100.0) |
| | Male, n (%) | 62,333 (35.0) |
| | Age, median (Q1;Q3) | 45.6 (35.3;54.7) |
| | CCI, n (%) | |
| | 0 | 146,397 (82.2) |
| | 1-2 | 27,330 (15.3) |
| | +3 | 4,388 (2.5) |
| | Education level, n (%) | |
| | No/Unknown education | 4,014 (2.3) |
| | Primary | 51,155 (28.7) |
| | Secondary | 74,041 (41.6) |
| | Tertiary | 48,905 (27.5) |
| Older people (65+ years) | Number of persons, n | 54,851 (100.0) |
| | Male, n (%) | 16,404 (29.9) |
| | Age, median (Q1;Q3) | 73.9 (69.1;79.8) |
| | CCI, n (%) | |
| | 0 | 26,434 (48.2) |
| | 1-2 | 20,117 (36.7) |
| | +3 | 8,300 (15.1) |
| | Education level, n (%) | |
| | No/Unknown education | 2,380 (4.3) |
| | Primary | 18,543 (33.8) |
| | Secondary | 24,091 (43.9) |
| | Tertiary | 9,837 (17.9) |

**Supplementary Table 26**
**Anxiety disorders - Prevalent cohort 2021**
**Patient characteristics - relatives - pooled**

| *Age strata of index patients* | *Closest relative type* | *Characteristic of family relatives by agestrata of index patients* | *Closest family relatives of brain disorder patients* | *Comparison relative* |
|---|---|---|---|---|
| Children and young people (0-24 years) | Fathers | Number of persons, n | 22,420 (100.0) | 22,420 (100.0) |
| | | Male, n (%) | 22,420 (100.0) | 22,420 (100.0) |
| | | Age, median (Q1;Q3) | 52.7 (48.5;56.5) | 52.6 (48.8;56.2) |
| | | CCI, n (%) | | |
| | | 0 | 19,089 (85.1) | 19,423 (86.6) |
| | | 1-2 | 2,888 (12.9) | 2,664 (11.9) |
| | | +3 | 443 (2.0) | 333 (1.5) |
| | | Education level, n (%) | | |
| | | No/Unknown education | 198 (0.9) | 198 (0.9) |
| | | Primary | 4,382 (19.5) | 4,382 (19.5) |
| | | Secondary | 11,846 (52.8) | 11,846 (52.8) |
| | | Tertiary | 5,994 (26.7) | 5,994 (26.7) |
| | | Relation to patient, n (%) | | |
| | | Father | 22,420 (100.0) | 22,420 (100.0) |
| Children and young people (0-24 years) | Mothers | Number of persons, n | 21,500 (100.0) | 21,500 (100.0) |
| | | Male, n (%) | 0 (0.0) | 0 (0.0) |
| | | Age, median (Q1;Q3) | 50.5 (46.8;54.4) | 50.6 (47.0;54.0) |
| | | CCI, n (%) | | |
| | | 0 | 18,181 (84.6) | 18,775 (87.3) |
| | | 1-2 | 3,012 (14.0) | 2,478 (11.5) |
| | | +3 | 307 (1.4) | 247 (1.1) |
| | | Education level, n (%) | | |
| | | No/Unknown education | 117 (0.5) | 117 (0.5) |
| | | Primary | 3,287 (15.3) | 3,287 (15.3) |
| | | Secondary | 9,715 (45.2) | 9,715 (45.2) |
| | | Tertiary | 8,381 (39.0) | 8,381 (39.0) |
| | | Relation to patient, n (%) | | |
| | | Mother | 21,500 (100.0) | 21,500 (100.0) |
| Young adults and adults (25-64 years) | Pooled | Number of persons, n | 178,115 (100.0) | 178,115 (100.0) |
| | | Male, n (%) | 100,959 (56.7) | 100,959 (56.7) |
| | | Age, median (Q1;Q3) | 48.0 (32.2;59.6) | 48.1 (32.2;59.6) |
| | | CCI, n (%) | | |
| | | 0 | 149,175 (83.8) | 152,042 (85.4) |
| | | 1-2 | 24,438 (13.7) | 22,029 (12.4) |
| | | +3 | 4,502 (2.5) | 4,044 (2.3) |
| | | Education level, n (%) | | |
| | | No/Unknown education | 11,985 (6.7) | 11,985 (6.7) |
| | | Primary | 46,022 (25.8) | 46,022 (25.8) |
| | | Secondary | 78,871 (44.3) | 78,871 (44.3) |
| | | Tertiary | 41,237 (23.2) | 41,237 (23.2) |
| | | Relation to patient, n (%) | | |
| | | Mother | 27,395 (15.4) | 27,395 (15.4) |
| | | Father | 9,887 (5.6) | 9,887 (5.6) |
| | | Partner | 92,556 (52.0) | 92,556 (52.0) |
| | | Child | 34,379 (19.3) | 34,379 (19.3) |

**Supplementary Table 26**
**Anxiety disorders - Prevalent cohort 2021**
**Patient characteristics - relatives - pooled**

| *Age strata of index patients* | *Closest relative type* | *Characteristic of family relatives by agestrata of index patients* | *Closest family relatives of brain disorder patients* | *Comparison relative* |
|---|---|---|---|---|
| | | Sibling | 13,898 (7.8) | 13,898 (7.8) |
| Older people (65+ years) | Pooled | Number of persons, n | 54,851 (100.0) | 54,851 (100.0) |
| | | Male, n (%) | 31,490 (57.4) | 31,490 (57.4) |
| | | Age, median (Q1;Q3) | 63.7 (52.2;73.0) | 63.8 (51.9;73.0) |
| | | CCI, n (%) | | |
| | | 0 | 39,337 (71.7) | 40,048 (73.0) |
| | | 1-2 | 12,047 (22.0) | 11,603 (21.2) |
| | | +3 | 3,467 (6.3) | 3,200 (5.8) |
| | | Education level, n (%) | | |
| | | No/Unknown education | 1,644 (3.0) | 1,644 (3.0) |
| | | Primary | 11,265 (20.5) | 11,265 (20.5) |
| | | Secondary | 27,434 (50.0) | 27,434 (50.0) |
| | | Tertiary | 14,508 (26.4) | 14,508 (26.4) |
| | | Relation to patient, n (%) | | |
| | | Partner | 27,151 (49.5) | 27,151 (49.5) |
| | | Child | 26,505 (48.3) | 26,505 (48.3) |
| | | Sibling | 1,195 (2.2) | 1,195 (2.2) |

**Supplementary Table 27**
**Anxiety disorders - Prevalent cohort 2021**
**Patient characteristics - relatives - not pooled**

| *Age strata of index patients* | *Closest relative type* | *Characteristic of family relatives by agestrata of index patients* | *Closest family relatives of brain disorder patients* | *Comparison relative* |
|---|---|---|---|---|
| Children and young people (0-24 years) | Fathers | Number of persons, n | 22,420 (100.0) | 22,420 (100.0) |
| | | Male, n (%) | 22,420 (100.0) | 22,420 (100.0) |
| | | Age, median (Q1;Q3) | 52.7 (48.5;56.5) | 52.6 (48.8;56.2) |
| | | CCI, n (%) | | |
| | | 0 | 19,089 (85.1) | 19,423 (86.6) |
| | | 1-2 | 2,888 (12.9) | 2,664 (11.9) |
| | | +3 | 443 (2.0) | 333 (1.5) |
| | | Education level, n (%) | | |
| | | No/Unknown education | 198 (0.9) | 198 (0.9) |
| | | Primary | 4,382 (19.5) | 4,382 (19.5) |
| | | Secondary | 11,846 (52.8) | 11,846 (52.8) |
| | | Tertiary | 5,994 (26.7) | 5,994 (26.7) |
| | | Relation to patient, n (%) | | |
| | | Father | 22,420 (100.0) | 22,420 (100.0) |
| Children and young people (0-24 years) | Mothers | Number of persons, n | 21,500 (100.0) | 21,500 (100.0) |
| | | Male, n (%) | 0 (0.0) | 0 (0.0) |
| | | Age, median (Q1;Q3) | 50.5 (46.8;54.4) | 50.6 (47.0;54.0) |
| | | CCI, n (%) | | |
| | | 0 | 18,181 (84.6) | 18,775 (87.3) |
| | | 1-2 | 3,012 (14.0) | 2,478 (11.5) |
| | | +3 | 307 (1.4) | 247 (1.1) |
| | | Education level, n (%) | | |
| | | No/Unknown education | 117 (0.5) | 117 (0.5) |
| | | Primary | 3,287 (15.3) | 3,287 (15.3) |
| | | Secondary | 9,715 (45.2) | 9,715 (45.2) |
| | | Tertiary | 8,381 (39.0) | 8,381 (39.0) |
| | | Relation to patient, n (%) | | |
| | | Mother | 21,500 (100.0) | 21,500 (100.0) |
| Young adults and adults (25-64 years) | Children | Number of persons, n | 34,379 (100.0) | 34,379 (100.0) |
| | | Male, n (%) | 18,271 (53.1) | 18,271 (53.1) |
| | | Age, median (Q1;Q3) | 19.9 (12.8;28.4) | 19.5 (12.5;27.9) |
| | | CCI, n (%) | | |
| | | 0 | 32,114 (93.4) | 32,504 (94.5) |
| | | 1-2 | 2,197 (6.4) | 1,802 (5.2) |
| | | +3 | 68 (0.2) | 73 (0.2) |
| | | Education level, n (%) | | |
| | | No/Unknown education | 9,027 (26.3) | 9,027 (26.3) |
| | | Primary | 14,032 (40.8) | 14,032 (40.8) |
| | | Secondary | 7,766 (22.6) | 7,766 (22.6) |
| | | Tertiary | 3,554 (10.3) | 3,554 (10.3) |
| | | Relation to patient, n (%) | | |
| | | Child | 34,379 (100.0) | 34,379 (100.0) |
| Young adults and adults (25-64 years) | Parents | Number of persons, n | 37,282 (100.0) | 37,282 (100.0) |
| | | Male, n (%) | 9,887 (26.5) | 9,887 (26.5) |
| | | Age, median (Q1;Q3) | 65.6 (58.9;73.6) | 65.5 (59.0;73.7) |

### Supplementary Table 27
### Anxiety disorders - Prevalent cohort 2021
### Patient characteristics - relatives - not pooled

| *Age strata of index patients* | *Closest relative type* | *Characteristic of family relatives by agestrata of index patients* | *Closest family relatives of brain disorder patients* | *Comparison relative* |
|---|---|---|---|---|
| | | CCI, n (%) | | |
| | | 0 | 25,354 (68.0) | 26,022 (69.8) |
| | | 1-2 | 9,403 (25.2) | 8,893 (23.9) |
| | | +3 | 2,525 (6.8) | 2,367 (6.3) |
| | | Education level, n (%) | | |
| | | No/Unknown education | 615 (1.6) | 615 (1.6) |
| | | Primary | 10,636 (28.5) | 10,636 (28.5) |
| | | Secondary | 18,166 (48.7) | 18,166 (48.7) |
| | | Tertiary | 7,865 (21.1) | 7,865 (21.1) |
| | | Relation to patient, n (%) | | |
| | | Mother | 27,395 (73.5) | 27,395 (73.5) |
| | | Father | 9,887 (26.5) | 9,887 (26.5) |
| Young adults and adults (25-64 years) | Partners | Number of persons, n | 92,556 (100.0) | 92,556 (100.0) |
| | | Male, n (%) | 65,027 (70.3) | 65,027 (70.3) |
| | | Age, median (Q1;Q3) | 47.0 (37.1;55.9) | 47.1 (37.1;55.9) |
| | | CCI, n (%) | | |
| | | 0 | 80,102 (86.5) | 81,757 (88.3) |
| | | 1-2 | 10,899 (11.8) | 9,498 (10.3) |
| | | +3 | 1,555 (1.7) | 1,301 (1.4) |
| | | Education level, n (%) | | |
| | | No/Unknown education | 2,295 (2.5) | 2,295 (2.5) |
| | | Primary | 17,946 (19.4) | 17,946 (19.4) |
| | | Secondary | 45,590 (49.3) | 45,590 (49.3) |
| | | Tertiary | 26,725 (28.9) | 26,725 (28.9) |
| | | Relation to patient, n (%) | | |
| | | Partner | 92,556 (100.0) | 92,556 (100.0) |
| Young adults and adults (25-64 years) | Siblings | Number of persons, n | 13,898 (100.0) | 13,898 (100.0) |
| | | Male, n (%) | 7,774 (55.9) | 7,774 (55.9) |
| | | Age, median (Q1;Q3) | 52.2 (35.7;59.3) | 52.2 (35.6;59.3) |
| | | CCI, n (%) | | |
| | | 0 | 11,605 (83.5) | 11,759 (84.6) |
| | | 1-2 | 1,939 (14.0) | 1,836 (13.2) |
| | | +3 | 354 (2.5) | 303 (2.2) |
| | | Education level, n (%) | | |
| | | No/Unknown education | 48 (0.3) | 48 (0.3) |
| | | Primary | 3,408 (24.5) | 3,408 (24.5) |
| | | Secondary | 7,349 (52.9) | 7,349 (52.9) |
| | | Tertiary | 3,093 (22.3) | 3,093 (22.3) |
| | | Relation to patient, n (%) | | |
| | | Sibling | 13,898 (100.0) | 13,898 (100.0) |
| Older people (65+ years) | Children | Number of persons, n | 26,505 (100.0) | 26,505 (100.0) |
| | | Male, n (%) | 13,770 (52.0) | 13,770 (52.0) |
| | | Age, median (Q1;Q3) | 51.8 (45.4;57.9) | 51.5 (45.3;57.7) |
| | | CCI, n (%) | | |
| | | 0 | 22,583 (85.2) | 22,726 (85.7) |

**Supplementary Table 27**
**Anxiety disorders - Prevalent cohort 2021**
**Patient characteristics - relatives - not pooled**

| *Age strata of index patients* | *Closest relative type* | *Characteristic of family relatives by agestrata of index patients* | *Closest family relatives of brain disorder patients* | *Comparison relative* |
|---|---|---|---|---|
| | | 1-2 | 3,407 (12.9) | 3,289 (12.4) |
| | | +3 | 515 (1.9) | 490 (1.8) |
| | | Education level, n (%) | | |
| | | No/Unknown education | 69 (0.3) | 69 (0.3) |
| | | Primary | 4,476 (16.9) | 4,476 (16.9) |
| | | Secondary | 13,083 (49.4) | 13,083 (49.4) |
| | | Tertiary | 8,877 (33.5) | 8,877 (33.5) |
| | | Relation to patient, n (%) | | |
| | | Child | 26,505 (100.0) | 26,505 (100.0) |
| Older people (65+ years) | Partners | Number of persons, n | 27,151 (100.0) | 27,151 (100.0) |
| | | Male, n (%) | 17,108 (63.0) | 17,108 (63.0) |
| | | Age, median (Q1;Q3) | 73.1 (68.5;77.6) | 73.0 (68.7;77.5) |
| | | CCI, n (%) | | |
| | | 0 | 15,891 (58.5) | 16,408 (60.4) |
| | | 1-2 | 8,371 (30.8) | 8,076 (29.7) |
| | | +3 | 2,889 (10.6) | 2,667 (9.8) |
| | | Education level, n (%) | | |
| | | No/Unknown education | 1,571 (5.8) | 1,571 (5.8) |
| | | Primary | 6,578 (24.2) | 6,578 (24.2) |
| | | Secondary | 13,633 (50.2) | 13,633 (50.2) |
| | | Tertiary | 5,369 (19.8) | 5,369 (19.8) |
| | | Relation to patient, n (%) | | |
| | | Partner | 27,151 (100.0) | 27,151 (100.0) |
| Older people (65+ years) | Siblings | Number of persons, n | 1,195 (100.0) | 1,195 (100.0) |
| | | Male, n (%) | 612 (51.2) | 612 (51.2) |
| | | Age, median (Q1;Q3) | 62.5 (59.7;65.2) | 62.6 (60.0;64.8) |
| | | CCI, n (%) | | |
| | | 0 | 863 (72.2) | 914 (76.5) |
| | | 1-2 | 269 (22.5) | 238 (19.9) |
| | | +3 | 63 (5.3) | 43 (3.6) |
| | | Education level, n (%) | | |
| | | No/Unknown education | * | * |
| | | Primary | * | * |
| | | Secondary | 718 (60.1) | 718 (60.1) |
| | | Tertiary | 262 (21.9) | 262 (21.9) |
| | | Relation to patient, n (%) | | |
| | | Sibling | 1,195 (100.0) | 1,195 (100.0) |

**Supplementary Table 28**
**Anxiety disorders - Prevalent cohort 2021**
**Cost analysis - pooled**

| *Age strata of index patients* | *Closest relative type* | *Number of index patients included in cost component calculations* | *Cost component* | *Actual costs* | | *Attributable costs* | |
|---|---|---|---|---|---|---|---|
| | | | | *Total (million EUR)* | *Per person (EUR)* | *Total (million EUR)* | *Per person (EUR)* |
| Children and young people (0-24 years) | Fathers | 22,420 | Primary sector | 5.7 | 253 | 0.5 | 22 |
| | | 22,420 | Hospital care | 40.3 | 1,800 | 7.0 | 313 |
| | | 22,420 | Filled prescriptions | 5.1 | 228 | 1.0 | 46 |
| | | 21,367 | Lost productivity (income loss) | . | . | 174.6 | 8,171 |
| Children and young people (0-24 years) | Mothers | 21,500 | Primary sector | 8.3 | 387 | 1.3 | 59 |
| | | 21,500 | Hospital care | 40.6 | 1,890 | 7.0 | 326 |
| | | 21,500 | Filled prescriptions | 5.6 | 263 | 1.8 | 82 |
| | | 21,284 | Lost productivity (income loss) | . | . | 129.0 | 6,062 |
| Young adults and adults (25-64 years) | Pooled | 178,115 | Primary sector | 53.9 | 304 | 4.8 | 27 |
| | | 178,115 | Hospital care | 349.3 | 1,969 | 38.1 | 215 |
| | | 178,115 | Filled prescriptions | 45.8 | 258 | 6.9 | 39 |
| | | 129,008 | Lost productivity (income loss) | . | . | 790.6 | 6,129 |
| Older people (65+ years) | Pooled | 54,851 | Primary sector | 22.6 | 415 | 1.7 | 30 |
| | | 54,851 | Hospital care | 167.2 | 3,074 | 14.4 | 265 |
| | | 54,851 | Filled prescriptions | 22.1 | 407 | 2.4 | 44 |
| | | 26,037 | Lost productivity (income loss) | . | . | 93.7 | 3,600 |

#### Supplementary Table 29
#### Anxiety disorders - Prevalent cohort 2021
#### Cost analysis - not pooled

| *Age strata of index patients* | *Closest relative type* | *Number of index patients included in cost component calculations* | *Cost component* | *Actual costs* | | *Attributable costs* | |
|---|---|---|---|---|---|---|---|
| | | | | *Total (million EUR)* | *Per person (EUR)* | *Total (million EUR)* | *Per person (EUR)* |
| Children and young people (0-24 years) | Fathers | 22,420 | Primary sector | 5.7 | 253 | 0.5 | 22 |
| | | 22,420 | Hospital care | 40.3 | 1,800 | 7.0 | 313 |
| | | 22,420 | Filled prescriptions | 5.1 | 228 | 1.0 | 46 |
| | | 21,367 | Lost productivity (income loss) | . | . | 174.6 | 8,171 |
| Children and young people (0-24 years) | Mothers | 21,500 | Primary sector | 8.3 | 387 | 1.3 | 59 |
| | | 21,500 | Hospital care | 40.6 | 1,890 | 7.0 | 326 |
| | | 21,500 | Filled prescriptions | 5.6 | 263 | 1.8 | 82 |
| | | 21,284 | Lost productivity (income loss) | . | . | 129.0 | 6,062 |
| Young adults and adults (25-64 years) | Children | 34,379 | Primary sector | 7.5 | 219 | 1.0 | 30 |
| | | 34,379 | Hospital care | 44.5 | 1,299 | 7.9 | 231 |
| | | 34,379 | Filled prescriptions | 3.8 | 112 | 0.3 | 9 |
| | | 17,786 | Lost productivity (income loss) | . | . | 48.1 | 2,706 |
| Young adults and adults (25-64 years) | Parents | 37,282 | Primary sector | 16.9 | 458 | 1.2 | 33 |
| | | 37,282 | Hospital care | 123.4 | 3,346 | 12.4 | 336 |
| | | 37,282 | Filled prescriptions | 18.2 | 493 | 2.6 | 71 |
| | | 14,769 | Lost productivity (income loss) | . | . | 64.5 | 4,365 |
| Young adults and adults (25-64 years) | Partners | 92,556 | Primary sector | 25.2 | 273 | 2.2 | 24 |
| | | 92,556 | Hospital care | 151.6 | 1,641 | 15.3 | 165 |
| | | 92,556 | Filled prescriptions | 20.2 | 218 | 3.6 | 38 |
| | | 84,549 | Lost productivity (income loss) | . | . | 620.4 | 7,338 |
| Young adults and adults (25-64 years) | Siblings | 13,898 | Primary sector | 4.3 | 310 | 0.4 | 28 |
| | | 13,898 | Hospital care | 29.7 | 2,145 | 2.6 | 186 |
| | | 13,898 | Filled prescriptions | 3.6 | 262 | 0.4 | 32 |
| | | 11,904 | Lost productivity (income loss) | . | . | 57.6 | 4,841 |
| Older people (65+ years) | Children | 26,505 | Primary sector | 8.4 | 316 | 0.8 | 31 |
| | | 26,505 | Hospital care | 47.7 | 1,805 | 3.1 | 118 |
| | | 26,505 | Filled prescriptions | 6.6 | 251 | 1.0 | 38 |
| | | 24,022 | Lost productivity (income loss) | . | . | 94.9 | 3,950 |
| Older people (65+ years) | Partners | 27,151 | Primary sector | 13.7 | 513 | 0.8 | 30 |
| | | 27,151 | Hospital care | 116.4 | 4,349 | 11.0 | 410 |
| | | 27,151 | Filled prescriptions | 15.0 | 562 | 1.3 | 50 |
| | | 1,353 | Lost productivity (income loss) | . | . | -0.2 | -117 |
| Older people (65+ years) | Siblings | 1,195 | Primary sector | 0.5 | 399 | 0.1 | 44 |
| | | 1,195 | Hospital care | 3.1 | 2,611 | 0.3 | 282 |
| | | 1,195 | Filled prescriptions | 0.4 | 378 | 0.1 | 62 |
| | | 662 | Lost productivity (income loss) | . | . | -1.0 | -1,501 |

**Supplementary Table 30**
**Bipolar disorder - Incident cohort 2016-2021**
**Identification of populations of relatives**

| *Agegroup of index patients in matched population* | | *Closest relative type identified* | | *Available control with same type of closest relative* |
|---|---|---|---|---|
| Children and young people (0-24 years) | n = 2,167 | Fathers | n = 1,713 (79%) | 95% |
| | | Mothers | n = 1,823 (84%) | 95% |
| | | No available closest relative | n = 163 (8%) | n/a |
| Young adults and adults (25-64 years) | n = 7,144 | Children | n = 1,054 (15%) | 94% |
| | | Parents | n = 1,581 (22%) | 91% |
| | | Partners | n = 3,186 (45%) | 95% |
| | | Siblings | n = 1,019 (14%) | 40% |
| | | Pooled | n = 6,840 (96%) | 86% |
| | | No available closest relative | n = 304 (4%) | n/a |
| Older people (65+ years) | n = 1,281 | Children | n = 459 (36%) | 94% |
| | | Partners | n = 610 (48%) | 95% |
| | | Siblings | n = 36 (3%) | 44% |
| | | Pooled | n = 1,105 (86%) | 93% |
| | | No available closest relative | n = 176 (14%) | n/a |

**Supplementary Table 31**
**Bipolar disorder - Incident cohort 2016-2021**
**Patient characteristics - cases**

| *Age strata of index patients* | *Characteristic* | |
|---|---|---|
| Children and young people (0-24 years) | Number of persons, n | 1,924 (100.0) |
| | Male, n (%) | 621 (32.3) |
| | Age, median (Q1;Q3) | 21.3 (19.1;23.1) |
| | CCI, n (%) | |
| | 0 | 1,790 (93.0) |
| | 1-2 | 129 (6.7) |
| | +3 | 5 (0.3) |
| | Education level, n (%) | |
| | No/Unknown education | 12 (0.6) |
| | Primary | 1,191 (61.9) |
| | Secondary | 679 (35.3) |
| | Tertiary | 42 (2.2) |
| Young adults and adults (25-64 years) | Number of persons, n | 5,906 (100.0) |
| | Male, n (%) | 2,464 (41.7) |
| | Age, median (Q1;Q3) | 41.8 (32.3;51.8) |
| | CCI, n (%) | |
| | 0 | 4,977 (84.3) |
| | 1-2 | 823 (13.9) |
| | +3 | 106 (1.8) |
| | Education level, n (%) | |
| | No/Unknown education | 166 (2.8) |
| | Primary | 1,367 (23.1) |
| | Secondary | 2,571 (43.5) |
| | Tertiary | 1,802 (30.5) |
| Older people (65+ years) | Number of persons, n | 1,033 (100.0) |
| | Male, n (%) | 430 (41.6) |
| | Age, median (Q1;Q3) | 73.1 (69.0;77.8) |
| | CCI, n (%) | |
| | 0 | 477 (46.2) |
| | 1-2 | 399 (38.6) |
| | +3 | 157 (15.2) |
| | Education level, n (%) | |
| | No/Unknown education | 83 (8.0) |
| | Primary | 285 (27.6) |
| | Secondary | 426 (41.2) |
| | Tertiary | 239 (23.1) |

**Supplementary Table 32**
**Bipolar disorder - Incident cohort 2016-2021**
**Patient characteristics - relatives - pooled**

| *Age strata of index patients* | *Closest relative type* | *Characteristic of family relatives by agestrata of index patients* | *Closest family relatives of brain disorder patients* | *Comparison relative* |
|---|---|---|---|---|
| Children and young people (0-24 years) | Fathers | Number of persons, n | 1,641 (100.0) | 1,641 (100.0) |
| | | Male, n (%) | 1,641 (100.0) | 1,641 (100.0) |
| | | Age, median (Q1;Q3) | 52.5 (48.6;56.3) | 52.7 (49.3;56.1) |
| | | CCI, n (%) | | |
| | | 0 | 1,382 (84.2) | 1,429 (87.1) |
| | | 1-2 | 224 (13.7) | 190 (11.6) |
| | | +3 | 35 (2.1) | 22 (1.3) |
| | | Education level, n (%) | | |
| | | No/Unknown education | 28 (1.7) | 28 (1.7) |
| | | Primary | 295 (18.0) | 295 (18.0) |
| | | Secondary | 845 (51.5) | 845 (51.5) |
| | | Tertiary | 473 (28.8) | 473 (28.8) |
| | | Relation to patient, n (%) | | |
| | | Father | 1,641 (100.0) | 1,641 (100.0) |
| Children and young people (0-24 years) | Mothers | Number of persons, n | 1,749 (100.0) | 1,749 (100.0) |
| | | Male, n (%) | 0 (0.0) | 0 (0.0) |
| | | Age, median (Q1;Q3) | 50.7 (46.9;54.3) | 50.7 (47.3;54.0) |
| | | CCI, n (%) | | |
| | | 0 | 1,483 (84.8) | 1,525 (87.2) |
| | | 1-2 | 239 (13.7) | 199 (11.4) |
| | | +3 | 27 (1.5) | 25 (1.4) |
| | | Education level, n (%) | | |
| | | No/Unknown education | 16 (0.9) | 16 (0.9) |
| | | Primary | 276 (15.8) | 276 (15.8) |
| | | Secondary | 772 (44.1) | 772 (44.1) |
| | | Tertiary | 685 (39.2) | 685 (39.2) |
| | | Relation to patient, n (%) | | |
| | | Mother | 1,749 (100.0) | 1,749 (100.0) |
| Young adults and adults (25-64 years) | Pooled | Number of persons, n | 5,906 (100.0) | 5,906 (100.0) |
| | | Male, n (%) | 2,965 (50.2) | 2,965 (50.2) |
| | | Age, median (Q1;Q3) | 46.5 (31.5;58.5) | 46.4 (31.4;58.3) |
| | | CCI, n (%) | | |
| | | 0 | 4,993 (84.5) | 5,031 (85.2) |
| | | 1-2 | 792 (13.4) | 726 (12.3) |
| | | +3 | 121 (2.0) | 149 (2.5) |
| | | Education level, n (%) | | |
| | | No/Unknown education | 475 (8.0) | 475 (8.0) |
| | | Primary | 1,403 (23.8) | 1,403 (23.8) |
| | | Secondary | 2,476 (41.9) | 2,476 (41.9) |
| | | Tertiary | 1,552 (26.3) | 1,552 (26.3) |
| | | Relation to patient, n (%) | | |
| | | Mother | 1,098 (18.6) | 1,098 (18.6) |
| | | Father | 349 (5.9) | 349 (5.9) |
| | | Partner | 3,048 (51.6) | 3,048 (51.6) |
| | | Child | 1,000 (16.9) | 1,000 (16.9) |

**Supplementary Table 32**
**Bipolar disorder - Incident cohort 2016-2021**
**Patient characteristics - relatives - pooled**

| *Age strata of index patients* | *Closest relative type* | *Characteristic of family relatives by agestrata of index patients* | *Closest family relatives of brain disorder patients* | *Comparison relative* |
|---|---|---|---|---|
| | | Sibling | 411 (7.0) | 411 (7.0) |
| Older people (65+ years) | Pooled | Number of persons, n | 1,033 (100.0) | 1,033 (100.0) |
| | | Male, n (%) | 509 (49.3) | 509 (49.3) |
| | | Age, median (Q1;Q3) | 65.0 (51.1;73.3) | 64.9 (51.1;73.1) |
| | | CCI, n (%) | | |
| | | 0 | 750 (72.6) | 747 (72.3) |
| | | 1-2 | 221 (21.4) | 218 (21.1) |
| | | +3 | 62 (6.0) | 68 (6.6) |
| | | Education level, n (%) | | |
| | | No/Unknown education | 48 (4.6) | 48 (4.6) |
| | | Primary | 202 (19.6) | 202 (19.6) |
| | | Secondary | 469 (45.4) | 469 (45.4) |
| | | Tertiary | 314 (30.4) | 314 (30.4) |
| | | Relation to patient, n (%) | | |
| | | Partner | 585 (56.6) | 585 (56.6) |
| | | Child | 432 (41.8) | 432 (41.8) |
| | | Sibling | 16 (1.5) | 16 (1.5) |

**Supplementary Table 33**
**Bipolar disorder - Incident cohort 2016-2021**
**Patient characteristics - relatives - not pooled**

| *Age strata of index patients* | *Closest relative type* | *Characteristic of family relatives by agestrata of index patients* | *Closest family relatives of brain disorder patients* | *Comparison relative* |
|---|---|---|---|---|
| Children and young people (0-24 years) | Fathers | Number of persons, n | 1,641 (100.0) | 1,641 (100.0) |
| | | Male, n (%) | 1,641 (100.0) | 1,641 (100.0) |
| | | Age, median (Q1;Q3) | 52.5 (48.6;56.3) | 52.7 (49.3;56.1) |
| | | CCI, n (%) | | |
| | | 0 | 1,382 (84.2) | 1,429 (87.1) |
| | | 1-2 | 224 (13.7) | 190 (11.6) |
| | | +3 | 35 (2.1) | 22 (1.3) |
| | | Education level, n (%) | | |
| | | No/Unknown education | 28 (1.7) | 28 (1.7) |
| | | Primary | 295 (18.0) | 295 (18.0) |
| | | Secondary | 845 (51.5) | 845 (51.5) |
| | | Tertiary | 473 (28.8) | 473 (28.8) |
| | | Relation to patient, n (%) | | |
| | | Father | 1,641 (100.0) | 1,641 (100.0) |
| Children and young people (0-24 years) | Mothers | Number of persons, n | 1,749 (100.0) | 1,749 (100.0) |
| | | Male, n (%) | 0 (0.0) | 0 (0.0) |
| | | Age, median (Q1;Q3) | 50.7 (46.9;54.3) | 50.7 (47.3;54.0) |
| | | CCI, n (%) | | |
| | | 0 | 1,483 (84.8) | 1,525 (87.2) |
| | | 1-2 | 239 (13.7) | 199 (11.4) |
| | | +3 | 27 (1.5) | 25 (1.4) |
| | | Education level, n (%) | | |
| | | No/Unknown education | 16 (0.9) | 16 (0.9) |
| | | Primary | 276 (15.8) | 276 (15.8) |
| | | Secondary | 772 (44.1) | 772 (44.1) |
| | | Tertiary | 685 (39.2) | 685 (39.2) |
| | | Relation to patient, n (%) | | |
| | | Mother | 1,749 (100.0) | 1,749 (100.0) |
| Young adults and adults (25-64 years) | Children | Number of persons, n | 1,000 (100.0) | 1,000 (100.0) |
| | | Male, n (%) | 518 (51.8) | 518 (51.8) |
| | | Age, median (Q1;Q3) | 17.6 (10.4;25.0) | 16.9 (9.9;24.6) |
| | | CCI, n (%) | | |
| | | 0 | 935 (93.5) | 936 (93.6) |
| | | 1-2 | * | * |
| | | +3 | * | * |
| | | Education level, n (%) | | |
| | | No/Unknown education | 349 (34.9) | 349 (34.9) |
| | | Primary | 383 (38.3) | 383 (38.3) |
| | | Secondary | 180 (18.0) | 180 (18.0) |
| | | Tertiary | 88 (8.8) | 88 (8.8) |
| | | Relation to patient, n (%) | | |
| | | Child | 1,000 (100.0) | 1,000 (100.0) |
| Young adults and adults (25-64 years) | Parents | Number of persons, n | 1,447 (100.0) | 1,447 (100.0) |
| | | Male, n (%) | 349 (24.1) | 349 (24.1) |
| | | Age, median (Q1;Q3) | 62.9 (57.0;71.2) | 62.7 (57.1;71.1) |

**Supplementary Table 33**
**Bipolar disorder - Incident cohort 2016-2021**
**Patient characteristics - relatives - not pooled**

| *Age strata of index patients* | *Closest relative type* | *Characteristic of family relatives by agestrata of index patients* | *Closest family relatives of brain disorder patients* | *Comparison relative* |
|---|---|---|---|---|
| | | CCI, n (%) | | |
| | | 0 | 1,021 (70.6) | 1,029 (71.1) |
| | | 1-2 | 353 (24.4) | 325 (22.5) |
| | | +3 | 73 (5.0) | 93 (6.4) |
| | | Education level, n (%) | | |
| | | No/Unknown education | 34 (2.3) | 34 (2.3) |
| | | Primary | 358 (24.7) | 358 (24.7) |
| | | Secondary | 694 (48.0) | 694 (48.0) |
| | | Tertiary | 361 (24.9) | 361 (24.9) |
| | | Relation to patient, n (%) | | |
| | | Mother | 1,098 (75.9) | 1,098 (75.9) |
| | | Father | 349 (24.1) | 349 (24.1) |
| Young adults and adults (25-64 years) | Partners | Number of persons, n | 3,048 (100.0) | 3,048 (100.0) |
| | | Male, n (%) | 1,848 (60.6) | 1,848 (60.6) |
| | | Age, median (Q1;Q3) | 43.8 (34.9;53.3) | 43.9 (34.6;53.2) |
| | | CCI, n (%) | | |
| | | 0 | 2,692 (88.3) | 2,714 (89.0) |
| | | 1-2 | 313 (10.3) | 288 (9.4) |
| | | +3 | 43 (1.4) | 46 (1.5) |
| | | Education level, n (%) | | |
| | | No/Unknown education | 91 (3.0) | 91 (3.0) |
| | | Primary | 585 (19.2) | 585 (19.2) |
| | | Secondary | 1,370 (44.9) | 1,370 (44.9) |
| | | Tertiary | 1,002 (32.9) | 1,002 (32.9) |
| | | Relation to patient, n (%) | | |
| | | Partner | 3,048 (100.0) | 3,048 (100.0) |
| Young adults and adults (25-64 years) | Siblings | Number of persons, n | 411 (100.0) | 411 (100.0) |
| | | Male, n (%) | 250 (60.8) | 250 (60.8) |
| | | Age, median (Q1;Q3) | 48.5 (33.3;57.7) | 48.0 (33.4;57.1) |
| | | CCI, n (%) | | |
| | | 0 | 345 (83.9) | 352 (85.6) |
| | | 1-2 | * | 51 (12.4) |
| | | +3 | * | 8 (1.9) |
| | | Education level, n (%) | | |
| | | No/Unknown education | * | * |
| | | Primary | * | * |
| | | Secondary | 232 (56.4) | 232 (56.4) |
| | | Tertiary | 101 (24.6) | 101 (24.6) |
| | | Relation to patient, n (%) | | |
| | | Sibling | 411 (100.0) | 411 (100.0) |
| Older people (65+ years) | Children | Number of persons, n | 432 (100.0) | 432 (100.0) |
| | | Male, n (%) | 211 (48.8) | 211 (48.8) |
| | | Age, median (Q1;Q3) | 49.3 (43.7;54.9) | 49.0 (43.8;54.4) |
| | | CCI, n (%) | | |
| | | 0 | 370 (85.6) | 372 (86.1) |

**Supplementary Table 33**
**Bipolar disorder - Incident cohort 2016-2021**
**Patient characteristics - relatives - not pooled**

| *Age strata of index patients* | *Closest relative type* | *Characteristic of family relatives by agestrata of index patients* | *Closest family relatives of brain disorder patients* | *Comparison relative* |
|---|---|---|---|---|
| | | 1-2 | 57 (13.2) | 51 (11.8) |
| | | +3 | 5 (1.2) | 9 (2.1) |
| | | Education level, n (%) | | |
| | | No/Unknown education | * | * |
| | | Primary | * | * |
| | | Secondary | 197 (45.6) | 197 (45.6) |
| | | Tertiary | 155 (35.9) | 155 (35.9) |
| | | Relation to patient, n (%) | | |
| | | Child | 432 (100.0) | 432 (100.0) |
| Older people (65+ years) | Partners | Number of persons, n | 585 (100.0) | 585 (100.0) |
| | | Male, n (%) | 286 (48.9) | 286 (48.9) |
| | | Age, median (Q1;Q3) | 72.5 (68.0;77.4) | 72.2 (67.9;76.8) |
| | | CCI, n (%) | | |
| | | 0 | 366 (62.6) | 365 (62.4) |
| | | 1-2 | 162 (27.7) | 161 (27.5) |
| | | +3 | 57 (9.7) | 59 (10.1) |
| | | Education level, n (%) | | |
| | | No/Unknown education | 46 (7.9) | 46 (7.9) |
| | | Primary | 122 (20.9) | 122 (20.9) |
| | | Secondary | 261 (44.6) | 261 (44.6) |
| | | Tertiary | 156 (26.7) | 156 (26.7) |
| | | Relation to patient, n (%) | | |
| | | Partner | 585 (100.0) | 585 (100.0) |
| Older people (65+ years) | Siblings | Number of persons, n | 16 (100.0) | 16 (100.0) |
| | | Male, n (%) | * | * |
| | | Age, median (Q1;Q3) | 61.9 (59.9;64.3) | 61.8 (60.2;63.5) |
| | | CCI, n (%) | | |
| | | 0 | * | 10 (62.5) |
| | | 1-2 | * | 6 (37.5) |
| | | +3 | 0 (0.0) | 0 (0.0) |
| | | Education level, n (%) | | |
| | | No/Unknown education | 0 (0.0) | 0 (0.0) |
| | | Primary | * | * |
| | | Secondary | * | * |
| | | Tertiary | * | * |
| | | Relation to patient, n (%) | | |
| | | Sibling | 16 (100.0) | 16 (100.0) |

**Supplementary Table 34**
**Bipolar disorder - Incident cohort 2016-2021**
**Cost analysis - pooled**

| *Age strata of index patients* | *Closest relative type* | *Number of index patients included in cost component calculations* | *Cost component* | *Actual costs* | | *Attributable costs* | |
|---|---|---|---|---|---|---|---|
| | | | | *Total (million EUR)* | *Per person (EUR)* | *Total (million EUR)* | *Per person (EUR)* |
| Children and young people (0-24 years) | Fathers | 1,641 | Primary sector | 0.4 | 268 | 0.0 | 18 |
| | | 1,641 | Hospital care | 2.9 | 1,800 | 0.5 | 312 |
| | | 1,641 | Filled prescriptions | 0.4 | 273 | 0.1 | 63 |
| | | 1,550 | Lost productivity (income loss) | . | . | -6.4 | -4,136 |
| Children and young people (0-24 years) | Mothers | 1,749 | Primary sector | 0.7 | 383 | 0.1 | 50 |
| | | 1,749 | Hospital care | 3.6 | 2,045 | 0.9 | 541 |
| | | 1,749 | Filled prescriptions | 0.5 | 270 | 0.1 | 72 |
| | | 1,729 | Lost productivity (income loss) | . | . | 1.2 | 676 |
| Young adults and adults (25-64 years) | Pooled | 5,906 | Primary sector | 1.9 | 316 | 0.2 | 30 |
| | | 5,906 | Hospital care | 11.4 | 1,946 | -0.2 | -38 |
| | | 5,906 | Filled prescriptions | 1.5 | 248 | 0.2 | 39 |
| | | 4,372 | Lost productivity (income loss) | . | . | 17.5 | 4,011 |
| Older people (65+ years) | Pooled | 1,033 | Primary sector | 0.5 | 455 | 0.1 | 68 |
| | | 1,033 | Hospital care | 2.7 | 2,678 | 0.3 | 301 |
| | | 1,033 | Filled prescriptions | 0.4 | 376 | 0.0 | 28 |
| | | 447 | Lost productivity (income loss) | . | . | 1.1 | 2,538 |

**Supplementary Table 35**
**Bipolar disorder - Incident cohort 2016-2021**
**Cost analysis - not pooled**

| | | | | *Actual costs* | | *Attributable costs* | |
|---|---|---|---|---|---|---|---|
| ***Age strata of index patients*** | ***Closest relative type*** | ***Number of index patients included in cost component calculations*** | ***Cost component*** | ***Total (million EUR)*** | ***Per person (EUR)*** | ***Total (million EUR)*** | ***Per person (EUR)*** |
| Children and young people (0-24 years) | Fathers | 1,641 | Primary sector | 0.4 | 268 | 0.0 | 18 |
| | | 1,641 | Hospital care | 2.9 | 1,800 | 0.5 | 312 |
| | | 1,641 | Filled prescriptions | 0.4 | 273 | 0.1 | 63 |
| | | 1,550 | Lost productivity (income loss) | . | . | -6.4 | -4,136 |
| Children and young people (0-24 years) | Mothers | 1,749 | Primary sector | 0.7 | 383 | 0.1 | 50 |
| | | 1,749 | Hospital care | 3.6 | 2,045 | 0.9 | 541 |
| | | 1,749 | Filled prescriptions | 0.5 | 270 | 0.1 | 72 |
| | | 1,729 | Lost productivity (income loss) | . | . | 1.2 | 676 |
| Young adults and adults (25-64 years) | Children | 1,000 | Primary sector | 0.2 | 218 | 0.0 | 33 |
| | | 1,000 | Hospital care | 1.0 | 1,005 | 0.0 | 16 |
| | | 1,000 | Filled prescriptions | 0.1 | 96 | -0.0 | -11 |
| | | 435 | Lost productivity (income loss) | . | . | 1.1 | 2,561 |
| Young adults and adults (25-64 years) | Parents | 1,447 | Primary sector | 0.6 | 440 | 0.0 | 31 |
| | | 1,447 | Hospital care | 4.3 | 3,021 | 0.2 | 168 |
| | | 1,447 | Filled prescriptions | 0.7 | 454 | 0.1 | 70 |
| | | 703 | Lost productivity (income loss) | . | . | 1.2 | 1,755 |
| Young adults and adults (25-64 years) | Partners | 3,048 | Primary sector | 0.9 | 292 | 0.1 | 30 |
| | | 3,048 | Hospital care | 4.9 | 1,603 | -0.8 | -263 |
| | | 3,048 | Filled prescriptions | 0.6 | 202 | 0.1 | 37 |
| | | 2,864 | Lost productivity (income loss) | . | . | 13.3 | 4,630 |
| Young adults and adults (25-64 years) | Siblings | 411 | Primary sector | 0.1 | 292 | 0.0 | 19 |
| | | 411 | Hospital care | 1.2 | 2,996 | 0.3 | 780 |
| | | 411 | Filled prescriptions | 0.1 | 235 | 0.0 | 62 |
| | | 370 | Lost productivity (income loss) | . | . | 1.9 | 5,212 |
| Older people (65+ years) | Children | 432 | Primary sector | 0.2 | 356 | 0.0 | 82 |
| | | 432 | Hospital care | 0.6 | 1,463 | 0.1 | 137 |
| | | 432 | Filled prescriptions | 0.1 | 189 | 0.0 | 6 |
| | | 416 | Lost productivity (income loss) | . | . | 1.7 | 3,978 |
| Older people (65+ years) | Partners | 585 | Primary sector | 0.3 | 530 | 0.0 | 57 |
| | | 585 | Hospital care | 2.1 | 3,649 | 0.2 | 432 |
| | | 585 | Filled prescriptions | 0.3 | 520 | 0.0 | 43 |
| | | 23 | Lost productivity (income loss) | . | . | -0.4 | -16,319 |
| Older people (65+ years) | Siblings | 16 | Primary sector | 0.0 | 416 | 0.0 | 107 |
| | | 16 | Hospital care | 0.0 | 629 | 0.0 | 27 |
| | | 16 | Filled prescriptions | 0.0 | 240 | 0.0 | 80 |
| | | 8 | Lost productivity (income loss) | . | . | -0.1 | -18,142 |

#### Supplementary Table 36
#### Bipolar disorder - Prevalent cohort 2021
#### Identification of populations of relatives

| ***Agegroup of index patients in matched population*** | | ***Closest relative type identified*** | | ***Available control with same type of closest relative*** |
|---|---|---|---|---|
| Children and young people (0-24 years) | n = 1,434 | Fathers | n = 1,140 (79%) | 95% |
| | | Mothers | n = 1,188 (83%) | 95% |
| | | No available closest relative | n = 78 (5%) | n/a |
| Young adults and adults (25-64 years) | n = 24,123 | Children | n = 4,172 (17%) | 94% |
| | | Parents | n = 5,447 (23%) | 88% |
| | | Partners | n = 9,904 (41%) | 95% |
| | | Siblings | n = 3,882 (16%) | 47% |
| | | Pooled | n = 23,405 (97%) | 85% |
| | | No available closest relative | n = 718 (3%) | n/a |
| Older people (65+ years) | n = 9,241 | Children | n = 3,779 (41%) | 94% |
| | | Partners | n = 3,758 (41%) | 96% |
| | | Siblings | n = 529 (6%) | 49% |
| | | Pooled | n = 8,066 (87%) | 92% |
| | | No available closest relative | n = 1,175 (13%) | n/a |

**Supplementary Table 37**
**Bipolar disorder - Prevalent cohort 2021**
**Patient characteristics - cases**

| ***Age strata of index patients*** | ***Characteristic*** | |
|---|---|---|
| Children and young people (0-24 years) | Number of persons, n | 1,301 (100.0) |
| | Male, n (%) | 440 (33.8) |
| | Age, median (Q1;Q3) | 22.4 (20.4;23.8) |
| | CCI, n (%) | |
| | 0 | 1,192 (91.6) |
| | 1-2 | 104 (8.0) |
| | +3 | 5 (0.4) |
| | Education level, n (%) | |
| | No/Unknown education | 6 (0.5) |
| | Primary | 794 (61.0) |
| | Secondary | 462 (35.5) |
| | Tertiary | 39 (3.0) |
| Young adults and adults (25-64 years) | Number of persons, n | 20,063 (100.0) |
| | Male, n (%) | 7,674 (38.2) |
| | Age, median (Q1;Q3) | 47.8 (37.7;56.4) |
| | CCI, n (%) | |
| | 0 | 15,827 (78.9) |
| | 1-2 | 3,598 (17.9) |
| | +3 | 638 (3.2) |
| | Education level, n (%) | |
| | No/Unknown education | 394 (2.0) |
| | Primary | 5,516 (27.5) |
| | Secondary | 8,655 (43.1) |
| | Tertiary | 5,498 (27.4) |
| Older people (65+ years) | Number of persons, n | 7,443 (100.0) |
| | Male, n (%) | 2,757 (37.0) |
| | Age, median (Q1;Q3) | 73.5 (68.8;78.3) |
| | CCI, n (%) | |
| | 0 | 3,373 (45.3) |
| | 1-2 | 2,874 (38.6) |
| | +3 | 1,196 (16.1) |
| | Education level, n (%) | |
| | No/Unknown education | 545 (7.3) |
| | Primary | 1,956 (26.3) |
| | Secondary | 3,132 (42.1) |
| | Tertiary | 1,810 (24.3) |

**Supplementary Table 38**
**Bipolar disorder - Prevalent cohort 2021**
**Patient characteristics - relatives - pooled**

| *Age strata of index patients* | *Closest relative type* | *Characteristic of family relatives by agestrata of index patients* | *Closest family relatives of brain disorder patients* | *Comparison relative* |
|---|---|---|---|---|
| Children and young people (0-24 years) | Fathers | Number of persons, n | 1,093 (100.0) | 1,093 (100.0) |
| | | Male, n (%) | 1,093 (100.0) | 1,093 (100.0) |
| | | Age, median (Q1;Q3) | 53.7 (49.6;57.6) | 53.6 (49.8;57.2) |
| | | CCI, n (%) | | |
| | | 0 | 904 (82.7) | 915 (83.7) |
| | | 1-2 | 163 (14.9) | 160 (14.6) |
| | | +3 | 26 (2.4) | 18 (1.6) |
| | | Education level, n (%) | | |
| | | No/Unknown education | 22 (2.0) | 22 (2.0) |
| | | Primary | 199 (18.2) | 199 (18.2) |
| | | Secondary | 547 (50.0) | 547 (50.0) |
| | | Tertiary | 325 (29.7) | 325 (29.7) |
| | | Relation to patient, n (%) | | |
| | | Father | 1,093 (100.0) | 1,093 (100.0) |
| Children and young people (0-24 years) | Mothers | Number of persons, n | 1,134 (100.0) | 1,134 (100.0) |
| | | Male, n (%) | 0 (0.0) | 0 (0.0) |
| | | Age, median (Q1;Q3) | 51.8 (47.9;55.4) | 51.8 (48.6;55.2) |
| | | CCI, n (%) | | |
| | | 0 | 942 (83.1) | 979 (86.3) |
| | | 1-2 | 170 (15.0) | 138 (12.2) |
| | | +3 | 22 (1.9) | 17 (1.5) |
| | | Education level, n (%) | | |
| | | No/Unknown education | 8 (0.7) | 8 (0.7) |
| | | Primary | 177 (15.6) | 177 (15.6) |
| | | Secondary | 507 (44.7) | 507 (44.7) |
| | | Tertiary | 442 (39.0) | 442 (39.0) |
| | | Relation to patient, n (%) | | |
| | | Mother | 1,134 (100.0) | 1,134 (100.0) |
| Young adults and adults (25-64 years) | Pooled | Number of persons, n | 20,063 (100.0) | 20,063 (100.0) |
| | | Male, n (%) | 10,319 (51.4) | 10,319 (51.4) |
| | | Age, median (Q1;Q3) | 51.0 (33.8;62.0) | 51.1 (33.8;61.9) |
| | | CCI, n (%) | | |
| | | 0 | 16,431 (81.9) | 16,618 (82.8) |
| | | 1-2 | 3,031 (15.1) | 2,865 (14.3) |
| | | +3 | 601 (3.0) | 580 (2.9) |
| | | Education level, n (%) | | |
| | | No/Unknown education | 1,080 (5.4) | 1,080 (5.4) |
| | | Primary | 5,170 (25.8) | 5,170 (25.8) |
| | | Secondary | 8,839 (44.1) | 8,839 (44.1) |
| | | Tertiary | 4,974 (24.8) | 4,974 (24.8) |
| | | Relation to patient, n (%) | | |
| | | Mother | 3,733 (18.6) | 3,733 (18.6) |
| | | Father | 1,110 (5.5) | 1,110 (5.5) |
| | | Partner | 9,455 (47.1) | 9,455 (47.1) |
| | | Child | 3,927 (19.6) | 3,927 (19.6) |

#### Supplementary Table 38
#### Bipolar disorder - Prevalent cohort 2021
#### Patient characteristics - relatives - pooled

| *Age strata of index patients* | *Closest relative type* | *Characteristic of family relatives by agestrata of index patients* | *Closest family relatives of brain disorder patients* | *Comparison relative* |
|---|---|---|---|---|
| | | Sibling | 1,838 (9.2) | 1,838 (9.2) |
| Older people (65+ years) | Pooled | Number of persons, n | 7,443 (100.0) | 7,443 (100.0) |
| | | Male, n (%) | 3,925 (52.7) | 3,925 (52.7) |
| | | Age, median (Q1;Q3) | 62.8 (49.8;72.6) | 62.8 (49.8;72.8) |
| | | CCI, n (%) | | |
| | | 0 | 5,418 (72.8) | 5,523 (74.2) |
| | | 1-2 | 1,617 (21.7) | 1,503 (20.2) |
| | | +3 | 408 (5.5) | 417 (5.6) |
| | | Education level, n (%) | | |
| | | No/Unknown education | 254 (3.4) | 254 (3.4) |
| | | Primary | 1,292 (17.4) | 1,292 (17.4) |
| | | Secondary | 3,463 (46.5) | 3,463 (46.5) |
| | | Tertiary | 2,434 (32.7) | 2,434 (32.7) |
| | | Relation to patient, n (%) | | |
| | | Partner | 3,610 (48.5) | 3,610 (48.5) |
| | | Child | 3,571 (48.0) | 3,571 (48.0) |
| | | Sibling | 262 (3.5) | 262 (3.5) |

**Supplementary Table 39**
**Bipolar disorder - Prevalent cohort 2021**
**Patient characteristics - relatives - not pooled**

| *Age strata of index patients* | *Closest relative type* | *Characteristic of family relatives by agestrata of index patients* | *Closest family relatives of brain disorder patients* | *Comparison relative* |
|---|---|---|---|---|
| Children and young people (0-24 years) | Fathers | Number of persons, n | 1,093 (100.0) | 1,093 (100.0) |
| | | Male, n (%) | 1,093 (100.0) | 1,093 (100.0) |
| | | Age, median (Q1;Q3) | 53.7 (49.6;57.6) | 53.6 (49.8;57.2) |
| | | CCI, n (%) | | |
| | | 0 | 904 (82.7) | 915 (83.7) |
| | | 1-2 | 163 (14.9) | 160 (14.6) |
| | | +3 | 26 (2.4) | 18 (1.6) |
| | | Education level, n (%) | | |
| | | No/Unknown education | 22 (2.0) | 22 (2.0) |
| | | Primary | 199 (18.2) | 199 (18.2) |
| | | Secondary | 547 (50.0) | 547 (50.0) |
| | | Tertiary | 325 (29.7) | 325 (29.7) |
| | | Relation to patient, n (%) | | |
| | | Father | 1,093 (100.0) | 1,093 (100.0) |
| Children and young people (0-24 years) | Mothers | Number of persons, n | 1,134 (100.0) | 1,134 (100.0) |
| | | Male, n (%) | 0 (0.0) | 0 (0.0) |
| | | Age, median (Q1;Q3) | 51.8 (47.9;55.4) | 51.8 (48.6;55.2) |
| | | CCI, n (%) | | |
| | | 0 | 942 (83.1) | 979 (86.3) |
| | | 1-2 | 170 (15.0) | 138 (12.2) |
| | | +3 | 22 (1.9) | 17 (1.5) |
| | | Education level, n (%) | | |
| | | No/Unknown education | 8 (0.7) | 8 (0.7) |
| | | Primary | 177 (15.6) | 177 (15.6) |
| | | Secondary | 507 (44.7) | 507 (44.7) |
| | | Tertiary | 442 (39.0) | 442 (39.0) |
| | | Relation to patient, n (%) | | |
| | | Mother | 1,134 (100.0) | 1,134 (100.0) |
| Young adults and adults (25-64 years) | Children | Number of persons, n | 3,927 (100.0) | 3,927 (100.0) |
| | | Male, n (%) | 2,053 (52.3) | 2,053 (52.3) |
| | | Age, median (Q1;Q3) | 22.5 (15.3;30.3) | 22.0 (15.0;30.3) |
| | | CCI, n (%) | | |
| | | 0 | 3,676 (93.6) | 3,688 (93.9) |
| | | 1-2 | 242 (6.2) | 226 (5.8) |
| | | +3 | 9 (0.2) | 13 (0.3) |
| | | Education level, n (%) | | |
| | | No/Unknown education | 756 (19.3) | 756 (19.3) |
| | | Primary | 1,612 (41.0) | 1,612 (41.0) |
| | | Secondary | 1,025 (26.1) | 1,025 (26.1) |
| | | Tertiary | 534 (13.6) | 534 (13.6) |
| | | Relation to patient, n (%) | | |
| | | Child | 3,927 (100.0) | 3,927 (100.0) |
| Young adults and adults (25-64 years) | Parents | Number of persons, n | 4,843 (100.0) | 4,843 (100.0) |
| | | Male, n (%) | 1,110 (22.9) | 1,110 (22.9) |
| | | Age, median (Q1;Q3) | 68.1 (60.8;75.2) | 68.1 (60.9;75.4) |

### Supplementary Table 39
### Bipolar disorder - Prevalent cohort 2021
### Patient characteristics - relatives - not pooled

| *Age strata of index patients* | *Closest relative type* | *Characteristic of family relatives by agestrata of index patients* | *Closest family relatives of brain disorder patients* | *Comparison relative* |
|---|---|---|---|---|
| | | CCI, n (%) | | |
| | | 0 | 3,166 (65.4) | 3,215 (66.4) |
| | | 1-2 | 1,311 (27.1) | 1,274 (26.3) |
| | | +3 | 366 (7.6) | 354 (7.3) |
| | | Education level, n (%) | | |
| | | No/Unknown education | 84 (1.7) | 84 (1.7) |
| | | Primary | 1,281 (26.5) | 1,281 (26.5) |
| | | Secondary | 2,329 (48.1) | 2,329 (48.1) |
| | | Tertiary | 1,149 (23.7) | 1,149 (23.7) |
| | | Relation to patient, n (%) | | |
| | | Mother | 3,733 (77.1) | 3,733 (77.1) |
| | | Father | 1,110 (22.9) | 1,110 (22.9) |
| Young adults and adults (25-64 years) | Partners | Number of persons, n | 9,455 (100.0) | 9,455 (100.0) |
| | | Male, n (%) | 6,098 (64.5) | 6,098 (64.5) |
| | | Age, median (Q1;Q3) | 48.7 (38.6;57.3) | 48.8 (38.7;57.2) |
| | | CCI, n (%) | | |
| | | 0 | 8,058 (85.2) | 8,212 (86.9) |
| | | 1-2 | 1,214 (12.8) | 1,090 (11.5) |
| | | +3 | 183 (1.9) | 153 (1.6) |
| | | Education level, n (%) | | |
| | | No/Unknown education | 235 (2.5) | 235 (2.5) |
| | | Primary | 1,885 (19.9) | 1,885 (19.9) |
| | | Secondary | 4,510 (47.7) | 4,510 (47.7) |
| | | Tertiary | 2,825 (29.9) | 2,825 (29.9) |
| | | Relation to patient, n (%) | | |
| | | Partner | 9,455 (100.0) | 9,455 (100.0) |
| Young adults and adults (25-64 years) | Siblings | Number of persons, n | 1,838 (100.0) | 1,838 (100.0) |
| | | Male, n (%) | 1,058 (57.6) | 1,058 (57.6) |
| | | Age, median (Q1;Q3) | 55.2 (44.7;60.8) | 55.3 (45.2;60.6) |
| | | CCI, n (%) | | |
| | | 0 | 1,531 (83.3) | 1,503 (81.8) |
| | | 1-2 | 264 (14.4) | 275 (15.0) |
| | | +3 | 43 (2.3) | 60 (3.3) |
| | | Education level, n (%) | | |
| | | No/Unknown education | 5 (0.3) | 5 (0.3) |
| | | Primary | 392 (21.3) | 392 (21.3) |
| | | Secondary | 975 (53.0) | 975 (53.0) |
| | | Tertiary | 466 (25.4) | 466 (25.4) |
| | | Relation to patient, n (%) | | |
| | | Sibling | 1,838 (100.0) | 1,838 (100.0) |
| Older people (65+ years) | Children | Number of persons, n | 3,571 (100.0) | 3,571 (100.0) |
| | | Male, n (%) | 1,853 (51.9) | 1,853 (51.9) |
| | | Age, median (Q1;Q3) | 49.3 (43.8;55.5) | 49.4 (43.5;55.3) |
| | | CCI, n (%) | | |
| | | 0 | 3,057 (85.6) | 3,096 (86.7) |

**Supplementary Table 39**
**Bipolar disorder - Prevalent cohort 2021**
**Patient characteristics - relatives - not pooled**

| *Age strata of index patients* | *Closest relative type* | *Characteristic of family relatives by agestrata of index patients* | *Closest family relatives of brain disorder patients* | *Comparison relative* |
|---|---|---|---|---|
| | | 1-2 | 447 (12.5) | 421 (11.8) |
| | | +3 | 67 (1.9) | 54 (1.5) |
| | | Education level, n (%) | | |
| | | No/Unknown education | 5 (0.1) | 5 (0.1) |
| | | Primary | 537 (15.0) | 537 (15.0) |
| | | Secondary | 1,570 (44.0) | 1,570 (44.0) |
| | | Tertiary | 1,459 (40.9) | 1,459 (40.9) |
| | | Relation to patient, n (%) | | |
| | | Child | 3,571 (100.0) | 3,571 (100.0) |
| Older people (65+ years) | Partners | Number of persons, n | 3,610 (100.0) | 3,610 (100.0) |
| | | Male, n (%) | 1,938 (53.7) | 1,938 (53.7) |
| | | Age, median (Q1;Q3) | 72.8 (68.3;77.3) | 72.9 (68.4;77.0) |
| | | CCI, n (%) | | |
| | | 0 | 2,164 (59.9) | 2,218 (61.4) |
| | | 1-2 | 1,112 (30.8) | 1,041 (28.8) |
| | | +3 | 334 (9.3) | 351 (9.7) |
| | | Education level, n (%) | | |
| | | No/Unknown education | 249 (6.9) | 249 (6.9) |
| | | Primary | 713 (19.8) | 713 (19.8) |
| | | Secondary | 1,750 (48.5) | 1,750 (48.5) |
| | | Tertiary | 898 (24.9) | 898 (24.9) |
| | | Relation to patient, n (%) | | |
| | | Partner | 3,610 (100.0) | 3,610 (100.0) |
| Older people (65+ years) | Siblings | Number of persons, n | 262 (100.0) | 262 (100.0) |
| | | Male, n (%) | 134 (51.1) | 134 (51.1) |
| | | Age, median (Q1;Q3) | 62.4 (59.0;65.1) | 62.5 (59.7;64.9) |
| | | CCI, n (%) | | |
| | | 0 | 197 (75.2) | 209 (79.8) |
| | | 1-2 | 58 (22.1) | 41 (15.6) |
| | | +3 | 7 (2.7) | 12 (4.6) |
| | | Education level, n (%) | | |
| | | No/Unknown education | 0 (0.0) | 0 (0.0) |
| | | Primary | 42 (16.0) | 42 (16.0) |
| | | Secondary | 143 (54.6) | 143 (54.6) |
| | | Tertiary | 77 (29.4) | 77 (29.4) |
| | | Relation to patient, n (%) | | |
| | | Sibling | 262 (100.0) | 262 (100.0) |

### Supplementary Table 40
### Bipolar disorder - Prevalent cohort 2021
### Cost analysis - pooled

| Age strata of index patients | Closest relative type | Number of index patients included in cost component calculations | Cost component | Actual costs | | Attributable costs | |
|---|---|---|---|---|---|---|---|
| | | | | Total (million EUR) | Per person (EUR) | Total (million EUR) | Per person (EUR) |
| Children and young people (0-24 years) | Fathers | 1,093 | Primary sector | 0.3 | 289 | 0.0 | 46 |
| | | 1,093 | Hospital care | 2.3 | 2,119 | 0.9 | 819 |
| | | 1,093 | Filled prescriptions | 0.3 | 255 | 0.0 | 32 |
| | | 1,021 | Lost productivity (income loss) | . | . | 6.6 | 6,493 |
| Children and young people (0-24 years) | Mothers | 1,134 | Primary sector | 0.5 | 399 | 0.1 | 56 |
| | | 1,134 | Hospital care | 2.3 | 1,999 | 0.9 | 788 |
| | | 1,134 | Filled prescriptions | 0.3 | 281 | 0.1 | 81 |
| | | 1,121 | Lost productivity (income loss) | . | . | 0.6 | 560 |
| Young adults and adults (25-64 years) | Pooled | 20,063 | Primary sector | 6.8 | 338 | 0.6 | 31 |
| | | 20,063 | Hospital care | 47.9 | 2,395 | 6.9 | 346 |
| | | 20,063 | Filled prescriptions | 6.2 | 308 | 1.1 | 55 |
| | | 13,832 | Lost productivity (income loss) | . | . | 76.3 | 5,516 |
| Older people (65+ years) | Pooled | 7,443 | Primary sector | 3.0 | 411 | 0.1 | 19 |
| | | 7,443 | Hospital care | 21.7 | 2,936 | 1.6 | 215 |
| | | 7,443 | Filled prescriptions | 3.0 | 400 | 0.3 | 44 |
| | | 3,703 | Lost productivity (income loss) | . | . | 3.1 | 837 |

**Supplementary Table 41**
**Bipolar disorder - Prevalent cohort 2021**
**Cost analysis - not pooled**

| | | | | *Actual costs* | | *Attributable costs* | |
|---|---|---|---|---|---|---|---|
| *Age strata of index patients* | *Closest relative type* | *Number of index patients included in cost component calculations* | *Cost component* | *Total (million EUR)* | *Per person (EUR)* | *Total (million EUR)* | *Per person (EUR)* |
| Children and young people (0-24 years) | Fathers | 1,093 | Primary sector | 0.3 | 289 | 0.0 | 46 |
| | | 1,093 | Hospital care | 2.3 | 2,119 | 0.9 | 819 |
| | | 1,093 | Filled prescriptions | 0.3 | 255 | 0.0 | 32 |
| | | 1,021 | Lost productivity (income loss) | . | . | 6.6 | 6,493 |
| Children and young people (0-24 years) | Mothers | 1,134 | Primary sector | 0.5 | 399 | 0.1 | 56 |
| | | 1,134 | Hospital care | 2.3 | 1,999 | 0.9 | 788 |
| | | 1,134 | Filled prescriptions | 0.3 | 281 | 0.1 | 81 |
| | | 1,121 | Lost productivity (income loss) | . | . | 0.6 | 560 |
| Young adults and adults (25-64 years) | Children | 3,927 | Primary sector | 0.9 | 240 | 0.1 | 28 |
| | | 3,927 | Hospital care | 7.1 | 1,814 | 2.0 | 517 |
| | | 3,927 | Filled prescriptions | 0.5 | 131 | 0.1 | 14 |
| | | 2,354 | Lost productivity (income loss) | . | . | 2.4 | 1,020 |
| Young adults and adults (25-64 years) | Parents | 4,843 | Primary sector | 2.4 | 496 | 0.2 | 33 |
| | | 4,843 | Hospital care | 17.6 | 3,666 | 1.4 | 294 |
| | | 4,843 | Filled prescriptions | 2.6 | 536 | 0.3 | 60 |
| | | 1,507 | Lost productivity (income loss) | . | . | 7.1 | 4,717 |
| Young adults and adults (25-64 years) | Partners | 9,455 | Primary sector | 2.8 | 301 | 0.3 | 34 |
| | | 9,455 | Hospital care | 18.5 | 1,959 | 3.6 | 384 |
| | | 9,455 | Filled prescriptions | 2.5 | 269 | 0.8 | 81 |
| | | 8,462 | Lost productivity (income loss) | . | . | 69.5 | 8,218 |
| Young adults and adults (25-64 years) | Siblings | 1,838 | Primary sector | 0.6 | 326 | 0.0 | 20 |
| | | 1,838 | Hospital care | 4.7 | 2,561 | -0.1 | -80 |
| | | 1,838 | Filled prescriptions | 0.5 | 290 | -0.0 | -6 |
| | | 1,509 | Lost productivity (income loss) | . | . | -2.8 | -1,824 |
| Older people (65+ years) | Children | 3,571 | Primary sector | 1.1 | 317 | 0.1 | 27 |
| | | 3,571 | Hospital care | 6.7 | 1,879 | 1.3 | 352 |
| | | 3,571 | Filled prescriptions | 0.9 | 240 | 0.1 | 25 |
| | | 3,380 | Lost productivity (income loss) | . | . | 5.9 | 1,736 |
| Older people (65+ years) | Partners | 3,610 | Primary sector | 1.8 | 507 | 0.0 | 11 |
| | | 3,610 | Hospital care | 14.5 | 4,079 | 0.4 | 110 |
| | | 3,610 | Filled prescriptions | 2.0 | 567 | 0.2 | 67 |
| | | 173 | Lost productivity (income loss) | . | . | -0.8 | -4,342 |
| Older people (65+ years) | Siblings | 262 | Primary sector | 0.1 | 395 | 0.0 | 18 |
| | | 262 | Hospital care | 0.5 | 1,748 | -0.1 | -207 |
| | | 262 | Filled prescriptions | 0.1 | 314 | -0.0 | -13 |
| | | 150 | Lost productivity (income loss) | . | . | -2.0 | -13,441 |

#### Supplementary Table 42
#### Brain tumors - Incident cohort 2016-2021
#### Identification of populations of relatives

| *Agegroup of index patients in matched population* | | *Closest relative type identified* | | *Available control with same type of closest relative* |
|---|---|---|---|---|
| Children and young people (0-24 years) | n = 591 | Fathers | n = 471 (80%) | 96% |
| | | Mothers | n = 501 (85%) | 96% |
| | | No available closest relative | n = 64 (11%) | n/a |
| Young adults and adults (25-64 years) | n = 5,582 | Children | n = 832 (15%) | 92% |
| | | Parents | n = 522 (9%) | 86% |
| | | Partners | n = 3,552 (64%) | 96% |
| | | Siblings | n = 515 (9%) | 53% |
| | | Pooled | n = 5,421 (97%) | 90% |
| | | No available closest relative | n = 161 (3%) | n/a |
| Older people (65+ years) | n = 6,532 | Children | n = 2,640 (40%) | 95% |
| | | Partners | n = 3,130 (48%) | 96% |
| | | Siblings | n = 99 (2%) | 38% |
| | | Pooled | n = 5,869 (90%) | 94% |
| | | No available closest relative | n = 663 (10%) | n/a |

**Supplementary Table 43**
**Brain tumors - Incident cohort 2016-2021**
**Patient characteristics - cases**

| ***Age strata of index patients*** | ***Characteristic*** | |
|---|---|---|
| Children and young people (0-24 years) | Number of persons, n | 510 (100.0) |
| | Male, n (%) | 264 (51.8) |
| | Age, median (Q1;Q3) | 15.2 (7.2;20.7) |
| | CCI, n (%) | |
| | 0 | 469 (92.0) |
| | 1-2 | * |
| | +3 | * |
| | Education level, n (%) | |
| | No/Unknown education | 206 (40.4) |
| | Primary | 214 (42.0) |
| | Secondary | 81 (15.9) |
| | Tertiary | 9 (1.8) |
| Young adults and adults (25-64 years) | Number of persons, n | 4,912 (100.0) |
| | Male, n (%) | 2,065 (42.0) |
| | Age, median (Q1;Q3) | 53.8 (45.5;59.6) |
| | CCI, n (%) | |
| | 0 | 3,698 (75.3) |
| | 1-2 | 954 (19.4) |
| | +3 | 260 (5.3) |
| | Education level, n (%) | |
| | No/Unknown education | 157 (3.2) |
| | Primary | 944 (19.2) |
| | Secondary | 2,300 (46.8) |
| | Tertiary | 1,511 (30.8) |
| Older people (65+ years) | Number of persons, n | 5,574 (100.0) |
| | Male, n (%) | 2,344 (42.1) |
| | Age, median (Q1;Q3) | 74.4 (70.0;79.7) |
| | CCI, n (%) | |
| | 0 | 2,664 (47.8) |
| | 1-2 | 1,970 (35.3) |
| | +3 | 940 (16.9) |
| | Education level, n (%) | |
| | No/Unknown education | 332 (6.0) |
| | Primary | 1,681 (30.2) |
| | Secondary | 2,507 (45.0) |
| | Tertiary | 1,054 (18.9) |

**Supplementary Table 44**
**Brain tumors - Incident cohort 2016-2021**
**Patient characteristics - relatives - pooled**

| *Age strata of index patients* | *Closest relative type* | *Characteristic of family relatives by agestrata of index patients* | *Closest family relatives of brain disorder patients* | *Comparison relative* |
|---|---|---|---|---|
| Children and young people (0-24 years) | Fathers | Number of persons, n | 454 (100.0) | 454 (100.0) |
| | | Male, n (%) | 454 (100.0) | 454 (100.0) |
| | | Age, median (Q1;Q3) | 47.3 (40.0;52.9) | 47.2 (39.8;52.7) |
| | | CCI, n (%) | | |
| | | 0 | 413 (91.0) | 405 (89.2) |
| | | 1-2 | 36 (7.9) | 43 (9.5) |
| | | +3 | 5 (1.1) | 6 (1.3) |
| | | Education level, n (%) | | |
| | | No/Unknown education | 5 (1.1) | 5 (1.1) |
| | | Primary | 90 (19.8) | 90 (19.8) |
| | | Secondary | 218 (48.0) | 218 (48.0) |
| | | Tertiary | 141 (31.1) | 141 (31.1) |
| | | Relation to patient, n (%) | | |
| | | Father | 454 (100.0) | 454 (100.0) |
| Children and young people (0-24 years) | Mothers | Number of persons, n | 485 (100.0) | 485 (100.0) |
| | | Male, n (%) | 0 (0.0) | 0 (0.0) |
| | | Age, median (Q1;Q3) | 45.2 (38.2;50.3) | 45.3 (38.1;50.5) |
| | | CCI, n (%) | | |
| | | 0 | 428 (88.2) | 427 (88.0) |
| | | 1-2 | 48 (9.9) | * |
| | | +3 | 9 (1.9) | * |
| | | Education level, n (%) | | |
| | | No/Unknown education | 9 (1.9) | 9 (1.9) |
| | | Primary | 80 (16.5) | 80 (16.5) |
| | | Secondary | 218 (44.9) | 218 (44.9) |
| | | Tertiary | 178 (36.7) | 178 (36.7) |
| | | Relation to patient, n (%) | | |
| | | Mother | 485 (100.0) | 485 (100.0) |
| Young adults and adults (25-64 years) | Pooled | Number of persons, n | 4,912 (100.0) | 4,912 (100.0) |
| | | Male, n (%) | 2,634 (53.6) | 2,634 (53.6) |
| | | Age, median (Q1;Q3) | 52.8 (40.1;60.6) | 52.7 (40.3;60.5) |
| | | CCI, n (%) | | |
| | | 0 | 4,088 (83.2) | 4,112 (83.7) |
| | | 1-2 | 688 (14.0) | 660 (13.4) |
| | | +3 | 136 (2.8) | 140 (2.9) |
| | | Education level, n (%) | | |
| | | No/Unknown education | 203 (4.1) | 203 (4.1) |
| | | Primary | 1,073 (21.8) | 1,073 (21.8) |
| | | Secondary | 2,315 (47.1) | 2,315 (47.1) |
| | | Tertiary | 1,321 (26.9) | 1,321 (26.9) |
| | | Relation to patient, n (%) | | |
| | | Mother | 358 (7.3) | 358 (7.3) |
| | | Father | 94 (1.9) | 94 (1.9) |
| | | Partner | 3,415 (69.5) | 3,415 (69.5) |
| | | Child | 772 (15.7) | 772 (15.7) |

**Supplementary Table 44**
**Brain tumors - Incident cohort 2016-2021**
**Patient characteristics - relatives - pooled**

| *Age strata of index patients* | *Closest relative type* | *Characteristic of family relatives by agestrata of index patients* | *Closest family relatives of brain disorder patients* | *Comparison relative* |
|---|---|---|---|---|
| | | Sibling | 273 (5.6) | 273 (5.6) |
| Older people (65+ years) | Pooled | Number of persons, n | 5,574 (100.0) | 5,574 (100.0) |
| | | Male, n (%) | 2,864 (51.4) | 2,864 (51.4) |
| | | Age, median (Q1;Q3) | 64.7 (52.8;73.2) | 65.1 (52.7;73.1) |
| | | CCI, n (%) | | |
| | | 0 | 4,006 (71.9) | 4,031 (72.3) |
| | | 1-2 | 1,205 (21.6) | 1,197 (21.5) |
| | | +3 | 363 (6.5) | 346 (6.2) |
| | | Education level, n (%) | | |
| | | No/Unknown education | 201 (3.6) | 201 (3.6) |
| | | Primary | 1,180 (21.2) | 1,180 (21.2) |
| | | Secondary | 2,758 (49.5) | 2,758 (49.5) |
| | | Tertiary | 1,435 (25.7) | 1,435 (25.7) |
| | | Relation to patient, n (%) | | |
| | | Partner | 3,027 (54.3) | 3,027 (54.3) |
| | | Child | 2,509 (45.0) | 2,509 (45.0) |
| | | Sibling | 38 (0.7) | 38 (0.7) |

**Supplementary Table 45**
**Brain tumors - Incident cohort 2016-2021**
**Patient characteristics - relatives - not pooled**

| *Age strata of index patients* | *Closest relative type* | *Characteristic of family relatives by agestrata of index patients* | *Closest family relatives of brain disorder patients* | *Comparison relative* |
|---|---|---|---|---|
| Children and young people (0-24 years) | Fathers | Number of persons, n | 454 (100.0) | 454 (100.0) |
| | | Male, n (%) | 454 (100.0) | 454 (100.0) |
| | | Age, median (Q1;Q3) | 47.3 (40.0;52.9) | 47.2 (39.8;52.7) |
| | | CCI, n (%) | | |
| | | 0 | 413 (91.0) | 405 (89.2) |
| | | 1-2 | 36 (7.9) | 43 (9.5) |
| | | +3 | 5 (1.1) | 6 (1.3) |
| | | Education level, n (%) | | |
| | | No/Unknown education | 5 (1.1) | 5 (1.1) |
| | | Primary | 90 (19.8) | 90 (19.8) |
| | | Secondary | 218 (48.0) | 218 (48.0) |
| | | Tertiary | 141 (31.1) | 141 (31.1) |
| | | Relation to patient, n (%) | | |
| | | Father | 454 (100.0) | 454 (100.0) |
| Children and young people (0-24 years) | Mothers | Number of persons, n | 485 (100.0) | 485 (100.0) |
| | | Male, n (%) | 0 (0.0) | 0 (0.0) |
| | | Age, median (Q1;Q3) | 45.2 (38.2;50.3) | 45.3 (38.1;50.5) |
| | | CCI, n (%) | | |
| | | 0 | 428 (88.2) | 427 (88.0) |
| | | 1-2 | 48 (9.9) | * |
| | | +3 | 9 (1.9) | * |
| | | Education level, n (%) | | |
| | | No/Unknown education | 9 (1.9) | 9 (1.9) |
| | | Primary | 80 (16.5) | 80 (16.5) |
| | | Secondary | 218 (44.9) | 218 (44.9) |
| | | Tertiary | 178 (36.7) | 178 (36.7) |
| | | Relation to patient, n (%) | | |
| | | Mother | 485 (100.0) | 485 (100.0) |
| Young adults and adults (25-64 years) | Children | Number of persons, n | 772 (100.0) | 772 (100.0) |
| | | Male, n (%) | 401 (51.9) | 401 (51.9) |
| | | Age, median (Q1;Q3) | 26.1 (17.9;34.0) | 25.5 (17.8;34.2) |
| | | CCI, n (%) | | |
| | | 0 | 727 (94.2) | 729 (94.4) |
| | | 1-2 | * | * |
| | | +3 | * | * |
| | | Education level, n (%) | | |
| | | No/Unknown education | 95 (12.3) | 95 (12.3) |
| | | Primary | 304 (39.4) | 304 (39.4) |
| | | Secondary | 248 (32.1) | 248 (32.1) |
| | | Tertiary | 125 (16.2) | 125 (16.2) |
| | | Relation to patient, n (%) | | |
| | | Child | 772 (100.0) | 772 (100.0) |
| Young adults and adults (25-64 years) | Parents | Number of persons, n | 452 (100.0) | 452 (100.0) |
| | | Male, n (%) | 94 (20.8) | 94 (20.8) |
| | | Age, median (Q1;Q3) | 72.2 (63.6;79.9) | 72.4 (63.2;80.0) |

**Supplementary Table 45**
**Brain tumors - Incident cohort 2016-2021**
**Patient characteristics - relatives - not pooled**

| *Age strata of index patients* | *Closest relative type* | *Characteristic of family relatives by agestrata of index patients* | *Closest family relatives of brain disorder patients* | *Comparison relative* |
|---|---|---|---|---|
| | | CCI, n (%) | | |
| | | 0 | 272 (60.2) | 296 (65.5) |
| | | 1-2 | 132 (29.2) | 112 (24.8) |
| | | +3 | 48 (10.6) | 44 (9.7) |
| | | Education level, n (%) | | |
| | | No/Unknown education | 6 (1.3) | 6 (1.3) |
| | | Primary | 141 (31.2) | 141 (31.2) |
| | | Secondary | 219 (48.5) | 219 (48.5) |
| | | Tertiary | 86 (19.0) | 86 (19.0) |
| | | Relation to patient, n (%) | | |
| | | Mother | 358 (79.2) | 358 (79.2) |
| | | Father | 94 (20.8) | 94 (20.8) |
| Young adults and adults (25-64 years) | Partners | Number of persons, n | 3,415 (100.0) | 3,415 (100.0) |
| | | Male, n (%) | 1,975 (57.8) | 1,975 (57.8) |
| | | Age, median (Q1;Q3) | 53.6 (45.7;59.9) | 53.5 (45.9;59.7) |
| | | CCI, n (%) | | |
| | | 0 | 2,863 (83.8) | 2,870 (84.0) |
| | | 1-2 | 475 (13.9) | 461 (13.5) |
| | | +3 | 77 (2.3) | 84 (2.5) |
| | | Education level, n (%) | | |
| | | No/Unknown education | 102 (3.0) | 102 (3.0) |
| | | Primary | 573 (16.8) | 573 (16.8) |
| | | Secondary | 1,682 (49.3) | 1,682 (49.3) |
| | | Tertiary | 1,058 (31.0) | 1,058 (31.0) |
| | | Relation to patient, n (%) | | |
| | | Partner | 3,415 (100.0) | 3,415 (100.0) |
| Young adults and adults (25-64 years) | Siblings | Number of persons, n | 273 (100.0) | 273 (100.0) |
| | | Male, n (%) | 164 (60.1) | 164 (60.1) |
| | | Age, median (Q1;Q3) | 56.2 (51.2;60.9) | 56.4 (51.8;60.8) |
| | | CCI, n (%) | | |
| | | 0 | 226 (82.8) | 217 (79.5) |
| | | 1-2 | 39 (14.3) | 47 (17.2) |
| | | +3 | 8 (2.9) | 9 (3.3) |
| | | Education level, n (%) | | |
| | | No/Unknown education | 0 (0.0) | 0 (0.0) |
| | | Primary | 55 (20.1) | 55 (20.1) |
| | | Secondary | 166 (60.8) | 166 (60.8) |
| | | Tertiary | 52 (19.0) | 52 (19.0) |
| | | Relation to patient, n (%) | | |
| | | Sibling | 273 (100.0) | 273 (100.0) |
| Older people (65+ years) | Children | Number of persons, n | 2,509 (100.0) | 2,509 (100.0) |
| | | Male, n (%) | 1,302 (51.9) | 1,302 (51.9) |
| | | Age, median (Q1;Q3) | 51.7 (45.9;56.8) | 51.5 (45.7;56.7) |
| | | CCI, n (%) | | |
| | | 0 | 2,135 (85.1) | 2,117 (84.4) |

**Supplementary Table 45**
**Brain tumors - Incident cohort 2016-2021**
**Patient characteristics - relatives - not pooled**

| *Age strata of index patients* | *Closest relative type* | *Characteristic of family relatives by agestrata of index patients* | *Closest family relatives of brain disorder patients* | *Comparison relative* |
|---|---|---|---|---|
| | | 1-2 | 326 (13.0) | 345 (13.8) |
| | | +3 | 48 (1.9) | 47 (1.9) |
| | | Education level, n (%) | | |
| | | No/Unknown education | 10 (0.4) | 10 (0.4) |
| | | Primary | 401 (16.0) | 401 (16.0) |
| | | Secondary | 1,302 (51.9) | 1,302 (51.9) |
| | | Tertiary | 796 (31.7) | 796 (31.7) |
| | | Relation to patient, n (%) | | |
| | | Child | 2,509 (100.0) | 2,509 (100.0) |
| Older people (65+ years) | Partners | Number of persons, n | 3,027 (100.0) | 3,027 (100.0) |
| | | Male, n (%) | 1,537 (50.8) | 1,537 (50.8) |
| | | Age, median (Q1;Q3) | 72.6 (68.5;77.0) | 72.4 (68.5;76.8) |
| | | CCI, n (%) | | |
| | | 0 | 1,842 (60.9) | 1,882 (62.2) |
| | | 1-2 | 872 (28.8) | 847 (28.0) |
| | | +3 | 313 (10.3) | 298 (9.8) |
| | | Education level, n (%) | | |
| | | No/Unknown education | 191 (6.3) | 191 (6.3) |
| | | Primary | 774 (25.6) | 774 (25.6) |
| | | Secondary | 1,431 (47.3) | 1,431 (47.3) |
| | | Tertiary | 631 (20.8) | 631 (20.8) |
| | | Relation to patient, n (%) | | |
| | | Partner | 3,027 (100.0) | 3,027 (100.0) |
| Older people (65+ years) | Siblings | Number of persons, n | 38 (100.0) | 38 (100.0) |
| | | Male, n (%) | 25 (65.8) | 25 (65.8) |
| | | Age, median (Q1;Q3) | 61.5 (59.3;65.1) | 61.6 (59.2;64.7) |
| | | CCI, n (%) | | |
| | | 0 | * | * |
| | | 1-2 | * | * |
| | | +3 | * | * |
| | | Education level, n (%) | | |
| | | No/Unknown education | 0 (0.0) | 0 (0.0) |
| | | Primary | 5 (13.2) | 5 (13.2) |
| | | Secondary | 25 (65.8) | 25 (65.8) |
| | | Tertiary | 8 (21.1) | 8 (21.1) |
| | | Relation to patient, n (%) | | |
| | | Sibling | 38 (100.0) | 38 (100.0) |

#### Supplementary Table 46
#### Brain tumors - Incident cohort 2016-2021
#### Cost analysis - pooled

| | | | | *Actual costs* | | *Attributable costs* | |
|---|---|---|---|---|---|---|---|
| ***Age strata of index patients*** | ***Closest relative type*** | ***Number of index patients included in cost component calculations*** | ***Cost component*** | ***Total (million EUR)*** | ***Per person (EUR)*** | ***Total (million EUR)*** | ***Per person (EUR)*** |
| Children and young people (0-24 years) | Fathers | 454 | Primary sector | 0.1 | 233 | 0.0 | 20 |
| | | 454 | Hospital care | 0.6 | 1,359 | 0.1 | 313 |
| | | 454 | Filled prescriptions | 0.1 | 161 | 0.0 | 17 |
| | | 444 | Lost productivity (income loss) | . | . | 0.9 | 1,984 |
| Children and young people (0-24 years) | Mothers | 485 | Primary sector | 0.2 | 334 | 0.0 | 7 |
| | | 485 | Hospital care | 0.8 | 1,648 | 0.2 | 446 |
| | | 485 | Filled prescriptions | 0.1 | 167 | 0.0 | 8 |
| | | 483 | Lost productivity (income loss) | . | . | 1.3 | 2,700 |
| Young adults and adults (25-64 years) | Pooled | 4,912 | Primary sector | 1.6 | 322 | 0.1 | 26 |
| | | 4,912 | Hospital care | 9.4 | 1,919 | 0.2 | 40 |
| | | 4,912 | Filled prescriptions | 1.2 | 250 | 0.0 | 6 |
| | | 3,735 | Lost productivity (income loss) | . | . | 2.1 | 564 |
| Older people (65+ years) | Pooled | 5,574 | Primary sector | 2.3 | 420 | 0.1 | 20 |
| | | 5,574 | Hospital care | 16.2 | 2,932 | -0.2 | -40 |
| | | 5,574 | Filled prescriptions | 2.1 | 378 | -0.2 | -31 |
| | | 2,494 | Lost productivity (income loss) | . | . | 8.5 | 3,404 |

#### Supplementary Table 47
#### Brain tumors - Incident cohort 2016-2021
#### Cost analysis - not pooled

| | | | | *Actual costs* | | *Attributable costs* | |
|---|---|---|---|---|---|---|---|
| *Age strata of index patients* | *Closest relative type* | *Number of index patients included in cost component calculations* | *Cost component* | *Total (million EUR)* | *Per person (EUR)* | *Total (million EUR)* | *Per person (EUR)* |
| Children and young people (0-24 years) | Fathers | 454 | Primary sector | 0.1 | 233 | 0.0 | 20 |
| | | 454 | Hospital care | 0.6 | 1,359 | 0.1 | 313 |
| | | 454 | Filled prescriptions | 0.1 | 161 | 0.0 | 17 |
| | | 444 | Lost productivity (income loss) | . | . | 0.9 | 1,984 |
| Children and young people (0-24 years) | Mothers | 485 | Primary sector | 0.2 | 334 | 0.0 | 7 |
| | | 485 | Hospital care | 0.8 | 1,648 | 0.2 | 446 |
| | | 485 | Filled prescriptions | 0.1 | 167 | 0.0 | 8 |
| | | 483 | Lost productivity (income loss) | . | . | 1.3 | 2,700 |
| Young adults and adults (25-64 years) | Children | 772 | Primary sector | 0.2 | 236 | 0.0 | 30 |
| | | 772 | Hospital care | 1.1 | 1,411 | 0.3 | 388 |
| | | 772 | Filled prescriptions | 0.1 | 126 | 0.0 | 30 |
| | | 553 | Lost productivity (income loss) | . | . | 0.8 | 1,435 |
| Young adults and adults (25-64 years) | Parents | 452 | Primary sector | 0.2 | 506 | 0.0 | 35 |
| | | 452 | Hospital care | 1.6 | 3,708 | 0.1 | 317 |
| | | 452 | Filled prescriptions | 0.3 | 643 | 0.0 | 80 |
| | | 98 | Lost productivity (income loss) | . | . | -0.3 | -2,855 |
| Young adults and adults (25-64 years) | Partners | 3,415 | Primary sector | 1.1 | 320 | 0.1 | 29 |
| | | 3,415 | Hospital care | 6.0 | 1,764 | -0.3 | -87 |
| | | 3,415 | Filled prescriptions | 0.8 | 222 | -0.0 | -13 |
| | | 2,859 | Lost productivity (income loss) | . | . | 3.0 | 1,045 |
| Young adults and adults (25-64 years) | Siblings | 273 | Primary sector | 0.1 | 299 | -0.0 | -42 |
| | | 273 | Hospital care | 0.6 | 2,387 | 0.1 | 201 |
| | | 273 | Filled prescriptions | 0.1 | 311 | 0.0 | 69 |
| | | 225 | Lost productivity (income loss) | . | . | -1.4 | -6,196 |
| Older people (65+ years) | Children | 2,509 | Primary sector | 0.8 | 311 | 0.1 | 23 |
| | | 2,509 | Hospital care | 4.9 | 1,947 | 0.6 | 250 |
| | | 2,509 | Filled prescriptions | 0.6 | 246 | 0.0 | 14 |
| | | 2,346 | Lost productivity (income loss) | . | . | 9.5 | 4,065 |
| Older people (65+ years) | Partners | 3,027 | Primary sector | 1.5 | 512 | 0.1 | 17 |
| | | 3,027 | Hospital care | 11.2 | 3,743 | -0.9 | -292 |
| | | 3,027 | Filled prescriptions | 1.5 | 489 | -0.2 | -69 |
| | | 126 | Lost productivity (income loss) | . | . | -1.0 | -8,189 |
| Older people (65+ years) | Siblings | 38 | Primary sector | 0.0 | 382 | 0.0 | 25 |
| | | 38 | Hospital care | 0.1 | 3,932 | 0.0 | 739 |
| | | 38 | Filled prescriptions | 0.0 | 283 | 0.0 | 43 |
| | | 22 | Lost productivity (income loss) | . | . | -0.0 | -710 |

#### Supplementary Table 48
#### Brain tumors - Prevalent cohort 2021
#### Identification of populations of relatives

| *Agegroup of index patients in matched population* | | *Closest relative type identified* | | *Available control with same type of closest relative* |
|---|---|---|---|---|
| Children and young people (0-24 years) | n = 877 | Fathers | n = 739 (84%) | 95% |
| | | Mothers | n = 780 (89%) | 96% |
| | | No available closest relative | n = 54 (6%) | n/a |
| Young adults and adults (25-64 years) | n = 8,370 | Children | n = 1,169 (14%) | 94% |
| | | Parents | n = 1,031 (12%) | 87% |
| | | Partners | n = 5,198 (62%) | 96% |
| | | Siblings | n = 807 (10%) | 50% |
| | | Pooled | n = 8,205 (98%) | 90% |
| | | No available closest relative | n = 165 (2%) | n/a |
| Older people (65+ years) | n = 9,492 | Children | n = 3,273 (34%) | 95% |
| | | Partners | n = 5,315 (56%) | 96% |
| | | Siblings | n = 173 (2%) | 46% |
| | | Pooled | n = 8,761 (92%) | 95% |
| | | No available closest relative | n = 731 (8%) | n/a |

**Supplementary Table 49**
**Brain tumors - Prevalent cohort 2021**
**Patient characteristics - cases**

| *Age strata of index patients* | *Characteristic* | |
|---|---|---|
| Children and young people (0-24 years) | Number of persons, n | 791 (100.0) |
| | Male, n (%) | 419 (53.0) |
| | Age, median (Q1;Q3) | 17.5 (12.1;21.6) |
| | CCI, n (%) | |
| | 0 | 510 (64.5) |
| | 1-2 | 221 (27.9) |
| | +3 | 60 (7.6) |
| | Education level, n (%) | |
| | No/Unknown education | 237 (30.0) |
| | Primary | 411 (52.0) |
| | Secondary | 131 (16.6) |
| | Tertiary | 12 (1.5) |
| Young adults and adults (25-64 years) | Number of persons, n | 7,454 (100.0) |
| | Male, n (%) | 3,043 (40.8) |
| | Age, median (Q1;Q3) | 53.8 (45.0;59.4) |
| | CCI, n (%) | |
| | 0 | 4,877 (65.4) |
| | 1-2 | 2,014 (27.0) |
| | +3 | 563 (7.6) |
| | Education level, n (%) | |
| | No/Unknown education | 175 (2.3) |
| | Primary | 1,476 (19.8) |
| | Secondary | 3,448 (46.3) |
| | Tertiary | 2,355 (31.6) |
| Older people (65+ years) | Number of persons, n | 8,351 (100.0) |
| | Male, n (%) | 2,950 (35.3) |
| | Age, median (Q1;Q3) | 75.0 (70.5;79.8) |
| | CCI, n (%) | |
| | 0 | 3,633 (43.5) |
| | 1-2 | 3,144 (37.6) |
| | +3 | 1,574 (18.8) |
| | Education level, n (%) | |
| | No/Unknown education | 533 (6.4) |
| | Primary | 2,276 (27.3) |
| | Secondary | 3,733 (44.7) |
| | Tertiary | 1,809 (21.7) |

**Supplementary Table 50**
**Brain tumors - Prevalent cohort 2021**
**Patient characteristics - relatives - pooled**

| *Age strata of index patients* | *Closest relative type* | *Characteristic of family relatives by agestrata of index patients* | *Closest family relatives of brain disorder patients* | *Comparison relative* |
|---|---|---|---|---|
| Children and young people (0-24 years) | Fathers | Number of persons, n | 708 (100.0) | 708 (100.0) |
| | | Male, n (%) | 708 (100.0) | 708 (100.0) |
| | | Age, median (Q1;Q3) | 49.4 (44.3;54.4) | 49.7 (44.2;54.1) |
| | | CCI, n (%) | | |
| | | 0 | 629 (88.8) | 627 (88.6) |
| | | 1-2 | 71 (10.0) | 73 (10.3) |
| | | +3 | 8 (1.1) | 8 (1.1) |
| | | Education level, n (%) | | |
| | | No/Unknown education | 10 (1.4) | 10 (1.4) |
| | | Primary | 137 (19.4) | 137 (19.4) |
| | | Secondary | 362 (51.1) | 362 (51.1) |
| | | Tertiary | 199 (28.1) | 199 (28.1) |
| | | Relation to patient, n (%) | | |
| | | Father | 708 (100.0) | 708 (100.0) |
| Children and young people (0-24 years) | Mothers | Number of persons, n | 749 (100.0) | 749 (100.0) |
| | | Male, n (%) | 0 (0.0) | 0 (0.0) |
| | | Age, median (Q1;Q3) | 47.4 (41.9;51.9) | 47.3 (42.1;52.0) |
| | | CCI, n (%) | | |
| | | 0 | 639 (85.3) | 665 (88.8) |
| | | 1-2 | 97 (13.0) | 79 (10.5) |
| | | +3 | 13 (1.7) | 5 (0.7) |
| | | Education level, n (%) | | |
| | | No/Unknown education | 8 (1.1) | 8 (1.1) |
| | | Primary | 109 (14.6) | 109 (14.6) |
| | | Secondary | 339 (45.3) | 339 (45.3) |
| | | Tertiary | 293 (39.1) | 293 (39.1) |
| | | Relation to patient, n (%) | | |
| | | Mother | 749 (100.0) | 749 (100.0) |
| Young adults and adults (25-64 years) | Pooled | Number of persons, n | 7,454 (100.0) | 7,454 (100.0) |
| | | Male, n (%) | 4,022 (54.0) | 4,022 (54.0) |
| | | Age, median (Q1;Q3) | 53.8 (40.8;61.1) | 54.0 (40.6;61.1) |
| | | CCI, n (%) | | |
| | | 0 | 6,154 (82.6) | 6,182 (82.9) |
| | | 1-2 | 1,107 (14.9) | 1,080 (14.5) |
| | | +3 | 193 (2.6) | 192 (2.6) |
| | | Education level, n (%) | | |
| | | No/Unknown education | 292 (3.9) | 292 (3.9) |
| | | Primary | 1,651 (22.1) | 1,651 (22.1) |
| | | Secondary | 3,486 (46.8) | 3,486 (46.8) |
| | | Tertiary | 2,025 (27.2) | 2,025 (27.2) |
| | | Relation to patient, n (%) | | |
| | | Mother | 694 (9.3) | 694 (9.3) |
| | | Father | 209 (2.8) | 209 (2.8) |
| | | Partner | 5,042 (67.6) | 5,042 (67.6) |
| | | Child | 1,100 (14.8) | 1,100 (14.8) |

**Supplementary Table 50**
**Brain tumors - Prevalent cohort 2021**
**Patient characteristics - relatives - pooled**

| *Age strata of index patients* | *Closest relative type* | *Characteristic of family relatives by agestrata of index patients* | *Closest family relatives of brain disorder patients* | *Comparison relative* |
|---|---|---|---|---|
| | | Sibling | 409 (5.5) | 409 (5.5) |
| Older people (65+ years) | Pooled | Number of persons, n | 8,351 (100.0) | 8,351 (100.0) |
| | | Male, n (%) | 4,482 (53.7) | 4,482 (53.7) |
| | | Age, median (Q1;Q3) | 68.2 (56.0;75.4) | 68.3 (55.8;75.4) |
| | | CCI, n (%) | | |
| | | 0 | 5,656 (67.7) | 5,741 (68.7) |
| | | 1-2 | 2,050 (24.5) | 1,987 (23.8) |
| | | +3 | 645 (7.7) | 623 (7.5) |
| | | Education level, n (%) | | |
| | | No/Unknown education | 379 (4.5) | 379 (4.5) |
| | | Primary | 1,652 (19.8) | 1,652 (19.8) |
| | | Secondary | 4,052 (48.5) | 4,052 (48.5) |
| | | Tertiary | 2,268 (27.2) | 2,268 (27.2) |
| | | Relation to patient, n (%) | | |
| | | Partner | 5,145 (61.6) | 5,145 (61.6) |
| | | Child | 3,126 (37.4) | 3,126 (37.4) |
| | | Sibling | 80 (1.0) | 80 (1.0) |

**Supplementary Table 51**
**Brain tumors - Prevalent cohort 2021**
**Patient characteristics - relatives - not pooled**

| *Age strata of index patients* | *Closest relative type* | *Characteristic of family relatives by agestrata of index patients* | *Closest family relatives of brain disorder patients* | *Comparison relative* |
|---|---|---|---|---|
| Children and young people (0-24 years) | Fathers | Number of persons, n | 708 (100.0) | 708 (100.0) |
| | | Male, n (%) | 708 (100.0) | 708 (100.0) |
| | | Age, median (Q1;Q3) | 49.4 (44.3;54.4) | 49.7 (44.2;54.1) |
| | | CCI, n (%) | | |
| | | 0 | 629 (88.8) | 627 (88.6) |
| | | 1-2 | 71 (10.0) | 73 (10.3) |
| | | +3 | 8 (1.1) | 8 (1.1) |
| | | Education level, n (%) | | |
| | | No/Unknown education | 10 (1.4) | 10 (1.4) |
| | | Primary | 137 (19.4) | 137 (19.4) |
| | | Secondary | 362 (51.1) | 362 (51.1) |
| | | Tertiary | 199 (28.1) | 199 (28.1) |
| | | Relation to patient, n (%) | | |
| | | Father | 708 (100.0) | 708 (100.0) |
| Children and young people (0-24 years) | Mothers | Number of persons, n | 749 (100.0) | 749 (100.0) |
| | | Male, n (%) | 0 (0.0) | 0 (0.0) |
| | | Age, median (Q1;Q3) | 47.4 (41.9;51.9) | 47.3 (42.1;52.0) |
| | | CCI, n (%) | | |
| | | 0 | 639 (85.3) | 665 (88.8) |
| | | 1-2 | 97 (13.0) | 79 (10.5) |
| | | +3 | 13 (1.7) | 5 (0.7) |
| | | Education level, n (%) | | |
| | | No/Unknown education | 8 (1.1) | 8 (1.1) |
| | | Primary | 109 (14.6) | 109 (14.6) |
| | | Secondary | 339 (45.3) | 339 (45.3) |
| | | Tertiary | 293 (39.1) | 293 (39.1) |
| | | Relation to patient, n (%) | | |
| | | Mother | 749 (100.0) | 749 (100.0) |
| Young adults and adults (25-64 years) | Children | Number of persons, n | 1,100 (100.0) | 1,100 (100.0) |
| | | Male, n (%) | 591 (53.7) | 591 (53.7) |
| | | Age, median (Q1;Q3) | 24.8 (16.9;31.9) | 24.3 (17.0;31.7) |
| | | CCI, n (%) | | |
| | | 0 | 1,031 (93.7) | 1,027 (93.4) |
| | | 1-2 | * | 68 (6.2) |
| | | +3 | * | 5 (0.5) |
| | | Education level, n (%) | | |
| | | No/Unknown education | 152 (13.8) | 152 (13.8) |
| | | Primary | 448 (40.7) | 448 (40.7) |
| | | Secondary | 319 (29.0) | 319 (29.0) |
| | | Tertiary | 181 (16.5) | 181 (16.5) |
| | | Relation to patient, n (%) | | |
| | | Child | 1,100 (100.0) | 1,100 (100.0) |
| Young adults and adults (25-64 years) | Parents | Number of persons, n | 903 (100.0) | 903 (100.0) |
| | | Male, n (%) | 209 (23.1) | 209 (23.1) |
| | | Age, median (Q1;Q3) | 69.9 (60.4;77.8) | 70.0 (60.5;77.6) |

### Supplementary Table 51
### Brain tumors - Prevalent cohort 2021
### Patient characteristics - relatives - not pooled

| *Age strata of index patients* | *Closest relative type* | *Characteristic of family relatives by agestrata of index patients* | *Closest family relatives of brain disorder patients* | *Comparison relative* |
|---|---|---|---|---|
| | | CCI, n (%) | | |
| | | 0 | 574 (63.6) | 595 (65.9) |
| | | 1-2 | 259 (28.7) | 247 (27.4) |
| | | +3 | 70 (7.8) | 61 (6.8) |
| | | Education level, n (%) | | |
| | | No/Unknown education | 17 (1.9) | 17 (1.9) |
| | | Primary | 293 (32.4) | 293 (32.4) |
| | | Secondary | 427 (47.3) | 427 (47.3) |
| | | Tertiary | 166 (18.4) | 166 (18.4) |
| | | Relation to patient, n (%) | | |
| | | Mother | 694 (76.9) | 694 (76.9) |
| | | Father | 209 (23.1) | 209 (23.1) |
| Young adults and adults (25-64 years) | Partners | Number of persons, n | 5,042 (100.0) | 5,042 (100.0) |
| | | Male, n (%) | 2,992 (59.3) | 2,992 (59.3) |
| | | Age, median (Q1;Q3) | 54.3 (46.3;60.4) | 54.5 (46.2;60.2) |
| | | CCI, n (%) | | |
| | | 0 | 4,224 (83.8) | 4,219 (83.7) |
| | | 1-2 | 712 (14.1) | 704 (14.0) |
| | | +3 | 106 (2.1) | 119 (2.4) |
| | | Education level, n (%) | | |
| | | No/Unknown education | 122 (2.4) | 122 (2.4) |
| | | Primary | 813 (16.1) | 813 (16.1) |
| | | Secondary | 2,500 (49.6) | 2,500 (49.6) |
| | | Tertiary | 1,607 (31.9) | 1,607 (31.9) |
| | | Relation to patient, n (%) | | |
| | | Partner | 5,042 (100.0) | 5,042 (100.0) |
| Young adults and adults (25-64 years) | Siblings | Number of persons, n | 409 (100.0) | 409 (100.0) |
| | | Male, n (%) | 230 (56.2) | 230 (56.2) |
| | | Age, median (Q1;Q3) | 56.6 (49.3;61.6) | 56.7 (48.8;61.5) |
| | | CCI, n (%) | | |
| | | 0 | 325 (79.5) | 341 (83.4) |
| | | 1-2 | 70 (17.1) | 61 (14.9) |
| | | +3 | 14 (3.4) | 7 (1.7) |
| | | Education level, n (%) | | |
| | | No/Unknown education | * | * |
| | | Primary | 97 (23.7) | 97 (23.7) |
| | | Secondary | 240 (58.7) | 240 (58.7) |
| | | Tertiary | * | * |
| | | Relation to patient, n (%) | | |
| | | Sibling | 409 (100.0) | 409 (100.0) |
| Older people (65+ years) | Children | Number of persons, n | 3,126 (100.0) | 3,126 (100.0) |
| | | Male, n (%) | 1,518 (48.6) | 1,518 (48.6) |
| | | Age, median (Q1;Q3) | 52.6 (46.6;58.0) | 52.8 (46.4;57.6) |
| | | CCI, n (%) | | |
| | | 0 | 2,621 (83.8) | 2,633 (84.2) |

**Supplementary Table 51**
**Brain tumors - Prevalent cohort 2021**
**Patient characteristics - relatives - not pooled**

| *Age strata of index patients* | *Closest relative type* | *Characteristic of family relatives by agestrata of index patients* | *Closest family relatives of brain disorder patients* | *Comparison relative* |
|---|---|---|---|---|
| | | 1-2 | 444 (14.2) | 426 (13.6) |
| | | +3 | 61 (2.0) | 67 (2.1) |
| | | Education level, n (%) | | |
| | | No/Unknown education | 13 (0.4) | 13 (0.4) |
| | | Primary | 501 (16.0) | 501 (16.0) |
| | | Secondary | 1,471 (47.1) | 1,471 (47.1) |
| | | Tertiary | 1,141 (36.5) | 1,141 (36.5) |
| | | Relation to patient, n (%) | | |
| | | Child | 3,126 (100.0) | 3,126 (100.0) |
| Older people (65+ years) | Partners | Number of persons, n | 5,145 (100.0) | 5,145 (100.0) |
| | | Male, n (%) | 2,915 (56.7) | 2,915 (56.7) |
| | | Age, median (Q1;Q3) | 73.9 (69.5;78.2) | 74.0 (69.5;78.2) |
| | | CCI, n (%) | | |
| | | 0 | 2,978 (57.9) | 3,044 (59.2) |
| | | 1-2 | 1,585 (30.8) | 1,547 (30.1) |
| | | +3 | 582 (11.3) | 554 (10.8) |
| | | Education level, n (%) | | |
| | | No/Unknown education | 366 (7.1) | 366 (7.1) |
| | | Primary | 1,141 (22.2) | 1,141 (22.2) |
| | | Secondary | 2,521 (49.0) | 2,521 (49.0) |
| | | Tertiary | 1,117 (21.7) | 1,117 (21.7) |
| | | Relation to patient, n (%) | | |
| | | Partner | 5,145 (100.0) | 5,145 (100.0) |
| Older people (65+ years) | Siblings | Number of persons, n | 80 (100.0) | 80 (100.0) |
| | | Male, n (%) | 49 (61.3) | 49 (61.3) |
| | | Age, median (Q1;Q3) | 62.5 (60.3;66.0) | 63.3 (60.1;65.0) |
| | | CCI, n (%) | | |
| | | 0 | 57 (71.3) | 64 (80.0) |
| | | 1-2 | * | * |
| | | +3 | * | * |
| | | Education level, n (%) | | |
| | | No/Unknown education | 0 (0.0) | 0 (0.0) |
| | | Primary | 10 (12.5) | 10 (12.5) |
| | | Secondary | 60 (75.0) | 60 (75.0) |
| | | Tertiary | 10 (12.5) | 10 (12.5) |
| | | Relation to patient, n (%) | | |
| | | Sibling | 80 (100.0) | 80 (100.0) |

**Supplementary Table 52**
**Brain tumors - Prevalent cohort 2021**
**Cost analysis - pooled**

| *Age strata of index patients* | *Closest relative type* | *Number of index patients included in cost component calculations* | *Cost component* | *Actual costs* | | *Attributable costs* | |
|---|---|---|---|---|---|---|---|
| | | | | *Total (million EUR)* | *Per person (EUR)* | *Total (million EUR)* | *Per person (EUR)* |
| Children and young people (0-24 years) | Fathers | 708 | Primary sector | 0.2 | 225 | 0.0 | 3 |
| | | 708 | Hospital care | 0.8 | 1,172 | -0.5 | -656 |
| | | 708 | Filled prescriptions | 0.1 | 149 | -0.0 | -7 |
| | | 689 | Lost productivity (income loss) | . | . | 6.2 | 8,954 |
| Children and young people (0-24 years) | Mothers | 749 | Primary sector | 0.3 | 350 | 0.0 | 4 |
| | | 749 | Hospital care | 1.3 | 1,724 | -0.1 | -175 |
| | | 749 | Filled prescriptions | 0.1 | 172 | -0.0 | -17 |
| | | 745 | Lost productivity (income loss) | . | . | -1.0 | -1,404 |
| Young adults and adults (25-64 years) | Pooled | 7,454 | Primary sector | 2.4 | 326 | 0.2 | 24 |
| | | 7,454 | Hospital care | 15.9 | 2,135 | 1.7 | 223 |
| | | 7,454 | Filled prescriptions | 2.1 | 281 | 0.2 | 29 |
| | | 5,619 | Lost productivity (income loss) | . | . | 10.2 | 1,819 |
| Older people (65+ years) | Pooled | 8,351 | Primary sector | 3.7 | 447 | 0.2 | 19 |
| | | 8,351 | Hospital care | 27.1 | 3,271 | 1.2 | 144 |
| | | 8,351 | Filled prescriptions | 3.6 | 431 | 0.0 | 3 |
| | | 3,082 | Lost productivity (income loss) | . | . | 2.0 | 656 |

#### Supplementary Table 53
#### Brain tumors - Prevalent cohort 2021
#### Cost analysis - not pooled

| | | | | *Actual costs* | | *Attributable costs* | |
|---|---|---|---|---|---|---|---|
| *Age strata of index patients* | *Closest relative type* | *Number of index patients included in cost component calculations* | *Cost component* | *Total (million EUR)* | *Per person (EUR)* | *Total (million EUR)* | *Per person (EUR)* |
| Children and young people (0-24 years) | Fathers | 708 | Primary sector | 0.2 | 225 | 0.0 | 3 |
| | | 708 | Hospital care | 0.8 | 1,172 | -0.5 | -656 |
| | | 708 | Filled prescriptions | 0.1 | 149 | -0.0 | -7 |
| | | 689 | Lost productivity (income loss) | . | . | 6.2 | 8,954 |
| Children and young people (0-24 years) | Mothers | 749 | Primary sector | 0.3 | 350 | 0.0 | 4 |
| | | 749 | Hospital care | 1.3 | 1,724 | -0.1 | -175 |
| | | 749 | Filled prescriptions | 0.1 | 172 | -0.0 | -17 |
| | | 745 | Lost productivity (income loss) | . | . | -1.0 | -1,404 |
| Young adults and adults (25-64 years) | Children | 1,100 | Primary sector | 0.3 | 244 | 0.0 | 41 |
| | | 1,100 | Hospital care | 2.2 | 1,966 | 0.5 | 462 |
| | | 1,100 | Filled prescriptions | 0.2 | 143 | 0.0 | 42 |
| | | 732 | Lost productivity (income loss) | . | . | 0.3 | 369 |
| Young adults and adults (25-64 years) | Parents | 903 | Primary sector | 0.4 | 493 | 0.0 | 50 |
| | | 903 | Hospital care | 3.4 | 3,815 | 0.7 | 806 |
| | | 903 | Filled prescriptions | 0.5 | 560 | 0.1 | 98 |
| | | 284 | Lost productivity (income loss) | . | . | 0.8 | 2,708 |
| Young adults and adults (25-64 years) | Partners | 5,042 | Primary sector | 1.6 | 314 | 0.1 | 14 |
| | | 5,042 | Hospital care | 9.4 | 1,873 | 0.2 | 41 |
| | | 5,042 | Filled prescriptions | 1.2 | 242 | -0.0 | -7 |
| | | 4,271 | Lost productivity (income loss) | . | . | 8.4 | 1,976 |
| Young adults and adults (25-64 years) | Siblings | 409 | Primary sector | 0.1 | 321 | 0.0 | 41 |
| | | 409 | Hospital care | 0.9 | 2,160 | 0.2 | 538 |
| | | 409 | Filled prescriptions | 0.2 | 533 | 0.1 | 279 |
| | | 332 | Lost productivity (income loss) | . | . | 0.7 | 2,229 |
| Older people (65+ years) | Children | 3,126 | Primary sector | 1.0 | 320 | 0.0 | 14 |
| | | 3,126 | Hospital care | 5.8 | 1,868 | 0.7 | 218 |
| | | 3,126 | Filled prescriptions | 0.8 | 255 | 0.0 | 2 |
| | | 2,857 | Lost productivity (income loss) | . | . | 1.2 | 403 |
| Older people (65+ years) | Partners | 5,145 | Primary sector | 2.7 | 527 | 0.1 | 23 |
| | | 5,145 | Hospital care | 20.9 | 4,117 | 0.3 | 54 |
| | | 5,145 | Filled prescriptions | 2.7 | 541 | 0.0 | 2 |
| | | 183 | Lost productivity (income loss) | . | . | 0.4 | 1,977 |
| Older people (65+ years) | Siblings | 80 | Primary sector | 0.0 | 393 | -0.0 | -11 |
| | | 80 | Hospital care | 0.3 | 4,293 | 0.2 | 3,037 |
| | | 80 | Filled prescriptions | 0.0 | 301 | 0.0 | 91 |
| | | 42 | Lost productivity (income loss) | . | . | 0.5 | 12,130 |

#### Supplementary Table 54
#### Cerebral palsy - Incident cohort 2016-2021
#### Identification of populations of relatives

| ***Agegroup of index patients in matched population*** | | ***Closest relative type identified*** | | ***Available control with same type of closest relative*** |
|---|---|---|---|---|
| Children and young people (0-24 years) | n = 1,214 | Fathers | n = 873 (72%) | 97% |
| | | Mothers | n = 923 (76%) | 96% |
| | | No available closest relative | n = 224 (18%) | n/a |
| Young adults and adults (25-64 years) | n = 572 | Children | n = 60 (10%) | 93% |
| | | Parents | n = 154 (27%) | 84% |
| | | Partners | n = 182 (32%) | 92% |
| | | Siblings | n = 141 (25%) | 52% |
| | | Pooled | n = 537 (94%) | 79% |
| | | No available closest relative | n = 35 (6%) | n/a |
| Older people (65+ years) | n = 339 | Children | n = 122 (36%) | 96% |
| | | Partners | n = 120 (35%) | 93% |
| | | Siblings | n = 7 (2%) | 57% |
| | | Pooled | n = 249 (73%) | 93% |
| | | No available closest relative | n = 90 (27%) | n/a |

### Supplementary Table 55
### Cerebral palsy - Incident cohort 2016-2021
### Patient characteristics - cases

| *Age strata of index patients* | *Characteristic* | |
|---|---|---|
| Children and young people (0-24 years) | Number of persons, n | 959 (100.0) |
| | Male, n (%) | 567 (59.1) |
| | Age, median (Q1;Q3) | 2.7 (1.3;8.1) |
| | CCI, n (%) | |
| | 0 | 750 (78.2) |
| | 1-2 | 173 (18.0) |
| | +3 | 36 (3.8) |
| | Education level, n (%) | |
| | No/Unknown education | 842 (87.8) |
| | Primary | 109 (11.4) |
| | Secondary | 8 (0.8) |
| | Tertiary | 0 (0.0) |
| Young adults and adults (25-64 years) | Number of persons, n | 429 (100.0) |
| | Male, n (%) | 218 (50.8) |
| | Age, median (Q1;Q3) | 50.7 (40.1;57.9) |
| | CCI, n (%) | |
| | 0 | 236 (55.0) |
| | 1-2 | 134 (31.2) |
| | +3 | 59 (13.8) |
| | Education level, n (%) | |
| | No/Unknown education | 17 (4.0) |
| | Primary | 209 (48.7) |
| | Secondary | 131 (30.5) |
| | Tertiary | 72 (16.8) |
| Older people (65+ years) | Number of persons, n | 234 (100.0) |
| | Male, n (%) | 124 (53.0) |
| | Age, median (Q1;Q3) | 75.2 (71.6;80.8) |
| | CCI, n (%) | |
| | 0 | 70 (29.9) |
| | 1-2 | 96 (41.0) |
| | +3 | 68 (29.1) |
| | Education level, n (%) | |
| | No/Unknown education | 15 (6.4) |
| | Primary | 90 (38.5) |
| | Secondary | 105 (44.9) |
| | Tertiary | 24 (10.3) |

**Supplementary Table 56**
**Cerebral palsy - Incident cohort 2016-2021**
**Patient characteristics - relatives - pooled**

| *Age strata of index patients* | *Closest relative type* | *Characteristic of family relatives by agestrata of index patients* | *Closest family relatives of brain disorder patients* | *Comparison relative* |
|---|---|---|---|---|
| Children and young people (0-24 years) | Fathers | Number of persons, n | 848 (100.0) | 848 (100.0) |
| | | Male, n (%) | 848 (100.0) | 848 (100.0) |
| | | Age, median (Q1;Q3) | 36.9 (32.2;43.5) | 37.0 (32.8;43.3) |
| | | CCI, n (%) | | |
| | | 0 | 791 (93.3) | 791 (93.3) |
| | | 1-2 | 52 (6.1) | * |
| | | +3 | 5 (0.6) | * |
| | | Education level, n (%) | | |
| | | No/Unknown education | 32 (3.8) | 32 (3.8) |
| | | Primary | 196 (23.1) | 196 (23.1) |
| | | Secondary | 378 (44.6) | 378 (44.6) |
| | | Tertiary | 242 (28.5) | 242 (28.5) |
| | | Relation to patient, n (%) | | |
| | | Father | 848 (100.0) | 848 (100.0) |
| Children and young people (0-24 years) | Mothers | Number of persons, n | 894 (100.0) | 894 (100.0) |
| | | Male, n (%) | 0 (0.0) | 0 (0.0) |
| | | Age, median (Q1;Q3) | 34.8 (30.2;40.4) | 34.7 (30.4;39.7) |
| | | CCI, n (%) | | |
| | | 0 | 806 (90.2) | 814 (91.1) |
| | | 1-2 | 82 (9.2) | * |
| | | +3 | 6 (0.7) | * |
| | | Education level, n (%) | | |
| | | No/Unknown education | 45 (5.0) | 45 (5.0) |
| | | Primary | 190 (21.3) | 190 (21.3) |
| | | Secondary | 309 (34.6) | 309 (34.6) |
| | | Tertiary | 350 (39.1) | 350 (39.1) |
| | | Relation to patient, n (%) | | |
| | | Mother | 894 (100.0) | 894 (100.0) |
| Young adults and adults (25-64 years) | Pooled | Number of persons, n | 429 (100.0) | 429 (100.0) |
| | | Male, n (%) | 191 (44.5) | 191 (44.5) |
| | | Age, median (Q1;Q3) | 56.0 (42.4;65.4) | 56.3 (42.1;64.4) |
| | | CCI, n (%) | | |
| | | 0 | 327 (76.2) | 345 (80.4) |
| | | 1-2 | 84 (19.6) | 71 (16.6) |
| | | +3 | 18 (4.2) | 13 (3.0) |
| | | Education level, n (%) | | |
| | | No/Unknown education | 14 (3.3) | 14 (3.3) |
| | | Primary | 110 (25.6) | 110 (25.6) |
| | | Secondary | 203 (47.3) | 203 (47.3) |
| | | Tertiary | 102 (23.8) | 102 (23.8) |
| | | Relation to patient, n (%) | | |
| | | Mother | 99 (23.1) | 99 (23.1) |
| | | Father | 31 (7.2) | 31 (7.2) |
| | | Partner | 169 (39.4) | 169 (39.4) |
| | | Child | 56 (13.1) | 56 (13.1) |

**Supplementary Table 56**
**Cerebral palsy - Incident cohort 2016-2021**
**Patient characteristics - relatives - pooled**

| *Age strata of index patients* | *Closest relative type* | *Characteristic of family relatives by agestrata of index patients* | *Closest family relatives of brain disorder patients* | *Comparison relative* |
|---|---|---|---|---|
| | | Sibling | 74 (17.2) | 74 (17.2) |
| Older people (65+ years) | Pooled | Number of persons, n | 234 (100.0) | 234 (100.0) |
| | | Male, n (%) | 106 (45.3) | 106 (45.3) |
| | | Age, median (Q1;Q3) | 61.1 (51.9;73.2) | 62.5 (51.4;72.8) |
| | | CCI, n (%) | | |
| | | 0 | 168 (71.8) | 173 (73.9) |
| | | 1-2 | 47 (20.1) | 54 (23.1) |
| | | +3 | 19 (8.1) | 7 (3.0) |
| | | Education level, n (%) | | |
| | | No/Unknown education | 7 (3.0) | 7 (3.0) |
| | | Primary | 59 (25.2) | 59 (25.2) |
| | | Secondary | 120 (51.3) | 120 (51.3) |
| | | Tertiary | 48 (20.5) | 48 (20.5) |
| | | Relation to patient, n (%) | | |
| | | Partner | * | * |
| | | Child | 118 (50.4) | 118 (50.4) |
| | | Sibling | * | * |

**Supplementary Table 57**
**Cerebral palsy - Incident cohort 2016-2021**
**Patient characteristics - relatives - not pooled**

| *Age strata of index patients* | *Closest relative type* | *Characteristic of family relatives by agestrata of index patients* | *Closest family relatives of brain disorder patients* | *Comparison relative* |
|---|---|---|---|---|
| Children and young people (0-24 years) | Fathers | Number of persons, n | 848 (100.0) | 848 (100.0) |
| | | Male, n (%) | 848 (100.0) | 848 (100.0) |
| | | Age, median (Q1;Q3) | 36.9 (32.2;43.5) | 37.0 (32.8;43.3) |
| | | CCI, n (%) | | |
| | | 0 | 791 (93.3) | 791 (93.3) |
| | | 1-2 | 52 (6.1) | * |
| | | +3 | 5 (0.6) | * |
| | | Education level, n (%) | | |
| | | No/Unknown education | 32 (3.8) | 32 (3.8) |
| | | Primary | 196 (23.1) | 196 (23.1) |
| | | Secondary | 378 (44.6) | 378 (44.6) |
| | | Tertiary | 242 (28.5) | 242 (28.5) |
| | | Relation to patient, n (%) | | |
| | | Father | 848 (100.0) | 848 (100.0) |
| Children and young people (0-24 years) | Mothers | Number of persons, n | 894 (100.0) | 894 (100.0) |
| | | Male, n (%) | 0 (0.0) | 0 (0.0) |
| | | Age, median (Q1;Q3) | 34.8 (30.2;40.4) | 34.7 (30.4;39.7) |
| | | CCI, n (%) | | |
| | | 0 | 806 (90.2) | 814 (91.1) |
| | | 1-2 | 82 (9.2) | * |
| | | +3 | 6 (0.7) | * |
| | | Education level, n (%) | | |
| | | No/Unknown education | 45 (5.0) | 45 (5.0) |
| | | Primary | 190 (21.3) | 190 (21.3) |
| | | Secondary | 309 (34.6) | 309 (34.6) |
| | | Tertiary | 350 (39.1) | 350 (39.1) |
| | | Relation to patient, n (%) | | |
| | | Mother | 894 (100.0) | 894 (100.0) |
| Young adults and adults (25-64 years) | Children | Number of persons, n | 56 (100.0) | 56 (100.0) |
| | | Male, n (%) | 27 (48.2) | 27 (48.2) |
| | | Age, median (Q1;Q3) | 28.3 (22.4;34.0) | 28.7 (22.1;33.4) |
| | | CCI, n (%) | | |
| | | 0 | * | * |
| | | 1-2 | * | * |
| | | +3 | 0 (0.0) | 0 (0.0) |
| | | Education level, n (%) | | |
| | | No/Unknown education | 6 (10.7) | 6 (10.7) |
| | | Primary | 20 (35.7) | 20 (35.7) |
| | | Secondary | 17 (30.4) | 17 (30.4) |
| | | Tertiary | 13 (23.2) | 13 (23.2) |
| | | Relation to patient, n (%) | | |
| | | Child | 56 (100.0) | 56 (100.0) |
| Young adults and adults (25-64 years) | Parents | Number of persons, n | 130 (100.0) | 130 (100.0) |
| | | Male, n (%) | 31 (23.8) | 31 (23.8) |
| | | Age, median (Q1;Q3) | 69.2 (62.3;76.5) | 69.7 (62.0;75.8) |

**Supplementary Table 57**
**Cerebral palsy - Incident cohort 2016-2021**
**Patient characteristics - relatives - not pooled**

| *Age strata of index patients* | *Closest relative type* | *Characteristic of family relatives by agestrata of index patients* | *Closest family relatives of brain disorder patients* | *Comparison relative* |
|---|---|---|---|---|
| | | CCI, n (%) | | |
| | | 0 | 82 (63.1) | 85 (65.4) |
| | | 1-2 | 35 (26.9) | 38 (29.2) |
| | | +3 | 13 (10.0) | 7 (5.4) |
| | | Education level, n (%) | | |
| | | No/Unknown education | * | * |
| | | Primary | 40 (30.8) | 40 (30.8) |
| | | Secondary | 69 (53.1) | 69 (53.1) |
| | | Tertiary | * | * |
| | | Relation to patient, n (%) | | |
| | | Mother | 99 (76.2) | 99 (76.2) |
| | | Father | 31 (23.8) | 31 (23.8) |
| Young adults and adults (25-64 years) | Partners | Number of persons, n | 169 (100.0) | 169 (100.0) |
| | | Male, n (%) | 94 (55.6) | 94 (55.6) |
| | | Age, median (Q1;Q3) | 53.3 (43.2;60.7) | 53.6 (43.6;59.8) |
| | | CCI, n (%) | | |
| | | 0 | 126 (74.6) | 145 (85.8) |
| | | 1-2 | * | 19 (11.2) |
| | | +3 | * | 5 (3.0) |
| | | Education level, n (%) | | |
| | | No/Unknown education | 6 (3.6) | 6 (3.6) |
| | | Primary | 32 (18.9) | 32 (18.9) |
| | | Secondary | 80 (47.3) | 80 (47.3) |
| | | Tertiary | 51 (30.2) | 51 (30.2) |
| | | Relation to patient, n (%) | | |
| | | Partner | 169 (100.0) | 169 (100.0) |
| Young adults and adults (25-64 years) | Siblings | Number of persons, n | 74 (100.0) | 74 (100.0) |
| | | Male, n (%) | 39 (52.7) | 39 (52.7) |
| | | Age, median (Q1;Q3) | 55.3 (46.0;59.3) | 54.6 (47.8;59.4) |
| | | CCI, n (%) | | |
| | | 0 | * | 61 (82.4) |
| | | 1-2 | * | * |
| | | +3 | * | * |
| | | Education level, n (%) | | |
| | | No/Unknown education | 0 (0.0) | 0 (0.0) |
| | | Primary | 18 (24.3) | 18 (24.3) |
| | | Secondary | 37 (50.0) | 37 (50.0) |
| | | Tertiary | 19 (25.7) | 19 (25.7) |
| | | Relation to patient, n (%) | | |
| | | Sibling | 74 (100.0) | 74 (100.0) |
| Older people (65+ years) | Children | Number of persons, n | 118 (100.0) | 118 (100.0) |
| | | Male, n (%) | 66 (55.9) | 66 (55.9) |
| | | Age, median (Q1;Q3) | 51.9 (47.8;56.8) | 51.7 (47.5;56.1) |
| | | CCI, n (%) | | |
| | | 0 | 102 (86.4) | 102 (86.4) |

#### Supplementary Table 57
#### Cerebral palsy - Incident cohort 2016-2021
#### Patient characteristics - relatives - not pooled

| *Age strata of index patients* | *Closest relative type* | *Characteristic of family relatives by agestrata of index patients* | *Closest family relatives of brain disorder patients* | *Comparison relative* |
|---|---|---|---|---|
| | | 1-2 | 11 (9.3) | * |
| | | +3 | 5 (4.2) | * |
| | | Education level, n (%) | | |
| | | No/Unknown education | 0 (0.0) | 0 (0.0) |
| | | Primary | 18 (15.3) | 18 (15.3) |
| | | Secondary | 69 (58.5) | 69 (58.5) |
| | | Tertiary | 31 (26.3) | 31 (26.3) |
| | | Relation to patient, n (%) | | |
| | | Child | 118 (100.0) | 118 (100.0) |
| Older people (65+ years) | Partners | Number of persons, n | 112 (100.0) | 112 (100.0) |
| | | Male, n (%) | 37 (33.0) | 37 (33.0) |
| | | Age, median (Q1;Q3) | 73.4 (68.6;79.3) | 73.0 (67.9;78.0) |
| | | CCI, n (%) | | |
| | | 0 | 62 (55.4) | 68 (60.7) |
| | | 1-2 | 36 (32.1) | 39 (34.8) |
| | | +3 | 14 (12.5) | 5 (4.5) |
| | | Education level, n (%) | | |
| | | No/Unknown education | 7 (6.3) | 7 (6.3) |
| | | Primary | 39 (34.8) | 39 (34.8) |
| | | Secondary | 49 (43.8) | 49 (43.8) |
| | | Tertiary | 17 (15.2) | 17 (15.2) |
| | | Relation to patient, n (%) | | |
| | | Partner | 112 (100.0) | 112 (100.0) |
| Older people (65+ years) | Siblings | Number of persons, n | * | * |
| | | Male, n (%) | * | * |
| | | Age, median (Q1;Q3) | 63.2 (59.2;70.0) | 63.5 (63.1;67.8) |
| | | CCI, n (%) | | |
| | | 0 | * | * |
| | | 1-2 | 0 (0.0) | * |
| | | +3 | 0 (0.0) | 0 (0.0) |
| | | Education level, n (%) | | |
| | | No/Unknown education | 0 (0.0) | 0 (0.0) |
| | | Primary | * | * |
| | | Secondary | * | * |
| | | Tertiary | 0 (0.0) | 0 (0.0) |
| | | Relation to patient, n (%) | | |
| | | Sibling | 4 (100.0) | 4 (100.0) |

#### Supplementary Table 58
#### Cerebral palsy - Incident cohort 2016-2021
#### Cost analysis - pooled

| Age strata of index patients | Closest relative type | Number of index patients included in cost component calculations | Cost component | Actual costs | | Attributable costs | |
|---|---|---|---|---|---|---|---|
| | | | | Total (million EUR) | Per person (EUR) | Total (million EUR) | Per person (EUR) |
| Children and young people (0-24 years) | Fathers | 848 | Primary sector | 0.2 | 195 | 0.0 | 19 |
| | | 848 | Hospital care | 0.8 | 934 | -0.1 | -135 |
| | | 848 | Filled prescriptions | 0.1 | 136 | 0.0 | 51 |
| | | 842 | Lost productivity (income loss) | . | . | 1.3 | 1,507 |
| Children and young people (0-24 years) | Mothers | 894 | Primary sector | 0.3 | 351 | 0.0 | 36 |
| | | 894 | Hospital care | 1.7 | 1,888 | 0.1 | 68 |
| | | 894 | Filled prescriptions | 0.1 | 133 | 0.0 | 10 |
| | | 893 | Lost productivity (income loss) | . | . | 1.9 | 2,115 |
| Young adults and adults (25-64 years) | Pooled | 429 | Primary sector | 0.2 | 392 | 0.0 | 48 |
| | | 429 | Hospital care | 1.0 | 2,232 | -0.0 | -116 |
| | | 429 | Filled prescriptions | 0.2 | 368 | 0.0 | 18 |
| | | 282 | Lost productivity (income loss) | . | . | 1.2 | 4,364 |
| Older people (65+ years) | Pooled | 234 | Primary sector | 0.1 | 402 | 0.0 | 7 |
| | | 234 | Hospital care | 0.5 | 2,028 | -0.0 | -161 |
| | | 234 | Filled prescriptions | 0.1 | 350 | -0.0 | -64 |
| | | 118 | Lost productivity (income loss) | . | . | 1.3 | 10,739 |

#### Supplementary Table 59
#### Cerebral palsy - Incident cohort 2016-2021
#### Cost analysis - not pooled

| *Age strata of index patients* | *Closest relative type* | *Number of index patients included in cost component calculations* | *Cost component* | *Actual costs* Total (million EUR) | *Actual costs* Per person (EUR) | *Attributable costs* Total (million EUR) | *Attributable costs* Per person (EUR) |
|---|---|---|---|---|---|---|---|
| Children and young people (0-24 years) | Fathers | 848 | Primary sector | 0.2 | 195 | 0.0 | 19 |
| | | 848 | Hospital care | 0.8 | 934 | -0.1 | -135 |
| | | 848 | Filled prescriptions | 0.1 | 136 | 0.0 | 51 |
| | | 842 | Lost productivity (income loss) | . | . | 1.3 | 1,507 |
| Children and young people (0-24 years) | Mothers | 894 | Primary sector | 0.3 | 351 | 0.0 | 36 |
| | | 894 | Hospital care | 1.7 | 1,888 | 0.1 | 68 |
| | | 894 | Filled prescriptions | 0.1 | 133 | 0.0 | 10 |
| | | 893 | Lost productivity (income loss) | . | . | 1.9 | 2,115 |
| Young adults and adults (25-64 years) | Children | 56 | Primary sector | 0.0 | 311 | 0.0 | 69 |
| | | 56 | Hospital care | 0.1 | 1,806 | 0.1 | 1,236 |
| | | 56 | Filled prescriptions | 0.0 | 106 | -0.0 | -182 |
| | | 45 | Lost productivity (income loss) | . | . | 0.4 | 8,272 |
| Young adults and adults (25-64 years) | Parents | 130 | Primary sector | 0.1 | 490 | 0.0 | 31 |
| | | 130 | Hospital care | 0.4 | 3,201 | -0.1 | -627 |
| | | 130 | Filled prescriptions | 0.1 | 564 | 0.0 | 69 |
| | | 30 | Lost productivity (income loss) | . | . | 0.1 | 2,589 |
| Young adults and adults (25-64 years) | Partners | 169 | Primary sector | 0.1 | 379 | 0.0 | 63 |
| | | 169 | Hospital care | 0.4 | 2,158 | -0.0 | -70 |
| | | 169 | Filled prescriptions | 0.1 | 383 | 0.0 | 137 |
| | | 141 | Lost productivity (income loss) | . | . | 1.3 | 9,252 |
| Young adults and adults (25-64 years) | Siblings | 74 | Primary sector | 0.0 | 316 | 0.0 | 23 |
| | | 74 | Hospital care | 0.1 | 1,040 | -0.0 | -355 |
| | | 74 | Filled prescriptions | 0.0 | 191 | -0.0 | -190 |
| | | 66 | Lost productivity (income loss) | . | . | -0.5 | -7,935 |
| Older people (65+ years) | Children | 118 | Primary sector | 0.0 | 269 | 0.0 | 9 |
| | | 118 | Hospital care | 0.1 | 1,214 | -0.0 | -131 |
| | | 118 | Filled prescriptions | 0.0 | 183 | -0.0 | -47 |
| | | 110 | Lost productivity (income loss) | . | . | 1.4 | 12,618 |
| Older people (65+ years) | Partners | 112 | Primary sector | 0.1 | 548 | 0.0 | 2 |
| | | 112 | Hospital care | 0.3 | 2,947 | -0.0 | -164 |
| | | 112 | Filled prescriptions | 0.1 | 536 | -0.0 | -83 |
| | | 6 | Lost productivity (income loss) | . | . | -0.1 | -16,535 |
| Older people (65+ years) | Siblings | . | Primary sector | . | . | . | . |
| | | . | Hospital care | . | . | . | . |
| | | . | Filled prescriptions | . | . | . | . |
| | | . | Lost productivity (income loss) | . | . | . | . |

### Supplementary Table 60
### Cerebral palsy - Prevalent cohort 2021
### Identification of populations of relatives

| *Agegroup of index patients in matched population* | | *Closest relative type identified* | | *Available control with same type of closest relative* |
|---|---|---|---|---|
| Children and young people (0-24 years) | n = 4,702 | Fathers | n = 4,061 (86%) | 96% |
| | | Mothers | n = 4,249 (90%) | 96% |
| | | No available closest relative | n = 194 (4%) | n/a |
| Young adults and adults (25-64 years) | n = 3,707 | Children | n = 153 (4%) | 93% |
| | | Parents | n = 1,638 (44%) | 92% |
| | | Partners | n = 674 (18%) | 95% |
| | | Siblings | n = 1,116 (30%) | 39% |
| | | Pooled | n = 3,581 (97%) | 76% |
| | | No available closest relative | n = 126 (3%) | n/a |
| Older people (65+ years) | n = 718 | Children | n = 186 (26%) | 91% |
| | | Partners | n = 212 (30%) | 96% |
| | | Siblings | n = 64 (9%) | 67% |
| | | Pooled | n = 462 (64%) | 90% |
| | | No available closest relative | n = 256 (36%) | n/a |

**Supplementary Table 61**
**Cerebral palsy - Prevalent cohort 2021**
**Patient characteristics - cases**

| *Age strata of index patients* | *Characteristic* | |
|---|---|---|
| Children and young people (0-24 years) | Number of persons, n | 4,333 (100.0) |
| | Male, n (%) | 2,543 (58.7) |
| | Age, median (Q1;Q3) | 15.4 (10.1;20.3) |
| | CCI, n (%) | |
| | 0 | 2,835 (65.4) |
| | 1-2 | 1,224 (28.2) |
| | +3 | 274 (6.3) |
| | Education level, n (%) | |
| | No/Unknown education | 1,763 (40.7) |
| | Primary | 2,287 (52.8) |
| | Secondary | 262 (6.0) |
| | Tertiary | 21 (0.5) |
| Young adults and adults (25-64 years) | Number of persons, n | 2,741 (100.0) |
| | Male, n (%) | 1,458 (53.2) |
| | Age, median (Q1;Q3) | 36.4 (29.5;51.0) |
| | CCI, n (%) | |
| | 0 | 1,839 (67.1) |
| | 1-2 | 694 (25.3) |
| | +3 | 208 (7.6) |
| | Education level, n (%) | |
| | No/Unknown education | 136 (5.0) |
| | Primary | 1,709 (62.3) |
| | Secondary | 521 (19.0) |
| | Tertiary | 375 (13.7) |
| Older people (65+ years) | Number of persons, n | 417 (100.0) |
| | Male, n (%) | 208 (49.9) |
| | Age, median (Q1;Q3) | 72.6 (67.5;78.2) |
| | CCI, n (%) | |
| | 0 | 127 (30.5) |
| | 1-2 | 165 (39.6) |
| | +3 | 125 (30.0) |
| | Education level, n (%) | |
| | No/Unknown education | 26 (6.2) |
| | Primary | 156 (37.4) |
| | Secondary | 174 (41.7) |
| | Tertiary | 61 (14.6) |

**Supplementary Table 62**
**Cerebral palsy - Prevalent cohort 2021**
**Patient characteristics - relatives - pooled**

| *Age strata of index patients* | *Closest relative type* | *Characteristic of family relatives by agestrata of index patients* | *Closest family relatives of brain disorder patients* | *Comparison relative* |
|---|---|---|---|---|
| Children and young people (0-24 years) | Fathers | Number of persons, n | 3,900 (100.0) | 3,900 (100.0) |
| | | Male, n (%) | 3,900 (100.0) | 3,900 (100.0) |
| | | Age, median (Q1;Q3) | 48.1 (42.1;53.7) | 48.3 (42.2;53.3) |
| | | CCI, n (%) | | |
| | | 0 | 3,433 (88.0) | 3,471 (89.0) |
| | | 1-2 | 407 (10.4) | 375 (9.6) |
| | | +3 | 60 (1.5) | 54 (1.4) |
| | | Education level, n (%) | | |
| | | No/Unknown education | 76 (1.9) | 76 (1.9) |
| | | Primary | 876 (22.5) | 876 (22.5) |
| | | Secondary | 1,919 (49.2) | 1,919 (49.2) |
| | | Tertiary | 1,029 (26.4) | 1,029 (26.4) |
| | | Relation to patient, n (%) | | |
| | | Father | 3,900 (100.0) | 3,900 (100.0) |
| Children and young people (0-24 years) | Mothers | Number of persons, n | 4,089 (100.0) | 4,089 (100.0) |
| | | Male, n (%) | 0 (0.0) | 0 (0.0) |
| | | Age, median (Q1;Q3) | 46.0 (40.2;51.3) | 46.1 (40.2;51.0) |
| | | CCI, n (%) | | |
| | | 0 | 3,478 (85.1) | 3,612 (88.3) |
| | | 1-2 | 542 (13.3) | 443 (10.8) |
| | | +3 | 69 (1.7) | 34 (0.8) |
| | | Education level, n (%) | | |
| | | No/Unknown education | 72 (1.8) | 72 (1.8) |
| | | Primary | 848 (20.7) | 848 (20.7) |
| | | Secondary | 1,655 (40.5) | 1,655 (40.5) |
| | | Tertiary | 1,514 (37.0) | 1,514 (37.0) |
| | | Relation to patient, n (%) | | |
| | | Mother | 4,089 (100.0) | 4,089 (100.0) |
| Young adults and adults (25-64 years) | Pooled | Number of persons, n | 2,741 (100.0) | 2,741 (100.0) |
| | | Male, n (%) | 1,096 (40.0) | 1,096 (40.0) |
| | | Age, median (Q1;Q3) | 57.1 (47.4;64.6) | 57.2 (47.3;64.1) |
| | | CCI, n (%) | | |
| | | 0 | 2,109 (76.9) | 2,161 (78.8) |
| | | 1-2 | 511 (18.6) | 485 (17.7) |
| | | +3 | 121 (4.4) | 95 (3.5) |
| | | Education level, n (%) | | |
| | | No/Unknown education | 72 (2.6) | 72 (2.6) |
| | | Primary | 707 (25.8) | 707 (25.8) |
| | | Secondary | 1,295 (47.2) | 1,295 (47.2) |
| | | Tertiary | 667 (24.3) | 667 (24.3) |
| | | Relation to patient, n (%) | | |
| | | Mother | 1,159 (42.3) | 1,159 (42.3) |
| | | Father | 355 (13.0) | 355 (13.0) |
| | | Partner | 646 (23.6) | 646 (23.6) |
| | | Child | 143 (5.2) | 143 (5.2) |

#### Supplementary Table 62
#### Cerebral palsy - Prevalent cohort 2021
#### Patient characteristics - relatives - pooled

| *Age strata of index patients* | *Closest relative type* | *Characteristic of family relatives by agestrata of index patients* | *Closest family relatives of brain disorder patients* | *Comparison relative* |
|---|---|---|---|---|
| | | Sibling | 438 (16.0) | 438 (16.0) |
| Older people (65+ years) | Pooled | Number of persons, n | 417 (100.0) | 417 (100.0) |
| | | Male, n (%) | 196 (47.0) | 196 (47.0) |
| | | Age, median (Q1;Q3) | 62.6 (52.7;71.0) | 63.2 (52.6;71.2) |
| | | CCI, n (%) | | |
| | | 0 | 307 (73.6) | 325 (77.9) |
| | | 1-2 | 79 (18.9) | 80 (19.2) |
| | | +3 | 31 (7.4) | 12 (2.9) |
| | | Education level, n (%) | | |
| | | No/Unknown education | 14 (3.4) | 14 (3.4) |
| | | Primary | 89 (21.3) | 89 (21.3) |
| | | Secondary | 202 (48.4) | 202 (48.4) |
| | | Tertiary | 112 (26.9) | 112 (26.9) |
| | | Relation to patient, n (%) | | |
| | | Partner | 204 (48.9) | 204 (48.9) |
| | | Child | 170 (40.8) | 170 (40.8) |
| | | Sibling | 43 (10.3) | 43 (10.3) |

### Supplementary Table 63
### Cerebral palsy - Prevalent cohort 2021
### Patient characteristics - relatives - not pooled

| *Age strata of index patients* | *Closest relative type* | *Characteristic of family relatives by agestrata of index patients* | *Closest family relatives of brain disorder patients* | *Comparison relative* |
|---|---|---|---|---|
| Children and young people (0-24 years) | Fathers | Number of persons, n | 3,900 (100.0) | 3,900 (100.0) |
| | | Male, n (%) | 3,900 (100.0) | 3,900 (100.0) |
| | | Age, median (Q1;Q3) | 48.1 (42.1;53.7) | 48.3 (42.2;53.3) |
| | | CCI, n (%) | | |
| | | 0 | 3,433 (88.0) | 3,471 (89.0) |
| | | 1-2 | 407 (10.4) | 375 (9.6) |
| | | +3 | 60 (1.5) | 54 (1.4) |
| | | Education level, n (%) | | |
| | | No/Unknown education | 76 (1.9) | 76 (1.9) |
| | | Primary | 876 (22.5) | 876 (22.5) |
| | | Secondary | 1,919 (49.2) | 1,919 (49.2) |
| | | Tertiary | 1,029 (26.4) | 1,029 (26.4) |
| | | Relation to patient, n (%) | | |
| | | Father | 3,900 (100.0) | 3,900 (100.0) |
| Children and young people (0-24 years) | Mothers | Number of persons, n | 4,089 (100.0) | 4,089 (100.0) |
| | | Male, n (%) | 0 (0.0) | 0 (0.0) |
| | | Age, median (Q1;Q3) | 46.0 (40.2;51.3) | 46.1 (40.2;51.0) |
| | | CCI, n (%) | | |
| | | 0 | 3,478 (85.1) | 3,612 (88.3) |
| | | 1-2 | 542 (13.3) | 443 (10.8) |
| | | +3 | 69 (1.7) | 34 (0.8) |
| | | Education level, n (%) | | |
| | | No/Unknown education | 72 (1.8) | 72 (1.8) |
| | | Primary | 848 (20.7) | 848 (20.7) |
| | | Secondary | 1,655 (40.5) | 1,655 (40.5) |
| | | Tertiary | 1,514 (37.0) | 1,514 (37.0) |
| | | Relation to patient, n (%) | | |
| | | Mother | 4,089 (100.0) | 4,089 (100.0) |
| Young adults and adults (25-64 years) | Children | Number of persons, n | 143 (100.0) | 143 (100.0) |
| | | Male, n (%) | 77 (53.8) | 77 (53.8) |
| | | Age, median (Q1;Q3) | 22.8 (13.1;32.0) | 22.7 (12.3;30.8) |
| | | CCI, n (%) | | |
| | | 0 | 133 (93.0) | 129 (90.2) |
| | | 1-2 | 10 (7.0) | 14 (9.8) |
| | | +3 | 0 (0.0) | 0 (0.0) |
| | | Education level, n (%) | | |
| | | No/Unknown education | 37 (25.9) | 37 (25.9) |
| | | Primary | 50 (35.0) | 50 (35.0) |
| | | Secondary | 36 (25.2) | 36 (25.2) |
| | | Tertiary | 20 (14.0) | 20 (14.0) |
| | | Relation to patient, n (%) | | |
| | | Child | 143 (100.0) | 143 (100.0) |
| Young adults and adults (25-64 years) | Parents | Number of persons, n | 1,514 (100.0) | 1,514 (100.0) |
| | | Male, n (%) | 355 (23.4) | 355 (23.4) |
| | | Age, median (Q1;Q3) | 61.9 (56.7;69.5) | 61.7 (56.7;69.4) |

### Supplementary Table 63
### Cerebral palsy - Prevalent cohort 2021
### Patient characteristics - relatives - not pooled

| *Age strata of index patients* | *Closest relative type* | *Characteristic of family relatives by agestrata of index patients* | *Closest family relatives of brain disorder patients* | *Comparison relative* |
|---|---|---|---|---|
| | | CCI, n (%) | | |
| | | 0 | 1,086 (71.7) | 1,087 (71.8) |
| | | 1-2 | 333 (22.0) | 347 (22.9) |
| | | +3 | 95 (6.3) | 80 (5.3) |
| | | Education level, n (%) | | |
| | | No/Unknown education | 24 (1.6) | 24 (1.6) |
| | | Primary | 373 (24.6) | 373 (24.6) |
| | | Secondary | 735 (48.5) | 735 (48.5) |
| | | Tertiary | 382 (25.2) | 382 (25.2) |
| | | Relation to patient, n (%) | | |
| | | Mother | 1,159 (76.6) | 1,159 (76.6) |
| | | Father | 355 (23.4) | 355 (23.4) |
| Young adults and adults (25-64 years) | Partners | Number of persons, n | 646 (100.0) | 646 (100.0) |
| | | Male, n (%) | 393 (60.8) | 393 (60.8) |
| | | Age, median (Q1;Q3) | 43.5 (32.5;55.7) | 43.9 (31.9;56.4) |
| | | CCI, n (%) | | |
| | | 0 | 532 (82.4) | 570 (88.2) |
| | | 1-2 | 99 (15.3) | 70 (10.8) |
| | | +3 | 15 (2.3) | 6 (0.9) |
| | | Education level, n (%) | | |
| | | No/Unknown education | 11 (1.7) | 11 (1.7) |
| | | Primary | 179 (27.7) | 179 (27.7) |
| | | Secondary | 283 (43.8) | 283 (43.8) |
| | | Tertiary | 173 (26.8) | 173 (26.8) |
| | | Relation to patient, n (%) | | |
| | | Partner | 646 (100.0) | 646 (100.0) |
| Young adults and adults (25-64 years) | Siblings | Number of persons, n | 438 (100.0) | 438 (100.0) |
| | | Male, n (%) | 271 (61.9) | 271 (61.9) |
| | | Age, median (Q1;Q3) | 52.1 (34.5;58.2) | 52.2 (34.5;59.0) |
| | | CCI, n (%) | | |
| | | 0 | 358 (81.7) | 375 (85.6) |
| | | 1-2 | 69 (15.8) | 54 (12.3) |
| | | +3 | 11 (2.5) | 9 (2.1) |
| | | Education level, n (%) | | |
| | | No/Unknown education | 0 (0.0) | 0 (0.0) |
| | | Primary | 105 (24.0) | 105 (24.0) |
| | | Secondary | 241 (55.0) | 241 (55.0) |
| | | Tertiary | 92 (21.0) | 92 (21.0) |
| | | Relation to patient, n (%) | | |
| | | Sibling | 438 (100.0) | 438 (100.0) |
| Older people (65+ years) | Children | Number of persons, n | 170 (100.0) | 170 (100.0) |
| | | Male, n (%) | 90 (52.9) | 90 (52.9) |
| | | Age, median (Q1;Q3) | 50.4 (42.7;55.7) | 49.9 (42.2;55.8) |
| | | CCI, n (%) | | |
| | | 0 | 155 (91.2) | 150 (88.2) |

**Supplementary Table 63**
**Cerebral palsy - Prevalent cohort 2021**
**Patient characteristics - relatives - not pooled**

| *Age strata of index patients* | *Closest relative type* | *Characteristic of family relatives by agestrata of index patients* | *Closest family relatives of brain disorder patients* | *Comparison relative* |
|---|---|---|---|---|
| | | 1-2 | * | * |
| | | +3 | * | * |
| | | Education level, n (%) | | |
| | | No/Unknown education | * | * |
| | | Primary | * | * |
| | | Secondary | 85 (50.0) | 85 (50.0) |
| | | Tertiary | 59 (34.7) | 59 (34.7) |
| | | Relation to patient, n (%) | | |
| | | Child | 170 (100.0) | 170 (100.0) |
| Older people (65+ years) | Partners | Number of persons, n | 204 (100.0) | 204 (100.0) |
| | | Male, n (%) | 87 (42.6) | 87 (42.6) |
| | | Age, median (Q1;Q3) | 71.0 (67.1;76.9) | 71.2 (67.2;76.9) |
| | | CCI, n (%) | | |
| | | 0 | 117 (57.4) | 140 (68.6) |
| | | 1-2 | 58 (28.4) | 54 (26.5) |
| | | +3 | 29 (14.2) | 10 (4.9) |
| | | Education level, n (%) | | |
| | | No/Unknown education | 12 (5.9) | 12 (5.9) |
| | | Primary | 59 (28.9) | 59 (28.9) |
| | | Secondary | 89 (43.6) | 89 (43.6) |
| | | Tertiary | 44 (21.6) | 44 (21.6) |
| | | Relation to patient, n (%) | | |
| | | Partner | 204 (100.0) | 204 (100.0) |
| Older people (65+ years) | Siblings | Number of persons, n | 43 (100.0) | 43 (100.0) |
| | | Male, n (%) | 19 (44.2) | 19 (44.2) |
| | | Age, median (Q1;Q3) | 62.1 (59.7;65.5) | 62.7 (60.7;64.4) |
| | | CCI, n (%) | | |
| | | 0 | 35 (81.4) | * |
| | | 1-2 | 8 (18.6) | * |
| | | +3 | 0 (0.0) | * |
| | | Education level, n (%) | | |
| | | No/Unknown education | * | * |
| | | Primary | * | * |
| | | Secondary | * | * |
| | | Tertiary | * | * |
| | | Relation to patient, n (%) | | |
| | | Sibling | 43 (100.0) | 43 (100.0) |

#### Supplementary Table 64
#### Cerebral palsy - Prevalent cohort 2021
#### Cost analysis - pooled

| | | | | Actual costs | | Attributable costs | |
|---|---|---|---|---|---|---|---|
| *Age strata of index patients* | *Closest relative type* | *Number of index patients included in cost component calculations* | *Cost component* | *Total (million EUR)* | *Per person (EUR)* | *Total (million EUR)* | *Per person (EUR)* |
| Children and young people (0-24 years) | Fathers | 3,900 | Primary sector | 0.9 | 218 | 0.0 | 2 |
| | | 3,900 | Hospital care | 6.0 | 1,539 | 0.8 | 202 |
| | | 3,900 | Filled prescriptions | 0.7 | 183 | 0.0 | 4 |
| | | 3,819 | Lost productivity (income loss) | . | . | 17.8 | 4,669 |
| Children and young people (0-24 years) | Mothers | 4,089 | Primary sector | 1.5 | 361 | 0.1 | 31 |
| | | 4,089 | Hospital care | 8.9 | 2,179 | 2.7 | 670 |
| | | 4,089 | Filled prescriptions | 0.9 | 224 | 0.2 | 48 |
| | | 4,060 | Lost productivity (income loss) | . | . | 23.7 | 5,847 |
| Young adults and adults (25-64 years) | Pooled | 2,741 | Primary sector | 1.0 | 373 | 0.0 | 10 |
| | | 2,741 | Hospital care | 7.2 | 2,642 | 0.6 | 202 |
| | | 2,741 | Filled prescriptions | 1.0 | 365 | 0.1 | 53 |
| | | 1,850 | Lost productivity (income loss) | . | . | 6.1 | 3,298 |
| Older people (65+ years) | Pooled | 417 | Primary sector | 0.2 | 411 | 0.0 | 4 |
| | | 417 | Hospital care | 1.4 | 3,393 | 0.5 | 1,171 |
| | | 417 | Filled prescriptions | 0.2 | 400 | 0.0 | 75 |
| | | 206 | Lost productivity (income loss) | . | . | 2.6 | 12,414 |

#### Supplementary Table 65
#### Cerebral palsy - Prevalent cohort 2021
#### Cost analysis - not pooled

| Age strata of index patients | Closest relative type | Number of index patients included in cost component calculations | Cost component | Actual costs: Total (million EUR) | Actual costs: Per person (EUR) | Attributable costs: Total (million EUR) | Attributable costs: Per person (EUR) |
|---|---|---|---|---|---|---|---|
| Children and young people (0-24 years) | Fathers | 3,900 | Primary sector | 0.9 | 218 | 0.0 | 2 |
| | | 3,900 | Hospital care | 6.0 | 1,539 | 0.8 | 202 |
| | | 3,900 | Filled prescriptions | 0.7 | 183 | 0.0 | 4 |
| | | 3,819 | Lost productivity (income loss) | . | . | 17.8 | 4,669 |
| Children and young people (0-24 years) | Mothers | 4,089 | Primary sector | 1.5 | 361 | 0.1 | 31 |
| | | 4,089 | Hospital care | 8.9 | 2,179 | 2.7 | 670 |
| | | 4,089 | Filled prescriptions | 0.9 | 224 | 0.2 | 48 |
| | | 4,060 | Lost productivity (income loss) | . | . | 23.7 | 5,847 |
| Young adults and adults (25-64 years) | Children | 143 | Primary sector | 0.0 | 255 | 0.0 | 29 |
| | | 143 | Hospital care | 0.3 | 2,125 | 0.1 | 373 |
| | | 143 | Filled prescriptions | 0.0 | 147 | 0.0 | 13 |
| | | 86 | Lost productivity (income loss) | . | . | 1.1 | 12,498 |
| Young adults and adults (25-64 years) | Parents | 1,514 | Primary sector | 0.6 | 427 | 0.0 | 8 |
| | | 1,514 | Hospital care | 5.3 | 3,555 | 0.9 | 607 |
| | | 1,514 | Filled prescriptions | 0.7 | 474 | 0.1 | 74 |
| | | 805 | Lost productivity (income loss) | . | . | -1.3 | -1,616 |
| Young adults and adults (25-64 years) | Partners | 646 | Primary sector | 0.2 | 306 | -0.0 | -0 |
| | | 646 | Hospital care | 0.9 | 1,444 | -0.2 | -235 |
| | | 646 | Filled prescriptions | 0.2 | 242 | 0.0 | 47 |
| | | 581 | Lost productivity (income loss) | . | . | 5.9 | 10,127 |
| Young adults and adults (25-64 years) | Siblings | 438 | Primary sector | 0.1 | 323 | 0.0 | 27 |
| | | 438 | Hospital care | 0.6 | 1,441 | -0.3 | -599 |
| | | 438 | Filled prescriptions | 0.1 | 244 | 0.0 | 6 |
| | | 378 | Lost productivity (income loss) | . | . | 0.4 | 1,175 |
| Older people (65+ years) | Children | 170 | Primary sector | 0.0 | 284 | -0.0 | -18 |
| | | 170 | Hospital care | 0.4 | 2,217 | 0.1 | 729 |
| | | 170 | Filled prescriptions | 0.0 | 181 | 0.0 | 2 |
| | | 163 | Lost productivity (income loss) | . | . | 2.6 | 16,124 |
| Older people (65+ years) | Partners | 204 | Primary sector | 0.1 | 521 | 0.0 | 27 |
| | | 204 | Hospital care | 1.0 | 4,919 | 0.4 | 2,190 |
| | | 204 | Filled prescriptions | 0.1 | 574 | 0.0 | 103 |
| | | 21 | Lost productivity (income loss) | . | . | 0.4 | 18,074 |
| Older people (65+ years) | Siblings | 43 | Primary sector | 0.0 | 402 | -0.0 | -20 |
| | | 43 | Hospital care | 0.0 | 931 | -0.1 | -1,818 |
| | | 43 | Filled prescriptions | 0.0 | 452 | 0.0 | 232 |
| | | 22 | Lost productivity (income loss) | . | . | -0.5 | -20,478 |

**Supplementary Table 66**
**Dementia - Incident cohort 2016-2021**
**Identification of populations of relatives**

| ***Agegroup of index patients in matched population*** | | ***Closest relative type identified*** | | ***Available control with same type of closest relative*** |
|---|---|---|---|---|
| Children and young people (0-24 years) | n = 33 | Fathers | n/a | n/a |
| | | Mothers | n/a | n/a |
| | | No available closest relative | n/a | n/a |
| Young adults and adults (25-64 years) | n = 3,055 | Children | n = 561 (18%) | 88% |
| | | Parents | n = 279 (9%) | 82% |
| | | Partners | n = 1,498 (49%) | 96% |
| | | Siblings | n = 543 (18%) | 61% |
| | | Pooled | n = 2,881 (94%) | 86% |
| | | No available closest relative | n = 174 (6%) | n/a |
| Older people (65+ years) | n = 49,243 | Children | n = 22,349 (45%) | 96% |
| | | Partners | n = 20,536 (42%) | 95% |
| | | Siblings | n = 391 (1%) | 26% |
| | | Pooled | n = 43,276 (88%) | 95% |
| | | No available closest relative | n = 5,967 (12%) | n/a |

**Supplementary Table 67**
**Dementia - Incident cohort 2016-2021**
**Patient characteristics - cases**

| ***Age strata of index patients*** | ***Characteristic*** | |
|---|---|---|
| Children and young people (0-24 years) | Number of persons, n | 30 (100.0) |
| | Male, n (%) | 10 (33.3) |
| | Age, median (Q1;Q3) | 18.2 (7.2;21.8) |
| | CCI, n (%) | |
| | 0 | * |
| | 1-2 | * |
| | +3 | * |
| | Education level, n (%) | |
| | No/Unknown education | 14 (46.7) |
| | Primary | 11 (36.7) |
| | Secondary | 5 (16.7) |
| | Tertiary | 0 (0.0) |
| Young adults and adults (25-64 years) | Number of persons, n | 2,499 (100.0) |
| | Male, n (%) | 1,430 (57.2) |
| | Age, median (Q1;Q3) | 59.8 (55.3;62.7) |
| | CCI, n (%) | |
| | 0 | 1,590 (63.6) |
| | 1-2 | 698 (27.9) |
| | +3 | 211 (8.4) |
| | Education level, n (%) | |
| | No/Unknown education | 150 (6.0) |
| | Primary | 745 (29.8) |
| | Secondary | 1,142 (45.7) |
| | Tertiary | 462 (18.5) |
| Older people (65+ years) | Number of persons, n | 41,243 (100.0) |
| | Male, n (%) | 18,268 (44.3) |
| | Age, median (Q1;Q3) | 81.0 (75.8;85.7) |
| | CCI, n (%) | |
| | 0 | 19,248 (46.7) |
| | 1-2 | 15,627 (37.9) |
| | +3 | 6,368 (15.4) |
| | Education level, n (%) | |
| | No/Unknown education | 2,459 (6.0) |
| | Primary | 15,906 (38.6) |
| | Secondary | 16,938 (41.1) |
| | Tertiary | 5,940 (14.4) |

**Supplementary Table 68**
**Dementia - Incident cohort 2016-2021**
**Patient characteristics - relatives - pooled**

| *Age strata of index patients* | *Closest relative type* | *Characteristic of family relatives by agestrata of index patients* | *Closest family relatives of brain disorder patients* | *Comparison relative* |
|---|---|---|---|---|
| Children and young people (0-24 years) | Fathers | Number of persons, n | 26 (100.0) | 26 (100.0) |
| | | Male, n (%) | 26 (100.0) | 26 (100.0) |
| | | Age, median (Q1;Q3) | 47.2 (39.2;54.2) | 45.8 (38.4;55.0) |
| | | CCI, n (%) | | |
| | | 0 | 21 (80.8) | * |
| | | 1-2 | 5 (19.2) | * |
| | | +3 | 0 (0.0) | 0 (0.0) |
| | | Education level, n (%) | | |
| | | No/Unknown education | 0 (0.0) | 0 (0.0) |
| | | Primary | 9 (34.6) | 9 (34.6) |
| | | Secondary | 10 (38.5) | 10 (38.5) |
| | | Tertiary | 7 (26.9) | 7 (26.9) |
| | | Relation to patient, n (%) | | |
| | | Father | 26 (100.0) | 26 (100.0) |
| Children and young people (0-24 years) | Mothers | Number of persons, n | 27 (100.0) | 27 (100.0) |
| | | Male, n (%) | 0 (0.0) | 0 (0.0) |
| | | Age, median (Q1;Q3) | 46.1 (36.2;51.6) | 47.2 (35.0;50.5) |
| | | CCI, n (%) | | |
| | | 0 | 22 (81.5) | * |
| | | 1-2 | 5 (18.5) | * |
| | | +3 | 0 (0.0) | * |
| | | Education level, n (%) | | |
| | | No/Unknown education | 0 (0.0) | 0 (0.0) |
| | | Primary | 6 (22.2) | 6 (22.2) |
| | | Secondary | 7 (25.9) | 7 (25.9) |
| | | Tertiary | 14 (51.9) | 14 (51.9) |
| | | Relation to patient, n (%) | | |
| | | Mother | 27 (100.0) | 27 (100.0) |
| Young adults and adults (25-64 years) | Pooled | Number of persons, n | 2,499 (100.0) | 2,499 (100.0) |
| | | Male, n (%) | 1,122 (44.9) | 1,122 (44.9) |
| | | Age, median (Q1;Q3) | 58.0 (47.8;63.4) | 57.7 (48.5;63.0) |
| | | CCI, n (%) | | |
| | | 0 | 1,952 (78.1) | 2,000 (80.0) |
| | | 1-2 | 445 (17.8) | 394 (15.8) |
| | | +3 | 102 (4.1) | 105 (4.2) |
| | | Education level, n (%) | | |
| | | No/Unknown education | 58 (2.3) | 58 (2.3) |
| | | Primary | 724 (29.0) | 724 (29.0) |
| | | Secondary | 1,159 (46.4) | 1,159 (46.4) |
| | | Tertiary | 558 (22.3) | 558 (22.3) |
| | | Relation to patient, n (%) | | |
| | | Mother | 185 (7.4) | 185 (7.4) |
| | | Father | 45 (1.8) | 45 (1.8) |
| | | Partner | 1,440 (57.6) | 1,440 (57.6) |
| | | Child | 494 (19.8) | 494 (19.8) |

**Supplementary Table 68**
**Dementia - Incident cohort 2016-2021**
**Patient characteristics - relatives - pooled**

| *Age strata of index patients* | *Closest relative type* | *Characteristic of family relatives by agestrata of index patients* | *Closest family relatives of brain disorder patients* | *Comparison relative* |
|---|---|---|---|---|
| | | Sibling | 335 (13.4) | 335 (13.4) |
| Older people (65+ years) | Pooled | Number of persons, n | 41,243 (100.0) | 41,243 (100.0) |
| | | Male, n (%) | 18,698 (45.3) | 18,698 (45.3) |
| | | Age, median (Q1;Q3) | 64.3 (56.2;76.4) | 64.1 (56.2;76.5) |
| | | CCI, n (%) | | |
| | | 0 | 29,137 (70.6) | 29,440 (71.4) |
| | | 1-2 | 9,308 (22.6) | 9,206 (22.3) |
| | | +3 | 2,798 (6.8) | 2,597 (6.3) |
| | | Education level, n (%) | | |
| | | No/Unknown education | 1,216 (2.9) | 1,216 (2.9) |
| | | Primary | 9,441 (22.9) | 9,441 (22.9) |
| | | Secondary | 20,495 (49.7) | 20,495 (49.7) |
| | | Tertiary | 10,091 (24.5) | 10,091 (24.5) |
| | | Relation to patient, n (%) | | |
| | | Partner | 19,660 (47.7) | 19,660 (47.7) |
| | | Child | 21,481 (52.1) | 21,481 (52.1) |
| | | Sibling | 102 (0.2) | 102 (0.2) |

**Supplementary Table 69**
**Dementia - Incident cohort 2016-2021**
**Patient characteristics - relatives - not pooled**

| *Age strata of index patients* | *Closest relative type* | *Characteristic of family relatives by agestrata of index patients* | *Closest family relatives of brain disorder patients* | *Comparison relative* |
|---|---|---|---|---|
| Children and young people (0-24 years) | Fathers | Number of persons, n | 26 (100.0) | 26 (100.0) |
| | | Male, n (%) | 26 (100.0) | 26 (100.0) |
| | | Age, median (Q1;Q3) | 47.2 (39.2;54.2) | 45.8 (38.4;55.0) |
| | | CCI, n (%) | | |
| | | 0 | 21 (80.8) | * |
| | | 1-2 | 5 (19.2) | * |
| | | +3 | 0 (0.0) | 0 (0.0) |
| | | Education level, n (%) | | |
| | | No/Unknown education | 0 (0.0) | 0 (0.0) |
| | | Primary | 9 (34.6) | 9 (34.6) |
| | | Secondary | 10 (38.5) | 10 (38.5) |
| | | Tertiary | 7 (26.9) | 7 (26.9) |
| | | Relation to patient, n (%) | | |
| | | Father | 26 (100.0) | 26 (100.0) |
| Children and young people (0-24 years) | Mothers | Number of persons, n | 27 (100.0) | 27 (100.0) |
| | | Male, n (%) | 0 (0.0) | 0 (0.0) |
| | | Age, median (Q1;Q3) | 46.1 (36.2;51.6) | 47.2 (35.0;50.5) |
| | | CCI, n (%) | | |
| | | 0 | 22 (81.5) | * |
| | | 1-2 | 5 (18.5) | * |
| | | +3 | 0 (0.0) | * |
| | | Education level, n (%) | | |
| | | No/Unknown education | 0 (0.0) | 0 (0.0) |
| | | Primary | 6 (22.2) | 6 (22.2) |
| | | Secondary | 7 (25.9) | 7 (25.9) |
| | | Tertiary | 14 (51.9) | 14 (51.9) |
| | | Relation to patient, n (%) | | |
| | | Mother | 27 (100.0) | 27 (100.0) |
| Young adults and adults (25-64 years) | Children | Number of persons, n | 494 (100.0) | 494 (100.0) |
| | | Male, n (%) | 260 (52.6) | 260 (52.6) |
| | | Age, median (Q1;Q3) | 31.6 (25.3;37.5) | 31.0 (25.1;37.1) |
| | | CCI, n (%) | | |
| | | 0 | 461 (93.3) | 458 (92.7) |
| | | 1-2 | * | * |
| | | +3 | * | * |
| | | Education level, n (%) | | |
| | | No/Unknown education | 9 (1.8) | 9 (1.8) |
| | | Primary | 194 (39.3) | 194 (39.3) |
| | | Secondary | 172 (34.8) | 172 (34.8) |
| | | Tertiary | 119 (24.1) | 119 (24.1) |
| | | Relation to patient, n (%) | | |
| | | Child | 494 (100.0) | 494 (100.0) |
| Young adults and adults (25-64 years) | Parents | Number of persons, n | 230 (100.0) | 230 (100.0) |
| | | Male, n (%) | 45 (19.6) | 45 (19.6) |
| | | Age, median (Q1;Q3) | 76.7 (70.5;81.6) | 76.9 (71.7;82.1) |

#### Supplementary Table 69
#### Dementia - Incident cohort 2016-2021
#### Patient characteristics - relatives - not pooled

| *Age strata of index patients* | *Closest relative type* | *Characteristic of family relatives by agestrata of index patients* | *Closest family relatives of brain disorder patients* | *Comparison relative* |
|---|---|---|---|---|
| | | CCI, n (%) | | |
| | | 0 | 139 (60.4) | 140 (60.9) |
| | | 1-2 | 64 (27.8) | 58 (25.2) |
| | | +3 | 27 (11.7) | 32 (13.9) |
| | | Education level, n (%) | | |
| | | No/Unknown education | * | * |
| | | Primary | 113 (49.1) | 113 (49.1) |
| | | Secondary | 88 (38.3) | 88 (38.3) |
| | | Tertiary | * | * |
| | | Relation to patient, n (%) | | |
| | | Mother | 185 (80.4) | 185 (80.4) |
| | | Father | 45 (19.6) | 45 (19.6) |
| Young adults and adults (25-64 years) | Partners | Number of persons, n | 1,440 (100.0) | 1,440 (100.0) |
| | | Male, n (%) | 617 (42.8) | 617 (42.8) |
| | | Age, median (Q1;Q3) | 59.7 (54.9;63.3) | 59.3 (55.0;63.1) |
| | | CCI, n (%) | | |
| | | 0 | 1,097 (76.2) | 1,158 (80.4) |
| | | 1-2 | 282 (19.6) | 229 (15.9) |
| | | +3 | 61 (4.2) | 53 (3.7) |
| | | Education level, n (%) | | |
| | | No/Unknown education | 45 (3.1) | 45 (3.1) |
| | | Primary | 340 (23.6) | 340 (23.6) |
| | | Secondary | 699 (48.5) | 699 (48.5) |
| | | Tertiary | 356 (24.7) | 356 (24.7) |
| | | Relation to patient, n (%) | | |
| | | Partner | 1,440 (100.0) | 1,440 (100.0) |
| Young adults and adults (25-64 years) | Siblings | Number of persons, n | 335 (100.0) | 335 (100.0) |
| | | Male, n (%) | 200 (59.7) | 200 (59.7) |
| | | Age, median (Q1;Q3) | 58.2 (54.4;61.6) | 57.9 (54.0;61.6) |
| | | CCI, n (%) | | |
| | | 0 | 255 (76.1) | 244 (72.8) |
| | | 1-2 | 70 (20.9) | 74 (22.1) |
| | | +3 | 10 (3.0) | 17 (5.1) |
| | | Education level, n (%) | | |
| | | No/Unknown education | 0 (0.0) | 0 (0.0) |
| | | Primary | 77 (23.0) | 77 (23.0) |
| | | Secondary | 200 (59.7) | 200 (59.7) |
| | | Tertiary | 58 (17.3) | 58 (17.3) |
| | | Relation to patient, n (%) | | |
| | | Sibling | 335 (100.0) | 335 (100.0) |
| Older people (65+ years) | Children | Number of persons, n | 21,481 (100.0) | 21,481 (100.0) |
| | | Male, n (%) | 11,077 (51.6) | 11,077 (51.6) |
| | | Age, median (Q1;Q3) | 56.6 (51.7;60.8) | 56.5 (51.8;60.4) |
| | | CCI, n (%) | | |
| | | 0 | 17,691 (82.4) | 17,875 (83.2) |

### Supplementary Table 69
### Dementia - Incident cohort 2016-2021
### Patient characteristics - relatives - not pooled

| *Age strata of index patients* | *Closest relative type* | *Characteristic of family relatives by agestrata of index patients* | *Closest family relatives of brain disorder patients* | *Comparison relative* |
|---|---|---|---|---|
| | | 1-2 | 3,229 (15.0) | 3,099 (14.4) |
| | | +3 | 561 (2.6) | 507 (2.4) |
| | | Education level, n (%) | | |
| | | No/Unknown education | 167 (0.8) | 167 (0.8) |
| | | Primary | 3,275 (15.2) | 3,275 (15.2) |
| | | Secondary | 11,415 (53.1) | 11,415 (53.1) |
| | | Tertiary | 6,624 (30.8) | 6,624 (30.8) |
| | | Relation to patient, n (%) | | |
| | | Child | 21,481 (100.0) | 21,481 (100.0) |
| Older people (65+ years) | Partners | Number of persons, n | 19,660 (100.0) | 19,660 (100.0) |
| | | Male, n (%) | 7,563 (38.5) | 7,563 (38.5) |
| | | Age, median (Q1;Q3) | 76.9 (72.1;81.7) | 76.8 (72.3;81.6) |
| | | CCI, n (%) | | |
| | | 0 | 11,370 (57.8) | 11,486 (58.4) |
| | | 1-2 | 6,055 (30.8) | 6,093 (31.0) |
| | | +3 | 2,235 (11.4) | 2,081 (10.6) |
| | | Education level, n (%) | | |
| | | No/Unknown education | 1,049 (5.3) | 1,049 (5.3) |
| | | Primary | 6,153 (31.3) | 6,153 (31.3) |
| | | Secondary | 9,013 (45.8) | 9,013 (45.8) |
| | | Tertiary | 3,445 (17.5) | 3,445 (17.5) |
| | | Relation to patient, n (%) | | |
| | | Partner | 19,660 (100.0) | 19,660 (100.0) |
| Older people (65+ years) | Siblings | Number of persons, n | 102 (100.0) | 102 (100.0) |
| | | Male, n (%) | 58 (56.9) | 58 (56.9) |
| | | Age, median (Q1;Q3) | 62.4 (59.0;65.7) | 62.6 (59.1;65.6) |
| | | CCI, n (%) | | |
| | | 0 | 76 (74.5) | 79 (77.5) |
| | | 1-2 | * | 14 (13.7) |
| | | +3 | * | 9 (8.8) |
| | | Education level, n (%) | | |
| | | No/Unknown education | 0 (0.0) | 0 (0.0) |
| | | Primary | 13 (12.7) | 13 (12.7) |
| | | Secondary | 67 (65.7) | 67 (65.7) |
| | | Tertiary | 22 (21.6) | 22 (21.6) |
| | | Relation to patient, n (%) | | |
| | | Sibling | 102 (100.0) | 102 (100.0) |

### Supplementary Table 70
### Dementia - Incident cohort 2016-2021
### Cost analysis - pooled

| Age strata of index patients | Closest relative type | Number of index patients included in cost component calculations | Cost component | Actual costs Total (million EUR) | Actual costs Per person (EUR) | Attributable costs Total (million EUR) | Attributable costs Per person (EUR) |
|---|---|---|---|---|---|---|---|
| Children and young people (0-24 years) | Fathers | 26 | Primary sector | 0.0 | 232 | 0.0 | 24 |
| | | 26 | Hospital care | 0.1 | 2,043 | 0.0 | 1,287 |
| | | 26 | Filled prescriptions | 0.0 | 128 | -0.0 | -16 |
| | | 24 | Lost productivity (income loss) | . | . | 0.0 | 1,046 |
| Children and young people (0-24 years) | Mothers | 27 | Primary sector | 0.0 | 393 | -0.0 | -24 |
| | | 27 | Hospital care | 0.1 | 2,062 | -0.1 | -3,439 |
| | | 27 | Filled prescriptions | 0.0 | 185 | -0.0 | -59 |
| | | 25 | Lost productivity (income loss) | . | . | 0.6 | 22,039 |
| Young adults and adults (25-64 years) | Pooled | 2,499 | Primary sector | 0.9 | 373 | 0.1 | 30 |
| | | 2,499 | Hospital care | 6.7 | 2,711 | 0.3 | 119 |
| | | 2,499 | Filled prescriptions | 0.8 | 313 | 0.1 | 23 |
| | | 1,738 | Lost productivity (income loss) | . | . | 11.2 | 6,458 |
| Older people (65+ years) | Pooled | 41,243 | Primary sector | 17.8 | 435 | 0.6 | 16 |
| | | 41,243 | Hospital care | 133.9 | 3,281 | 8.7 | 214 |
| | | 41,243 | Filled prescriptions | 17.2 | 421 | 0.9 | 22 |
| | | 18,587 | Lost productivity (income loss) | . | . | 52.0 | 2,798 |

#### Supplementary Table 71
#### Dementia - Incident cohort 2016-2021
#### Cost analysis - not pooled

| | | | | *Actual costs* | | *Attributable costs* | |
|---|---|---|---|---|---|---|---|
| *Age strata of index patients* | *Closest relative type* | *Number of index patients included in cost component calculations* | *Cost component* | *Total (million EUR)* | *Per person (EUR)* | *Total (million EUR)* | *Per person (EUR)* |
| Children and young people (0-24 years) | Fathers | 26 | Primary sector | 0.0 | 232 | 0.0 | 24 |
| | | 26 | Hospital care | 0.1 | 2,043 | 0.0 | 1,287 |
| | | 26 | Filled prescriptions | 0.0 | 128 | -0.0 | -16 |
| | | 24 | Lost productivity (income loss) | . | . | 0.0 | 1,046 |
| Children and young people (0-24 years) | Mothers | 27 | Primary sector | 0.0 | 393 | -0.0 | -24 |
| | | 27 | Hospital care | 0.1 | 2,062 | -0.1 | -3,439 |
| | | 27 | Filled prescriptions | 0.0 | 185 | -0.0 | -59 |
| | | 25 | Lost productivity (income loss) | . | . | 0.6 | 22,039 |
| Young adults and adults (25-64 years) | Children | 494 | Primary sector | 0.1 | 292 | 0.0 | 36 |
| | | 494 | Hospital care | 1.6 | 3,299 | 0.9 | 1,780 |
| | | 494 | Filled prescriptions | 0.1 | 118 | -0.0 | -38 |
| | | 453 | Lost productivity (income loss) | . | . | 1.4 | 3,009 |
| Young adults and adults (25-64 years) | Parents | 230 | Primary sector | 0.1 | 534 | 0.0 | 13 |
| | | 230 | Hospital care | 1.0 | 4,481 | -0.1 | -453 |
| | | 230 | Filled prescriptions | 0.1 | 672 | 0.0 | 85 |
| | | 20 | Lost productivity (income loss) | . | . | 0.1 | 5,319 |
| Young adults and adults (25-64 years) | Partners | 1,440 | Primary sector | 0.5 | 385 | 0.1 | 40 |
| | | 1,440 | Hospital care | 3.6 | 2,510 | 0.1 | 98 |
| | | 1,440 | Filled prescriptions | 0.4 | 309 | 0.0 | 25 |
| | | 1,009 | Lost productivity (income loss) | . | . | 9.1 | 9,018 |
| Young adults and adults (25-64 years) | Siblings | 335 | Primary sector | 0.1 | 335 | -0.0 | -14 |
| | | 335 | Hospital care | 0.5 | 1,524 | -0.6 | -1,861 |
| | | 335 | Filled prescriptions | 0.1 | 377 | 0.0 | 60 |
| | | 256 | Lost productivity (income loss) | . | . | 0.7 | 2,557 |
| Older people (65+ years) | Children | 21,481 | Primary sector | 7.0 | 329 | 0.4 | 18 |
| | | 21,481 | Hospital care | 46.3 | 2,162 | 4.8 | 225 |
| | | 21,481 | Filled prescriptions | 5.8 | 270 | 0.3 | 13 |
| | | 18,121 | Lost productivity (income loss) | . | . | 51.5 | 2,843 |
| Older people (65+ years) | Partners | 19,660 | Primary sector | 10.7 | 555 | 0.3 | 14 |
| | | 19,660 | Hospital care | 87.4 | 4,534 | 4.1 | 210 |
| | | 19,660 | Filled prescriptions | 11.4 | 589 | 0.6 | 32 |
| | | 413 | Lost productivity (income loss) | . | . | -0.6 | -1,536 |
| Older people (65+ years) | Siblings | 102 | Primary sector | 0.0 | 324 | -0.0 | -23 |
| | | 102 | Hospital care | 0.1 | 1,258 | -0.2 | -1,501 |
| | | 102 | Filled prescriptions | 0.0 | 273 | -0.0 | -54 |
| | | 53 | Lost productivity (income loss) | . | . | 1.1 | 21,169 |

#### Supplementary Table 72
#### Dementia - Prevalent cohort 2021
#### Identification of populations of relatives

| *Agegroup of index patients in matched population* | | *Closest relative type identified* | | *Available control with same type of closest relative* |
|---|---|---|---|---|
| Children and young people (0-24 years) | n = 69 | Fathers | n/a | n/a |
| | | Mothers | n/a | n/a |
| | | No available closest relative | n/a | n/a |
| Young adults and adults (25-64 years) | n = 3,525 | Children | n = 674 (19%) | 90% |
| | | Parents | n = 438 (12%) | 86% |
| | | Partners | n = 1,423 (40%) | 96% |
| | | Siblings | n = 769 (22%) | 61% |
| | | Pooled | n = 3,304 (94%) | 85% |
| | | No available closest relative | n = 221 (6%) | n/a |
| Older people (65+ years) | n = 39,540 | Children | n = 20,532 (52%) | 96% |
| | | Partners | n = 13,573 (34%) | 95% |
| | | Siblings | n = 508 (1%) | 35% |
| | | Pooled | n = 34,613 (88%) | 95% |
| | | No available closest relative | n = 4,927 (12%) | n/a |

**Supplementary Table 73**
**Dementia - Prevalent cohort 2021**
**Patient characteristics - cases**

| *Age strata of index patients* | *Characteristic* | |
|---|---|---|
| Children and young people (0-24 years) | Number of persons, n | 61 (100.0) |
| | Male, n (%) | 31 (50.8) |
| | Age, median (Q1;Q3) | 17.1 (11.5;21.6) |
| | CCI, n (%) | |
| | 0 | 31 (50.8) |
| | 1-2 | 23 (37.7) |
| | +3 | 7 (11.5) |
| | Education level, n (%) | |
| | No/Unknown education | 21 (34.4) |
| | Primary | 32 (52.5) |
| | Secondary | 8 (13.1) |
| | Tertiary | 0 (0.0) |
| Young adults and adults (25-64 years) | Number of persons, n | 2,834 (100.0) |
| | Male, n (%) | 1,581 (55.8) |
| | Age, median (Q1;Q3) | 59.2 (54.4;62.5) |
| | CCI, n (%) | |
| | 0 | 750 (26.5) |
| | 1-2 | 1,668 (58.9) |
| | +3 | 416 (14.7) |
| | Education level, n (%) | |
| | No/Unknown education | 174 (6.1) |
| | Primary | 1,007 (35.5) |
| | Secondary | 1,182 (41.7) |
| | Tertiary | 471 (16.6) |
| Older people (65+ years) | Number of persons, n | 32,919 (100.0) |
| | Male, n (%) | 13,713 (41.7) |
| | Age, median (Q1;Q3) | 81.7 (76.4;86.6) |
| | CCI, n (%) | |
| | 0 | 3,205 (9.7) |
| | 1-2 | 20,459 (62.1) |
| | +3 | 9,255 (28.1) |
| | Education level, n (%) | |
| | No/Unknown education | 1,853 (5.6) |
| | Primary | 12,472 (37.9) |
| | Secondary | 13,730 (41.7) |
| | Tertiary | 4,864 (14.8) |

### Supplementary Table 74
### Dementia - Prevalent cohort 2021
### Patient characteristics - relatives - pooled

| *Age strata of index patients* | *Closest relative type* | *Characteristic of family relatives by agestrata of index patients* | *Closest family relatives of brain disorder patients* | *Comparison relative* |
|---|---|---|---|---|
| Children and young people (0-24 years) | Fathers | Number of persons, n | 59 (100.0) | 59 (100.0) |
| | | Male, n (%) | 59 (100.0) | 59 (100.0) |
| | | Age, median (Q1;Q3) | 49.5 (42.8;55.7) | 51.5 (42.9;53.6) |
| | | CCI, n (%) | | |
| | | 0 | * | 54 (91.5) |
| | | 1-2 | * | 5 (8.5) |
| | | +3 | * | 0 (0.0) |
| | | Education level, n (%) | | |
| | | No/Unknown education | 0 (0.0) | 0 (0.0) |
| | | Primary | 12 (20.3) | 12 (20.3) |
| | | Secondary | 27 (45.8) | 27 (45.8) |
| | | Tertiary | 20 (33.9) | 20 (33.9) |
| | | Relation to patient, n (%) | | |
| | | Father | 59 (100.0) | 59 (100.0) |
| Children and young people (0-24 years) | Mothers | Number of persons, n | 55 (100.0) | 55 (100.0) |
| | | Male, n (%) | 0 (0.0) | 0 (0.0) |
| | | Age, median (Q1;Q3) | 48.7 (43.3;52.9) | 47.6 (42.9;51.9) |
| | | CCI, n (%) | | |
| | | 0 | 45 (81.8) | * |
| | | 1-2 | 10 (18.2) | * |
| | | +3 | 0 (0.0) | * |
| | | Education level, n (%) | | |
| | | No/Unknown education | * | * |
| | | Primary | * | * |
| | | Secondary | * | * |
| | | Tertiary | 24 (43.6) | 24 (43.6) |
| | | Relation to patient, n (%) | | |
| | | Mother | 55 (100.0) | 55 (100.0) |
| Young adults and adults (25-64 years) | Pooled | Number of persons, n | 2,834 (100.0) | 2,834 (100.0) |
| | | Male, n (%) | 1,298 (45.8) | 1,298 (45.8) |
| | | Age, median (Q1;Q3) | 57.4 (43.6;63.9) | 57.5 (44.0;63.6) |
| | | CCI, n (%) | | |
| | | 0 | 2,209 (77.9) | 2,303 (81.3) |
| | | 1-2 | 528 (18.6) | 441 (15.6) |
| | | +3 | 97 (3.4) | 90 (3.2) |
| | | Education level, n (%) | | |
| | | No/Unknown education | 73 (2.6) | 73 (2.6) |
| | | Primary | 786 (27.7) | 786 (27.7) |
| | | Secondary | 1,376 (48.6) | 1,376 (48.6) |
| | | Tertiary | 599 (21.1) | 599 (21.1) |
| | | Relation to patient, n (%) | | |
| | | Mother | 309 (10.9) | 309 (10.9) |
| | | Father | 70 (2.5) | 70 (2.5) |
| | | Partner | 1,371 (48.4) | 1,371 (48.4) |
| | | Child | 609 (21.5) | 609 (21.5) |

**Supplementary Table 74**
**Dementia - Prevalent cohort 2021**
**Patient characteristics - relatives - pooled**

| *Age strata of index patients* | *Closest relative type* | *Characteristic of family relatives by agestrata of index patients* | *Closest family relatives of brain disorder patients* | *Comparison relative* |
|---|---|---|---|---|
| | | Sibling | 475 (16.8) | 475 (16.8) |
| Older people (65+ years) | Pooled | Number of persons, n | 32,919 (100.0) | 32,919 (100.0) |
| | | Male, n (%) | 15,275 (46.4) | 15,275 (46.4) |
| | | Age, median (Q1;Q3) | 63.2 (55.9;74.8) | 62.9 (56.0;74.9) |
| | | CCI, n (%) | | |
| | | 0 | 23,613 (71.7) | 24,042 (73.0) |
| | | 1-2 | 7,261 (22.1) | 6,944 (21.1) |
| | | +3 | 2,045 (6.2) | 1,933 (5.9) |
| | | Education level, n (%) | | |
| | | No/Unknown education | 804 (2.4) | 804 (2.4) |
| | | Primary | 6,728 (20.4) | 6,728 (20.4) |
| | | Secondary | 16,525 (50.2) | 16,525 (50.2) |
| | | Tertiary | 8,862 (26.9) | 8,862 (26.9) |
| | | Relation to patient, n (%) | | |
| | | Partner | 12,996 (39.5) | 12,996 (39.5) |
| | | Child | 19,745 (60.0) | 19,745 (60.0) |
| | | Sibling | 178 (0.5) | 178 (0.5) |

**Supplementary Table 75**
**Dementia - Prevalent cohort 2021**
**Patient characteristics - relatives - not pooled**

| *Age strata of index patients* | *Closest relative type* | *Characteristic of family relatives by agestrata of index patients* | *Closest family relatives of brain disorder patients* | *Comparison relative* |
|---|---|---|---|---|
| Children and young people (0-24 years) | Fathers | Number of persons, n | 59 (100.0) | 59 (100.0) |
| | | Male, n (%) | 59 (100.0) | 59 (100.0) |
| | | Age, median (Q1;Q3) | 49.5 (42.8;55.7) | 51.5 (42.9;53.6) |
| | | CCI, n (%) | | |
| | | 0 | * | 54 (91.5) |
| | | 1-2 | * | 5 (8.5) |
| | | +3 | * | 0 (0.0) |
| | | Education level, n (%) | | |
| | | No/Unknown education | 0 (0.0) | 0 (0.0) |
| | | Primary | 12 (20.3) | 12 (20.3) |
| | | Secondary | 27 (45.8) | 27 (45.8) |
| | | Tertiary | 20 (33.9) | 20 (33.9) |
| | | Relation to patient, n (%) | | |
| | | Father | 59 (100.0) | 59 (100.0) |
| Children and young people (0-24 years) | Mothers | Number of persons, n | 55 (100.0) | 55 (100.0) |
| | | Male, n (%) | 0 (0.0) | 0 (0.0) |
| | | Age, median (Q1;Q3) | 48.7 (43.3;52.9) | 47.6 (42.9;51.9) |
| | | CCI, n (%) | | |
| | | 0 | 45 (81.8) | * |
| | | 1-2 | 10 (18.2) | * |
| | | +3 | 0 (0.0) | * |
| | | Education level, n (%) | | |
| | | No/Unknown education | * | * |
| | | Primary | * | * |
| | | Secondary | * | * |
| | | Tertiary | 24 (43.6) | 24 (43.6) |
| | | Relation to patient, n (%) | | |
| | | Mother | 55 (100.0) | 55 (100.0) |
| Young adults and adults (25-64 years) | Children | Number of persons, n | 609 (100.0) | 609 (100.0) |
| | | Male, n (%) | 333 (54.7) | 333 (54.7) |
| | | Age, median (Q1;Q3) | 32.2 (26.0;36.6) | 31.5 (26.2;36.2) |
| | | CCI, n (%) | | |
| | | 0 | 555 (91.1) | 569 (93.4) |
| | | 1-2 | 49 (8.0) | 34 (5.6) |
| | | +3 | 5 (0.8) | 6 (1.0) |
| | | Education level, n (%) | | |
| | | No/Unknown education | 17 (2.8) | 17 (2.8) |
| | | Primary | 210 (34.5) | 210 (34.5) |
| | | Secondary | 248 (40.7) | 248 (40.7) |
| | | Tertiary | 134 (22.0) | 134 (22.0) |
| | | Relation to patient, n (%) | | |
| | | Child | 609 (100.0) | 609 (100.0) |
| Young adults and adults (25-64 years) | Parents | Number of persons, n | 379 (100.0) | 379 (100.0) |
| | | Male, n (%) | 70 (18.5) | 70 (18.5) |
| | | Age, median (Q1;Q3) | 76.2 (69.3;82.3) | 76.9 (68.2;82.3) |

### Supplementary Table 75
### Dementia - Prevalent cohort 2021
### Patient characteristics - relatives - not pooled

| *Age strata of index patients* | *Closest relative type* | *Characteristic of family relatives by agestrata of index patients* | *Closest family relatives of brain disorder patients* | *Comparison relative* |
|---|---|---|---|---|
| | | CCI, n (%) | | |
| | | 0 | 233 (61.5) | 234 (61.7) |
| | | 1-2 | 117 (30.9) | 103 (27.2) |
| | | +3 | 29 (7.7) | 42 (11.1) |
| | | Education level, n (%) | | |
| | | No/Unknown education | 6 (1.6) | 6 (1.6) |
| | | Primary | 157 (41.4) | 157 (41.4) |
| | | Secondary | 155 (40.9) | 155 (40.9) |
| | | Tertiary | 61 (16.1) | 61 (16.1) |
| | | Relation to patient, n (%) | | |
| | | Mother | 309 (81.5) | 309 (81.5) |
| | | Father | 70 (18.5) | 70 (18.5) |
| Young adults and adults (25-64 years) | Partners | Number of persons, n | 1,371 (100.0) | 1,371 (100.0) |
| | | Male, n (%) | 626 (45.7) | 626 (45.7) |
| | | Age, median (Q1;Q3) | 58.7 (53.3;63.0) | 58.7 (53.8;63.0) |
| | | CCI, n (%) | | |
| | | 0 | 1,054 (76.9) | 1,122 (81.8) |
| | | 1-2 | 269 (19.6) | 218 (15.9) |
| | | +3 | 48 (3.5) | 31 (2.3) |
| | | Education level, n (%) | | |
| | | No/Unknown education | 47 (3.4) | 47 (3.4) |
| | | Primary | 317 (23.1) | 317 (23.1) |
| | | Secondary | 703 (51.3) | 703 (51.3) |
| | | Tertiary | 304 (22.2) | 304 (22.2) |
| | | Relation to patient, n (%) | | |
| | | Partner | 1,371 (100.0) | 1,371 (100.0) |
| Young adults and adults (25-64 years) | Siblings | Number of persons, n | 475 (100.0) | 475 (100.0) |
| | | Male, n (%) | 269 (56.6) | 269 (56.6) |
| | | Age, median (Q1;Q3) | 59.5 (55.5;63.3) | 59.4 (54.9;63.0) |
| | | CCI, n (%) | | |
| | | 0 | 367 (77.3) | 378 (79.6) |
| | | 1-2 | 93 (19.6) | 86 (18.1) |
| | | +3 | 15 (3.2) | 11 (2.3) |
| | | Education level, n (%) | | |
| | | No/Unknown education | * | * |
| | | Primary | 102 (21.5) | 102 (21.5) |
| | | Secondary | 270 (56.8) | 270 (56.8) |
| | | Tertiary | * | * |
| | | Relation to patient, n (%) | | |
| | | Sibling | 475 (100.0) | 475 (100.0) |
| Older people (65+ years) | Children | Number of persons, n | 19,745 (100.0) | 19,745 (100.0) |
| | | Male, n (%) | 10,009 (50.7) | 10,009 (50.7) |
| | | Age, median (Q1;Q3) | 57.5 (52.3;62.0) | 57.5 (52.3;61.7) |
| | | CCI, n (%) | | |
| | | 0 | 16,055 (81.3) | 16,271 (82.4) |

**Supplementary Table 75**
**Dementia - Prevalent cohort 2021**
**Patient characteristics - relatives - not pooled**

| *Age strata of index patients* | *Closest relative type* | *Characteristic of family relatives by agestrata of index patients* | *Closest family relatives of brain disorder patients* | *Comparison relative* |
|---|---|---|---|---|
| | | 1-2 | 3,157 (16.0) | 2,984 (15.1) |
| | | +3 | 533 (2.7) | 490 (2.5) |
| | | Education level, n (%) | | |
| | | No/Unknown education | 105 (0.5) | 105 (0.5) |
| | | Primary | 2,960 (15.0) | 2,960 (15.0) |
| | | Secondary | 10,308 (52.2) | 10,308 (52.2) |
| | | Tertiary | 6,372 (32.3) | 6,372 (32.3) |
| | | Relation to patient, n (%) | | |
| | | Child | 19,745 (100.0) | 19,745 (100.0) |
| Older people (65+ years) | Partners | Number of persons, n | 12,996 (100.0) | 12,996 (100.0) |
| | | Male, n (%) | 5,168 (39.8) | 5,168 (39.8) |
| | | Age, median (Q1;Q3) | 77.0 (72.5;82.0) | 77.2 (72.7;81.8) |
| | | CCI, n (%) | | |
| | | 0 | 7,428 (57.2) | 7,642 (58.8) |
| | | 1-2 | 4,064 (31.3) | 3,922 (30.2) |
| | | +3 | 1,504 (11.6) | 1,432 (11.0) |
| | | Education level, n (%) | | |
| | | No/Unknown education | 697 (5.4) | 697 (5.4) |
| | | Primary | 3,740 (28.8) | 3,740 (28.8) |
| | | Secondary | 6,109 (47.0) | 6,109 (47.0) |
| | | Tertiary | 2,450 (18.9) | 2,450 (18.9) |
| | | Relation to patient, n (%) | | |
| | | Partner | 12,996 (100.0) | 12,996 (100.0) |
| Older people (65+ years) | Siblings | Number of persons, n | 178 (100.0) | 178 (100.0) |
| | | Male, n (%) | 98 (55.1) | 98 (55.1) |
| | | Age, median (Q1;Q3) | 63.2 (59.6;66.2) | 62.9 (60.4;66.1) |
| | | CCI, n (%) | | |
| | | 0 | 130 (73.0) | 129 (72.5) |
| | | 1-2 | 40 (22.5) | 38 (21.3) |
| | | +3 | 8 (4.5) | 11 (6.2) |
| | | Education level, n (%) | | |
| | | No/Unknown education | * | * |
| | | Primary | * | * |
| | | Secondary | 108 (60.7) | 108 (60.7) |
| | | Tertiary | 40 (22.5) | 40 (22.5) |
| | | Relation to patient, n (%) | | |
| | | Sibling | 178 (100.0) | 178 (100.0) |

#### Supplementary Table 76
#### Dementia - Prevalent cohort 2021
#### Cost analysis - pooled

| *Age strata of index patients* | *Closest relative type* | *Number of index patients included in cost component calculations* | *Cost component* | *Actual costs* | | *Attributable costs* | |
|---|---|---|---|---|---|---|---|
| | | | | *Total (million EUR)* | *Per person (EUR)* | *Total (million EUR)* | *Per person (EUR)* |
| Children and young people (0-24 years) | Fathers | 59 | Primary sector | 0.0 | 255 | 0.0 | 57 |
| | | 59 | Hospital care | 0.1 | 2,167 | 0.1 | 1,204 |
| | | 59 | Filled prescriptions | 0.0 | 127 | -0.0 | -49 |
| | | 58 | Lost productivity (income loss) | . | . | 1.0 | 16,432 |
| Children and young people (0-24 years) | Mothers | 55 | Primary sector | 0.0 | 430 | 0.0 | 33 |
| | | 55 | Hospital care | 0.1 | 1,099 | -0.0 | -47 |
| | | 55 | Filled prescriptions | 0.0 | 238 | 0.0 | 93 |
| | | 54 | Lost productivity (income loss) | . | . | -0.2 | -2,863 |
| Young adults and adults (25-64 years) | Pooled | 2,834 | Primary sector | 1.1 | 381 | 0.1 | 35 |
| | | 2,834 | Hospital care | 6.9 | 2,464 | 1.1 | 391 |
| | | 2,834 | Filled prescriptions | 1.0 | 363 | 0.3 | 99 |
| | | 1,960 | Lost productivity (income loss) | . | . | 15.3 | 7,812 |
| Older people (65+ years) | Pooled | 32,919 | Primary sector | 14.4 | 440 | 1.1 | 34 |
| | | 32,919 | Hospital care | 106.0 | 3,249 | 12.6 | 386 |
| | | 32,919 | Filled prescriptions | 13.8 | 424 | 1.8 | 55 |
| | | 16,183 | Lost productivity (income loss) | . | . | 42.1 | 2,599 |

**Supplementary Table 77**
**Dementia - Prevalent cohort 2021**
**Cost analysis - not pooled**

| *Age strata of index patients* | *Closest relative type* | *Number of index patients included in cost component calculations* | *Cost component* | *Actual costs* | | *Attributable costs* | |
|---|---|---|---|---|---|---|---|
| | | | | *Total (million EUR)* | *Per person (EUR)* | *Total (million EUR)* | *Per person (EUR)* |
| Children and young people (0-24 years) | Fathers | 59 | Primary sector | 0.0 | 255 | 0.0 | 57 |
| | | 59 | Hospital care | 0.1 | 2,167 | 0.1 | 1,204 |
| | | 59 | Filled prescriptions | 0.0 | 127 | -0.0 | -49 |
| | | 58 | Lost productivity (income loss) | . | . | 1.0 | 16,432 |
| Children and young people (0-24 years) | Mothers | 55 | Primary sector | 0.0 | 430 | 0.0 | 33 |
| | | 55 | Hospital care | 0.1 | 1,099 | -0.0 | -47 |
| | | 55 | Filled prescriptions | 0.0 | 238 | 0.0 | 93 |
| | | 54 | Lost productivity (income loss) | . | . | -0.2 | -2,863 |
| Young adults and adults (25-64 years) | Children | 609 | Primary sector | 0.2 | 285 | 0.0 | 30 |
| | | 609 | Hospital care | 1.1 | 1,774 | 0.2 | 282 |
| | | 609 | Filled prescriptions | 0.1 | 176 | 0.0 | 75 |
| | | 548 | Lost productivity (income loss) | . | . | 1.2 | 2,138 |
| Young adults and adults (25-64 years) | Parents | 379 | Primary sector | 0.2 | 534 | 0.0 | 35 |
| | | 379 | Hospital care | 1.3 | 3,417 | -0.1 | -300 |
| | | 379 | Filled prescriptions | 0.2 | 653 | 0.0 | 131 |
| | | 58 | Lost productivity (income loss) | . | . | 0.2 | 3,288 |
| Young adults and adults (25-64 years) | Partners | 1,371 | Primary sector | 0.5 | 391 | 0.1 | 49 |
| | | 1,371 | Hospital care | 3.4 | 2,513 | 0.9 | 678 |
| | | 1,371 | Filled prescriptions | 0.5 | 359 | 0.1 | 107 |
| | | 1,017 | Lost productivity (income loss) | . | . | 11.6 | 11,364 |
| Young adults and adults (25-64 years) | Siblings | 475 | Primary sector | 0.2 | 356 | -0.0 | -0 |
| | | 475 | Hospital care | 1.2 | 2,460 | 0.1 | 245 |
| | | 475 | Filled prescriptions | 0.2 | 386 | 0.0 | 80 |
| | | 337 | Lost productivity (income loss) | . | . | 2.4 | 7,098 |
| Older people (65+ years) | Children | 19,745 | Primary sector | 6.9 | 352 | 0.6 | 28 |
| | | 19,745 | Hospital care | 46.7 | 2,373 | 6.3 | 318 |
| | | 19,745 | Filled prescriptions | 5.9 | 302 | 0.9 | 46 |
| | | 15,792 | Lost productivity (income loss) | . | . | 40.0 | 2,535 |
| Older people (65+ years) | Partners | 12,996 | Primary sector | 7.4 | 577 | 0.6 | 45 |
| | | 12,996 | Hospital care | 58.6 | 4,592 | 6.3 | 493 |
| | | 12,996 | Filled prescriptions | 7.8 | 613 | 0.9 | 67 |
| | | 297 | Lost productivity (income loss) | . | . | 0.9 | 2,919 |
| Older people (65+ years) | Siblings | 178 | Primary sector | 0.1 | 385 | -0.0 | -12 |
| | | 178 | Hospital care | 0.7 | 3,778 | 0.1 | 321 |
| | | 178 | Filled prescriptions | 0.1 | 420 | 0.0 | 155 |
| | | 94 | Lost productivity (income loss) | . | . | 1.2 | 12,279 |

**Supplementary Table 78**
**Depression - Incident cohort 2016-2021**
**Identification of populations of relatives**

| ***Agegroup of index patients in matched population*** | | ***Closest relative type identified*** | | ***Available control with same type of closest relative*** |
|---|---|---|---|---|
| Children and young people (0-24 years) | n = 35,182 | Fathers | n = 23,140 (66%) | 96% |
| | | Mothers | n = 20,475 (58%) | 96% |
| | | No available closest relative | n = 6,558 (19%) | n/a |
| Young adults and adults (25-64 years) | n = 94,037 | Children | n = 19,023 (20%) | 94% |
| | | Parents | n = 15,290 (16%) | 89% |
| | | Partners | n = 41,183 (44%) | 95% |
| | | Siblings | n = 12,188 (13%) | 58% |
| | | Pooled | n = 87,684 (93%) | 89% |
| | | No available closest relative | n = 6,353 (7%) | n/a |
| Older people (65+ years) | n = 62,987 | Children | n = 30,287 (48%) | 95% |
| | | Partners | n = 21,587 (34%) | 95% |
| | | Siblings | n = 839 (1%) | 32% |
| | | Pooled | n = 52,713 (84%) | 94% |
| | | No available closest relative | n = 10,274 (16%) | n/a |

**Supplementary Table 79**
**Depression - Incident cohort 2016-2021**
**Patient characteristics - cases**

| ***Age strata of index patients*** | ***Characteristic*** | |
|---|---|---|
| Children and young people (0-24 years) | Number of persons, n | 27,584 (100.0) |
| | Male, n (%) | 9,792 (35.5) |
| | Age, median (Q1;Q3) | 20.4 (18.0;22.7) |
| | CCI, n (%) | |
| | 0 | 25,695 (93.2) |
| | 1-2 | 1,826 (6.6) |
| | +3 | 63 (0.2) |
| | Education level, n (%) | |
| | No/Unknown education | 662 (2.4) |
| | Primary | 18,639 (67.6) |
| | Secondary | 7,689 (27.9) |
| | Tertiary | 594 (2.2) |
| Young adults and adults (25-64 years) | Number of persons, n | 78,169 (100.0) |
| | Male, n (%) | 35,934 (46.0) |
| | Age, median (Q1;Q3) | 43.7 (33.0;53.8) |
| | CCI, n (%) | |
| | 0 | 63,830 (81.7) |
| | 1-2 | 11,789 (15.1) |
| | +3 | 2,550 (3.3) |
| | Education level, n (%) | |
| | No/Unknown education | 3,568 (4.6) |
| | Primary | 18,834 (24.1) |
| | Secondary | 34,781 (44.5) |
| | Tertiary | 20,986 (26.8) |
| Older people (65+ years) | Number of persons, n | 49,846 (100.0) |
| | Male, n (%) | 22,232 (44.6) |
| | Age, median (Q1;Q3) | 78.3 (72.8;84.0) |
| | CCI, n (%) | |
| | 0 | 16,974 (34.1) |
| | 1-2 | 21,263 (42.7) |
| | +3 | 11,609 (23.3) |
| | Education level, n (%) | |
| | No/Unknown education | 2,695 (5.4) |
| | Primary | 18,432 (37.0) |
| | Secondary | 21,394 (42.9) |
| | Tertiary | 7,325 (14.7) |

**Supplementary Table 80**
**Depression - Incident cohort 2016-2021**
**Patient characteristics - relatives - pooled**

| *Age strata of index patients* | *Closest relative type* | *Characteristic of family relatives by agestrata of index patients* | *Closest family relatives of brain disorder patients* | *Comparison relative* |
|---|---|---|---|---|
| Children and young people (0-24 years) | Fathers | Number of persons, n | 22,255 (100.0) | 22,255 (100.0) |
| | | Male, n (%) | 22,255 (100.0) | 22,255 (100.0) |
| | | Age, median (Q1;Q3) | 52.0 (48.3;55.9) | 52.1 (48.5;55.5) |
| | | CCI, n (%) | | |
| | | 0 | 19,264 (86.6) | 19,654 (88.3) |
| | | 1-2 | 2,671 (12.0) | 2,269 (10.2) |
| | | +3 | 320 (1.4) | 332 (1.5) |
| | | Education level, n (%) | | |
| | | No/Unknown education | 241 (1.1) | 241 (1.1) |
| | | Primary | 4,342 (19.5) | 4,342 (19.5) |
| | | Secondary | 11,846 (53.2) | 11,846 (53.2) |
| | | Tertiary | 5,826 (26.2) | 5,826 (26.2) |
| | | Relation to patient, n (%) | | |
| | | Father | 22,255 (100.0) | 22,255 (100.0) |
| Children and young people (0-24 years) | Mothers | Number of persons, n | 19,713 (100.0) | 19,713 (100.0) |
| | | Male, n (%) | 0 (0.0) | 0 (0.0) |
| | | Age, median (Q1;Q3) | 50.0 (46.3;53.6) | 50.0 (46.7;53.3) |
| | | CCI, n (%) | | |
| | | 0 | 17,113 (86.8) | 17,418 (88.4) |
| | | 1-2 | 2,365 (12.0) | 2,084 (10.6) |
| | | +3 | 235 (1.2) | 211 (1.1) |
| | | Education level, n (%) | | |
| | | No/Unknown education | 138 (0.7) | 138 (0.7) |
| | | Primary | 2,919 (14.8) | 2,919 (14.8) |
| | | Secondary | 9,109 (46.2) | 9,109 (46.2) |
| | | Tertiary | 7,547 (38.3) | 7,547 (38.3) |
| | | Relation to patient, n (%) | | |
| | | Mother | 19,713 (100.0) | 19,713 (100.0) |
| Young adults and adults (25-64 years) | Pooled | Number of persons, n | 78,169 (100.0) | 78,169 (100.0) |
| | | Male, n (%) | 41,518 (53.1) | 41,518 (53.1) |
| | | Age, median (Q1;Q3) | 43.3 (28.4;56.9) | 43.3 (28.3;57.0) |
| | | CCI, n (%) | | |
| | | 0 | 67,642 (86.5) | 68,680 (87.9) |
| | | 1-2 | 9,066 (11.6) | 8,214 (10.5) |
| | | +3 | 1,461 (1.9) | 1,275 (1.6) |
| | | Education level, n (%) | | |
| | | No/Unknown education | 8,355 (10.7) | 8,355 (10.7) |
| | | Primary | 19,646 (25.1) | 19,646 (25.1) |
| | | Secondary | 32,405 (41.5) | 32,405 (41.5) |
| | | Tertiary | 17,763 (22.7) | 17,763 (22.7) |
| | | Relation to patient, n (%) | | |
| | | Mother | 9,306 (11.9) | 9,306 (11.9) |
| | | Father | 4,396 (5.6) | 4,396 (5.6) |
| | | Partner | 39,235 (50.2) | 39,235 (50.2) |
| | | Child | 18,069 (23.1) | 18,069 (23.1) |

#### Supplementary Table 80
#### Depression - Incident cohort 2016-2021
#### Patient characteristics - relatives - pooled

| *Age strata of index patients* | *Closest relative type* | *Characteristic of family relatives by agestrata of index patients* | *Closest family relatives of brain disorder patients* | *Comparison relative* |
|---|---|---|---|---|
| | | Sibling | 7,163 (9.2) | 7,163 (9.2) |
| Older people (65+ years) | Pooled | Number of persons, n | 49,846 (100.0) | 49,846 (100.0) |
| | | Male, n (%) | 25,256 (50.7) | 25,256 (50.7) |
| | | Age, median (Q1;Q3) | 61.1 (52.7;73.2) | 60.8 (52.6;73.3) |
| | | CCI, n (%) | | |
| | | 0 | 36,837 (73.9) | 37,905 (76.0) |
| | | 1-2 | 10,122 (20.3) | 9,563 (19.2) |
| | | +3 | 2,887 (5.8) | 2,378 (4.8) |
| | | Education level, n (%) | | |
| | | No/Unknown education | 1,256 (2.5) | 1,256 (2.5) |
| | | Primary | 10,342 (20.7) | 10,342 (20.7) |
| | | Secondary | 25,133 (50.4) | 25,133 (50.4) |
| | | Tertiary | 13,115 (26.3) | 13,115 (26.3) |
| | | Relation to patient, n (%) | | |
| | | Partner | 20,644 (41.4) | 20,644 (41.4) |
| | | Child | 28,928 (58.0) | 28,928 (58.0) |
| | | Sibling | 274 (0.5) | 274 (0.5) |

**Supplementary Table 81**
**Depression - Incident cohort 2016-2021**
**Patient characteristics - relatives - not pooled**

| *Age strata of index patients* | *Closest relative type* | *Characteristic of family relatives by agestrata of index patients* | *Closest family relatives of brain disorder patients* | *Comparison relative* |
|---|---|---|---|---|
| Children and young people (0-24 years) | Fathers | Number of persons, n | 22,255 (100.0) | 22,255 (100.0) |
| | | Male, n (%) | 22,255 (100.0) | 22,255 (100.0) |
| | | Age, median (Q1;Q3) | 52.0 (48.3;55.9) | 52.1 (48.5;55.5) |
| | | CCI, n (%) | | |
| | | 0 | 19,264 (86.6) | 19,654 (88.3) |
| | | 1-2 | 2,671 (12.0) | 2,269 (10.2) |
| | | +3 | 320 (1.4) | 332 (1.5) |
| | | Education level, n (%) | | |
| | | No/Unknown education | 241 (1.1) | 241 (1.1) |
| | | Primary | 4,342 (19.5) | 4,342 (19.5) |
| | | Secondary | 11,846 (53.2) | 11,846 (53.2) |
| | | Tertiary | 5,826 (26.2) | 5,826 (26.2) |
| | | Relation to patient, n (%) | | |
| | | Father | 22,255 (100.0) | 22,255 (100.0) |
| Children and young people (0-24 years) | Mothers | Number of persons, n | 19,713 (100.0) | 19,713 (100.0) |
| | | Male, n (%) | 0 (0.0) | 0 (0.0) |
| | | Age, median (Q1;Q3) | 50.0 (46.3;53.6) | 50.0 (46.7;53.3) |
| | | CCI, n (%) | | |
| | | 0 | 17,113 (86.8) | 17,418 (88.4) |
| | | 1-2 | 2,365 (12.0) | 2,084 (10.6) |
| | | +3 | 235 (1.2) | 211 (1.1) |
| | | Education level, n (%) | | |
| | | No/Unknown education | 138 (0.7) | 138 (0.7) |
| | | Primary | 2,919 (14.8) | 2,919 (14.8) |
| | | Secondary | 9,109 (46.2) | 9,109 (46.2) |
| | | Tertiary | 7,547 (38.3) | 7,547 (38.3) |
| | | Relation to patient, n (%) | | |
| | | Mother | 19,713 (100.0) | 19,713 (100.0) |
| Young adults and adults (25-64 years) | Children | Number of persons, n | 18,069 (100.0) | 18,069 (100.0) |
| | | Male, n (%) | 9,605 (53.2) | 9,605 (53.2) |
| | | Age, median (Q1;Q3) | 17.5 (9.8;26.0) | 17.3 (9.3;25.6) |
| | | CCI, n (%) | | |
| | | 0 | 16,955 (93.8) | 17,098 (94.6) |
| | | 1-2 | 1,075 (5.9) | 939 (5.2) |
| | | +3 | 39 (0.2) | 32 (0.2) |
| | | Education level, n (%) | | |
| | | No/Unknown education | 6,393 (35.4) | 6,393 (35.4) |
| | | Primary | 6,497 (36.0) | 6,497 (36.0) |
| | | Secondary | 3,469 (19.2) | 3,469 (19.2) |
| | | Tertiary | 1,710 (9.5) | 1,710 (9.5) |
| | | Relation to patient, n (%) | | |
| | | Child | 18,069 (100.0) | 18,069 (100.0) |
| Young adults and adults (25-64 years) | Parents | Number of persons, n | 13,702 (100.0) | 13,702 (100.0) |
| | | Male, n (%) | 4,396 (32.1) | 4,396 (32.1) |
| | | Age, median (Q1;Q3) | 63.0 (56.8;71.3) | 62.9 (57.0;71.4) |

### Supplementary Table 81
### Depression - Incident cohort 2016-2021
### Patient characteristics - relatives - not pooled

| *Age strata of index patients* | *Closest relative type* | *Characteristic of family relatives by agestrata of index patients* | *Closest family relatives of brain disorder patients* | *Comparison relative* |
|---|---|---|---|---|
| | | CCI, n (%) | | |
| | | 0 | 9,952 (72.6) | 10,217 (74.6) |
| | | 1-2 | 2,997 (21.9) | 2,844 (20.8) |
| | | +3 | 753 (5.5) | 641 (4.7) |
| | | Education level, n (%) | | |
| | | No/Unknown education | 233 (1.7) | 233 (1.7) |
| | | Primary | 3,778 (27.6) | 3,778 (27.6) |
| | | Secondary | 6,808 (49.7) | 6,808 (49.7) |
| | | Tertiary | 2,883 (21.0) | 2,883 (21.0) |
| | | Relation to patient, n (%) | | |
| | | Mother | 9,306 (67.9) | 9,306 (67.9) |
| | | Father | 4,396 (32.1) | 4,396 (32.1) |
| Young adults and adults (25-64 years) | Partners | Number of persons, n | 39,235 (100.0) | 39,235 (100.0) |
| | | Male, n (%) | 23,465 (59.8) | 23,465 (59.8) |
| | | Age, median (Q1;Q3) | 45.4 (35.4;54.9) | 45.5 (35.6;54.9) |
| | | CCI, n (%) | | |
| | | 0 | 34,465 (87.8) | 35,046 (89.3) |
| | | 1-2 | 4,212 (10.7) | 3,717 (9.5) |
| | | +3 | 558 (1.4) | 472 (1.2) |
| | | Education level, n (%) | | |
| | | No/Unknown education | 1,694 (4.3) | 1,694 (4.3) |
| | | Primary | 7,451 (19.0) | 7,451 (19.0) |
| | | Secondary | 18,481 (47.1) | 18,481 (47.1) |
| | | Tertiary | 11,609 (29.6) | 11,609 (29.6) |
| | | Relation to patient, n (%) | | |
| | | Partner | 39,235 (100.0) | 39,235 (100.0) |
| Young adults and adults (25-64 years) | Siblings | Number of persons, n | 7,163 (100.0) | 7,163 (100.0) |
| | | Male, n (%) | 4,052 (56.6) | 4,052 (56.6) |
| | | Age, median (Q1;Q3) | 41.8 (28.2;56.2) | 42.1 (28.0;56.2) |
| | | CCI, n (%) | | |
| | | 0 | 6,270 (87.5) | 6,319 (88.2) |
| | | 1-2 | 782 (10.9) | 714 (10.0) |
| | | +3 | 111 (1.5) | 130 (1.8) |
| | | Education level, n (%) | | |
| | | No/Unknown education | 35 (0.5) | 35 (0.5) |
| | | Primary | 1,920 (26.8) | 1,920 (26.8) |
| | | Secondary | 3,647 (50.9) | 3,647 (50.9) |
| | | Tertiary | 1,561 (21.8) | 1,561 (21.8) |
| | | Relation to patient, n (%) | | |
| | | Sibling | 7,163 (100.0) | 7,163 (100.0) |
| Older people (65+ years) | Children | Number of persons, n | 28,928 (100.0) | 28,928 (100.0) |
| | | Male, n (%) | 15,461 (53.4) | 15,461 (53.4) |
| | | Age, median (Q1;Q3) | 54.1 (48.2;59.1) | 53.9 (48.2;58.8) |
| | | CCI, n (%) | | |
| | | 0 | 24,673 (85.3) | 25,069 (86.7) |

**Supplementary Table 81**
**Depression - Incident cohort 2016-2021**
**Patient characteristics - relatives - not pooled**

| *Age strata of index patients* | *Closest relative type* | *Characteristic of family relatives by agestrata of index patients* | *Closest family relatives of brain disorder patients* | *Comparison relative* |
|---|---|---|---|---|
| | | 1-2 | 3,702 (12.8) | 3,399 (11.7) |
| | | +3 | 553 (1.9) | 460 (1.6) |
| | | Education level, n (%) | | |
| | | No/Unknown education | 131 (0.5) | 131 (0.5) |
| | | Primary | 4,225 (14.6) | 4,225 (14.6) |
| | | Secondary | 15,223 (52.6) | 15,223 (52.6) |
| | | Tertiary | 9,349 (32.3) | 9,349 (32.3) |
| | | Relation to patient, n (%) | | |
| | | Child | 28,928 (100.0) | 28,928 (100.0) |
| Older people (65+ years) | Partners | Number of persons, n | 20,644 (100.0) | 20,644 (100.0) |
| | | Male, n (%) | 9,642 (46.7) | 9,642 (46.7) |
| | | Age, median (Q1;Q3) | 75.2 (70.3;80.4) | 75.1 (70.4;80.2) |
| | | CCI, n (%) | | |
| | | 0 | 11,957 (57.9) | 12,628 (61.2) |
| | | 1-2 | 6,365 (30.8) | 6,109 (29.6) |
| | | +3 | 2,322 (11.2) | 1,907 (9.2) |
| | | Education level, n (%) | | |
| | | No/Unknown education | 1,125 (5.4) | 1,125 (5.4) |
| | | Primary | 6,069 (29.4) | 6,069 (29.4) |
| | | Secondary | 9,740 (47.2) | 9,740 (47.2) |
| | | Tertiary | 3,710 (18.0) | 3,710 (18.0) |
| | | Relation to patient, n (%) | | |
| | | Partner | 20,644 (100.0) | 20,644 (100.0) |
| Older people (65+ years) | Siblings | Number of persons, n | 274 (100.0) | 274 (100.0) |
| | | Male, n (%) | 153 (55.8) | 153 (55.8) |
| | | Age, median (Q1;Q3) | 62.5 (59.3;65.0) | 62.0 (59.0;65.2) |
| | | CCI, n (%) | | |
| | | 0 | 207 (75.5) | 208 (75.9) |
| | | 1-2 | 55 (20.1) | 55 (20.1) |
| | | +3 | 12 (4.4) | 11 (4.0) |
| | | Education level, n (%) | | |
| | | No/Unknown education | 0 (0.0) | 0 (0.0) |
| | | Primary | 48 (17.5) | 48 (17.5) |
| | | Secondary | 170 (62.0) | 170 (62.0) |
| | | Tertiary | 56 (20.4) | 56 (20.4) |
| | | Relation to patient, n (%) | | |
| | | Sibling | 274 (100.0) | 274 (100.0) |

#### Supplementary Table 82
#### Depression - Incident cohort 2016-2021
#### Cost analysis - pooled

| | | | | Actual costs | | Attributable costs | |
|---|---|---|---|---|---|---|---|
| *Age strata of index patients* | *Closest relative type* | *Number of index patients included in cost component calculations* | *Cost component* | *Total (million EUR)* | *Per person (EUR)* | *Total (million EUR)* | *Per person (EUR)* |
| Children and young people (0-24 years) | Fathers | 22,255 | Primary sector | 5.0 | 226 | 0.3 | 13 |
| | | 22,255 | Hospital care | 33.6 | 1,511 | 2.4 | 110 |
| | | 22,255 | Filled prescriptions | 4.3 | 192 | 0.8 | 36 |
| | | 21,220 | Lost productivity (income loss) | . | . | 78.2 | 3,687 |
| Children and young people (0-24 years) | Mothers | 19,713 | Primary sector | 6.6 | 335 | 0.7 | 38 |
| | | 19,713 | Hospital care | 30.6 | 1,554 | 4.9 | 249 |
| | | 19,713 | Filled prescriptions | 3.8 | 191 | 0.8 | 43 |
| | | 19,506 | Lost productivity (income loss) | . | . | 49.9 | 2,558 |
| Young adults and adults (25-64 years) | Pooled | 78,169 | Primary sector | 21.8 | 280 | 2.4 | 31 |
| | | 78,169 | Hospital care | 134.8 | 1,732 | 19.7 | 253 |
| | | 78,169 | Filled prescriptions | 15.5 | 199 | 2.4 | 31 |
| | | 57,155 | Lost productivity (income loss) | . | . | 271.6 | 4,753 |
| Older people (65+ years) | Pooled | 49,846 | Primary sector | 19.2 | 390 | 1.5 | 30 |
| | | 49,846 | Hospital care | 142.3 | 2,889 | 17.7 | 359 |
| | | 49,846 | Filled prescriptions | 17.2 | 350 | 2.0 | 40 |
| | | 26,556 | Lost productivity (income loss) | . | . | 32.9 | 1,240 |

### Supplementary Table 83
### Depression - Incident cohort 2016-2021
### Cost analysis - not pooled

| Age strata of index patients | Closest relative type | Number of index patients included in cost component calculations | Cost component | Actual costs | | Attributable costs | |
|---|---|---|---|---|---|---|---|
| | | | | Total (million EUR) | Per person (EUR) | Total (million EUR) | Per person (EUR) |
| Children and young people (0-24 years) | Fathers | 22,255 | Primary sector | 5.0 | 226 | 0.3 | 13 |
| | | 22,255 | Hospital care | 33.6 | 1,511 | 2.4 | 110 |
| | | 22,255 | Filled prescriptions | 4.3 | 192 | 0.8 | 36 |
| | | 21,220 | Lost productivity (income loss) | . | . | 78.2 | 3,687 |
| Children and young people (0-24 years) | Mothers | 19,713 | Primary sector | 6.6 | 335 | 0.7 | 38 |
| | | 19,713 | Hospital care | 30.6 | 1,554 | 4.9 | 249 |
| | | 19,713 | Filled prescriptions | 3.8 | 191 | 0.8 | 43 |
| | | 19,506 | Lost productivity (income loss) | . | . | 49.9 | 2,558 |
| Young adults and adults (25-64 years) | Children | 18,069 | Primary sector | 3.6 | 200 | 0.5 | 26 |
| | | 18,069 | Hospital care | 19.6 | 1,088 | 3.4 | 192 |
| | | 18,069 | Filled prescriptions | 1.8 | 98 | 0.4 | 23 |
| | | 8,137 | Lost productivity (income loss) | . | . | 20.9 | 2,567 |
| Young adults and adults (25-64 years) | Parents | 13,702 | Primary sector | 5.4 | 399 | 0.4 | 26 |
| | | 13,702 | Hospital care | 39.2 | 2,883 | 3.9 | 289 |
| | | 13,702 | Filled prescriptions | 5.5 | 402 | 0.8 | 60 |
| | | 6,473 | Lost productivity (income loss) | . | . | 10.5 | 1,617 |
| Young adults and adults (25-64 years) | Partners | 39,235 | Primary sector | 10.9 | 278 | 1.5 | 38 |
| | | 39,235 | Hospital care | 64.4 | 1,648 | 11.1 | 284 |
| | | 39,235 | Filled prescriptions | 6.9 | 176 | 1.1 | 28 |
| | | 36,119 | Lost productivity (income loss) | . | . | 232.4 | 6,433 |
| Young adults and adults (25-64 years) | Siblings | 7,163 | Primary sector | 1.9 | 265 | 0.1 | 14 |
| | | 7,163 | Hospital care | 11.5 | 1,613 | 1.2 | 166 |
| | | 7,163 | Filled prescriptions | 1.3 | 189 | 0.1 | 15 |
| | | 6,426 | Lost productivity (income loss) | . | . | 7.9 | 1,232 |
| Older people (65+ years) | Children | 28,928 | Primary sector | 8.4 | 289 | 0.6 | 22 |
| | | 28,928 | Hospital care | 49.1 | 1,702 | 4.9 | 171 |
| | | 28,928 | Filled prescriptions | 6.1 | 211 | 0.6 | 22 |
| | | 25,745 | Lost productivity (income loss) | . | . | 34.4 | 1,335 |
| Older people (65+ years) | Partners | 20,644 | Primary sector | 10.8 | 536 | 0.9 | 43 |
| | | 20,644 | Hospital care | 92.4 | 4,586 | 12.4 | 614 |
| | | 20,644 | Filled prescriptions | 11.1 | 549 | 1.3 | 66 |
| | | 672 | Lost productivity (income loss) | . | . | -1.8 | -2,697 |
| Older people (65+ years) | Siblings | 274 | Primary sector | 0.1 | 340 | 0.0 | 20 |
| | | 274 | Hospital care | 0.9 | 3,149 | 0.4 | 1,423 |
| | | 274 | Filled prescriptions | 0.1 | 351 | 0.0 | 45 |
| | | 139 | Lost productivity (income loss) | . | . | 0.4 | 2,578 |

#### Supplementary Table 84
#### Depression - Prevalent cohort 2021
#### Identification of populations of relatives

| *Agegroup of index patients in matched population* | | *Closest relative type identified* | | *Available control with same type of closest relative* |
|---|---|---|---|---|
| Children and young people (0-24 years) | n = 29,108 | Fathers | n = 18,831 (65%) | 96% |
| | | Mothers | n = 16,253 (56%) | 96% |
| | | No available closest relative | n = 5,519 (19%) | n/a |
| Young adults and adults (25-64 years) | n = 491,154 | Children | n = 119,641 (24%) | 94% |
| | | Parents | n = 82,168 (17%) | 87% |
| | | Partners | n = 199,086 (41%) | 95% |
| | | Siblings | n = 69,007 (14%) | 56% |
| | | Pooled | n = 469,902 (96%) | 88% |
| | | No available closest relative | n = 21,252 (4%) | n/a |
| Older people (65+ years) | n = 223,742 | Children | n = 100,368 (45%) | 94% |
| | | Partners | n = 78,109 (35%) | 95% |
| | | Siblings | n = 7,446 (3%) | 49% |
| | | Pooled | n = 185,923 (83%) | 93% |
| | | No available closest relative | n = 37,819 (17%) | n/a |

**Supplementary Table 85**
**Depression - Prevalent cohort 2021**
**Patient characteristics - cases**

| *Age strata of index patients* | *Characteristic* | |
|---|---|---|
| Children and young people (0-24 years) | Number of persons, n | 22,751 (100.0) |
| | Male, n (%) | 7,723 (33.9) |
| | Age, median (Q1;Q3) | 22.1 (19.9;23.7) |
| | CCI, n (%) | |
| | 0 | 21,001 (92.3) |
| | 1-2 | 1,661 (7.3) |
| | +3 | 89 (0.4) |
| | Education level, n (%) | |
| | No/Unknown education | 149 (0.7) |
| | Primary | 14,474 (63.6) |
| | Secondary | 7,541 (33.1) |
| | Tertiary | 587 (2.6) |
| Young adults and adults (25-64 years) | Number of persons, n | 413,547 (100.0) |
| | Male, n (%) | 156,959 (38.0) |
| | Age, median (Q1;Q3) | 48.0 (37.8;56.2) |
| | CCI, n (%) | |
| | 0 | 335,414 (81.1) |
| | 1-2 | 66,812 (16.2) |
| | +3 | 11,321 (2.7) |
| | Education level, n (%) | |
| | No/Unknown education | 9,915 (2.4) |
| | Primary | 108,185 (26.2) |
| | Secondary | 185,145 (44.8) |
| | Tertiary | 110,302 (26.7) |
| Older people (65+ years) | Number of persons, n | 173,810 (100.0) |
| | Male, n (%) | 61,791 (35.6) |
| | Age, median (Q1;Q3) | 74.7 (69.6;80.9) |
| | CCI, n (%) | |
| | 0 | 80,304 (46.2) |
| | 1-2 | 65,628 (37.8) |
| | +3 | 27,878 (16.0) |
| | Education level, n (%) | |
| | No/Unknown education | 8,303 (4.8) |
| | Primary | 57,711 (33.2) |
| | Secondary | 76,048 (43.8) |
| | Tertiary | 31,748 (18.3) |

**Supplementary Table 86**
**Depression - Prevalent cohort 2021**
**Patient characteristics - relatives - pooled**

| *Age strata of index patients* | *Closest relative type* | *Characteristic of family relatives by agestrata of index patients* | *Closest family relatives of brain disorder patients* | *Comparison relative* |
|---|---|---|---|---|
| Children and young people (0-24 years) | Fathers | Number of persons, n | 18,117 (100.0) | 18,117 (100.0) |
| | | Male, n (%) | 18,117 (100.0) | 18,117 (100.0) |
| | | Age, median (Q1;Q3) | 53.7 (49.9;57.3) | 53.7 (50.3;56.9) |
| | | CCI, n (%) | | |
| | | 0 | 15,508 (85.6) | 15,760 (87.0) |
| | | 1-2 | 2,308 (12.7) | 2,064 (11.4) |
| | | +3 | 301 (1.7) | 293 (1.6) |
| | | Education level, n (%) | | |
| | | No/Unknown education | 163 (0.9) | 163 (0.9) |
| | | Primary | 3,320 (18.3) | 3,320 (18.3) |
| | | Secondary | 9,775 (54.0) | 9,775 (54.0) |
| | | Tertiary | 4,859 (26.8) | 4,859 (26.8) |
| | | Relation to patient, n (%) | | |
| | | Father | 18,117 (100.0) | 18,117 (100.0) |
| Children and young people (0-24 years) | Mothers | Number of persons, n | 15,674 (100.0) | 15,674 (100.0) |
| | | Male, n (%) | 0 (0.0) | 0 (0.0) |
| | | Age, median (Q1;Q3) | 51.7 (48.1;55.2) | 51.6 (48.4;54.7) |
| | | CCI, n (%) | | |
| | | 0 | 13,397 (85.5) | 13,672 (87.2) |
| | | 1-2 | 2,064 (13.2) | 1,812 (11.6) |
| | | +3 | 213 (1.4) | 190 (1.2) |
| | | Education level, n (%) | | |
| | | No/Unknown education | 73 (0.5) | 73 (0.5) |
| | | Primary | 2,117 (13.5) | 2,117 (13.5) |
| | | Secondary | 7,304 (46.6) | 7,304 (46.6) |
| | | Tertiary | 6,180 (39.4) | 6,180 (39.4) |
| | | Relation to patient, n (%) | | |
| | | Mother | 15,674 (100.0) | 15,674 (100.0) |
| Young adults and adults (25-64 years) | Pooled | Number of persons, n | 413,547 (100.0) | 413,547 (100.0) |
| | | Male, n (%) | 236,431 (57.2) | 236,431 (57.2) |
| | | Age, median (Q1;Q3) | 46.0 (29.0;59.3) | 46.1 (28.9;59.3) |
| | | CCI, n (%) | | |
| | | 0 | 352,270 (85.2) | 357,768 (86.5) |
| | | 1-2 | 52,123 (12.6) | 47,586 (11.5) |
| | | +3 | 9,154 (2.2) | 8,193 (2.0) |
| | | Education level, n (%) | | |
| | | No/Unknown education | 32,860 (7.9) | 32,860 (7.9) |
| | | Primary | 111,195 (26.9) | 111,195 (26.9) |
| | | Secondary | 179,585 (43.4) | 179,585 (43.4) |
| | | Tertiary | 89,907 (21.7) | 89,907 (21.7) |
| | | Relation to patient, n (%) | | |
| | | Mother | 48,788 (11.8) | 48,788 (11.8) |
| | | Father | 22,974 (5.6) | 22,974 (5.6) |
| | | Partner | 190,095 (46.0) | 190,095 (46.0) |
| | | Child | 112,999 (27.3) | 112,999 (27.3) |

**Supplementary Table 86**
**Depression - Prevalent cohort 2021**
**Patient characteristics - relatives - pooled**

| *Age strata of index patients* | *Closest relative type* | *Characteristic of family relatives by agestrata of index patients* | *Closest family relatives of brain disorder patients* | *Comparison relative* |
|---|---|---|---|---|
| | | Sibling | 38,691 (9.4) | 38,691 (9.4) |
| Older people (65+ years) | Pooled | Number of persons, n | 173,810 (100.0) | 173,810 (100.0) |
| | | Male, n (%) | 96,705 (55.6) | 96,705 (55.6) |
| | | Age, median (Q1;Q3) | 61.5 (50.7;71.9) | 61.4 (50.6;71.9) |
| | | CCI, n (%) | | |
| | | 0 | 130,801 (75.3) | 132,866 (76.4) |
| | | 1-2 | 34,265 (19.7) | 32,808 (18.9) |
| | | +3 | 8,744 (5.0) | 8,136 (4.7) |
| | | Education level, n (%) | | |
| | | No/Unknown education | 4,585 (2.6) | 4,585 (2.6) |
| | | Primary | 32,882 (18.9) | 32,882 (18.9) |
| | | Secondary | 86,523 (49.8) | 86,523 (49.8) |
| | | Tertiary | 49,820 (28.7) | 49,820 (28.7) |
| | | Relation to patient, n (%) | | |
| | | Partner | 74,964 (43.1) | 74,964 (43.1) |
| | | Child | 95,196 (54.8) | 95,196 (54.8) |
| | | Sibling | 3,650 (2.1) | 3,650 (2.1) |

**Supplementary Table 87**
**Depression - Prevalent cohort 2021**
**Patient characteristics - relatives - not pooled**

| *Age strata of index patients* | *Closest relative type* | *Characteristic of family relatives by agestrata of index patients* | *Closest family relatives of brain disorder patients* | *Comparison relative* |
|---|---|---|---|---|
| Children and young people (0-24 years) | Fathers | Number of persons, n | 18,117 (100.0) | 18,117 (100.0) |
| | | Male, n (%) | 18,117 (100.0) | 18,117 (100.0) |
| | | Age, median (Q1;Q3) | 53.7 (49.9;57.3) | 53.7 (50.3;56.9) |
| | | CCI, n (%) | | |
| | | 0 | 15,508 (85.6) | 15,760 (87.0) |
| | | 1-2 | 2,308 (12.7) | 2,064 (11.4) |
| | | +3 | 301 (1.7) | 293 (1.6) |
| | | Education level, n (%) | | |
| | | No/Unknown education | 163 (0.9) | 163 (0.9) |
| | | Primary | 3,320 (18.3) | 3,320 (18.3) |
| | | Secondary | 9,775 (54.0) | 9,775 (54.0) |
| | | Tertiary | 4,859 (26.8) | 4,859 (26.8) |
| | | Relation to patient, n (%) | | |
| | | Father | 18,117 (100.0) | 18,117 (100.0) |
| Children and young people (0-24 years) | Mothers | Number of persons, n | 15,674 (100.0) | 15,674 (100.0) |
| | | Male, n (%) | 0 (0.0) | 0 (0.0) |
| | | Age, median (Q1;Q3) | 51.7 (48.1;55.2) | 51.6 (48.4;54.7) |
| | | CCI, n (%) | | |
| | | 0 | 13,397 (85.5) | 13,672 (87.2) |
| | | 1-2 | 2,064 (13.2) | 1,812 (11.6) |
| | | +3 | 213 (1.4) | 190 (1.2) |
| | | Education level, n (%) | | |
| | | No/Unknown education | 73 (0.5) | 73 (0.5) |
| | | Primary | 2,117 (13.5) | 2,117 (13.5) |
| | | Secondary | 7,304 (46.6) | 7,304 (46.6) |
| | | Tertiary | 6,180 (39.4) | 6,180 (39.4) |
| | | Relation to patient, n (%) | | |
| | | Mother | 15,674 (100.0) | 15,674 (100.0) |
| Young adults and adults (25-64 years) | Children | Number of persons, n | 112,999 (100.0) | 112,999 (100.0) |
| | | Male, n (%) | 61,560 (54.5) | 61,560 (54.5) |
| | | Age, median (Q1;Q3) | 20.3 (13.6;28.3) | 19.9 (13.5;27.8) |
| | | CCI, n (%) | | |
| | | 0 | 105,862 (93.7) | 107,267 (94.9) |
| | | 1-2 | 6,904 (6.1) | 5,533 (4.9) |
| | | +3 | 233 (0.2) | 199 (0.2) |
| | | Education level, n (%) | | |
| | | No/Unknown education | 26,943 (23.8) | 26,943 (23.8) |
| | | Primary | 46,164 (40.9) | 46,164 (40.9) |
| | | Secondary | 26,844 (23.8) | 26,844 (23.8) |
| | | Tertiary | 13,048 (11.5) | 13,048 (11.5) |
| | | Relation to patient, n (%) | | |
| | | Child | 112,999 (100.0) | 112,999 (100.0) |
| Young adults and adults (25-64 years) | Parents | Number of persons, n | 71,762 (100.0) | 71,762 (100.0) |
| | | Male, n (%) | 22,974 (32.0) | 22,974 (32.0) |
| | | Age, median (Q1;Q3) | 67.9 (60.6;75.4) | 68.0 (60.8;75.5) |

### Supplementary Table 87
### Depression - Prevalent cohort 2021
### Patient characteristics - relatives - not pooled

| *Age strata of index patients* | *Closest relative type* | *Characteristic of family relatives by agestrata of index patients* | *Closest family relatives of brain disorder patients* | *Comparison relative* |
|---|---|---|---|---|
| | | CCI, n (%) | | |
| | | 0 | 48,495 (67.6) | 49,950 (69.6) |
| | | 1-2 | 18,300 (25.5) | 17,196 (24.0) |
| | | +3 | 4,967 (6.9) | 4,616 (6.4) |
| | | Education level, n (%) | | |
| | | No/Unknown education | 1,128 (1.6) | 1,128 (1.6) |
| | | Primary | 21,406 (29.8) | 21,406 (29.8) |
| | | Secondary | 35,805 (49.9) | 35,805 (49.9) |
| | | Tertiary | 13,423 (18.7) | 13,423 (18.7) |
| | | Relation to patient, n (%) | | |
| | | Mother | 48,788 (68.0) | 48,788 (68.0) |
| | | Father | 22,974 (32.0) | 22,974 (32.0) |
| Young adults and adults (25-64 years) | Partners | Number of persons, n | 190,095 (100.0) | 190,095 (100.0) |
| | | Male, n (%) | 130,414 (68.6) | 130,414 (68.6) |
| | | Age, median (Q1;Q3) | 49.1 (38.9;57.3) | 49.2 (39.0;57.3) |
| | | CCI, n (%) | | |
| | | 0 | 165,002 (86.8) | 167,293 (88.0) |
| | | 1-2 | 21,953 (11.5) | 20,133 (10.6) |
| | | +3 | 3,140 (1.7) | 2,669 (1.4) |
| | | Education level, n (%) | | |
| | | No/Unknown education | 4,683 (2.5) | 4,683 (2.5) |
| | | Primary | 34,807 (18.3) | 34,807 (18.3) |
| | | Secondary | 96,162 (50.6) | 96,162 (50.6) |
| | | Tertiary | 54,443 (28.6) | 54,443 (28.6) |
| | | Relation to patient, n (%) | | |
| | | Partner | 190,095 (100.0) | 190,095 (100.0) |
| Young adults and adults (25-64 years) | Siblings | Number of persons, n | 38,691 (100.0) | 38,691 (100.0) |
| | | Male, n (%) | 21,483 (55.5) | 21,483 (55.5) |
| | | Age, median (Q1;Q3) | 52.4 (35.2;59.4) | 52.5 (35.1;59.4) |
| | | CCI, n (%) | | |
| | | 0 | 32,911 (85.1) | 33,258 (86.0) |
| | | 1-2 | 4,966 (12.8) | 4,724 (12.2) |
| | | +3 | 814 (2.1) | 709 (1.8) |
| | | Education level, n (%) | | |
| | | No/Unknown education | 106 (0.3) | 106 (0.3) |
| | | Primary | 8,818 (22.8) | 8,818 (22.8) |
| | | Secondary | 20,774 (53.7) | 20,774 (53.7) |
| | | Tertiary | 8,993 (23.2) | 8,993 (23.2) |
| | | Relation to patient, n (%) | | |
| | | Sibling | 38,691 (100.0) | 38,691 (100.0) |
| Older people (65+ years) | Children | Number of persons, n | 95,196 (100.0) | 95,196 (100.0) |
| | | Male, n (%) | 51,239 (53.8) | 51,239 (53.8) |
| | | Age, median (Q1;Q3) | 51.9 (45.2;58.0) | 51.8 (45.1;57.7) |
| | | CCI, n (%) | | |
| | | 0 | 82,118 (86.3) | 83,030 (87.2) |

**Supplementary Table 87**
**Depression - Prevalent cohort 2021**
**Patient characteristics - relatives - not pooled**

| *Age strata of index patients* | *Closest relative type* | *Characteristic of family relatives by agestrata of index patients* | *Closest family relatives of brain disorder patients* | *Comparison relative* |
|---|---|---|---|---|
| | | 1-2 | 11,519 (12.1) | 10,752 (11.3) |
| | | +3 | 1,559 (1.6) | 1,414 (1.5) |
| | | Education level, n (%) | | |
| | | No/Unknown education | 247 (0.3) | 247 (0.3) |
| | | Primary | 14,093 (14.8) | 14,093 (14.8) |
| | | Secondary | 47,297 (49.7) | 47,297 (49.7) |
| | | Tertiary | 33,559 (35.3) | 33,559 (35.3) |
| | | Relation to patient, n (%) | | |
| | | Child | 95,196 (100.0) | 95,196 (100.0) |
| Older people (65+ years) | Partners | Number of persons, n | 74,964 (100.0) | 74,964 (100.0) |
| | | Male, n (%) | 43,463 (58.0) | 43,463 (58.0) |
| | | Age, median (Q1;Q3) | 73.3 (68.6;77.8) | 73.3 (68.8;77.7) |
| | | CCI, n (%) | | |
| | | 0 | 45,894 (61.2) | 46,975 (62.7) |
| | | 1-2 | 22,023 (29.4) | 21,397 (28.5) |
| | | +3 | 7,047 (9.4) | 6,592 (8.8) |
| | | Education level, n (%) | | |
| | | No/Unknown education | 4,321 (5.8) | 4,321 (5.8) |
| | | Primary | 18,223 (24.3) | 18,223 (24.3) |
| | | Secondary | 36,946 (49.3) | 36,946 (49.3) |
| | | Tertiary | 15,474 (20.6) | 15,474 (20.6) |
| | | Relation to patient, n (%) | | |
| | | Partner | 74,964 (100.0) | 74,964 (100.0) |
| Older people (65+ years) | Siblings | Number of persons, n | 3,650 (100.0) | 3,650 (100.0) |
| | | Male, n (%) | 2,003 (54.9) | 2,003 (54.9) |
| | | Age, median (Q1;Q3) | 62.6 (59.6;65.2) | 62.6 (59.9;65.2) |
| | | CCI, n (%) | | |
| | | 0 | 2,789 (76.4) | 2,861 (78.4) |
| | | 1-2 | 723 (19.8) | 659 (18.1) |
| | | +3 | 138 (3.8) | 130 (3.6) |
| | | Education level, n (%) | | |
| | | No/Unknown education | 17 (0.5) | 17 (0.5) |
| | | Primary | 566 (15.5) | 566 (15.5) |
| | | Secondary | 2,280 (62.5) | 2,280 (62.5) |
| | | Tertiary | 787 (21.6) | 787 (21.6) |
| | | Relation to patient, n (%) | | |
| | | Sibling | 3,650 (100.0) | 3,650 (100.0) |

#### Supplementary Table 88
#### Depression - Prevalent cohort 2021
#### Cost analysis - pooled

| *Age strata of index patients* | *Closest relative type* | *Number of index patients included in cost component calculations* | *Cost component* | *Actual costs* | | *Attributable costs* | |
|---|---|---|---|---|---|---|---|
| | | | | *Total (million EUR)* | *Per person (EUR)* | *Total (million EUR)* | *Per person (EUR)* |
| Children and young people (0-24 years) | Fathers | 18,117 | Primary sector | 4.3 | 237 | 0.2 | 12 |
| | | 18,117 | Hospital care | 28.3 | 1,568 | 2.5 | 137 |
| | | 18,117 | Filled prescriptions | 3.8 | 208 | 0.6 | 31 |
| | | 17,105 | Lost productivity (income loss) | . | . | 132.7 | 7,760 |
| Children and young people (0-24 years) | Mothers | 15,674 | Primary sector | 5.5 | 353 | 0.7 | 47 |
| | | 15,674 | Hospital care | 26.0 | 1,664 | 4.4 | 280 |
| | | 15,674 | Filled prescriptions | 3.2 | 205 | 0.6 | 42 |
| | | 15,487 | Lost productivity (income loss) | . | . | 37.9 | 2,447 |
| Young adults and adults (25-64 years) | Pooled | 413,547 | Primary sector | 115.6 | 281 | 9.0 | 22 |
| | | 413,547 | Hospital care | 740.4 | 1,797 | 73.7 | 179 |
| | | 413,547 | Filled prescriptions | 92.2 | 224 | 13.0 | 31 |
| | | 287,037 | Lost productivity (income loss) | . | . | 1,401.6 | 4,883 |
| Older people (65+ years) | Pooled | 173,810 | Primary sector | 65.3 | 378 | 3.8 | 22 |
| | | 173,810 | Hospital care | 458.3 | 2,654 | 28.1 | 163 |
| | | 173,810 | Filled prescriptions | 59.2 | 343 | 5.8 | 33 |
| | | 91,956 | Lost productivity (income loss) | . | . | 249.7 | 2,716 |

#### Supplementary Table 89
#### Depression - Prevalent cohort 2021
#### Cost analysis - not pooled

| *Age strata of index patients* | *Closest relative type* | *Number of index patients included in cost component calculations* | *Cost component* | *Actual costs* | | *Attributable costs* | |
|---|---|---|---|---|---|---|---|
| | | | | *Total (million EUR)* | *Per person (EUR)* | *Total (million EUR)* | *Per person (EUR)* |
| Children and young people (0-24 years) | Fathers | 18,117 | Primary sector | 4.3 | 237 | 0.2 | 12 |
| | | 18,117 | Hospital care | 28.3 | 1,568 | 2.5 | 137 |
| | | 18,117 | Filled prescriptions | 3.8 | 208 | 0.6 | 31 |
| | | 17,105 | Lost productivity (income loss) | . | . | 132.7 | 7,760 |
| Children and young people (0-24 years) | Mothers | 15,674 | Primary sector | 5.5 | 353 | 0.7 | 47 |
| | | 15,674 | Hospital care | 26.0 | 1,664 | 4.4 | 280 |
| | | 15,674 | Filled prescriptions | 3.2 | 205 | 0.6 | 42 |
| | | 15,487 | Lost productivity (income loss) | . | . | 37.9 | 2,447 |
| Young adults and adults (25-64 years) | Children | 112,999 | Primary sector | 23.9 | 212 | 3.4 | 30 |
| | | 112,999 | Hospital care | 130.0 | 1,154 | 30.6 | 271 |
| | | 112,999 | Filled prescriptions | 11.9 | 106 | 1.8 | 16 |
| | | 60,549 | Lost productivity (income loss) | . | . | 130.3 | 2,152 |
| Young adults and adults (25-64 years) | Parents | 71,762 | Primary sector | 30.8 | 434 | 1.4 | 19 |
| | | 71,762 | Hospital care | 236.0 | 3,323 | 10.5 | 148 |
| | | 71,762 | Filled prescriptions | 33.5 | 471 | 4.2 | 60 |
| | | 22,978 | Lost productivity (income loss) | . | . | 93.4 | 4,066 |
| Young adults and adults (25-64 years) | Partners | 190,095 | Primary sector | 49.8 | 262 | 3.6 | 19 |
| | | 190,095 | Hospital care | 300.5 | 1,584 | 28.9 | 152 |
| | | 190,095 | Filled prescriptions | 38.2 | 201 | 6.3 | 33 |
| | | 170,485 | Lost productivity (income loss) | . | . | 1,109.2 | 6,506 |
| Young adults and adults (25-64 years) | Siblings | 38,691 | Primary sector | 11.1 | 287 | 0.6 | 16 |
| | | 38,691 | Hospital care | 73.8 | 1,912 | 3.8 | 98 |
| | | 38,691 | Filled prescriptions | 8.7 | 225 | 0.7 | 17 |
| | | 33,025 | Lost productivity (income loss) | . | . | 68.8 | 2,083 |
| Older people (65+ years) | Children | 95,196 | Primary sector | 27.5 | 289 | 2.2 | 23 |
| | | 95,196 | Hospital care | 154.8 | 1,629 | 10.9 | 115 |
| | | 95,196 | Filled prescriptions | 19.7 | 207 | 2.2 | 23 |
| | | 86,359 | Lost productivity (income loss) | . | . | 245.7 | 2,846 |
| Older people (65+ years) | Partners | 74,964 | Primary sector | 36.5 | 493 | 1.5 | 21 |
| | | 74,964 | Hospital care | 294.2 | 3,975 | 16.7 | 225 |
| | | 74,964 | Filled prescriptions | 38.3 | 518 | 3.5 | 47 |
| | | 3,635 | Lost productivity (income loss) | . | . | -3.3 | -921 |
| Older people (65+ years) | Siblings | 3,650 | Primary sector | 1.3 | 355 | 0.1 | 23 |
| | | 3,650 | Hospital care | 9.2 | 2,536 | 0.6 | 159 |
| | | 3,650 | Filled prescriptions | 1.1 | 315 | 0.1 | 23 |
| | | 1,962 | Lost productivity (income loss) | . | . | 7.3 | 3,742 |

**Supplementary Table 90**
**Developmental and behavioural disorders - Incident cohort 2016-2021**
**Identification of populations of relatives**

| ***Agegroup of index patients in matched population*** | | ***Closest relative type identified*** | | ***Available control with same type of closest relative*** |
|---|---|---|---|---|
| Children and young people (0-24 years) | n = 60,667 | Fathers | n = 49,390 (81%) | 96% |
| | | Mothers | n = 51,687 (85%) | 96% |
| | | No available closest relative | n = 3,599 (6%) | n/a |
| Young adults and adults (25-64 years) | n = 24,563 | Children | n = 2,844 (12%) | 94% |
| | | Parents | n = 7,434 (30%) | 92% |
| | | Partners | n = 9,691 (39%) | 95% |
| | | Siblings | n = 3,747 (15%) | 36% |
| | | Pooled | n = 23,716 (97%) | 85% |
| | | No available closest relative | n = 847 (3%) | n/a |
| Older people (65+ years) | n = 924 | Children | n = 374 (40%) | 93% |
| | | Partners | n = 365 (40%) | 95% |
| | | Siblings | n = 33 (4%) | 48% |
| | | Pooled | n = 772 (84%) | 92% |
| | | No available closest relative | n = 152 (16%) | n/a |

**Supplementary Table 91**
**Developmental and behavioural disorders - Incident cohort 2016-2021**
**Patient characteristics - cases**

| *Age strata of index patients* | *Characteristic* | |
|---|---|---|
| Children and young people (0-24 years) | Number of persons, n | 55,312 (100.0) |
| | Male, n (%) | 32,242 (58.3) |
| | Age, median (Q1;Q3) | 13.0 (8.7;17.3) |
| | CCI, n (%) | |
| | 0 | 50,756 (91.8) |
| | 1-2 | 4,467 (8.1) |
| | +3 | 89 (0.2) |
| | Education level, n (%) | |
| | No/Unknown education | 28,329 (51.2) |
| | Primary | 23,701 (42.8) |
| | Secondary | 3,108 (5.6) |
| | Tertiary | 174 (0.3) |
| Young adults and adults (25-64 years) | Number of persons, n | 20,266 (100.0) |
| | Male, n (%) | 11,012 (54.3) |
| | Age, median (Q1;Q3) | 36.4 (29.9;44.7) |
| | CCI, n (%) | |
| | 0 | 17,834 (88.0) |
| | 1-2 | 2,181 (10.8) |
| | +3 | 251 (1.2) |
| | Education level, n (%) | |
| | No/Unknown education | 411 (2.0) |
| | Primary | 5,964 (29.4) |
| | Secondary | 8,792 (43.4) |
| | Tertiary | 5,099 (25.2) |
| Older people (65+ years) | Number of persons, n | 711 (100.0) |
| | Male, n (%) | 364 (51.2) |
| | Age, median (Q1;Q3) | 73.7 (69.1;80.0) |
| | CCI, n (%) | |
| | 0 | 228 (32.1) |
| | 1-2 | 293 (41.2) |
| | +3 | 190 (26.7) |
| | Education level, n (%) | |
| | No/Unknown education | 48 (6.8) |
| | Primary | 207 (29.1) |
| | Secondary | 307 (43.2) |
| | Tertiary | 149 (21.0) |

**Supplementary Table 92**
**Developmental and behavioural disorders - Incident cohort 2016-2021**
**Patient characteristics - relatives - pooled**

| *Age strata of index patients* | *Closest relative type* | *Characteristic of family relatives by agestrata of index patients* | *Closest family relatives of brain disorder patients* | *Comparison relative* |
|---|---|---|---|---|
| Children and young people (0-24 years) | Fathers | Number of persons, n | 47,830 (100.0) | 47,830 (100.0) |
| | | Male, n (%) | 47,830 (100.0) | 47,830 (100.0) |
| | | Age, median (Q1;Q3) | 45.7 (40.4;51.1) | 45.8 (40.7;50.9) |
| | | CCI, n (%) | | |
| | | 0 | 42,738 (89.4) | 43,299 (90.5) |
| | | 1-2 | 4,537 (9.5) | 4,041 (8.4) |
| | | +3 | 555 (1.2) | 490 (1.0) |
| | | Education level, n (%) | | |
| | | No/Unknown education | 876 (1.8) | 876 (1.8) |
| | | Primary | 10,502 (22.0) | 10,502 (22.0) |
| | | Secondary | 23,841 (49.8) | 23,841 (49.8) |
| | | Tertiary | 12,611 (26.4) | 12,611 (26.4) |
| | | Relation to patient, n (%) | | |
| | | Father | 47,830 (100.0) | 47,830 (100.0) |
| Children and young people (0-24 years) | Mothers | Number of persons, n | 50,091 (100.0) | 50,091 (100.0) |
| | | Male, n (%) | 0 (0.0) | 0 (0.0) |
| | | Age, median (Q1;Q3) | 43.3 (37.9;48.5) | 43.4 (38.3;48.3) |
| | | CCI, n (%) | | |
| | | 0 | 43,775 (87.4) | 45,014 (89.9) |
| | | 1-2 | 5,811 (11.6) | 4,644 (9.3) |
| | | +3 | 505 (1.0) | 433 (0.9) |
| | | Education level, n (%) | | |
| | | No/Unknown education | 877 (1.8) | 877 (1.8) |
| | | Primary | 9,091 (18.1) | 9,091 (18.1) |
| | | Secondary | 21,090 (42.1) | 21,090 (42.1) |
| | | Tertiary | 19,033 (38.0) | 19,033 (38.0) |
| | | Relation to patient, n (%) | | |
| | | Mother | 50,091 (100.0) | 50,091 (100.0) |
| Young adults and adults (25-64 years) | Pooled | Number of persons, n | 20,266 (100.0) | 20,266 (100.0) |
| | | Male, n (%) | 8,182 (40.4) | 8,182 (40.4) |
| | | Age, median (Q1;Q3) | 45.0 (30.9;57.7) | 45.1 (31.0;57.8) |
| | | CCI, n (%) | | |
| | | 0 | 17,105 (84.4) | 17,406 (85.9) |
| | | 1-2 | 2,654 (13.1) | 2,447 (12.1) |
| | | +3 | 507 (2.5) | 413 (2.0) |
| | | Education level, n (%) | | |
| | | No/Unknown education | 1,844 (9.1) | 1,844 (9.1) |
| | | Primary | 4,547 (22.4) | 4,547 (22.4) |
| | | Secondary | 8,568 (42.3) | 8,568 (42.3) |
| | | Tertiary | 5,307 (26.2) | 5,307 (26.2) |
| | | Relation to patient, n (%) | | |
| | | Mother | 5,321 (26.3) | 5,321 (26.3) |
| | | Father | 1,591 (7.9) | 1,591 (7.9) |
| | | Partner | 9,279 (45.8) | 9,279 (45.8) |
| | | Child | 2,699 (13.3) | 2,699 (13.3) |

#### Supplementary Table 92
#### Developmental and behavioural disorders - Incident cohort 2016-2021
#### Patient characteristics - relatives - pooled

| *Age strata of index patients* | *Closest relative type* | *Characteristic of family relatives by agestrata of index patients* | *Closest family relatives of brain disorder patients* | *Comparison relative* |
|---|---|---|---|---|
| | | Sibling | 1,376 (6.8) | 1,376 (6.8) |
| Older people (65+ years) | Pooled | Number of persons, n | 711 (100.0) | 711 (100.0) |
| | | Male, n (%) | 304 (42.8) | 304 (42.8) |
| | | Age, median (Q1;Q3) | 62.1 (50.1;71.9) | 62.1 (50.5;72.4) |
| | | CCI, n (%) | | |
| | | 0 | 508 (71.4) | 531 (74.7) |
| | | 1-2 | 167 (23.5) | 140 (19.7) |
| | | +3 | 36 (5.1) | 40 (5.6) |
| | | Education level, n (%) | | |
| | | No/Unknown education | 27 (3.8) | 27 (3.8) |
| | | Primary | 144 (20.3) | 144 (20.3) |
| | | Secondary | 323 (45.4) | 323 (45.4) |
| | | Tertiary | 217 (30.5) | 217 (30.5) |
| | | Relation to patient, n (%) | | |
| | | Partner | 347 (48.8) | 347 (48.8) |
| | | Child | 348 (48.9) | 348 (48.9) |
| | | Sibling | 16 (2.3) | 16 (2.3) |

**Supplementary Table 93**
**Developmental and behavioural disorders - Incident cohort 2016-2021**
**Patient characteristics - relatives - not pooled**

| *Age strata of index patients* | *Closest relative type* | *Characteristic of family relatives by agestrata of index patients* | *Closest family relatives of brain disorder patients* | *Comparison relative* |
|---|---|---|---|---|
| Children and young people (0-24 years) | Fathers | Number of persons, n | 47,830 (100.0) | 47,830 (100.0) |
| | | Male, n (%) | 47,830 (100.0) | 47,830 (100.0) |
| | | Age, median (Q1;Q3) | 45.7 (40.4;51.1) | 45.8 (40.7;50.9) |
| | | CCI, n (%) | | |
| | | 0 | 42,738 (89.4) | 43,299 (90.5) |
| | | 1-2 | 4,537 (9.5) | 4,041 (8.4) |
| | | +3 | 555 (1.2) | 490 (1.0) |
| | | Education level, n (%) | | |
| | | No/Unknown education | 876 (1.8) | 876 (1.8) |
| | | Primary | 10,502 (22.0) | 10,502 (22.0) |
| | | Secondary | 23,841 (49.8) | 23,841 (49.8) |
| | | Tertiary | 12,611 (26.4) | 12,611 (26.4) |
| | | Relation to patient, n (%) | | |
| | | Father | 47,830 (100.0) | 47,830 (100.0) |
| Children and young people (0-24 years) | Mothers | Number of persons, n | 50,091 (100.0) | 50,091 (100.0) |
| | | Male, n (%) | 0 (0.0) | 0 (0.0) |
| | | Age, median (Q1;Q3) | 43.3 (37.9;48.5) | 43.4 (38.3;48.3) |
| | | CCI, n (%) | | |
| | | 0 | 43,775 (87.4) | 45,014 (89.9) |
| | | 1-2 | 5,811 (11.6) | 4,644 (9.3) |
| | | +3 | 505 (1.0) | 433 (0.9) |
| | | Education level, n (%) | | |
| | | No/Unknown education | 877 (1.8) | 877 (1.8) |
| | | Primary | 9,091 (18.1) | 9,091 (18.1) |
| | | Secondary | 21,090 (42.1) | 21,090 (42.1) |
| | | Tertiary | 19,033 (38.0) | 19,033 (38.0) |
| | | Relation to patient, n (%) | | |
| | | Mother | 50,091 (100.0) | 50,091 (100.0) |
| Young adults and adults (25-64 years) | Children | Number of persons, n | 2,699 (100.0) | 2,699 (100.0) |
| | | Male, n (%) | 1,316 (48.8) | 1,316 (48.8) |
| | | Age, median (Q1;Q3) | 11.6 (6.8;18.4) | 11.2 (6.6;18.3) |
| | | CCI, n (%) | | |
| | | 0 | 2,522 (93.4) | 2,561 (94.9) |
| | | 1-2 | 172 (6.4) | 133 (4.9) |
| | | +3 | 5 (0.2) | 5 (0.2) |
| | | Education level, n (%) | | |
| | | No/Unknown education | 1,530 (56.7) | 1,530 (56.7) |
| | | Primary | 811 (30.0) | 811 (30.0) |
| | | Secondary | 278 (10.3) | 278 (10.3) |
| | | Tertiary | 80 (3.0) | 80 (3.0) |
| | | Relation to patient, n (%) | | |
| | | Child | 2,699 (100.0) | 2,699 (100.0) |
| Young adults and adults (25-64 years) | Parents | Number of persons, n | 6,912 (100.0) | 6,912 (100.0) |
| | | Male, n (%) | 1,591 (23.0) | 1,591 (23.0) |
| | | Age, median (Q1;Q3) | 60.5 (55.6;67.1) | 60.6 (56.0;67.0) |

**Supplementary Table 93**
**Developmental and behavioural disorders - Incident cohort 2016-2021**
**Patient characteristics - relatives - not pooled**

| *Age strata of index patients* | *Closest relative type* | *Characteristic of family relatives by agestrata of index patients* | *Closest family relatives of brain disorder patients* | *Comparison relative* |
|---|---|---|---|---|
| | | CCI, n (%) | | |
| | | 0 | 4,946 (71.6) | 5,148 (74.5) |
| | | 1-2 | 1,586 (22.9) | 1,449 (21.0) |
| | | +3 | 380 (5.5) | 315 (4.6) |
| | | Education level, n (%) | | |
| | | No/Unknown education | 127 (1.8) | 127 (1.8) |
| | | Primary | 1,780 (25.8) | 1,780 (25.8) |
| | | Secondary | 3,345 (48.4) | 3,345 (48.4) |
| | | Tertiary | 1,660 (24.0) | 1,660 (24.0) |
| | | Relation to patient, n (%) | | |
| | | Mother | 5,321 (77.0) | 5,321 (77.0) |
| | | Father | 1,591 (23.0) | 1,591 (23.0) |
| Young adults and adults (25-64 years) | Partners | Number of persons, n | 9,279 (100.0) | 9,279 (100.0) |
| | | Male, n (%) | 4,416 (47.6) | 4,416 (47.6) |
| | | Age, median (Q1;Q3) | 38.6 (31.6;46.4) | 38.6 (31.7;46.5) |
| | | CCI, n (%) | | |
| | | 0 | 8,432 (90.9) | 8,495 (91.6) |
| | | 1-2 | 759 (8.2) | 711 (7.7) |
| | | +3 | 88 (0.9) | 73 (0.8) |
| | | Education level, n (%) | | |
| | | No/Unknown education | 184 (2.0) | 184 (2.0) |
| | | Primary | 1,600 (17.2) | 1,600 (17.2) |
| | | Secondary | 4,241 (45.7) | 4,241 (45.7) |
| | | Tertiary | 3,254 (35.1) | 3,254 (35.1) |
| | | Relation to patient, n (%) | | |
| | | Partner | 9,279 (100.0) | 9,279 (100.0) |
| Young adults and adults (25-64 years) | Siblings | Number of persons, n | 1,376 (100.0) | 1,376 (100.0) |
| | | Male, n (%) | 859 (62.4) | 859 (62.4) |
| | | Age, median (Q1;Q3) | 38.8 (29.2;52.4) | 38.6 (29.3;52.0) |
| | | CCI, n (%) | | |
| | | 0 | 1,205 (87.6) | 1,202 (87.4) |
| | | 1-2 | 137 (10.0) | 154 (11.2) |
| | | +3 | 34 (2.5) | 20 (1.5) |
| | | Education level, n (%) | | |
| | | No/Unknown education | * | * |
| | | Primary | 356 (25.9) | 356 (25.9) |
| | | Secondary | 704 (51.2) | 704 (51.2) |
| | | Tertiary | * | * |
| | | Relation to patient, n (%) | | |
| | | Sibling | 1,376 (100.0) | 1,376 (100.0) |
| Older people (65+ years) | Children | Number of persons, n | 348 (100.0) | 348 (100.0) |
| | | Male, n (%) | 168 (48.3) | 168 (48.3) |
| | | Age, median (Q1;Q3) | 50.0 (43.6;56.9) | 50.1 (42.9;56.3) |
| | | CCI, n (%) | | |
| | | 0 | 290 (83.3) | 301 (86.5) |

**Supplementary Table 93**
**Developmental and behavioural disorders - Incident cohort 2016-2021**
**Patient characteristics - relatives - not pooled**

| *Age strata of index patients* | *Closest relative type* | *Characteristic of family relatives by agestrata of index patients* | *Closest family relatives of brain disorder patients* | *Comparison relative* |
|---|---|---|---|---|
| | | 1-2 | 53 (15.2) | 38 (10.9) |
| | | +3 | 5 (1.4) | 9 (2.6) |
| | | Education level, n (%) | | |
| | | No/Unknown education | * | * |
| | | Primary | * | * |
| | | Secondary | 172 (49.4) | 172 (49.4) |
| | | Tertiary | 118 (33.9) | 118 (33.9) |
| | | Relation to patient, n (%) | | |
| | | Child | 348 (100.0) | 348 (100.0) |
| Older people (65+ years) | Partners | Number of persons, n | 347 (100.0) | 347 (100.0) |
| | | Male, n (%) | 130 (37.5) | 130 (37.5) |
| | | Age, median (Q1;Q3) | 72.1 (67.0;77.6) | 72.6 (67.2;77.4) |
| | | CCI, n (%) | | |
| | | 0 | 204 (58.8) | 216 (62.2) |
| | | 1-2 | 112 (32.3) | 101 (29.1) |
| | | +3 | 31 (8.9) | 30 (8.6) |
| | | Education level, n (%) | | |
| | | No/Unknown education | 26 (7.5) | 26 (7.5) |
| | | Primary | 84 (24.2) | 84 (24.2) |
| | | Secondary | 142 (40.9) | 142 (40.9) |
| | | Tertiary | 95 (27.4) | 95 (27.4) |
| | | Relation to patient, n (%) | | |
| | | Partner | 347 (100.0) | 347 (100.0) |
| Older people (65+ years) | Siblings | Number of persons, n | 16 (100.0) | 16 (100.0) |
| | | Male, n (%) | 6 (37.5) | 6 (37.5) |
| | | Age, median (Q1;Q3) | 62.4 (59.5;64.6) | 62.6 (60.1;64.1) |
| | | CCI, n (%) | | |
| | | 0 | * | * |
| | | 1-2 | * | * |
| | | +3 | 0 (0.0) | * |
| | | Education level, n (%) | | |
| | | No/Unknown education | 0 (0.0) | 0 (0.0) |
| | | Primary | * | * |
| | | Secondary | * | * |
| | | Tertiary | * | * |
| | | Relation to patient, n (%) | | |
| | | Sibling | 16 (100.0) | 16 (100.0) |

### Supplementary Table 94
#### Developmental and behavioural disorders - Incident cohort 2016-2021
#### Cost analysis - pooled

| | | | | *Actual costs* | | *Attributable costs* | |
|---|---|---|---|---|---|---|---|
| *Age strata of index patients* | *Closest relative type* | *Number of index patients included in cost component calculations* | *Cost component* | *Total (million EUR)* | *Per person (EUR)* | *Total (million EUR)* | *Per person (EUR)* |
| Children and young people (0-24 years) | Fathers | 47,830 | Primary sector | 10.4 | 217 | 1.0 | 21 |
| | | 47,830 | Hospital care | 65.9 | 1,381 | 8.7 | 183 |
| | | 47,830 | Filled prescriptions | 7.7 | 162 | 1.5 | 32 |
| | | 46,914 | Lost productivity (income loss) | . | . | 355.8 | 7,584 |
| Children and young people (0-24 years) | Mothers | 50,091 | Primary sector | 18.8 | 375 | 3.0 | 61 |
| | | 50,091 | Hospital care | 89.6 | 1,790 | 13.7 | 274 |
| | | 50,091 | Filled prescriptions | 10.3 | 206 | 2.6 | 51 |
| | | 49,849 | Lost productivity (income loss) | . | . | 292.0 | 5,857 |
| Young adults and adults (25-64 years) | Pooled | 20,266 | Primary sector | 6.9 | 343 | 1.1 | 52 |
| | | 20,266 | Hospital care | 42.7 | 2,114 | 6.9 | 343 |
| | | 20,266 | Filled prescriptions | 5.3 | 261 | 1.0 | 48 |
| | | 14,984 | Lost productivity (income loss) | . | . | 47.2 | 3,151 |
| Older people (65+ years) | Pooled | 711 | Primary sector | 0.3 | 486 | 0.0 | 68 |
| | | 711 | Hospital care | 1.9 | 2,732 | 0.1 | 160 |
| | | 711 | Filled prescriptions | 0.3 | 377 | -0.0 | -30 |
| | | 351 | Lost productivity (income loss) | . | . | 1.3 | 3,804 |

#### Supplementary Table 95
#### Developmental and behavioural disorders - Incident cohort 2016-2021
#### Cost analysis - not pooled

| | | | | *Actual costs* | | *Attributable costs* | |
|---|---|---|---|---|---|---|---|
| ***Age strata of index patients*** | ***Closest relative type*** | ***Number of index patients included in cost component calculations*** | ***Cost component*** | ***Total (million EUR)*** | ***Per person (EUR)*** | ***Total (million EUR)*** | ***Per person (EUR)*** |
| Children and young people (0-24 years) | Fathers | 47,830 | Primary sector | 10.4 | 217 | 1.0 | 21 |
| | | 47,830 | Hospital care | 65.9 | 1,381 | 8.7 | 183 |
| | | 47,830 | Filled prescriptions | 7.7 | 162 | 1.5 | 32 |
| | | 46,914 | Lost productivity (income loss) | . | . | 355.8 | 7,584 |
| Children and young people (0-24 years) | Mothers | 50,091 | Primary sector | 18.8 | 375 | 3.0 | 61 |
| | | 50,091 | Hospital care | 89.6 | 1,790 | 13.7 | 274 |
| | | 50,091 | Filled prescriptions | 10.3 | 206 | 2.6 | 51 |
| | | 49,849 | Lost productivity (income loss) | . | . | 292.0 | 5,857 |
| Young adults and adults (25-64 years) | Children | 2,699 | Primary sector | 0.6 | 220 | 0.1 | 53 |
| | | 2,699 | Hospital care | 2.1 | 787 | 0.4 | 162 |
| | | 2,699 | Filled prescriptions | 0.2 | 76 | 0.1 | 20 |
| | | 625 | Lost productivity (income loss) | . | . | 1.4 | 2,212 |
| Young adults and adults (25-64 years) | Parents | 6,912 | Primary sector | 3.0 | 436 | 0.3 | 40 |
| | | 6,912 | Hospital care | 20.7 | 3,016 | 2.5 | 371 |
| | | 6,912 | Filled prescriptions | 3.1 | 445 | 0.4 | 65 |
| | | 3,971 | Lost productivity (income loss) | . | . | 7.9 | 1,991 |
| Young adults and adults (25-64 years) | Partners | 9,279 | Primary sector | 3.0 | 321 | 0.6 | 65 |
| | | 9,279 | Hospital care | 16.2 | 1,752 | 2.4 | 255 |
| | | 9,279 | Filled prescriptions | 1.7 | 186 | 0.4 | 43 |
| | | 9,108 | Lost productivity (income loss) | . | . | 42.7 | 4,688 |
| Young adults and adults (25-64 years) | Siblings | 1,376 | Primary sector | 0.4 | 273 | 0.0 | 26 |
| | | 1,376 | Hospital care | 3.6 | 2,655 | 1.6 | 1,155 |
| | | 1,376 | Filled prescriptions | 0.3 | 216 | 0.1 | 47 |
| | | 1,280 | Lost productivity (income loss) | . | . | -4.8 | -3,726 |
| Older people (65+ years) | Children | 348 | Primary sector | 0.1 | 376 | 0.0 | 84 |
| | | 348 | Hospital care | 0.5 | 1,440 | 0.1 | 291 |
| | | 348 | Filled prescriptions | 0.1 | 205 | -0.0 | -21 |
| | | 323 | Lost productivity (income loss) | . | . | 0.8 | 2,583 |
| Older people (65+ years) | Partners | 347 | Primary sector | 0.2 | 596 | 0.0 | 53 |
| | | 347 | Hospital care | 1.4 | 3,998 | -0.0 | -76 |
| | | 347 | Filled prescriptions | 0.2 | 547 | -0.0 | -54 |
| | | 21 | Lost productivity (income loss) | . | . | 0.4 | 18,060 |
| Older people (65+ years) | Siblings | 16 | Primary sector | 0.0 | 506 | 0.0 | 48 |
| | | 16 | Hospital care | 0.1 | 3,548 | 0.0 | 2,364 |
| | | 16 | Filled prescriptions | 0.0 | 453 | 0.0 | 275 |
| | | 7 | Lost productivity (income loss) | . | . | 0.1 | 17,362 |

**Supplementary Table 96**
**Developmental and behavioural disorders - Prevalent cohort 2021**
**Identification of populations of relatives**

| ***Agegroup of index patients in matched population*** | | ***Closest relative type identified*** | | ***Available control with same type of closest relative*** |
|---|---|---|---|---|
| Children and young people (0-24 years) | n = 103,684 | Fathers | n = 83,499 (81%) | 96% |
| | | Mothers | n = 87,349 (84%) | 96% |
| | | No available closest relative | n = 5,636 (5%) | n/a |
| Young adults and adults (25-64 years) | n = 82,932 | Children | n = 8,242 (10%) | 94% |
| | | Parents | n = 30,210 (36%) | 93% |
| | | Partners | n = 27,512 (33%) | 95% |
| | | Siblings | n = 15,440 (19%) | 36% |
| | | Pooled | n = 81,404 (98%) | 83% |
| | | No available closest relative | n = 1,528 (2%) | n/a |
| Older people (65+ years) | n = 2,222 | Children | n = 717 (32%) | 90% |
| | | Partners | n = 838 (38%) | 95% |
| | | Siblings | n = 163 (7%) | 53% |
| | | Pooled | n = 1,718 (77%) | 89% |
| | | No available closest relative | n = 504 (23%) | n/a |

**Supplementary Table 97**
**Developmental and behavioural disorders - Prevalent cohort 2021**
**Patient characteristics - cases**

| *Age strata of index patients* | *Characteristic* | |
|---|---|---|
| Children and young people (0-24 years) | Number of persons, n | 95,079 (100.0) |
| | Male, n (%) | 62,139 (65.4) |
| | Age, median (Q1;Q3) | 17.6 (13.5;21.3) |
| | CCI, n (%) | |
| | 0 | 87,918 (92.5) |
| | 1-2 | 6,958 (7.3) |
| | +3 | 203 (0.2) |
| | Education level, n (%) | |
| | No/Unknown education | 22,782 (24.0) |
| | Primary | 61,736 (64.9) |
| | Secondary | 10,014 (10.5) |
| | Tertiary | 547 (0.6) |
| Young adults and adults (25-64 years) | Number of persons, n | 68,117 (100.0) |
| | Male, n (%) | 40,194 (59.0) |
| | Age, median (Q1;Q3) | 33.2 (28.3;43.3) |
| | CCI, n (%) | |
| | 0 | 59,855 (87.9) |
| | 1-2 | 7,429 (10.9) |
| | +3 | 833 (1.2) |
| | Education level, n (%) | |
| | No/Unknown education | 1,124 (1.7) |
| | Primary | 29,184 (42.8) |
| | Secondary | 26,461 (38.8) |
| | Tertiary | 11,348 (16.7) |
| Older people (65+ years) | Number of persons, n | 1,538 (100.0) |
| | Male, n (%) | 726 (47.2) |
| | Age, median (Q1;Q3) | 70.7 (67.3;76.3) |
| | CCI, n (%) | |
| | 0 | 739 (48.0) |
| | 1-2 | 556 (36.2) |
| | +3 | 243 (15.8) |
| | Education level, n (%) | |
| | No/Unknown education | 112 (7.3) |
| | Primary | 406 (26.4) |
| | Secondary | 622 (40.4) |
| | Tertiary | 398 (25.9) |

**Supplementary Table 98**
**Developmental and behavioural disorders - Prevalent cohort 2021**
**Patient characteristics - relatives - pooled**

| *Age strata of index patients* | *Closest relative type* | *Characteristic of family relatives by agestrata of index patients* | *Closest family relatives of brain disorder patients* | *Comparison relative* |
|---|---|---|---|---|
| Children and young people (0-24 years) | Fathers | Number of persons, n | 80,831 (100.0) | 80,831 (100.0) |
| | | Male, n (%) | 80,831 (100.0) | 80,831 (100.0) |
| | | Age, median (Q1;Q3) | 49.5 (44.7;54.3) | 49.6 (45.1;54.1) |
| | | CCI, n (%) | | |
| | | 0 | 69,807 (86.4) | 71,602 (88.6) |
| | | 1-2 | 9,650 (11.9) | 8,147 (10.1) |
| | | +3 | 1,374 (1.7) | 1,082 (1.3) |
| | | Education level, n (%) | | |
| | | No/Unknown education | 1,158 (1.4) | 1,158 (1.4) |
| | | Primary | 18,620 (23.0) | 18,620 (23.0) |
| | | Secondary | 41,226 (51.0) | 41,226 (51.0) |
| | | Tertiary | 19,827 (24.5) | 19,827 (24.5) |
| | | Relation to patient, n (%) | | |
| | | Father | 80,831 (100.0) | 80,831 (100.0) |
| Children and young people (0-24 years) | Mothers | Number of persons, n | 84,691 (100.0) | 84,691 (100.0) |
| | | Male, n (%) | 0 (0.0) | 0 (0.0) |
| | | Age, median (Q1;Q3) | 47.1 (42.4;51.7) | 47.4 (42.8;51.6) |
| | | CCI, n (%) | | |
| | | 0 | 72,037 (85.1) | 74,767 (88.3) |
| | | 1-2 | 11,512 (13.6) | 9,075 (10.7) |
| | | +3 | 1,142 (1.3) | 849 (1.0) |
| | | Education level, n (%) | | |
| | | No/Unknown education | 945 (1.1) | 945 (1.1) |
| | | Primary | 16,064 (19.0) | 16,064 (19.0) |
| | | Secondary | 37,641 (44.4) | 37,641 (44.4) |
| | | Tertiary | 30,041 (35.5) | 30,041 (35.5) |
| | | Relation to patient, n (%) | | |
| | | Mother | 84,691 (100.0) | 84,691 (100.0) |
| Young adults and adults (25-64 years) | Pooled | Number of persons, n | 68,117 (100.0) | 68,117 (100.0) |
| | | Male, n (%) | 26,467 (38.9) | 26,467 (38.9) |
| | | Age, median (Q1;Q3) | 48.8 (30.2;58.9) | 49.2 (30.3;59.1) |
| | | CCI, n (%) | | |
| | | 0 | 56,521 (83.0) | 57,938 (85.1) |
| | | 1-2 | 9,842 (14.4) | 8,624 (12.7) |
| | | +3 | 1,754 (2.6) | 1,555 (2.3) |
| | | Education level, n (%) | | |
| | | No/Unknown education | 5,078 (7.5) | 5,078 (7.5) |
| | | Primary | 17,360 (25.5) | 17,360 (25.5) |
| | | Secondary | 30,244 (44.4) | 30,244 (44.4) |
| | | Tertiary | 15,435 (22.7) | 15,435 (22.7) |
| | | Relation to patient, n (%) | | |
| | | Mother | 21,319 (31.3) | 21,319 (31.3) |
| | | Father | 7,036 (10.3) | 7,036 (10.3) |
| | | Partner | 26,331 (38.7) | 26,331 (38.7) |
| | | Child | 7,784 (11.4) | 7,784 (11.4) |

**Supplementary Table 98**
**Developmental and behavioural disorders - Prevalent cohort 2021**
**Patient characteristics - relatives - pooled**

| *Age strata of index patients* | *Closest relative type* | *Characteristic of family relatives by agestrata of index patients* | *Closest family relatives of brain disorder patients* | *Comparison relative* |
|---|---|---|---|---|
| | | Sibling | 5,647 (8.3) | 5,647 (8.3) |
| Older people (65+ years) | Pooled | Number of persons, n | 1,538 (100.0) | 1,538 (100.0) |
| | | Male, n (%) | 719 (46.7) | 719 (46.7) |
| | | Age, median (Q1;Q3) | 62.8 (48.8;70.7) | 62.9 (48.4;71.0) |
| | | CCI, n (%) | | |
| | | 0 | 1,137 (73.9) | 1,166 (75.8) |
| | | 1-2 | 313 (20.4) | 294 (19.1) |
| | | +3 | 88 (5.7) | 78 (5.1) |
| | | Education level, n (%) | | |
| | | No/Unknown education | 64 (4.2) | 64 (4.2) |
| | | Primary | 286 (18.6) | 286 (18.6) |
| | | Secondary | 689 (44.8) | 689 (44.8) |
| | | Tertiary | 499 (32.4) | 499 (32.4) |
| | | Relation to patient, n (%) | | |
| | | Partner | 798 (51.9) | 798 (51.9) |
| | | Child | 652 (42.4) | 652 (42.4) |
| | | Sibling | 88 (5.7) | 88 (5.7) |

#### Supplementary Table 99
#### Developmental and behavioural disorders - Prevalent cohort 2021
#### Patient characteristics - relatives - not pooled

| *Age strata of index patients* | *Closest relative type* | *Characteristic of family relatives by agestrata of index patients* | *Closest family relatives of brain disorder patients* | *Comparison relative* |
|---|---|---|---|---|
| Children and young people (0-24 years) | Fathers | Number of persons, n | 80,831 (100.0) | 80,831 (100.0) |
| | | Male, n (%) | 80,831 (100.0) | 80,831 (100.0) |
| | | Age, median (Q1;Q3) | 49.5 (44.7;54.3) | 49.6 (45.1;54.1) |
| | | CCI, n (%) | | |
| | | 0 | 69,807 (86.4) | 71,602 (88.6) |
| | | 1-2 | 9,650 (11.9) | 8,147 (10.1) |
| | | +3 | 1,374 (1.7) | 1,082 (1.3) |
| | | Education level, n (%) | | |
| | | No/Unknown education | 1,158 (1.4) | 1,158 (1.4) |
| | | Primary | 18,620 (23.0) | 18,620 (23.0) |
| | | Secondary | 41,226 (51.0) | 41,226 (51.0) |
| | | Tertiary | 19,827 (24.5) | 19,827 (24.5) |
| | | Relation to patient, n (%) | | |
| | | Father | 80,831 (100.0) | 80,831 (100.0) |
| Children and young people (0-24 years) | Mothers | Number of persons, n | 84,691 (100.0) | 84,691 (100.0) |
| | | Male, n (%) | 0 (0.0) | 0 (0.0) |
| | | Age, median (Q1;Q3) | 47.1 (42.4;51.7) | 47.4 (42.8;51.6) |
| | | CCI, n (%) | | |
| | | 0 | 72,037 (85.1) | 74,767 (88.3) |
| | | 1-2 | 11,512 (13.6) | 9,075 (10.7) |
| | | +3 | 1,142 (1.3) | 849 (1.0) |
| | | Education level, n (%) | | |
| | | No/Unknown education | 945 (1.1) | 945 (1.1) |
| | | Primary | 16,064 (19.0) | 16,064 (19.0) |
| | | Secondary | 37,641 (44.4) | 37,641 (44.4) |
| | | Tertiary | 30,041 (35.5) | 30,041 (35.5) |
| | | Relation to patient, n (%) | | |
| | | Mother | 84,691 (100.0) | 84,691 (100.0) |
| Young adults and adults (25-64 years) | Children | Number of persons, n | 7,784 (100.0) | 7,784 (100.0) |
| | | Male, n (%) | 3,844 (49.4) | 3,844 (49.4) |
| | | Age, median (Q1;Q3) | 12.0 (6.1;19.5) | 11.4 (5.5;19.2) |
| | | CCI, n (%) | | |
| | | 0 | 7,260 (93.3) | 7,386 (94.9) |
| | | 1-2 | 512 (6.6) | 384 (4.9) |
| | | +3 | 12 (0.2) | 14 (0.2) |
| | | Education level, n (%) | | |
| | | No/Unknown education | 4,283 (55.0) | 4,283 (55.0) |
| | | Primary | 2,307 (29.6) | 2,307 (29.6) |
| | | Secondary | 886 (11.4) | 886 (11.4) |
| | | Tertiary | 308 (4.0) | 308 (4.0) |
| | | Relation to patient, n (%) | | |
| | | Child | 7,784 (100.0) | 7,784 (100.0) |
| Young adults and adults (25-64 years) | Parents | Number of persons, n | 28,355 (100.0) | 28,355 (100.0) |
| | | Male, n (%) | 7,036 (24.8) | 7,036 (24.8) |
| | | Age, median (Q1;Q3) | 59.7 (55.2;65.7) | 59.8 (55.6;65.8) |

**Supplementary Table 99**
**Developmental and behavioural disorders - Prevalent cohort 2021**
**Patient characteristics - relatives - not pooled**

| *Age strata of index patients* | *Closest relative type* | *Characteristic of family relatives by agestrata of index patients* | *Closest family relatives of brain disorder patients* | *Comparison relative* |
|---|---|---|---|---|
| | | CCI, n (%) | | |
| | | 0 | 20,479 (72.2) | 21,415 (75.5) |
| | | 1-2 | 6,452 (22.8) | 5,676 (20.0) |
| | | +3 | 1,424 (5.0) | 1,264 (4.5) |
| | | Education level, n (%) | | |
| | | No/Unknown education | 396 (1.4) | 396 (1.4) |
| | | Primary | 7,582 (26.7) | 7,582 (26.7) |
| | | Secondary | 13,843 (48.8) | 13,843 (48.8) |
| | | Tertiary | 6,534 (23.0) | 6,534 (23.0) |
| | | Relation to patient, n (%) | | |
| | | Mother | 21,319 (75.2) | 21,319 (75.2) |
| | | Father | 7,036 (24.8) | 7,036 (24.8) |
| Young adults and adults (25-64 years) | Partners | Number of persons, n | 26,331 (100.0) | 26,331 (100.0) |
| | | Male, n (%) | 12,011 (45.6) | 12,011 (45.6) |
| | | Age, median (Q1;Q3) | 35.0 (29.0;45.4) | 34.9 (29.2;45.3) |
| | | CCI, n (%) | | |
| | | 0 | 23,783 (90.3) | 24,180 (91.8) |
| | | 1-2 | 2,317 (8.8) | 1,969 (7.5) |
| | | +3 | 231 (0.9) | 182 (0.7) |
| | | Education level, n (%) | | |
| | | No/Unknown education | 381 (1.4) | 381 (1.4) |
| | | Primary | 5,852 (22.2) | 5,852 (22.2) |
| | | Secondary | 12,683 (48.2) | 12,683 (48.2) |
| | | Tertiary | 7,415 (28.2) | 7,415 (28.2) |
| | | Relation to patient, n (%) | | |
| | | Partner | 26,331 (100.0) | 26,331 (100.0) |
| Young adults and adults (25-64 years) | Siblings | Number of persons, n | 5,647 (100.0) | 5,647 (100.0) |
| | | Male, n (%) | 3,576 (63.3) | 3,576 (63.3) |
| | | Age, median (Q1;Q3) | 37.8 (28.6;52.5) | 37.6 (28.7;52.6) |
| | | CCI, n (%) | | |
| | | 0 | 4,999 (88.5) | 4,957 (87.8) |
| | | 1-2 | 561 (9.9) | 595 (10.5) |
| | | +3 | 87 (1.5) | 95 (1.7) |
| | | Education level, n (%) | | |
| | | No/Unknown education | 18 (0.3) | 18 (0.3) |
| | | Primary | 1,619 (28.7) | 1,619 (28.7) |
| | | Secondary | 2,832 (50.2) | 2,832 (50.2) |
| | | Tertiary | 1,178 (20.9) | 1,178 (20.9) |
| | | Relation to patient, n (%) | | |
| | | Sibling | 5,647 (100.0) | 5,647 (100.0) |
| Older people (65+ years) | Children | Number of persons, n | 652 (100.0) | 652 (100.0) |
| | | Male, n (%) | 335 (51.4) | 335 (51.4) |
| | | Age, median (Q1;Q3) | 46.8 (41.0;53.5) | 46.6 (40.9;53.2) |
| | | CCI, n (%) | | |
| | | 0 | 567 (87.0) | 562 (86.2) |

**Supplementary Table 99**
**Developmental and behavioural disorders - Prevalent cohort 2021**
**Patient characteristics - relatives - not pooled**

| *Age strata of index patients* | *Closest relative type* | *Characteristic of family relatives by agestrata of index patients* | *Closest family relatives of brain disorder patients* | *Comparison relative* |
|---|---|---|---|---|
| | | 1-2 | 69 (10.6) | 78 (12.0) |
| | | +3 | 16 (2.5) | 12 (1.8) |
| | | Education level, n (%) | | |
| | | No/Unknown education | * | * |
| | | Primary | * | * |
| | | Secondary | 274 (42.0) | 274 (42.0) |
| | | Tertiary | 259 (39.7) | 259 (39.7) |
| | | Relation to patient, n (%) | | |
| | | Child | 652 (100.0) | 652 (100.0) |
| Older people (65+ years) | Partners | Number of persons, n | 798 (100.0) | 798 (100.0) |
| | | Male, n (%) | 342 (42.9) | 342 (42.9) |
| | | Age, median (Q1;Q3) | 70.3 (65.8;75.6) | 70.7 (66.1;75.3) |
| | | CCI, n (%) | | |
| | | 0 | 503 (63.0) | 530 (66.4) |
| | | 1-2 | 227 (28.4) | 208 (26.1) |
| | | +3 | 68 (8.5) | 60 (7.5) |
| | | Education level, n (%) | | |
| | | No/Unknown education | 62 (7.8) | 62 (7.8) |
| | | Primary | 147 (18.4) | 147 (18.4) |
| | | Secondary | 365 (45.7) | 365 (45.7) |
| | | Tertiary | 224 (28.1) | 224 (28.1) |
| | | Relation to patient, n (%) | | |
| | | Partner | 798 (100.0) | 798 (100.0) |
| Older people (65+ years) | Siblings | Number of persons, n | 88 (100.0) | 88 (100.0) |
| | | Male, n (%) | 42 (47.7) | 42 (47.7) |
| | | Age, median (Q1;Q3) | 62.2 (59.8;65.0) | 61.9 (59.2;64.7) |
| | | CCI, n (%) | | |
| | | 0 | 67 (76.1) | 74 (84.1) |
| | | 1-2 | * | 8 (9.1) |
| | | +3 | * | 6 (6.8) |
| | | Education level, n (%) | | |
| | | No/Unknown education | * | * |
| | | Primary | 21 (23.9) | 21 (23.9) |
| | | Secondary | 50 (56.8) | 50 (56.8) |
| | | Tertiary | * | * |
| | | Relation to patient, n (%) | | |
| | | Sibling | 88 (100.0) | 88 (100.0) |

#### Supplementary Table 100
#### Developmental and behavioural disorders - Prevalent cohort 2021
#### Cost analysis - pooled

| | | | | *Actual costs* | | *Attributable costs* | |
|---|---|---|---|---|---|---|---|
| ***Age strata of index patients*** | ***Closest relative type*** | ***Number of index patients included in cost component calculations*** | ***Cost component*** | ***Total (million EUR)*** | ***Per person (EUR)*** | ***Total (million EUR)*** | ***Per person (EUR)*** |
| Children and young people (0-24 years) | Fathers | 80,831 | Primary sector | 18.9 | 234 | 1.5 | 18 |
| | | 80,831 | Hospital care | 134.3 | 1,665 | 23.8 | 295 |
| | | 80,831 | Filled prescriptions | 16.6 | 205 | 3.5 | 44 |
| | | 78,468 | Lost productivity (income loss) | . | . | 824.8 | 10,511 |
| Children and young people (0-24 years) | Mothers | 84,691 | Primary sector | 32.6 | 386 | 4.8 | 57 |
| | | 84,691 | Hospital care | 170.8 | 2,019 | 45.6 | 539 |
| | | 84,691 | Filled prescriptions | 20.8 | 246 | 6.1 | 72 |
| | | 84,214 | Lost productivity (income loss) | . | . | 631.7 | 7,501 |
| Young adults and adults (25-64 years) | Pooled | 68,117 | Primary sector | 23.0 | 339 | 2.3 | 35 |
| | | 68,117 | Hospital care | 152.8 | 2,252 | 21.3 | 314 |
| | | 68,117 | Filled prescriptions | 19.3 | 284 | 3.8 | 56 |
| | | 51,063 | Lost productivity (income loss) | . | . | 290.6 | 5,691 |
| Older people (65+ years) | Pooled | 1,538 | Primary sector | 0.7 | 443 | 0.1 | 39 |
| | | 1,538 | Hospital care | 4.2 | 2,740 | 0.2 | 162 |
| | | 1,538 | Filled prescriptions | 0.6 | 376 | 0.0 | 11 |
| | | 755 | Lost productivity (income loss) | . | . | 2.5 | 3,298 |

#### Supplementary Table 101
#### Developmental and behavioural disorders - Prevalent cohort 2021
#### Cost analysis - not pooled

| | | | | *Actual costs* | | *Attributable costs* | |
|---|---|---|---|---|---|---|---|
| ***Age strata of index patients*** | ***Closest relative type*** | ***Number of index patients included in cost component calculations*** | ***Cost component*** | ***Total (million EUR)*** | ***Per person (EUR)*** | ***Total (million EUR)*** | ***Per person (EUR)*** |
| Children and young people (0-24 years) | Fathers | 80,831 | Primary sector | 18.9 | 234 | 1.5 | 18 |
| | | 80,831 | Hospital care | 134.3 | 1,665 | 23.8 | 295 |
| | | 80,831 | Filled prescriptions | 16.6 | 205 | 3.5 | 44 |
| | | 78,468 | Lost productivity (income loss) | . | . | 824.8 | 10,511 |
| Children and young people (0-24 years) | Mothers | 84,691 | Primary sector | 32.6 | 386 | 4.8 | 57 |
| | | 84,691 | Hospital care | 170.8 | 2,019 | 45.6 | 539 |
| | | 84,691 | Filled prescriptions | 20.8 | 246 | 6.1 | 72 |
| | | 84,214 | Lost productivity (income loss) | . | . | 631.7 | 7,501 |
| Young adults and adults (25-64 years) | Children | 7,784 | Primary sector | 1.6 | 204 | 0.3 | 36 |
| | | 7,784 | Hospital care | 6.7 | 864 | 0.9 | 122 |
| | | 7,784 | Filled prescriptions | 0.5 | 70 | 0.1 | 13 |
| | | 1,956 | Lost productivity (income loss) | . | . | 7.7 | 3,923 |
| Young adults and adults (25-64 years) | Parents | 28,355 | Primary sector | 12.2 | 431 | 1.2 | 43 |
| | | 28,355 | Hospital care | 86.9 | 3,085 | 11.6 | 413 |
| | | 28,355 | Filled prescriptions | 12.8 | 453 | 2.4 | 84 |
| | | 18,102 | Lost productivity (income loss) | . | . | 126.4 | 6,980 |
| Young adults and adults (25-64 years) | Partners | 26,331 | Primary sector | 7.7 | 293 | 0.8 | 29 |
| | | 26,331 | Hospital care | 48.5 | 1,844 | 8.7 | 330 |
| | | 26,331 | Filled prescriptions | 4.9 | 185 | 1.3 | 49 |
| | | 25,854 | Lost productivity (income loss) | . | . | 153.4 | 5,935 |
| Young adults and adults (25-64 years) | Siblings | 5,647 | Primary sector | 1.5 | 273 | 0.1 | 15 |
| | | 5,647 | Hospital care | 10.7 | 1,899 | 0.0 | 7 |
| | | 5,647 | Filled prescriptions | 1.1 | 202 | 0.1 | 12 |
| | | 5,151 | Lost productivity (income loss) | . | . | 3.1 | 605 |
| Older people (65+ years) | Children | 652 | Primary sector | 0.2 | 336 | 0.0 | 39 |
| | | 652 | Hospital care | 1.1 | 1,678 | 0.1 | 133 |
| | | 652 | Filled prescriptions | 0.1 | 203 | 0.0 | 3 |
| | | 623 | Lost productivity (income loss) | . | . | 3.3 | 5,262 |
| Older people (65+ years) | Partners | 798 | Primary sector | 0.4 | 527 | 0.0 | 43 |
| | | 798 | Hospital care | 2.9 | 3,711 | 0.3 | 383 |
| | | 798 | Filled prescriptions | 0.4 | 527 | 0.1 | 118 |
| | | 83 | Lost productivity (income loss) | . | . | -1.2 | -14,213 |
| Older people (65+ years) | Siblings | 88 | Primary sector | 0.0 | 476 | 0.0 | 8 |
| | | 88 | Hospital care | 0.2 | 1,809 | -0.1 | -1,638 |
| | | 88 | Filled prescriptions | 0.0 | 300 | -0.1 | -912 |
| | | 49 | Lost productivity (income loss) | . | . | 0.4 | 7,988 |

**Supplementary Table 102**
**Drug use disorder - Incident cohort 2016-2021**
**Identification of populations of relatives**

| ***Agegroup of index patients in matched population*** | | ***Closest relative type identified*** | | ***Available control with same type of closest relative*** |
|---|---|---|---|---|
| Children and young people (0-24 years) | n = 8,650 | Fathers | n = 6,647 (77%) | 95% |
| | | Mothers | n = 7,631 (88%) | 95% |
| | | No available closest relative | n = 543 (6%) | n/a |
| Young adults and adults (25-64 years) | n = 9,851 | Children | n = 1,344 (14%) | 92% |
| | | Parents | n = 3,356 (34%) | 92% |
| | | Partners | n = 2,293 (23%) | 94% |
| | | Siblings | n = 2,283 (23%) | 37% |
| | | Pooled | n = 9,276 (94%) | 79% |
| | | No available closest relative | n = 575 (6%) | n/a |
| Older people (65+ years) | n = 1,112 | Children | n = 514 (46%) | 93% |
| | | Partners | n = 399 (36%) | 94% |
| | | Siblings | n = 45 (4%) | 33% |
| | | Pooled | n = 958 (86%) | 91% |
| | | No available closest relative | n = 154 (14%) | n/a |

**Supplementary Table 103**
**Drug use disorder - Incident cohort 2016-2021**
**Patient characteristics - cases**

| ***Age strata of index patients*** | ***Characteristic*** | |
|---|---|---|
| Children and young people (0-24 years) | Number of persons, n | 7,768 (100.0) |
| | Male, n (%) | 5,121 (65.9) |
| | Age, median (Q1;Q3) | 20.2 (17.9;22.4) |
| | CCI, n (%) | |
| | 0 | 7,266 (93.5) |
| | 1-2 | 479 (6.2) |
| | +3 | 23 (0.3) |
| | Education level, n (%) | |
| | No/Unknown education | 66 (0.8) |
| | Primary | 6,551 (84.3) |
| | Secondary | 1,125 (14.5) |
| | Tertiary | 26 (0.3) |
| Young adults and adults (25-64 years) | Number of persons, n | 7,363 (100.0) |
| | Male, n (%) | 5,239 (71.2) |
| | Age, median (Q1;Q3) | 35.0 (28.7;46.3) |
| | CCI, n (%) | |
| | 0 | 6,044 (82.1) |
| | 1-2 | 1,116 (15.2) |
| | +3 | 203 (2.8) |
| | Education level, n (%) | |
| | No/Unknown education | 213 (2.9) |
| | Primary | 3,483 (47.3) |
| | Secondary | 2,881 (39.1) |
| | Tertiary | 786 (10.7) |
| Older people (65+ years) | Number of persons, n | 874 (100.0) |
| | Male, n (%) | 296 (33.9) |
| | Age, median (Q1;Q3) | 74.1 (69.4;80.3) |
| | CCI, n (%) | |
| | 0 | 329 (37.6) |
| | 1-2 | 355 (40.6) |
| | +3 | 190 (21.7) |
| | Education level, n (%) | |
| | No/Unknown education | 54 (6.2) |
| | Primary | 327 (37.4) |
| | Secondary | 375 (42.9) |
| | Tertiary | 118 (13.5) |

**Supplementary Table 104**
**Drug use disorder - Incident cohort 2016-2021**
**Patient characteristics - relatives - pooled**

| *Age strata of index patients* | *Closest relative type* | *Characteristic of family relatives by agestrata of index patients* | *Closest family relatives of brain disorder patients* | *Comparison relative* |
|---|---|---|---|---|
| Children and young people (0-24 years) | Fathers | Number of persons, n | 6,363 (100.0) | 6,363 (100.0) |
| | | Male, n (%) | 6,363 (100.0) | 6,363 (100.0) |
| | | Age, median (Q1;Q3) | 51.3 (47.0;55.4) | 51.3 (47.6;55.1) |
| | | CCI, n (%) | | |
| | | 0 | 5,321 (83.6) | 5,450 (85.7) |
| | | 1-2 | 886 (13.9) | 804 (12.6) |
| | | +3 | 156 (2.5) | 109 (1.7) |
| | | Education level, n (%) | | |
| | | No/Unknown education | 104 (1.6) | 104 (1.6) |
| | | Primary | 1,891 (29.7) | 1,891 (29.7) |
| | | Secondary | 3,247 (51.0) | 3,247 (51.0) |
| | | Tertiary | 1,121 (17.6) | 1,121 (17.6) |
| | | Relation to patient, n (%) | | |
| | | Father | 6,363 (100.0) | 6,363 (100.0) |
| Children and young people (0-24 years) | Mothers | Number of persons, n | 7,319 (100.0) | 7,319 (100.0) |
| | | Male, n (%) | 0 (0.0) | 0 (0.0) |
| | | Age, median (Q1;Q3) | 48.5 (44.4;52.7) | 48.8 (45.3;52.4) |
| | | CCI, n (%) | | |
| | | 0 | 6,054 (82.7) | 6,376 (87.1) |
| | | 1-2 | 1,133 (15.5) | 852 (11.6) |
| | | +3 | 132 (1.8) | 91 (1.2) |
| | | Education level, n (%) | | |
| | | No/Unknown education | 59 (0.8) | 59 (0.8) |
| | | Primary | 1,972 (26.9) | 1,972 (26.9) |
| | | Secondary | 3,391 (46.3) | 3,391 (46.3) |
| | | Tertiary | 1,897 (25.9) | 1,897 (25.9) |
| | | Relation to patient, n (%) | | |
| | | Mother | 7,319 (100.0) | 7,319 (100.0) |
| Young adults and adults (25-64 years) | Pooled | Number of persons, n | 7,363 (100.0) | 7,363 (100.0) |
| | | Male, n (%) | 2,681 (36.4) | 2,681 (36.4) |
| | | Age, median (Q1;Q3) | 49.7 (28.9;59.2) | 50.2 (28.9;59.5) |
| | | CCI, n (%) | | |
| | | 0 | 5,957 (80.9) | 6,181 (83.9) |
| | | 1-2 | 1,166 (15.8) | 979 (13.3) |
| | | +3 | 240 (3.3) | 203 (2.8) |
| | | Education level, n (%) | | |
| | | No/Unknown education | 631 (8.6) | 631 (8.6) |
| | | Primary | 2,356 (32.0) | 2,356 (32.0) |
| | | Secondary | 3,125 (42.4) | 3,125 (42.4) |
| | | Tertiary | 1,251 (17.0) | 1,251 (17.0) |
| | | Relation to patient, n (%) | | |
| | | Mother | 2,353 (32.0) | 2,353 (32.0) |
| | | Father | 755 (10.3) | 755 (10.3) |
| | | Partner | 2,162 (29.4) | 2,162 (29.4) |
| | | Child | 1,244 (16.9) | 1,244 (16.9) |

**Supplementary Table 104**
**Drug use disorder - Incident cohort 2016-2021**
**Patient characteristics - relatives - pooled**

| *Age strata of index patients* | *Closest relative type* | *Characteristic of family relatives by agestrata of index patients* | *Closest family relatives of brain disorder patients* | *Comparison relative* |
|---|---|---|---|---|
| | | Sibling | 849 (11.5) | 849 (11.5) |
| Older people (65+ years) | Pooled | Number of persons, n | 874 (100.0) | 874 (100.0) |
| | | Male, n (%) | 471 (53.9) | 471 (53.9) |
| | | Age, median (Q1;Q3) | 60.0 (49.7;71.7) | 60.3 (49.8;71.9) |
| | | CCI, n (%) | | |
| | | 0 | 626 (71.6) | 639 (73.1) |
| | | 1-2 | 187 (21.4) | 188 (21.5) |
| | | +3 | 61 (7.0) | 47 (5.4) |
| | | Education level, n (%) | | |
| | | No/Unknown education | 24 (2.7) | 24 (2.7) |
| | | Primary | 188 (21.5) | 188 (21.5) |
| | | Secondary | 458 (52.4) | 458 (52.4) |
| | | Tertiary | 204 (23.3) | 204 (23.3) |
| | | Relation to patient, n (%) | | |
| | | Partner | 377 (43.1) | 377 (43.1) |
| | | Child | 482 (55.1) | 482 (55.1) |
| | | Sibling | 15 (1.7) | 15 (1.7) |

#### Supplementary Table 105
#### Drug use disorder - Incident cohort 2016-2021
#### Patient characteristics - relatives - not pooled

| *Age strata of index patients* | *Closest relative type* | *Characteristic of family relatives by agestrata of index patients* | *Closest family relatives of brain disorder patients* | *Comparison relative* |
|---|---|---|---|---|
| Children and young people (0-24 years) | Fathers | Number of persons, n | 6,363 (100.0) | 6,363 (100.0) |
| | | Male, n (%) | 6,363 (100.0) | 6,363 (100.0) |
| | | Age, median (Q1;Q3) | 51.3 (47.0;55.4) | 51.3 (47.6;55.1) |
| | | CCI, n (%) | | |
| | | 0 | 5,321 (83.6) | 5,450 (85.7) |
| | | 1-2 | 886 (13.9) | 804 (12.6) |
| | | +3 | 156 (2.5) | 109 (1.7) |
| | | Education level, n (%) | | |
| | | No/Unknown education | 104 (1.6) | 104 (1.6) |
| | | Primary | 1,891 (29.7) | 1,891 (29.7) |
| | | Secondary | 3,247 (51.0) | 3,247 (51.0) |
| | | Tertiary | 1,121 (17.6) | 1,121 (17.6) |
| | | Relation to patient, n (%) | | |
| | | Father | 6,363 (100.0) | 6,363 (100.0) |
| Children and young people (0-24 years) | Mothers | Number of persons, n | 7,319 (100.0) | 7,319 (100.0) |
| | | Male, n (%) | 0 (0.0) | 0 (0.0) |
| | | Age, median (Q1;Q3) | 48.5 (44.4;52.7) | 48.8 (45.3;52.4) |
| | | CCI, n (%) | | |
| | | 0 | 6,054 (82.7) | 6,376 (87.1) |
| | | 1-2 | 1,133 (15.5) | 852 (11.6) |
| | | +3 | 132 (1.8) | 91 (1.2) |
| | | Education level, n (%) | | |
| | | No/Unknown education | 59 (0.8) | 59 (0.8) |
| | | Primary | 1,972 (26.9) | 1,972 (26.9) |
| | | Secondary | 3,391 (46.3) | 3,391 (46.3) |
| | | Tertiary | 1,897 (25.9) | 1,897 (25.9) |
| | | Relation to patient, n (%) | | |
| | | Mother | 7,319 (100.0) | 7,319 (100.0) |
| Young adults and adults (25-64 years) | Children | Number of persons, n | 1,244 (100.0) | 1,244 (100.0) |
| | | Male, n (%) | 658 (52.9) | 658 (52.9) |
| | | Age, median (Q1;Q3) | 16.5 (9.1;25.1) | 16.3 (8.0;24.1) |
| | | CCI, n (%) | | |
| | | 0 | 1,154 (92.8) | 1,171 (94.1) |
| | | 1-2 | * | * |
| | | +3 | * | * |
| | | Education level, n (%) | | |
| | | No/Unknown education | 486 (39.1) | 486 (39.1) |
| | | Primary | 478 (38.4) | 478 (38.4) |
| | | Secondary | 198 (15.9) | 198 (15.9) |
| | | Tertiary | 82 (6.6) | 82 (6.6) |
| | | Relation to patient, n (%) | | |
| | | Child | 1,244 (100.0) | 1,244 (100.0) |
| Young adults and adults (25-64 years) | Parents | Number of persons, n | 3,108 (100.0) | 3,108 (100.0) |
| | | Male, n (%) | 755 (24.3) | 755 (24.3) |
| | | Age, median (Q1;Q3) | 59.4 (54.8;66.3) | 59.7 (55.2;66.4) |

**Supplementary Table 105**
**Drug use disorder - Incident cohort 2016-2021**
**Patient characteristics - relatives - not pooled**

| *Age strata of index patients* | *Closest relative type* | *Characteristic of family relatives by agestrata of index patients* | *Closest family relatives of brain disorder patients* | *Comparison relative* |
|---|---|---|---|---|
| | | CCI, n (%) | | |
| | | 0 | 2,159 (69.5) | 2,319 (74.6) |
| | | 1-2 | 751 (24.2) | 630 (20.3) |
| | | +3 | 198 (6.4) | 159 (5.1) |
| | | Education level, n (%) | | |
| | | No/Unknown education | 67 (2.2) | 67 (2.2) |
| | | Primary | 967 (31.1) | 967 (31.1) |
| | | Secondary | 1,497 (48.2) | 1,497 (48.2) |
| | | Tertiary | 577 (18.6) | 577 (18.6) |
| | | Relation to patient, n (%) | | |
| | | Mother | 2,353 (75.7) | 2,353 (75.7) |
| | | Father | 755 (24.3) | 755 (24.3) |
| Young adults and adults (25-64 years) | Partners | Number of persons, n | 2,162 (100.0) | 2,162 (100.0) |
| | | Male, n (%) | 687 (31.8) | 687 (31.8) |
| | | Age, median (Q1;Q3) | 35.3 (28.4;46.7) | 35.4 (28.6;46.7) |
| | | CCI, n (%) | | |
| | | 0 | 1,927 (89.1) | 1,954 (90.4) |
| | | 1-2 | 211 (9.8) | 180 (8.3) |
| | | +3 | 24 (1.1) | 28 (1.3) |
| | | Education level, n (%) | | |
| | | No/Unknown education | 71 (3.3) | 71 (3.3) |
| | | Primary | 620 (28.7) | 620 (28.7) |
| | | Secondary | 1,014 (46.9) | 1,014 (46.9) |
| | | Tertiary | 457 (21.1) | 457 (21.1) |
| | | Relation to patient, n (%) | | |
| | | Partner | 2,162 (100.0) | 2,162 (100.0) |
| Young adults and adults (25-64 years) | Siblings | Number of persons, n | 849 (100.0) | 849 (100.0) |
| | | Male, n (%) | 581 (68.4) | 581 (68.4) |
| | | Age, median (Q1;Q3) | 41.5 (29.1;54.8) | 41.5 (29.2;55.0) |
| | | CCI, n (%) | | |
| | | 0 | 717 (84.5) | 737 (86.8) |
| | | 1-2 | 118 (13.9) | 97 (11.4) |
| | | +3 | 14 (1.6) | 15 (1.8) |
| | | Education level, n (%) | | |
| | | No/Unknown education | 7 (0.8) | 7 (0.8) |
| | | Primary | 291 (34.3) | 291 (34.3) |
| | | Secondary | 416 (49.0) | 416 (49.0) |
| | | Tertiary | 135 (15.9) | 135 (15.9) |
| | | Relation to patient, n (%) | | |
| | | Sibling | 849 (100.0) | 849 (100.0) |
| Older people (65+ years) | Children | Number of persons, n | 482 (100.0) | 482 (100.0) |
| | | Male, n (%) | 245 (50.8) | 245 (50.8) |
| | | Age, median (Q1;Q3) | 51.0 (44.8;56.5) | 51.0 (44.1;56.6) |
| | | CCI, n (%) | | |
| | | 0 | 407 (84.4) | 404 (83.8) |

#### Supplementary Table 105
#### Drug use disorder - Incident cohort 2016-2021
#### Patient characteristics - relatives - not pooled

| *Age strata of index patients* | *Closest relative type* | *Characteristic of family relatives by agestrata of index patients* | *Closest family relatives of brain disorder patients* | *Comparison relative* |
|---|---|---|---|---|
| | | 1-2 | 62 (12.9) | 69 (14.3) |
| | | +3 | 13 (2.7) | 9 (1.9) |
| | | Education level, n (%) | | |
| | | No/Unknown education | * | * |
| | | Primary | * | * |
| | | Secondary | 253 (52.5) | 253 (52.5) |
| | | Tertiary | 146 (30.3) | 146 (30.3) |
| | | Relation to patient, n (%) | | |
| | | Child | 482 (100.0) | 482 (100.0) |
| Older people (65+ years) | Partners | Number of persons, n | 377 (100.0) | 377 (100.0) |
| | | Male, n (%) | 217 (57.6) | 217 (57.6) |
| | | Age, median (Q1;Q3) | 73.5 (68.6;78.6) | 73.3 (68.6;78.4) |
| | | CCI, n (%) | | |
| | | 0 | 207 (54.9) | 223 (59.2) |
| | | 1-2 | 122 (32.4) | 116 (30.8) |
| | | +3 | 48 (12.7) | 38 (10.1) |
| | | Education level, n (%) | | |
| | | No/Unknown education | 23 (6.1) | 23 (6.1) |
| | | Primary | 103 (27.3) | 103 (27.3) |
| | | Secondary | 196 (52.0) | 196 (52.0) |
| | | Tertiary | 55 (14.6) | 55 (14.6) |
| | | Relation to patient, n (%) | | |
| | | Partner | 377 (100.0) | 377 (100.0) |
| Older people (65+ years) | Siblings | Number of persons, n | 15 (100.0) | 15 (100.0) |
| | | Male, n (%) | 9 (60.0) | 9 (60.0) |
| | | Age, median (Q1;Q3) | 60.0 (58.2;61.1) | 59.7 (58.2;61.5) |
| | | CCI, n (%) | | |
| | | 0 | * | * |
| | | 1-2 | * | * |
| | | +3 | 0 (0.0) | 0 (0.0) |
| | | Education level, n (%) | | |
| | | No/Unknown education | 0 (0.0) | 0 (0.0) |
| | | Primary | * | * |
| | | Secondary | * | * |
| | | Tertiary | * | * |
| | | Relation to patient, n (%) | | |
| | | Sibling | 15 (100.0) | 15 (100.0) |

#### Supplementary Table 106
#### Drug use disorder - Incident cohort 2016-2021
#### Cost analysis - pooled

| *Age strata of index patients* | *Closest relative type* | *Number of index patients included in cost component calculations* | *Cost component* | *Actual costs* | | *Attributable costs* | |
|---|---|---|---|---|---|---|---|
| | | | | *Total (million EUR)* | *Per person (EUR)* | *Total (million EUR)* | *Per person (EUR)* |
| Children and young people (0-24 years) | Fathers | 6,363 | Primary sector | 1.6 | 253 | 0.1 | 16 |
| | | 6,363 | Hospital care | 12.8 | 2,021 | 3.3 | 519 |
| | | 6,363 | Filled prescriptions | 1.7 | 273 | 0.5 | 73 |
| | | 6,072 | Lost productivity (income loss) | . | . | 109.2 | 17,978 |
| Children and young people (0-24 years) | Mothers | 7,319 | Primary sector | 2.8 | 383 | 0.3 | 41 |
| | | 7,319 | Hospital care | 17.9 | 2,453 | 6.0 | 821 |
| | | 7,319 | Filled prescriptions | 2.1 | 281 | 0.5 | 73 |
| | | 7,240 | Lost productivity (income loss) | . | . | 68.0 | 9,386 |
| Young adults and adults (25-64 years) | Pooled | 7,363 | Primary sector | 2.5 | 340 | 0.2 | 30 |
| | | 7,363 | Hospital care | 17.2 | 2,350 | 2.8 | 377 |
| | | 7,363 | Filled prescriptions | 2.1 | 289 | 0.4 | 55 |
| | | 5,265 | Lost productivity (income loss) | . | . | 34.1 | 6,471 |
| Older people (65+ years) | Pooled | 874 | Primary sector | 0.4 | 426 | 0.1 | 65 |
| | | 874 | Hospital care | 2.8 | 3,269 | 0.3 | 397 |
| | | 874 | Filled prescriptions | 0.4 | 452 | 0.1 | 107 |
| | | 477 | Lost productivity (income loss) | . | . | -2.9 | -6,153 |

#### Supplementary Table 107
#### Drug use disorder - Incident cohort 2016-2021
#### Cost analysis - not pooled

| Age strata of index patients | Closest relative type | Number of index patients included in cost component calculations | Cost component | Actual costs: Total (million EUR) | Actual costs: Per person (EUR) | Attributable costs: Total (million EUR) | Attributable costs: Per person (EUR) |
|---|---|---|---|---|---|---|---|
| Children and young people (0-24 years) | Fathers | 6,363 | Primary sector | 1.6 | 253 | 0.1 | 16 |
| | | 6,363 | Hospital care | 12.8 | 2,021 | 3.3 | 519 |
| | | 6,363 | Filled prescriptions | 1.7 | 273 | 0.5 | 73 |
| | | 6,072 | Lost productivity (income loss) | . | . | 109.2 | 17,978 |
| Children and young people (0-24 years) | Mothers | 7,319 | Primary sector | 2.8 | 383 | 0.3 | 41 |
| | | 7,319 | Hospital care | 17.9 | 2,453 | 6.0 | 821 |
| | | 7,319 | Filled prescriptions | 2.1 | 281 | 0.5 | 73 |
| | | 7,240 | Lost productivity (income loss) | . | . | 68.0 | 9,386 |
| Young adults and adults (25-64 years) | Children | 1,244 | Primary sector | 0.2 | 201 | 0.0 | 10 |
| | | 1,244 | Hospital care | 1.3 | 1,059 | 0.3 | 223 |
| | | 1,244 | Filled prescriptions | 0.1 | 104 | 0.0 | 27 |
| | | 513 | Lost productivity (income loss) | . | . | 0.8 | 1,469 |
| Young adults and adults (25-64 years) | Parents | 3,108 | Primary sector | 1.3 | 421 | 0.1 | 38 |
| | | 3,108 | Hospital care | 9.8 | 3,191 | 1.7 | 561 |
| | | 3,108 | Filled prescriptions | 1.4 | 446 | 0.2 | 80 |
| | | 1,902 | Lost productivity (income loss) | . | . | 10.0 | 5,251 |
| Young adults and adults (25-64 years) | Partners | 2,162 | Primary sector | 0.7 | 336 | 0.1 | 45 |
| | | 2,162 | Hospital care | 4.6 | 2,133 | 0.6 | 270 |
| | | 2,162 | Filled prescriptions | 0.4 | 204 | 0.1 | 60 |
| | | 2,070 | Lost productivity (income loss) | . | . | 20.6 | 9,967 |
| Young adults and adults (25-64 years) | Siblings | 849 | Primary sector | 0.2 | 258 | -0.0 | -11 |
| | | 849 | Hospital care | 1.5 | 1,727 | 0.2 | 206 |
| | | 849 | Filled prescriptions | 0.2 | 201 | -0.0 | -4 |
| | | 780 | Lost productivity (income loss) | . | . | 2.7 | 3,460 |
| Older people (65+ years) | Children | 482 | Primary sector | 0.2 | 324 | 0.0 | 46 |
| | | 482 | Hospital care | 0.9 | 1,932 | -0.0 | -56 |
| | | 482 | Filled prescriptions | 0.1 | 296 | 0.0 | 59 |
| | | 435 | Lost productivity (income loss) | . | . | -0.8 | -1,802 |
| Older people (65+ years) | Partners | 377 | Primary sector | 0.2 | 561 | 0.0 | 88 |
| | | 377 | Hospital care | 1.9 | 5,036 | 0.3 | 922 |
| | | 377 | Filled prescriptions | 0.2 | 654 | 0.1 | 163 |
| | | 29 | Lost productivity (income loss) | . | . | -1.3 | -46,339 |
| Older people (65+ years) | Siblings | 15 | Primary sector | 0.0 | 380 | 0.0 | 89 |
| | | 15 | Hospital care | 0.0 | 2,210 | 0.0 | 1,870 |
| | | 15 | Filled prescriptions | 0.0 | 402 | 0.0 | 277 |
| | | 13 | Lost productivity (income loss) | . | . | -0.8 | -62,068 |

**Supplementary Table 108**
**Drug use disorder - Prevalent cohort 2021**
**Identification of populations of relatives**

| *Agegroup of index patients in matched population* | | *Closest relative type identified* | | *Available control with same type of closest relative* |
|---|---|---|---|---|
| Children and young people (0-24 years) | n = 7,149 | Fathers | n = 5,459 (76%) | 95% |
| | | Mothers | n = 6,326 (88%) | 95% |
| | | No available closest relative | n = 415 (6%) | n/a |
| Young adults and adults (25-64 years) | n = 43,933 | Children | n = 6,761 (15%) | 91% |
| | | Parents | n = 14,574 (33%) | 90% |
| | | Partners | n = 10,461 (24%) | 93% |
| | | Siblings | n = 10,517 (24%) | 38% |
| | | Pooled | n = 42,313 (96%) | 78% |
| | | No available closest relative | n = 1,620 (4%) | n/a |
| Older people (65+ years) | n = 4,067 | Children | n = 1,914 (47%) | 92% |
| | | Partners | n = 1,153 (28%) | 94% |
| | | Siblings | n = 333 (8%) | 48% |
| | | Pooled | n = 3,400 (84%) | 88% |
| | | No available closest relative | n = 667 (16%) | n/a |

**Supplementary Table 109**
**Drug use disorder - Prevalent cohort 2021**
**Patient characteristics - cases**

| ***Age strata of index patients*** | ***Characteristic*** | |
|---|---|---|
| Children and young people (0-24 years) | Number of persons, n | 6,458 (100.0) |
| | Male, n (%) | 4,005 (62.0) |
| | Age, median (Q1;Q3) | 22.2 (20.1;23.7) |
| | CCI, n (%) | |
| | 0 | 5,947 (92.1) |
| | 1-2 | 486 (7.5) |
| | +3 | 25 (0.4) |
| | Education level, n (%) | |
| | No/Unknown education | 110 (1.7) |
| | Primary | 5,322 (82.4) |
| | Secondary | 984 (15.2) |
| | Tertiary | 42 (0.7) |
| Young adults and adults (25-64 years) | Number of persons, n | 33,371 (100.0) |
| | Male, n (%) | 22,641 (67.8) |
| | Age, median (Q1;Q3) | 38.1 (31.0;48.4) |
| | CCI, n (%) | |
| | 0 | 26,226 (78.6) |
| | 1-2 | 5,981 (17.9) |
| | +3 | 1,164 (3.5) |
| | Education level, n (%) | |
| | No/Unknown education | 754 (2.3) |
| | Primary | 17,848 (53.5) |
| | Secondary | 11,501 (34.5) |
| | Tertiary | 3,268 (9.8) |
| Older people (65+ years) | Number of persons, n | 3,016 (100.0) |
| | Male, n (%) | 1,056 (35.0) |
| | Age, median (Q1;Q3) | 72.5 (68.1;78.1) |
| | CCI, n (%) | |
| | 0 | 1,128 (37.4) |
| | 1-2 | 1,211 (40.2) |
| | +3 | 677 (22.4) |
| | Education level, n (%) | |
| | No/Unknown education | 175 (5.8) |
| | Primary | 1,081 (35.8) |
| | Secondary | 1,241 (41.1) |
| | Tertiary | 519 (17.2) |

**Supplementary Table 110**
**Drug use disorder - Prevalent cohort 2021**
**Patient characteristics - relatives - pooled**

| *Age strata of index patients* | *Closest relative type* | *Characteristic of family relatives by agestrata of index patients* | *Closest family relatives of brain disorder patients* | *Comparison relative* |
|---|---|---|---|---|
| Children and young people (0-24 years) | Fathers | Number of persons, n | 5,223 (100.0) | 5,223 (100.0) |
| | | Male, n (%) | 5,223 (100.0) | 5,223 (100.0) |
| | | Age, median (Q1;Q3) | 53.0 (48.9;56.9) | 53.1 (49.4;56.7) |
| | | CCI, n (%) | | |
| | | 0 | 4,247 (81.3) | 4,435 (84.9) |
| | | 1-2 | 837 (16.0) | 686 (13.1) |
| | | +3 | 139 (2.7) | 102 (2.0) |
| | | Education level, n (%) | | |
| | | No/Unknown education | 70 (1.3) | 70 (1.3) |
| | | Primary | 1,551 (29.7) | 1,551 (29.7) |
| | | Secondary | 2,666 (51.0) | 2,666 (51.0) |
| | | Tertiary | 936 (17.9) | 936 (17.9) |
| | | Relation to patient, n (%) | | |
| | | Father | 5,223 (100.0) | 5,223 (100.0) |
| Children and young people (0-24 years) | Mothers | Number of persons, n | 6,067 (100.0) | 6,067 (100.0) |
| | | Male, n (%) | 0 (0.0) | 0 (0.0) |
| | | Age, median (Q1;Q3) | 50.2 (46.2;54.4) | 50.5 (47.0;54.0) |
| | | CCI, n (%) | | |
| | | 0 | 4,913 (81.0) | 5,163 (85.1) |
| | | 1-2 | 1,043 (17.2) | 824 (13.6) |
| | | +3 | 111 (1.8) | 80 (1.3) |
| | | Education level, n (%) | | |
| | | No/Unknown education | 50 (0.8) | 50 (0.8) |
| | | Primary | 1,629 (26.9) | 1,629 (26.9) |
| | | Secondary | 2,788 (46.0) | 2,788 (46.0) |
| | | Tertiary | 1,600 (26.4) | 1,600 (26.4) |
| | | Relation to patient, n (%) | | |
| | | Mother | 6,067 (100.0) | 6,067 (100.0) |
| Young adults and adults (25-64 years) | Pooled | Number of persons, n | 33,371 (100.0) | 33,371 (100.0) |
| | | Male, n (%) | 12,471 (37.4) | 12,471 (37.4) |
| | | Age, median (Q1;Q3) | 49.3 (29.8;61.0) | 49.8 (29.9;61.1) |
| | | CCI, n (%) | | |
| | | 0 | 26,759 (80.2) | 27,893 (83.6) |
| | | 1-2 | 5,411 (16.2) | 4,636 (13.9) |
| | | +3 | 1,201 (3.6) | 842 (2.5) |
| | | Education level, n (%) | | |
| | | No/Unknown education | 2,823 (8.5) | 2,823 (8.5) |
| | | Primary | 10,830 (32.5) | 10,830 (32.5) |
| | | Secondary | 14,102 (42.3) | 14,102 (42.3) |
| | | Tertiary | 5,616 (16.8) | 5,616 (16.8) |
| | | Relation to patient, n (%) | | |
| | | Mother | 10,125 (30.3) | 10,125 (30.3) |
| | | Father | 3,119 (9.3) | 3,119 (9.3) |
| | | Partner | 9,817 (29.4) | 9,817 (29.4) |
| | | Child | 6,212 (18.6) | 6,212 (18.6) |

#### Supplementary Table 110
#### Drug use disorder - Prevalent cohort 2021
#### Patient characteristics - relatives - pooled

| *Age strata of index patients* | *Closest relative type* | *Characteristic of family relatives by agestrata of index patients* | *Closest family relatives of brain disorder patients* | *Comparison relative* |
|---|---|---|---|---|
| | | Sibling | 4,098 (12.3) | 4,098 (12.3) |
| Older people (65+ years) | Pooled | Number of persons, n | 3,016 (100.0) | 3,016 (100.0) |
| | | Male, n (%) | 1,577 (52.3) | 1,577 (52.3) |
| | | Age, median (Q1;Q3) | 57.8 (47.0;69.3) | 58.0 (46.8;68.9) |
| | | CCI, n (%) | | |
| | | 0 | 2,260 (74.9) | 2,328 (77.2) |
| | | 1-2 | 571 (18.9) | 557 (18.5) |
| | | +3 | 185 (6.1) | 131 (4.3) |
| | | Education level, n (%) | | |
| | | No/Unknown education | 72 (2.4) | 72 (2.4) |
| | | Primary | 636 (21.1) | 636 (21.1) |
| | | Secondary | 1,471 (48.8) | 1,471 (48.8) |
| | | Tertiary | 837 (27.8) | 837 (27.8) |
| | | Relation to patient, n (%) | | |
| | | Partner | 1,085 (36.0) | 1,085 (36.0) |
| | | Child | 1,769 (58.7) | 1,769 (58.7) |
| | | Sibling | 162 (5.4) | 162 (5.4) |

**Supplementary Table 111**
**Drug use disorder - Prevalent cohort 2021**
**Patient characteristics - relatives - not pooled**

| *Age strata of index patients* | *Closest relative type* | *Characteristic of family relatives by agestrata of index patients* | *Closest family relatives of brain disorder patients* | *Comparison relative* |
|---|---|---|---|---|
| Children and young people (0-24 years) | Fathers | Number of persons, n | 5,223 (100.0) | 5,223 (100.0) |
| | | Male, n (%) | 5,223 (100.0) | 5,223 (100.0) |
| | | Age, median (Q1;Q3) | 53.0 (48.9;56.9) | 53.1 (49.4;56.7) |
| | | CCI, n (%) | | |
| | | 0 | 4,247 (81.3) | 4,435 (84.9) |
| | | 1-2 | 837 (16.0) | 686 (13.1) |
| | | +3 | 139 (2.7) | 102 (2.0) |
| | | Education level, n (%) | | |
| | | No/Unknown education | 70 (1.3) | 70 (1.3) |
| | | Primary | 1,551 (29.7) | 1,551 (29.7) |
| | | Secondary | 2,666 (51.0) | 2,666 (51.0) |
| | | Tertiary | 936 (17.9) | 936 (17.9) |
| | | Relation to patient, n (%) | | |
| | | Father | 5,223 (100.0) | 5,223 (100.0) |
| Children and young people (0-24 years) | Mothers | Number of persons, n | 6,067 (100.0) | 6,067 (100.0) |
| | | Male, n (%) | 0 (0.0) | 0 (0.0) |
| | | Age, median (Q1;Q3) | 50.2 (46.2;54.4) | 50.5 (47.0;54.0) |
| | | CCI, n (%) | | |
| | | 0 | 4,913 (81.0) | 5,163 (85.1) |
| | | 1-2 | 1,043 (17.2) | 824 (13.6) |
| | | +3 | 111 (1.8) | 80 (1.3) |
| | | Education level, n (%) | | |
| | | No/Unknown education | 50 (0.8) | 50 (0.8) |
| | | Primary | 1,629 (26.9) | 1,629 (26.9) |
| | | Secondary | 2,788 (46.0) | 2,788 (46.0) |
| | | Tertiary | 1,600 (26.4) | 1,600 (26.4) |
| | | Relation to patient, n (%) | | |
| | | Mother | 6,067 (100.0) | 6,067 (100.0) |
| Young adults and adults (25-64 years) | Children | Number of persons, n | 6,212 (100.0) | 6,212 (100.0) |
| | | Male, n (%) | 3,240 (52.2) | 3,240 (52.2) |
| | | Age, median (Q1;Q3) | 16.9 (9.6;25.9) | 16.6 (8.5;24.8) |
| | | CCI, n (%) | | |
| | | 0 | 5,781 (93.1) | 5,875 (94.6) |
| | | 1-2 | 422 (6.8) | 324 (5.2) |
| | | +3 | 9 (0.1) | 13 (0.2) |
| | | Education level, n (%) | | |
| | | No/Unknown education | 2,358 (38.0) | 2,358 (38.0) |
| | | Primary | 2,395 (38.6) | 2,395 (38.6) |
| | | Secondary | 1,093 (17.6) | 1,093 (17.6) |
| | | Tertiary | 366 (5.9) | 366 (5.9) |
| | | Relation to patient, n (%) | | |
| | | Child | 6,212 (100.0) | 6,212 (100.0) |
| Young adults and adults (25-64 years) | Parents | Number of persons, n | 13,244 (100.0) | 13,244 (100.0) |
| | | Male, n (%) | 3,119 (23.6) | 3,119 (23.6) |
| | | Age, median (Q1;Q3) | 62.5 (56.8;69.6) | 62.5 (57.2;69.8) |

### Supplementary Table 111
### Drug use disorder - Prevalent cohort 2021
### Patient characteristics - relatives - not pooled

| *Age strata of index patients* | *Closest relative type* | *Characteristic of family relatives by agestrata of index patients* | *Closest family relatives of brain disorder patients* | *Comparison relative* |
|---|---|---|---|---|
| | | CCI, n (%) | | |
| | | 0 | 8,909 (67.3) | 9,618 (72.6) |
| | | 1-2 | 3,382 (25.5) | 2,981 (22.5) |
| | | +3 | 953 (7.2) | 645 (4.9) |
| | | Education level, n (%) | | |
| | | No/Unknown education | 221 (1.7) | 221 (1.7) |
| | | Primary | 4,162 (31.4) | 4,162 (31.4) |
| | | Secondary | 6,362 (48.0) | 6,362 (48.0) |
| | | Tertiary | 2,499 (18.9) | 2,499 (18.9) |
| | | Relation to patient, n (%) | | |
| | | Mother | 10,125 (76.4) | 10,125 (76.4) |
| | | Father | 3,119 (23.6) | 3,119 (23.6) |
| Young adults and adults (25-64 years) | Partners | Number of persons, n | 9,817 (100.0) | 9,817 (100.0) |
| | | Male, n (%) | 3,470 (35.3) | 3,470 (35.3) |
| | | Age, median (Q1;Q3) | 35.8 (29.8;45.9) | 35.9 (30.1;45.8) |
| | | CCI, n (%) | | |
| | | 0 | 8,655 (88.2) | 8,896 (90.6) |
| | | 1-2 | 1,029 (10.5) | 835 (8.5) |
| | | +3 | 133 (1.4) | 86 (0.9) |
| | | Education level, n (%) | | |
| | | No/Unknown education | 233 (2.4) | 233 (2.4) |
| | | Primary | 2,930 (29.8) | 2,930 (29.8) |
| | | Secondary | 4,601 (46.9) | 4,601 (46.9) |
| | | Tertiary | 2,053 (20.9) | 2,053 (20.9) |
| | | Relation to patient, n (%) | | |
| | | Partner | 9,817 (100.0) | 9,817 (100.0) |
| Young adults and adults (25-64 years) | Siblings | Number of persons, n | 4,098 (100.0) | 4,098 (100.0) |
| | | Male, n (%) | 2,642 (64.5) | 2,642 (64.5) |
| | | Age, median (Q1;Q3) | 48.1 (33.3;57.4) | 48.5 (33.2;57.5) |
| | | CCI, n (%) | | |
| | | 0 | 3,414 (83.3) | 3,504 (85.5) |
| | | 1-2 | 578 (14.1) | 496 (12.1) |
| | | +3 | 106 (2.6) | 98 (2.4) |
| | | Education level, n (%) | | |
| | | No/Unknown education | 11 (0.3) | 11 (0.3) |
| | | Primary | 1,343 (32.8) | 1,343 (32.8) |
| | | Secondary | 2,046 (49.9) | 2,046 (49.9) |
| | | Tertiary | 698 (17.0) | 698 (17.0) |
| | | Relation to patient, n (%) | | |
| | | Sibling | 4,098 (100.0) | 4,098 (100.0) |
| Older people (65+ years) | Children | Number of persons, n | 1,769 (100.0) | 1,769 (100.0) |
| | | Male, n (%) | 870 (49.2) | 870 (49.2) |
| | | Age, median (Q1;Q3) | 48.6 (43.0;55.6) | 48.5 (42.7;55.2) |
| | | CCI, n (%) | | |
| | | 0 | 1,535 (86.8) | 1,545 (87.3) |

#### Supplementary Table 111
#### Drug use disorder - Prevalent cohort 2021
#### Patient characteristics - relatives - not pooled

| *Age strata of index patients* | *Closest relative type* | *Characteristic of family relatives by agestrata of index patients* | *Closest family relatives of brain disorder patients* | *Comparison relative* |
|---|---|---|---|---|
| | | 1-2 | 191 (10.8) | 199 (11.2) |
| | | +3 | 43 (2.4) | 25 (1.4) |
| | | Education level, n (%) | | |
| | | No/Unknown education | * | * |
| | | Primary | * | * |
| | | Secondary | 836 (47.3) | 836 (47.3) |
| | | Tertiary | 598 (33.8) | 598 (33.8) |
| | | Relation to patient, n (%) | | |
| | | Child | 1,769 (100.0) | 1,769 (100.0) |
| Older people (65+ years) | Partners | Number of persons, n | 1,085 (100.0) | 1,085 (100.0) |
| | | Male, n (%) | 628 (57.9) | 628 (57.9) |
| | | Age, median (Q1;Q3) | 72.4 (67.5;77.5) | 72.5 (67.5;77.5) |
| | | CCI, n (%) | | |
| | | 0 | 606 (55.9) | 664 (61.2) |
| | | 1-2 | 349 (32.2) | 322 (29.7) |
| | | +3 | 130 (12.0) | 99 (9.1) |
| | | Education level, n (%) | | |
| | | No/Unknown education | 69 (6.4) | 69 (6.4) |
| | | Primary | 277 (25.5) | 277 (25.5) |
| | | Secondary | 541 (49.9) | 541 (49.9) |
| | | Tertiary | 198 (18.2) | 198 (18.2) |
| | | Relation to patient, n (%) | | |
| | | Partner | 1,085 (100.0) | 1,085 (100.0) |
| Older people (65+ years) | Siblings | Number of persons, n | 162 (100.0) | 162 (100.0) |
| | | Male, n (%) | 79 (48.8) | 79 (48.8) |
| | | Age, median (Q1;Q3) | 62.6 (59.4;65.1) | 62.4 (60.0;64.9) |
| | | CCI, n (%) | | |
| | | 0 | 119 (73.5) | 119 (73.5) |
| | | 1-2 | 31 (19.1) | 36 (22.2) |
| | | +3 | 12 (7.4) | 7 (4.3) |
| | | Education level, n (%) | | |
| | | No/Unknown education | * | * |
| | | Primary | * | * |
| | | Secondary | 94 (58.0) | 94 (58.0) |
| | | Tertiary | 41 (25.3) | 41 (25.3) |
| | | Relation to patient, n (%) | | |
| | | Sibling | 162 (100.0) | 162 (100.0) |

#### Supplementary Table 112
#### Drug use disorder - Prevalent cohort 2021
#### Cost analysis - pooled

| Age strata of index patients | Closest relative type | Number of index patients included in cost component calculations | Cost component | Actual costs Total (million EUR) | Actual costs Per person (EUR) | Attributable costs Total (million EUR) | Attributable costs Per person (EUR) |
|---|---|---|---|---|---|---|---|
| Children and young people (0-24 years) | Fathers | 5,223 | Primary sector | 1.4 | 264 | 0.1 | 23 |
| | | 5,223 | Hospital care | 12.0 | 2,300 | 4.0 | 772 |
| | | 5,223 | Filled prescriptions | 1.6 | 314 | 0.6 | 112 |
| | | 4,927 | Lost productivity (income loss) | . | . | 85.8 | 17,420 |
| Children and young people (0-24 years) | Mothers | 6,067 | Primary sector | 2.4 | 400 | 0.3 | 46 |
| | | 6,067 | Hospital care | 14.6 | 2,403 | 4.0 | 662 |
| | | 6,067 | Filled prescriptions | 2.0 | 324 | 0.5 | 87 |
| | | 6,007 | Lost productivity (income loss) | . | . | 56.4 | 9,392 |
| Young adults and adults (25-64 years) | Pooled | 33,371 | Primary sector | 11.4 | 345 | 1.0 | 30 |
| | | 33,371 | Hospital care | 83.6 | 2,519 | 18.8 | 567 |
| | | 33,371 | Filled prescriptions | 10.9 | 329 | 2.2 | 65 |
| | | 22,517 | Lost productivity (income loss) | . | . | 192.4 | 8,546 |
| Older people (65+ years) | Pooled | 3,016 | Primary sector | 1.2 | 397 | 0.1 | 30 |
| | | 3,016 | Hospital care | 9.5 | 3,181 | 1.8 | 604 |
| | | 3,016 | Filled prescriptions | 1.3 | 437 | 0.3 | 108 |
| | | 1,843 | Lost productivity (income loss) | . | . | 8.0 | 4,343 |

#### Supplementary Table 113
#### Drug use disorder - Prevalent cohort 2021
#### Cost analysis - not pooled

| Age strata of index patients | Closest relative type | Number of index patients included in cost component calculations | Cost component | Actual costs: Total (million EUR) | Actual costs: Per person (EUR) | Attributable costs: Total (million EUR) | Attributable costs: Per person (EUR) |
|---|---|---|---|---|---|---|---|
| Children and young people (0-24 years) | Fathers | 5,223 | Primary sector | 1.4 | 264 | 0.1 | 23 |
| | | 5,223 | Hospital care | 12.0 | 2,300 | 4.0 | 772 |
| | | 5,223 | Filled prescriptions | 1.6 | 314 | 0.6 | 112 |
| | | 4,927 | Lost productivity (income loss) | . | . | 85.8 | 17,420 |
| Children and young people (0-24 years) | Mothers | 6,067 | Primary sector | 2.4 | 400 | 0.3 | 46 |
| | | 6,067 | Hospital care | 14.6 | 2,403 | 4.0 | 662 |
| | | 6,067 | Filled prescriptions | 2.0 | 324 | 0.5 | 87 |
| | | 6,007 | Lost productivity (income loss) | . | . | 56.4 | 9,392 |
| Young adults and adults (25-64 years) | Children | 6,212 | Primary sector | 1.3 | 216 | 0.2 | 34 |
| | | 6,212 | Hospital care | 7.1 | 1,143 | 1.4 | 219 |
| | | 6,212 | Filled prescriptions | 0.9 | 139 | 0.3 | 51 |
| | | 2,579 | Lost productivity (income loss) | . | . | 11.3 | 4,368 |
| Young adults and adults (25-64 years) | Parents | 13,244 | Primary sector | 5.8 | 445 | 0.4 | 33 |
| | | 13,244 | Hospital care | 45.9 | 3,499 | 11.1 | 847 |
| | | 13,244 | Filled prescriptions | 6.5 | 495 | 1.0 | 79 |
| | | 6,762 | Lost productivity (income loss) | . | . | 53.3 | 7,888 |
| Young adults and adults (25-64 years) | Partners | 9,817 | Primary sector | 3.1 | 312 | 0.2 | 25 |
| | | 9,817 | Hospital care | 21.4 | 2,186 | 5.1 | 519 |
| | | 9,817 | Filled prescriptions | 2.4 | 247 | 0.6 | 63 |
| | | 9,496 | Lost productivity (income loss) | . | . | 103.2 | 10,866 |
| Young adults and adults (25-64 years) | Siblings | 4,098 | Primary sector | 1.2 | 299 | 0.1 | 24 |
| | | 4,098 | Hospital care | 9.2 | 2,263 | 1.3 | 308 |
| | | 4,098 | Filled prescriptions | 1.1 | 279 | 0.2 | 49 |
| | | 3,680 | Lost productivity (income loss) | . | . | 24.6 | 6,693 |
| Older people (65+ years) | Children | 1,769 | Primary sector | 0.6 | 329 | 0.1 | 32 |
| | | 1,769 | Hospital care | 4.0 | 2,246 | 0.5 | 273 |
| | | 1,769 | Filled prescriptions | 0.5 | 280 | 0.1 | 58 |
| | | 1,666 | Lost productivity (income loss) | . | . | 8.1 | 4,851 |
| Older people (65+ years) | Partners | 1,085 | Primary sector | 0.5 | 508 | 0.0 | 30 |
| | | 1,085 | Hospital care | 4.9 | 4,602 | 1.0 | 905 |
| | | 1,085 | Filled prescriptions | 0.7 | 688 | 0.2 | 186 |
| | | 87 | Lost productivity (income loss) | . | . | 0.7 | 7,696 |
| Older people (65+ years) | Siblings | 162 | Primary sector | 0.1 | 409 | 0.0 | 13 |
| | | 162 | Hospital care | 0.6 | 3,994 | 0.4 | 2,227 |
| | | 162 | Filled prescriptions | 0.1 | 493 | 0.0 | 146 |
| | | 90 | Lost productivity (income loss) | . | . | -0.7 | -8,312 |

**Supplementary Table 114**
**Eating disorders - Incident cohort 2016-2021**
**Identification of populations of relatives**

| *Agegroup of index patients in matched population* | | *Closest relative type identified* | | *Available control with same type of closest relative* |
|---|---|---|---|---|
| Children and young people (0-24 years) | n = 5,098 | Fathers | n = 4,481 (88%) | 97% |
| | | Mothers | n = 4,686 (92%) | 97% |
| | | No available closest relative | n = 205 (4%) | n/a |
| Young adults and adults (25-64 years) | n = 1,372 | Children | n = 134 (10%) | 97% |
| | | Parents | n = 432 (31%) | 94% |
| | | Partners | n = 513 (37%) | 96% |
| | | Siblings | n = 233 (17%) | 31% |
| | | Pooled | n = 1,312 (96%) | 84% |
| | | No available closest relative | n = 60 (4%) | n/a |
| Older people (65+ years) | n = 25 | Children | n/a | n/a |
| | | Partners | n/a | n/a |
| | | Siblings | n/a | n/a |
| | | Pooled | n/a | n/a |
| | | No available closest relative | n/a | n/a |

#### Supplementary Table 115
#### Eating disorders - Incident cohort 2016-2021
#### Patient characteristics - cases

| *Age strata of index patients* | *Characteristic* | |
|---|---|---|
| Children and young people (0-24 years) | Number of persons, n | 4,767 (100.0) |
| | Male, n (%) | 305 (6.4) |
| | Age, median (Q1;Q3) | 16.9 (14.6;20.3) |
| | CCI, n (%) | |
| | 0 | 4,477 (93.9) |
| | 1-2 | 284 (6.0) |
| | +3 | 6 (0.1) |
| | Education level, n (%) | |
| | No/Unknown education | 451 (9.5) |
| | Primary | 3,342 (70.1) |
| | Secondary | 886 (18.6) |
| | Tertiary | 88 (1.8) |
| Young adults and adults (25-64 years) | Number of persons, n | 1,108 (100.0) |
| | Male, n (%) | 60 (5.4) |
| | Age, median (Q1;Q3) | 31.2 (27.2;38.7) |
| | CCI, n (%) | |
| | 0 | 979 (88.4) |
| | 1-2 | 121 (10.9) |
| | +3 | 8 (0.7) |
| | Education level, n (%) | |
| | No/Unknown education | 26 (2.3) |
| | Primary | 206 (18.6) |
| | Secondary | 450 (40.6) |
| | Tertiary | 426 (38.4) |
| Older people (65+ years) | Number of persons, n | 23 (100.0) |
| | Male, n (%) | * |
| | Age, median (Q1;Q3) | 68.5 (66.4;76.2) |
| | CCI, n (%) | |
| | 0 | * |
| | 1-2 | 11 (47.8) |
| | +3 | * |
| | Education level, n (%) | |
| | No/Unknown education | * |
| | Primary | * |
| | Secondary | * |
| | Tertiary | * |

#### Supplementary Table 116
#### Eating disorders - Incident cohort 2016-2021
#### Patient characteristics - relatives - pooled

| *Age strata of index patients* | *Closest relative type* | *Characteristic of family relatives by agestrata of index patients* | *Closest family relatives of brain disorder patients* | *Comparison relative* |
|---|---|---|---|---|
| Children and young people (0-24 years) | Fathers | Number of persons, n | 4,367 (100.0) | 4,367 (100.0) |
| | | Male, n (%) | 4,367 (100.0) | 4,367 (100.0) |
| | | Age, median (Q1;Q3) | 50.2 (46.3;54.4) | 50.1 (46.4;54.1) |
| | | CCI, n (%) | | |
| | | 0 | 3,805 (87.1) | 3,833 (87.8) |
| | | 1-2 | 496 (11.4) | 481 (11.0) |
| | | +3 | 66 (1.5) | 53 (1.2) |
| | | Education level, n (%) | | |
| | | No/Unknown education | 47 (1.1) | 47 (1.1) |
| | | Primary | 557 (12.8) | 557 (12.8) |
| | | Secondary | 2,282 (52.3) | 2,282 (52.3) |
| | | Tertiary | 1,481 (33.9) | 1,481 (33.9) |
| | | Relation to patient, n (%) | | |
| | | Father | 4,367 (100.0) | 4,367 (100.0) |
| Children and young people (0-24 years) | Mothers | Number of persons, n | 4,564 (100.0) | 4,564 (100.0) |
| | | Male, n (%) | 0 (0.0) | 0 (0.0) |
| | | Age, median (Q1;Q3) | 48.0 (44.4;51.7) | 47.9 (44.4;51.5) |
| | | CCI, n (%) | | |
| | | 0 | 3,990 (87.4) | 4,040 (88.5) |
| | | 1-2 | 533 (11.7) | 483 (10.6) |
| | | +3 | 41 (0.9) | 41 (0.9) |
| | | Education level, n (%) | | |
| | | No/Unknown education | 31 (0.7) | 31 (0.7) |
| | | Primary | 528 (11.6) | 528 (11.6) |
| | | Secondary | 1,934 (42.4) | 1,934 (42.4) |
| | | Tertiary | 2,071 (45.4) | 2,071 (45.4) |
| | | Relation to patient, n (%) | | |
| | | Mother | 4,564 (100.0) | 4,564 (100.0) |
| Young adults and adults (25-64 years) | Pooled | Number of persons, n | 1,108 (100.0) | 1,108 (100.0) |
| | | Male, n (%) | 660 (59.6) | 660 (59.6) |
| | | Age, median (Q1;Q3) | 41.8 (29.6;56.8) | 41.0 (29.7;56.6) |
| | | CCI, n (%) | | |
| | | 0 | 962 (86.8) | 974 (87.9) |
| | | 1-2 | 123 (11.1) | 115 (10.4) |
| | | +3 | 23 (2.1) | 19 (1.7) |
| | | Education level, n (%) | | |
| | | No/Unknown education | 81 (7.3) | 81 (7.3) |
| | | Primary | 240 (21.7) | 240 (21.7) |
| | | Secondary | 473 (42.7) | 473 (42.7) |
| | | Tertiary | 314 (28.3) | 314 (28.3) |
| | | Relation to patient, n (%) | | |
| | | Mother | 327 (29.5) | 327 (29.5) |
| | | Father | 81 (7.3) | 81 (7.3) |
| | | Partner | 495 (44.7) | 495 (44.7) |
| | | Child | 131 (11.8) | 131 (11.8) |

#### Supplementary Table 116
#### Eating disorders - Incident cohort 2016-2021
#### Patient characteristics - relatives - pooled

| *Age strata of index patients* | *Closest relative type* | *Characteristic of family relatives by agestrata of index patients* | *Closest family relatives of brain disorder patients* | *Comparison relative* |
|---|---|---|---|---|
| | | Sibling | 74 (6.7) | 74 (6.7) |
| Older people (65+ years) | Pooled | Number of persons, n | 23 (100.0) | 23 (100.0) |
| | | Male, n (%) | 12 (52.2) | 12 (52.2) |
| | | Age, median (Q1;Q3) | 57.1 (48.1;68.9) | 57.8 (45.4;69.9) |
| | | CCI, n (%) | | |
| | | 0 | 18 (78.3) | 18 (78.3) |
| | | 1-2 | 5 (21.7) | 5 (21.7) |
| | | +3 | 0 (0.0) | 0 (0.0) |
| | | Education level, n (%) | | |
| | | No/Unknown education | * | * |
| | | Primary | * | * |
| | | Secondary | * | * |
| | | Tertiary | * | * |
| | | Relation to patient, n (%) | | |
| | | Partner | * | * |
| | | Child | * | * |
| | | Sibling | * | * |

**Supplementary Table 117**
**Eating disorders - Incident cohort 2016-2021**
**Patient characteristics - relatives - not pooled**

| *Age strata of index patients* | *Closest relative type* | *Characteristic of family relatives by agestrata of index patients* | *Closest family relatives of brain disorder patients* | *Comparison relative* |
|---|---|---|---|---|
| Children and young people (0-24 years) | Fathers | Number of persons, n | 4,367 (100.0) | 4,367 (100.0) |
| | | Male, n (%) | 4,367 (100.0) | 4,367 (100.0) |
| | | Age, median (Q1;Q3) | 50.2 (46.3;54.4) | 50.1 (46.4;54.1) |
| | | CCI, n (%) | | |
| | | 0 | 3,805 (87.1) | 3,833 (87.8) |
| | | 1-2 | 496 (11.4) | 481 (11.0) |
| | | +3 | 66 (1.5) | 53 (1.2) |
| | | Education level, n (%) | | |
| | | No/Unknown education | 47 (1.1) | 47 (1.1) |
| | | Primary | 557 (12.8) | 557 (12.8) |
| | | Secondary | 2,282 (52.3) | 2,282 (52.3) |
| | | Tertiary | 1,481 (33.9) | 1,481 (33.9) |
| | | Relation to patient, n (%) | | |
| | | Father | 4,367 (100.0) | 4,367 (100.0) |
| Children and young people (0-24 years) | Mothers | Number of persons, n | 4,564 (100.0) | 4,564 (100.0) |
| | | Male, n (%) | 0 (0.0) | 0 (0.0) |
| | | Age, median (Q1;Q3) | 48.0 (44.4;51.7) | 47.9 (44.4;51.5) |
| | | CCI, n (%) | | |
| | | 0 | 3,990 (87.4) | 4,040 (88.5) |
| | | 1-2 | 533 (11.7) | 483 (10.6) |
| | | +3 | 41 (0.9) | 41 (0.9) |
| | | Education level, n (%) | | |
| | | No/Unknown education | 31 (0.7) | 31 (0.7) |
| | | Primary | 528 (11.6) | 528 (11.6) |
| | | Secondary | 1,934 (42.4) | 1,934 (42.4) |
| | | Tertiary | 2,071 (45.4) | 2,071 (45.4) |
| | | Relation to patient, n (%) | | |
| | | Mother | 4,564 (100.0) | 4,564 (100.0) |
| Young adults and adults (25-64 years) | Children | Number of persons, n | 131 (100.0) | 131 (100.0) |
| | | Male, n (%) | 59 (45.0) | 59 (45.0) |
| | | Age, median (Q1;Q3) | 14.1 (8.4;19.6) | 14.4 (7.1;19.0) |
| | | CCI, n (%) | | |
| | | 0 | 120 (91.6) | 124 (94.7) |
| | | 1-2 | 11 (8.4) | 7 (5.3) |
| | | +3 | 0 (0.0) | 0 (0.0) |
| | | Education level, n (%) | | |
| | | No/Unknown education | 64 (48.9) | 64 (48.9) |
| | | Primary | 48 (36.6) | 48 (36.6) |
| | | Secondary | 12 (9.2) | 12 (9.2) |
| | | Tertiary | 7 (5.3) | 7 (5.3) |
| | | Relation to patient, n (%) | | |
| | | Child | 131 (100.0) | 131 (100.0) |
| Young adults and adults (25-64 years) | Parents | Number of persons, n | 408 (100.0) | 408 (100.0) |
| | | Male, n (%) | 81 (19.9) | 81 (19.9) |
| | | Age, median (Q1;Q3) | 58.6 (54.3;63.8) | 58.5 (54.7;63.3) |

**Supplementary Table 117**
**Eating disorders - Incident cohort 2016-2021**
**Patient characteristics - relatives - not pooled**

| *Age strata of index patients* | *Closest relative type* | *Characteristic of family relatives by agestrata of index patients* | *Closest family relatives of brain disorder patients* | *Comparison relative* |
|---|---|---|---|---|
| | | CCI, n (%) | | |
| | | 0 | 313 (76.7) | 324 (79.4) |
| | | 1-2 | 77 (18.9) | 71 (17.4) |
| | | +3 | 18 (4.4) | 13 (3.2) |
| | | Education level, n (%) | | |
| | | No/Unknown education | 5 (1.2) | 5 (1.2) |
| | | Primary | 85 (20.8) | 85 (20.8) |
| | | Secondary | 196 (48.0) | 196 (48.0) |
| | | Tertiary | 122 (29.9) | 122 (29.9) |
| | | Relation to patient, n (%) | | |
| | | Mother | 327 (80.1) | 327 (80.1) |
| | | Father | 81 (19.9) | 81 (19.9) |
| Young adults and adults (25-64 years) | Partners | Number of persons, n | 495 (100.0) | 495 (100.0) |
| | | Male, n (%) | 477 (96.4) | 477 (96.4) |
| | | Age, median (Q1;Q3) | 35.0 (29.8;42.2) | 34.6 (29.9;41.8) |
| | | CCI, n (%) | | |
| | | 0 | 459 (92.7) | 458 (92.5) |
| | | 1-2 | 31 (6.3) | 32 (6.5) |
| | | +3 | 5 (1.0) | 5 (1.0) |
| | | Education level, n (%) | | |
| | | No/Unknown education | 12 (2.4) | 12 (2.4) |
| | | Primary | 89 (18.0) | 89 (18.0) |
| | | Secondary | 229 (46.3) | 229 (46.3) |
| | | Tertiary | 165 (33.3) | 165 (33.3) |
| | | Relation to patient, n (%) | | |
| | | Partner | 495 (100.0) | 495 (100.0) |
| Young adults and adults (25-64 years) | Siblings | Number of persons, n | 74 (100.0) | 74 (100.0) |
| | | Male, n (%) | 43 (58.1) | 43 (58.1) |
| | | Age, median (Q1;Q3) | 30.0 (24.0;38.1) | 29.3 (24.6;38.1) |
| | | CCI, n (%) | | |
| | | 0 | * | * |
| | | 1-2 | * | * |
| | | +3 | 0 (0.0) | * |
| | | Education level, n (%) | | |
| | | No/Unknown education | 0 (0.0) | 0 (0.0) |
| | | Primary | 18 (24.3) | 18 (24.3) |
| | | Secondary | 36 (48.6) | 36 (48.6) |
| | | Tertiary | 20 (27.0) | 20 (27.0) |
| | | Relation to patient, n (%) | | |
| | | Sibling | 74 (100.0) | 74 (100.0) |
| Older people (65+ years) | Children | Number of persons, n | 14 (100.0) | 14 (100.0) |
| | | Male, n (%) | * | * |
| | | Age, median (Q1;Q3) | 50.5 (46.5;57.0) | 48.1 (42.6;57.2) |
| | | CCI, n (%) | | |
| | | 0 | * | * |

#### Supplementary Table 117
#### Eating disorders - Incident cohort 2016-2021
#### Patient characteristics - relatives - not pooled

| *Age strata of index patients* | *Closest relative type* | *Characteristic of family relatives by agestrata of index patients* | *Closest family relatives of brain disorder patients* | *Comparison relative* |
|---|---|---|---|---|
| | | 1-2 | * | * |
| | | +3 | 0 (0.0) | 0 (0.0) |
| | | Education level, n (%) | | |
| | | No/Unknown education | * | * |
| | | Primary | * | * |
| | | Secondary | * | * |
| | | Tertiary | * | * |
| | | Relation to patient, n (%) | | |
| | | Child | 14 (100.0) | 14 (100.0) |
| Older people (65+ years) | Partners | Number of persons, n | 8 (100.0) | 8 (100.0) |
| | | Male, n (%) | * | * |
| | | Age, median (Q1;Q3) | 69.4 (68.0;76.6) | 72.5 (68.4;76.7) |
| | | CCI, n (%) | | |
| | | 0 | * | * |
| | | 1-2 | * | * |
| | | +3 | 0 (0.0) | 0 (0.0) |
| | | Education level, n (%) | | |
| | | No/Unknown education | 0 (0.0) | 0 (0.0) |
| | | Primary | * | * |
| | | Secondary | * | * |
| | | Tertiary | * | * |
| | | Relation to patient, n (%) | | |
| | | Partner | 8 (100.0) | 8 (100.0) |
| Older people (65+ years) | Siblings | Number of persons, n | * | * |
| | | Male, n (%) | * | * |
| | | Age, median (Q1;Q3) | 62.8 (62.8;62.8) | 63.2 (63.2;63.2) |
| | | CCI, n (%) | | |
| | | 0 | * | * |
| | | 1-2 | 0 (0.0) | 0 (0.0) |
| | | +3 | 0 (0.0) | 0 (0.0) |
| | | Education level, n (%) | | |
| | | No/Unknown education | 0 (0.0) | 0 (0.0) |
| | | Primary | 0 (0.0) | 0 (0.0) |
| | | Secondary | * | * |
| | | Tertiary | 0 (0.0) | 0 (0.0) |
| | | Relation to patient, n (%) | | |
| | | Sibling | 1 (100.0) | 1 (100.0) |

#### Supplementary Table 118
#### Eating disorders - Incident cohort 2016-2021
#### Cost analysis - pooled

| *Age strata of index patients* | *Closest relative type* | *Number of index patients included in cost component calculations* | *Cost component* | *Actual costs* | | *Attributable costs* | |
|---|---|---|---|---|---|---|---|
| | | | | *Total (million EUR)* | *Per person (EUR)* | *Total (million EUR)* | *Per person (EUR)* |
| Children and young people (0-24 years) | Fathers | 4,367 | Primary sector | 1.0 | 224 | -0.0 | -4 |
| | | 4,367 | Hospital care | 6.1 | 1,404 | -0.4 | -102 |
| | | 4,367 | Filled prescriptions | 0.8 | 184 | 0.1 | 17 |
| | | 4,220 | Lost productivity (income loss) | . | . | -16.2 | -3,836 |
| Children and young people (0-24 years) | Mothers | 4,564 | Primary sector | 1.6 | 341 | 0.1 | 15 |
| | | 4,564 | Hospital care | 7.4 | 1,631 | 0.5 | 116 |
| | | 4,564 | Filled prescriptions | 0.9 | 188 | 0.0 | 8 |
| | | 4,530 | Lost productivity (income loss) | . | . | -10.1 | -2,219 |
| Young adults and adults (25-64 years) | Pooled | 1,108 | Primary sector | 0.3 | 273 | 0.0 | 24 |
| | | 1,108 | Hospital care | 1.6 | 1,448 | -0.0 | -8 |
| | | 1,108 | Filled prescriptions | 0.2 | 216 | 0.0 | 34 |
| | | 870 | Lost productivity (income loss) | . | . | 1.0 | 1,166 |
| Older people (65+ years) | Pooled | 23 | Primary sector | 0.0 | 329 | -0.0 | -62 |
| | | 23 | Hospital care | 0.0 | 1,477 | -0.1 | -4,678 |
| | | 23 | Filled prescriptions | 0.0 | 407 | -0.0 | -65 |
| | | 14 | Lost productivity (income loss) | . | . | 0.1 | 9,564 |

#### Supplementary Table 119
#### Eating disorders - Incident cohort 2016-2021
#### Cost analysis - not pooled

| *Age strata of index patients* | *Closest relative type* | *Number of index patients included in cost component calculations* | *Cost component* | *Actual costs* | | *Attributable costs* | |
|---|---|---|---|---|---|---|---|
| | | | | *Total (million EUR)* | *Per person (EUR)* | *Total (million EUR)* | *Per person (EUR)* |
| Children and young people (0-24 years) | Fathers | 4,367 | Primary sector | 1.0 | 224 | -0.0 | -4 |
| | | 4,367 | Hospital care | 6.1 | 1,404 | -0.4 | -102 |
| | | 4,367 | Filled prescriptions | 0.8 | 184 | 0.1 | 17 |
| | | 4,220 | Lost productivity (income loss) | . | . | -16.2 | -3,836 |
| Children and young people (0-24 years) | Mothers | 4,564 | Primary sector | 1.6 | 341 | 0.1 | 15 |
| | | 4,564 | Hospital care | 7.4 | 1,631 | 0.5 | 116 |
| | | 4,564 | Filled prescriptions | 0.9 | 188 | 0.0 | 8 |
| | | 4,530 | Lost productivity (income loss) | . | . | -10.1 | -2,219 |
| Young adults and adults (25-64 years) | Children | 131 | Primary sector | 0.0 | 171 | -0.0 | -5 |
| | | 131 | Hospital care | 0.1 | 486 | -0.0 | -377 |
| | | 131 | Filled prescriptions | 0.0 | 82 | 0.0 | 10 |
| | | 33 | Lost productivity (income loss) | . | . | -0.1 | -2,672 |
| Young adults and adults (25-64 years) | Parents | 408 | Primary sector | 0.2 | 397 | 0.0 | 25 |
| | | 408 | Hospital care | 0.9 | 2,217 | 0.1 | 143 |
| | | 408 | Filled prescriptions | 0.2 | 384 | 0.0 | 86 |
| | | 276 | Lost productivity (income loss) | . | . | 2.1 | 7,776 |
| Young adults and adults (25-64 years) | Partners | 495 | Primary sector | 0.1 | 210 | 0.0 | 37 |
| | | 495 | Hospital care | 0.5 | 941 | -0.2 | -312 |
| | | 495 | Filled prescriptions | 0.0 | 96 | -0.0 | -16 |
| | | 489 | Lost productivity (income loss) | . | . | -0.6 | -1,283 |
| Young adults and adults (25-64 years) | Siblings | 74 | Primary sector | 0.0 | 195 | -0.0 | -25 |
| | | 74 | Hospital care | 0.2 | 2,300 | 0.1 | 1,837 |
| | | 74 | Filled prescriptions | 0.0 | 331 | 0.0 | 118 |
| | | 72 | Lost productivity (income loss) | . | . | -0.4 | -5,780 |
| Older people (65+ years) | Children | 14 | Primary sector | 0.0 | 347 | -0.0 | -48 |
| | | 14 | Hospital care | 0.0 | 1,850 | -0.1 | -6,020 |
| | | 14 | Filled prescriptions | 0.0 | 419 | 0.0 | 40 |
| | | 14 | Lost productivity (income loss) | . | . | 0.1 | 9,564 |
| Older people (65+ years) | Partners | 8 | Primary sector | 0.0 | 277 | -0.0 | -120 |
| | | 8 | Hospital care | 0.0 | 1,006 | -0.0 | -2,907 |
| | | 8 | Filled prescriptions | 0.0 | 438 | -0.0 | -258 |
| Older people (65+ years) | Siblings | . | Primary sector | . | . | . | . |
| | | . | Hospital care | . | . | . | . |
| | | . | Filled prescriptions | . | . | . | . |

**Supplementary Table 120**
**Eating disorders - Prevalent cohort 2021**
**Identification of populations of relatives**

| ***Agegroup of index patients in matched population*** | | ***Closest relative type identified*** | | ***Available control with same type of closest relative*** |
|---|---|---|---|---|
| Children and young people (0-24 years) | n = 5,748 | Fathers | n = 5,073 (88%) | 97% |
| | | Mothers | n = 5,324 (93%) | 97% |
| | | No available closest relative | n = 202 (4%) | n/a |
| Young adults and adults (25-64 years) | n = 11,904 | Children | n = 1,223 (10%) | 96% |
| | | Parents | n = 3,029 (25%) | 92% |
| | | Partners | n = 5,929 (50%) | 96% |
| | | Siblings | n = 1,597 (13%) | 30% |
| | | Pooled | n = 11,778 (99%) | 86% |
| | | No available closest relative | n = 126 (1%) | n/a |
| Older people (65+ years) | n = 128 | Children | n = 57 (45%) | 100% |
| | | Partners | n = 37 (29%) | 100% |
| | | Siblings | n = 13 (10%) | 69% |
| | | Pooled | n = 107 (84%) | 96% |
| | | No available closest relative | n = 21 (16%) | n/a |

**Supplementary Table 121**
**Eating disorders - Prevalent cohort 2021**
**Patient characteristics - cases**

| *Age strata of index patients* | *Characteristic* | |
|---|---|---|
| Children and young people (0-24 years) | Number of persons, n | 5,417 (100.0) |
| | Male, n (%) | 408 (7.5) |
| | Age, median (Q1;Q3) | 20.8 (17.8;23.0) |
| | CCI, n (%) | |
| | 0 | 5,080 (93.8) |
| | 1-2 | 324 (6.0) |
| | +3 | 13 (0.2) |
| | Education level, n (%) | |
| | No/Unknown education | 105 (1.9) |
| | Primary | 3,210 (59.3) |
| | Secondary | 1,901 (35.1) |
| | Tertiary | 201 (3.7) |
| Young adults and adults (25-64 years) | Number of persons, n | 10,199 (100.0) |
| | Male, n (%) | 403 (4.0) |
| | Age, median (Q1;Q3) | 33.8 (29.3;40.5) |
| | CCI, n (%) | |
| | 0 | 8,987 (88.1) |
| | 1-2 | 1,103 (10.8) |
| | +3 | 109 (1.1) |
| | Education level, n (%) | |
| | No/Unknown education | 58 (0.6) |
| | Primary | 1,804 (17.7) |
| | Secondary | 3,496 (34.3) |
| | Tertiary | 4,841 (47.5) |
| Older people (65+ years) | Number of persons, n | 103 (100.0) |
| | Male, n (%) | 8 (7.8) |
| | Age, median (Q1;Q3) | 70.3 (66.8;74.7) |
| | CCI, n (%) | |
| | 0 | 44 (42.7) |
| | 1-2 | 42 (40.8) |
| | +3 | 17 (16.5) |
| | Education level, n (%) | |
| | No/Unknown education | * |
| | Primary | 29 (28.2) |
| | Secondary | 46 (44.7) |
| | Tertiary | * |

**Supplementary Table 122**
**Eating disorders - Prevalent cohort 2021**
**Patient characteristics - relatives - pooled**

| *Age strata of index patients* | *Closest relative type* | *Characteristic of family relatives by agestrata of index patients* | *Closest family relatives of brain disorder patients* | *Comparison relative* |
|---|---|---|---|---|
| Children and young people (0-24 years) | Fathers | Number of persons, n | 4,957 (100.0) | 4,957 (100.0) |
| | | Male, n (%) | 4,957 (100.0) | 4,957 (100.0) |
| | | Age, median (Q1;Q3) | 53.0 (49.1;56.9) | 53.1 (49.4;56.5) |
| | | CCI, n (%) | | |
| | | 0 | 4,311 (87.0) | 4,305 (86.8) |
| | | 1-2 | 568 (11.5) | 572 (11.5) |
| | | +3 | 78 (1.6) | 80 (1.6) |
| | | Education level, n (%) | | |
| | | No/Unknown education | 48 (1.0) | 48 (1.0) |
| | | Primary | 622 (12.5) | 622 (12.5) |
| | | Secondary | 2,643 (53.3) | 2,643 (53.3) |
| | | Tertiary | 1,644 (33.2) | 1,644 (33.2) |
| | | Relation to patient, n (%) | | |
| | | Father | 4,957 (100.0) | 4,957 (100.0) |
| Children and young people (0-24 years) | Mothers | Number of persons, n | 5,201 (100.0) | 5,201 (100.0) |
| | | Male, n (%) | 0 (0.0) | 0 (0.0) |
| | | Age, median (Q1;Q3) | 50.9 (47.3;54.5) | 50.7 (47.3;54.0) |
| | | CCI, n (%) | | |
| | | 0 | 4,514 (86.8) | 4,488 (86.3) |
| | | 1-2 | 636 (12.2) | 652 (12.5) |
| | | +3 | 51 (1.0) | 61 (1.2) |
| | | Education level, n (%) | | |
| | | No/Unknown education | 21 (0.4) | 21 (0.4) |
| | | Primary | 601 (11.6) | 601 (11.6) |
| | | Secondary | 2,302 (44.3) | 2,302 (44.3) |
| | | Tertiary | 2,277 (43.8) | 2,277 (43.8) |
| | | Relation to patient, n (%) | | |
| | | Mother | 5,201 (100.0) | 5,201 (100.0) |
| Young adults and adults (25-64 years) | Pooled | Number of persons, n | 10,199 (100.0) | 10,199 (100.0) |
| | | Male, n (%) | 7,008 (68.7) | 7,008 (68.7) |
| | | Age, median (Q1;Q3) | 39.2 (30.4;55.7) | 39.1 (30.5;55.7) |
| | | CCI, n (%) | | |
| | | 0 | 8,975 (88.0) | 9,027 (88.5) |
| | | 1-2 | 1,051 (10.3) | 1,014 (9.9) |
| | | +3 | 173 (1.7) | 158 (1.5) |
| | | Education level, n (%) | | |
| | | No/Unknown education | 803 (7.9) | 803 (7.9) |
| | | Primary | 1,754 (17.2) | 1,754 (17.2) |
| | | Secondary | 4,113 (40.3) | 4,113 (40.3) |
| | | Tertiary | 3,529 (34.6) | 3,529 (34.6) |
| | | Relation to patient, n (%) | | |
| | | Mother | 2,216 (21.7) | 2,216 (21.7) |
| | | Father | 594 (5.8) | 594 (5.8) |
| | | Partner | 5,726 (56.1) | 5,726 (56.1) |
| | | Child | 1,177 (11.5) | 1,177 (11.5) |

**Supplementary Table 122**
**Eating disorders - Prevalent cohort 2021**
**Patient characteristics - relatives - pooled**

| *Age strata of index patients* | *Closest relative type* | *Characteristic of family relatives by agestrata of index patients* | *Closest family relatives of brain disorder patients* | *Comparison relative* |
|---|---|---|---|---|
| | | Sibling | 486 (4.8) | 486 (4.8) |
| Older people (65+ years) | Pooled | Number of persons, n | 103 (100.0) | 103 (100.0) |
| | | Male, n (%) | 62 (60.2) | 62 (60.2) |
| | | Age, median (Q1;Q3) | 54.4 (46.1;68.1) | 54.1 (45.0;68.8) |
| | | CCI, n (%) | | |
| | | 0 | 78 (75.7) | 81 (78.6) |
| | | 1-2 | 20 (19.4) | 14 (13.6) |
| | | +3 | 5 (4.9) | 8 (7.8) |
| | | Education level, n (%) | | |
| | | No/Unknown education | * | * |
| | | Primary | * | * |
| | | Secondary | 44 (42.7) | 44 (42.7) |
| | | Tertiary | 38 (36.9) | 38 (36.9) |
| | | Relation to patient, n (%) | | |
| | | Partner | 37 (35.9) | 37 (35.9) |
| | | Child | 57 (55.3) | 57 (55.3) |
| | | Sibling | 9 (8.7) | 9 (8.7) |

#### Supplementary Table 123
#### Eating disorders - Prevalent cohort 2021
#### Patient characteristics - relatives - not pooled

| *Age strata of index patients* | *Closest relative type* | *Characteristic of family relatives by agestrata of index patients* | *Closest family relatives of brain disorder patients* | *Comparison relative* |
|---|---|---|---|---|
| Children and young people (0-24 years) | Fathers | Number of persons, n | 4,957 (100.0) | 4,957 (100.0) |
| | | Male, n (%) | 4,957 (100.0) | 4,957 (100.0) |
| | | Age, median (Q1;Q3) | 53.0 (49.1;56.9) | 53.1 (49.4;56.5) |
| | | CCI, n (%) | | |
| | | 0 | 4,311 (87.0) | 4,305 (86.8) |
| | | 1-2 | 568 (11.5) | 572 (11.5) |
| | | +3 | 78 (1.6) | 80 (1.6) |
| | | Education level, n (%) | | |
| | | No/Unknown education | 48 (1.0) | 48 (1.0) |
| | | Primary | 622 (12.5) | 622 (12.5) |
| | | Secondary | 2,643 (53.3) | 2,643 (53.3) |
| | | Tertiary | 1,644 (33.2) | 1,644 (33.2) |
| | | Relation to patient, n (%) | | |
| | | Father | 4,957 (100.0) | 4,957 (100.0) |
| Children and young people (0-24 years) | Mothers | Number of persons, n | 5,201 (100.0) | 5,201 (100.0) |
| | | Male, n (%) | 0 (0.0) | 0 (0.0) |
| | | Age, median (Q1;Q3) | 50.9 (47.3;54.5) | 50.7 (47.3;54.0) |
| | | CCI, n (%) | | |
| | | 0 | 4,514 (86.8) | 4,488 (86.3) |
| | | 1-2 | 636 (12.2) | 652 (12.5) |
| | | +3 | 51 (1.0) | 61 (1.2) |
| | | Education level, n (%) | | |
| | | No/Unknown education | 21 (0.4) | 21 (0.4) |
| | | Primary | 601 (11.6) | 601 (11.6) |
| | | Secondary | 2,302 (44.3) | 2,302 (44.3) |
| | | Tertiary | 2,277 (43.8) | 2,277 (43.8) |
| | | Relation to patient, n (%) | | |
| | | Mother | 5,201 (100.0) | 5,201 (100.0) |
| Young adults and adults (25-64 years) | Children | Number of persons, n | 1,177 (100.0) | 1,177 (100.0) |
| | | Male, n (%) | 551 (46.8) | 551 (46.8) |
| | | Age, median (Q1;Q3) | 12.0 (6.2;18.0) | 11.7 (6.4;18.1) |
| | | CCI, n (%) | | |
| | | 0 | 1,105 (93.9) | 1,117 (94.9) |
| | | 1-2 | 72 (6.1) | * |
| | | +3 | 0 (0.0) | * |
| | | Education level, n (%) | | |
| | | No/Unknown education | 662 (56.2) | 662 (56.2) |
| | | Primary | 382 (32.5) | 382 (32.5) |
| | | Secondary | 87 (7.4) | 87 (7.4) |
| | | Tertiary | 46 (3.9) | 46 (3.9) |
| | | Relation to patient, n (%) | | |
| | | Child | 1,177 (100.0) | 1,177 (100.0) |
| Young adults and adults (25-64 years) | Parents | Number of persons, n | 2,810 (100.0) | 2,810 (100.0) |
| | | Male, n (%) | 594 (21.1) | 594 (21.1) |
| | | Age, median (Q1;Q3) | 61.1 (56.5;66.7) | 60.9 (56.7;66.6) |

### Supplementary Table 123
### Eating disorders - Prevalent cohort 2021
### Patient characteristics - relatives - not pooled

| *Age strata of index patients* | *Closest relative type* | *Characteristic of family relatives by agestrata of index patients* | *Closest family relatives of brain disorder patients* | *Comparison relative* |
|---|---|---|---|---|
| | | CCI, n (%) | | |
| | | 0 | 2,081 (74.1) | 2,083 (74.1) |
| | | 1-2 | 603 (21.5) | 606 (21.6) |
| | | +3 | 126 (4.5) | 121 (4.3) |
| | | Education level, n (%) | | |
| | | No/Unknown education | 48 (1.7) | 48 (1.7) |
| | | Primary | 488 (17.4) | 488 (17.4) |
| | | Secondary | 1,354 (48.2) | 1,354 (48.2) |
| | | Tertiary | 920 (32.7) | 920 (32.7) |
| | | Relation to patient, n (%) | | |
| | | Mother | 2,216 (78.9) | 2,216 (78.9) |
| | | Father | 594 (21.1) | 594 (21.1) |
| Young adults and adults (25-64 years) | Partners | Number of persons, n | 5,726 (100.0) | 5,726 (100.0) |
| | | Male, n (%) | 5,590 (97.6) | 5,590 (97.6) |
| | | Age, median (Q1;Q3) | 35.8 (30.9;42.5) | 35.8 (31.0;42.4) |
| | | CCI, n (%) | | |
| | | 0 | 5,347 (93.4) | 5,394 (94.2) |
| | | 1-2 | 335 (5.9) | 303 (5.3) |
| | | +3 | 44 (0.8) | 29 (0.5) |
| | | Education level, n (%) | | |
| | | No/Unknown education | 90 (1.6) | 90 (1.6) |
| | | Primary | 782 (13.7) | 782 (13.7) |
| | | Secondary | 2,443 (42.7) | 2,443 (42.7) |
| | | Tertiary | 2,411 (42.1) | 2,411 (42.1) |
| | | Relation to patient, n (%) | | |
| | | Partner | 5,726 (100.0) | 5,726 (100.0) |
| Young adults and adults (25-64 years) | Siblings | Number of persons, n | 486 (100.0) | 486 (100.0) |
| | | Male, n (%) | 273 (56.2) | 273 (56.2) |
| | | Age, median (Q1;Q3) | 35.4 (28.6;48.9) | 35.3 (28.4;48.6) |
| | | CCI, n (%) | | |
| | | 0 | 442 (90.9) | 433 (89.1) |
| | | 1-2 | * | 47 (9.7) |
| | | +3 | * | 6 (1.2) |
| | | Education level, n (%) | | |
| | | No/Unknown education | * | * |
| | | Primary | * | * |
| | | Secondary | 229 (47.1) | 229 (47.1) |
| | | Tertiary | 152 (31.3) | 152 (31.3) |
| | | Relation to patient, n (%) | | |
| | | Sibling | 486 (100.0) | 486 (100.0) |
| Older people (65+ years) | Children | Number of persons, n | 57 (100.0) | 57 (100.0) |
| | | Male, n (%) | 29 (50.9) | 29 (50.9) |
| | | Age, median (Q1;Q3) | 46.9 (41.3;50.3) | 45.8 (41.9;51.7) |
| | | CCI, n (%) | | |
| | | 0 | 52 (91.2) | 50 (87.7) |

**Supplementary Table 123**
**Eating disorders - Prevalent cohort 2021**
**Patient characteristics - relatives - not pooled**

| *Age strata of index patients* | *Closest relative type* | *Characteristic of family relatives by agestrata of index patients* | *Closest family relatives of brain disorder patients* | *Comparison relative* |
|---|---|---|---|---|
| | | 1-2 | 5 (8.8) | 7 (12.3) |
| | | +3 | 0 (0.0) | 0 (0.0) |
| | | Education level, n (%) | | |
| | | No/Unknown education | 0 (0.0) | 0 (0.0) |
| | | Primary | 11 (19.3) | 11 (19.3) |
| | | Secondary | 21 (36.8) | 21 (36.8) |
| | | Tertiary | 25 (43.9) | 25 (43.9) |
| | | Relation to patient, n (%) | | |
| | | Child | 57 (100.0) | 57 (100.0) |
| Older people (65+ years) | Partners | Number of persons, n | 37 (100.0) | 37 (100.0) |
| | | Male, n (%) | 32 (86.5) | 32 (86.5) |
| | | Age, median (Q1;Q3) | 70.0 (67.5;74.6) | 70.8 (68.0;73.3) |
| | | CCI, n (%) | | |
| | | 0 | 21 (56.8) | 25 (67.6) |
| | | 1-2 | 11 (29.7) | 6 (16.2) |
| | | +3 | 5 (13.5) | 6 (16.2) |
| | | Education level, n (%) | | |
| | | No/Unknown education | * | * |
| | | Primary | * | * |
| | | Secondary | 19 (51.4) | 19 (51.4) |
| | | Tertiary | * | * |
| | | Relation to patient, n (%) | | |
| | | Partner | 37 (100.0) | 37 (100.0) |
| Older people (65+ years) | Siblings | Number of persons, n | 9 (100.0) | 9 (100.0) |
| | | Male, n (%) | * | * |
| | | Age, median (Q1;Q3) | 66.1 (64.0;67.8) | 67.1 (63.5;68.3) |
| | | CCI, n (%) | | |
| | | 0 | * | * |
| | | 1-2 | * | * |
| | | +3 | 0 (0.0) | * |
| | | Education level, n (%) | | |
| | | No/Unknown education | 0 (0.0) | 0 (0.0) |
| | | Primary | * | * |
| | | Secondary | * | * |
| | | Tertiary | * | * |
| | | Relation to patient, n (%) | | |
| | | Sibling | 9 (100.0) | 9 (100.0) |

#### Supplementary Table 124
#### Eating disorders - Prevalent cohort 2021
#### Cost analysis - pooled

| | | | | *Actual costs* | | *Attributable costs* | |
|---|---|---|---|---|---|---|---|
| *Age strata of index patients* | *Closest relative type* | *Number of index patients included in cost component calculations* | *Cost component* | *Total (million EUR)* | *Per person (EUR)* | *Total (million EUR)* | *Per person (EUR)* |
| Children and young people (0-24 years) | Fathers | 4,957 | Primary sector | 1.2 | 250 | 0.0 | 9 |
| | | 4,957 | Hospital care | 8.0 | 1,607 | 0.2 | 40 |
| | | 4,957 | Filled prescriptions | 1.0 | 211 | 0.1 | 28 |
| | | 4,713 | Lost productivity (income loss) | . | . | -37.0 | -7,859 |
| Children and young people (0-24 years) | Mothers | 5,201 | Primary sector | 1.9 | 372 | 0.1 | 26 |
| | | 5,201 | Hospital care | 8.3 | 1,596 | 0.4 | 73 |
| | | 5,201 | Filled prescriptions | 1.1 | 218 | 0.1 | 10 |
| | | 5,154 | Lost productivity (income loss) | . | . | -10.1 | -1,954 |
| Young adults and adults (25-64 years) | Pooled | 10,199 | Primary sector | 2.7 | 265 | 0.2 | 20 |
| | | 10,199 | Hospital care | 15.2 | 1,489 | 0.4 | 44 |
| | | 10,199 | Filled prescriptions | 1.9 | 189 | 0.0 | 1 |
| | | 8,017 | Lost productivity (income loss) | . | . | -2.6 | -329 |
| Older people (65+ years) | Pooled | 103 | Primary sector | 0.0 | 347 | -0.0 | -12 |
| | | 103 | Hospital care | 0.2 | 1,959 | -0.1 | -1,399 |
| | | 103 | Filled prescriptions | 0.0 | 382 | 0.0 | 34 |
| | | 60 | Lost productivity (income loss) | . | . | 0.7 | 11,931 |

#### Supplementary Table 125
#### Eating disorders - Prevalent cohort 2021
#### Cost analysis - not pooled

| *Age strata of index patients* | *Closest relative type* | *Number of index patients included in cost component calculations* | *Cost component* | *Actual costs* | | *Attributable costs* | |
|---|---|---|---|---|---|---|---|
| | | | | *Total (million EUR)* | *Per person (EUR)* | *Total (million EUR)* | *Per person (EUR)* |
| Children and young people (0-24 years) | Fathers | 4,957 | Primary sector | 1.2 | 250 | 0.0 | 9 |
| | | 4,957 | Hospital care | 8.0 | 1,607 | 0.2 | 40 |
| | | 4,957 | Filled prescriptions | 1.0 | 211 | 0.1 | 28 |
| | | 4,713 | Lost productivity (income loss) | . | . | -37.0 | -7,859 |
| Children and young people (0-24 years) | Mothers | 5,201 | Primary sector | 1.9 | 372 | 0.1 | 26 |
| | | 5,201 | Hospital care | 8.3 | 1,596 | 0.4 | 73 |
| | | 5,201 | Filled prescriptions | 1.1 | 218 | 0.1 | 10 |
| | | 5,154 | Lost productivity (income loss) | . | . | -10.1 | -1,954 |
| Young adults and adults (25-64 years) | Children | 1,177 | Primary sector | 0.2 | 201 | 0.0 | 35 |
| | | 1,177 | Hospital care | 1.0 | 823 | -0.3 | -294 |
| | | 1,177 | Filled prescriptions | 0.1 | 104 | 0.0 | 31 |
| | | 246 | Lost productivity (income loss) | . | . | 0.7 | 2,888 |
| Young adults and adults (25-64 years) | Parents | 2,810 | Primary sector | 1.2 | 434 | 0.1 | 32 |
| | | 2,810 | Hospital care | 7.6 | 2,718 | 0.7 | 237 |
| | | 2,810 | Filled prescriptions | 1.1 | 391 | -0.1 | -25 |
| | | 1,655 | Lost productivity (income loss) | . | . | -3.5 | -2,104 |
| Young adults and adults (25-64 years) | Partners | 5,726 | Primary sector | 1.1 | 195 | 0.1 | 13 |
| | | 5,726 | Hospital care | 5.4 | 936 | -0.2 | -29 |
| | | 5,726 | Filled prescriptions | 0.6 | 110 | 0.1 | 11 |
| | | 5,662 | Lost productivity (income loss) | . | . | 1.3 | 229 |
| Young adults and adults (25-64 years) | Siblings | 486 | Primary sector | 0.1 | 263 | -0.0 | -2 |
| | | 486 | Hospital care | 1.2 | 2,542 | 0.3 | 605 |
| | | 486 | Filled prescriptions | 0.1 | 159 | -0.0 | -31 |
| | | 454 | Lost productivity (income loss) | . | . | -1.2 | -2,566 |
| Older people (65+ years) | Children | 57 | Primary sector | 0.0 | 312 | 0.0 | 55 |
| | | 57 | Hospital care | 0.0 | 716 | -0.0 | -677 |
| | | 57 | Filled prescriptions | 0.0 | 230 | -0.0 | -44 |
| | | 56 | Lost productivity (income loss) | . | . | 0.5 | 9,093 |
| Older people (65+ years) | Partners | 37 | Primary sector | 0.0 | 392 | -0.0 | -77 |
| | | 37 | Hospital care | 0.1 | 3,895 | -0.0 | -1,341 |
| | | 37 | Filled prescriptions | 0.0 | 554 | 0.0 | 100 |
| | | . | Lost productivity (income loss) | . | . | . | . |
| Older people (65+ years) | Siblings | 9 | Primary sector | 0.0 | 398 | -0.0 | -196 |
| | | 9 | Hospital care | 0.0 | 2,247 | -0.1 | -6,485 |
| | | 9 | Filled prescriptions | 0.0 | 689 | 0.0 | 285 |
| | | . | Lost productivity (income loss) | . | . | . | . |

**Supplementary Table 126**
**Epilepsy - Incident cohort 2016-2021**
**Identification of populations of relatives**

| ***Agegroup of index patients in matched population*** | | ***Closest relative type identified*** | | ***Available control with same type of closest relative*** |
|---|---|---|---|---|
| Children and young people (0-24 years) | n = 6,420 | Fathers | n = 4,919 (77%) | 96% |
| | | Mothers | n = 5,113 (80%) | 96% |
| | | No available closest relative | n = 843 (13%) | n/a |
| Young adults and adults (25-64 years) | n = 7,007 | Children | n = 1,073 (15%) | 91% |
| | | Parents | n = 1,016 (14%) | 88% |
| | | Partners | n = 3,407 (49%) | 96% |
| | | Siblings | n = 1,088 (16%) | 52% |
| | | Pooled | n = 6,584 (94%) | 86% |
| | | No available closest relative | n = 423 (6%) | n/a |
| Older people (65+ years) | n = 8,357 | Children | n = 3,100 (37%) | 94% |
| | | Partners | n = 4,113 (49%) | 96% |
| | | Siblings | n = 138 (2%) | 38% |
| | | Pooled | n = 7,351 (88%) | 94% |
| | | No available closest relative | n = 1,006 (12%) | n/a |

**Supplementary Table 127**
**Epilepsy - Incident cohort 2016-2021**
**Patient characteristics - cases**

| ***Age strata of index patients*** | ***Characteristic*** | |
|---|---|---|
| Children and young people (0-24 years) | Number of persons, n | 5,389 (100.0) |
| | Male, n (%) | 2,882 (53.5) |
| | Age, median (Q1;Q3) | 11.0 (5.6;17.0) |
| | CCI, n (%) | |
| | 0 | 4,817 (89.4) |
| | 1-2 | 527 (9.8) |
| | +3 | 45 (0.8) |
| | Education level, n (%) | |
| | No/Unknown education | 3,233 (60.0) |
| | Primary | 1,816 (33.7) |
| | Secondary | 314 (5.8) |
| | Tertiary | 26 (0.5) |
| Young adults and adults (25-64 years) | Number of persons, n | 5,728 (100.0) |
| | Male, n (%) | 3,092 (54.0) |
| | Age, median (Q1;Q3) | 52.1 (40.8;58.9) |
| | CCI, n (%) | |
| | 0 | 3,220 (56.2) |
| | 1-2 | 1,824 (31.8) |
| | +3 | 684 (11.9) |
| | Education level, n (%) | |
| | No/Unknown education | 278 (4.9) |
| | Primary | 1,677 (29.3) |
| | Secondary | 2,479 (43.3) |
| | Tertiary | 1,294 (22.6) |
| Older people (65+ years) | Number of persons, n | 6,933 (100.0) |
| | Male, n (%) | 3,688 (53.2) |
| | Age, median (Q1;Q3) | 75.8 (71.0;81.3) |
| | CCI, n (%) | |
| | 0 | 1,760 (25.4) |
| | 1-2 | 3,246 (46.8) |
| | +3 | 1,927 (27.8) |
| | Education level, n (%) | |
| | No/Unknown education | 425 (6.1) |
| | Primary | 2,273 (32.8) |
| | Secondary | 3,077 (44.4) |
| | Tertiary | 1,158 (16.7) |

**Supplementary Table 128**
**Epilepsy - Incident cohort 2016-2021**
**Patient characteristics - relatives - pooled**

| *Age strata of index patients* | *Closest relative type* | *Characteristic of family relatives by agestrata of index patients* | *Closest family relatives of brain disorder patients* | *Comparison relative* |
|---|---|---|---|---|
| Children and young people (0-24 years) | Fathers | Number of persons, n | 4,746 (100.0) | 4,746 (100.0) |
| | | Male, n (%) | 4,746 (100.0) | 4,746 (100.0) |
| | | Age, median (Q1;Q3) | 44.3 (38.1;50.3) | 44.2 (38.2;50.0) |
| | | CCI, n (%) | | |
| | | 0 | 4,272 (90.0) | 4,318 (91.0) |
| | | 1-2 | 424 (8.9) | 375 (7.9) |
| | | +3 | 50 (1.1) | 53 (1.1) |
| | | Education level, n (%) | | |
| | | No/Unknown education | 107 (2.3) | 107 (2.3) |
| | | Primary | 1,093 (23.0) | 1,093 (23.0) |
| | | Secondary | 2,274 (47.9) | 2,274 (47.9) |
| | | Tertiary | 1,272 (26.8) | 1,272 (26.8) |
| | | Relation to patient, n (%) | | |
| | | Father | 4,746 (100.0) | 4,746 (100.0) |
| Children and young people (0-24 years) | Mothers | Number of persons, n | 4,937 (100.0) | 4,937 (100.0) |
| | | Male, n (%) | 0 (0.0) | 0 (0.0) |
| | | Age, median (Q1;Q3) | 41.8 (35.5;47.7) | 42.1 (35.7;47.6) |
| | | CCI, n (%) | | |
| | | 0 | 4,356 (88.2) | 4,446 (90.1) |
| | | 1-2 | 533 (10.8) | 451 (9.1) |
| | | +3 | 48 (1.0) | 40 (0.8) |
| | | Education level, n (%) | | |
| | | No/Unknown education | 130 (2.6) | 130 (2.6) |
| | | Primary | 919 (18.6) | 919 (18.6) |
| | | Secondary | 2,024 (41.0) | 2,024 (41.0) |
| | | Tertiary | 1,864 (37.8) | 1,864 (37.8) |
| | | Relation to patient, n (%) | | |
| | | Mother | 4,937 (100.0) | 4,937 (100.0) |
| Young adults and adults (25-64 years) | Pooled | Number of persons, n | 5,728 (100.0) | 5,728 (100.0) |
| | | Male, n (%) | 2,621 (45.8) | 2,621 (45.8) |
| | | Age, median (Q1;Q3) | 52.2 (36.1;60.4) | 52.2 (36.1;60.5) |
| | | CCI, n (%) | | |
| | | 0 | 4,686 (81.8) | 4,770 (83.3) |
| | | 1-2 | 852 (14.9) | 797 (13.9) |
| | | +3 | 190 (3.3) | 161 (2.8) |
| | | Education level, n (%) | | |
| | | No/Unknown education | 342 (6.0) | 342 (6.0) |
| | | Primary | 1,548 (27.0) | 1,548 (27.0) |
| | | Secondary | 2,588 (45.2) | 2,588 (45.2) |
| | | Tertiary | 1,250 (21.8) | 1,250 (21.8) |
| | | Relation to patient, n (%) | | |
| | | Mother | 709 (12.4) | 709 (12.4) |
| | | Father | 188 (3.3) | 188 (3.3) |
| | | Partner | 3,271 (57.1) | 3,271 (57.1) |
| | | Child | 986 (17.2) | 986 (17.2) |

**Supplementary Table 128**
**Epilepsy - Incident cohort 2016-2021**
**Patient characteristics - relatives - pooled**

| *Age strata of index patients* | *Closest relative type* | *Characteristic of family relatives by agestrata of index patients* | *Closest family relatives of brain disorder patients* | *Comparison relative* |
|---|---|---|---|---|
| | | Sibling | 574 (10.0) | 574 (10.0) |
| Older people (65+ years) | Pooled | Number of persons, n | 6,933 (100.0) | 6,933 (100.0) |
| | | Male, n (%) | 2,921 (42.1) | 2,921 (42.1) |
| | | Age, median (Q1;Q3) | 66.0 (54.3;74.4) | 66.1 (54.2;74.2) |
| | | CCI, n (%) | | |
| | | 0 | 4,971 (71.7) | 5,015 (72.3) |
| | | 1-2 | 1,561 (22.5) | 1,522 (22.0) |
| | | +3 | 401 (5.8) | 396 (5.7) |
| | | Education level, n (%) | | |
| | | No/Unknown education | 209 (3.0) | 209 (3.0) |
| | | Primary | 1,578 (22.8) | 1,578 (22.8) |
| | | Secondary | 3,418 (49.3) | 3,418 (49.3) |
| | | Tertiary | 1,728 (24.9) | 1,728 (24.9) |
| | | Relation to patient, n (%) | | |
| | | Partner | 3,963 (57.2) | 3,963 (57.2) |
| | | Child | 2,917 (42.1) | 2,917 (42.1) |
| | | Sibling | 53 (0.8) | 53 (0.8) |

**Supplementary Table 129**
**Epilepsy - Incident cohort 2016-2021**
**Patient characteristics - relatives - not pooled**

| *Age strata of index patients* | *Closest relative type* | *Characteristic of family relatives by agestrata of index patients* | *Closest family relatives of brain disorder patients* | *Comparison relative* |
|---|---|---|---|---|
| Children and young people (0-24 years) | Fathers | Number of persons, n | 4,746 (100.0) | 4,746 (100.0) |
| | | Male, n (%) | 4,746 (100.0) | 4,746 (100.0) |
| | | Age, median (Q1;Q3) | 44.3 (38.1;50.3) | 44.2 (38.2;50.0) |
| | | CCI, n (%) | | |
| | | 0 | 4,272 (90.0) | 4,318 (91.0) |
| | | 1-2 | 424 (8.9) | 375 (7.9) |
| | | +3 | 50 (1.1) | 53 (1.1) |
| | | Education level, n (%) | | |
| | | No/Unknown education | 107 (2.3) | 107 (2.3) |
| | | Primary | 1,093 (23.0) | 1,093 (23.0) |
| | | Secondary | 2,274 (47.9) | 2,274 (47.9) |
| | | Tertiary | 1,272 (26.8) | 1,272 (26.8) |
| | | Relation to patient, n (%) | | |
| | | Father | 4,746 (100.0) | 4,746 (100.0) |
| Children and young people (0-24 years) | Mothers | Number of persons, n | 4,937 (100.0) | 4,937 (100.0) |
| | | Male, n (%) | 0 (0.0) | 0 (0.0) |
| | | Age, median (Q1;Q3) | 41.8 (35.5;47.7) | 42.1 (35.7;47.6) |
| | | CCI, n (%) | | |
| | | 0 | 4,356 (88.2) | 4,446 (90.1) |
| | | 1-2 | 533 (10.8) | 451 (9.1) |
| | | +3 | 48 (1.0) | 40 (0.8) |
| | | Education level, n (%) | | |
| | | No/Unknown education | 130 (2.6) | 130 (2.6) |
| | | Primary | 919 (18.6) | 919 (18.6) |
| | | Secondary | 2,024 (41.0) | 2,024 (41.0) |
| | | Tertiary | 1,864 (37.8) | 1,864 (37.8) |
| | | Relation to patient, n (%) | | |
| | | Mother | 4,937 (100.0) | 4,937 (100.0) |
| Young adults and adults (25-64 years) | Children | Number of persons, n | 986 (100.0) | 986 (100.0) |
| | | Male, n (%) | 510 (51.7) | 510 (51.7) |
| | | Age, median (Q1;Q3) | 24.4 (16.1;32.1) | 24.1 (15.9;32.0) |
| | | CCI, n (%) | | |
| | | 0 | 924 (93.7) | 925 (93.8) |
| | | 1-2 | * | * |
| | | +3 | * | * |
| | | Education level, n (%) | | |
| | | No/Unknown education | 193 (19.6) | 193 (19.6) |
| | | Primary | 386 (39.1) | 386 (39.1) |
| | | Secondary | 264 (26.8) | 264 (26.8) |
| | | Tertiary | 143 (14.5) | 143 (14.5) |
| | | Relation to patient, n (%) | | |
| | | Child | 986 (100.0) | 986 (100.0) |
| Young adults and adults (25-64 years) | Parents | Number of persons, n | 897 (100.0) | 897 (100.0) |
| | | Male, n (%) | 188 (21.0) | 188 (21.0) |
| | | Age, median (Q1;Q3) | 67.7 (58.2;76.8) | 67.6 (58.7;76.7) |

#### Supplementary Table 129
#### Epilepsy - Incident cohort 2016-2021
#### Patient characteristics - relatives - not pooled

| *Age strata of index patients* | *Closest relative type* | *Characteristic of family relatives by agestrata of index patients* | *Closest family relatives of brain disorder patients* | *Comparison relative* |
|---|---|---|---|---|
| | | CCI, n (%) | | |
| | | 0 | 559 (62.3) | 607 (67.7) |
| | | 1-2 | 248 (27.6) | 223 (24.9) |
| | | +3 | 90 (10.0) | 67 (7.5) |
| | | Education level, n (%) | | |
| | | No/Unknown education | 20 (2.2) | 20 (2.2) |
| | | Primary | 318 (35.5) | 318 (35.5) |
| | | Secondary | 401 (44.7) | 401 (44.7) |
| | | Tertiary | 158 (17.6) | 158 (17.6) |
| | | Relation to patient, n (%) | | |
| | | Mother | 709 (79.0) | 709 (79.0) |
| | | Father | 188 (21.0) | 188 (21.0) |
| Young adults and adults (25-64 years) | Partners | Number of persons, n | 3,271 (100.0) | 3,271 (100.0) |
| | | Male, n (%) | 1,557 (47.6) | 1,557 (47.6) |
| | | Age, median (Q1;Q3) | 52.5 (42.4;59.2) | 52.2 (42.1;59.2) |
| | | CCI, n (%) | | |
| | | 0 | 2,738 (83.7) | 2,764 (84.5) |
| | | 1-2 | 452 (13.8) | 429 (13.1) |
| | | +3 | 81 (2.5) | 78 (2.4) |
| | | Education level, n (%) | | |
| | | No/Unknown education | 129 (3.9) | 129 (3.9) |
| | | Primary | 695 (21.2) | 695 (21.2) |
| | | Secondary | 1,603 (49.0) | 1,603 (49.0) |
| | | Tertiary | 844 (25.8) | 844 (25.8) |
| | | Relation to patient, n (%) | | |
| | | Partner | 3,271 (100.0) | 3,271 (100.0) |
| Young adults and adults (25-64 years) | Siblings | Number of persons, n | 574 (100.0) | 574 (100.0) |
| | | Male, n (%) | 366 (63.8) | 366 (63.8) |
| | | Age, median (Q1;Q3) | 55.3 (48.1;60.0) | 55.2 (48.4;59.8) |
| | | CCI, n (%) | | |
| | | 0 | 465 (81.0) | 474 (82.6) |
| | | 1-2 | 93 (16.2) | 86 (15.0) |
| | | +3 | 16 (2.8) | 14 (2.4) |
| | | Education level, n (%) | | |
| | | No/Unknown education | 0 (0.0) | 0 (0.0) |
| | | Primary | 149 (26.0) | 149 (26.0) |
| | | Secondary | 320 (55.7) | 320 (55.7) |
| | | Tertiary | 105 (18.3) | 105 (18.3) |
| | | Relation to patient, n (%) | | |
| | | Sibling | 574 (100.0) | 574 (100.0) |
| Older people (65+ years) | Children | Number of persons, n | 2,917 (100.0) | 2,917 (100.0) |
| | | Male, n (%) | 1,447 (49.6) | 1,447 (49.6) |
| | | Age, median (Q1;Q3) | 52.6 (46.8;58.0) | 52.4 (47.0;57.7) |
| | | CCI, n (%) | | |
| | | 0 | 2,464 (84.5) | 2,495 (85.5) |

**Supplementary Table 129**
**Epilepsy - Incident cohort 2016-2021**
**Patient characteristics - relatives - not pooled**

| *Age strata of index patients* | *Closest relative type* | *Characteristic of family relatives by agestrata of index patients* | *Closest family relatives of brain disorder patients* | *Comparison relative* |
|---|---|---|---|---|
| | | 1-2 | 402 (13.8) | 372 (12.8) |
| | | +3 | 51 (1.7) | 50 (1.7) |
| | | Education level, n (%) | | |
| | | No/Unknown education | 16 (0.5) | 16 (0.5) |
| | | Primary | 470 (16.1) | 470 (16.1) |
| | | Secondary | 1,519 (52.1) | 1,519 (52.1) |
| | | Tertiary | 912 (31.3) | 912 (31.3) |
| | | Relation to patient, n (%) | | |
| | | Child | 2,917 (100.0) | 2,917 (100.0) |
| Older people (65+ years) | Partners | Number of persons, n | 3,963 (100.0) | 3,963 (100.0) |
| | | Male, n (%) | 1,448 (36.5) | 1,448 (36.5) |
| | | Age, median (Q1;Q3) | 73.3 (68.7;78.0) | 73.2 (68.8;77.9) |
| | | CCI, n (%) | | |
| | | 0 | 2,470 (62.3) | 2,478 (62.5) |
| | | 1-2 | 1,146 (28.9) | 1,141 (28.8) |
| | | +3 | 347 (8.8) | 344 (8.7) |
| | | Education level, n (%) | | |
| | | No/Unknown education | 193 (4.9) | 193 (4.9) |
| | | Primary | 1,099 (27.7) | 1,099 (27.7) |
| | | Secondary | 1,873 (47.3) | 1,873 (47.3) |
| | | Tertiary | 798 (20.1) | 798 (20.1) |
| | | Relation to patient, n (%) | | |
| | | Partner | 3,963 (100.0) | 3,963 (100.0) |
| Older people (65+ years) | Siblings | Number of persons, n | 53 (100.0) | 53 (100.0) |
| | | Male, n (%) | 26 (49.1) | 26 (49.1) |
| | | Age, median (Q1;Q3) | 61.6 (60.3;63.8) | 62.3 (59.5;64.0) |
| | | CCI, n (%) | | |
| | | 0 | 37 (69.8) | 42 (79.2) |
| | | 1-2 | * | * |
| | | +3 | * | * |
| | | Education level, n (%) | | |
| | | No/Unknown education | 0 (0.0) | 0 (0.0) |
| | | Primary | 9 (17.0) | 9 (17.0) |
| | | Secondary | 26 (49.1) | 26 (49.1) |
| | | Tertiary | 18 (34.0) | 18 (34.0) |
| | | Relation to patient, n (%) | | |
| | | Sibling | 53 (100.0) | 53 (100.0) |

#### Supplementary Table 130
#### Epilepsy - Incident cohort 2016-2021
#### Cost analysis - pooled

| *Age strata of index patients* | *Closest relative type* | *Number of index patients included in cost component calculations* | *Cost component* | *Actual costs* | | *Attributable costs* | |
|---|---|---|---|---|---|---|---|
| | | | | *Total (million EUR)* | *Per person (EUR)* | *Total (million EUR)* | *Per person (EUR)* |
| Children and young people (0-24 years) | Fathers | 4,746 | Primary sector | 1.0 | 209 | 0.1 | 12 |
| | | 4,746 | Hospital care | 5.6 | 1,179 | -0.2 | -50 |
| | | 4,746 | Filled prescriptions | 0.7 | 148 | 0.0 | 5 |
| | | 4,666 | Lost productivity (income loss) | . | . | 12.0 | 2,569 |
| Children and young people (0-24 years) | Mothers | 4,937 | Primary sector | 1.7 | 341 | 0.1 | 26 |
| | | 4,937 | Hospital care | 8.5 | 1,719 | 1.2 | 235 |
| | | 4,937 | Filled prescriptions | 0.9 | 173 | 0.1 | 18 |
| | | 4,914 | Lost productivity (income loss) | . | . | 18.8 | 3,821 |
| Young adults and adults (25-64 years) | Pooled | 5,728 | Primary sector | 1.9 | 329 | 0.2 | 28 |
| | | 5,728 | Hospital care | 14.2 | 2,492 | 2.1 | 368 |
| | | 5,728 | Filled prescriptions | 1.6 | 279 | 0.1 | 26 |
| | | 4,256 | Lost productivity (income loss) | . | . | 13.0 | 3,062 |
| Older people (65+ years) | Pooled | 6,933 | Primary sector | 2.9 | 425 | 0.1 | 15 |
| | | 6,933 | Hospital care | 21.7 | 3,154 | 1.2 | 176 |
| | | 6,933 | Filled prescriptions | 2.8 | 407 | 0.2 | 30 |
| | | 2,876 | Lost productivity (income loss) | . | . | 3.0 | 1,048 |

### Supplementary Table 131
### Epilepsy - Incident cohort 2016-2021
### Cost analysis - not pooled

| Age strata of index patients | Closest relative type | Number of index patients included in cost component calculations | Cost component | Actual costs: Total (million EUR) | Actual costs: Per person (EUR) | Attributable costs: Total (million EUR) | Attributable costs: Per person (EUR) |
|---|---|---|---|---|---|---|---|
| Children and young people (0-24 years) | Fathers | 4,746 | Primary sector | 1.0 | 209 | 0.1 | 12 |
| | | 4,746 | Hospital care | 5.6 | 1,179 | -0.2 | -50 |
| | | 4,746 | Filled prescriptions | 0.7 | 148 | 0.0 | 5 |
| | | 4,666 | Lost productivity (income loss) | . | . | 12.0 | 2,569 |
| Children and young people (0-24 years) | Mothers | 4,937 | Primary sector | 1.7 | 341 | 0.1 | 26 |
| | | 4,937 | Hospital care | 8.5 | 1,719 | 1.2 | 235 |
| | | 4,937 | Filled prescriptions | 0.9 | 173 | 0.1 | 18 |
| | | 4,914 | Lost productivity (income loss) | . | . | 18.8 | 3,821 |
| Young adults and adults (25-64 years) | Children | 986 | Primary sector | 0.2 | 238 | 0.0 | 25 |
| | | 986 | Hospital care | 2.0 | 2,037 | 0.7 | 720 |
| | | 986 | Filled prescriptions | 0.1 | 132 | 0.0 | 19 |
| | | 660 | Lost productivity (income loss) | . | . | 0.9 | 1,417 |
| Young adults and adults (25-64 years) | Parents | 897 | Primary sector | 0.4 | 484 | 0.0 | 47 |
| | | 897 | Hospital care | 3.0 | 3,398 | -0.1 | -93 |
| | | 897 | Filled prescriptions | 0.5 | 568 | 0.1 | 81 |
| | | 322 | Lost productivity (income loss) | . | . | 1.7 | 5,240 |
| Young adults and adults (25-64 years) | Partners | 3,271 | Primary sector | 1.0 | 320 | 0.1 | 28 |
| | | 3,271 | Hospital care | 7.4 | 2,262 | 0.9 | 262 |
| | | 3,271 | Filled prescriptions | 0.8 | 248 | 0.1 | 21 |
| | | 2,781 | Lost productivity (income loss) | . | . | 9.0 | 3,241 |
| Young adults and adults (25-64 years) | Siblings | 574 | Primary sector | 0.2 | 296 | 0.0 | 7 |
| | | 574 | Hospital care | 1.8 | 3,179 | 0.6 | 1,080 |
| | | 574 | Filled prescriptions | 0.1 | 256 | -0.0 | -18 |
| | | 493 | Lost productivity (income loss) | . | . | 1.4 | 2,832 |
| Older people (65+ years) | Children | 2,917 | Primary sector | 0.9 | 309 | 0.1 | 20 |
| | | 2,917 | Hospital care | 5.4 | 1,870 | 0.6 | 197 |
| | | 2,917 | Filled prescriptions | 0.7 | 250 | 0.1 | 35 |
| | | 2,661 | Lost productivity (income loss) | . | . | 2.9 | 1,079 |
| Older people (65+ years) | Partners | 3,963 | Primary sector | 2.0 | 513 | 0.0 | 12 |
| | | 3,963 | Hospital care | 16.2 | 4,125 | 0.7 | 181 |
| | | 3,963 | Filled prescriptions | 2.1 | 523 | 0.1 | 28 |
| | | 183 | Lost productivity (income loss) | . | . | 0.2 | 1,038 |
| Older people (65+ years) | Siblings | 53 | Primary sector | 0.0 | 337 | -0.0 | -77 |
| | | 53 | Hospital care | 0.1 | 1,783 | -0.1 | -1,326 |
| | | 53 | Filled prescriptions | 0.0 | 456 | -0.0 | -63 |
| | | 32 | Lost productivity (income loss) | . | . | -0.0 | -1,532 |

**Supplementary Table 132**
**Epilepsy - Prevalent cohort 2021**
**Identification of populations of relatives**

| ***Agegroup of index patients in matched population*** | | ***Closest relative type identified*** | | ***Available control with same type of closest relative*** |
|---|---|---|---|---|
| Children and young people (0-24 years) | n = 16,102 | Fathers | n = 13,765 (85%) | 96% |
| | | Mothers | n = 14,300 (89%) | 96% |
| | | No available closest relative | n = 679 (4%) | n/a |
| Young adults and adults (25-64 years) | n = 34,324 | Children | n = 3,909 (11%) | 93% |
| | | Parents | n = 7,954 (23%) | 90% |
| | | Partners | n = 15,519 (45%) | 96% |
| | | Siblings | n = 5,984 (17%) | 47% |
| | | Pooled | n = 33,366 (97%) | 85% |
| | | No available closest relative | n = 958 (3%) | n/a |
| Older people (65+ years) | n = 17,254 | Children | n = 5,944 (34%) | 94% |
| | | Partners | n = 8,527 (49%) | 96% |
| | | Siblings | n = 628 (4%) | 50% |
| | | Pooled | n = 15,099 (88%) | 93% |
| | | No available closest relative | n = 2,155 (12%) | n/a |

### Supplementary Table 133
### Epilepsy - Prevalent cohort 2021
### Patient characteristics - cases

| *Age strata of index patients* | *Characteristic* | |
|---|---|---|
| Children and young people (0-24 years) | Number of persons, n | 14,927 (100.0) |
| | Male, n (%) | 7,903 (52.9) |
| | Age, median (Q1;Q3) | 16.9 (11.8;21.1) |
| | CCI, n (%) | |
| | 0 | 13,074 (87.6) |
| | 1-2 | 1,624 (10.9) |
| | +3 | 229 (1.5) |
| | Education level, n (%) | |
| | No/Unknown education | 4,746 (31.8) |
| | Primary | 8,088 (54.2) |
| | Secondary | 1,916 (12.8) |
| | Tertiary | 177 (1.2) |
| Young adults and adults (25-64 years) | Number of persons, n | 28,621 (100.0) |
| | Male, n (%) | 14,050 (49.1) |
| | Age, median (Q1;Q3) | 45.9 (34.3;55.7) |
| | CCI, n (%) | |
| | 0 | 21,155 (73.9) |
| | 1-2 | 5,937 (20.7) |
| | +3 | 1,529 (5.3) |
| | Education level, n (%) | |
| | No/Unknown education | 905 (3.2) |
| | Primary | 10,021 (35.0) |
| | Secondary | 11,121 (38.9) |
| | Tertiary | 6,574 (23.0) |
| Older people (65+ years) | Number of persons, n | 14,177 (100.0) |
| | Male, n (%) | 7,200 (50.8) |
| | Age, median (Q1;Q3) | 75.3 (70.3;80.7) |
| | CCI, n (%) | |
| | 0 | 4,687 (33.1) |
| | 1-2 | 6,251 (44.1) |
| | +3 | 3,239 (22.8) |
| | Education level, n (%) | |
| | No/Unknown education | 810 (5.7) |
| | Primary | 4,479 (31.6) |
| | Secondary | 6,385 (45.0) |
| | Tertiary | 2,503 (17.7) |

#### Supplementary Table 134
#### Epilepsy - Prevalent cohort 2021
#### Patient characteristics - relatives - pooled

| *Age strata of index patients* | *Closest relative type* | *Characteristic of family relatives by agestrata of index patients* | *Closest family relatives of brain disorder patients* | *Comparison relative* |
|---|---|---|---|---|
| Children and young people (0-24 years) | Fathers | Number of persons, n | 13,311 (100.0) | 13,311 (100.0) |
| | | Male, n (%) | 13,311 (100.0) | 13,311 (100.0) |
| | | Age, median (Q1;Q3) | 49.0 (43.5;54.1) | 49.0 (43.7;53.9) |
| | | CCI, n (%) | | |
| | | 0 | 11,583 (87.0) | 11,895 (89.4) |
| | | 1-2 | 1,533 (11.5) | 1,247 (9.4) |
| | | +3 | 195 (1.5) | 169 (1.3) |
| | | Education level, n (%) | | |
| | | No/Unknown education | 208 (1.6) | 208 (1.6) |
| | | Primary | 2,836 (21.3) | 2,836 (21.3) |
| | | Secondary | 6,644 (49.9) | 6,644 (49.9) |
| | | Tertiary | 3,623 (27.2) | 3,623 (27.2) |
| | | Relation to patient, n (%) | | |
| | | Father | 13,311 (100.0) | 13,311 (100.0) |
| Children and young people (0-24 years) | Mothers | Number of persons, n | 13,833 (100.0) | 13,833 (100.0) |
| | | Male, n (%) | 0 (0.0) | 0 (0.0) |
| | | Age, median (Q1;Q3) | 46.7 (41.3;51.6) | 46.8 (41.6;51.4) |
| | | CCI, n (%) | | |
| | | 0 | 11,836 (85.6) | 12,253 (88.6) |
| | | 1-2 | 1,840 (13.3) | 1,458 (10.5) |
| | | +3 | 157 (1.1) | 122 (0.9) |
| | | Education level, n (%) | | |
| | | No/Unknown education | 208 (1.5) | 208 (1.5) |
| | | Primary | 2,566 (18.5) | 2,566 (18.5) |
| | | Secondary | 5,875 (42.5) | 5,875 (42.5) |
| | | Tertiary | 5,184 (37.5) | 5,184 (37.5) |
| | | Relation to patient, n (%) | | |
| | | Mother | 13,833 (100.0) | 13,833 (100.0) |
| Young adults and adults (25-64 years) | Pooled | Number of persons, n | 28,621 (100.0) | 28,621 (100.0) |
| | | Male, n (%) | 13,546 (47.3) | 13,546 (47.3) |
| | | Age, median (Q1;Q3) | 51.8 (35.2;61.1) | 51.9 (35.3;61.0) |
| | | CCI, n (%) | | |
| | | 0 | 23,268 (81.3) | 23,810 (83.2) |
| | | 1-2 | 4,438 (15.5) | 4,001 (14.0) |
| | | +3 | 915 (3.2) | 810 (2.8) |
| | | Education level, n (%) | | |
| | | No/Unknown education | 1,425 (5.0) | 1,425 (5.0) |
| | | Primary | 7,447 (26.0) | 7,447 (26.0) |
| | | Secondary | 12,923 (45.2) | 12,923 (45.2) |
| | | Tertiary | 6,826 (23.8) | 6,826 (23.8) |
| | | Relation to patient, n (%) | | |
| | | Mother | 5,604 (19.6) | 5,604 (19.6) |
| | | Father | 1,629 (5.7) | 1,629 (5.7) |
| | | Partner | 14,911 (52.1) | 14,911 (52.1) |
| | | Child | 3,640 (12.7) | 3,640 (12.7) |

**Supplementary Table 134**
**Epilepsy - Prevalent cohort 2021**
**Patient characteristics - relatives - pooled**

| *Age strata of index patients* | *Closest relative type* | *Characteristic of family relatives by agestrata of index patients* | *Closest family relatives of brain disorder patients* | *Comparison relative* |
|---|---|---|---|---|
| | | Sibling | 2,837 (9.9) | 2,837 (9.9) |
| Older people (65+ years) | Pooled | Number of persons, n | 14,177 (100.0) | 14,177 (100.0) |
| | | Male, n (%) | 6,311 (44.5) | 6,311 (44.5) |
| | | Age, median (Q1;Q3) | 66.7 (55.0;74.8) | 66.7 (54.9;74.8) |
| | | CCI, n (%) | | |
| | | 0 | 9,888 (69.7) | 10,188 (71.9) |
| | | 1-2 | 3,394 (23.9) | 3,078 (21.7) |
| | | +3 | 895 (6.3) | 911 (6.4) |
| | | Education level, n (%) | | |
| | | No/Unknown education | 418 (2.9) | 418 (2.9) |
| | | Primary | 3,191 (22.5) | 3,191 (22.5) |
| | | Secondary | 6,893 (48.6) | 6,893 (48.6) |
| | | Tertiary | 3,675 (25.9) | 3,675 (25.9) |
| | | Relation to patient, n (%) | | |
| | | Partner | 8,222 (58.0) | 8,222 (58.0) |
| | | Child | 5,640 (39.8) | 5,640 (39.8) |
| | | Sibling | 315 (2.2) | 315 (2.2) |

#### Supplementary Table 135
#### Epilepsy - Prevalent cohort 2021
#### Patient characteristics - relatives - not pooled

| *Age strata of index patients* | *Closest relative type* | *Characteristic of family relatives by agestrata of index patients* | *Closest family relatives of brain disorder patients* | *Comparison relative* |
|---|---|---|---|---|
| Children and young people (0-24 years) | Fathers | Number of persons, n | 13,311 (100.0) | 13,311 (100.0) |
| | | Male, n (%) | 13,311 (100.0) | 13,311 (100.0) |
| | | Age, median (Q1;Q3) | 49.0 (43.5;54.1) | 49.0 (43.7;53.9) |
| | | CCI, n (%) | | |
| | | 0 | 11,583 (87.0) | 11,895 (89.4) |
| | | 1-2 | 1,533 (11.5) | 1,247 (9.4) |
| | | +3 | 195 (1.5) | 169 (1.3) |
| | | Education level, n (%) | | |
| | | No/Unknown education | 208 (1.6) | 208 (1.6) |
| | | Primary | 2,836 (21.3) | 2,836 (21.3) |
| | | Secondary | 6,644 (49.9) | 6,644 (49.9) |
| | | Tertiary | 3,623 (27.2) | 3,623 (27.2) |
| | | Relation to patient, n (%) | | |
| | | Father | 13,311 (100.0) | 13,311 (100.0) |
| Children and young people (0-24 years) | Mothers | Number of persons, n | 13,833 (100.0) | 13,833 (100.0) |
| | | Male, n (%) | 0 (0.0) | 0 (0.0) |
| | | Age, median (Q1;Q3) | 46.7 (41.3;51.6) | 46.8 (41.6;51.4) |
| | | CCI, n (%) | | |
| | | 0 | 11,836 (85.6) | 12,253 (88.6) |
| | | 1-2 | 1,840 (13.3) | 1,458 (10.5) |
| | | +3 | 157 (1.1) | 122 (0.9) |
| | | Education level, n (%) | | |
| | | No/Unknown education | 208 (1.5) | 208 (1.5) |
| | | Primary | 2,566 (18.5) | 2,566 (18.5) |
| | | Secondary | 5,875 (42.5) | 5,875 (42.5) |
| | | Tertiary | 5,184 (37.5) | 5,184 (37.5) |
| | | Relation to patient, n (%) | | |
| | | Mother | 13,833 (100.0) | 13,833 (100.0) |
| Young adults and adults (25-64 years) | Children | Number of persons, n | 3,640 (100.0) | 3,640 (100.0) |
| | | Male, n (%) | 1,868 (51.3) | 1,868 (51.3) |
| | | Age, median (Q1;Q3) | 21.6 (13.0;30.5) | 21.2 (13.0;30.3) |
| | | CCI, n (%) | | |
| | | 0 | 3,402 (93.5) | 3,393 (93.2) |
| | | 1-2 | 233 (6.4) | 235 (6.5) |
| | | +3 | 5 (0.1) | 12 (0.3) |
| | | Education level, n (%) | | |
| | | No/Unknown education | 922 (25.3) | 922 (25.3) |
| | | Primary | 1,421 (39.0) | 1,421 (39.0) |
| | | Secondary | 888 (24.4) | 888 (24.4) |
| | | Tertiary | 409 (11.2) | 409 (11.2) |
| | | Relation to patient, n (%) | | |
| | | Child | 3,640 (100.0) | 3,640 (100.0) |
| Young adults and adults (25-64 years) | Parents | Number of persons, n | 7,233 (100.0) | 7,233 (100.0) |
| | | Male, n (%) | 1,629 (22.5) | 1,629 (22.5) |
| | | Age, median (Q1;Q3) | 65.0 (58.0;73.6) | 64.9 (58.2;73.8) |

#### Supplementary Table 135
#### Epilepsy - Prevalent cohort 2021
#### Patient characteristics - relatives - not pooled

| *Age strata of index patients* | *Closest relative type* | *Characteristic of family relatives by agestrata of index patients* | *Closest family relatives of brain disorder patients* | *Comparison relative* |
|---|---|---|---|---|
| | | CCI, n (%) | | |
| | | 0 | 4,851 (67.1) | 4,989 (69.0) |
| | | 1-2 | 1,878 (26.0) | 1,747 (24.2) |
| | | +3 | 504 (7.0) | 497 (6.9) |
| | | Education level, n (%) | | |
| | | No/Unknown education | 125 (1.7) | 125 (1.7) |
| | | Primary | 2,124 (29.4) | 2,124 (29.4) |
| | | Secondary | 3,482 (48.1) | 3,482 (48.1) |
| | | Tertiary | 1,502 (20.8) | 1,502 (20.8) |
| | | Relation to patient, n (%) | | |
| | | Mother | 5,604 (77.5) | 5,604 (77.5) |
| | | Father | 1,629 (22.5) | 1,629 (22.5) |
| Young adults and adults (25-64 years) | Partners | Number of persons, n | 14,911 (100.0) | 14,911 (100.0) |
| | | Male, n (%) | 8,306 (55.7) | 8,306 (55.7) |
| | | Age, median (Q1;Q3) | 47.4 (35.8;56.7) | 47.3 (36.0;56.5) |
| | | CCI, n (%) | | |
| | | 0 | 12,710 (85.2) | 13,068 (87.6) |
| | | 1-2 | 1,886 (12.6) | 1,601 (10.7) |
| | | +3 | 315 (2.1) | 242 (1.6) |
| | | Education level, n (%) | | |
| | | No/Unknown education | 362 (2.4) | 362 (2.4) |
| | | Primary | 3,184 (21.4) | 3,184 (21.4) |
| | | Secondary | 7,000 (46.9) | 7,000 (46.9) |
| | | Tertiary | 4,365 (29.3) | 4,365 (29.3) |
| | | Relation to patient, n (%) | | |
| | | Partner | 14,911 (100.0) | 14,911 (100.0) |
| Young adults and adults (25-64 years) | Siblings | Number of persons, n | 2,837 (100.0) | 2,837 (100.0) |
| | | Male, n (%) | 1,743 (61.4) | 1,743 (61.4) |
| | | Age, median (Q1;Q3) | 54.6 (43.3;60.3) | 54.8 (42.9;60.1) |
| | | CCI, n (%) | | |
| | | 0 | 2,305 (81.2) | 2,360 (83.2) |
| | | 1-2 | 441 (15.5) | 418 (14.7) |
| | | +3 | 91 (3.2) | 59 (2.1) |
| | | Education level, n (%) | | |
| | | No/Unknown education | 16 (0.6) | 16 (0.6) |
| | | Primary | 718 (25.3) | 718 (25.3) |
| | | Secondary | 1,553 (54.7) | 1,553 (54.7) |
| | | Tertiary | 550 (19.4) | 550 (19.4) |
| | | Relation to patient, n (%) | | |
| | | Sibling | 2,837 (100.0) | 2,837 (100.0) |
| Older people (65+ years) | Children | Number of persons, n | 5,640 (100.0) | 5,640 (100.0) |
| | | Male, n (%) | 2,819 (50.0) | 2,819 (50.0) |
| | | Age, median (Q1;Q3) | 52.4 (46.0;58.1) | 52.3 (46.0;57.9) |
| | | CCI, n (%) | | |
| | | 0 | 4,764 (84.5) | 4,861 (86.2) |

**Supplementary Table 135**
**Epilepsy - Prevalent cohort 2021**
**Patient characteristics - relatives - not pooled**

| *Age strata of index patients* | *Closest relative type* | *Characteristic of family relatives by agestrata of index patients* | *Closest family relatives of brain disorder patients* | *Comparison relative* |
|---|---|---|---|---|
| | | 1-2 | 770 (13.7) | 665 (11.8) |
| | | +3 | 106 (1.9) | 114 (2.0) |
| | | Education level, n (%) | | |
| | | No/Unknown education | 19 (0.3) | 19 (0.3) |
| | | Primary | 1,007 (17.9) | 1,007 (17.9) |
| | | Secondary | 2,720 (48.2) | 2,720 (48.2) |
| | | Tertiary | 1,894 (33.6) | 1,894 (33.6) |
| | | Relation to patient, n (%) | | |
| | | Child | 5,640 (100.0) | 5,640 (100.0) |
| Older people (65+ years) | Partners | Number of persons, n | 8,222 (100.0) | 8,222 (100.0) |
| | | Male, n (%) | 3,313 (40.3) | 3,313 (40.3) |
| | | Age, median (Q1;Q3) | 73.6 (68.7;78.1) | 73.7 (68.9;78.0) |
| | | CCI, n (%) | | |
| | | 0 | 4,910 (59.7) | 5,090 (61.9) |
| | | 1-2 | 2,534 (30.8) | 2,355 (28.6) |
| | | +3 | 778 (9.5) | 777 (9.5) |
| | | Education level, n (%) | | |
| | | No/Unknown education | 396 (4.8) | 396 (4.8) |
| | | Primary | 2,121 (25.8) | 2,121 (25.8) |
| | | Secondary | 3,994 (48.6) | 3,994 (48.6) |
| | | Tertiary | 1,711 (20.8) | 1,711 (20.8) |
| | | Relation to patient, n (%) | | |
| | | Partner | 8,222 (100.0) | 8,222 (100.0) |
| Older people (65+ years) | Siblings | Number of persons, n | 315 (100.0) | 315 (100.0) |
| | | Male, n (%) | 179 (56.8) | 179 (56.8) |
| | | Age, median (Q1;Q3) | 62.6 (60.0;65.3) | 62.7 (59.7;64.6) |
| | | CCI, n (%) | | |
| | | 0 | 214 (67.9) | 237 (75.2) |
| | | 1-2 | 90 (28.6) | 58 (18.4) |
| | | +3 | 11 (3.5) | 20 (6.3) |
| | | Education level, n (%) | | |
| | | No/Unknown education | * | * |
| | | Primary | * | * |
| | | Secondary | 179 (56.8) | 179 (56.8) |
| | | Tertiary | 70 (22.2) | 70 (22.2) |
| | | Relation to patient, n (%) | | |
| | | Sibling | 315 (100.0) | 315 (100.0) |

#### Supplementary Table 136
#### Epilepsy - Prevalent cohort 2021
#### Cost analysis - pooled

| | | | | *Actual costs* | | *Attributable costs* | |
|---|---|---|---|---|---|---|---|
| ***Age strata of index patients*** | ***Closest relative type*** | ***Number of index patients included in cost component calculations*** | ***Cost component*** | ***Total (million EUR)*** | ***Per person (EUR)*** | ***Total (million EUR)*** | ***Per person (EUR)*** |
| Children and young people (0-24 years) | Fathers | 13,311 | Primary sector | 3.0 | 229 | 0.2 | 14 |
| | | 13,311 | Hospital care | 20.6 | 1,550 | 2.4 | 180 |
| | | 13,311 | Filled prescriptions | 2.6 | 197 | 0.6 | 43 |
| | | 12,979 | Lost productivity (income loss) | . | . | 65.2 | 5,021 |
| Children and young people (0-24 years) | Mothers | 13,833 | Primary sector | 5.1 | 367 | 0.5 | 38 |
| | | 13,833 | Hospital care | 25.6 | 1,852 | 5.0 | 361 |
| | | 13,833 | Filled prescriptions | 3.0 | 216 | 0.5 | 38 |
| | | 13,746 | Lost productivity (income loss) | . | . | 44.3 | 3,222 |
| Young adults and adults (25-64 years) | Pooled | 28,621 | Primary sector | 9.4 | 329 | 0.5 | 19 |
| | | 28,621 | Hospital care | 67.8 | 2,379 | 11.7 | 409 |
| | | 28,621 | Filled prescriptions | 8.3 | 292 | 1.2 | 42 |
| | | 21,071 | Lost productivity (income loss) | . | . | 104.9 | 4,978 |
| Older people (65+ years) | Pooled | 14,177 | Primary sector | 6.3 | 445 | 0.5 | 34 |
| | | 14,177 | Hospital care | 45.6 | 3,244 | 3.1 | 221 |
| | | 14,177 | Filled prescriptions | 6.1 | 434 | 0.7 | 51 |
| | | 5,675 | Lost productivity (income loss) | . | . | 27.7 | 4,883 |

#### Supplementary Table 137
#### Epilepsy - Prevalent cohort 2021
#### Cost analysis - not pooled

| *Age strata of index patients* | *Closest relative type* | *Number of index patients included in cost component calculations* | *Cost component* | *Actual costs* | | *Attributable costs* | |
|---|---|---|---|---|---|---|---|
| | | | | *Total (million EUR)* | *Per person (EUR)* | *Total (million EUR)* | *Per person (EUR)* |
| Children and young people (0-24 years) | Fathers | 13,311 | Primary sector | 3.0 | 229 | 0.2 | 14 |
| | | 13,311 | Hospital care | 20.6 | 1,550 | 2.4 | 180 |
| | | 13,311 | Filled prescriptions | 2.6 | 197 | 0.6 | 43 |
| | | 12,979 | Lost productivity (income loss) | . | . | 65.2 | 5,021 |
| Children and young people (0-24 years) | Mothers | 13,833 | Primary sector | 5.1 | 367 | 0.5 | 38 |
| | | 13,833 | Hospital care | 25.6 | 1,852 | 5.0 | 361 |
| | | 13,833 | Filled prescriptions | 3.0 | 216 | 0.5 | 38 |
| | | 13,746 | Lost productivity (income loss) | . | . | 44.3 | 3,222 |
| Young adults and adults (25-64 years) | Children | 3,640 | Primary sector | 0.8 | 232 | 0.1 | 28 |
| | | 3,640 | Hospital care | 6.9 | 1,899 | 2.7 | 748 |
| | | 3,640 | Filled prescriptions | 0.4 | 113 | 0.0 | 8 |
| | | 2,046 | Lost productivity (income loss) | . | . | 3.8 | 1,866 |
| Young adults and adults (25-64 years) | Parents | 7,233 | Primary sector | 3.3 | 457 | 0.2 | 22 |
| | | 7,233 | Hospital care | 25.2 | 3,524 | 3.3 | 467 |
| | | 7,233 | Filled prescriptions | 3.5 | 489 | 0.4 | 54 |
| | | 3,070 | Lost productivity (income loss) | . | . | 9.8 | 3,186 |
| Young adults and adults (25-64 years) | Partners | 14,911 | Primary sector | 4.4 | 296 | 0.3 | 18 |
| | | 14,911 | Hospital care | 29.0 | 1,950 | 4.3 | 287 |
| | | 14,911 | Filled prescriptions | 3.6 | 240 | 0.7 | 47 |
| | | 13,556 | Lost productivity (income loss) | . | . | 81.1 | 5,984 |
| Young adults and adults (25-64 years) | Siblings | 2,837 | Primary sector | 0.9 | 305 | 0.0 | 8 |
| | | 2,837 | Hospital care | 6.6 | 2,352 | 1.3 | 474 |
| | | 2,837 | Filled prescriptions | 0.8 | 296 | 0.1 | 30 |
| | | 2,399 | Lost productivity (income loss) | . | . | 10.2 | 4,241 |
| Older people (65+ years) | Children | 5,640 | Primary sector | 1.8 | 324 | 0.2 | 27 |
| | | 5,640 | Hospital care | 11.5 | 2,040 | 1.9 | 331 |
| | | 5,640 | Filled prescriptions | 1.4 | 250 | 0.1 | 20 |
| | | 5,115 | Lost productivity (income loss) | . | . | 26.6 | 5,197 |
| Older people (65+ years) | Partners | 8,222 | Primary sector | 4.3 | 533 | 0.3 | 39 |
| | | 8,222 | Hospital care | 33.1 | 4,082 | 1.1 | 133 |
| | | 8,222 | Filled prescriptions | 4.6 | 563 | 0.6 | 74 |
| | | 390 | Lost productivity (income loss) | . | . | 1.0 | 2,462 |
| Older people (65+ years) | Siblings | 315 | Primary sector | 0.1 | 369 | 0.0 | 2 |
| | | 315 | Hospital care | 1.0 | 3,146 | 0.2 | 515 |
| | | 315 | Filled prescriptions | 0.1 | 406 | 0.0 | 26 |
| | | 170 | Lost productivity (income loss) | . | . | 0.2 | 988 |

**Supplementary Table 138**
**Headache - Incident cohort 2016-2021**
**Identification of populations of relatives**

| *Agegroup of index patients in matched population* | | *Closest relative type identified* | | *Available control with same type of closest relative* |
|---|---|---|---|---|
| Children and young people (0-24 years) | n = 30,324 | Fathers | n = 24,075 (79%) | 96% |
| | | Mothers | n = 19,926 (66%) | 96% |
| | | No available closest relative | n = 2,852 (9%) | n/a |
| Young adults and adults (25-64 years) | n = 72,494 | Children | n = 10,968 (15%) | 96% |
| | | Parents | n = 10,337 (14%) | 91% |
| | | Partners | n = 42,432 (59%) | 96% |
| | | Siblings | n = 6,344 (9%) | 50% |
| | | Pooled | n = 70,081 (97%) | 91% |
| | | No available closest relative | n = 2,413 (3%) | n/a |
| Older people (65+ years) | n = 7,395 | Children | n = 2,623 (35%) | 94% |
| | | Partners | n = 3,972 (54%) | 96% |
| | | Siblings | n = 130 (2%) | 46% |
| | | Pooled | n = 6,725 (91%) | 94% |
| | | No available closest relative | n = 670 (9%) | n/a |

**Supplementary Table 139**
**Headache - Incident cohort 2016-2021**
**Patient characteristics - cases**

| *Age strata of index patients* | *Characteristic* | |
|---|---|---|
| Children and young people (0-24 years) | Number of persons, n | 26,618 (100.0) |
| | Male, n (%) | 7,759 (29.1) |
| | Age, median (Q1;Q3) | 18.6 (14.6;22.0) |
| | CCI, n (%) | |
| | 0 | 24,639 (92.6) |
| | 1-2 | 1,917 (7.2) |
| | +3 | 62 (0.2) |
| | Education level, n (%) | |
| | No/Unknown education | 4,566 (17.2) |
| | Primary | 14,921 (56.1) |
| | Secondary | 6,332 (23.8) |
| | Tertiary | 799 (3.0) |
| Young adults and adults (25-64 years) | Number of persons, n | 63,911 (100.0) |
| | Male, n (%) | 17,208 (26.9) |
| | Age, median (Q1;Q3) | 41.9 (32.9;50.5) |
| | CCI, n (%) | |
| | 0 | 55,250 (86.4) |
| | 1-2 | 7,710 (12.1) |
| | +3 | 951 (1.5) |
| | Education level, n (%) | |
| | No/Unknown education | 2,635 (4.1) |
| | Primary | 12,656 (19.8) |
| | Secondary | 26,740 (41.8) |
| | Tertiary | 21,880 (34.2) |
| Older people (65+ years) | Number of persons, n | 6,371 (100.0) |
| | Male, n (%) | 2,172 (34.1) |
| | Age, median (Q1;Q3) | 72.4 (68.4;77.2) |
| | CCI, n (%) | |
| | 0 | 3,260 (51.2) |
| | 1-2 | 2,254 (35.4) |
| | +3 | 857 (13.5) |
| | Education level, n (%) | |
| | No/Unknown education | 386 (6.1) |
| | Primary | 1,904 (29.9) |
| | Secondary | 2,915 (45.8) |
| | Tertiary | 1,166 (18.3) |

#### Supplementary Table 140
#### Headache - Incident cohort 2016-2021
#### Patient characteristics - relatives - pooled

| *Age strata of index patients* | *Closest relative type* | *Characteristic of family relatives by agestrata of index patients* | *Closest family relatives of brain disorder patients* | *Comparison relative* |
|---|---|---|---|---|
| Children and young people (0-24 years) | Fathers | Number of persons, n | 23,317 (100.0) | 23,317 (100.0) |
| | | Male, n (%) | 23,317 (100.0) | 23,317 (100.0) |
| | | Age, median (Q1;Q3) | 50.2 (45.6;54.4) | 50.2 (46.0;54.2) |
| | | CCI, n (%) | | |
| | | 0 | 20,268 (86.9) | 20,572 (88.2) |
| | | 1-2 | 2,691 (11.5) | 2,388 (10.2) |
| | | +3 | 358 (1.5) | 357 (1.5) |
| | | Education level, n (%) | | |
| | | No/Unknown education | 326 (1.4) | 326 (1.4) |
| | | Primary | 4,615 (19.8) | 4,615 (19.8) |
| | | Secondary | 12,498 (53.6) | 12,498 (53.6) |
| | | Tertiary | 5,878 (25.2) | 5,878 (25.2) |
| | | Relation to patient, n (%) | | |
| | | Father | 23,317 (100.0) | 23,317 (100.0) |
| Children and young people (0-24 years) | Mothers | Number of persons, n | 19,253 (100.0) | 19,253 (100.0) |
| | | Male, n (%) | 0 (0.0) | 0 (0.0) |
| | | Age, median (Q1;Q3) | 48.0 (43.6;52.1) | 48.0 (43.9;51.9) |
| | | CCI, n (%) | | |
| | | 0 | 16,613 (86.3) | 17,003 (88.3) |
| | | 1-2 | 2,371 (12.3) | 2,035 (10.6) |
| | | +3 | 269 (1.4) | 215 (1.1) |
| | | Education level, n (%) | | |
| | | No/Unknown education | 159 (0.8) | 159 (0.8) |
| | | Primary | 3,262 (16.9) | 3,262 (16.9) |
| | | Secondary | 8,943 (46.4) | 8,943 (46.4) |
| | | Tertiary | 6,889 (35.8) | 6,889 (35.8) |
| | | Relation to patient, n (%) | | |
| | | Mother | 19,253 (100.0) | 19,253 (100.0) |
| Young adults and adults (25-64 years) | Pooled | Number of persons, n | 63,911 (100.0) | 63,911 (100.0) |
| | | Male, n (%) | 40,975 (64.1) | 40,975 (64.1) |
| | | Age, median (Q1;Q3) | 43.3 (30.7;54.9) | 43.4 (30.6;54.9) |
| | | CCI, n (%) | | |
| | | 0 | 55,762 (87.2) | 56,383 (88.2) |
| | | 1-2 | 7,001 (11.0) | 6,491 (10.2) |
| | | +3 | 1,148 (1.8) | 1,037 (1.6) |
| | | Education level, n (%) | | |
| | | No/Unknown education | 5,911 (9.2) | 5,911 (9.2) |
| | | Primary | 14,987 (23.4) | 14,987 (23.4) |
| | | Secondary | 27,188 (42.5) | 27,188 (42.5) |
| | | Tertiary | 15,825 (24.8) | 15,825 (24.8) |
| | | Relation to patient, n (%) | | |
| | | Mother | 6,514 (10.2) | 6,514 (10.2) |
| | | Father | 2,899 (4.5) | 2,899 (4.5) |
| | | Partner | 40,793 (63.8) | 40,793 (63.8) |
| | | Child | 10,530 (16.5) | 10,530 (16.5) |

**Supplementary Table 140**
**Headache - Incident cohort 2016-2021**
**Patient characteristics - relatives - pooled**

| *Age strata of index patients* | *Closest relative type* | *Characteristic of family relatives by agestrata of index patients* | *Closest family relatives of brain disorder patients* | *Comparison relative* |
|---|---|---|---|---|
| | | Sibling | 3,175 (5.0) | 3,175 (5.0) |
| Older people (65+ years) | Pooled | Number of persons, n | 6,371 (100.0) | 6,371 (100.0) |
| | | Male, n (%) | 3,845 (60.4) | 3,845 (60.4) |
| | | Age, median (Q1;Q3) | 66.6 (52.4;73.8) | 66.9 (52.1;73.6) |
| | | CCI, n (%) | | |
| | | 0 | 4,461 (70.0) | 4,574 (71.8) |
| | | 1-2 | 1,432 (22.5) | 1,375 (21.6) |
| | | +3 | 478 (7.5) | 422 (6.6) |
| | | Education level, n (%) | | |
| | | No/Unknown education | 269 (4.2) | 269 (4.2) |
| | | Primary | 1,330 (20.9) | 1,330 (20.9) |
| | | Secondary | 3,191 (50.1) | 3,191 (50.1) |
| | | Tertiary | 1,581 (24.8) | 1,581 (24.8) |
| | | Relation to patient, n (%) | | |
| | | Partner | 3,837 (60.2) | 3,837 (60.2) |
| | | Child | 2,473 (38.8) | 2,473 (38.8) |
| | | Sibling | 61 (1.0) | 61 (1.0) |

#### Supplementary Table 141
#### Headache - Incident cohort 2016-2021
#### Patient characteristics - relatives - not pooled

| *Age strata of index patients* | *Closest relative type* | *Characteristic of family relatives by agestrata of index patients* | *Closest family relatives of brain disorder patients* | *Comparison relative* |
|---|---|---|---|---|
| Children and young people (0-24 years) | Fathers | Number of persons, n | 23,317 (100.0) | 23,317 (100.0) |
| | | Male, n (%) | 23,317 (100.0) | 23,317 (100.0) |
| | | Age, median (Q1;Q3) | 50.2 (45.6;54.4) | 50.2 (46.0;54.2) |
| | | CCI, n (%) | | |
| | | 0 | 20,268 (86.9) | 20,572 (88.2) |
| | | 1-2 | 2,691 (11.5) | 2,388 (10.2) |
| | | +3 | 358 (1.5) | 357 (1.5) |
| | | Education level, n (%) | | |
| | | No/Unknown education | 326 (1.4) | 326 (1.4) |
| | | Primary | 4,615 (19.8) | 4,615 (19.8) |
| | | Secondary | 12,498 (53.6) | 12,498 (53.6) |
| | | Tertiary | 5,878 (25.2) | 5,878 (25.2) |
| | | Relation to patient, n (%) | | |
| | | Father | 23,317 (100.0) | 23,317 (100.0) |
| Children and young people (0-24 years) | Mothers | Number of persons, n | 19,253 (100.0) | 19,253 (100.0) |
| | | Male, n (%) | 0 (0.0) | 0 (0.0) |
| | | Age, median (Q1;Q3) | 48.0 (43.6;52.1) | 48.0 (43.9;51.9) |
| | | CCI, n (%) | | |
| | | 0 | 16,613 (86.3) | 17,003 (88.3) |
| | | 1-2 | 2,371 (12.3) | 2,035 (10.6) |
| | | +3 | 269 (1.4) | 215 (1.1) |
| | | Education level, n (%) | | |
| | | No/Unknown education | 159 (0.8) | 159 (0.8) |
| | | Primary | 3,262 (16.9) | 3,262 (16.9) |
| | | Secondary | 8,943 (46.4) | 8,943 (46.4) |
| | | Tertiary | 6,889 (35.8) | 6,889 (35.8) |
| | | Relation to patient, n (%) | | |
| | | Mother | 19,253 (100.0) | 19,253 (100.0) |
| Young adults and adults (25-64 years) | Children | Number of persons, n | 10,530 (100.0) | 10,530 (100.0) |
| | | Male, n (%) | 5,543 (52.6) | 5,543 (52.6) |
| | | Age, median (Q1;Q3) | 15.8 (9.2;22.7) | 15.8 (8.6;22.5) |
| | | CCI, n (%) | | |
| | | 0 | 9,866 (93.7) | 9,961 (94.6) |
| | | 1-2 | 647 (6.1) | 554 (5.3) |
| | | +3 | 17 (0.2) | 15 (0.1) |
| | | Education level, n (%) | | |
| | | No/Unknown education | 4,192 (39.8) | 4,192 (39.8) |
| | | Primary | 4,030 (38.3) | 4,030 (38.3) |
| | | Secondary | 1,682 (16.0) | 1,682 (16.0) |
| | | Tertiary | 626 (5.9) | 626 (5.9) |
| | | Relation to patient, n (%) | | |
| | | Child | 10,530 (100.0) | 10,530 (100.0) |
| Young adults and adults (25-64 years) | Parents | Number of persons, n | 9,413 (100.0) | 9,413 (100.0) |
| | | Male, n (%) | 2,899 (30.8) | 2,899 (30.8) |
| | | Age, median (Q1;Q3) | 63.3 (57.0;71.3) | 63.2 (57.3;71.4) |

**Supplementary Table 141**
**Headache - Incident cohort 2016-2021**
**Patient characteristics - relatives - not pooled**

| *Age strata of index patients* | *Closest relative type* | *Characteristic of family relatives by agestrata of index patients* | *Closest family relatives of brain disorder patients* | *Comparison relative* |
|---|---|---|---|---|
| | | CCI, n (%) | | |
| | | 0 | 6,603 (70.1) | 6,822 (72.5) |
| | | 1-2 | 2,221 (23.6) | 2,066 (21.9) |
| | | +3 | 589 (6.3) | 525 (5.6) |
| | | Education level, n (%) | | |
| | | No/Unknown education | 174 (1.8) | 174 (1.8) |
| | | Primary | 2,692 (28.6) | 2,692 (28.6) |
| | | Secondary | 4,616 (49.0) | 4,616 (49.0) |
| | | Tertiary | 1,931 (20.5) | 1,931 (20.5) |
| | | Relation to patient, n (%) | | |
| | | Mother | 6,514 (69.2) | 6,514 (69.2) |
| | | Father | 2,899 (30.8) | 2,899 (30.8) |
| Young adults and adults (25-64 years) | Partners | Number of persons, n | 40,793 (100.0) | 40,793 (100.0) |
| | | Male, n (%) | 30,904 (75.8) | 30,904 (75.8) |
| | | Age, median (Q1;Q3) | 43.9 (35.1;52.4) | 44.0 (35.1;52.4) |
| | | CCI, n (%) | | |
| | | 0 | 36,499 (89.5) | 36,760 (90.1) |
| | | 1-2 | 3,799 (9.3) | 3,589 (8.8) |
| | | +3 | 495 (1.2) | 444 (1.1) |
| | | Education level, n (%) | | |
| | | No/Unknown education | 1,535 (3.8) | 1,535 (3.8) |
| | | Primary | 7,451 (18.3) | 7,451 (18.3) |
| | | Secondary | 19,282 (47.3) | 19,282 (47.3) |
| | | Tertiary | 12,525 (30.7) | 12,525 (30.7) |
| | | Relation to patient, n (%) | | |
| | | Partner | 40,793 (100.0) | 40,793 (100.0) |
| Young adults and adults (25-64 years) | Siblings | Number of persons, n | 3,175 (100.0) | 3,175 (100.0) |
| | | Male, n (%) | 1,629 (51.3) | 1,629 (51.3) |
| | | Age, median (Q1;Q3) | 38.1 (28.2;53.2) | 38.3 (28.0;53.1) |
| | | CCI, n (%) | | |
| | | 0 | 2,794 (88.0) | 2,840 (89.4) |
| | | 1-2 | 334 (10.5) | 282 (8.9) |
| | | +3 | 47 (1.5) | 53 (1.7) |
| | | Education level, n (%) | | |
| | | No/Unknown education | 10 (0.3) | 10 (0.3) |
| | | Primary | 814 (25.6) | 814 (25.6) |
| | | Secondary | 1,608 (50.6) | 1,608 (50.6) |
| | | Tertiary | 743 (23.4) | 743 (23.4) |
| | | Relation to patient, n (%) | | |
| | | Sibling | 3,175 (100.0) | 3,175 (100.0) |
| Older people (65+ years) | Children | Number of persons, n | 2,473 (100.0) | 2,473 (100.0) |
| | | Male, n (%) | 1,350 (54.6) | 1,350 (54.6) |
| | | Age, median (Q1;Q3) | 49.0 (43.4;54.9) | 49.0 (43.2;54.5) |
| | | CCI, n (%) | | |
| | | 0 | 2,123 (85.8) | 2,181 (88.2) |

**Supplementary Table 141**
**Headache - Incident cohort 2016-2021**
**Patient characteristics - relatives - not pooled**

| *Age strata of index patients* | *Closest relative type* | *Characteristic of family relatives by agestrata of index patients* | *Closest family relatives of brain disorder patients* | *Comparison relative* |
|---|---|---|---|---|
| | | 1-2 | 314 (12.7) | 254 (10.3) |
| | | +3 | 36 (1.5) | 38 (1.5) |
| | | Education level, n (%) | | |
| | | No/Unknown education | * | * |
| | | Primary | * | * |
| | | Secondary | 1,257 (50.8) | 1,257 (50.8) |
| | | Tertiary | 793 (32.1) | 793 (32.1) |
| | | Relation to patient, n (%) | | |
| | | Child | 2,473 (100.0) | 2,473 (100.0) |
| Older people (65+ years) | Partners | Number of persons, n | 3,837 (100.0) | 3,837 (100.0) |
| | | Male, n (%) | 2,458 (64.1) | 2,458 (64.1) |
| | | Age, median (Q1;Q3) | 72.3 (68.0;76.7) | 72.2 (68.2;76.5) |
| | | CCI, n (%) | | |
| | | 0 | 2,291 (59.7) | 2,349 (61.2) |
| | | 1-2 | 1,105 (28.8) | 1,109 (28.9) |
| | | +3 | 441 (11.5) | 379 (9.9) |
| | | Education level, n (%) | | |
| | | No/Unknown education | 264 (6.9) | 264 (6.9) |
| | | Primary | 898 (23.4) | 898 (23.4) |
| | | Secondary | 1,900 (49.5) | 1,900 (49.5) |
| | | Tertiary | 775 (20.2) | 775 (20.2) |
| | | Relation to patient, n (%) | | |
| | | Partner | 3,837 (100.0) | 3,837 (100.0) |
| Older people (65+ years) | Siblings | Number of persons, n | 61 (100.0) | 61 (100.0) |
| | | Male, n (%) | 37 (60.7) | 37 (60.7) |
| | | Age, median (Q1;Q3) | 61.9 (58.5;66.6) | 63.6 (58.5;66.5) |
| | | CCI, n (%) | | |
| | | 0 | 47 (77.0) | 44 (72.1) |
| | | 1-2 | * | 12 (19.7) |
| | | +3 | * | 5 (8.2) |
| | | Education level, n (%) | | |
| | | No/Unknown education | * | * |
| | | Primary | 13 (21.3) | 13 (21.3) |
| | | Secondary | 34 (55.7) | 34 (55.7) |
| | | Tertiary | * | * |
| | | Relation to patient, n (%) | | |
| | | Sibling | 61 (100.0) | 61 (100.0) |

#### Supplementary Table 142
#### Headache - Incident cohort 2016-2021
#### Cost analysis - pooled

| *Age strata of index patients* | *Closest relative type* | *Number of index patients included in cost component calculations* | *Cost component* | *Actual costs* | | *Attributable costs* | |
|---|---|---|---|---|---|---|---|
| | | | | *Total (million EUR)* | *Per person (EUR)* | *Total (million EUR)* | *Per person (EUR)* |
| Children and young people (0-24 years) | Fathers | 23,317 | Primary sector | 5.5 | 236 | 0.5 | 22 |
| | | 23,317 | Hospital care | 36.8 | 1,582 | 3.5 | 150 |
| | | 23,317 | Filled prescriptions | 4.5 | 195 | 0.6 | 25 |
| | | 22,483 | Lost productivity (income loss) | . | . | 24.6 | 1,093 |
| Children and young people (0-24 years) | Mothers | 19,253 | Primary sector | 6.7 | 347 | 0.7 | 35 |
| | | 19,253 | Hospital care | 35.6 | 1,850 | 6.9 | 360 |
| | | 19,253 | Filled prescriptions | 3.9 | 203 | 0.7 | 38 |
| | | 19,119 | Lost productivity (income loss) | . | . | 36.6 | 1,914 |
| Young adults and adults (25-64 years) | Pooled | 63,911 | Primary sector | 17.1 | 269 | 1.6 | 24 |
| | | 63,911 | Hospital care | 104.9 | 1,647 | 6.8 | 107 |
| | | 63,911 | Filled prescriptions | 12.5 | 195 | 1.2 | 19 |
| | | 50,081 | Lost productivity (income loss) | . | . | 65.1 | 1,300 |
| Older people (65+ years) | Pooled | 6,371 | Primary sector | 2.8 | 437 | 0.2 | 39 |
| | | 6,371 | Hospital care | 20.2 | 3,192 | -0.4 | -56 |
| | | 6,371 | Filled prescriptions | 2.6 | 418 | 0.1 | 19 |
| | | 2,533 | Lost productivity (income loss) | . | . | 6.9 | 2,738 |

### Supplementary Table 143
### Headache - Incident cohort 2016-2021
### Cost analysis - not pooled

| Age strata of index patients | Closest relative type | Number of index patients included in cost component calculations | Cost component | Actual costs: Total (million EUR) | Actual costs: Per person (EUR) | Attributable costs: Total (million EUR) | Attributable costs: Per person (EUR) |
|---|---|---|---|---|---|---|---|
| Children and young people (0-24 years) | Fathers | 23,317 | Primary sector | 5.5 | 236 | 0.5 | 22 |
| | | 23,317 | Hospital care | 36.8 | 1,582 | 3.5 | 150 |
| | | 23,317 | Filled prescriptions | 4.5 | 195 | 0.6 | 25 |
| | | 22,483 | Lost productivity (income loss) | . | . | 24.6 | 1,093 |
| Children and young people (0-24 years) | Mothers | 19,253 | Primary sector | 6.7 | 347 | 0.7 | 35 |
| | | 19,253 | Hospital care | 35.6 | 1,850 | 6.9 | 360 |
| | | 19,253 | Filled prescriptions | 3.9 | 203 | 0.7 | 38 |
| | | 19,119 | Lost productivity (income loss) | . | . | 36.6 | 1,914 |
| Young adults and adults (25-64 years) | Children | 10,530 | Primary sector | 2.0 | 193 | 0.2 | 22 |
| | | 10,530 | Hospital care | 11.0 | 1,046 | 1.8 | 176 |
| | | 10,530 | Filled prescriptions | 1.0 | 91 | 0.1 | 12 |
| | | 3,962 | Lost productivity (income loss) | . | . | 2.1 | 532 |
| Young adults and adults (25-64 years) | Parents | 9,413 | Primary sector | 4.0 | 426 | 0.2 | 24 |
| | | 9,413 | Hospital care | 30.5 | 3,270 | 3.7 | 392 |
| | | 9,413 | Filled prescriptions | 4.3 | 460 | 0.5 | 56 |
| | | 4,350 | Lost productivity (income loss) | . | . | 12.1 | 2,792 |
| Young adults and adults (25-64 years) | Partners | 40,793 | Primary sector | 10.3 | 252 | 1.1 | 27 |
| | | 40,793 | Hospital care | 57.5 | 1,412 | 1.8 | 45 |
| | | 40,793 | Filled prescriptions | 6.6 | 162 | 0.5 | 13 |
| | | 38,821 | Lost productivity (income loss) | . | . | 52.3 | 1,347 |
| Young adults and adults (25-64 years) | Siblings | 3,175 | Primary sector | 0.9 | 274 | 0.0 | 1 |
| | | 3,175 | Hospital care | 5.9 | 1,868 | -0.5 | -163 |
| | | 3,175 | Filled prescriptions | 0.6 | 187 | 0.0 | 9 |
| | | 2,948 | Lost productivity (income loss) | . | . | -1.5 | -493 |
| Older people (65+ years) | Children | 2,473 | Primary sector | 0.7 | 295 | 0.1 | 32 |
| | | 2,473 | Hospital care | 5.2 | 2,119 | 1.1 | 452 |
| | | 2,473 | Filled prescriptions | 0.6 | 230 | -0.0 | -4 |
| | | 2,349 | Lost productivity (income loss) | . | . | 8.6 | 3,665 |
| Older people (65+ years) | Partners | 3,837 | Primary sector | 2.0 | 529 | 0.2 | 41 |
| | | 3,837 | Hospital care | 14.8 | 3,906 | -0.9 | -230 |
| | | 3,837 | Filled prescriptions | 2.1 | 542 | 0.1 | 36 |
| | | 156 | Lost productivity (income loss) | . | . | -1.7 | -11,072 |
| Older people (65+ years) | Siblings | 61 | Primary sector | 0.0 | 443 | 0.0 | 149 |
| | | 61 | Hospital care | 0.1 | 2,154 | -0.6 | -9,846 |
| | | 61 | Filled prescriptions | 0.0 | 323 | -0.0 | -70 |
| | | 28 | Lost productivity (income loss) | . | . | 0.1 | 1,857 |

**Supplementary Table 144**
**Headache - Prevalent cohort 2021**
**Identification of populations of relatives**

| *Agegroup of index patients in matched population* | | *Closest relative type identified* | | *Available control with same type of closest relative* |
|---|---|---|---|---|
| Children and young people (0-24 years) | n = 34,797 | Fathers | n = 27,689 (80%) | 97% |
| | | Mothers | n = 22,382 (64%) | 96% |
| | | No available closest relative | n = 3,059 (9%) | n/a |
| Young adults and adults (25-64 years) | n = 302,175 | Children | n = 51,252 (17%) | 95% |
| | | Parents | n = 38,453 (13%) | 89% |
| | | Partners | n = 181,066 (60%) | 96% |
| | | Siblings | n = 26,049 (9%) | 54% |
| | | Pooled | n = 296,820 (98%) | 91% |
| | | No available closest relative | n = 5,355 (2%) | n/a |
| Older people (65+ years) | n = 94,824 | Children | n = 31,591 (33%) | 95% |
| | | Partners | n = 51,758 (55%) | 96% |
| | | Siblings | n = 2,492 (3%) | 48% |
| | | Pooled | n = 85,841 (91%) | 94% |
| | | No available closest relative | n = 8,983 (9%) | n/a |

**Supplementary Table 145**
**Headache - Prevalent cohort 2021**
**Patient characteristics - cases**

| *Age strata of index patients* | *Characteristic* | |
|---|---|---|
| Children and young people (0-24 years) | Number of persons, n | 30,831 (100.0) |
| | Male, n (%) | 11,319 (36.7) |
| | Age, median (Q1;Q3) | 20.6 (17.4;23.0) |
| | CCI, n (%) | |
| | 0 | 28,287 (91.7) |
| | 1-2 | 2,469 (8.0) |
| | +3 | 75 (0.2) |
| | Education level, n (%) | |
| | No/Unknown education | 2,235 (7.2) |
| | Primary | 17,395 (56.4) |
| | Secondary | 10,049 (32.6) |
| | Tertiary | 1,152 (3.7) |
| Young adults and adults (25-64 years) | Number of persons, n | 272,292 (100.0) |
| | Male, n (%) | 63,769 (23.4) |
| | Age, median (Q1;Q3) | 48.6 (38.4;56.4) |
| | CCI, n (%) | |
| | 0 | 225,571 (82.8) |
| | 1-2 | 41,224 (15.1) |
| | +3 | 5,497 (2.0) |
| | Education level, n (%) | |
| | No/Unknown education | 5,907 (2.2) |
| | Primary | 51,021 (18.7) |
| | Secondary | 122,965 (45.2) |
| | Tertiary | 92,399 (33.9) |
| Older people (65+ years) | Number of persons, n | 81,235 (100.0) |
| | Male, n (%) | 15,748 (19.4) |
| | Age, median (Q1;Q3) | 72.2 (68.3;76.9) |
| | CCI, n (%) | |
| | 0 | 46,731 (57.5) |
| | 1-2 | 26,018 (32.0) |
| | +3 | 8,486 (10.4) |
| | Education level, n (%) | |
| | No/Unknown education | 4,222 (5.2) |
| | Primary | 22,447 (27.6) |
| | Secondary | 36,516 (45.0) |
| | Tertiary | 18,050 (22.2) |

**Supplementary Table 146**
**Headache - Prevalent cohort 2021**
**Patient characteristics - relatives - pooled**

| *Age strata of index patients* | *Closest relative type* | *Characteristic of family relatives by agestrata of index patients* | *Closest family relatives of brain disorder patients* | *Comparison relative* |
|---|---|---|---|---|
| Children and young people (0-24 years) | Fathers | Number of persons, n | 26,878 (100.0) | 26,878 (100.0) |
| | | Male, n (%) | 26,878 (100.0) | 26,878 (100.0) |
| | | Age, median (Q1;Q3) | 52.1 (47.9;56.0) | 52.1 (48.2;55.8) |
| | | CCI, n (%) | | |
| | | 0 | 23,078 (85.9) | 23,391 (87.0) |
| | | 1-2 | 3,309 (12.3) | 3,048 (11.3) |
| | | +3 | 491 (1.8) | 439 (1.6) |
| | | Education level, n (%) | | |
| | | No/Unknown education | 273 (1.0) | 273 (1.0) |
| | | Primary | 5,055 (18.8) | 5,055 (18.8) |
| | | Secondary | 14,719 (54.8) | 14,719 (54.8) |
| | | Tertiary | 6,831 (25.4) | 6,831 (25.4) |
| | | Relation to patient, n (%) | | |
| | | Father | 26,878 (100.0) | 26,878 (100.0) |
| Children and young people (0-24 years) | Mothers | Number of persons, n | 21,692 (100.0) | 21,692 (100.0) |
| | | Male, n (%) | 0 (0.0) | 0 (0.0) |
| | | Age, median (Q1;Q3) | 49.8 (45.9;53.7) | 49.9 (46.2;53.5) |
| | | CCI, n (%) | | |
| | | 0 | 18,525 (85.4) | 19,020 (87.7) |
| | | 1-2 | 2,878 (13.3) | 2,425 (11.2) |
| | | +3 | 289 (1.3) | 247 (1.1) |
| | | Education level, n (%) | | |
| | | No/Unknown education | 133 (0.6) | 133 (0.6) |
| | | Primary | 3,338 (15.4) | 3,338 (15.4) |
| | | Secondary | 10,052 (46.3) | 10,052 (46.3) |
| | | Tertiary | 8,169 (37.7) | 8,169 (37.7) |
| | | Relation to patient, n (%) | | |
| | | Mother | 21,692 (100.0) | 21,692 (100.0) |
| Young adults and adults (25-64 years) | Pooled | Number of persons, n | 272,292 (100.0) | 272,292 (100.0) |
| | | Male, n (%) | 182,894 (67.2) | 182,894 (67.2) |
| | | Age, median (Q1;Q3) | 48.9 (33.5;59.3) | 48.9 (33.4;59.3) |
| | | CCI, n (%) | | |
| | | 0 | 230,322 (84.6) | 233,370 (85.7) |
| | | 1-2 | 35,506 (13.0) | 32,986 (12.1) |
| | | +3 | 6,464 (2.4) | 5,936 (2.2) |
| | | Education level, n (%) | | |
| | | No/Unknown education | 16,537 (6.1) | 16,537 (6.1) |
| | | Primary | 63,326 (23.3) | 63,326 (23.3) |
| | | Secondary | 125,614 (46.1) | 125,614 (46.1) |
| | | Tertiary | 66,815 (24.5) | 66,815 (24.5) |
| | | Relation to patient, n (%) | | |
| | | Mother | 24,180 (8.9) | 24,180 (8.9) |
| | | Father | 10,218 (3.8) | 10,218 (3.8) |
| | | Partner | 174,753 (64.2) | 174,753 (64.2) |
| | | Child | 48,947 (18.0) | 48,947 (18.0) |

**Supplementary Table 146**
**Headache - Prevalent cohort 2021**
**Patient characteristics - relatives - pooled**

| *Age strata of index patients* | *Closest relative type* | *Characteristic of family relatives by agestrata of index patients* | *Closest family relatives of brain disorder patients* | *Comparison relative* |
|---|---|---|---|---|
| | | Sibling | 14,194 (5.2) | 14,194 (5.2) |
| Older people (65+ years) | Pooled | Number of persons, n | 81,235 (100.0) | 81,235 (100.0) |
| | | Male, n (%) | 56,971 (70.1) | 56,971 (70.1) |
| | | Age, median (Q1;Q3) | 67.8 (53.6;74.3) | 67.9 (53.4;74.2) |
| | | CCI, n (%) | | |
| | | 0 | 55,893 (68.8) | 57,195 (70.4) |
| | | 1-2 | 19,284 (23.7) | 18,285 (22.5) |
| | | +3 | 6,058 (7.5) | 5,755 (7.1) |
| | | Education level, n (%) | | |
| | | No/Unknown education | 3,699 (4.6) | 3,699 (4.6) |
| | | Primary | 15,313 (18.9) | 15,313 (18.9) |
| | | Secondary | 40,904 (50.4) | 40,904 (50.4) |
| | | Tertiary | 21,319 (26.2) | 21,319 (26.2) |
| | | Relation to patient, n (%) | | |
| | | Partner | 49,892 (61.4) | 49,892 (61.4) |
| | | Child | 30,133 (37.1) | 30,133 (37.1) |
| | | Sibling | 1,210 (1.5) | 1,210 (1.5) |

#### Supplementary Table 147
#### Headache - Prevalent cohort 2021
#### Patient characteristics - relatives - not pooled

| *Age strata of index patients* | *Closest relative type* | *Characteristic of family relatives by agestrata of index patients* | *Closest family relatives of brain disorder patients* | *Comparison relative* |
|---|---|---|---|---|
| Children and young people (0-24 years) | Fathers | Number of persons, n | 26,878 (100.0) | 26,878 (100.0) |
| | | Male, n (%) | 26,878 (100.0) | 26,878 (100.0) |
| | | Age, median (Q1;Q3) | 52.1 (47.9;56.0) | 52.1 (48.2;55.8) |
| | | CCI, n (%) | | |
| | | 0 | 23,078 (85.9) | 23,391 (87.0) |
| | | 1-2 | 3,309 (12.3) | 3,048 (11.3) |
| | | +3 | 491 (1.8) | 439 (1.6) |
| | | Education level, n (%) | | |
| | | No/Unknown education | 273 (1.0) | 273 (1.0) |
| | | Primary | 5,055 (18.8) | 5,055 (18.8) |
| | | Secondary | 14,719 (54.8) | 14,719 (54.8) |
| | | Tertiary | 6,831 (25.4) | 6,831 (25.4) |
| | | Relation to patient, n (%) | | |
| | | Father | 26,878 (100.0) | 26,878 (100.0) |
| Children and young people (0-24 years) | Mothers | Number of persons, n | 21,692 (100.0) | 21,692 (100.0) |
| | | Male, n (%) | 0 (0.0) | 0 (0.0) |
| | | Age, median (Q1;Q3) | 49.8 (45.9;53.7) | 49.9 (46.2;53.5) |
| | | CCI, n (%) | | |
| | | 0 | 18,525 (85.4) | 19,020 (87.7) |
| | | 1-2 | 2,878 (13.3) | 2,425 (11.2) |
| | | +3 | 289 (1.3) | 247 (1.1) |
| | | Education level, n (%) | | |
| | | No/Unknown education | 133 (0.6) | 133 (0.6) |
| | | Primary | 3,338 (15.4) | 3,338 (15.4) |
| | | Secondary | 10,052 (46.3) | 10,052 (46.3) |
| | | Tertiary | 8,169 (37.7) | 8,169 (37.7) |
| | | Relation to patient, n (%) | | |
| | | Mother | 21,692 (100.0) | 21,692 (100.0) |
| Young adults and adults (25-64 years) | Children | Number of persons, n | 48,947 (100.0) | 48,947 (100.0) |
| | | Male, n (%) | 26,160 (53.4) | 26,160 (53.4) |
| | | Age, median (Q1;Q3) | 20.3 (13.4;28.8) | 20.0 (13.4;28.5) |
| | | CCI, n (%) | | |
| | | 0 | 45,794 (93.6) | 46,374 (94.7) |
| | | 1-2 | 3,057 (6.2) | 2,454 (5.0) |
| | | +3 | 96 (0.2) | 119 (0.2) |
| | | Education level, n (%) | | |
| | | No/Unknown education | 11,781 (24.1) | 11,781 (24.1) |
| | | Primary | 20,077 (41.0) | 20,077 (41.0) |
| | | Secondary | 11,454 (23.4) | 11,454 (23.4) |
| | | Tertiary | 5,635 (11.5) | 5,635 (11.5) |
| | | Relation to patient, n (%) | | |
| | | Child | 48,947 (100.0) | 48,947 (100.0) |
| Young adults and adults (25-64 years) | Parents | Number of persons, n | 34,398 (100.0) | 34,398 (100.0) |
| | | Male, n (%) | 10,218 (29.7) | 10,218 (29.7) |
| | | Age, median (Q1;Q3) | 68.6 (60.3;76.4) | 68.7 (60.4;76.5) |

**Supplementary Table 147**
**Headache - Prevalent cohort 2021**
**Patient characteristics - relatives - not pooled**

| *Age strata of index patients* | *Closest relative type* | *Characteristic of family relatives by agestrata of index patients* | *Closest family relatives of brain disorder patients* | *Comparison relative* |
|---|---|---|---|---|
| | | CCI, n (%) | | |
| | | 0 | 22,272 (64.7) | 22,958 (66.7) |
| | | 1-2 | 9,363 (27.2) | 8,925 (25.9) |
| | | +3 | 2,763 (8.0) | 2,515 (7.3) |
| | | Education level, n (%) | | |
| | | No/Unknown education | 541 (1.6) | 541 (1.6) |
| | | Primary | 10,388 (30.2) | 10,388 (30.2) |
| | | Secondary | 17,234 (50.1) | 17,234 (50.1) |
| | | Tertiary | 6,235 (18.1) | 6,235 (18.1) |
| | | Relation to patient, n (%) | | |
| | | Mother | 24,180 (70.3) | 24,180 (70.3) |
| | | Father | 10,218 (29.7) | 10,218 (29.7) |
| Young adults and adults (25-64 years) | Partners | Number of persons, n | 174,753 (100.0) | 174,753 (100.0) |
| | | Male, n (%) | 138,916 (79.5) | 138,916 (79.5) |
| | | Age, median (Q1;Q3) | 50.2 (40.1;58.2) | 50.3 (40.2;58.1) |
| | | CCI, n (%) | | |
| | | 0 | 150,386 (86.1) | 152,037 (87.0) |
| | | 1-2 | 21,117 (12.1) | 19,737 (11.3) |
| | | +3 | 3,250 (1.9) | 2,979 (1.7) |
| | | Education level, n (%) | | |
| | | No/Unknown education | 4,170 (2.4) | 4,170 (2.4) |
| | | Primary | 29,580 (16.9) | 29,580 (16.9) |
| | | Secondary | 89,375 (51.1) | 89,375 (51.1) |
| | | Tertiary | 51,628 (29.5) | 51,628 (29.5) |
| | | Relation to patient, n (%) | | |
| | | Partner | 174,753 (100.0) | 174,753 (100.0) |
| Young adults and adults (25-64 years) | Siblings | Number of persons, n | 14,194 (100.0) | 14,194 (100.0) |
| | | Male, n (%) | 7,600 (53.5) | 7,600 (53.5) |
| | | Age, median (Q1;Q3) | 52.6 (34.7;59.5) | 52.6 (34.7;59.4) |
| | | CCI, n (%) | | |
| | | 0 | 11,870 (83.6) | 12,001 (84.5) |
| | | 1-2 | 1,969 (13.9) | 1,870 (13.2) |
| | | +3 | 355 (2.5) | 323 (2.3) |
| | | Education level, n (%) | | |
| | | No/Unknown education | 45 (0.3) | 45 (0.3) |
| | | Primary | 3,281 (23.1) | 3,281 (23.1) |
| | | Secondary | 7,551 (53.2) | 7,551 (53.2) |
| | | Tertiary | 3,317 (23.4) | 3,317 (23.4) |
| | | Relation to patient, n (%) | | |
| | | Sibling | 14,194 (100.0) | 14,194 (100.0) |
| Older people (65+ years) | Children | Number of persons, n | 30,133 (100.0) | 30,133 (100.0) |
| | | Male, n (%) | 16,658 (55.3) | 16,658 (55.3) |
| | | Age, median (Q1;Q3) | 49.4 (43.6;55.5) | 49.3 (43.6;55.3) |
| | | CCI, n (%) | | |
| | | 0 | 25,883 (85.9) | 26,369 (87.5) |

**Supplementary Table 147**
**Headache - Prevalent cohort 2021**
**Patient characteristics - relatives - not pooled**

| *Age strata of index patients* | *Closest relative type* | *Characteristic of family relatives by agestrata of index patients* | *Closest family relatives of brain disorder patients* | *Comparison relative* |
|---|---|---|---|---|
| | | 1-2 | 3,708 (12.3) | 3,291 (10.9) |
| | | +3 | 542 (1.8) | 473 (1.6) |
| | | Education level, n (%) | | |
| | | No/Unknown education | 49 (0.2) | 49 (0.2) |
| | | Primary | 5,049 (16.8) | 5,049 (16.8) |
| | | Secondary | 14,360 (47.7) | 14,360 (47.7) |
| | | Tertiary | 10,675 (35.4) | 10,675 (35.4) |
| | | Relation to patient, n (%) | | |
| | | Child | 30,133 (100.0) | 30,133 (100.0) |
| Older people (65+ years) | Partners | Number of persons, n | 49,892 (100.0) | 49,892 (100.0) |
| | | Male, n (%) | 39,655 (79.5) | 39,655 (79.5) |
| | | Age, median (Q1;Q3) | 72.9 (68.8;76.9) | 72.8 (68.9;76.7) |
| | | CCI, n (%) | | |
| | | 0 | 29,108 (58.3) | 29,893 (59.9) |
| | | 1-2 | 15,331 (30.7) | 14,771 (29.6) |
| | | +3 | 5,453 (10.9) | 5,228 (10.5) |
| | | Education level, n (%) | | |
| | | No/Unknown education | 3,644 (7.3) | 3,644 (7.3) |
| | | Primary | 10,079 (20.2) | 10,079 (20.2) |
| | | Secondary | 25,802 (51.7) | 25,802 (51.7) |
| | | Tertiary | 10,367 (20.8) | 10,367 (20.8) |
| | | Relation to patient, n (%) | | |
| | | Partner | 49,892 (100.0) | 49,892 (100.0) |
| Older people (65+ years) | Siblings | Number of persons, n | 1,210 (100.0) | 1,210 (100.0) |
| | | Male, n (%) | 658 (54.4) | 658 (54.4) |
| | | Age, median (Q1;Q3) | 62.8 (59.8;65.2) | 62.9 (60.0;65.2) |
| | | CCI, n (%) | | |
| | | 0 | 902 (74.5) | 933 (77.1) |
| | | 1-2 | 245 (20.2) | 223 (18.4) |
| | | +3 | 63 (5.2) | 54 (4.5) |
| | | Education level, n (%) | | |
| | | No/Unknown education | 6 (0.5) | 6 (0.5) |
| | | Primary | 185 (15.3) | 185 (15.3) |
| | | Secondary | 742 (61.3) | 742 (61.3) |
| | | Tertiary | 277 (22.9) | 277 (22.9) |
| | | Relation to patient, n (%) | | |
| | | Sibling | 1,210 (100.0) | 1,210 (100.0) |

### Supplementary Table 148
### Headache - Prevalent cohort 2021
### Cost analysis - pooled

| *Age strata of index patients* | *Closest relative type* | *Number of index patients included in cost component calculations* | *Cost component* | *Actual costs* | | *Attributable costs* | |
|---|---|---|---|---|---|---|---|
| | | | | *Total (million EUR)* | *Per person (EUR)* | *Total (million EUR)* | *Per person (EUR)* |
| Children and young people (0-24 years) | Fathers | 26,878 | Primary sector | 6.6 | 246 | 0.5 | 19 |
| | | 26,878 | Hospital care | 44.5 | 1,658 | 5.0 | 188 |
| | | 26,878 | Filled prescriptions | 5.8 | 217 | 1.1 | 40 |
| | | 25,818 | Lost productivity (income loss) | . | . | 42.9 | 1,662 |
| Children and young people (0-24 years) | Mothers | 21,692 | Primary sector | 8.0 | 368 | 1.1 | 51 |
| | | 21,692 | Hospital care | 39.2 | 1,810 | 8.8 | 406 |
| | | 21,692 | Filled prescriptions | 4.9 | 225 | 1.0 | 47 |
| | | 21,540 | Lost productivity (income loss) | . | . | 43.9 | 2,038 |
| Young adults and adults (25-64 years) | Pooled | 272,292 | Primary sector | 78.1 | 288 | 6.2 | 23 |
| | | 272,292 | Hospital care | 511.9 | 1,886 | 40.2 | 148 |
| | | 272,292 | Filled prescriptions | 64.7 | 238 | 6.1 | 23 |
| | | 203,170 | Lost productivity (income loss) | . | . | 223.8 | 1,101 |
| Older people (65+ years) | Pooled | 81,235 | Primary sector | 34.5 | 428 | 2.4 | 30 |
| | | 81,235 | Hospital care | 269.7 | 3,349 | 11.9 | 148 |
| | | 81,235 | Filled prescriptions | 35.2 | 437 | 3.1 | 38 |
| | | 30,926 | Lost productivity (income loss) | . | . | 69.5 | 2,246 |

### Supplementary Table 149
### Headache - Prevalent cohort 2021
### Cost analysis - not pooled

| Age strata of index patients | Closest relative type | Number of index patients included in cost component calculations | Cost component | Actual costs: Total (million EUR) | Actual costs: Per person (EUR) | Attributable costs: Total (million EUR) | Attributable costs: Per person (EUR) |
|---|---|---|---|---|---|---|---|
| Children and young people (0-24 years) | Fathers | 26,878 | Primary sector | 6.6 | 246 | 0.5 | 19 |
| | | 26,878 | Hospital care | 44.5 | 1,658 | 5.0 | 188 |
| | | 26,878 | Filled prescriptions | 5.8 | 217 | 1.1 | 40 |
| | | 25,818 | Lost productivity (income loss) | . | . | 42.9 | 1,662 |
| Children and young people (0-24 years) | Mothers | 21,692 | Primary sector | 8.0 | 368 | 1.1 | 51 |
| | | 21,692 | Hospital care | 39.2 | 1,810 | 8.8 | 406 |
| | | 21,692 | Filled prescriptions | 4.9 | 225 | 1.0 | 47 |
| | | 21,540 | Lost productivity (income loss) | . | . | 43.9 | 2,038 |
| Young adults and adults (25-64 years) | Children | 48,947 | Primary sector | 10.7 | 219 | 1.3 | 27 |
| | | 48,947 | Hospital care | 60.2 | 1,233 | 3.1 | 64 |
| | | 48,947 | Filled prescriptions | 5.6 | 115 | 0.7 | 13 |
| | | 26,220 | Lost productivity (income loss) | . | . | 21.4 | 816 |
| Young adults and adults (25-64 years) | Parents | 34,398 | Primary sector | 16.2 | 477 | 1.4 | 41 |
| | | 34,398 | Hospital care | 124.3 | 3,663 | 8.3 | 244 |
| | | 34,398 | Filled prescriptions | 18.4 | 541 | 2.5 | 74 |
| | | 11,213 | Lost productivity (income loss) | . | . | 23.1 | 2,062 |
| Young adults and adults (25-64 years) | Partners | 174,753 | Primary sector | 46.8 | 268 | 3.2 | 18 |
| | | 174,753 | Hospital care | 294.4 | 1,688 | 25.5 | 146 |
| | | 174,753 | Filled prescriptions | 37.0 | 212 | 2.7 | 16 |
| | | 153,590 | Lost productivity (income loss) | . | . | 172.6 | 1,124 |
| Young adults and adults (25-64 years) | Siblings | 14,194 | Primary sector | 4.4 | 309 | 0.3 | 18 |
| | | 14,194 | Hospital care | 33.0 | 2,331 | 3.4 | 237 |
| | | 14,194 | Filled prescriptions | 3.7 | 262 | 0.2 | 16 |
| | | 12,147 | Lost productivity (income loss) | . | . | 6.6 | 545 |
| Older people (65+ years) | Children | 30,133 | Primary sector | 8.9 | 296 | 0.7 | 23 |
| | | 30,133 | Hospital care | 55.5 | 1,847 | 6.2 | 206 |
| | | 30,133 | Filled prescriptions | 6.9 | 230 | 0.6 | 21 |
| | | 28,474 | Lost productivity (income loss) | . | . | 72.6 | 2,550 |
| Older people (65+ years) | Partners | 49,892 | Primary sector | 25.1 | 510 | 1.8 | 36 |
| | | 49,892 | Hospital care | 211.2 | 4,286 | 5.9 | 121 |
| | | 49,892 | Filled prescriptions | 27.8 | 565 | 2.4 | 49 |
| | | 1,828 | Lost productivity (income loss) | . | . | -6.0 | -3,281 |
| Older people (65+ years) | Siblings | 1,210 | Primary sector | 0.5 | 380 | -0.0 | -1 |
| | | 1,210 | Hospital care | 3.0 | 2,516 | -0.2 | -201 |
| | | 1,210 | Filled prescriptions | 0.5 | 375 | 0.0 | 28 |
| | | 624 | Lost productivity (income loss) | . | . | 2.9 | 4,600 |

**Supplementary Table 150**
**Infections of the CNS - Incident cohort 2016-2021**
**Identification of populations of relatives**

| ***Agegroup of index patients in matched population*** | | ***Closest relative type identified*** | | ***Available control with same type of closest relative*** |
|---|---|---|---|---|
| Children and young people (0-24 years) | n = 1,817 | Fathers | n = 1,147 (63%) | 97% |
| | | Mothers | n = 1,241 (68%) | 97% |
| | | No available closest relative | n = 512 (28%) | n/a |
| Young adults and adults (25-64 years) | n = 3,353 | Children | n = 453 (14%) | 92% |
| | | Parents | n = 438 (13%) | 92% |
| | | Partners | n = 1,961 (58%) | 96% |
| | | Siblings | n = 383 (11%) | 50% |
| | | Pooled | n = 3,235 (96%) | 89% |
| | | No available closest relative | n = 118 (4%) | n/a |
| Older people (65+ years) | n = 2,092 | Children | n = 849 (41%) | 94% |
| | | Partners | n = 994 (48%) | 95% |
| | | Siblings | n = 41 (2%) | 36% |
| | | Pooled | n = 1,884 (90%) | 94% |
| | | No available closest relative | n = 208 (10%) | n/a |

**Supplementary Table 151**
**Infections of the CNS - Incident cohort 2016-2021**
**Patient characteristics - cases**

| ***Age strata of index patients*** | ***Characteristic*** | |
|---|---|---|
| Children and young people (0-24 years) | Number of persons, n | 1,268 (100.0) |
| | Male, n (%) | 663 (52.3) |
| | Age, median (Q1;Q3) | 15.2 (6.0;20.1) |
| | CCI, n (%) | |
| | 0 | 1,147 (90.5) |
| | 1-2 | 113 (8.9) |
| | +3 | 8 (0.6) |
| | Education level, n (%) | |
| | No/Unknown education | 559 (44.1) |
| | Primary | 506 (39.9) |
| | Secondary | 184 (14.5) |
| | Tertiary | 19 (1.5) |
| Young adults and adults (25-64 years) | Number of persons, n | 2,901 (100.0) |
| | Male, n (%) | 1,500 (51.7) |
| | Age, median (Q1;Q3) | 45.5 (33.7;55.6) |
| | CCI, n (%) | |
| | 0 | 2,133 (73.5) |
| | 1-2 | 577 (19.9) |
| | +3 | 191 (6.6) |
| | Education level, n (%) | |
| | No/Unknown education | 90 (3.1) |
| | Primary | 574 (19.8) |
| | Secondary | 1,323 (45.6) |
| | Tertiary | 914 (31.5) |
| Older people (65+ years) | Number of persons, n | 1,773 (100.0) |
| | Male, n (%) | 874 (49.3) |
| | Age, median (Q1;Q3) | 74.9 (70.1;80.6) |
| | CCI, n (%) | |
| | 0 | 702 (39.6) |
| | 1-2 | 661 (37.3) |
| | +3 | 410 (23.1) |
| | Education level, n (%) | |
| | No/Unknown education | 124 (7.0) |
| | Primary | 558 (31.5) |
| | Secondary | 791 (44.6) |
| | Tertiary | 300 (16.9) |

**Supplementary Table 152**
**Infections of the CNS - Incident cohort 2016-2021**
**Patient characteristics - relatives - pooled**

| *Age strata of index patients* | *Closest relative type* | *Characteristic of family relatives by agestrata of index patients* | *Closest family relatives of brain disorder patients* | *Comparison relative* |
|---|---|---|---|---|
| Children and young people (0-24 years) | Fathers | Number of persons, n | 1,115 (100.0) | 1,115 (100.0) |
| | | Male, n (%) | 1,115 (100.0) | 1,115 (100.0) |
| | | Age, median (Q1;Q3) | 46.9 (38.7;53.3) | 47.2 (39.4;53.1) |
| | | CCI, n (%) | | |
| | | 0 | 998 (89.5) | 1,004 (90.0) |
| | | 1-2 | 102 (9.1) | 98 (8.8) |
| | | +3 | 15 (1.3) | 13 (1.2) |
| | | Education level, n (%) | | |
| | | No/Unknown education | 25 (2.2) | 25 (2.2) |
| | | Primary | 207 (18.6) | 207 (18.6) |
| | | Secondary | 555 (49.8) | 555 (49.8) |
| | | Tertiary | 328 (29.4) | 328 (29.4) |
| | | Relation to patient, n (%) | | |
| | | Father | 1,115 (100.0) | 1,115 (100.0) |
| Children and young people (0-24 years) | Mothers | Number of persons, n | 1,205 (100.0) | 1,205 (100.0) |
| | | Male, n (%) | 0 (0.0) | 0 (0.0) |
| | | Age, median (Q1;Q3) | 44.3 (37.3;50.3) | 44.5 (36.9;50.1) |
| | | CCI, n (%) | | |
| | | 0 | 1,044 (86.6) | 1,073 (89.0) |
| | | 1-2 | 150 (12.4) | 122 (10.1) |
| | | +3 | 11 (0.9) | 10 (0.8) |
| | | Education level, n (%) | | |
| | | No/Unknown education | 29 (2.4) | 29 (2.4) |
| | | Primary | 205 (17.0) | 205 (17.0) |
| | | Secondary | 508 (42.2) | 508 (42.2) |
| | | Tertiary | 463 (38.4) | 463 (38.4) |
| | | Relation to patient, n (%) | | |
| | | Mother | 1,205 (100.0) | 1,205 (100.0) |
| Young adults and adults (25-64 years) | Pooled | Number of persons, n | 2,901 (100.0) | 2,901 (100.0) |
| | | Male, n (%) | 1,337 (46.1) | 1,337 (46.1) |
| | | Age, median (Q1;Q3) | 45.7 (31.7;57.5) | 46.0 (31.7;57.4) |
| | | CCI, n (%) | | |
| | | 0 | 2,463 (84.9) | 2,549 (87.9) |
| | | 1-2 | 376 (13.0) | 283 (9.8) |
| | | +3 | 62 (2.1) | 69 (2.4) |
| | | Education level, n (%) | | |
| | | No/Unknown education | 179 (6.2) | 179 (6.2) |
| | | Primary | 617 (21.3) | 617 (21.3) |
| | | Secondary | 1,270 (43.8) | 1,270 (43.8) |
| | | Tertiary | 835 (28.8) | 835 (28.8) |
| | | Relation to patient, n (%) | | |
| | | Mother | 317 (10.9) | 317 (10.9) |
| | | Father | 87 (3.0) | 87 (3.0) |
| | | Partner | 1,887 (65.0) | 1,887 (65.0) |
| | | Child | 417 (14.4) | 417 (14.4) |

#### Supplementary Table 152
#### Infections of the CNS - Incident cohort 2016-2021
#### Patient characteristics - relatives - pooled

| *Age strata of index patients* | *Closest relative type* | *Characteristic of family relatives by agestrata of index patients* | *Closest family relatives of brain disorder patients* | *Comparison relative* |
|---|---|---|---|---|
| | | Sibling | 193 (6.7) | 193 (6.7) |
| Older people (65+ years) | Pooled | Number of persons, n | 1,773 (100.0) | 1,773 (100.0) |
| | | Male, n (%) | 819 (46.2) | 819 (46.2) |
| | | Age, median (Q1;Q3) | 64.5 (53.1;73.3) | 64.4 (52.8;73.2) |
| | | CCI, n (%) | | |
| | | 0 | 1,280 (72.2) | 1,262 (71.2) |
| | | 1-2 | 374 (21.1) | 399 (22.5) |
| | | +3 | 119 (6.7) | 112 (6.3) |
| | | Education level, n (%) | | |
| | | No/Unknown education | 57 (3.2) | 57 (3.2) |
| | | Primary | 366 (20.6) | 366 (20.6) |
| | | Secondary | 854 (48.2) | 854 (48.2) |
| | | Tertiary | 496 (28.0) | 496 (28.0) |
| | | Relation to patient, n (%) | | |
| | | Partner | 954 (53.8) | 954 (53.8) |
| | | Child | 804 (45.3) | 804 (45.3) |
| | | Sibling | 15 (0.8) | 15 (0.8) |

#### Supplementary Table 153
#### Infections of the CNS - Incident cohort 2016-2021
#### Patient characteristics - relatives - not pooled

| *Age strata of index patients* | *Closest relative type* | *Characteristic of family relatives by agestrata of index patients* | *Closest family relatives of brain disorder patients* | *Comparison relative* |
|---|---|---|---|---|
| Children and young people (0-24 years) | Fathers | Number of persons, n | 1,115 (100.0) | 1,115 (100.0) |
| | | Male, n (%) | 1,115 (100.0) | 1,115 (100.0) |
| | | Age, median (Q1;Q3) | 46.9 (38.7;53.3) | 47.2 (39.4;53.1) |
| | | CCI, n (%) | | |
| | | 0 | 998 (89.5) | 1,004 (90.0) |
| | | 1-2 | 102 (9.1) | 98 (8.8) |
| | | +3 | 15 (1.3) | 13 (1.2) |
| | | Education level, n (%) | | |
| | | No/Unknown education | 25 (2.2) | 25 (2.2) |
| | | Primary | 207 (18.6) | 207 (18.6) |
| | | Secondary | 555 (49.8) | 555 (49.8) |
| | | Tertiary | 328 (29.4) | 328 (29.4) |
| | | Relation to patient, n (%) | | |
| | | Father | 1,115 (100.0) | 1,115 (100.0) |
| Children and young people (0-24 years) | Mothers | Number of persons, n | 1,205 (100.0) | 1,205 (100.0) |
| | | Male, n (%) | 0 (0.0) | 0 (0.0) |
| | | Age, median (Q1;Q3) | 44.3 (37.3;50.3) | 44.5 (36.9;50.1) |
| | | CCI, n (%) | | |
| | | 0 | 1,044 (86.6) | 1,073 (89.0) |
| | | 1-2 | 150 (12.4) | 122 (10.1) |
| | | +3 | 11 (0.9) | 10 (0.8) |
| | | Education level, n (%) | | |
| | | No/Unknown education | 29 (2.4) | 29 (2.4) |
| | | Primary | 205 (17.0) | 205 (17.0) |
| | | Secondary | 508 (42.2) | 508 (42.2) |
| | | Tertiary | 463 (38.4) | 463 (38.4) |
| | | Relation to patient, n (%) | | |
| | | Mother | 1,205 (100.0) | 1,205 (100.0) |
| Young adults and adults (25-64 years) | Children | Number of persons, n | 417 (100.0) | 417 (100.0) |
| | | Male, n (%) | 217 (52.0) | 217 (52.0) |
| | | Age, median (Q1;Q3) | 19.7 (11.5;29.0) | 19.7 (11.4;28.3) |
| | | CCI, n (%) | | |
| | | 0 | 392 (94.0) | 397 (95.2) |
| | | 1-2 | 25 (6.0) | 20 (4.8) |
| | | +3 | 0 (0.0) | 0 (0.0) |
| | | Education level, n (%) | | |
| | | No/Unknown education | 121 (29.0) | 121 (29.0) |
| | | Primary | 149 (35.7) | 149 (35.7) |
| | | Secondary | 103 (24.7) | 103 (24.7) |
| | | Tertiary | 44 (10.6) | 44 (10.6) |
| | | Relation to patient, n (%) | | |
| | | Child | 417 (100.0) | 417 (100.0) |
| Young adults and adults (25-64 years) | Parents | Number of persons, n | 404 (100.0) | 404 (100.0) |
| | | Male, n (%) | 87 (21.5) | 87 (21.5) |
| | | Age, median (Q1;Q3) | 63.8 (56.3;72.7) | 63.6 (56.9;72.6) |

### Supplementary Table 153
### Infections of the CNS - Incident cohort 2016-2021
### Patient characteristics - relatives - not pooled

| *Age strata of index patients* | *Closest relative type* | *Characteristic of family relatives by agestrata of index patients* | *Closest family relatives of brain disorder patients* | *Comparison relative* |
|---|---|---|---|---|
| | | CCI, n (%) | | |
| | | 0 | 266 (65.8) | 297 (73.5) |
| | | 1-2 | 103 (25.5) | 74 (18.3) |
| | | +3 | 35 (8.7) | 33 (8.2) |
| | | Education level, n (%) | | |
| | | No/Unknown education | * | * |
| | | Primary | 125 (30.9) | 125 (30.9) |
| | | Secondary | 196 (48.5) | 196 (48.5) |
| | | Tertiary | * | * |
| | | Relation to patient, n (%) | | |
| | | Mother | 317 (78.5) | 317 (78.5) |
| | | Father | 87 (21.5) | 87 (21.5) |
| Young adults and adults (25-64 years) | Partners | Number of persons, n | 1,887 (100.0) | 1,887 (100.0) |
| | | Male, n (%) | 918 (48.6) | 918 (48.6) |
| | | Age, median (Q1;Q3) | 45.1 (33.8;55.6) | 45.5 (33.9;55.4) |
| | | CCI, n (%) | | |
| | | 0 | 1,654 (87.7) | 1,687 (89.4) |
| | | 1-2 | 208 (11.0) | 169 (9.0) |
| | | +3 | 25 (1.3) | 31 (1.6) |
| | | Education level, n (%) | | |
| | | No/Unknown education | 54 (2.9) | 54 (2.9) |
| | | Primary | 300 (15.9) | 300 (15.9) |
| | | Secondary | 865 (45.8) | 865 (45.8) |
| | | Tertiary | 668 (35.4) | 668 (35.4) |
| | | Relation to patient, n (%) | | |
| | | Partner | 1,887 (100.0) | 1,887 (100.0) |
| Young adults and adults (25-64 years) | Siblings | Number of persons, n | 193 (100.0) | 193 (100.0) |
| | | Male, n (%) | 115 (59.6) | 115 (59.6) |
| | | Age, median (Q1;Q3) | 54.2 (40.4;59.8) | 54.1 (42.6;59.2) |
| | | CCI, n (%) | | |
| | | 0 | 151 (78.2) | 168 (87.0) |
| | | 1-2 | * | 20 (10.4) |
| | | +3 | * | 5 (2.6) |
| | | Education level, n (%) | | |
| | | No/Unknown education | 0 (0.0) | 0 (0.0) |
| | | Primary | 43 (22.3) | 43 (22.3) |
| | | Secondary | 106 (54.9) | 106 (54.9) |
| | | Tertiary | 44 (22.8) | 44 (22.8) |
| | | Relation to patient, n (%) | | |
| | | Sibling | 193 (100.0) | 193 (100.0) |
| Older people (65+ years) | Children | Number of persons, n | 804 (100.0) | 804 (100.0) |
| | | Male, n (%) | 402 (50.0) | 402 (50.0) |
| | | Age, median (Q1;Q3) | 51.9 (45.8;57.4) | 52.0 (46.0;57.3) |
| | | CCI, n (%) | | |
| | | 0 | 684 (85.1) | 678 (84.3) |

### Supplementary Table 153
### Infections of the CNS - Incident cohort 2016-2021
### Patient characteristics - relatives - not pooled

| *Age strata of index patients* | *Closest relative type* | *Characteristic of family relatives by agestrata of index patients* | *Closest family relatives of brain disorder patients* | *Comparison relative* |
|---|---|---|---|---|
| | | 1-2 | 101 (12.6) | 105 (13.1) |
| | | +3 | 19 (2.4) | 21 (2.6) |
| | | Education level, n (%) | | |
| | | No/Unknown education | 8 (1.0) | 8 (1.0) |
| | | Primary | 115 (14.3) | 115 (14.3) |
| | | Secondary | 398 (49.5) | 398 (49.5) |
| | | Tertiary | 283 (35.2) | 283 (35.2) |
| | | Relation to patient, n (%) | | |
| | | Child | 804 (100.0) | 804 (100.0) |
| Older people (65+ years) | Partners | Number of persons, n | 954 (100.0) | 954 (100.0) |
| | | Male, n (%) | 409 (42.9) | 409 (42.9) |
| | | Age, median (Q1;Q3) | 72.7 (68.0;77.5) | 72.5 (67.9;77.3) |
| | | CCI, n (%) | | |
| | | 0 | 583 (61.1) | 573 (60.1) |
| | | 1-2 | 272 (28.5) | 290 (30.4) |
| | | +3 | 99 (10.4) | 91 (9.5) |
| | | Education level, n (%) | | |
| | | No/Unknown education | 49 (5.1) | 49 (5.1) |
| | | Primary | 249 (26.1) | 249 (26.1) |
| | | Secondary | 445 (46.6) | 445 (46.6) |
| | | Tertiary | 211 (22.1) | 211 (22.1) |
| | | Relation to patient, n (%) | | |
| | | Partner | 954 (100.0) | 954 (100.0) |
| Older people (65+ years) | Siblings | Number of persons, n | 15 (100.0) | 15 (100.0) |
| | | Male, n (%) | 8 (53.3) | 8 (53.3) |
| | | Age, median (Q1;Q3) | 60.5 (59.2;62.7) | 60.8 (58.0;61.9) |
| | | CCI, n (%) | | |
| | | 0 | * | * |
| | | 1-2 | * | * |
| | | +3 | * | 0 (0.0) |
| | | Education level, n (%) | | |
| | | No/Unknown education | 0 (0.0) | 0 (0.0) |
| | | Primary | * | * |
| | | Secondary | * | * |
| | | Tertiary | * | * |
| | | Relation to patient, n (%) | | |
| | | Sibling | 15 (100.0) | 15 (100.0) |

#### Supplementary Table 154
#### Infections of the CNS - Incident cohort 2016-2021
#### Cost analysis - pooled

| | | | | Actual costs | | Attributable costs | |
|---|---|---|---|---|---|---|---|
| *Age strata of index patients* | *Closest relative type* | *Number of index patients included in cost component calculations* | *Cost component* | *Total (million EUR)* | *Per person (EUR)* | *Total (million EUR)* | *Per person (EUR)* |
| Children and young people (0-24 years) | Fathers | 1,115 | Primary sector | 0.3 | 232 | 0.0 | 30 |
| | | 1,115 | Hospital care | 1.9 | 1,668 | 0.6 | 509 |
| | | 1,115 | Filled prescriptions | 0.2 | 205 | 0.1 | 54 |
| | | 1,085 | Lost productivity (income loss) | . | . | 1.3 | 1,174 |
| Children and young people (0-24 years) | Mothers | 1,205 | Primary sector | 0.5 | 375 | 0.1 | 48 |
| | | 1,205 | Hospital care | 2.6 | 2,173 | 1.0 | 862 |
| | | 1,205 | Filled prescriptions | 0.2 | 203 | 0.0 | 31 |
| | | 1,199 | Lost productivity (income loss) | . | . | -3.1 | -2,583 |
| Young adults and adults (25-64 years) | Pooled | 2,901 | Primary sector | 0.9 | 316 | 0.1 | 25 |
| | | 2,901 | Hospital care | 6.6 | 2,286 | 2.1 | 728 |
| | | 2,901 | Filled prescriptions | 0.7 | 232 | 0.0 | 14 |
| | | 2,284 | Lost productivity (income loss) | . | . | 1.0 | 453 |
| Older people (65+ years) | Pooled | 1,773 | Primary sector | 0.7 | 419 | 0.0 | 27 |
| | | 1,773 | Hospital care | 5.6 | 3,162 | 0.2 | 130 |
| | | 1,773 | Filled prescriptions | 0.7 | 396 | 0.0 | 11 |
| | | 811 | Lost productivity (income loss) | . | . | -4.0 | -4,979 |

#### Supplementary Table 155
#### Infections of the CNS - Incident cohort 2016-2021
#### Cost analysis - not pooled

| Age strata of index patients | Closest relative type | Number of index patients included in cost component calculations | Cost component | Actual costs: Total (million EUR) | Actual costs: Per person (EUR) | Attributable costs: Total (million EUR) | Attributable costs: Per person (EUR) |
|---|---|---|---|---|---|---|---|
| Children and young people (0-24 years) | Fathers | 1,115 | Primary sector | 0.3 | 232 | 0.0 | 30 |
| | | 1,115 | Hospital care | 1.9 | 1,668 | 0.6 | 509 |
| | | 1,115 | Filled prescriptions | 0.2 | 205 | 0.1 | 54 |
| | | 1,085 | Lost productivity (income loss) | . | . | 1.3 | 1,174 |
| Children and young people (0-24 years) | Mothers | 1,205 | Primary sector | 0.5 | 375 | 0.1 | 48 |
| | | 1,205 | Hospital care | 2.6 | 2,173 | 1.0 | 862 |
| | | 1,205 | Filled prescriptions | 0.2 | 203 | 0.0 | 31 |
| | | 1,199 | Lost productivity (income loss) | . | . | -3.1 | -2,583 |
| Young adults and adults (25-64 years) | Children | 417 | Primary sector | 0.1 | 230 | 0.0 | 11 |
| | | 417 | Hospital care | 0.3 | 774 | -0.1 | -282 |
| | | 417 | Filled prescriptions | 0.0 | 102 | 0.0 | 20 |
| | | 226 | Lost productivity (income loss) | . | . | -0.7 | -3,163 |
| Young adults and adults (25-64 years) | Parents | 404 | Primary sector | 0.2 | 452 | 0.0 | 23 |
| | | 404 | Hospital care | 1.0 | 2,511 | -0.0 | -9 |
| | | 404 | Filled prescriptions | 0.2 | 439 | -0.0 | -75 |
| | | 183 | Lost productivity (income loss) | . | . | 0.9 | 4,945 |
| Young adults and adults (25-64 years) | Partners | 1,887 | Primary sector | 0.6 | 308 | 0.1 | 30 |
| | | 1,887 | Hospital care | 4.3 | 2,258 | 1.5 | 818 |
| | | 1,887 | Filled prescriptions | 0.4 | 211 | 0.0 | 24 |
| | | 1,713 | Lost productivity (income loss) | . | . | 1.4 | 846 |
| Young adults and adults (25-64 years) | Siblings | 193 | Primary sector | 0.1 | 299 | 0.0 | 2 |
| | | 193 | Hospital care | 1.0 | 5,348 | 0.7 | 3,554 |
| | | 193 | Filled prescriptions | 0.1 | 292 | 0.0 | 86 |
| | | 162 | Lost productivity (income loss) | . | . | -0.6 | -3,734 |
| Older people (65+ years) | Children | 804 | Primary sector | 0.2 | 302 | 0.0 | 24 |
| | | 804 | Hospital care | 1.6 | 1,989 | -0.1 | -77 |
| | | 804 | Filled prescriptions | 0.2 | 268 | 0.0 | 52 |
| | | 747 | Lost productivity (income loss) | . | . | -3.1 | -4,179 |
| Older people (65+ years) | Partners | 954 | Primary sector | 0.5 | 524 | 0.0 | 33 |
| | | 954 | Hospital care | 4.0 | 4,195 | 0.3 | 313 |
| | | 954 | Filled prescriptions | 0.5 | 508 | -0.0 | -25 |
| | | 53 | Lost productivity (income loss) | . | . | -0.9 | -16,346 |
| Older people (65+ years) | Siblings | 15 | Primary sector | 0.0 | 158 | -0.0 | -136 |
| | | 15 | Hospital care | 0.0 | 1,151 | -0.0 | -233 |
| | | 15 | Filled prescriptions | 0.0 | 282 | 0.0 | 40 |
| | | 11 | Lost productivity (income loss) | . | . | -0.1 | -4,568 |

### Supplementary Table 156
#### Infections of the CNS - Prevalent cohort 2021
#### Identification of populations of relatives

| *Agegroup of index patients in matched population* | | *Closest relative type identified* | | *Available control with same type of closest relative* |
|---|---|---|---|---|
| Children and young people (0-24 years) | n = 4,871 | Fathers | n = 4,194 (86%) | 97% |
| | | Mothers | n = 4,420 (91%) | 97% |
| | | No available closest relative | n = 172 (4%) | n/a |
| Young adults and adults (25-64 years) | n = 9,295 | Children | n = 1,201 (13%) | 94% |
| | | Parents | n = 1,408 (15%) | 92% |
| | | Partners | n = 5,604 (60%) | 96% |
| | | Siblings | n = 934 (10%) | 43% |
| | | Pooled | n = 9,147 (98%) | 90% |
| | | No available closest relative | n = 148 (2%) | n/a |
| Older people (65+ years) | n = 3,825 | Children | n = 1,257 (33%) | 94% |
| | | Partners | n = 2,155 (56%) | 96% |
| | | Siblings | n = 90 (2%) | 53% |
| | | Pooled | n = 3,502 (92%) | 94% |
| | | No available closest relative | n = 323 (8%) | n/a |

#### Supplementary Table 157
#### Infections of the CNS - Prevalent cohort 2021
#### Patient characteristics - cases

| *Age strata of index patients* | *Characteristic* | |
|---|---|---|
| Children and young people (0-24 years) | Number of persons, n | 4,573 (100.0) |
| | Male, n (%) | 2,635 (57.6) |
| | Age, median (Q1;Q3) | 15.6 (10.0;20.1) |
| | CCI, n (%) | |
| | 0 | 4,059 (88.8) |
| | 1-2 | 456 (10.0) |
| | +3 | 58 (1.3) |
| | Education level, n (%) | |
| | No/Unknown education | 1,735 (37.9) |
| | Primary | 2,165 (47.3) |
| | Secondary | 619 (13.5) |
| | Tertiary | 54 (1.2) |
| Young adults and adults (25-64 years) | Number of persons, n | 8,252 (100.0) |
| | Male, n (%) | 4,149 (50.3) |
| | Age, median (Q1;Q3) | 45.0 (34.7;54.5) |
| | CCI, n (%) | |
| | 0 | 6,307 (76.4) |
| | 1-2 | 1,481 (17.9) |
| | +3 | 464 (5.6) |
| | Education level, n (%) | |
| | No/Unknown education | 166 (2.0) |
| | Primary | 1,632 (19.8) |
| | Secondary | 3,672 (44.5) |
| | Tertiary | 2,782 (33.7) |
| Older people (65+ years) | Number of persons, n | 3,314 (100.0) |
| | Male, n (%) | 1,600 (48.3) |
| | Age, median (Q1;Q3) | 74.5 (69.9;79.9) |
| | CCI, n (%) | |
| | 0 | 1,265 (38.2) |
| | 1-2 | 1,299 (39.2) |
| | +3 | 750 (22.6) |
| | Education level, n (%) | |
| | No/Unknown education | 216 (6.5) |
| | Primary | 905 (27.3) |
| | Secondary | 1,525 (46.0) |
| | Tertiary | 668 (20.2) |

**Supplementary Table 158**
**Infections of the CNS - Prevalent cohort 2021**
**Patient characteristics - relatives - pooled**

| *Age strata of index patients* | *Closest relative type* | *Characteristic of family relatives by agestrata of index patients* | *Closest family relatives of brain disorder patients* | *Comparison relative* |
|---|---|---|---|---|
| Children and young people (0-24 years) | Fathers | Number of persons, n | 4,080 (100.0) | 4,080 (100.0) |
| | | Male, n (%) | 4,080 (100.0) | 4,080 (100.0) |
| | | Age, median (Q1;Q3) | 47.9 (42.0;53.3) | 47.9 (42.1;53.1) |
| | | CCI, n (%) | | |
| | | 0 | 3,597 (88.2) | 3,655 (89.6) |
| | | 1-2 | 426 (10.4) | 380 (9.3) |
| | | +3 | 57 (1.4) | 45 (1.1) |
| | | Education level, n (%) | | |
| | | No/Unknown education | 45 (1.1) | 45 (1.1) |
| | | Primary | 762 (18.7) | 762 (18.7) |
| | | Secondary | 2,042 (50.0) | 2,042 (50.0) |
| | | Tertiary | 1,231 (30.2) | 1,231 (30.2) |
| | | Relation to patient, n (%) | | |
| | | Father | 4,080 (100.0) | 4,080 (100.0) |
| Children and young people (0-24 years) | Mothers | Number of persons, n | 4,299 (100.0) | 4,299 (100.0) |
| | | Male, n (%) | 0 (0.0) | 0 (0.0) |
| | | Age, median (Q1;Q3) | 45.9 (39.8;50.8) | 45.9 (40.2;50.7) |
| | | CCI, n (%) | | |
| | | 0 | 3,704 (86.2) | 3,800 (88.4) |
| | | 1-2 | 535 (12.4) | 469 (10.9) |
| | | +3 | 60 (1.4) | 30 (0.7) |
| | | Education level, n (%) | | |
| | | No/Unknown education | 48 (1.1) | 48 (1.1) |
| | | Primary | 699 (16.3) | 699 (16.3) |
| | | Secondary | 1,737 (40.4) | 1,737 (40.4) |
| | | Tertiary | 1,815 (42.2) | 1,815 (42.2) |
| | | Relation to patient, n (%) | | |
| | | Mother | 4,299 (100.0) | 4,299 (100.0) |
| Young adults and adults (25-64 years) | Pooled | Number of persons, n | 8,252 (100.0) | 8,252 (100.0) |
| | | Male, n (%) | 3,811 (46.2) | 3,811 (46.2) |
| | | Age, median (Q1;Q3) | 46.4 (32.7;57.7) | 46.6 (32.9;57.6) |
| | | CCI, n (%) | | |
| | | 0 | 7,001 (84.8) | 7,094 (86.0) |
| | | 1-2 | 1,074 (13.0) | 989 (12.0) |
| | | +3 | 177 (2.1) | 169 (2.0) |
| | | Education level, n (%) | | |
| | | No/Unknown education | 478 (5.8) | 478 (5.8) |
| | | Primary | 1,755 (21.3) | 1,755 (21.3) |
| | | Secondary | 3,515 (42.6) | 3,515 (42.6) |
| | | Tertiary | 2,504 (30.3) | 2,504 (30.3) |
| | | Relation to patient, n (%) | | |
| | | Mother | 1,017 (12.3) | 1,017 (12.3) |
| | | Father | 284 (3.4) | 284 (3.4) |
| | | Partner | 5,403 (65.5) | 5,403 (65.5) |
| | | Child | 1,138 (13.8) | 1,138 (13.8) |

#### Supplementary Table 158
#### Infections of the CNS - Prevalent cohort 2021
#### Patient characteristics - relatives - pooled

| *Age strata of index patients* | *Closest relative type* | *Characteristic of family relatives by agestrata of index patients* | *Closest family relatives of brain disorder patients* | *Comparison relative* |
|---|---|---|---|---|
| | | Sibling | 410 (5.0) | 410 (5.0) |
| Older people (65+ years) | Pooled | Number of persons, n | 3,314 (100.0) | 3,314 (100.0) |
| | | Male, n (%) | 1,483 (44.7) | 1,483 (44.7) |
| | | Age, median (Q1;Q3) | 67.2 (55.9;74.6) | 67.3 (56.1;74.5) |
| | | CCI, n (%) | | |
| | | 0 | 2,317 (69.9) | 2,310 (69.7) |
| | | 1-2 | 755 (22.8) | 772 (23.3) |
| | | +3 | 242 (7.3) | 232 (7.0) |
| | | Education level, n (%) | | |
| | | No/Unknown education | 103 (3.1) | 103 (3.1) |
| | | Primary | 675 (20.4) | 675 (20.4) |
| | | Secondary | 1,610 (48.6) | 1,610 (48.6) |
| | | Tertiary | 926 (27.9) | 926 (27.9) |
| | | Relation to patient, n (%) | | |
| | | Partner | 2,076 (62.6) | 2,076 (62.6) |
| | | Child | 1,190 (35.9) | 1,190 (35.9) |
| | | Sibling | 48 (1.4) | 48 (1.4) |

**Supplementary Table 159**
**Infections of the CNS - Prevalent cohort 2021**
**Patient characteristics - relatives - not pooled**

| *Age strata of index patients* | *Closest relative type* | *Characteristic of family relatives by agestrata of index patients* | *Closest family relatives of brain disorder patients* | *Comparison relative* |
|---|---|---|---|---|
| Children and young people (0-24 years) | Fathers | Number of persons, n | 4,080 (100.0) | 4,080 (100.0) |
| | | Male, n (%) | 4,080 (100.0) | 4,080 (100.0) |
| | | Age, median (Q1;Q3) | 47.9 (42.0;53.3) | 47.9 (42.1;53.1) |
| | | CCI, n (%) | | |
| | | 0 | 3,597 (88.2) | 3,655 (89.6) |
| | | 1-2 | 426 (10.4) | 380 (9.3) |
| | | +3 | 57 (1.4) | 45 (1.1) |
| | | Education level, n (%) | | |
| | | No/Unknown education | 45 (1.1) | 45 (1.1) |
| | | Primary | 762 (18.7) | 762 (18.7) |
| | | Secondary | 2,042 (50.0) | 2,042 (50.0) |
| | | Tertiary | 1,231 (30.2) | 1,231 (30.2) |
| | | Relation to patient, n (%) | | |
| | | Father | 4,080 (100.0) | 4,080 (100.0) |
| Children and young people (0-24 years) | Mothers | Number of persons, n | 4,299 (100.0) | 4,299 (100.0) |
| | | Male, n (%) | 0 (0.0) | 0 (0.0) |
| | | Age, median (Q1;Q3) | 45.9 (39.8;50.8) | 45.9 (40.2;50.7) |
| | | CCI, n (%) | | |
| | | 0 | 3,704 (86.2) | 3,800 (88.4) |
| | | 1-2 | 535 (12.4) | 469 (10.9) |
| | | +3 | 60 (1.4) | 30 (0.7) |
| | | Education level, n (%) | | |
| | | No/Unknown education | 48 (1.1) | 48 (1.1) |
| | | Primary | 699 (16.3) | 699 (16.3) |
| | | Secondary | 1,737 (40.4) | 1,737 (40.4) |
| | | Tertiary | 1,815 (42.2) | 1,815 (42.2) |
| | | Relation to patient, n (%) | | |
| | | Mother | 4,299 (100.0) | 4,299 (100.0) |
| Young adults and adults (25-64 years) | Children | Number of persons, n | 1,138 (100.0) | 1,138 (100.0) |
| | | Male, n (%) | 584 (51.3) | 584 (51.3) |
| | | Age, median (Q1;Q3) | 18.5 (12.0;26.9) | 18.5 (11.4;26.5) |
| | | CCI, n (%) | | |
| | | 0 | 1,054 (92.6) | 1,075 (94.5) |
| | | 1-2 | * | * |
| | | +3 | * | * |
| | | Education level, n (%) | | |
| | | No/Unknown education | 339 (29.8) | 339 (29.8) |
| | | Primary | 464 (40.8) | 464 (40.8) |
| | | Secondary | 223 (19.6) | 223 (19.6) |
| | | Tertiary | 112 (9.8) | 112 (9.8) |
| | | Relation to patient, n (%) | | |
| | | Child | 1,138 (100.0) | 1,138 (100.0) |
| Young adults and adults (25-64 years) | Parents | Number of persons, n | 1,301 (100.0) | 1,301 (100.0) |
| | | Male, n (%) | 284 (21.8) | 284 (21.8) |
| | | Age, median (Q1;Q3) | 64.5 (57.8;73.6) | 64.5 (57.8;73.7) |

#### Supplementary Table 159
#### Infections of the CNS - Prevalent cohort 2021
#### Patient characteristics - relatives - not pooled

| *Age strata of index patients* | *Closest relative type* | *Characteristic of family relatives by agestrata of index patients* | *Closest family relatives of brain disorder patients* | *Comparison relative* |
|---|---|---|---|---|
| | | CCI, n (%) | | |
| | | 0 | 879 (67.6) | 914 (70.3) |
| | | 1-2 | 341 (26.2) | 309 (23.8) |
| | | +3 | 81 (6.2) | 78 (6.0) |
| | | Education level, n (%) | | |
| | | No/Unknown education | 20 (1.5) | 20 (1.5) |
| | | Primary | 362 (27.8) | 362 (27.8) |
| | | Secondary | 632 (48.6) | 632 (48.6) |
| | | Tertiary | 287 (22.1) | 287 (22.1) |
| | | Relation to patient, n (%) | | |
| | | Mother | 1,017 (78.2) | 1,017 (78.2) |
| | | Father | 284 (21.8) | 284 (21.8) |
| Young adults and adults (25-64 years) | Partners | Number of persons, n | 5,403 (100.0) | 5,403 (100.0) |
| | | Male, n (%) | 2,693 (49.8) | 2,693 (49.8) |
| | | Age, median (Q1;Q3) | 45.5 (35.6;54.9) | 45.6 (35.6;54.7) |
| | | CCI, n (%) | | |
| | | 0 | 4,723 (87.4) | 4,752 (88.0) |
| | | 1-2 | 596 (11.0) | 575 (10.6) |
| | | +3 | 84 (1.6) | 76 (1.4) |
| | | Education level, n (%) | | |
| | | No/Unknown education | 118 (2.2) | 118 (2.2) |
| | | Primary | 816 (15.1) | 816 (15.1) |
| | | Secondary | 2,442 (45.2) | 2,442 (45.2) |
| | | Tertiary | 2,027 (37.5) | 2,027 (37.5) |
| | | Relation to patient, n (%) | | |
| | | Partner | 5,403 (100.0) | 5,403 (100.0) |
| Young adults and adults (25-64 years) | Siblings | Number of persons, n | 410 (100.0) | 410 (100.0) |
| | | Male, n (%) | 250 (61.0) | 250 (61.0) |
| | | Age, median (Q1;Q3) | 54.3 (36.5;59.6) | 54.6 (38.3;59.7) |
| | | CCI, n (%) | | |
| | | 0 | 345 (84.1) | 353 (86.1) |
| | | 1-2 | 57 (13.9) | 43 (10.5) |
| | | +3 | 8 (2.0) | 14 (3.4) |
| | | Education level, n (%) | | |
| | | No/Unknown education | * | * |
| | | Primary | 113 (27.6) | 113 (27.6) |
| | | Secondary | 218 (53.2) | 218 (53.2) |
| | | Tertiary | * | * |
| | | Relation to patient, n (%) | | |
| | | Sibling | 410 (100.0) | 410 (100.0) |
| Older people (65+ years) | Children | Number of persons, n | 1,190 (100.0) | 1,190 (100.0) |
| | | Male, n (%) | 579 (48.7) | 579 (48.7) |
| | | Age, median (Q1;Q3) | 52.3 (45.8;58.1) | 52.1 (45.9;57.5) |
| | | CCI, n (%) | | |
| | | 0 | 1,002 (84.2) | 997 (83.8) |

### Supplementary Table 159
#### Infections of the CNS - Prevalent cohort 2021
#### Patient characteristics - relatives - not pooled

| *Age strata of index patients* | *Closest relative type* | *Characteristic of family relatives by agestrata of index patients* | *Closest family relatives of brain disorder patients* | *Comparison relative* |
|---|---|---|---|---|
| | | 1-2 | 162 (13.6) | 159 (13.4) |
| | | +3 | 26 (2.2) | 34 (2.9) |
| | | Education level, n (%) | | |
| | | No/Unknown education | * | * |
| | | Primary | * | * |
| | | Secondary | 576 (48.4) | 576 (48.4) |
| | | Tertiary | 421 (35.4) | 421 (35.4) |
| | | Relation to patient, n (%) | | |
| | | Child | 1,190 (100.0) | 1,190 (100.0) |
| Older people (65+ years) | Partners | Number of persons, n | 2,076 (100.0) | 2,076 (100.0) |
| | | Male, n (%) | 878 (42.3) | 878 (42.3) |
| | | Age, median (Q1;Q3) | 73.0 (67.8;77.2) | 72.9 (68.3;77.1) |
| | | CCI, n (%) | | |
| | | 0 | 1,280 (61.7) | 1,276 (61.5) |
| | | 1-2 | 582 (28.0) | 604 (29.1) |
| | | +3 | 214 (10.3) | 196 (9.4) |
| | | Education level, n (%) | | |
| | | No/Unknown education | 99 (4.8) | 99 (4.8) |
| | | Primary | 483 (23.3) | 483 (23.3) |
| | | Secondary | 1,003 (48.3) | 1,003 (48.3) |
| | | Tertiary | 491 (23.7) | 491 (23.7) |
| | | Relation to patient, n (%) | | |
| | | Partner | 2,076 (100.0) | 2,076 (100.0) |
| Older people (65+ years) | Siblings | Number of persons, n | 48 (100.0) | 48 (100.0) |
| | | Male, n (%) | 26 (54.2) | 26 (54.2) |
| | | Age, median (Q1;Q3) | 62.1 (59.1;64.8) | 61.5 (60.1;64.1) |
| | | CCI, n (%) | | |
| | | 0 | 35 (72.9) | 37 (77.1) |
| | | 1-2 | * | * |
| | | +3 | * | * |
| | | Education level, n (%) | | |
| | | No/Unknown education | 0 (0.0) | 0 (0.0) |
| | | Primary | * | * |
| | | Secondary | 31 (64.6) | 31 (64.6) |
| | | Tertiary | * | * |
| | | Relation to patient, n (%) | | |
| | | Sibling | 48 (100.0) | 48 (100.0) |

### Supplementary Table 160
### Infections of the CNS - Prevalent cohort 2021
### Cost analysis - pooled

| *Age strata of index patients* | *Closest relative type* | *Number of index patients included in cost component calculations* | *Cost component* | *Actual costs* | | *Attributable costs* | |
|---|---|---|---|---|---|---|---|
| | | | | *Total (million EUR)* | *Per person (EUR)* | *Total (million EUR)* | *Per person (EUR)* |
| Children and young people (0-24 years) | Fathers | 4,080 | Primary sector | 0.9 | 225 | 0.1 | 14 |
| | | 4,080 | Hospital care | 6.1 | 1,508 | 0.8 | 208 |
| | | 4,080 | Filled prescriptions | 0.7 | 178 | 0.1 | 22 |
| | | 3,983 | Lost productivity (income loss) | . | . | 2.5 | 624 |
| Children and young people (0-24 years) | Mothers | 4,299 | Primary sector | 1.6 | 368 | 0.1 | 32 |
| | | 4,299 | Hospital care | 7.9 | 1,851 | 2.0 | 459 |
| | | 4,299 | Filled prescriptions | 0.9 | 211 | 0.1 | 29 |
| | | 4,276 | Lost productivity (income loss) | . | . | 0.1 | 30 |
| Young adults and adults (25-64 years) | Pooled | 8,252 | Primary sector | 2.5 | 304 | 0.1 | 9 |
| | | 8,252 | Hospital care | 17.8 | 2,157 | 4.1 | 493 |
| | | 8,252 | Filled prescriptions | 1.9 | 233 | 0.2 | 25 |
| | | 6,453 | Lost productivity (income loss) | . | . | -0.5 | -76 |
| Older people (65+ years) | Pooled | 3,314 | Primary sector | 1.5 | 449 | 0.1 | 25 |
| | | 3,314 | Hospital care | 10.5 | 3,191 | -0.2 | -75 |
| | | 3,314 | Filled prescriptions | 1.4 | 427 | 0.1 | 40 |
| | | 1,241 | Lost productivity (income loss) | . | . | 2.3 | 1,843 |

#### Supplementary Table 161
#### Infections of the CNS - Prevalent cohort 2021
#### Cost analysis - not pooled

| | | | | *Actual costs* | | *Attributable costs* | |
|---|---|---|---|---|---|---|---|
| *Age strata of index patients* | *Closest relative type* | *Number of index patients included in cost component calculations* | *Cost component* | *Total (million EUR)* | *Per person (EUR)* | *Total (million EUR)* | *Per person (EUR)* |
| Children and young people (0-24 years) | Fathers | 4,080 | Primary sector | 0.9 | 225 | 0.1 | 14 |
| | | 4,080 | Hospital care | 6.1 | 1,508 | 0.8 | 208 |
| | | 4,080 | Filled prescriptions | 0.7 | 178 | 0.1 | 22 |
| | | 3,983 | Lost productivity (income loss) | . | . | 2.5 | 624 |
| Children and young people (0-24 years) | Mothers | 4,299 | Primary sector | 1.6 | 368 | 0.1 | 32 |
| | | 4,299 | Hospital care | 7.9 | 1,851 | 2.0 | 459 |
| | | 4,299 | Filled prescriptions | 0.9 | 211 | 0.1 | 29 |
| | | 4,276 | Lost productivity (income loss) | . | . | 0.1 | 30 |
| Young adults and adults (25-64 years) | Children | 1,138 | Primary sector | 0.2 | 204 | 0.0 | 12 |
| | | 1,138 | Hospital care | 1.2 | 1,040 | 0.1 | 80 |
| | | 1,138 | Filled prescriptions | 0.1 | 120 | 0.0 | 39 |
| | | 529 | Lost productivity (income loss) | . | . | 1.1 | 2,071 |
| Young adults and adults (25-64 years) | Parents | 1,301 | Primary sector | 0.6 | 456 | 0.0 | 14 |
| | | 1,301 | Hospital care | 4.5 | 3,448 | 0.8 | 587 |
| | | 1,301 | Filled prescriptions | 0.6 | 467 | 0.0 | 28 |
| | | 568 | Lost productivity (income loss) | . | . | -7.2 | -12,733 |
| Young adults and adults (25-64 years) | Partners | 5,403 | Primary sector | 1.6 | 288 | 0.0 | 4 |
| | | 5,403 | Hospital care | 11.0 | 2,040 | 3.2 | 590 |
| | | 5,403 | Filled prescriptions | 1.1 | 203 | 0.1 | 23 |
| | | 5,009 | Lost productivity (income loss) | . | . | 4.8 | 968 |
| Young adults and adults (25-64 years) | Siblings | 410 | Primary sector | 0.1 | 304 | 0.0 | 44 |
| | | 410 | Hospital care | 1.1 | 2,719 | 0.0 | 65 |
| | | 410 | Filled prescriptions | 0.1 | 191 | -0.0 | -1 |
| | | 347 | Lost productivity (income loss) | . | . | 0.8 | 2,309 |
| Older people (65+ years) | Children | 1,190 | Primary sector | 0.4 | 327 | 0.0 | 30 |
| | | 1,190 | Hospital care | 2.7 | 2,293 | 0.6 | 465 |
| | | 1,190 | Filled prescriptions | 0.3 | 271 | 0.1 | 56 |
| | | 1,078 | Lost productivity (income loss) | . | . | 3.3 | 3,106 |
| Older people (65+ years) | Partners | 2,076 | Primary sector | 1.1 | 521 | 0.0 | 24 |
| | | 2,076 | Hospital care | 7.7 | 3,765 | -0.6 | -308 |
| | | 2,076 | Filled prescriptions | 1.1 | 518 | 0.1 | 32 |
| | | 131 | Lost productivity (income loss) | . | . | -1.3 | -10,192 |
| Older people (65+ years) | Siblings | 48 | Primary sector | 0.0 | 364 | 0.0 | 1 |
| | | 48 | Hospital care | 0.0 | 845 | -0.2 | -3,456 |
| | | 48 | Filled prescriptions | 0.0 | 377 | -0.0 | -44 |
| | | 32 | Lost productivity (income loss) | . | . | 0.3 | 8,567 |

#### Supplementary Table 162
#### Intellectual disability - Incident cohort 2016-2021
#### Identification of populations of relatives

| ***Agegroup of index patients in matched population*** | | ***Closest relative type identified*** | | ***Available control with same type of closest relative*** |
|---|---|---|---|---|
| Children and young people (0-24 years) | n = 5,074 | Fathers | n = 4,174 (82%) | 94% |
| | | Mothers | n = 4,411 (87%) | 94% |
| | | No available closest relative | n = 357 (7%) | n/a |
| Young adults and adults (25-64 years) | n = 2,140 | Children | n = 104 (5%) | 92% |
| | | Parents | n = 878 (41%) | 91% |
| | | Partners | n = 320 (15%) | 93% |
| | | Siblings | n = 686 (32%) | 41% |
| | | Pooled | n = 1,988 (93%) | 74% |
| | | No available closest relative | n = 152 (7%) | n/a |
| Older people (65+ years) | n = 378 | Children | n = 32 (8%) | 96% |
| | | Partners | n = 32 (8%) | 90% |
| | | Siblings | n = 31 (8%) | 38% |
| | | Pooled | n = 95 (25%) | 75% |
| | | No available closest relative | n = 283 (75%) | n/a |

**Supplementary Table 163**
**Intellectual disability - Incident cohort 2016-2021**
**Patient characteristics - cases**

| ***Age strata of index patients*** | ***Characteristic*** | |
|---|---|---|
| Children and young people (0-24 years) | Number of persons, n | 4,475 (100.0) |
| | Male, n (%) | 2,801 (62.6) |
| | Age, median (Q1;Q3) | 12.4 (6.2;17.3) |
| | CCI, n (%) | |
| | 0 | 3,958 (88.4) |
| | 1-2 | 480 (10.7) |
| | +3 | 37 (0.8) |
| | Education level, n (%) | |
| | No/Unknown education | 2,473 (55.3) |
| | Primary | 1,966 (43.9) |
| | Secondary | 36 (0.8) |
| | Tertiary | 0 (0.0) |
| Young adults and adults (25-64 years) | Number of persons, n | 1,481 (100.0) |
| | Male, n (%) | 746 (50.4) |
| | Age, median (Q1;Q3) | 38.8 (30.2;48.8) |
| | CCI, n (%) | |
| | 0 | 1,233 (83.3) |
| | 1-2 | 207 (14.0) |
| | +3 | 41 (2.8) |
| | Education level, n (%) | |
| | No/Unknown education | 134 (9.0) |
| | Primary | 1,127 (76.1) |
| | Secondary | 173 (11.7) |
| | Tertiary | 47 (3.2) |
| Older people (65+ years) | Number of persons, n | 72 (100.0) |
| | Male, n (%) | 34 (47.2) |
| | Age, median (Q1;Q3) | 69.6 (66.8;75.6) |
| | CCI, n (%) | |
| | 0 | 33 (45.8) |
| | 1-2 | 30 (41.7) |
| | +3 | 9 (12.5) |
| | Education level, n (%) | |
| | No/Unknown education | * |
| | Primary | 51 (70.8) |
| | Secondary | * |
| | Tertiary | * |

**Supplementary Table 164**
**Intellectual disability - Incident cohort 2016-2021**
**Patient characteristics - relatives - pooled**

| *Age strata of index patients* | *Closest relative type* | *Characteristic of family relatives by agestrata of index patients* | *Closest family relatives of brain disorder patients* | *Comparison relative* |
|---|---|---|---|---|
| Children and young people (0-24 years) | Fathers | Number of persons, n | 3,953 (100.0) | 3,953 (100.0) |
| | | Male, n (%) | 3,953 (100.0) | 3,953 (100.0) |
| | | Age, median (Q1;Q3) | 45.0 (38.8;50.6) | 44.9 (39.1;50.4) |
| | | CCI, n (%) | | |
| | | 0 | 3,444 (87.1) | 3,549 (89.8) |
| | | 1-2 | 444 (11.2) | 370 (9.4) |
| | | +3 | 65 (1.6) | 34 (0.9) |
| | | Education level, n (%) | | |
| | | No/Unknown education | 151 (3.8) | 151 (3.8) |
| | | Primary | 1,452 (36.7) | 1,452 (36.7) |
| | | Secondary | 1,725 (43.6) | 1,725 (43.6) |
| | | Tertiary | 625 (15.8) | 625 (15.8) |
| | | Relation to patient, n (%) | | |
| | | Father | 3,953 (100.0) | 3,953 (100.0) |
| Children and young people (0-24 years) | Mothers | Number of persons, n | 4,183 (100.0) | 4,183 (100.0) |
| | | Male, n (%) | 0 (0.0) | 0 (0.0) |
| | | Age, median (Q1;Q3) | 41.9 (36.0;47.3) | 42.1 (36.3;47.5) |
| | | CCI, n (%) | | |
| | | 0 | 3,590 (85.8) | 3,761 (89.9) |
| | | 1-2 | 539 (12.9) | 396 (9.5) |
| | | +3 | 54 (1.3) | 26 (0.6) |
| | | Education level, n (%) | | |
| | | No/Unknown education | 146 (3.5) | 146 (3.5) |
| | | Primary | 1,458 (34.9) | 1,458 (34.9) |
| | | Secondary | 1,705 (40.8) | 1,705 (40.8) |
| | | Tertiary | 874 (20.9) | 874 (20.9) |
| | | Relation to patient, n (%) | | |
| | | Mother | 4,183 (100.0) | 4,183 (100.0) |
| Young adults and adults (25-64 years) | Pooled | Number of persons, n | 1,481 (100.0) | 1,481 (100.0) |
| | | Male, n (%) | 605 (40.9) | 605 (40.9) |
| | | Age, median (Q1;Q3) | 56.5 (42.9;65.2) | 56.6 (42.8;65.3) |
| | | CCI, n (%) | | |
| | | 0 | 1,148 (77.5) | 1,191 (80.4) |
| | | 1-2 | 256 (17.3) | 232 (15.7) |
| | | +3 | 77 (5.2) | 58 (3.9) |
| | | Education level, n (%) | | |
| | | No/Unknown education | 56 (3.8) | 56 (3.8) |
| | | Primary | 638 (43.1) | 638 (43.1) |
| | | Secondary | 585 (39.5) | 585 (39.5) |
| | | Tertiary | 202 (13.6) | 202 (13.6) |
| | | Relation to patient, n (%) | | |
| | | Mother | 628 (42.4) | 628 (42.4) |
| | | Father | 172 (11.6) | 172 (11.6) |
| | | Partner | 299 (20.2) | 299 (20.2) |
| | | Child | 96 (6.5) | 96 (6.5) |

**Supplementary Table 164**
**Intellectual disability - Incident cohort 2016-2021**
**Patient characteristics - relatives - pooled**

| *Age strata of index patients* | *Closest relative type* | *Characteristic of family relatives by agestrata of index patients* | *Closest family relatives of brain disorder patients* | *Comparison relative* |
|---|---|---|---|---|
| | | Sibling | 286 (19.3) | 286 (19.3) |
| Older people (65+ years) | Pooled | Number of persons, n | 72 (100.0) | 72 (100.0) |
| | | Male, n (%) | 32 (44.4) | 32 (44.4) |
| | | Age, median (Q1;Q3) | 61.3 (48.7;68.9) | 60.3 (49.5;68.9) |
| | | CCI, n (%) | | |
| | | 0 | 60 (83.3) | 53 (73.6) |
| | | 1-2 | * | * |
| | | +3 | * | * |
| | | Education level, n (%) | | |
| | | No/Unknown education | 0 (0.0) | 0 (0.0) |
| | | Primary | 28 (38.9) | 28 (38.9) |
| | | Secondary | 30 (41.7) | 30 (41.7) |
| | | Tertiary | 14 (19.4) | 14 (19.4) |
| | | Relation to patient, n (%) | | |
| | | Partner | 29 (40.3) | 29 (40.3) |
| | | Child | 31 (43.1) | 31 (43.1) |
| | | Sibling | 12 (16.7) | 12 (16.7) |

**Supplementary Table 165**
**Intellectual disability - Incident cohort 2016-2021**
**Patient characteristics - relatives - not pooled**

| *Age strata of index patients* | *Closest relative type* | *Characteristic of family relatives by agestrata of index patients* | *Closest family relatives of brain disorder patients* | *Comparison relative* |
|---|---|---|---|---|
| Children and young people (0-24 years) | Fathers | Number of persons, n | 3,953 (100.0) | 3,953 (100.0) |
| | | Male, n (%) | 3,953 (100.0) | 3,953 (100.0) |
| | | Age, median (Q1;Q3) | 45.0 (38.8;50.6) | 44.9 (39.1;50.4) |
| | | CCI, n (%) | | |
| | | 0 | 3,444 (87.1) | 3,549 (89.8) |
| | | 1-2 | 444 (11.2) | 370 (9.4) |
| | | +3 | 65 (1.6) | 34 (0.9) |
| | | Education level, n (%) | | |
| | | No/Unknown education | 151 (3.8) | 151 (3.8) |
| | | Primary | 1,452 (36.7) | 1,452 (36.7) |
| | | Secondary | 1,725 (43.6) | 1,725 (43.6) |
| | | Tertiary | 625 (15.8) | 625 (15.8) |
| | | Relation to patient, n (%) | | |
| | | Father | 3,953 (100.0) | 3,953 (100.0) |
| Children and young people (0-24 years) | Mothers | Number of persons, n | 4,183 (100.0) | 4,183 (100.0) |
| | | Male, n (%) | 0 (0.0) | 0 (0.0) |
| | | Age, median (Q1;Q3) | 41.9 (36.0;47.3) | 42.1 (36.3;47.5) |
| | | CCI, n (%) | | |
| | | 0 | 3,590 (85.8) | 3,761 (89.9) |
| | | 1-2 | 539 (12.9) | 396 (9.5) |
| | | +3 | 54 (1.3) | 26 (0.6) |
| | | Education level, n (%) | | |
| | | No/Unknown education | 146 (3.5) | 146 (3.5) |
| | | Primary | 1,458 (34.9) | 1,458 (34.9) |
| | | Secondary | 1,705 (40.8) | 1,705 (40.8) |
| | | Tertiary | 874 (20.9) | 874 (20.9) |
| | | Relation to patient, n (%) | | |
| | | Mother | 4,183 (100.0) | 4,183 (100.0) |
| Young adults and adults (25-64 years) | Children | Number of persons, n | 96 (100.0) | 96 (100.0) |
| | | Male, n (%) | 42 (43.8) | 42 (43.8) |
| | | Age, median (Q1;Q3) | 15.2 (9.5;24.8) | 15.6 (7.7;24.1) |
| | | CCI, n (%) | | |
| | | 0 | 91 (94.8) | * |
| | | 1-2 | 5 (5.2) | * |
| | | +3 | 0 (0.0) | 0 (0.0) |
| | | Education level, n (%) | | |
| | | No/Unknown education | 40 (41.7) | 40 (41.7) |
| | | Primary | 36 (37.5) | 36 (37.5) |
| | | Secondary | * | * |
| | | Tertiary | * | * |
| | | Relation to patient, n (%) | | |
| | | Child | 96 (100.0) | 96 (100.0) |
| Young adults and adults (25-64 years) | Parents | Number of persons, n | 800 (100.0) | 800 (100.0) |
| | | Male, n (%) | 172 (21.5) | 172 (21.5) |
| | | Age, median (Q1;Q3) | 63.6 (57.2;70.7) | 64.1 (57.3;71.4) |

### Supplementary Table 165
#### Intellectual disability - Incident cohort 2016-2021
#### Patient characteristics - relatives - not pooled

| *Age strata of index patients* | *Closest relative type* | *Characteristic of family relatives by agestrata of index patients* | *Closest family relatives of brain disorder patients* | *Comparison relative* |
|---|---|---|---|---|
| | | CCI, n (%) | | |
| | | 0 | 553 (69.1) | 569 (71.1) |
| | | 1-2 | 184 (23.0) | 177 (22.1) |
| | | +3 | 63 (7.9) | 54 (6.8) |
| | | Education level, n (%) | | |
| | | No/Unknown education | 10 (1.3) | 10 (1.3) |
| | | Primary | 350 (43.8) | 350 (43.8) |
| | | Secondary | 322 (40.3) | 322 (40.3) |
| | | Tertiary | 118 (14.8) | 118 (14.8) |
| | | Relation to patient, n (%) | | |
| | | Mother | 628 (78.5) | 628 (78.5) |
| | | Father | 172 (21.5) | 172 (21.5) |
| Young adults and adults (25-64 years) | Partners | Number of persons, n | 299 (100.0) | 299 (100.0) |
| | | Male, n (%) | 211 (70.6) | 211 (70.6) |
| | | Age, median (Q1;Q3) | 39.5 (32.3;47.2) | 39.8 (31.7;47.8) |
| | | CCI, n (%) | | |
| | | 0 | 261 (87.3) | 275 (92.0) |
| | | 1-2 | 31 (10.4) | * |
| | | +3 | 7 (2.3) | * |
| | | Education level, n (%) | | |
| | | No/Unknown education | 6 (2.0) | 6 (2.0) |
| | | Primary | 150 (50.2) | 150 (50.2) |
| | | Secondary | 107 (35.8) | 107 (35.8) |
| | | Tertiary | 36 (12.0) | 36 (12.0) |
| | | Relation to patient, n (%) | | |
| | | Partner | 299 (100.0) | 299 (100.0) |
| Young adults and adults (25-64 years) | Siblings | Number of persons, n | 286 (100.0) | 286 (100.0) |
| | | Male, n (%) | 180 (62.9) | 180 (62.9) |
| | | Age, median (Q1;Q3) | 52.4 (38.2;58.1) | 52.5 (38.5;58.3) |
| | | CCI, n (%) | | |
| | | 0 | 243 (85.0) | 254 (88.8) |
| | | 1-2 | 36 (12.6) | * |
| | | +3 | 7 (2.4) | * |
| | | Education level, n (%) | | |
| | | No/Unknown education | 0 (0.0) | 0 (0.0) |
| | | Primary | 102 (35.7) | 102 (35.7) |
| | | Secondary | 140 (49.0) | 140 (49.0) |
| | | Tertiary | 44 (15.4) | 44 (15.4) |
| | | Relation to patient, n (%) | | |
| | | Sibling | 286 (100.0) | 286 (100.0) |
| Older people (65+ years) | Children | Number of persons, n | 31 (100.0) | 31 (100.0) |
| | | Male, n (%) | 11 (35.5) | 11 (35.5) |
| | | Age, median (Q1;Q3) | 47.1 (42.0;52.7) | 47.5 (43.5;52.3) |
| | | CCI, n (%) | | |
| | | 0 | * | * |

**Supplementary Table 165**
**Intellectual disability - Incident cohort 2016-2021**
**Patient characteristics - relatives - not pooled**

| *Age strata of index patients* | *Closest relative type* | *Characteristic of family relatives by agestrata of index patients* | *Closest family relatives of brain disorder patients* | *Comparison relative* |
|---|---|---|---|---|
| | | 1-2 | * | * |
| | | +3 | * | 0 (0.0) |
| | | Education level, n (%) | | |
| | | No/Unknown education | 0 (0.0) | 0 (0.0) |
| | | Primary | 8 (25.8) | 8 (25.8) |
| | | Secondary | 13 (41.9) | 13 (41.9) |
| | | Tertiary | 10 (32.3) | 10 (32.3) |
| | | Relation to patient, n (%) | | |
| | | Child | 31 (100.0) | 31 (100.0) |
| Older people (65+ years) | Partners | Number of persons, n | 29 (100.0) | 29 (100.0) |
| | | Male, n (%) | 14 (48.3) | 14 (48.3) |
| | | Age, median (Q1;Q3) | 71.3 (65.9;72.6) | 70.1 (66.4;73.5) |
| | | CCI, n (%) | | |
| | | 0 | * | 16 (55.2) |
| | | 1-2 | * | * |
| | | +3 | * | * |
| | | Education level, n (%) | | |
| | | No/Unknown education | 0 (0.0) | 0 (0.0) |
| | | Primary | 17 (58.6) | 17 (58.6) |
| | | Secondary | * | * |
| | | Tertiary | * | * |
| | | Relation to patient, n (%) | | |
| | | Partner | 29 (100.0) | 29 (100.0) |
| Older people (65+ years) | Siblings | Number of persons, n | 12 (100.0) | 12 (100.0) |
| | | Male, n (%) | 7 (58.3) | 7 (58.3) |
| | | Age, median (Q1;Q3) | 63.1 (58.7;64.6) | 60.6 (58.6;64.7) |
| | | CCI, n (%) | | |
| | | 0 | 12 (100.0) | * |
| | | 1-2 | 0 (0.0) | * |
| | | +3 | 0 (0.0) | 0 (0.0) |
| | | Education level, n (%) | | |
| | | No/Unknown education | 0 (0.0) | 0 (0.0) |
| | | Primary | * | * |
| | | Secondary | * | * |
| | | Tertiary | * | * |
| | | Relation to patient, n (%) | | |
| | | Sibling | 12 (100.0) | 12 (100.0) |

#### Supplementary Table 166
#### Intellectual disability - Incident cohort 2016-2021
#### Cost analysis - pooled

| | | | | *Actual costs* | | *Attributable costs* | |
|---|---|---|---|---|---|---|---|
| *Age strata of index patients* | *Closest relative type* | *Number of index patients included in cost component calculations* | *Cost component* | *Total (million EUR)* | *Per person (EUR)* | *Total (million EUR)* | *Per person (EUR)* |
| Children and young people (0-24 years) | Fathers | 3,953 | Primary sector | 0.9 | 221 | 0.1 | 26 |
| | | 3,953 | Hospital care | 7.2 | 1,817 | 1.8 | 468 |
| | | 3,953 | Filled prescriptions | 0.8 | 213 | 0.3 | 71 |
| | | 3,863 | Lost productivity (income loss) | . | . | 64.8 | 16,776 |
| Children and young people (0-24 years) | Mothers | 4,183 | Primary sector | 1.5 | 368 | 0.1 | 28 |
| | | 4,183 | Hospital care | 9.2 | 2,193 | 2.7 | 646 |
| | | 4,183 | Filled prescriptions | 1.0 | 239 | 0.3 | 76 |
| | | 4,159 | Lost productivity (income loss) | . | . | 53.4 | 12,844 |
| Young adults and adults (25-64 years) | Pooled | 1,481 | Primary sector | 0.5 | 352 | -0.0 | -1 |
| | | 1,481 | Hospital care | 4.6 | 3,117 | 1.1 | 761 |
| | | 1,481 | Filled prescriptions | 0.6 | 427 | 0.1 | 87 |
| | | 931 | Lost productivity (income loss) | . | . | 10.3 | 11,014 |
| Older people (65+ years) | Pooled | 72 | Primary sector | 0.0 | 417 | 0.0 | 55 |
| | | 72 | Hospital care | 0.2 | 3,105 | 0.1 | 1,692 |
| | | 72 | Filled prescriptions | 0.0 | 477 | 0.0 | 244 |
| | | 39 | Lost productivity (income loss) | . | . | -0.3 | -6,484 |

#### Supplementary Table 167
#### Intellectual disability - Incident cohort 2016-2021
#### Cost analysis - not pooled

| | | | | *Actual costs* | | *Attributable costs* | |
|---|---|---|---|---|---|---|---|
| ***Age strata of index patients*** | ***Closest relative type*** | ***Number of index patients included in cost component calculations*** | ***Cost component*** | ***Total (million EUR)*** | ***Per person (EUR)*** | ***Total (million EUR)*** | ***Per person (EUR)*** |
| Children and young people (0-24 years) | Fathers | 3,953 | Primary sector | 0.9 | 221 | 0.1 | 26 |
| | | 3,953 | Hospital care | 7.2 | 1,817 | 1.8 | 468 |
| | | 3,953 | Filled prescriptions | 0.8 | 213 | 0.3 | 71 |
| | | 3,863 | Lost productivity (income loss) | . | . | 64.8 | 16,776 |
| Children and young people (0-24 years) | Mothers | 4,183 | Primary sector | 1.5 | 368 | 0.1 | 28 |
| | | 4,183 | Hospital care | 9.2 | 2,193 | 2.7 | 646 |
| | | 4,183 | Filled prescriptions | 1.0 | 239 | 0.3 | 76 |
| | | 4,159 | Lost productivity (income loss) | . | . | 53.4 | 12,844 |
| Young adults and adults (25-64 years) | Children | 96 | Primary sector | 0.0 | 194 | 0.0 | 31 |
| | | 96 | Hospital care | 0.1 | 1,483 | 0.1 | 784 |
| | | 96 | Filled prescriptions | 0.0 | 57 | -0.0 | -9 |
| | | 37 | Lost productivity (income loss) | . | . | 0.1 | 3,003 |
| Young adults and adults (25-64 years) | Parents | 800 | Primary sector | 0.3 | 431 | -0.0 | -8 |
| | | 800 | Hospital care | 3.1 | 3,897 | 0.7 | 861 |
| | | 800 | Filled prescriptions | 0.4 | 553 | 0.1 | 66 |
| | | 352 | Lost productivity (income loss) | . | . | 4.3 | 12,228 |
| Young adults and adults (25-64 years) | Partners | 299 | Primary sector | 0.1 | 271 | 0.0 | 37 |
| | | 299 | Hospital care | 0.7 | 2,366 | 0.3 | 1,126 |
| | | 299 | Filled prescriptions | 0.1 | 292 | 0.0 | 149 |
| | | 289 | Lost productivity (income loss) | . | . | 4.7 | 16,109 |
| Young adults and adults (25-64 years) | Siblings | 286 | Primary sector | 0.1 | 272 | -0.0 | -33 |
| | | 286 | Hospital care | 0.7 | 2,294 | 0.0 | 90 |
| | | 286 | Filled prescriptions | 0.1 | 346 | 0.0 | 113 |
| | | 253 | Lost productivity (income loss) | . | . | 1.2 | 4,678 |
| Older people (65+ years) | Children | 31 | Primary sector | 0.0 | 262 | -0.0 | -40 |
| | | 31 | Hospital care | 0.0 | 869 | -0.0 | -523 |
| | | 31 | Filled prescriptions | 0.0 | 296 | 0.0 | 162 |
| | | 30 | Lost productivity (income loss) | . | . | -0.3 | -9,043 |
| Older people (65+ years) | Partners | 29 | Primary sector | 0.0 | 611 | 0.0 | 138 |
| | | 29 | Hospital care | 0.2 | 6,481 | 0.1 | 4,671 |
| | | 29 | Filled prescriptions | 0.0 | 806 | 0.0 | 485 |
| | | . | Lost productivity (income loss) | . | . | . | . |
| Older people (65+ years) | Siblings | 12 | Primary sector | 0.0 | 368 | 0.0 | 109 |
| | | 12 | Hospital care | 0.0 | 1,056 | 0.0 | 516 |
| | | 12 | Filled prescriptions | 0.0 | 184 | -0.0 | -104 |
| | | 6 | Lost productivity (income loss) | . | . | -0.0 | -7,547 |

#### Supplementary Table 168
#### Intellectual disability - Prevalent cohort 2021
#### Identification of populations of relatives

| ***Agegroup of index patients in matched population*** | | ***Closest relative type identified*** | | ***Available control with same type of closest relative*** |
|---|---|---|---|---|
| Children and young people (0-24 years) | n = 13,152 | Fathers | n = 11,137 (85%) | 94% |
| | | Mothers | n = 11,984 (91%) | 95% |
| | | No available closest relative | n = 444 (3%) | n/a |
| Young adults and adults (25-64 years) | n = 15,195 | Children | n = 545 (4%) | 92% |
| | | Parents | n = 6,617 (44%) | 91% |
| | | Partners | n = 2,100 (14%) | 92% |
| | | Siblings | n = 5,176 (34%) | 41% |
| | | Pooled | n = 14,438 (95%) | 74% |
| | | No available closest relative | n = 757 (5%) | n/a |
| Older people (65+ years) | n = 1,866 | Children | n = 179 (10%) | 91% |
| | | Partners | n = 97 (5%) | 93% |
| | | Siblings | n = 313 (17%) | 50% |
| | | Pooled | n = 589 (32%) | 70% |
| | | No available closest relative | n = 1,277 (68%) | n/a |

#### Supplementary Table 169
#### Intellectual disability - Prevalent cohort 2021
#### Patient characteristics - cases

| *Age strata of index patients* | *Characteristic* | |
|---|---|---|
| Children and young people (0-24 years) | Number of persons, n | 12,087 (100.0) |
| | Male, n (%) | 7,856 (65.0) |
| | Age, median (Q1;Q3) | 18.5 (13.6;21.8) |
| | CCI, n (%) | |
| | 0 | 10,693 (88.5) |
| | 1-2 | 1,270 (10.5) |
| | +3 | 124 (1.0) |
| | Education level, n (%) | |
| | No/Unknown education | 3,004 (24.9) |
| | Primary | 8,721 (72.2) |
| | Secondary | 354 (2.9) |
| | Tertiary | 8 (0.1) |
| Young adults and adults (25-64 years) | Number of persons, n | 10,695 (100.0) |
| | Male, n (%) | 5,720 (53.5) |
| | Age, median (Q1;Q3) | 35.2 (28.8;48.6) |
| | CCI, n (%) | |
| | 0 | 8,729 (81.6) |
| | 1-2 | 1,701 (15.9) |
| | +3 | 265 (2.5) |
| | Education level, n (%) | |
| | No/Unknown education | 874 (8.2) |
| | Primary | 8,606 (80.5) |
| | Secondary | 1,009 (9.4) |
| | Tertiary | 206 (1.9) |
| Older people (65+ years) | Number of persons, n | 414 (100.0) |
| | Male, n (%) | 168 (40.6) |
| | Age, median (Q1;Q3) | 68.4 (66.6;74.6) |
| | CCI, n (%) | |
| | 0 | 199 (48.1) |
| | 1-2 | 158 (38.2) |
| | +3 | 57 (13.8) |
| | Education level, n (%) | |
| | No/Unknown education | 22 (5.3) |
| | Primary | 305 (73.7) |
| | Secondary | 65 (15.7) |
| | Tertiary | 22 (5.3) |

#### Supplementary Table 170
#### Intellectual disability - Prevalent cohort 2021
#### Patient characteristics - relatives - pooled

| *Age strata of index patients* | *Closest relative type* | *Characteristic of family relatives by agestrata of index patients* | *Closest family relatives of brain disorder patients* | *Comparison relative* |
|---|---|---|---|---|
| Children and young people (0-24 years) | Fathers | Number of persons, n | 10,580 (100.0) | 10,580 (100.0) |
| | | Male, n (%) | 10,580 (100.0) | 10,580 (100.0) |
| | | Age, median (Q1;Q3) | 50.1 (44.9;55.1) | 50.2 (45.2;54.7) |
| | | CCI, n (%) | | |
| | | 0 | 8,866 (83.8) | 9,246 (87.4) |
| | | 1-2 | 1,475 (13.9) | 1,167 (11.0) |
| | | +3 | 239 (2.3) | 167 (1.6) |
| | | Education level, n (%) | | |
| | | No/Unknown education | 270 (2.6) | 270 (2.6) |
| | | Primary | 3,435 (32.5) | 3,435 (32.5) |
| | | Secondary | 5,011 (47.4) | 5,011 (47.4) |
| | | Tertiary | 1,864 (17.6) | 1,864 (17.6) |
| | | Relation to patient, n (%) | | |
| | | Father | 10,580 (100.0) | 10,580 (100.0) |
| Children and young people (0-24 years) | Mothers | Number of persons, n | 11,399 (100.0) | 11,399 (100.0) |
| | | Male, n (%) | 0 (0.0) | 0 (0.0) |
| | | Age, median (Q1;Q3) | 47.3 (42.5;52.2) | 47.5 (42.8;51.9) |
| | | CCI, n (%) | | |
| | | 0 | 9,471 (83.1) | 9,912 (87.0) |
| | | 1-2 | 1,750 (15.4) | 1,345 (11.8) |
| | | +3 | 178 (1.6) | 142 (1.2) |
| | | Education level, n (%) | | |
| | | No/Unknown education | 221 (1.9) | 221 (1.9) |
| | | Primary | 3,403 (29.9) | 3,403 (29.9) |
| | | Secondary | 4,984 (43.7) | 4,984 (43.7) |
| | | Tertiary | 2,791 (24.5) | 2,791 (24.5) |
| | | Relation to patient, n (%) | | |
| | | Mother | 11,399 (100.0) | 11,399 (100.0) |
| Young adults and adults (25-64 years) | Pooled | Number of persons, n | 10,695 (100.0) | 10,695 (100.0) |
| | | Male, n (%) | 4,185 (39.1) | 4,185 (39.1) |
| | | Age, median (Q1;Q3) | 56.9 (44.6;64.8) | 57.2 (44.9;64.5) |
| | | CCI, n (%) | | |
| | | 0 | 8,073 (75.5) | 8,448 (79.0) |
| | | 1-2 | 2,132 (19.9) | 1,849 (17.3) |
| | | +3 | 490 (4.6) | 398 (3.7) |
| | | Education level, n (%) | | |
| | | No/Unknown education | 376 (3.5) | 376 (3.5) |
| | | Primary | 3,964 (37.1) | 3,964 (37.1) |
| | | Secondary | 4,653 (43.5) | 4,653 (43.5) |
| | | Tertiary | 1,702 (15.9) | 1,702 (15.9) |
| | | Relation to patient, n (%) | | |
| | | Mother | 4,753 (44.4) | 4,753 (44.4) |
| | | Father | 1,328 (12.4) | 1,328 (12.4) |
| | | Partner | 1,952 (18.3) | 1,952 (18.3) |
| | | Child | 505 (4.7) | 505 (4.7) |

**Supplementary Table 170**
**Intellectual disability - Prevalent cohort 2021**
**Patient characteristics - relatives - pooled**

| *Age strata of index patients* | *Closest relative type* | *Characteristic of family relatives by agestrata of index patients* | *Closest family relatives of brain disorder patients* | *Comparison relative* |
|---|---|---|---|---|
| | | Sibling | 2,157 (20.2) | 2,157 (20.2) |
| Older people (65+ years) | Pooled | Number of persons, n | 414 (100.0) | 414 (100.0) |
| | | Male, n (%) | 199 (48.1) | 199 (48.1) |
| | | Age, median (Q1;Q3) | 60.7 (50.9;66.3) | 60.4 (52.0;66.5) |
| | | CCI, n (%) | | |
| | | 0 | 316 (76.3) | 311 (75.1) |
| | | 1-2 | 85 (20.5) | 85 (20.5) |
| | | +3 | 13 (3.1) | 18 (4.3) |
| | | Education level, n (%) | | |
| | | No/Unknown education | * | * |
| | | Primary | 145 (35.0) | 145 (35.0) |
| | | Secondary | 194 (46.9) | 194 (46.9) |
| | | Tertiary | * | * |
| | | Relation to patient, n (%) | | |
| | | Partner | 91 (22.0) | 91 (22.0) |
| | | Child | 164 (39.6) | 164 (39.6) |
| | | Sibling | 159 (38.4) | 159 (38.4) |

**Supplementary Table 171**
**Intellectual disability - Prevalent cohort 2021**
**Patient characteristics - relatives - not pooled**

| *Age strata of index patients* | *Closest relative type* | *Characteristic of family relatives by agestrata of index patients* | *Closest family relatives of brain disorder patients* | *Comparison relative* |
|---|---|---|---|---|
| Children and young people (0-24 years) | Fathers | Number of persons, n | 10,580 (100.0) | 10,580 (100.0) |
| | | Male, n (%) | 10,580 (100.0) | 10,580 (100.0) |
| | | Age, median (Q1;Q3) | 50.1 (44.9;55.1) | 50.2 (45.2;54.7) |
| | | CCI, n (%) | | |
| | | 0 | 8,866 (83.8) | 9,246 (87.4) |
| | | 1-2 | 1,475 (13.9) | 1,167 (11.0) |
| | | +3 | 239 (2.3) | 167 (1.6) |
| | | Education level, n (%) | | |
| | | No/Unknown education | 270 (2.6) | 270 (2.6) |
| | | Primary | 3,435 (32.5) | 3,435 (32.5) |
| | | Secondary | 5,011 (47.4) | 5,011 (47.4) |
| | | Tertiary | 1,864 (17.6) | 1,864 (17.6) |
| | | Relation to patient, n (%) | | |
| | | Father | 10,580 (100.0) | 10,580 (100.0) |
| Children and young people (0-24 years) | Mothers | Number of persons, n | 11,399 (100.0) | 11,399 (100.0) |
| | | Male, n (%) | 0 (0.0) | 0 (0.0) |
| | | Age, median (Q1;Q3) | 47.3 (42.5;52.2) | 47.5 (42.8;51.9) |
| | | CCI, n (%) | | |
| | | 0 | 9,471 (83.1) | 9,912 (87.0) |
| | | 1-2 | 1,750 (15.4) | 1,345 (11.8) |
| | | +3 | 178 (1.6) | 142 (1.2) |
| | | Education level, n (%) | | |
| | | No/Unknown education | 221 (1.9) | 221 (1.9) |
| | | Primary | 3,403 (29.9) | 3,403 (29.9) |
| | | Secondary | 4,984 (43.7) | 4,984 (43.7) |
| | | Tertiary | 2,791 (24.5) | 2,791 (24.5) |
| | | Relation to patient, n (%) | | |
| | | Mother | 11,399 (100.0) | 11,399 (100.0) |
| Young adults and adults (25-64 years) | Children | Number of persons, n | 505 (100.0) | 505 (100.0) |
| | | Male, n (%) | 261 (51.7) | 261 (51.7) |
| | | Age, median (Q1;Q3) | 15.3 (7.2;26.4) | 15.2 (6.3;25.2) |
| | | CCI, n (%) | | |
| | | 0 | 470 (93.1) | 476 (94.3) |
| | | 1-2 | * | 29 (5.7) |
| | | +3 | * | 0 (0.0) |
| | | Education level, n (%) | | |
| | | No/Unknown education | 227 (45.0) | 227 (45.0) |
| | | Primary | 181 (35.8) | 181 (35.8) |
| | | Secondary | 76 (15.0) | 76 (15.0) |
| | | Tertiary | 21 (4.2) | 21 (4.2) |
| | | Relation to patient, n (%) | | |
| | | Child | 505 (100.0) | 505 (100.0) |
| Young adults and adults (25-64 years) | Parents | Number of persons, n | 6,081 (100.0) | 6,081 (100.0) |
| | | Male, n (%) | 1,328 (21.8) | 1,328 (21.8) |
| | | Age, median (Q1;Q3) | 62.3 (56.4;70.2) | 62.1 (56.7;70.4) |

**Supplementary Table 171**
**Intellectual disability - Prevalent cohort 2021**
**Patient characteristics - relatives - not pooled**

| *Age strata of index patients* | *Closest relative type* | *Characteristic of family relatives by agestrata of index patients* | *Closest family relatives of brain disorder patients* | *Comparison relative* |
|---|---|---|---|---|
| | | CCI, n (%) | | |
| | | 0 | 4,130 (67.9) | 4,407 (72.5) |
| | | 1-2 | 1,557 (25.6) | 1,354 (22.3) |
| | | +3 | 394 (6.5) | 320 (5.3) |
| | | Education level, n (%) | | |
| | | No/Unknown education | 105 (1.7) | 105 (1.7) |
| | | Primary | 2,130 (35.0) | 2,130 (35.0) |
| | | Secondary | 2,735 (45.0) | 2,735 (45.0) |
| | | Tertiary | 1,111 (18.3) | 1,111 (18.3) |
| | | Relation to patient, n (%) | | |
| | | Mother | 4,753 (78.2) | 4,753 (78.2) |
| | | Father | 1,328 (21.8) | 1,328 (21.8) |
| Young adults and adults (25-64 years) | Partners | Number of persons, n | 1,952 (100.0) | 1,952 (100.0) |
| | | Male, n (%) | 1,274 (65.3) | 1,274 (65.3) |
| | | Age, median (Q1;Q3) | 35.3 (28.7;46.6) | 34.9 (28.8;46.5) |
| | | CCI, n (%) | | |
| | | 0 | 1,703 (87.2) | 1,749 (89.6) |
| | | 1-2 | 214 (11.0) | 184 (9.4) |
| | | +3 | 35 (1.8) | 19 (1.0) |
| | | Education level, n (%) | | |
| | | No/Unknown education | 35 (1.8) | 35 (1.8) |
| | | Primary | 969 (49.6) | 969 (49.6) |
| | | Secondary | 741 (38.0) | 741 (38.0) |
| | | Tertiary | 207 (10.6) | 207 (10.6) |
| | | Relation to patient, n (%) | | |
| | | Partner | 1,952 (100.0) | 1,952 (100.0) |
| Young adults and adults (25-64 years) | Siblings | Number of persons, n | 2,157 (100.0) | 2,157 (100.0) |
| | | Male, n (%) | 1,322 (61.3) | 1,322 (61.3) |
| | | Age, median (Q1;Q3) | 52.4 (36.4;59.9) | 52.7 (36.3;60.0) |
| | | CCI, n (%) | | |
| | | 0 | 1,770 (82.1) | 1,816 (84.2) |
| | | 1-2 | 327 (15.2) | 282 (13.1) |
| | | +3 | 60 (2.8) | 59 (2.7) |
| | | Education level, n (%) | | |
| | | No/Unknown education | 9 (0.4) | 9 (0.4) |
| | | Primary | 684 (31.7) | 684 (31.7) |
| | | Secondary | 1,101 (51.0) | 1,101 (51.0) |
| | | Tertiary | 363 (16.8) | 363 (16.8) |
| | | Relation to patient, n (%) | | |
| | | Sibling | 2,157 (100.0) | 2,157 (100.0) |
| Older people (65+ years) | Children | Number of persons, n | 164 (100.0) | 164 (100.0) |
| | | Male, n (%) | 75 (45.7) | 75 (45.7) |
| | | Age, median (Q1;Q3) | 48.9 (45.1;54.9) | 49.4 (44.7;54.4) |
| | | CCI, n (%) | | |
| | | 0 | 140 (85.4) | 140 (85.4) |

### Supplementary Table 171
### Intellectual disability - Prevalent cohort 2021
### Patient characteristics - relatives - not pooled

| *Age strata of index patients* | *Closest relative type* | *Characteristic of family relatives by agestrata of index patients* | *Closest family relatives of brain disorder patients* | *Comparison relative* |
|---|---|---|---|---|
| | | 1-2 | * | * |
| | | +3 | * | * |
| | | Education level, n (%) | | |
| | | No/Unknown education | 0 (0.0) | 0 (0.0) |
| | | Primary | 67 (40.9) | 67 (40.9) |
| | | Secondary | 68 (41.5) | 68 (41.5) |
| | | Tertiary | 29 (17.7) | 29 (17.7) |
| | | Relation to patient, n (%) | | |
| | | Child | 164 (100.0) | 164 (100.0) |
| Older people (65+ years) | Partners | Number of persons, n | 91 (100.0) | 91 (100.0) |
| | | Male, n (%) | 50 (54.9) | 50 (54.9) |
| | | Age, median (Q1;Q3) | 71.4 (66.0;74.5) | 71.0 (66.5;75.3) |
| | | CCI, n (%) | | |
| | | 0 | 57 (62.6) | 56 (61.5) |
| | | 1-2 | 29 (31.9) | 27 (29.7) |
| | | +3 | 5 (5.5) | 8 (8.8) |
| | | Education level, n (%) | | |
| | | No/Unknown education | 0 (0.0) | 0 (0.0) |
| | | Primary | 41 (45.1) | 41 (45.1) |
| | | Secondary | 35 (38.5) | 35 (38.5) |
| | | Tertiary | 15 (16.5) | 15 (16.5) |
| | | Relation to patient, n (%) | | |
| | | Partner | 91 (100.0) | 91 (100.0) |
| Older people (65+ years) | Siblings | Number of persons, n | 159 (100.0) | 159 (100.0) |
| | | Male, n (%) | 74 (46.5) | 74 (46.5) |
| | | Age, median (Q1;Q3) | 63.0 (59.2;66.0) | 62.6 (59.3;65.0) |
| | | CCI, n (%) | | |
| | | 0 | 119 (74.8) | 115 (72.3) |
| | | 1-2 | 34 (21.4) | 37 (23.3) |
| | | +3 | 6 (3.8) | 7 (4.4) |
| | | Education level, n (%) | | |
| | | No/Unknown education | * | * |
| | | Primary | 37 (23.3) | 37 (23.3) |
| | | Secondary | 91 (57.2) | 91 (57.2) |
| | | Tertiary | * | * |
| | | Relation to patient, n (%) | | |
| | | Sibling | 159 (100.0) | 159 (100.0) |

#### Supplementary Table 172
#### Intellectual disability - Prevalent cohort 2021
#### Cost analysis - pooled

| *Age strata of index patients* | *Closest relative type* | *Number of index patients included in cost component calculations* | *Cost component* | *Actual costs* | | *Attributable costs* | |
|---|---|---|---|---|---|---|---|
| | | | | *Total (million EUR)* | *Per person (EUR)* | *Total (million EUR)* | *Per person (EUR)* |
| Children and young people (0-24 years) | Fathers | 10,580 | Primary sector | 2.6 | 244 | 0.2 | 20 |
| | | 10,580 | Hospital care | 20.4 | 1,930 | 3.1 | 294 |
| | | 10,580 | Filled prescriptions | 2.8 | 268 | 0.8 | 79 |
| | | 10,207 | Lost productivity (income loss) | . | . | 183.2 | 17,951 |
| Children and young people (0-24 years) | Mothers | 11,399 | Primary sector | 4.4 | 383 | 0.5 | 40 |
| | | 11,399 | Hospital care | 24.5 | 2,152 | 4.9 | 427 |
| | | 11,399 | Filled prescriptions | 3.2 | 284 | 0.9 | 83 |
| | | 11,310 | Lost productivity (income loss) | . | . | 128.4 | 11,357 |
| Young adults and adults (25-64 years) | Pooled | 10,695 | Primary sector | 4.0 | 380 | 0.3 | 29 |
| | | 10,695 | Hospital care | 30.6 | 2,877 | 5.8 | 543 |
| | | 10,695 | Filled prescriptions | 4.4 | 417 | 1.1 | 104 |
| | | 7,116 | Lost productivity (income loss) | . | . | 80.2 | 11,267 |
| Older people (65+ years) | Pooled | 414 | Primary sector | 0.2 | 390 | 0.0 | 45 |
| | | 414 | Hospital care | 1.3 | 3,170 | 0.4 | 1,069 |
| | | 414 | Filled prescriptions | 0.2 | 409 | 0.0 | 52 |
| | | 252 | Lost productivity (income loss) | . | . | 3.1 | 12,157 |

#### Supplementary Table 173
#### Intellectual disability - Prevalent cohort 2021
#### Cost analysis - not pooled

| | | | | Actual costs | | Attributable costs | |
|---|---|---|---|---|---|---|---|
| *Age strata of index patients* | *Closest relative type* | *Number of index patients included in cost component calculations* | *Cost component* | *Total (million EUR)* | *Per person (EUR)* | *Total (million EUR)* | *Per person (EUR)* |
| Children and young people (0-24 years) | Fathers | 10,580 | Primary sector | 2.6 | 244 | 0.2 | 20 |
| | | 10,580 | Hospital care | 20.4 | 1,930 | 3.1 | 294 |
| | | 10,580 | Filled prescriptions | 2.8 | 268 | 0.8 | 79 |
| | | 10,207 | Lost productivity (income loss) | . | . | 183.2 | 17,951 |
| Children and young people (0-24 years) | Mothers | 11,399 | Primary sector | 4.4 | 383 | 0.5 | 40 |
| | | 11,399 | Hospital care | 24.5 | 2,152 | 4.9 | 427 |
| | | 11,399 | Filled prescriptions | 3.2 | 284 | 0.9 | 83 |
| | | 11,310 | Lost productivity (income loss) | . | . | 128.4 | 11,357 |
| Young adults and adults (25-64 years) | Children | 505 | Primary sector | 0.1 | 226 | 0.0 | 21 |
| | | 505 | Hospital care | 0.6 | 1,245 | 0.0 | 74 |
| | | 505 | Filled prescriptions | 0.0 | 94 | 0.0 | 6 |
| | | 195 | Lost productivity (income loss) | . | . | 1.9 | 9,968 |
| Young adults and adults (25-64 years) | Parents | 6,081 | Primary sector | 2.7 | 451 | 0.2 | 33 |
| | | 6,081 | Hospital care | 20.7 | 3,435 | 3.9 | 652 |
| | | 6,081 | Filled prescriptions | 3.2 | 538 | 0.8 | 133 |
| | | 3,200 | Lost productivity (income loss) | . | . | 34.7 | 10,851 |
| Young adults and adults (25-64 years) | Partners | 1,952 | Primary sector | 0.6 | 288 | 0.1 | 44 |
| | | 1,952 | Hospital care | 3.7 | 1,900 | 1.0 | 500 |
| | | 1,952 | Filled prescriptions | 0.5 | 274 | 0.3 | 133 |
| | | 1,886 | Lost productivity (income loss) | . | . | 34.9 | 18,523 |
| Young adults and adults (25-64 years) | Siblings | 2,157 | Primary sector | 0.6 | 302 | 0.0 | 7 |
| | | 2,157 | Hospital care | 5.5 | 2,579 | 0.8 | 385 |
| | | 2,157 | Filled prescriptions | 0.6 | 283 | 0.0 | 18 |
| | | 1,835 | Lost productivity (income loss) | . | . | 8.6 | 4,671 |
| Older people (65+ years) | Children | 164 | Primary sector | 0.1 | 337 | 0.0 | 60 |
| | | 164 | Hospital care | 0.2 | 1,381 | -0.0 | -38 |
| | | 164 | Filled prescriptions | 0.1 | 308 | 0.0 | 69 |
| | | 154 | Lost productivity (income loss) | . | . | 1.7 | 10,910 |
| Older people (65+ years) | Partners | 91 | Primary sector | 0.0 | 463 | -0.0 | -2 |
| | | 91 | Hospital care | 0.4 | 4,514 | 0.1 | 1,300 |
| | | 91 | Filled prescriptions | 0.1 | 630 | 0.0 | 116 |
| | | 11 | Lost productivity (income loss) | . | . | 0.3 | 24,294 |
| Older people (65+ years) | Siblings | 159 | Primary sector | 0.1 | 405 | 0.0 | 57 |
| | | 159 | Hospital care | 0.7 | 4,268 | 0.3 | 2,087 |
| | | 159 | Filled prescriptions | 0.1 | 390 | -0.0 | -2 |
| | | 87 | Lost productivity (income loss) | . | . | 1.1 | 12,828 |

**Supplementary Table 174**
**Multiple sclerosis - Incident cohort 2016-2021**
**Identification of populations of relatives**

| *Agegroup of index patients in matched population* | | *Closest relative type identified* | | *Available control with same type of closest relative* |
|---|---|---|---|---|
| Children and young people (0-24 years) | n = 471 | Fathers | n = 393 (83%) | 94% |
| | | Mothers | n = 405 (86%) | 94% |
| | | No available closest relative | n = 37 (8%) | n/a |
| Young adults and adults (25-64 years) | n = 3,500 | Children | n = 358 (10%) | 96% |
| | | Parents | n = 519 (15%) | 92% |
| | | Partners | n = 2,223 (64%) | 96% |
| | | Siblings | n = 294 (8%) | 41% |
| | | Pooled | n = 3,394 (97%) | 91% |
| | | No available closest relative | n = 106 (3%) | n/a |
| Older people (65+ years) | n = 367 | Children | n = 112 (31%) | 90% |
| | | Partners | n = 217 (59%) | 97% |
| | | Siblings | n = 10 (3%) | 30% |
| | | Pooled | n = 339 (92%) | 93% |
| | | No available closest relative | n = 28 (8%) | n/a |

**Supplementary Table 175**
**Multiple sclerosis - Incident cohort 2016-2021**
**Patient characteristics - cases**

| *Age strata of index patients* | *Characteristic* | |
|---|---|---|
| Children and young people (0-24 years) | Number of persons, n | 411 (100.0) |
| | Male, n (%) | 121 (29.4) |
| | Age, median (Q1;Q3) | 22.1 (19.4;23.8) |
| | CCI, n (%) | |
| | 0 | 370 (90.0) |
| | 1-2 | * |
| | +3 | * |
| | Education level, n (%) | |
| | No/Unknown education | 7 (1.7) |
| | Primary | 232 (56.4) |
| | Secondary | 145 (35.3) |
| | Tertiary | 27 (6.6) |
| Young adults and adults (25-64 years) | Number of persons, n | 3,091 (100.0) |
| | Male, n (%) | 1,043 (33.7) |
| | Age, median (Q1;Q3) | 42.9 (34.2;51.4) |
| | CCI, n (%) | |
| | 0 | 2,692 (87.1) |
| | 1-2 | 348 (11.3) |
| | +3 | 51 (1.6) |
| | Education level, n (%) | |
| | No/Unknown education | 95 (3.1) |
| | Primary | 518 (16.8) |
| | Secondary | 1,405 (45.5) |
| | Tertiary | 1,073 (34.7) |
| Older people (65+ years) | Number of persons, n | 316 (100.0) |
| | Male, n (%) | 124 (39.2) |
| | Age, median (Q1;Q3) | 71.5 (68.1;75.5) |
| | CCI, n (%) | |
| | 0 | 189 (59.8) |
| | 1-2 | 91 (28.8) |
| | +3 | 36 (11.4) |
| | Education level, n (%) | |
| | No/Unknown education | 13 (4.1) |
| | Primary | 86 (27.2) |
| | Secondary | 154 (48.7) |
| | Tertiary | 63 (19.9) |

#### Supplementary Table 176
#### Multiple sclerosis - Incident cohort 2016-2021
#### Patient characteristics - relatives - pooled

| *Age strata of index patients* | *Closest relative type* | *Characteristic of family relatives by agestrata of index patients* | *Closest family relatives of brain disorder patients* | *Comparison relative* |
|---|---|---|---|---|
| Children and young people (0-24 years) | Fathers | Number of persons, n | 371 (100.0) | 371 (100.0) |
| | | Male, n (%) | 371 (100.0) | 371 (100.0) |
| | | Age, median (Q1;Q3) | 52.6 (49.4;56.7) | 52.5 (49.5;56.3) |
| | | CCI, n (%) | | |
| | | 0 | 316 (85.2) | 323 (87.1) |
| | | 1-2 | 48 (12.9) | 40 (10.8) |
| | | +3 | 7 (1.9) | 8 (2.2) |
| | | Education level, n (%) | | |
| | | No/Unknown education | 9 (2.4) | 9 (2.4) |
| | | Primary | 98 (26.4) | 98 (26.4) |
| | | Secondary | 212 (57.1) | 212 (57.1) |
| | | Tertiary | 52 (14.0) | 52 (14.0) |
| | | Relation to patient, n (%) | | |
| | | Father | 371 (100.0) | 371 (100.0) |
| Children and young people (0-24 years) | Mothers | Number of persons, n | 384 (100.0) | 384 (100.0) |
| | | Male, n (%) | 0 (0.0) | 0 (0.0) |
| | | Age, median (Q1;Q3) | 50.0 (46.3;54.2) | 50.3 (46.8;54.0) |
| | | CCI, n (%) | | |
| | | 0 | 326 (84.9) | 339 (88.3) |
| | | 1-2 | * | 40 (10.4) |
| | | +3 | * | 5 (1.3) |
| | | Education level, n (%) | | |
| | | No/Unknown education | * | * |
| | | Primary | * | * |
| | | Secondary | 184 (47.9) | 184 (47.9) |
| | | Tertiary | 110 (28.6) | 110 (28.6) |
| | | Relation to patient, n (%) | | |
| | | Mother | 384 (100.0) | 384 (100.0) |
| Young adults and adults (25-64 years) | Pooled | Number of persons, n | 3,091 (100.0) | 3,091 (100.0) |
| | | Male, n (%) | 1,806 (58.4) | 1,806 (58.4) |
| | | Age, median (Q1;Q3) | 45.7 (33.7;56.3) | 45.6 (33.8;56.3) |
| | | CCI, n (%) | | |
| | | 0 | 2,672 (86.4) | 2,640 (85.4) |
| | | 1-2 | 346 (11.2) | 379 (12.3) |
| | | +3 | 73 (2.4) | 72 (2.3) |
| | | Education level, n (%) | | |
| | | No/Unknown education | 201 (6.5) | 201 (6.5) |
| | | Primary | 667 (21.6) | 667 (21.6) |
| | | Secondary | 1,428 (46.2) | 1,428 (46.2) |
| | | Tertiary | 795 (25.7) | 795 (25.7) |
| | | Relation to patient, n (%) | | |
| | | Mother | 366 (11.8) | 366 (11.8) |
| | | Father | 112 (3.6) | 112 (3.6) |
| | | Partner | 2,145 (69.4) | 2,145 (69.4) |
| | | Child | 346 (11.2) | 346 (11.2) |

#### Supplementary Table 176
#### Multiple sclerosis - Incident cohort 2016-2021
#### Patient characteristics - relatives - pooled

| *Age strata of index patients* | *Closest relative type* | *Characteristic of family relatives by agestrata of index patients* | *Closest family relatives of brain disorder patients* | *Comparison relative* |
|---|---|---|---|---|
| | | Sibling | 122 (3.9) | 122 (3.9) |
| Older people (65+ years) | Pooled | Number of persons, n | 316 (100.0) | 316 (100.0) |
| | | Male, n (%) | 170 (53.8) | 170 (53.8) |
| | | Age, median (Q1;Q3) | 67.2 (55.3;72.7) | 66.8 (55.2;72.8) |
| | | CCI, n (%) | | |
| | | 0 | 221 (69.9) | 215 (68.0) |
| | | 1-2 | 76 (24.1) | 85 (26.9) |
| | | +3 | 19 (6.0) | 16 (5.1) |
| | | Education level, n (%) | | |
| | | No/Unknown education | 16 (5.1) | 16 (5.1) |
| | | Primary | 75 (23.7) | 75 (23.7) |
| | | Secondary | 148 (46.8) | 148 (46.8) |
| | | Tertiary | 77 (24.4) | 77 (24.4) |
| | | Relation to patient, n (%) | | |
| | | Partner | 212 (67.1) | 212 (67.1) |
| | | Child | * | * |
| | | Sibling | * | * |

#### Supplementary Table 177
#### Multiple sclerosis - Incident cohort 2016-2021
#### Patient characteristics - relatives - not pooled

| *Age strata of index patients* | *Closest relative type* | *Characteristic of family relatives by agestrata of index patients* | *Closest family relatives of brain disorder patients* | *Comparison relative* |
|---|---|---|---|---|
| Children and young people (0-24 years) | Fathers | Number of persons, n | 371 (100.0) | 371 (100.0) |
| | | Male, n (%) | 371 (100.0) | 371 (100.0) |
| | | Age, median (Q1;Q3) | 52.6 (49.4;56.7) | 52.5 (49.5;56.3) |
| | | CCI, n (%) | | |
| | | 0 | 316 (85.2) | 323 (87.1) |
| | | 1-2 | 48 (12.9) | 40 (10.8) |
| | | +3 | 7 (1.9) | 8 (2.2) |
| | | Education level, n (%) | | |
| | | No/Unknown education | 9 (2.4) | 9 (2.4) |
| | | Primary | 98 (26.4) | 98 (26.4) |
| | | Secondary | 212 (57.1) | 212 (57.1) |
| | | Tertiary | 52 (14.0) | 52 (14.0) |
| | | Relation to patient, n (%) | | |
| | | Father | 371 (100.0) | 371 (100.0) |
| Children and young people (0-24 years) | Mothers | Number of persons, n | 384 (100.0) | 384 (100.0) |
| | | Male, n (%) | 0 (0.0) | 0 (0.0) |
| | | Age, median (Q1;Q3) | 50.0 (46.3;54.2) | 50.3 (46.8;54.0) |
| | | CCI, n (%) | | |
| | | 0 | 326 (84.9) | 339 (88.3) |
| | | 1-2 | * | 40 (10.4) |
| | | +3 | * | 5 (1.3) |
| | | Education level, n (%) | | |
| | | No/Unknown education | * | * |
| | | Primary | * | * |
| | | Secondary | 184 (47.9) | 184 (47.9) |
| | | Tertiary | 110 (28.6) | 110 (28.6) |
| | | Relation to patient, n (%) | | |
| | | Mother | 384 (100.0) | 384 (100.0) |
| Young adults and adults (25-64 years) | Children | Number of persons, n | 346 (100.0) | 346 (100.0) |
| | | Male, n (%) | 170 (49.1) | 170 (49.1) |
| | | Age, median (Q1;Q3) | 17.0 (10.9;24.4) | 16.6 (9.8;23.6) |
| | | CCI, n (%) | | |
| | | 0 | 329 (95.1) | 323 (93.4) |
| | | 1-2 | * | 23 (6.6) |
| | | +3 | * | 0 (0.0) |
| | | Education level, n (%) | | |
| | | No/Unknown education | 128 (37.0) | 128 (37.0) |
| | | Primary | 137 (39.6) | 137 (39.6) |
| | | Secondary | 48 (13.9) | 48 (13.9) |
| | | Tertiary | 33 (9.5) | 33 (9.5) |
| | | Relation to patient, n (%) | | |
| | | Child | 346 (100.0) | 346 (100.0) |
| Young adults and adults (25-64 years) | Parents | Number of persons, n | 478 (100.0) | 478 (100.0) |
| | | Male, n (%) | 112 (23.4) | 112 (23.4) |
| | | Age, median (Q1;Q3) | 64.6 (57.9;72.4) | 64.8 (57.7;71.9) |

### Supplementary Table 177
### Multiple sclerosis - Incident cohort 2016-2021
### Patient characteristics - relatives - not pooled

| *Age strata of index patients* | *Closest relative type* | *Characteristic of family relatives by agestrata of index patients* | *Closest family relatives of brain disorder patients* | *Comparison relative* |
|---|---|---|---|---|
| | | CCI, n (%) | | |
| | | 0 | 323 (67.6) | 323 (67.6) |
| | | 1-2 | 113 (23.6) | 124 (25.9) |
| | | +3 | 42 (8.8) | 31 (6.5) |
| | | Education level, n (%) | | |
| | | No/Unknown education | 7 (1.5) | 7 (1.5) |
| | | Primary | 139 (29.1) | 139 (29.1) |
| | | Secondary | 245 (51.3) | 245 (51.3) |
| | | Tertiary | 87 (18.2) | 87 (18.2) |
| | | Relation to patient, n (%) | | |
| | | Mother | 366 (76.6) | 366 (76.6) |
| | | Father | 112 (23.4) | 112 (23.4) |
| Young adults and adults (25-64 years) | Partners | Number of persons, n | 2,145 (100.0) | 2,145 (100.0) |
| | | Male, n (%) | 1,449 (67.6) | 1,449 (67.6) |
| | | Age, median (Q1;Q3) | 44.5 (35.6;53.2) | 44.5 (35.6;53.0) |
| | | CCI, n (%) | | |
| | | 0 | 1,909 (89.0) | 1,894 (88.3) |
| | | 1-2 | 209 (9.7) | 216 (10.1) |
| | | +3 | 27 (1.3) | 35 (1.6) |
| | | Education level, n (%) | | |
| | | No/Unknown education | 66 (3.1) | 66 (3.1) |
| | | Primary | 367 (17.1) | 367 (17.1) |
| | | Secondary | 1,067 (49.7) | 1,067 (49.7) |
| | | Tertiary | 645 (30.1) | 645 (30.1) |
| | | Relation to patient, n (%) | | |
| | | Partner | 2,145 (100.0) | 2,145 (100.0) |
| Young adults and adults (25-64 years) | Siblings | Number of persons, n | 122 (100.0) | 122 (100.0) |
| | | Male, n (%) | 75 (61.5) | 75 (61.5) |
| | | Age, median (Q1;Q3) | 45.7 (32.7;55.1) | 46.5 (33.3;55.0) |
| | | CCI, n (%) | | |
| | | 0 | 111 (91.0) | 100 (82.0) |
| | | 1-2 | * | 16 (13.1) |
| | | +3 | * | 6 (4.9) |
| | | Education level, n (%) | | |
| | | No/Unknown education | 0 (0.0) | 0 (0.0) |
| | | Primary | 24 (19.7) | 24 (19.7) |
| | | Secondary | 68 (55.7) | 68 (55.7) |
| | | Tertiary | 30 (24.6) | 30 (24.6) |
| | | Relation to patient, n (%) | | |
| | | Sibling | 122 (100.0) | 122 (100.0) |
| Older people (65+ years) | Children | Number of persons, n | 101 (100.0) | 101 (100.0) |
| | | Male, n (%) | 55 (54.5) | 55 (54.5) |
| | | Age, median (Q1;Q3) | 51.0 (44.6;54.9) | 49.9 (44.1;54.7) |
| | | CCI, n (%) | | |
| | | 0 | 85 (84.2) | 84 (83.2) |

#### Supplementary Table 177
#### Multiple sclerosis - Incident cohort 2016-2021
#### Patient characteristics - relatives - not pooled

| *Age strata of index patients* | *Closest relative type* | *Characteristic of family relatives by agestrata of index patients* | *Closest family relatives of brain disorder patients* | *Comparison relative* |
|---|---|---|---|---|
| | | 1-2 | * | * |
| | | +3 | * | * |
| | | Education level, n (%) | | |
| | | No/Unknown education | 0 (0.0) | 0 (0.0) |
| | | Primary | 20 (19.8) | 20 (19.8) |
| | | Secondary | 47 (46.5) | 47 (46.5) |
| | | Tertiary | 34 (33.7) | 34 (33.7) |
| | | Relation to patient, n (%) | | |
| | | Child | 101 (100.0) | 101 (100.0) |
| Older people (65+ years) | Partners | Number of persons, n | 212 (100.0) | 212 (100.0) |
| | | Male, n (%) | 112 (52.8) | 112 (52.8) |
| | | Age, median (Q1;Q3) | 70.7 (67.1;75.4) | 70.7 (66.7;74.4) |
| | | CCI, n (%) | | |
| | | 0 | 134 (63.2) | 130 (61.3) |
| | | 1-2 | 62 (29.2) | 67 (31.6) |
| | | +3 | 16 (7.5) | 15 (7.1) |
| | | Education level, n (%) | | |
| | | No/Unknown education | 16 (7.5) | 16 (7.5) |
| | | Primary | 55 (25.9) | 55 (25.9) |
| | | Secondary | 98 (46.2) | 98 (46.2) |
| | | Tertiary | 43 (20.3) | 43 (20.3) |
| | | Relation to patient, n (%) | | |
| | | Partner | 212 (100.0) | 212 (100.0) |
| Older people (65+ years) | Siblings | Number of persons, n | * | * |
| | | Male, n (%) | * | * |
| | | Age, median (Q1;Q3) | 66.4 (61.2;67.1) | 67.0 (64.7;71.2) |
| | | CCI, n (%) | | |
| | | 0 | * | * |
| | | 1-2 | * | * |
| | | +3 | 0 (0.0) | 0 (0.0) |
| | | Education level, n (%) | | |
| | | No/Unknown education | 0 (0.0) | 0 (0.0) |
| | | Primary | 0 (0.0) | 0 (0.0) |
| | | Secondary | * | * |
| | | Tertiary | 0 (0.0) | 0 (0.0) |
| | | Relation to patient, n (%) | | |
| | | Sibling | 3 (100.0) | 3 (100.0) |

#### Supplementary Table 178
#### Multiple sclerosis - Incident cohort 2016-2021
#### Cost analysis - pooled

| *Age strata of index patients* | *Closest relative type* | *Number of index patients included in cost component calculations* | *Cost component* | *Actual costs* | | *Attributable costs* | |
|---|---|---|---|---|---|---|---|
| | | | | *Total (million EUR)* | *Per person (EUR)* | *Total (million EUR)* | *Per person (EUR)* |
| Children and young people (0-24 years) | Fathers | 371 | Primary sector | 0.1 | 249 | 0.0 | 6 |
| | | 371 | Hospital care | 0.7 | 1,900 | -0.0 | -63 |
| | | 371 | Filled prescriptions | 0.1 | 301 | 0.0 | 118 |
| | | 342 | Lost productivity (income loss) | . | . | 2.5 | 7,239 |
| Children and young people (0-24 years) | Mothers | 384 | Primary sector | 0.1 | 356 | 0.0 | 21 |
| | | 384 | Hospital care | 0.8 | 2,060 | 0.2 | 620 |
| | | 384 | Filled prescriptions | 0.1 | 211 | -0.0 | -35 |
| | | 375 | Lost productivity (income loss) | . | . | 2.3 | 6,111 |
| Young adults and adults (25-64 years) | Pooled | 3,091 | Primary sector | 0.9 | 282 | 0.0 | 13 |
| | | 3,091 | Hospital care | 5.9 | 1,911 | 0.8 | 267 |
| | | 3,091 | Filled prescriptions | 0.7 | 214 | 0.0 | 12 |
| | | 2,465 | Lost productivity (income loss) | . | . | -0.3 | -137 |
| Older people (65+ years) | Pooled | 316 | Primary sector | 0.1 | 409 | -0.0 | -3 |
| | | 316 | Hospital care | 0.8 | 2,698 | 0.1 | 182 |
| | | 316 | Filled prescriptions | 0.1 | 459 | 0.0 | 32 |
| | | 115 | Lost productivity (income loss) | . | . | 0.2 | 2,122 |

**Supplementary Table 179**
**Multiple sclerosis - Incident cohort 2016-2021**
**Cost analysis - not pooled**

| *Age strata of index patients* | *Closest relative type* | *Number of index patients included in cost component calculations* | *Cost component* | *Actual costs* Total (million EUR) | *Actual costs* Per person (EUR) | *Attributable costs* Total (million EUR) | *Attributable costs* Per person (EUR) |
|---|---|---|---|---|---|---|---|
| Children and young people (0-24 years) | Fathers | 371 | Primary sector | 0.1 | 249 | 0.0 | 6 |
| | | 371 | Hospital care | 0.7 | 1,900 | -0.0 | -63 |
| | | 371 | Filled prescriptions | 0.1 | 301 | 0.0 | 118 |
| | | 342 | Lost productivity (income loss) | . | . | 2.5 | 7,239 |
| Children and young people (0-24 years) | Mothers | 384 | Primary sector | 0.1 | 356 | 0.0 | 21 |
| | | 384 | Hospital care | 0.8 | 2,060 | 0.2 | 620 |
| | | 384 | Filled prescriptions | 0.1 | 211 | -0.0 | -35 |
| | | 375 | Lost productivity (income loss) | . | . | 2.3 | 6,111 |
| Young adults and adults (25-64 years) | Children | 346 | Primary sector | 0.1 | 192 | 0.0 | 15 |
| | | 346 | Hospital care | 0.3 | 860 | -0.0 | -74 |
| | | 346 | Filled prescriptions | 0.0 | 104 | -0.0 | -10 |
| | | 140 | Lost productivity (income loss) | . | . | -0.3 | -1,995 |
| Young adults and adults (25-64 years) | Parents | 478 | Primary sector | 0.2 | 442 | 0.0 | 23 |
| | | 478 | Hospital care | 2.0 | 4,243 | 0.6 | 1,163 |
| | | 478 | Filled prescriptions | 0.2 | 463 | 0.0 | 28 |
| | | 199 | Lost productivity (income loss) | . | . | 0.9 | 4,575 |
| Young adults and adults (25-64 years) | Partners | 2,145 | Primary sector | 0.6 | 261 | 0.0 | 13 |
| | | 2,145 | Hospital care | 3.4 | 1,601 | 0.5 | 254 |
| | | 2,145 | Filled prescriptions | 0.4 | 180 | 0.0 | 17 |
| | | 2,014 | Lost productivity (income loss) | . | . | -0.9 | -440 |
| Young adults and adults (25-64 years) | Siblings | 122 | Primary sector | 0.0 | 278 | -0.0 | -32 |
| | | 122 | Hospital care | 0.2 | 1,239 | -0.2 | -2,037 |
| | | 122 | Filled prescriptions | 0.0 | 152 | -0.0 | -85 |
| | | 112 | Lost productivity (income loss) | . | . | -0.1 | -737 |
| Older people (65+ years) | Children | 101 | Primary sector | 0.0 | 269 | 0.0 | 8 |
| | | 101 | Hospital care | 0.3 | 3,056 | 0.1 | 1,473 |
| | | 101 | Filled prescriptions | 0.0 | 253 | 0.0 | 27 |
| | | 98 | Lost productivity (income loss) | . | . | 0.4 | 4,032 |
| Older people (65+ years) | Partners | 212 | Primary sector | 0.1 | 475 | -0.0 | -10 |
| | | 212 | Hospital care | 0.5 | 2,565 | -0.1 | -423 |
| | | 212 | Filled prescriptions | 0.1 | 556 | 0.0 | 33 |
| | | 17 | Lost productivity (income loss) | . | . | -0.2 | -8,888 |
| Older people (65+ years) | Siblings | . | Primary sector | . | . | . | . |
| | | . | Hospital care | . | . | . | . |
| | | . | Filled prescriptions | . | . | . | . |

**Supplementary Table 180**
**Multiple sclerosis - Prevalent cohort 2021**
**Identification of populations of relatives**

| ***Agegroup of index patients in matched population*** | | ***Closest relative type identified*** | | ***Available control with same type of closest relative*** |
|---|---|---|---|---|
| Children and young people (0-24 years) | n = 267 | Fathers | n = 227 (85%) | 95% |
| | | Mothers | n = 238 (89%) | 95% |
| | | No available closest relative | n = 14 (5%) | n/a |
| Young adults and adults (25-64 years) | n = 12,259 | Children | n = 1,632 (13%) | 95% |
| | | Parents | n = 1,663 (14%) | 88% |
| | | Partners | n = 7,559 (62%) | 96% |
| | | Siblings | n = 1,181 (10%) | 49% |
| | | Pooled | n = 12,035 (98%) | 90% |
| | | No available closest relative | n = 224 (2%) | n/a |
| Older people (65+ years) | n = 3,784 | Children | n = 1,170 (31%) | 95% |
| | | Partners | n = 2,167 (57%) | 96% |
| | | Siblings | n = 132 (3%) | 50% |
| | | Pooled | n = 3,469 (92%) | 94% |
| | | No available closest relative | n = 315 (8%) | n/a |

**Supplementary Table 181**
**Multiple sclerosis - Prevalent cohort 2021**
**Patient characteristics - cases**

| ***Age strata of index patients*** | ***Characteristic*** | |
|---|---|---|
| Children and young people (0-24 years) | Number of persons, n | 243 (100.0) |
| | Male, n (%) | 68 (28.0) |
| | Age, median (Q1;Q3) | 22.8 (20.4;24.0) |
| | CCI, n (%) | |
| | 0 | 219 (90.1) |
| | 1-2 | * |
| | +3 | * |
| | Education level, n (%) | |
| | No/Unknown education | * |
| | Primary | 116 (47.7) |
| | Secondary | 108 (44.4) |
| | Tertiary | * |
| Young adults and adults (25-64 years) | Number of persons, n | 10,917 (100.0) |
| | Male, n (%) | 3,270 (30.0) |
| | Age, median (Q1;Q3) | 50.1 (42.0;57.2) |
| | CCI, n (%) | |
| | 0 | 9,016 (82.6) |
| | 1-2 | 1,641 (15.0) |
| | +3 | 260 (2.4) |
| | Education level, n (%) | |
| | No/Unknown education | 162 (1.5) |
| | Primary | 2,048 (18.8) |
| | Secondary | 5,184 (47.5) |
| | Tertiary | 3,523 (32.3) |
| Older people (65+ years) | Number of persons, n | 3,279 (100.0) |
| | Male, n (%) | 1,041 (31.7) |
| | Age, median (Q1;Q3) | 71.6 (67.9;75.7) |
| | CCI, n (%) | |
| | 0 | 1,937 (59.1) |
| | 1-2 | 1,011 (30.8) |
| | +3 | 331 (10.1) |
| | Education level, n (%) | |
| | No/Unknown education | 121 (3.7) |
| | Primary | 845 (25.8) |
| | Secondary | 1,605 (48.9) |
| | Tertiary | 708 (21.6) |

**Supplementary Table 182**
**Multiple sclerosis - Prevalent cohort 2021**
**Patient characteristics - relatives - pooled**

| *Age strata of index patients* | *Closest relative type* | *Characteristic of family relatives by agestrata of index patients* | *Closest family relatives of brain disorder patients* | *Comparison relative* |
|---|---|---|---|---|
| Children and young people (0-24 years) | Fathers | Number of persons, n | 217 (100.0) | 217 (100.0) |
| | | Male, n (%) | 217 (100.0) | 217 (100.0) |
| | | Age, median (Q1;Q3) | 53.9 (50.3;57.7) | 54.1 (50.4;56.9) |
| | | CCI, n (%) | | |
| | | 0 | 183 (84.3) | 186 (85.7) |
| | | 1-2 | 29 (13.4) | 25 (11.5) |
| | | +3 | 5 (2.3) | 6 (2.8) |
| | | Education level, n (%) | | |
| | | No/Unknown education | * | * |
| | | Primary | 58 (26.7) | 58 (26.7) |
| | | Secondary | 123 (56.7) | 123 (56.7) |
| | | Tertiary | * | * |
| | | Relation to patient, n (%) | | |
| | | Father | 217 (100.0) | 217 (100.0) |
| Children and young people (0-24 years) | Mothers | Number of persons, n | 228 (100.0) | 228 (100.0) |
| | | Male, n (%) | 0 (0.0) | 0 (0.0) |
| | | Age, median (Q1;Q3) | 51.1 (47.4;54.8) | 51.0 (48.0;54.3) |
| | | CCI, n (%) | | |
| | | 0 | 183 (80.3) | 201 (88.2) |
| | | 1-2 | * | * |
| | | +3 | * | * |
| | | Education level, n (%) | | |
| | | No/Unknown education | * | * |
| | | Primary | * | * |
| | | Secondary | 103 (45.2) | 103 (45.2) |
| | | Tertiary | 70 (30.7) | 70 (30.7) |
| | | Relation to patient, n (%) | | |
| | | Mother | 228 (100.0) | 228 (100.0) |
| Young adults and adults (25-64 years) | Pooled | Number of persons, n | 10,917 (100.0) | 10,917 (100.0) |
| | | Male, n (%) | 6,606 (60.5) | 6,606 (60.5) |
| | | Age, median (Q1;Q3) | 51.6 (38.7;60.6) | 51.6 (38.7;60.6) |
| | | CCI, n (%) | | |
| | | 0 | 9,093 (83.3) | 9,155 (83.9) |
| | | 1-2 | 1,510 (13.8) | 1,496 (13.7) |
| | | +3 | 314 (2.9) | 266 (2.4) |
| | | Education level, n (%) | | |
| | | No/Unknown education | 460 (4.2) | 460 (4.2) |
| | | Primary | 2,441 (22.4) | 2,441 (22.4) |
| | | Secondary | 5,280 (48.4) | 5,280 (48.4) |
| | | Tertiary | 2,736 (25.1) | 2,736 (25.1) |
| | | Relation to patient, n (%) | | |
| | | Mother | 1,191 (10.9) | 1,191 (10.9) |
| | | Father | 288 (2.6) | 288 (2.6) |
| | | Partner | 7,298 (66.8) | 7,298 (66.8) |
| | | Child | 1,555 (14.2) | 1,555 (14.2) |

**Supplementary Table 182**
**Multiple sclerosis - Prevalent cohort 2021**
**Patient characteristics - relatives - pooled**

| *Age strata of index patients* | *Closest relative type* | *Characteristic of family relatives by agestrata of index patients* | *Closest family relatives of brain disorder patients* | *Comparison relative* |
|---|---|---|---|---|
| | | Sibling | 585 (5.4) | 585 (5.4) |
| Older people (65+ years) | Pooled | Number of persons, n | 3,279 (100.0) | 3,279 (100.0) |
| | | Male, n (%) | 1,929 (58.8) | 1,929 (58.8) |
| | | Age, median (Q1;Q3) | 67.4 (53.3;73.8) | 67.5 (52.9;73.4) |
| | | CCI, n (%) | | |
| | | 0 | 2,294 (70.0) | 2,366 (72.2) |
| | | 1-2 | 745 (22.7) | 712 (21.7) |
| | | +3 | 240 (7.3) | 201 (6.1) |
| | | Education level, n (%) | | |
| | | No/Unknown education | 121 (3.7) | 121 (3.7) |
| | | Primary | 627 (19.1) | 627 (19.1) |
| | | Secondary | 1,593 (48.6) | 1,593 (48.6) |
| | | Tertiary | 938 (28.6) | 938 (28.6) |
| | | Relation to patient, n (%) | | |
| | | Partner | 2,097 (64.0) | 2,097 (64.0) |
| | | Child | 1,116 (34.0) | 1,116 (34.0) |
| | | Sibling | 66 (2.0) | 66 (2.0) |

**Supplementary Table 183**
**Multiple sclerosis - Prevalent cohort 2021**
**Patient characteristics - relatives - not pooled**

| *Age strata of index patients* | *Closest relative type* | *Characteristic of family relatives by agestrata of index patients* | *Closest family relatives of brain disorder patients* | *Comparison relative* |
|---|---|---|---|---|
| Children and young people (0-24 years) | Fathers | Number of persons, n | 217 (100.0) | 217 (100.0) |
| | | Male, n (%) | 217 (100.0) | 217 (100.0) |
| | | Age, median (Q1;Q3) | 53.9 (50.3;57.7) | 54.1 (50.4;56.9) |
| | | CCI, n (%) | | |
| | | 0 | 183 (84.3) | 186 (85.7) |
| | | 1-2 | 29 (13.4) | 25 (11.5) |
| | | +3 | 5 (2.3) | 6 (2.8) |
| | | Education level, n (%) | | |
| | | No/Unknown education | * | * |
| | | Primary | 58 (26.7) | 58 (26.7) |
| | | Secondary | 123 (56.7) | 123 (56.7) |
| | | Tertiary | * | * |
| | | Relation to patient, n (%) | | |
| | | Father | 217 (100.0) | 217 (100.0) |
| Children and young people (0-24 years) | Mothers | Number of persons, n | 228 (100.0) | 228 (100.0) |
| | | Male, n (%) | 0 (0.0) | 0 (0.0) |
| | | Age, median (Q1;Q3) | 51.1 (47.4;54.8) | 51.0 (48.0;54.3) |
| | | CCI, n (%) | | |
| | | 0 | 183 (80.3) | 201 (88.2) |
| | | 1-2 | * | * |
| | | +3 | * | * |
| | | Education level, n (%) | | |
| | | No/Unknown education | * | * |
| | | Primary | * | * |
| | | Secondary | 103 (45.2) | 103 (45.2) |
| | | Tertiary | 70 (30.7) | 70 (30.7) |
| | | Relation to patient, n (%) | | |
| | | Mother | 228 (100.0) | 228 (100.0) |
| Young adults and adults (25-64 years) | Children | Number of persons, n | 1,555 (100.0) | 1,555 (100.0) |
| | | Male, n (%) | 802 (51.6) | 802 (51.6) |
| | | Age, median (Q1;Q3) | 22.3 (15.4;30.4) | 22.0 (15.4;30.6) |
| | | CCI, n (%) | | |
| | | 0 | 1,465 (94.2) | 1,478 (95.0) |
| | | 1-2 | 83 (5.3) | * |
| | | +3 | 7 (0.5) | * |
| | | Education level, n (%) | | |
| | | No/Unknown education | 285 (18.3) | 285 (18.3) |
| | | Primary | 628 (40.4) | 628 (40.4) |
| | | Secondary | 412 (26.5) | 412 (26.5) |
| | | Tertiary | 230 (14.8) | 230 (14.8) |
| | | Relation to patient, n (%) | | |
| | | Child | 1,555 (100.0) | 1,555 (100.0) |
| Young adults and adults (25-64 years) | Parents | Number of persons, n | 1,479 (100.0) | 1,479 (100.0) |
| | | Male, n (%) | 288 (19.5) | 288 (19.5) |
| | | Age, median (Q1;Q3) | 71.8 (63.9;78.2) | 71.6 (63.8;77.8) |

### Supplementary Table 183
### Multiple sclerosis - Prevalent cohort 2021
### Patient characteristics - relatives - not pooled

| *Age strata of index patients* | *Closest relative type* | *Characteristic of family relatives by agestrata of index patients* | *Closest family relatives of brain disorder patients* | *Comparison relative* |
|---|---|---|---|---|
| | | CCI, n (%) | | |
| | | 0 | 919 (62.1) | 923 (62.4) |
| | | 1-2 | 422 (28.5) | 442 (29.9) |
| | | +3 | 138 (9.3) | 114 (7.7) |
| | | Education level, n (%) | | |
| | | No/Unknown education | 31 (2.1) | 31 (2.1) |
| | | Primary | 488 (33.0) | 488 (33.0) |
| | | Secondary | 714 (48.3) | 714 (48.3) |
| | | Tertiary | 246 (16.6) | 246 (16.6) |
| | | Relation to patient, n (%) | | |
| | | Mother | 1,191 (80.5) | 1,191 (80.5) |
| | | Father | 288 (19.5) | 288 (19.5) |
| Young adults and adults (25-64 years) | Partners | Number of persons, n | 7,298 (100.0) | 7,298 (100.0) |
| | | Male, n (%) | 5,213 (71.4) | 5,213 (71.4) |
| | | Age, median (Q1;Q3) | 51.4 (42.8;58.6) | 51.3 (42.7;58.4) |
| | | CCI, n (%) | | |
| | | 0 | 6,242 (85.5) | 6,282 (86.1) |
| | | 1-2 | 904 (12.4) | 884 (12.1) |
| | | +3 | 152 (2.1) | 132 (1.8) |
| | | Education level, n (%) | | |
| | | No/Unknown education | 144 (2.0) | 144 (2.0) |
| | | Primary | 1,214 (16.6) | 1,214 (16.6) |
| | | Secondary | 3,813 (52.2) | 3,813 (52.2) |
| | | Tertiary | 2,127 (29.1) | 2,127 (29.1) |
| | | Relation to patient, n (%) | | |
| | | Partner | 7,298 (100.0) | 7,298 (100.0) |
| Young adults and adults (25-64 years) | Siblings | Number of persons, n | 585 (100.0) | 585 (100.0) |
| | | Male, n (%) | 303 (51.8) | 303 (51.8) |
| | | Age, median (Q1;Q3) | 56.2 (49.8;61.4) | 57.0 (49.6;61.7) |
| | | CCI, n (%) | | |
| | | 0 | 467 (79.8) | 472 (80.7) |
| | | 1-2 | 101 (17.3) | 96 (16.4) |
| | | +3 | 17 (2.9) | 17 (2.9) |
| | | Education level, n (%) | | |
| | | No/Unknown education | 0 (0.0) | 0 (0.0) |
| | | Primary | 111 (19.0) | 111 (19.0) |
| | | Secondary | 341 (58.3) | 341 (58.3) |
| | | Tertiary | 133 (22.7) | 133 (22.7) |
| | | Relation to patient, n (%) | | |
| | | Sibling | 585 (100.0) | 585 (100.0) |
| Older people (65+ years) | Children | Number of persons, n | 1,116 (100.0) | 1,116 (100.0) |
| | | Male, n (%) | 569 (51.0) | 569 (51.0) |
| | | Age, median (Q1;Q3) | 48.6 (43.4;53.8) | 48.1 (43.3;53.5) |
| | | CCI, n (%) | | |
| | | 0 | 956 (85.7) | 992 (88.9) |

### Supplementary Table 183
### Multiple sclerosis - Prevalent cohort 2021
### Patient characteristics - relatives - not pooled

| *Age strata of index patients* | *Closest relative type* | *Characteristic of family relatives by agestrata of index patients* | *Closest family relatives of brain disorder patients* | *Comparison relative* |
|---|---|---|---|---|
| | | 1-2 | 138 (12.4) | 117 (10.5) |
| | | +3 | 22 (2.0) | 7 (0.6) |
| | | Education level, n (%) | | |
| | | No/Unknown education | * | * |
| | | Primary | * | * |
| | | Secondary | 516 (46.2) | 516 (46.2) |
| | | Tertiary | 439 (39.3) | 439 (39.3) |
| | | Relation to patient, n (%) | | |
| | | Child | 1,116 (100.0) | 1,116 (100.0) |
| Older people (65+ years) | Partners | Number of persons, n | 2,097 (100.0) | 2,097 (100.0) |
| | | Male, n (%) | 1,326 (63.2) | 1,326 (63.2) |
| | | Age, median (Q1;Q3) | 71.9 (67.8;76.0) | 71.7 (67.9;75.6) |
| | | CCI, n (%) | | |
| | | 0 | 1,291 (61.6) | 1,322 (63.0) |
| | | 1-2 | 591 (28.2) | 584 (27.8) |
| | | +3 | 215 (10.3) | 191 (9.1) |
| | | Education level, n (%) | | |
| | | No/Unknown education | 119 (5.7) | 119 (5.7) |
| | | Primary | 456 (21.7) | 456 (21.7) |
| | | Secondary | 1,043 (49.7) | 1,043 (49.7) |
| | | Tertiary | 479 (22.8) | 479 (22.8) |
| | | Relation to patient, n (%) | | |
| | | Partner | 2,097 (100.0) | 2,097 (100.0) |
| Older people (65+ years) | Siblings | Number of persons, n | 66 (100.0) | 66 (100.0) |
| | | Male, n (%) | 34 (51.5) | 34 (51.5) |
| | | Age, median (Q1;Q3) | 62.8 (60.8;65.8) | 62.8 (60.1;65.8) |
| | | CCI, n (%) | | |
| | | 0 | 47 (71.2) | 52 (78.8) |
| | | 1-2 | * | * |
| | | +3 | * | * |
| | | Education level, n (%) | | |
| | | No/Unknown education | 0 (0.0) | 0 (0.0) |
| | | Primary | 12 (18.2) | 12 (18.2) |
| | | Secondary | 34 (51.5) | 34 (51.5) |
| | | Tertiary | 20 (30.3) | 20 (30.3) |
| | | Relation to patient, n (%) | | |
| | | Sibling | 66 (100.0) | 66 (100.0) |

#### Supplementary Table 184
#### Multiple sclerosis - Prevalent cohort 2021
#### Cost analysis - pooled

| | | | | *Actual costs* | | *Attributable costs* | |
|---|---|---|---|---|---|---|---|
| *Age strata of index patients* | *Closest relative type* | *Number of index patients included in cost component calculations* | *Cost component* | *Total (million EUR)* | *Per person (EUR)* | *Total (million EUR)* | *Per person (EUR)* |
| Children and young people (0-24 years) | Fathers | 217 | Primary sector | 0.1 | 252 | 0.0 | 26 |
| | | 217 | Hospital care | 0.3 | 1,555 | 0.0 | 159 |
| | | 217 | Filled prescriptions | 0.1 | 357 | 0.0 | 164 |
| | | 203 | Lost productivity (income loss) | . | . | 3.7 | 18,114 |
| Children and young people (0-24 years) | Mothers | 228 | Primary sector | 0.1 | 408 | 0.0 | 95 |
| | | 228 | Hospital care | 0.6 | 2,461 | 0.4 | 1,600 |
| | | 228 | Filled prescriptions | 0.1 | 263 | 0.0 | 5 |
| | | 227 | Lost productivity (income loss) | . | . | 2.4 | 10,452 |
| Young adults and adults (25-64 years) | Pooled | 10,917 | Primary sector | 3.3 | 306 | 0.1 | 10 |
| | | 10,917 | Hospital care | 21.7 | 1,994 | 0.9 | 84 |
| | | 10,917 | Filled prescriptions | 3.0 | 278 | 0.4 | 36 |
| | | 8,145 | Lost productivity (income loss) | . | . | 9.3 | 1,145 |
| Older people (65+ years) | Pooled | 3,279 | Primary sector | 1.3 | 413 | 0.0 | 10 |
| | | 3,279 | Hospital care | 10.6 | 3,264 | 0.2 | 47 |
| | | 3,279 | Filled prescriptions | 1.4 | 420 | 0.0 | 4 |
| | | 1,217 | Lost productivity (income loss) | . | . | 0.3 | 259 |

### Supplementary Table 185
### Multiple sclerosis - Prevalent cohort 2021
### Cost analysis - not pooled

| Age strata of index patients | Closest relative type | Number of index patients included in cost component calculations | Cost component | Actual costs: Total (million EUR) | Actual costs: Per person (EUR) | Attributable costs: Total (million EUR) | Attributable costs: Per person (EUR) |
|---|---|---|---|---|---|---|---|
| Children and young people (0-24 years) | Fathers | 217 | Primary sector | 0.1 | 252 | 0.0 | 26 |
| | | 217 | Hospital care | 0.3 | 1,555 | 0.0 | 159 |
| | | 217 | Filled prescriptions | 0.1 | 357 | 0.0 | 164 |
| | | 203 | Lost productivity (income loss) | . | . | 3.7 | 18,114 |
| Children and young people (0-24 years) | Mothers | 228 | Primary sector | 0.1 | 408 | 0.0 | 95 |
| | | 228 | Hospital care | 0.6 | 2,461 | 0.4 | 1,600 |
| | | 228 | Filled prescriptions | 0.1 | 263 | 0.0 | 5 |
| | | 227 | Lost productivity (income loss) | . | . | 2.4 | 10,452 |
| Young adults and adults (25-64 years) | Children | 1,555 | Primary sector | 0.4 | 231 | 0.0 | 24 |
| | | 1,555 | Hospital care | 1.7 | 1,078 | 0.1 | 73 |
| | | 1,555 | Filled prescriptions | 0.2 | 132 | 0.1 | 39 |
| | | 943 | Lost productivity (income loss) | . | . | -1.1 | -1,128 |
| Young adults and adults (25-64 years) | Parents | 1,479 | Primary sector | 0.7 | 490 | -0.0 | -22 |
| | | 1,479 | Hospital care | 5.5 | 3,769 | -0.6 | -433 |
| | | 1,479 | Filled prescriptions | 0.8 | 579 | 0.1 | 44 |
| | | 319 | Lost productivity (income loss) | . | . | 0.5 | 1,480 |
| Young adults and adults (25-64 years) | Partners | 7,298 | Primary sector | 2.1 | 282 | 0.1 | 13 |
| | | 7,298 | Hospital care | 12.4 | 1,700 | 0.3 | 42 |
| | | 7,298 | Filled prescriptions | 1.8 | 247 | 0.3 | 36 |
| | | 6,414 | Lost productivity (income loss) | . | . | 10.5 | 1,641 |
| Young adults and adults (25-64 years) | Siblings | 585 | Primary sector | 0.2 | 345 | 0.0 | 25 |
| | | 585 | Hospital care | 2.1 | 3,667 | 1.1 | 1,920 |
| | | 585 | Filled prescriptions | 0.2 | 298 | 0.0 | 6 |
| | | 469 | Lost productivity (income loss) | . | . | -0.6 | -1,293 |
| Older people (65+ years) | Children | 1,116 | Primary sector | 0.3 | 292 | 0.0 | 12 |
| | | 1,116 | Hospital care | 1.8 | 1,658 | 0.1 | 133 |
| | | 1,116 | Filled prescriptions | 0.2 | 190 | -0.0 | -44 |
| | | 1,082 | Lost productivity (income loss) | . | . | 0.8 | 735 |
| Older people (65+ years) | Partners | 2,097 | Primary sector | 1.0 | 480 | 0.0 | 12 |
| | | 2,097 | Hospital care | 8.4 | 4,081 | -0.2 | -75 |
| | | 2,097 | Filled prescriptions | 1.1 | 544 | 0.1 | 32 |
| | | 103 | Lost productivity (income loss) | . | . | -0.9 | -8,456 |
| Older people (65+ years) | Siblings | 66 | Primary sector | 0.0 | 351 | -0.0 | -84 |
| | | 66 | Hospital care | 0.3 | 4,735 | 0.2 | 2,430 |
| | | 66 | Filled prescriptions | 0.0 | 400 | -0.0 | -51 |
| | | 32 | Lost productivity (income loss) | . | . | 0.4 | 12,224 |

**Supplementary Table 186**
**Neuromuscular disorders - Incident cohort 2016-2021**
**Identification of populations of relatives**

| ***Agegroup of index patients in matched population*** | | ***Closest relative type identified*** | | ***Available control with same type of closest relative*** |
|---|---|---|---|---|
| Children and young people (0-24 years) | n = 518 | Fathers | n = 384 (74%) | 94% |
| | | Mothers | n = 406 (78%) | 94% |
| | | No available closest relative | n = 58 (11%) | n/a |
| Young adults and adults (25-64 years) | n = 1,544 | Children | n = 197 (13%) | 92% |
| | | Parents | n = 198 (13%) | 90% |
| | | Partners | n = 942 (61%) | 96% |
| | | Siblings | n = 160 (10%) | 54% |
| | | Pooled | n = 1,497 (97%) | 90% |
| | | No available closest relative | n = 47 (3%) | n/a |
| Older people (65+ years) | n = 1,106 | Children | n = 346 (31%) | 96% |
| | | Partners | n = 641 (58%) | 97% |
| | | Siblings | n = 16 (1%) | 31% |
| | | Pooled | n = 1,003 (91%) | 96% |
| | | No available closest relative | n = 103 (9%) | n/a |

**Supplementary Table 187**
**Neuromuscular disorders - Incident cohort 2016-2021**
**Patient characteristics - cases**

| *Age strata of index patients* | *Characteristic* | |
|---|---|---|
| Children and young people (0-24 years) | Number of persons, n | 438 (100.0) |
| | Male, n (%) | 254 (58.0) |
| | Age, median (Q1;Q3) | 13.1 (6.1;19.0) |
| | CCI, n (%) | |
| | 0 | 365 (83.3) |
| | 1-2 | 66 (15.1) |
| | +3 | 7 (1.6) |
| | Education level, n (%) | |
| | No/Unknown education | 218 (49.8) |
| | Primary | 153 (34.9) |
| | Secondary | 56 (12.8) |
| | Tertiary | 11 (2.5) |
| Young adults and adults (25-64 years) | Number of persons, n | 1,355 (100.0) |
| | Male, n (%) | 641 (47.3) |
| | Age, median (Q1;Q3) | 50.0 (39.4;57.5) |
| | CCI, n (%) | |
| | 0 | 918 (67.7) |
| | 1-2 | 321 (23.7) |
| | +3 | 116 (8.6) |
| | Education level, n (%) | |
| | No/Unknown education | 44 (3.2) |
| | Primary | 297 (21.9) |
| | Secondary | 601 (44.4) |
| | Tertiary | 413 (30.5) |
| Older people (65+ years) | Number of persons, n | 964 (100.0) |
| | Male, n (%) | 504 (52.3) |
| | Age, median (Q1;Q3) | 74.0 (70.1;78.9) |
| | CCI, n (%) | |
| | 0 | 443 (46.0) |
| | 1-2 | 354 (36.7) |
| | +3 | 167 (17.3) |
| | Education level, n (%) | |
| | No/Unknown education | 70 (7.3) |
| | Primary | 309 (32.1) |
| | Secondary | 442 (45.9) |
| | Tertiary | 143 (14.8) |

### Supplementary Table 188
### Neuromuscular disorders - Incident cohort 2016-2021
### Patient characteristics - relatives - pooled

| *Age strata of index patients* | *Closest relative type* | *Characteristic of family relatives by agestrata of index patients* | *Closest family relatives of brain disorder patients* | *Comparison relative* |
|---|---|---|---|---|
| Children and young people (0-24 years) | Fathers | Number of persons, n | 363 (100.0) | 363 (100.0) |
| | | Male, n (%) | 363 (100.0) | 363 (100.0) |
| | | Age, median (Q1;Q3) | 45.7 (39.1;51.6) | 45.8 (39.2;51.7) |
| | | CCI, n (%) | | |
| | | 0 | 329 (90.6) | 332 (91.5) |
| | | 1-2 | * | * |
| | | +3 | * | * |
| | | Education level, n (%) | | |
| | | No/Unknown education | * | * |
| | | Primary | * | * |
| | | Secondary | 173 (47.7) | 173 (47.7) |
| | | Tertiary | 110 (30.3) | 110 (30.3) |
| | | Relation to patient, n (%) | | |
| | | Father | 363 (100.0) | 363 (100.0) |
| Children and young people (0-24 years) | Mothers | Number of persons, n | 385 (100.0) | 385 (100.0) |
| | | Male, n (%) | 0 (0.0) | 0 (0.0) |
| | | Age, median (Q1;Q3) | 42.7 (36.2;49.1) | 43.3 (36.2;49.0) |
| | | CCI, n (%) | | |
| | | 0 | 340 (88.3) | 348 (90.4) |
| | | 1-2 | 38 (9.9) | * |
| | | +3 | 7 (1.8) | * |
| | | Education level, n (%) | | |
| | | No/Unknown education | 8 (2.1) | 8 (2.1) |
| | | Primary | 65 (16.9) | 65 (16.9) |
| | | Secondary | 153 (39.7) | 153 (39.7) |
| | | Tertiary | 159 (41.3) | 159 (41.3) |
| | | Relation to patient, n (%) | | |
| | | Mother | 385 (100.0) | 385 (100.0) |
| Young adults and adults (25-64 years) | Pooled | Number of persons, n | 1,355 (100.0) | 1,355 (100.0) |
| | | Male, n (%) | 681 (50.3) | 681 (50.3) |
| | | Age, median (Q1;Q3) | 50.5 (36.7;59.0) | 50.5 (36.8;58.9) |
| | | CCI, n (%) | | |
| | | 0 | 1,129 (83.3) | 1,146 (84.6) |
| | | 1-2 | 193 (14.2) | 176 (13.0) |
| | | +3 | 33 (2.4) | 33 (2.4) |
| | | Education level, n (%) | | |
| | | No/Unknown education | 55 (4.1) | 55 (4.1) |
| | | Primary | 302 (22.3) | 302 (22.3) |
| | | Secondary | 642 (47.4) | 642 (47.4) |
| | | Tertiary | 356 (26.3) | 356 (26.3) |
| | | Relation to patient, n (%) | | |
| | | Mother | 150 (11.1) | 150 (11.1) |
| | | Father | 29 (2.1) | 29 (2.1) |
| | | Partner | 907 (66.9) | 907 (66.9) |
| | | Child | 182 (13.4) | 182 (13.4) |

**Supplementary Table 188**
**Neuromuscular disorders - Incident cohort 2016-2021**
**Patient characteristics - relatives - pooled**

| *Age strata of index patients* | *Closest relative type* | *Characteristic of family relatives by agestrata of index patients* | *Closest family relatives of brain disorder patients* | *Comparison relative* |
|---|---|---|---|---|
| | | Sibling | 87 (6.4) | 87 (6.4) |
| Older people (65+ years) | Pooled | Number of persons, n | 964 (100.0) | 964 (100.0) |
| | | Male, n (%) | 415 (43.0) | 415 (43.0) |
| | | Age, median (Q1;Q3) | 67.8 (56.0;74.3) | 67.6 (55.4;73.9) |
| | | CCI, n (%) | | |
| | | 0 | 681 (70.6) | 704 (73.0) |
| | | 1-2 | 217 (22.5) | 192 (19.9) |
| | | +3 | 66 (6.8) | 68 (7.1) |
| | | Education level, n (%) | | |
| | | No/Unknown education | 28 (2.9) | 28 (2.9) |
| | | Primary | 197 (20.4) | 197 (20.4) |
| | | Secondary | 505 (52.4) | 505 (52.4) |
| | | Tertiary | 234 (24.3) | 234 (24.3) |
| | | Relation to patient, n (%) | | |
| | | Partner | 624 (64.7) | 624 (64.7) |
| | | Child | 335 (34.8) | 335 (34.8) |
| | | Sibling | 5 (0.5) | 5 (0.5) |

#### Supplementary Table 189
#### Neuromuscular disorders - Incident cohort 2016-2021
#### Patient characteristics - relatives - not pooled

| *Age strata of index patients* | *Closest relative type* | *Characteristic of family relatives by agestrata of index patients* | *Closest family relatives of brain disorder patients* | *Comparison relative* |
|---|---|---|---|---|
| Children and young people (0-24 years) | Fathers | Number of persons, n | 363 (100.0) | 363 (100.0) |
| | | Male, n (%) | 363 (100.0) | 363 (100.0) |
| | | Age, median (Q1;Q3) | 45.7 (39.1;51.6) | 45.8 (39.2;51.7) |
| | | CCI, n (%) | | |
| | | 0 | 329 (90.6) | 332 (91.5) |
| | | 1-2 | * | * |
| | | +3 | * | * |
| | | Education level, n (%) | | |
| | | No/Unknown education | * | * |
| | | Primary | * | * |
| | | Secondary | 173 (47.7) | 173 (47.7) |
| | | Tertiary | 110 (30.3) | 110 (30.3) |
| | | Relation to patient, n (%) | | |
| | | Father | 363 (100.0) | 363 (100.0) |
| Children and young people (0-24 years) | Mothers | Number of persons, n | 385 (100.0) | 385 (100.0) |
| | | Male, n (%) | 0 (0.0) | 0 (0.0) |
| | | Age, median (Q1;Q3) | 42.7 (36.2;49.1) | 43.3 (36.2;49.0) |
| | | CCI, n (%) | | |
| | | 0 | 340 (88.3) | 348 (90.4) |
| | | 1-2 | 38 (9.9) | * |
| | | +3 | 7 (1.8) | * |
| | | Education level, n (%) | | |
| | | No/Unknown education | 8 (2.1) | 8 (2.1) |
| | | Primary | 65 (16.9) | 65 (16.9) |
| | | Secondary | 153 (39.7) | 153 (39.7) |
| | | Tertiary | 159 (41.3) | 159 (41.3) |
| | | Relation to patient, n (%) | | |
| | | Mother | 385 (100.0) | 385 (100.0) |
| Young adults and adults (25-64 years) | Children | Number of persons, n | 182 (100.0) | 182 (100.0) |
| | | Male, n (%) | 103 (56.6) | 103 (56.6) |
| | | Age, median (Q1;Q3) | 24.9 (16.5;32.5) | 24.6 (16.6;31.8) |
| | | CCI, n (%) | | |
| | | 0 | 168 (92.3) | 171 (94.0) |
| | | 1-2 | * | 11 (6.0) |
| | | +3 | * | 0 (0.0) |
| | | Education level, n (%) | | |
| | | No/Unknown education | 34 (18.7) | 34 (18.7) |
| | | Primary | 68 (37.4) | 68 (37.4) |
| | | Secondary | 56 (30.8) | 56 (30.8) |
| | | Tertiary | 24 (13.2) | 24 (13.2) |
| | | Relation to patient, n (%) | | |
| | | Child | 182 (100.0) | 182 (100.0) |
| Young adults and adults (25-64 years) | Parents | Number of persons, n | 179 (100.0) | 179 (100.0) |
| | | Male, n (%) | 29 (16.2) | 29 (16.2) |
| | | Age, median (Q1;Q3) | 64.8 (57.2;74.6) | 65.6 (58.2;75.3) |

**Supplementary Table 189**
**Neuromuscular disorders - Incident cohort 2016-2021**
**Patient characteristics - relatives - not pooled**

| *Age strata of index patients* | *Closest relative type* | *Characteristic of family relatives by agestrata of index patients* | *Closest family relatives of brain disorder patients* | *Comparison relative* |
|---|---|---|---|---|
| | | CCI, n (%) | | |
| | | 0 | 127 (70.9) | 128 (71.5) |
| | | 1-2 | 40 (22.3) | 35 (19.6) |
| | | +3 | 12 (6.7) | 16 (8.9) |
| | | Education level, n (%) | | |
| | | No/Unknown education | * | * |
| | | Primary | 60 (33.5) | 60 (33.5) |
| | | Secondary | 79 (44.1) | 79 (44.1) |
| | | Tertiary | * | * |
| | | Relation to patient, n (%) | | |
| | | Mother | 150 (83.8) | 150 (83.8) |
| | | Father | 29 (16.2) | 29 (16.2) |
| Young adults and adults (25-64 years) | Partners | Number of persons, n | 907 (100.0) | 907 (100.0) |
| | | Male, n (%) | 485 (53.5) | 485 (53.5) |
| | | Age, median (Q1;Q3) | 50.3 (41.3;57.5) | 50.3 (40.9;57.7) |
| | | CCI, n (%) | | |
| | | 0 | 764 (84.2) | 774 (85.3) |
| | | 1-2 | 126 (13.9) | 121 (13.3) |
| | | +3 | 17 (1.9) | 12 (1.3) |
| | | Education level, n (%) | | |
| | | No/Unknown education | 19 (2.1) | 19 (2.1) |
| | | Primary | 152 (16.8) | 152 (16.8) |
| | | Secondary | 455 (50.2) | 455 (50.2) |
| | | Tertiary | 281 (31.0) | 281 (31.0) |
| | | Relation to patient, n (%) | | |
| | | Partner | 907 (100.0) | 907 (100.0) |
| Young adults and adults (25-64 years) | Siblings | Number of persons, n | 87 (100.0) | 87 (100.0) |
| | | Male, n (%) | 64 (73.6) | 64 (73.6) |
| | | Age, median (Q1;Q3) | 55.0 (45.8;58.6) | 54.3 (45.4;59.4) |
| | | CCI, n (%) | | |
| | | 0 | 70 (80.5) | 73 (83.9) |
| | | 1-2 | * | 9 (10.3) |
| | | +3 | * | 5 (5.7) |
| | | Education level, n (%) | | |
| | | No/Unknown education | 0 (0.0) | 0 (0.0) |
| | | Primary | 22 (25.3) | 22 (25.3) |
| | | Secondary | 52 (59.8) | 52 (59.8) |
| | | Tertiary | 13 (14.9) | 13 (14.9) |
| | | Relation to patient, n (%) | | |
| | | Sibling | 87 (100.0) | 87 (100.0) |
| Older people (65+ years) | Children | Number of persons, n | 335 (100.0) | 335 (100.0) |
| | | Male, n (%) | 179 (53.4) | 179 (53.4) |
| | | Age, median (Q1;Q3) | 51.6 (45.9;57.1) | 51.5 (45.5;56.7) |
| | | CCI, n (%) | | |
| | | 0 | 281 (83.9) | 291 (86.9) |

#### Supplementary Table 189
#### Neuromuscular disorders - Incident cohort 2016-2021
#### Patient characteristics - relatives - not pooled

| *Age strata of index patients* | *Closest relative type* | *Characteristic of family relatives by agestrata of index patients* | *Closest family relatives of brain disorder patients* | *Comparison relative* |
|---|---|---|---|---|
| | | 1-2 | 48 (14.3) | 37 (11.0) |
| | | +3 | 6 (1.8) | 7 (2.1) |
| | | Education level, n (%) | | |
| | | No/Unknown education | 0 (0.0) | 0 (0.0) |
| | | Primary | 33 (9.9) | 33 (9.9) |
| | | Secondary | 192 (57.3) | 192 (57.3) |
| | | Tertiary | 110 (32.8) | 110 (32.8) |
| | | Relation to patient, n (%) | | |
| | | Child | 335 (100.0) | 335 (100.0) |
| Older people (65+ years) | Partners | Number of persons, n | 624 (100.0) | 624 (100.0) |
| | | Male, n (%) | 233 (37.3) | 233 (37.3) |
| | | Age, median (Q1;Q3) | 72.3 (68.0;76.7) | 72.2 (68.2;76.3) |
| | | CCI, n (%) | | |
| | | 0 | 398 (63.8) | 409 (65.5) |
| | | 1-2 | 167 (26.8) | 154 (24.7) |
| | | +3 | 59 (9.5) | 61 (9.8) |
| | | Education level, n (%) | | |
| | | No/Unknown education | 28 (4.5) | 28 (4.5) |
| | | Primary | 164 (26.3) | 164 (26.3) |
| | | Secondary | 309 (49.5) | 309 (49.5) |
| | | Tertiary | 123 (19.7) | 123 (19.7) |
| | | Relation to patient, n (%) | | |
| | | Partner | 624 (100.0) | 624 (100.0) |
| Older people (65+ years) | Siblings | Number of persons, n | 5 (100.0) | 5 (100.0) |
| | | Male, n (%) | * | * |
| | | Age, median (Q1;Q3) | 55.8 (55.2;60.6) | 57.4 (53.2;64.5) |
| | | CCI, n (%) | | |
| | | 0 | * | * |
| | | 1-2 | * | * |
| | | +3 | * | 0 (0.0) |
| | | Education level, n (%) | | |
| | | No/Unknown education | 0 (0.0) | 0 (0.0) |
| | | Primary | 0 (0.0) | 0 (0.0) |
| | | Secondary | * | * |
| | | Tertiary | * | * |
| | | Relation to patient, n (%) | | |
| | | Sibling | 5 (100.0) | 5 (100.0) |

#### Supplementary Table 190
#### Neuromuscular disorders - Incident cohort 2016-2021
#### Cost analysis - pooled

| *Age strata of index patients* | *Closest relative type* | *Number of index patients included in cost component calculations* | *Cost component* | *Actual costs* | | *Attributable costs* | |
|---|---|---|---|---|---|---|---|
| | | | | *Total (million EUR)* | *Per person (EUR)* | *Total (million EUR)* | *Per person (EUR)* |
| Children and young people (0-24 years) | Fathers | 363 | Primary sector | 0.1 | 227 | 0.0 | 35 |
| | | 363 | Hospital care | 0.5 | 1,324 | -0.2 | -640 |
| | | 363 | Filled prescriptions | 0.1 | 179 | 0.0 | 60 |
| | | 356 | Lost productivity (income loss) | . | . | 2.5 | 7,001 |
| Children and young people (0-24 years) | Mothers | 385 | Primary sector | 0.2 | 391 | 0.0 | 77 |
| | | 385 | Hospital care | 0.9 | 2,243 | 0.4 | 1,119 |
| | | 385 | Filled prescriptions | 0.1 | 209 | 0.0 | 47 |
| | | 384 | Lost productivity (income loss) | . | . | 1.4 | 3,712 |
| Young adults and adults (25-64 years) | Pooled | 1,355 | Primary sector | 0.4 | 324 | 0.1 | 38 |
| | | 1,355 | Hospital care | 2.6 | 1,941 | 0.2 | 140 |
| | | 1,355 | Filled prescriptions | 0.3 | 250 | 0.0 | 29 |
| | | 1,077 | Lost productivity (income loss) | . | . | -2.3 | -2,110 |
| Older people (65+ years) | Pooled | 964 | Primary sector | 0.4 | 469 | 0.1 | 64 |
| | | 964 | Hospital care | 3.6 | 3,745 | 0.5 | 530 |
| | | 964 | Filled prescriptions | 0.4 | 448 | 0.0 | 52 |
| | | 359 | Lost productivity (income loss) | . | . | 2.0 | 5,442 |

#### Supplementary Table 191
#### Neuromuscular disorders - Incident cohort 2016-2021
#### Cost analysis - not pooled

| *Age strata of index patients* | *Closest relative type* | *Number of index patients included in cost component calculations* | *Cost component* | *Actual costs* | | *Attributable costs* | |
|---|---|---|---|---|---|---|---|
| | | | | *Total (million EUR)* | *Per person (EUR)* | *Total (million EUR)* | *Per person (EUR)* |
| Children and young people (0-24 years) | Fathers | 363 | Primary sector | 0.1 | 227 | 0.0 | 35 |
| | | 363 | Hospital care | 0.5 | 1,324 | -0.2 | -640 |
| | | 363 | Filled prescriptions | 0.1 | 179 | 0.0 | 60 |
| | | 356 | Lost productivity (income loss) | . | . | 2.5 | 7,001 |
| Children and young people (0-24 years) | Mothers | 385 | Primary sector | 0.2 | 391 | 0.0 | 77 |
| | | 385 | Hospital care | 0.9 | 2,243 | 0.4 | 1,119 |
| | | 385 | Filled prescriptions | 0.1 | 209 | 0.0 | 47 |
| | | 384 | Lost productivity (income loss) | . | . | 1.4 | 3,712 |
| Young adults and adults (25-64 years) | Children | 182 | Primary sector | 0.0 | 256 | 0.0 | 68 |
| | | 182 | Hospital care | 0.3 | 1,704 | 0.1 | 410 |
| | | 182 | Filled prescriptions | 0.0 | 89 | -0.0 | -11 |
| | | 125 | Lost productivity (income loss) | . | . | 0.6 | 5,184 |
| Young adults and adults (25-64 years) | Parents | 179 | Primary sector | 0.1 | 457 | 0.0 | 6 |
| | | 179 | Hospital care | 0.7 | 3,828 | 0.3 | 1,512 |
| | | 179 | Filled prescriptions | 0.1 | 456 | 0.0 | 31 |
| | | 75 | Lost productivity (income loss) | . | . | 2.4 | 31,889 |
| Young adults and adults (25-64 years) | Partners | 907 | Primary sector | 0.3 | 313 | 0.0 | 35 |
| | | 907 | Hospital care | 1.5 | 1,709 | -0.1 | -73 |
| | | 907 | Filled prescriptions | 0.2 | 238 | 0.0 | 30 |
| | | 801 | Lost productivity (income loss) | . | . | -4.4 | -5,518 |
| Young adults and adults (25-64 years) | Siblings | 87 | Primary sector | 0.0 | 316 | 0.0 | 64 |
| | | 87 | Hospital care | 0.1 | 1,015 | -0.1 | -999 |
| | | 87 | Filled prescriptions | 0.0 | 303 | 0.0 | 89 |
| | | 76 | Lost productivity (income loss) | . | . | -0.9 | -11,744 |
| Older people (65+ years) | Children | 335 | Primary sector | 0.1 | 335 | 0.0 | 67 |
| | | 335 | Hospital care | 0.8 | 2,430 | -0.1 | -409 |
| | | 335 | Filled prescriptions | 0.1 | 322 | 0.0 | 112 |
| | | 314 | Lost productivity (income loss) | . | . | 2.5 | 7,821 |
| Older people (65+ years) | Partners | 624 | Primary sector | 0.3 | 541 | 0.0 | 60 |
| | | 624 | Hospital care | 2.7 | 4,370 | 0.6 | 928 |
| | | 624 | Filled prescriptions | 0.3 | 518 | 0.0 | 20 |
| | | 42 | Lost productivity (income loss) | . | . | -0.6 | -14,788 |
| Older people (65+ years) | Siblings | 5 | Primary sector | 0.0 | 551 | 0.0 | 300 |
| | | 5 | Hospital care | 0.1 | 14,469 | 0.1 | 14,222 |
| | | 5 | Filled prescriptions | 0.0 | 308 | -0.0 | -97 |
| | | . | Lost productivity (income loss) | . | . | . | . |

### Supplementary Table 192
### Neuromuscular disorders - Prevalent cohort 2021
### Identification of populations of relatives

| ***Agegroup of index patients in matched population*** | | ***Closest relative type identified*** | | ***Available control with same type of closest relative*** |
|---|---|---|---|---|
| Children and young people (0-24 years) | n = 1,183 | Fathers | n = 970 (82%) | 96% |
| | | Mothers | n = 985 (83%) | 97% |
| | | No available closest relative | n = 47 (4%) | n/a |
| Young adults and adults (25-64 years) | n = 4,378 | Children | n = 540 (12%) | 94% |
| | | Parents | n = 762 (17%) | 89% |
| | | Partners | n = 2,414 (55%) | 97% |
| | | Siblings | n = 537 (12%) | 49% |
| | | Pooled | n = 4,253 (97%) | 89% |
| | | No available closest relative | n = 125 (3%) | n/a |
| Older people (65+ years) | n = 2,637 | Children | n = 834 (32%) | 95% |
| | | Partners | n = 1,524 (58%) | 97% |
| | | Siblings | n = 45 (2%) | 44% |
| | | Pooled | n = 2,403 (91%) | 95% |
| | | No available closest relative | n = 234 (9%) | n/a |

**Supplementary Table 193**
**Neuromuscular disorders - Prevalent cohort 2021**
**Patient characteristics - cases**

| *Age strata of index patients* | *Characteristic* | |
|---|---|---|
| Children and young people (0-24 years) | Number of persons, n | 1,105 (100.0) |
| | Male, n (%) | 678 (61.4) |
| | Age, median (Q1;Q3) | 16.5 (11.5;20.9) |
| | CCI, n (%) | |
| | 0 | 782 (70.8) |
| | 1-2 | 284 (25.7) |
| | +3 | 39 (3.5) |
| | Education level, n (%) | |
| | No/Unknown education | 369 (33.4) |
| | Primary | 559 (50.6) |
| | Secondary | 159 (14.4) |
| | Tertiary | 18 (1.6) |
| Young adults and adults (25-64 years) | Number of persons, n | 3,810 (100.0) |
| | Male, n (%) | 1,862 (48.9) |
| | Age, median (Q1;Q3) | 49.5 (39.0;57.6) |
| | CCI, n (%) | |
| | 0 | 2,331 (61.2) |
| | 1-2 | 1,148 (30.1) |
| | +3 | 331 (8.7) |
| | Education level, n (%) | |
| | No/Unknown education | 80 (2.1) |
| | Primary | 931 (24.4) |
| | Secondary | 1,673 (43.9) |
| | Tertiary | 1,126 (29.6) |
| Older people (65+ years) | Number of persons, n | 2,298 (100.0) |
| | Male, n (%) | 1,137 (49.5) |
| | Age, median (Q1;Q3) | 74.5 (70.1;79.4) |
| | CCI, n (%) | |
| | 0 | 878 (38.2) |
| | 1-2 | 915 (39.8) |
| | +3 | 505 (22.0) |
| | Education level, n (%) | |
| | No/Unknown education | 149 (6.5) |
| | Primary | 647 (28.2) |
| | Secondary | 1,086 (47.3) |
| | Tertiary | 416 (18.1) |

#### Supplementary Table 194
#### Neuromuscular disorders - Prevalent cohort 2021
#### Patient characteristics - relatives - pooled

| *Age strata of index patients* | *Closest relative type* | *Characteristic of family relatives by agestrata of index patients* | *Closest family relatives of brain disorder patients* | *Comparison relative* |
|---|---|---|---|---|
| Children and young people (0-24 years) | Fathers | Number of persons, n | 940 (100.0) | 940 (100.0) |
| | | Male, n (%) | 940 (100.0) | 940 (100.0) |
| | | Age, median (Q1;Q3) | 48.8 (43.6;53.7) | 48.7 (43.8;53.3) |
| | | CCI, n (%) | | |
| | | 0 | 825 (87.8) | 841 (89.5) |
| | | 1-2 | 102 (10.9) | 87 (9.3) |
| | | +3 | 13 (1.4) | 12 (1.3) |
| | | Education level, n (%) | | |
| | | No/Unknown education | 12 (1.3) | 12 (1.3) |
| | | Primary | 202 (21.5) | 202 (21.5) |
| | | Secondary | 463 (49.3) | 463 (49.3) |
| | | Tertiary | 263 (28.0) | 263 (28.0) |
| | | Relation to patient, n (%) | | |
| | | Father | 940 (100.0) | 940 (100.0) |
| Children and young people (0-24 years) | Mothers | Number of persons, n | 957 (100.0) | 957 (100.0) |
| | | Male, n (%) | 0 (0.0) | 0 (0.0) |
| | | Age, median (Q1;Q3) | 46.7 (41.6;51.3) | 46.7 (41.4;50.9) |
| | | CCI, n (%) | | |
| | | 0 | 831 (86.8) | 847 (88.5) |
| | | 1-2 | 112 (11.7) | 101 (10.6) |
| | | +3 | 14 (1.5) | 9 (0.9) |
| | | Education level, n (%) | | |
| | | No/Unknown education | 18 (1.9) | 18 (1.9) |
| | | Primary | 169 (17.7) | 169 (17.7) |
| | | Secondary | 372 (38.9) | 372 (38.9) |
| | | Tertiary | 398 (41.6) | 398 (41.6) |
| | | Relation to patient, n (%) | | |
| | | Mother | 957 (100.0) | 957 (100.0) |
| Young adults and adults (25-64 years) | Pooled | Number of persons, n | 3,810 (100.0) | 3,810 (100.0) |
| | | Male, n (%) | 1,881 (49.4) | 1,881 (49.4) |
| | | Age, median (Q1;Q3) | 52.2 (37.3;60.6) | 52.6 (37.4;60.3) |
| | | CCI, n (%) | | |
| | | 0 | 3,147 (82.6) | 3,187 (83.6) |
| | | 1-2 | 557 (14.6) | 518 (13.6) |
| | | +3 | 106 (2.8) | 105 (2.8) |
| | | Education level, n (%) | | |
| | | No/Unknown education | 158 (4.1) | 158 (4.1) |
| | | Primary | 875 (23.0) | 875 (23.0) |
| | | Secondary | 1,799 (47.2) | 1,799 (47.2) |
| | | Tertiary | 978 (25.7) | 978 (25.7) |
| | | Relation to patient, n (%) | | |
| | | Mother | 512 (13.4) | 512 (13.4) |
| | | Father | 170 (4.5) | 170 (4.5) |
| | | Partner | 2,354 (61.8) | 2,354 (61.8) |
| | | Child | 509 (13.4) | 509 (13.4) |

**Supplementary Table 194**
**Neuromuscular disorders - Prevalent cohort 2021**
**Patient characteristics - relatives - pooled**

| *Age strata of index patients* | *Closest relative type* | *Characteristic of family relatives by agestrata of index patients* | *Closest family relatives of brain disorder patients* | *Comparison relative* |
|---|---|---|---|---|
| | | Sibling | 265 (7.0) | 265 (7.0) |
| Older people (65+ years) | Pooled | Number of persons, n | 2,298 (100.0) | 2,298 (100.0) |
| | | Male, n (%) | 1,044 (45.4) | 1,044 (45.4) |
| | | Age, median (Q1;Q3) | 67.9 (56.7;74.8) | 68.0 (56.3;74.6) |
| | | CCI, n (%) | | |
| | | 0 | 1,607 (69.9) | 1,606 (69.9) |
| | | 1-2 | 526 (22.9) | 546 (23.8) |
| | | +3 | 165 (7.2) | 146 (6.4) |
| | | Education level, n (%) | | |
| | | No/Unknown education | 83 (3.6) | 83 (3.6) |
| | | Primary | 499 (21.7) | 499 (21.7) |
| | | Secondary | 1,131 (49.2) | 1,131 (49.2) |
| | | Tertiary | 585 (25.5) | 585 (25.5) |
| | | Relation to patient, n (%) | | |
| | | Partner | 1,483 (64.5) | 1,483 (64.5) |
| | | Child | 795 (34.6) | 795 (34.6) |
| | | Sibling | 20 (0.9) | 20 (0.9) |

#### Supplementary Table 195
#### Neuromuscular disorders - Prevalent cohort 2021
#### Patient characteristics - relatives - not pooled

| *Age strata of index patients* | *Closest relative type* | *Characteristic of family relatives by agestrata of index patients* | *Closest family relatives of brain disorder patients* | *Comparison relative* |
|---|---|---|---|---|
| Children and young people (0-24 years) | Fathers | Number of persons, n | 940 (100.0) | 940 (100.0) |
| | | Male, n (%) | 940 (100.0) | 940 (100.0) |
| | | Age, median (Q1;Q3) | 48.8 (43.6;53.7) | 48.7 (43.8;53.3) |
| | | CCI, n (%) | | |
| | | 0 | 825 (87.8) | 841 (89.5) |
| | | 1-2 | 102 (10.9) | 87 (9.3) |
| | | +3 | 13 (1.4) | 12 (1.3) |
| | | Education level, n (%) | | |
| | | No/Unknown education | 12 (1.3) | 12 (1.3) |
| | | Primary | 202 (21.5) | 202 (21.5) |
| | | Secondary | 463 (49.3) | 463 (49.3) |
| | | Tertiary | 263 (28.0) | 263 (28.0) |
| | | Relation to patient, n (%) | | |
| | | Father | 940 (100.0) | 940 (100.0) |
| Children and young people (0-24 years) | Mothers | Number of persons, n | 957 (100.0) | 957 (100.0) |
| | | Male, n (%) | 0 (0.0) | 0 (0.0) |
| | | Age, median (Q1;Q3) | 46.7 (41.6;51.3) | 46.7 (41.4;50.9) |
| | | CCI, n (%) | | |
| | | 0 | 831 (86.8) | 847 (88.5) |
| | | 1-2 | 112 (11.7) | 101 (10.6) |
| | | +3 | 14 (1.5) | 9 (0.9) |
| | | Education level, n (%) | | |
| | | No/Unknown education | 18 (1.9) | 18 (1.9) |
| | | Primary | 169 (17.7) | 169 (17.7) |
| | | Secondary | 372 (38.9) | 372 (38.9) |
| | | Tertiary | 398 (41.6) | 398 (41.6) |
| | | Relation to patient, n (%) | | |
| | | Mother | 957 (100.0) | 957 (100.0) |
| Young adults and adults (25-64 years) | Children | Number of persons, n | 509 (100.0) | 509 (100.0) |
| | | Male, n (%) | 276 (54.2) | 276 (54.2) |
| | | Age, median (Q1;Q3) | 22.9 (15.1;30.3) | 22.9 (15.1;30.3) |
| | | CCI, n (%) | | |
| | | 0 | 467 (91.7) | 471 (92.5) |
| | | 1-2 | * | * |
| | | +3 | * | * |
| | | Education level, n (%) | | |
| | | No/Unknown education | 96 (18.9) | 96 (18.9) |
| | | Primary | 209 (41.1) | 209 (41.1) |
| | | Secondary | 146 (28.7) | 146 (28.7) |
| | | Tertiary | 58 (11.4) | 58 (11.4) |
| | | Relation to patient, n (%) | | |
| | | Child | 509 (100.0) | 509 (100.0) |
| Young adults and adults (25-64 years) | Parents | Number of persons, n | 682 (100.0) | 682 (100.0) |
| | | Male, n (%) | 170 (24.9) | 170 (24.9) |
| | | Age, median (Q1;Q3) | 65.9 (59.4;74.3) | 66.0 (59.2;73.9) |

**Supplementary Table 195**
**Neuromuscular disorders - Prevalent cohort 2021**
**Patient characteristics - relatives - not pooled**

| *Age strata of index patients* | *Closest relative type* | *Characteristic of family relatives by agestrata of index patients* | *Closest family relatives of brain disorder patients* | *Comparison relative* |
|---|---|---|---|---|
| | | CCI, n (%) | | |
| | | 0 | 461 (67.6) | 473 (69.4) |
| | | 1-2 | 175 (25.7) | 157 (23.0) |
| | | +3 | 46 (6.7) | 52 (7.6) |
| | | Education level, n (%) | | |
| | | No/Unknown education | 8 (1.2) | 8 (1.2) |
| | | Primary | 204 (29.9) | 204 (29.9) |
| | | Secondary | 309 (45.3) | 309 (45.3) |
| | | Tertiary | 161 (23.6) | 161 (23.6) |
| | | Relation to patient, n (%) | | |
| | | Mother | 512 (75.1) | 512 (75.1) |
| | | Father | 170 (24.9) | 170 (24.9) |
| Young adults and adults (25-64 years) | Partners | Number of persons, n | 2,354 (100.0) | 2,354 (100.0) |
| | | Male, n (%) | 1,274 (54.1) | 1,274 (54.1) |
| | | Age, median (Q1;Q3) | 51.1 (41.3;58.3) | 51.5 (41.4;58.2) |
| | | CCI, n (%) | | |
| | | 0 | 1,998 (84.9) | 2,021 (85.9) |
| | | 1-2 | 300 (12.7) | 284 (12.1) |
| | | +3 | 56 (2.4) | 49 (2.1) |
| | | Education level, n (%) | | |
| | | No/Unknown education | 54 (2.3) | 54 (2.3) |
| | | Primary | 402 (17.1) | 402 (17.1) |
| | | Secondary | 1,188 (50.5) | 1,188 (50.5) |
| | | Tertiary | 710 (30.2) | 710 (30.2) |
| | | Relation to patient, n (%) | | |
| | | Partner | 2,354 (100.0) | 2,354 (100.0) |
| Young adults and adults (25-64 years) | Siblings | Number of persons, n | 265 (100.0) | 265 (100.0) |
| | | Male, n (%) | 161 (60.8) | 161 (60.8) |
| | | Age, median (Q1;Q3) | 55.6 (44.5;60.5) | 55.0 (42.5;60.5) |
| | | CCI, n (%) | | |
| | | 0 | 221 (83.4) | 222 (83.8) |
| | | 1-2 | * | * |
| | | +3 | * | * |
| | | Education level, n (%) | | |
| | | No/Unknown education | 0 (0.0) | 0 (0.0) |
| | | Primary | 60 (22.6) | 60 (22.6) |
| | | Secondary | 156 (58.9) | 156 (58.9) |
| | | Tertiary | 49 (18.5) | 49 (18.5) |
| | | Relation to patient, n (%) | | |
| | | Sibling | 265 (100.0) | 265 (100.0) |
| Older people (65+ years) | Children | Number of persons, n | 795 (100.0) | 795 (100.0) |
| | | Male, n (%) | 419 (52.7) | 419 (52.7) |
| | | Age, median (Q1;Q3) | 52.2 (46.3;57.7) | 52.0 (46.0;57.2) |
| | | CCI, n (%) | | |
| | | 0 | 673 (84.7) | 690 (86.8) |

#### Supplementary Table 195
#### Neuromuscular disorders - Prevalent cohort 2021
#### Patient characteristics - relatives - not pooled

| *Age strata of index patients* | *Closest relative type* | *Characteristic of family relatives by agestrata of index patients* | *Closest family relatives of brain disorder patients* | *Comparison relative* |
|---|---|---|---|---|
| | | 1-2 | 104 (13.1) | 95 (11.9) |
| | | +3 | 18 (2.3) | 10 (1.3) |
| | | Education level, n (%) | | |
| | | No/Unknown education | * | * |
| | | Primary | * | * |
| | | Secondary | 396 (49.8) | 396 (49.8) |
| | | Tertiary | 273 (34.3) | 273 (34.3) |
| | | Relation to patient, n (%) | | |
| | | Child | 795 (100.0) | 795 (100.0) |
| Older people (65+ years) | Partners | Number of persons, n | 1,483 (100.0) | 1,483 (100.0) |
| | | Male, n (%) | 614 (41.4) | 614 (41.4) |
| | | Age, median (Q1;Q3) | 73.0 (68.2;77.5) | 72.9 (68.4;77.3) |
| | | CCI, n (%) | | |
| | | 0 | 920 (62.0) | 902 (60.8) |
| | | 1-2 | 416 (28.1) | 446 (30.1) |
| | | +3 | 147 (9.9) | 135 (9.1) |
| | | Education level, n (%) | | |
| | | No/Unknown education | 81 (5.5) | 81 (5.5) |
| | | Primary | 371 (25.0) | 371 (25.0) |
| | | Secondary | 721 (48.6) | 721 (48.6) |
| | | Tertiary | 310 (20.9) | 310 (20.9) |
| | | Relation to patient, n (%) | | |
| | | Partner | 1,483 (100.0) | 1,483 (100.0) |
| Older people (65+ years) | Siblings | Number of persons, n | 20 (100.0) | 20 (100.0) |
| | | Male, n (%) | 11 (55.0) | 11 (55.0) |
| | | Age, median (Q1;Q3) | 61.1 (58.8;62.9) | 61.7 (60.1;64.8) |
| | | CCI, n (%) | | |
| | | 0 | 14 (70.0) | * |
| | | 1-2 | 6 (30.0) | * |
| | | +3 | 0 (0.0) | * |
| | | Education level, n (%) | | |
| | | No/Unknown education | 0 (0.0) | 0 (0.0) |
| | | Primary | * | * |
| | | Secondary | * | * |
| | | Tertiary | * | * |
| | | Relation to patient, n (%) | | |
| | | Sibling | 20 (100.0) | 20 (100.0) |

#### Supplementary Table 196
#### Neuromuscular disorders - Prevalent cohort 2021
#### Cost analysis - pooled

| | | | | *Actual costs* | | *Attributable costs* | |
|---|---|---|---|---|---|---|---|
| *Age strata of index patients* | *Closest relative type* | *Number of index patients included in cost component calculations* | *Cost component* | *Total (million EUR)* | *Per person (EUR)* | *Total (million EUR)* | *Per person (EUR)* |
| Children and young people (0-24 years) | Fathers | 940 | Primary sector | 0.2 | 231 | 0.0 | 17 |
| | | 940 | Hospital care | 1.5 | 1,569 | 0.3 | 307 |
| | | 940 | Filled prescriptions | 0.1 | 141 | -0.0 | -28 |
| | | 921 | Lost productivity (income loss) | . | . | 6.2 | 6,720 |
| Children and young people (0-24 years) | Mothers | 957 | Primary sector | 0.4 | 371 | 0.0 | 20 |
| | | 957 | Hospital care | 1.7 | 1,756 | 0.0 | 5 |
| | | 957 | Filled prescriptions | 0.2 | 216 | 0.1 | 55 |
| | | 954 | Lost productivity (income loss) | . | . | 5.1 | 5,301 |
| Young adults and adults (25-64 years) | Pooled | 3,810 | Primary sector | 1.2 | 325 | 0.1 | 26 |
| | | 3,810 | Hospital care | 7.1 | 1,857 | -0.9 | -239 |
| | | 3,810 | Filled prescriptions | 1.0 | 264 | 0.1 | 27 |
| | | 2,923 | Lost productivity (income loss) | . | . | -0.4 | -125 |
| Older people (65+ years) | Pooled | 2,298 | Primary sector | 1.1 | 463 | 0.1 | 38 |
| | | 2,298 | Hospital care | 8.1 | 3,546 | 0.5 | 210 |
| | | 2,298 | Filled prescriptions | 1.1 | 465 | 0.0 | 18 |
| | | 823 | Lost productivity (income loss) | . | . | 0.8 | 1,026 |

#### Supplementary Table 197
#### Neuromuscular disorders - Prevalent cohort 2021
#### Cost analysis - not pooled

| Age strata of index patients | Closest relative type | Number of index patients included in cost component calculations | Cost component | Actual costs: Total (million EUR) | Actual costs: Per person (EUR) | Attributable costs: Total (million EUR) | Attributable costs: Per person (EUR) |
|---|---|---|---|---|---|---|---|
| Children and young people (0-24 years) | Fathers | 940 | Primary sector | 0.2 | 231 | 0.0 | 17 |
| | | 940 | Hospital care | 1.5 | 1,569 | 0.3 | 307 |
| | | 940 | Filled prescriptions | 0.1 | 141 | -0.0 | -28 |
| | | 921 | Lost productivity (income loss) | . | . | 6.2 | 6,720 |
| Children and young people (0-24 years) | Mothers | 957 | Primary sector | 0.4 | 371 | 0.0 | 20 |
| | | 957 | Hospital care | 1.7 | 1,756 | 0.0 | 5 |
| | | 957 | Filled prescriptions | 0.2 | 216 | 0.1 | 55 |
| | | 954 | Lost productivity (income loss) | . | . | 5.1 | 5,301 |
| Young adults and adults (25-64 years) | Children | 509 | Primary sector | 0.1 | 239 | 0.0 | 56 |
| | | 509 | Hospital care | 0.5 | 1,009 | -0.2 | -423 |
| | | 509 | Filled prescriptions | 0.1 | 102 | 0.0 | 8 |
| | | 317 | Lost productivity (income loss) | . | . | -0.2 | -542 |
| Young adults and adults (25-64 years) | Parents | 682 | Primary sector | 0.3 | 466 | 0.0 | 51 |
| | | 682 | Hospital care | 2.1 | 3,101 | -0.3 | -469 |
| | | 682 | Filled prescriptions | 0.3 | 489 | 0.0 | 38 |
| | | 265 | Lost productivity (income loss) | . | . | -0.4 | -1,511 |
| Young adults and adults (25-64 years) | Partners | 2,354 | Primary sector | 0.7 | 302 | 0.0 | 11 |
| | | 2,354 | Hospital care | 4.0 | 1,706 | -0.3 | -127 |
| | | 2,354 | Filled prescriptions | 0.5 | 233 | 0.1 | 29 |
| | | 2,113 | Lost productivity (income loss) | . | . | -0.5 | -232 |
| Young adults and adults (25-64 years) | Siblings | 265 | Primary sector | 0.1 | 329 | 0.0 | 41 |
| | | 265 | Hospital care | 0.4 | 1,639 | -0.1 | -290 |
| | | 265 | Filled prescriptions | 0.1 | 278 | 0.0 | 15 |
| | | 228 | Lost productivity (income loss) | . | . | 0.7 | 3,056 |
| Older people (65+ years) | Children | 795 | Primary sector | 0.3 | 330 | 0.0 | 34 |
| | | 795 | Hospital care | 1.6 | 2,045 | 0.3 | 386 |
| | | 795 | Filled prescriptions | 0.2 | 297 | -0.0 | -24 |
| | | 737 | Lost productivity (income loss) | . | . | 2.8 | 3,769 |
| Older people (65+ years) | Partners | 1,483 | Primary sector | 0.8 | 534 | 0.1 | 37 |
| | | 1,483 | Hospital care | 6.4 | 4,391 | 0.2 | 118 |
| | | 1,483 | Filled prescriptions | 0.8 | 557 | 0.1 | 37 |
| | | 74 | Lost productivity (income loss) | . | . | -0.2 | -2,372 |
| Older people (65+ years) | Siblings | 20 | Primary sector | 0.0 | 535 | 0.0 | 268 |
| | | 20 | Hospital care | 0.0 | 1,214 | -0.0 | -96 |
| | | 20 | Filled prescriptions | 0.0 | 442 | 0.0 | 339 |
| | | 12 | Lost productivity (income loss) | . | . | -1.8 | -146,514 |

### Supplementary Table 198
#### Other neurodegenerative disorders - Incident cohort 2016-2021
#### Identification of populations of relatives

| *Agegroup of index patients in matched population* | | *Closest relative type identified* | | *Available control with same type of closest relative* |
|---|---|---|---|---|
| Children and young people (0-24 years) | n = 152 | Fathers | n = 109 (72%) | 91% |
| | | Mothers | n = 114 (75%) | 93% |
| | | No available closest relative | n = 23 (15%) | n/a |
| Young adults and adults (25-64 years) | n = 978 | Children | n = 163 (17%) | 93% |
| | | Parents | n = 88 (9%) | 89% |
| | | Partners | n = 568 (58%) | 96% |
| | | Siblings | n = 113 (12%) | 58% |
| | | Pooled | n = 932 (95%) | 90% |
| | | No available closest relative | n = 46 (5%) | n/a |
| Older people (65+ years) | n = 1,490 | Children | n = 522 (35%) | 94% |
| | | Partners | n = 812 (54%) | 96% |
| | | Siblings | n = 27 (2%) | 33% |
| | | Pooled | n = 1,361 (91%) | 94% |
| | | No available closest relative | n = 129 (9%) | n/a |

### Supplementary Table 199
### Other neurodegenerative disorders - Incident cohort 2016-2021
### Patient characteristics - cases

| *Age strata of index patients* | *Characteristic* | |
|---|---|---|
| Children and young people (0-24 years) | Number of persons, n | 120 (100.0) |
| | Male, n (%) | 57 (47.5) |
| | Age, median (Q1;Q3) | 12.6 (5.3;18.8) |
| | CCI, n (%) | |
| | 0 | 99 (82.5) |
| | 1-2 | * |
| | +3 | * |
| | Education level, n (%) | |
| | No/Unknown education | 64 (53.3) |
| | Primary | 45 (37.5) |
| | Secondary | * |
| | Tertiary | * |
| Young adults and adults (25-64 years) | Number of persons, n | 844 (100.0) |
| | Male, n (%) | 491 (58.2) |
| | Age, median (Q1;Q3) | 55.6 (47.5;60.9) |
| | CCI, n (%) | |
| | 0 | 612 (72.5) |
| | 1-2 | 194 (23.0) |
| | +3 | 38 (4.5) |
| | Education level, n (%) | |
| | No/Unknown education | 31 (3.7) |
| | Primary | 196 (23.2) |
| | Secondary | 413 (48.9) |
| | Tertiary | 204 (24.2) |
| Older people (65+ years) | Number of persons, n | 1,287 (100.0) |
| | Male, n (%) | 655 (50.9) |
| | Age, median (Q1;Q3) | 73.2 (69.5;77.8) |
| | CCI, n (%) | |
| | 0 | 697 (54.2) |
| | 1-2 | 451 (35.0) |
| | +3 | 139 (10.8) |
| | Education level, n (%) | |
| | No/Unknown education | 88 (6.8) |
| | Primary | 316 (24.6) |
| | Secondary | 600 (46.6) |
| | Tertiary | 283 (22.0) |

**Supplementary Table 200**
**Other neurodegenerative disorders - Incident cohort 2016-2021**
**Patient characteristics - relatives - pooled**

| *Age strata of index patients* | *Closest relative type* | *Characteristic of family relatives by agestrata of index patients* | *Closest family relatives of brain disorder patients* | *Comparison relative* |
|---|---|---|---|---|
| Children and young people (0-24 years) | Fathers | Number of persons, n | 100 (100.0) | 100 (100.0) |
| | | Male, n (%) | 100 (100.0) | 100 (100.0) |
| | | Age, median (Q1;Q3) | 44.3 (38.0;50.7) | 45.8 (37.2;50.8) |
| | | CCI, n (%) | | |
| | | 0 | 87 (87.0) | * |
| | | 1-2 | * | * |
| | | +3 | * | * |
| | | Education level, n (%) | | |
| | | No/Unknown education | * | * |
| | | Primary | 30 (30.0) | 30 (30.0) |
| | | Secondary | 46 (46.0) | 46 (46.0) |
| | | Tertiary | * | * |
| | | Relation to patient, n (%) | | |
| | | Father | 100 (100.0) | 100 (100.0) |
| Children and young people (0-24 years) | Mothers | Number of persons, n | 107 (100.0) | 107 (100.0) |
| | | Male, n (%) | 0 (0.0) | 0 (0.0) |
| | | Age, median (Q1;Q3) | 42.2 (35.4;49.2) | 42.0 (34.7;49.2) |
| | | CCI, n (%) | | |
| | | 0 | 95 (88.8) | * |
| | | 1-2 | * | * |
| | | +3 | * | * |
| | | Education level, n (%) | | |
| | | No/Unknown education | * | * |
| | | Primary | * | * |
| | | Secondary | 39 (36.4) | 39 (36.4) |
| | | Tertiary | 38 (35.5) | 38 (35.5) |
| | | Relation to patient, n (%) | | |
| | | Mother | 107 (100.0) | 107 (100.0) |
| Young adults and adults (25-64 years) | Pooled | Number of persons, n | 844 (100.0) | 844 (100.0) |
| | | Male, n (%) | 368 (43.6) | 368 (43.6) |
| | | Age, median (Q1;Q3) | 53.2 (38.3;60.5) | 53.1 (38.8;60.7) |
| | | CCI, n (%) | | |
| | | 0 | 691 (81.9) | 693 (82.1) |
| | | 1-2 | 123 (14.6) | 134 (15.9) |
| | | +3 | 30 (3.6) | 17 (2.0) |
| | | Education level, n (%) | | |
| | | No/Unknown education | 41 (4.9) | 41 (4.9) |
| | | Primary | 218 (25.8) | 218 (25.8) |
| | | Secondary | 375 (44.4) | 375 (44.4) |
| | | Tertiary | 210 (24.9) | 210 (24.9) |
| | | Relation to patient, n (%) | | |
| | | Mother | 60 (7.1) | 60 (7.1) |
| | | Father | 19 (2.3) | 19 (2.3) |
| | | Partner | 546 (64.7) | 546 (64.7) |
| | | Child | 153 (18.1) | 153 (18.1) |

**Supplementary Table 200**
**Other neurodegenerative disorders - Incident cohort 2016-2021**
**Patient characteristics - relatives - pooled**

| *Age strata of index patients* | *Closest relative type* | *Characteristic of family relatives by agestrata of index patients* | *Closest family relatives of brain disorder patients* | *Comparison relative* |
|---|---|---|---|---|
| | | Sibling | 66 (7.8) | 66 (7.8) |
| Older people (65+ years) | Pooled | Number of persons, n | 1,287 (100.0) | 1,287 (100.0) |
| | | Male, n (%) | 591 (45.9) | 591 (45.9) |
| | | Age, median (Q1;Q3) | 65.8 (53.2;73.0) | 66.2 (52.8;72.6) |
| | | CCI, n (%) | | |
| | | 0 | 956 (74.3) | 917 (71.3) |
| | | 1-2 | 264 (20.5) | 280 (21.8) |
| | | +3 | 67 (5.2) | 90 (7.0) |
| | | Education level, n (%) | | |
| | | No/Unknown education | 55 (4.3) | 55 (4.3) |
| | | Primary | 253 (19.7) | 253 (19.7) |
| | | Secondary | 627 (48.7) | 627 (48.7) |
| | | Tertiary | 352 (27.4) | 352 (27.4) |
| | | Relation to patient, n (%) | | |
| | | Partner | 784 (60.9) | 784 (60.9) |
| | | Child | 494 (38.4) | 494 (38.4) |
| | | Sibling | 9 (0.7) | 9 (0.7) |

**Supplementary Table 201**
**Other neurodegenerative disorders - Incident cohort 2016-2021**
**Patient characteristics - relatives - not pooled**

| *Age strata of index patients* | *Closest relative type* | *Characteristic of family relatives by agestrata of index patients* | *Closest family relatives of brain disorder patients* | *Comparison relative* |
|---|---|---|---|---|
| Children and young people (0-24 years) | Fathers | Number of persons, n | 100 (100.0) | 100 (100.0) |
| | | Male, n (%) | 100 (100.0) | 100 (100.0) |
| | | Age, median (Q1;Q3) | 44.3 (38.0;50.7) | 45.8 (37.2;50.8) |
| | | CCI, n (%) | | |
| | | 0 | 87 (87.0) | * |
| | | 1-2 | * | * |
| | | +3 | * | * |
| | | Education level, n (%) | | |
| | | No/Unknown education | * | * |
| | | Primary | 30 (30.0) | 30 (30.0) |
| | | Secondary | 46 (46.0) | 46 (46.0) |
| | | Tertiary | * | * |
| | | Relation to patient, n (%) | | |
| | | Father | 100 (100.0) | 100 (100.0) |
| Children and young people (0-24 years) | Mothers | Number of persons, n | 107 (100.0) | 107 (100.0) |
| | | Male, n (%) | 0 (0.0) | 0 (0.0) |
| | | Age, median (Q1;Q3) | 42.2 (35.4;49.2) | 42.0 (34.7;49.2) |
| | | CCI, n (%) | | |
| | | 0 | 95 (88.8) | * |
| | | 1-2 | * | * |
| | | +3 | * | * |
| | | Education level, n (%) | | |
| | | No/Unknown education | * | * |
| | | Primary | * | * |
| | | Secondary | 39 (36.4) | 39 (36.4) |
| | | Tertiary | 38 (35.5) | 38 (35.5) |
| | | Relation to patient, n (%) | | |
| | | Mother | 107 (100.0) | 107 (100.0) |
| Young adults and adults (25-64 years) | Children | Number of persons, n | 153 (100.0) | 153 (100.0) |
| | | Male, n (%) | 81 (52.9) | 81 (52.9) |
| | | Age, median (Q1;Q3) | 27.4 (20.6;33.4) | 27.8 (19.5;34.1) |
| | | CCI, n (%) | | |
| | | 0 | * | 148 (96.7) |
| | | 1-2 | * | 5 (3.3) |
| | | +3 | * | 0 (0.0) |
| | | Education level, n (%) | | |
| | | No/Unknown education | 16 (10.5) | 16 (10.5) |
| | | Primary | 62 (40.5) | 62 (40.5) |
| | | Secondary | 49 (32.0) | 49 (32.0) |
| | | Tertiary | 26 (17.0) | 26 (17.0) |
| | | Relation to patient, n (%) | | |
| | | Child | 153 (100.0) | 153 (100.0) |
| Young adults and adults (25-64 years) | Parents | Number of persons, n | 79 (100.0) | 79 (100.0) |
| | | Male, n (%) | 19 (24.1) | 19 (24.1) |
| | | Age, median (Q1;Q3) | 68.7 (61.5;77.3) | 70.5 (62.1;78.5) |

**Supplementary Table 201**
**Other neurodegenerative disorders - Incident cohort 2016-2021**
**Patient characteristics - relatives - not pooled**

| *Age strata of index patients* | *Closest relative type* | *Characteristic of family relatives by agestrata of index patients* | *Closest family relatives of brain disorder patients* | *Comparison relative* |
|---|---|---|---|---|
| | | CCI, n (%) | | |
| | | 0 | 52 (65.8) | 50 (63.3) |
| | | 1-2 | 18 (22.8) | * |
| | | +3 | 9 (11.4) | * |
| | | Education level, n (%) | | |
| | | No/Unknown education | * | * |
| | | Primary | 33 (41.8) | 33 (41.8) |
| | | Secondary | 31 (39.2) | 31 (39.2) |
| | | Tertiary | * | * |
| | | Relation to patient, n (%) | | |
| | | Mother | 60 (75.9) | 60 (75.9) |
| | | Father | 19 (24.1) | 19 (24.1) |
| Young adults and adults (25-64 years) | Partners | Number of persons, n | 546 (100.0) | 546 (100.0) |
| | | Male, n (%) | 236 (43.2) | 236 (43.2) |
| | | Age, median (Q1;Q3) | 54.6 (45.9;60.5) | 55.1 (46.4;60.7) |
| | | CCI, n (%) | | |
| | | 0 | 444 (81.3) | 446 (81.7) |
| | | 1-2 | 83 (15.2) | 88 (16.1) |
| | | +3 | 19 (3.5) | 12 (2.2) |
| | | Education level, n (%) | | |
| | | No/Unknown education | 23 (4.2) | 23 (4.2) |
| | | Primary | 113 (20.7) | 113 (20.7) |
| | | Secondary | 250 (45.8) | 250 (45.8) |
| | | Tertiary | 160 (29.3) | 160 (29.3) |
| | | Relation to patient, n (%) | | |
| | | Partner | 546 (100.0) | 546 (100.0) |
| Young adults and adults (25-64 years) | Siblings | Number of persons, n | 66 (100.0) | 66 (100.0) |
| | | Male, n (%) | 32 (48.5) | 32 (48.5) |
| | | Age, median (Q1;Q3) | 55.9 (50.1;60.1) | 56.1 (50.4;59.4) |
| | | CCI, n (%) | | |
| | | 0 | 52 (78.8) | 49 (74.2) |
| | | 1-2 | * | * |
| | | +3 | * | * |
| | | Education level, n (%) | | |
| | | No/Unknown education | 0 (0.0) | 0 (0.0) |
| | | Primary | 10 (15.2) | 10 (15.2) |
| | | Secondary | 45 (68.2) | 45 (68.2) |
| | | Tertiary | 11 (16.7) | 11 (16.7) |
| | | Relation to patient, n (%) | | |
| | | Sibling | 66 (100.0) | 66 (100.0) |
| Older people (65+ years) | Children | Number of persons, n | 494 (100.0) | 494 (100.0) |
| | | Male, n (%) | 262 (53.0) | 262 (53.0) |
| | | Age, median (Q1;Q3) | 50.1 (44.7;55.3) | 50.2 (44.8;54.4) |
| | | CCI, n (%) | | |
| | | 0 | 428 (86.6) | 428 (86.6) |

#### Supplementary Table 201
#### Other neurodegenerative disorders - Incident cohort 2016-2021
#### Patient characteristics - relatives - not pooled

| *Age strata of index patients* | *Closest relative type* | *Characteristic of family relatives by agestrata of index patients* | *Closest family relatives of brain disorder patients* | *Comparison relative* |
|---|---|---|---|---|
| | | 1-2 | 56 (11.3) | 52 (10.5) |
| | | +3 | 10 (2.0) | 14 (2.8) |
| | | Education level, n (%) | | |
| | | No/Unknown education | * | * |
| | | Primary | * | * |
| | | Secondary | 249 (50.4) | 249 (50.4) |
| | | Tertiary | 156 (31.6) | 156 (31.6) |
| | | Relation to patient, n (%) | | |
| | | Child | 494 (100.0) | 494 (100.0) |
| Older people (65+ years) | Partners | Number of persons, n | 784 (100.0) | 784 (100.0) |
| | | Male, n (%) | 325 (41.5) | 325 (41.5) |
| | | Age, median (Q1;Q3) | 71.3 (67.3;76.1) | 71.4 (67.4;75.4) |
| | | CCI, n (%) | | |
| | | 0 | 522 (66.6) | 484 (61.7) |
| | | 1-2 | 207 (26.4) | 225 (28.7) |
| | | +3 | 55 (7.0) | 75 (9.6) |
| | | Education level, n (%) | | |
| | | No/Unknown education | 54 (6.9) | 54 (6.9) |
| | | Primary | 164 (20.9) | 164 (20.9) |
| | | Secondary | 371 (47.3) | 371 (47.3) |
| | | Tertiary | 195 (24.9) | 195 (24.9) |
| | | Relation to patient, n (%) | | |
| | | Partner | 784 (100.0) | 784 (100.0) |
| Older people (65+ years) | Siblings | Number of persons, n | 9 (100.0) | 9 (100.0) |
| | | Male, n (%) | * | * |
| | | Age, median (Q1;Q3) | 63.6 (60.9;66.3) | 62.2 (58.8;63.3) |
| | | CCI, n (%) | | |
| | | 0 | * | * |
| | | 1-2 | * | * |
| | | +3 | * | * |
| | | Education level, n (%) | | |
| | | No/Unknown education | 0 (0.0) | 0 (0.0) |
| | | Primary | * | * |
| | | Secondary | * | * |
| | | Tertiary | * | * |
| | | Relation to patient, n (%) | | |
| | | Sibling | 9 (100.0) | 9 (100.0) |

#### Supplementary Table 202
#### Other neurodegenerative disorders - Incident cohort 2016-2021
#### Cost analysis - pooled

| Age strata of index patients | Closest relative type | Number of index patients included in cost component calculations | Cost component | Actual costs | | Attributable costs | |
|---|---|---|---|---|---|---|---|
| | | | | Total (million EUR) | Per person (EUR) | Total (million EUR) | Per person (EUR) |
| Children and young people (0-24 years) | Fathers | 100 | Primary sector | 0.0 | 223 | 0.0 | 9 |
| | | 100 | Hospital care | 0.2 | 2,118 | 0.1 | 1,147 |
| | | 100 | Filled prescriptions | 0.0 | 176 | 0.0 | 22 |
| | | 97 | Lost productivity (income loss) | . | . | 0.3 | 3,513 |
| Children and young people (0-24 years) | Mothers | 107 | Primary sector | 0.0 | 358 | 0.0 | 14 |
| | | 107 | Hospital care | 0.3 | 2,440 | 0.1 | 711 |
| | | 107 | Filled prescriptions | 0.0 | 179 | -0.0 | -103 |
| | | 106 | Lost productivity (income loss) | . | . | 0.1 | 975 |
| Young adults and adults (25-64 years) | Pooled | 844 | Primary sector | 0.3 | 340 | 0.0 | 32 |
| | | 844 | Hospital care | 2.3 | 2,747 | 0.2 | 231 |
| | | 844 | Filled prescriptions | 0.2 | 268 | 0.0 | 11 |
| | | 639 | Lost productivity (income loss) | . | . | 0.0 | 21 |
| Older people (65+ years) | Pooled | 1,287 | Primary sector | 0.5 | 398 | -0.0 | -27 |
| | | 1,287 | Hospital care | 3.8 | 3,003 | -0.4 | -320 |
| | | 1,287 | Filled prescriptions | 0.5 | 364 | -0.1 | -48 |
| | | 527 | Lost productivity (income loss) | . | . | 2.3 | 4,393 |

#### Supplementary Table 203
#### Other neurodegenerative disorders - Incident cohort 2016-2021
#### Cost analysis - not pooled

| | | | | Actual costs | | Attributable costs | |
|---|---|---|---|---|---|---|---|
| *Age strata of index patients* | *Closest relative type* | *Number of index patients included in cost component calculations* | *Cost component* | *Total (million EUR)* | *Per person (EUR)* | *Total (million EUR)* | *Per person (EUR)* |
| Children and young people (0-24 years) | Fathers | 100 | Primary sector | 0.0 | 223 | 0.0 | 9 |
| | | 100 | Hospital care | 0.2 | 2,118 | 0.1 | 1,147 |
| | | 100 | Filled prescriptions | 0.0 | 176 | 0.0 | 22 |
| | | 97 | Lost productivity (income loss) | . | . | 0.3 | 3,513 |
| Children and young people (0-24 years) | Mothers | 107 | Primary sector | 0.0 | 358 | 0.0 | 14 |
| | | 107 | Hospital care | 0.3 | 2,440 | 0.1 | 711 |
| | | 107 | Filled prescriptions | 0.0 | 179 | -0.0 | -103 |
| | | 106 | Lost productivity (income loss) | . | . | 0.1 | 975 |
| Young adults and adults (25-64 years) | Children | 153 | Primary sector | 0.0 | 267 | 0.0 | 65 |
| | | 153 | Hospital care | 0.1 | 860 | -0.0 | -135 |
| | | 153 | Filled prescriptions | 0.0 | 121 | -0.0 | -12 |
| | | 117 | Lost productivity (income loss) | . | . | 0.4 | 3,535 |
| Young adults and adults (25-64 years) | Parents | 79 | Primary sector | 0.0 | 467 | 0.0 | 17 |
| | | 79 | Hospital care | 0.4 | 5,157 | 0.1 | 1,953 |
| | | 79 | Filled prescriptions | 0.0 | 460 | 0.0 | 2 |
| | | 20 | Lost productivity (income loss) | . | . | -0.2 | -9,484 |
| Young adults and adults (25-64 years) | Partners | 546 | Primary sector | 0.2 | 345 | 0.0 | 23 |
| | | 546 | Hospital care | 1.6 | 2,909 | 0.1 | 182 |
| | | 546 | Filled prescriptions | 0.2 | 283 | 0.0 | 25 |
| | | 446 | Lost productivity (income loss) | . | . | -0.7 | -1,496 |
| Young adults and adults (25-64 years) | Siblings | 66 | Primary sector | 0.0 | 318 | 0.0 | 41 |
| | | 66 | Hospital care | 0.2 | 2,994 | -0.0 | -527 |
| | | 66 | Filled prescriptions | 0.0 | 264 | -0.0 | -33 |
| | | 56 | Lost productivity (income loss) | . | . | 0.5 | 8,152 |
| Older people (65+ years) | Children | 494 | Primary sector | 0.1 | 288 | -0.0 | -17 |
| | | 494 | Hospital care | 1.0 | 1,984 | 0.2 | 423 |
| | | 494 | Filled prescriptions | 0.1 | 214 | 0.0 | 28 |
| | | 483 | Lost productivity (income loss) | . | . | 2.9 | 5,966 |
| Older people (65+ years) | Partners | 784 | Primary sector | 0.4 | 468 | -0.0 | -30 |
| | | 784 | Hospital care | 2.8 | 3,637 | -0.6 | -793 |
| | | 784 | Filled prescriptions | 0.4 | 461 | -0.1 | -96 |
| | | 41 | Lost productivity (income loss) | . | . | -0.8 | -19,831 |
| Older people (65+ years) | Siblings | 9 | Primary sector | 0.0 | 325 | -0.0 | -246 |
| | | 9 | Hospital care | 0.0 | 4,060 | -0.0 | -176 |
| | | 9 | Filled prescriptions | 0.0 | 277 | 0.0 | 28 |
| | | . | Lost productivity (income loss) | . | . | . | . |

**Supplementary Table 204**
**Other neurodegenerative disorders - Prevalent cohort 2021**
**Identification of populations of relatives**

| ***Agegroup of index patients in matched population*** | | ***Closest relative type identified*** | | ***Available control with same type of closest relative*** |
|---|---|---|---|---|
| Children and young people (0-24 years) | n = 368 | Fathers | n = 314 (85%) | 94% |
| | | Mothers | n = 324 (88%) | 95% |
| | | No available closest relative | n = 8 (2%) | n/a |
| Young adults and adults (25-64 years) | n = 1,711 | Children | n = 288 (17%) | 93% |
| | | Parents | n = 303 (18%) | 91% |
| | | Partners | n = 799 (47%) | 97% |
| | | Siblings | n = 254 (15%) | 53% |
| | | Pooled | n = 1,644 (96%) | 88% |
| | | No available closest relative | n = 67 (4%) | n/a |
| Older people (65+ years) | n = 1,802 | Children | n = 603 (33%) | 94% |
| | | Partners | n = 972 (54%) | 96% |
| | | Siblings | n = 46 (3%) | 45% |
| | | Pooled | n = 1,621 (90%) | 94% |
| | | No available closest relative | n = 181 (10%) | n/a |

#### Supplementary Table 205
#### Other neurodegenerative disorders - Prevalent cohort 2021
#### Patient characteristics - cases

| *Age strata of index patients* | *Characteristic* | |
|---|---|---|
| Children and young people (0-24 years) | Number of persons, n | 341 (100.0) |
| | Male, n (%) | 182 (53.4) |
| | Age, median (Q1;Q3) | 16.8 (11.8;20.8) |
| | CCI, n (%) | |
| | 0 | 217 (63.6) |
| | 1-2 | 105 (30.8) |
| | +3 | 19 (5.6) |
| | Education level, n (%) | |
| | No/Unknown education | 99 (29.0) |
| | Primary | 219 (64.2) |
| | Secondary | * |
| | Tertiary | * |
| Young adults and adults (25-64 years) | Number of persons, n | 1,462 (100.0) |
| | Male, n (%) | 821 (56.2) |
| | Age, median (Q1;Q3) | 53.1 (42.4;59.3) |
| | CCI, n (%) | |
| | 0 | 948 (64.8) |
| | 1-2 | 402 (27.5) |
| | +3 | 112 (7.7) |
| | Education level, n (%) | |
| | No/Unknown education | 45 (3.1) |
| | Primary | 404 (27.6) |
| | Secondary | 641 (43.8) |
| | Tertiary | 372 (25.4) |
| Older people (65+ years) | Number of persons, n | 1,527 (100.0) |
| | Male, n (%) | 808 (52.9) |
| | Age, median (Q1;Q3) | 74.1 (70.0;78.7) |
| | CCI, n (%) | |
| | 0 | 687 (45.0) |
| | 1-2 | 596 (39.0) |
| | +3 | 244 (16.0) |
| | Education level, n (%) | |
| | No/Unknown education | 99 (6.5) |
| | Primary | 392 (25.7) |
| | Secondary | 688 (45.1) |
| | Tertiary | 348 (22.8) |

**Supplementary Table 206**
**Other neurodegenerative disorders - Prevalent cohort 2021**
**Patient characteristics - relatives - pooled**

| *Age strata of index patients* | *Closest relative type* | *Characteristic of family relatives by agestrata of index patients* | *Closest family relatives of brain disorder patients* | *Comparison relative* |
|---|---|---|---|---|
| Children and young people (0-24 years) | Fathers | Number of persons, n | 296 (100.0) | 296 (100.0) |
| | | Male, n (%) | 296 (100.0) | 296 (100.0) |
| | | Age, median (Q1;Q3) | 49.0 (43.4;54.3) | 49.1 (43.7;53.7) |
| | | CCI, n (%) | | |
| | | 0 | 251 (84.8) | 263 (88.9) |
| | | 1-2 | * | 27 (9.1) |
| | | +3 | * | 6 (2.0) |
| | | Education level, n (%) | | |
| | | No/Unknown education | 7 (2.4) | 7 (2.4) |
| | | Primary | 74 (25.0) | 74 (25.0) |
| | | Secondary | 146 (49.3) | 146 (49.3) |
| | | Tertiary | 69 (23.3) | 69 (23.3) |
| | | Relation to patient, n (%) | | |
| | | Father | 296 (100.0) | 296 (100.0) |
| Children and young people (0-24 years) | Mothers | Number of persons, n | 309 (100.0) | 309 (100.0) |
| | | Male, n (%) | 0 (0.0) | 0 (0.0) |
| | | Age, median (Q1;Q3) | 45.9 (40.7;51.2) | 45.9 (40.8;51.0) |
| | | CCI, n (%) | | |
| | | 0 | 272 (88.0) | 273 (88.3) |
| | | 1-2 | 37 (12.0) | 30 (9.7) |
| | | +3 | 0 (0.0) | 6 (1.9) |
| | | Education level, n (%) | | |
| | | No/Unknown education | * | * |
| | | Primary | * | * |
| | | Secondary | 133 (43.0) | 133 (43.0) |
| | | Tertiary | 123 (39.8) | 123 (39.8) |
| | | Relation to patient, n (%) | | |
| | | Mother | 309 (100.0) | 309 (100.0) |
| Young adults and adults (25-64 years) | Pooled | Number of persons, n | 1,462 (100.0) | 1,462 (100.0) |
| | | Male, n (%) | 635 (43.4) | 635 (43.4) |
| | | Age, median (Q1;Q3) | 53.4 (36.9;61.2) | 53.6 (36.6;60.9) |
| | | CCI, n (%) | | |
| | | 0 | 1,169 (80.0) | 1,203 (82.3) |
| | | 1-2 | 246 (16.8) | 214 (14.6) |
| | | +3 | 47 (3.2) | 45 (3.1) |
| | | Education level, n (%) | | |
| | | No/Unknown education | 65 (4.4) | 65 (4.4) |
| | | Primary | 359 (24.6) | 359 (24.6) |
| | | Secondary | 698 (47.7) | 698 (47.7) |
| | | Tertiary | 340 (23.3) | 340 (23.3) |
| | | Relation to patient, n (%) | | |
| | | Mother | 204 (14.0) | 204 (14.0) |
| | | Father | 72 (4.9) | 72 (4.9) |
| | | Partner | 779 (53.3) | 779 (53.3) |
| | | Child | 270 (18.5) | 270 (18.5) |

**Supplementary Table 206**
**Other neurodegenerative disorders - Prevalent cohort 2021**
**Patient characteristics - relatives - pooled**

| *Age strata of index patients* | *Closest relative type* | *Characteristic of family relatives by agestrata of index patients* | *Closest family relatives of brain disorder patients* | *Comparison relative* |
|---|---|---|---|---|
| | | Sibling | 137 (9.4) | 137 (9.4) |
| Older people (65+ years) | Pooled | Number of persons, n | 1,527 (100.0) | 1,527 (100.0) |
| | | Male, n (%) | 673 (44.1) | 673 (44.1) |
| | | Age, median (Q1;Q3) | 67.0 (53.7;74.6) | 67.2 (53.8;74.5) |
| | | CCI, n (%) | | |
| | | 0 | 1,075 (70.4) | 1,099 (72.0) |
| | | 1-2 | 360 (23.6) | 349 (22.9) |
| | | +3 | 92 (6.0) | 79 (5.2) |
| | | Education level, n (%) | | |
| | | No/Unknown education | 63 (4.1) | 63 (4.1) |
| | | Primary | 326 (21.3) | 326 (21.3) |
| | | Secondary | 715 (46.8) | 715 (46.8) |
| | | Tertiary | 423 (27.7) | 423 (27.7) |
| | | Relation to patient, n (%) | | |
| | | Partner | 938 (61.4) | 938 (61.4) |
| | | Child | 568 (37.2) | 568 (37.2) |
| | | Sibling | 21 (1.4) | 21 (1.4) |

**Supplementary Table 207**
**Other neurodegenerative disorders - Prevalent cohort 2021**
**Patient characteristics - relatives - not pooled**

| *Age strata of index patients* | *Closest relative type* | *Characteristic of family relatives by agestrata of index patients* | *Closest family relatives of brain disorder patients* | *Comparison relative* |
|---|---|---|---|---|
| Children and young people (0-24 years) | Fathers | Number of persons, n | 296 (100.0) | 296 (100.0) |
| | | Male, n (%) | 296 (100.0) | 296 (100.0) |
| | | Age, median (Q1;Q3) | 49.0 (43.4;54.3) | 49.1 (43.7;53.7) |
| | | CCI, n (%) | | |
| | | 0 | 251 (84.8) | 263 (88.9) |
| | | 1-2 | * | 27 (9.1) |
| | | +3 | * | 6 (2.0) |
| | | Education level, n (%) | | |
| | | No/Unknown education | 7 (2.4) | 7 (2.4) |
| | | Primary | 74 (25.0) | 74 (25.0) |
| | | Secondary | 146 (49.3) | 146 (49.3) |
| | | Tertiary | 69 (23.3) | 69 (23.3) |
| | | Relation to patient, n (%) | | |
| | | Father | 296 (100.0) | 296 (100.0) |
| Children and young people (0-24 years) | Mothers | Number of persons, n | 309 (100.0) | 309 (100.0) |
| | | Male, n (%) | 0 (0.0) | 0 (0.0) |
| | | Age, median (Q1;Q3) | 45.9 (40.7;51.2) | 45.9 (40.8;51.0) |
| | | CCI, n (%) | | |
| | | 0 | 272 (88.0) | 273 (88.3) |
| | | 1-2 | 37 (12.0) | 30 (9.7) |
| | | +3 | 0 (0.0) | 6 (1.9) |
| | | Education level, n (%) | | |
| | | No/Unknown education | * | * |
| | | Primary | * | * |
| | | Secondary | 133 (43.0) | 133 (43.0) |
| | | Tertiary | 123 (39.8) | 123 (39.8) |
| | | Relation to patient, n (%) | | |
| | | Mother | 309 (100.0) | 309 (100.0) |
| Young adults and adults (25-64 years) | Children | Number of persons, n | 270 (100.0) | 270 (100.0) |
| | | Male, n (%) | 134 (49.6) | 134 (49.6) |
| | | Age, median (Q1;Q3) | 27.8 (18.6;33.7) | 27.3 (19.2;33.5) |
| | | CCI, n (%) | | |
| | | 0 | 250 (92.6) | 250 (92.6) |
| | | 1-2 | * | 20 (7.4) |
| | | +3 | * | 0 (0.0) |
| | | Education level, n (%) | | |
| | | No/Unknown education | 30 (11.1) | 30 (11.1) |
| | | Primary | 94 (34.8) | 94 (34.8) |
| | | Secondary | 91 (33.7) | 91 (33.7) |
| | | Tertiary | 55 (20.4) | 55 (20.4) |
| | | Relation to patient, n (%) | | |
| | | Child | 270 (100.0) | 270 (100.0) |
| Young adults and adults (25-64 years) | Parents | Number of persons, n | 276 (100.0) | 276 (100.0) |
| | | Male, n (%) | 72 (26.1) | 72 (26.1) |
| | | Age, median (Q1;Q3) | 67.7 (60.8;75.2) | 67.9 (60.2;75.8) |

**Supplementary Table 207**
**Other neurodegenerative disorders - Prevalent cohort 2021**
**Patient characteristics - relatives - not pooled**

| *Age strata of index patients* | *Closest relative type* | *Characteristic of family relatives by agestrata of index patients* | *Closest family relatives of brain disorder patients* | *Comparison relative* |
|---|---|---|---|---|
| | | CCI, n (%) | | |
| | | 0 | 174 (63.0) | 184 (66.7) |
| | | 1-2 | 80 (29.0) | 73 (26.4) |
| | | +3 | 22 (8.0) | 19 (6.9) |
| | | Education level, n (%) | | |
| | | No/Unknown education | 8 (2.9) | 8 (2.9) |
| | | Primary | 83 (30.1) | 83 (30.1) |
| | | Secondary | 129 (46.7) | 129 (46.7) |
| | | Tertiary | 56 (20.3) | 56 (20.3) |
| | | Relation to patient, n (%) | | |
| | | Mother | 204 (73.9) | 204 (73.9) |
| | | Father | 72 (26.1) | 72 (26.1) |
| Young adults and adults (25-64 years) | Partners | Number of persons, n | 779 (100.0) | 779 (100.0) |
| | | Male, n (%) | 349 (44.8) | 349 (44.8) |
| | | Age, median (Q1;Q3) | 53.5 (43.8;59.5) | 53.7 (44.8;59.3) |
| | | CCI, n (%) | | |
| | | 0 | 630 (80.9) | 649 (83.3) |
| | | 1-2 | 127 (16.3) | 108 (13.9) |
| | | +3 | 22 (2.8) | 22 (2.8) |
| | | Education level, n (%) | | |
| | | No/Unknown education | 26 (3.3) | 26 (3.3) |
| | | Primary | 149 (19.1) | 149 (19.1) |
| | | Secondary | 395 (50.7) | 395 (50.7) |
| | | Tertiary | 209 (26.8) | 209 (26.8) |
| | | Relation to patient, n (%) | | |
| | | Partner | 779 (100.0) | 779 (100.0) |
| Young adults and adults (25-64 years) | Siblings | Number of persons, n | 137 (100.0) | 137 (100.0) |
| | | Male, n (%) | 80 (58.4) | 80 (58.4) |
| | | Age, median (Q1;Q3) | 55.3 (46.5;59.6) | 54.9 (46.4;59.6) |
| | | CCI, n (%) | | |
| | | 0 | 115 (83.9) | 120 (87.6) |
| | | 1-2 | * | * |
| | | +3 | * | * |
| | | Education level, n (%) | | |
| | | No/Unknown education | * | * |
| | | Primary | 33 (24.1) | 33 (24.1) |
| | | Secondary | 83 (60.6) | 83 (60.6) |
| | | Tertiary | * | * |
| | | Relation to patient, n (%) | | |
| | | Sibling | 137 (100.0) | 137 (100.0) |
| Older people (65+ years) | Children | Number of persons, n | 568 (100.0) | 568 (100.0) |
| | | Male, n (%) | 292 (51.4) | 292 (51.4) |
| | | Age, median (Q1;Q3) | 50.6 (44.7;55.9) | 50.0 (44.5;55.5) |
| | | CCI, n (%) | | |
| | | 0 | 473 (83.3) | 490 (86.3) |

**Supplementary Table 207**
**Other neurodegenerative disorders - Prevalent cohort 2021**
**Patient characteristics - relatives - not pooled**

| *Age strata of index patients* | *Closest relative type* | *Characteristic of family relatives by agestrata of index patients* | *Closest family relatives of brain disorder patients* | *Comparison relative* |
|---|---|---|---|---|
| | | 1-2 | 85 (15.0) | 69 (12.1) |
| | | +3 | 10 (1.8) | 9 (1.6) |
| | | Education level, n (%) | | |
| | | No/Unknown education | 0 (0.0) | 0 (0.0) |
| | | Primary | 99 (17.4) | 99 (17.4) |
| | | Secondary | 280 (49.3) | 280 (49.3) |
| | | Tertiary | 189 (33.3) | 189 (33.3) |
| | | Relation to patient, n (%) | | |
| | | Child | 568 (100.0) | 568 (100.0) |
| Older people (65+ years) | Partners | Number of persons, n | 938 (100.0) | 938 (100.0) |
| | | Male, n (%) | 371 (39.6) | 371 (39.6) |
| | | Age, median (Q1;Q3) | 72.9 (68.3;77.6) | 72.9 (68.4;77.3) |
| | | CCI, n (%) | | |
| | | 0 | 586 (62.5) | 595 (63.4) |
| | | 1-2 | 271 (28.9) | 273 (29.1) |
| | | +3 | 81 (8.6) | 70 (7.5) |
| | | Education level, n (%) | | |
| | | No/Unknown education | 63 (6.7) | 63 (6.7) |
| | | Primary | 222 (23.7) | 222 (23.7) |
| | | Secondary | 422 (45.0) | 422 (45.0) |
| | | Tertiary | 231 (24.6) | 231 (24.6) |
| | | Relation to patient, n (%) | | |
| | | Partner | 938 (100.0) | 938 (100.0) |
| Older people (65+ years) | Siblings | Number of persons, n | 21 (100.0) | 21 (100.0) |
| | | Male, n (%) | 10 (47.6) | 10 (47.6) |
| | | Age, median (Q1;Q3) | 63.4 (59.7;67.5) | 63.1 (60.7;65.7) |
| | | CCI, n (%) | | |
| | | 0 | * | 14 (66.7) |
| | | 1-2 | * | 7 (33.3) |
| | | +3 | * | 0 (0.0) |
| | | Education level, n (%) | | |
| | | No/Unknown education | 0 (0.0) | 0 (0.0) |
| | | Primary | * | * |
| | | Secondary | * | * |
| | | Tertiary | * | * |
| | | Relation to patient, n (%) | | |
| | | Sibling | 21 (100.0) | 21 (100.0) |

#### Supplementary Table 208
#### Other neurodegenerative disorders - Prevalent cohort 2021
#### Cost analysis - pooled

| | | | | *Actual costs* | | *Attributable costs* | |
|---|---|---|---|---|---|---|---|
| ***Age strata of index patients*** | ***Closest relative type*** | ***Number of index patients included in cost component calculations*** | ***Cost component*** | ***Total (million EUR)*** | ***Per person (EUR)*** | ***Total (million EUR)*** | ***Per person (EUR)*** |
| Children and young people (0-24 years) | Fathers | 296 | Primary sector | 0.1 | 280 | 0.0 | 72 |
| | | 296 | Hospital care | 0.4 | 1,466 | 0.2 | 646 |
| | | 296 | Filled prescriptions | 0.1 | 215 | 0.0 | 99 |
| | | 289 | Lost productivity (income loss) | . | . | 0.9 | 2,995 |
| Children and young people (0-24 years) | Mothers | 309 | Primary sector | 0.1 | 403 | 0.0 | 49 |
| | | 309 | Hospital care | 1.0 | 3,134 | 0.5 | 1,667 |
| | | 309 | Filled prescriptions | 0.1 | 244 | 0.0 | 41 |
| | | 308 | Lost productivity (income loss) | . | . | 0.8 | 2,448 |
| Young adults and adults (25-64 years) | Pooled | 1,462 | Primary sector | 0.5 | 343 | 0.0 | 10 |
| | | 1,462 | Hospital care | 3.8 | 2,606 | 0.1 | 38 |
| | | 1,462 | Filled prescriptions | 0.4 | 292 | 0.0 | 22 |
| | | 1,083 | Lost productivity (income loss) | . | . | 0.3 | 239 |
| Older people (65+ years) | Pooled | 1,527 | Primary sector | 0.7 | 453 | 0.1 | 34 |
| | | 1,527 | Hospital care | 5.1 | 3,386 | 0.3 | 170 |
| | | 1,527 | Filled prescriptions | 0.6 | 423 | 0.1 | 66 |
| | | 596 | Lost productivity (income loss) | . | . | 0.2 | 340 |

#### Supplementary Table 209
#### Other neurodegenerative disorders - Prevalent cohort 2021
#### Cost analysis - not pooled

| *Age strata of index patients* | *Closest relative type* | *Number of index patients included in cost component calculations* | *Cost component* | *Actual costs* Total (million EUR) | *Actual costs* Per person (EUR) | *Attributable costs* Total (million EUR) | *Attributable costs* Per person (EUR) |
|---|---|---|---|---|---|---|---|
| Children and young people (0-24 years) | Fathers | 296 | Primary sector | 0.1 | 280 | 0.0 | 72 |
| | | 296 | Hospital care | 0.4 | 1,466 | 0.2 | 646 |
| | | 296 | Filled prescriptions | 0.1 | 215 | 0.0 | 99 |
| | | 289 | Lost productivity (income loss) | . | . | 0.9 | 2,995 |
| Children and young people (0-24 years) | Mothers | 309 | Primary sector | 0.1 | 403 | 0.0 | 49 |
| | | 309 | Hospital care | 1.0 | 3,134 | 0.5 | 1,667 |
| | | 309 | Filled prescriptions | 0.1 | 244 | 0.0 | 41 |
| | | 308 | Lost productivity (income loss) | . | . | 0.8 | 2,448 |
| Young adults and adults (25-64 years) | Children | 270 | Primary sector | 0.1 | 258 | -0.0 | -27 |
| | | 270 | Hospital care | 0.4 | 1,582 | -0.5 | -1,698 |
| | | 270 | Filled prescriptions | 0.0 | 145 | 0.0 | 49 |
| | | 198 | Lost productivity (income loss) | . | . | 0.1 | 725 |
| Young adults and adults (25-64 years) | Parents | 276 | Primary sector | 0.1 | 466 | -0.0 | -18 |
| | | 276 | Hospital care | 1.0 | 3,620 | 0.1 | 282 |
| | | 276 | Filled prescriptions | 0.1 | 452 | -0.0 | -52 |
| | | 87 | Lost productivity (income loss) | . | . | -1.0 | -11,906 |
| Young adults and adults (25-64 years) | Partners | 779 | Primary sector | 0.3 | 330 | 0.0 | 25 |
| | | 779 | Hospital care | 2.1 | 2,693 | 0.3 | 383 |
| | | 779 | Filled prescriptions | 0.2 | 293 | 0.0 | 32 |
| | | 677 | Lost productivity (income loss) | . | . | 1.6 | 2,394 |
| Young adults and adults (25-64 years) | Siblings | 137 | Primary sector | 0.0 | 339 | 0.0 | 59 |
| | | 137 | Hospital care | 0.3 | 2,131 | 0.1 | 1,013 |
| | | 137 | Filled prescriptions | 0.0 | 257 | 0.0 | 57 |
| | | 121 | Lost productivity (income loss) | . | . | -0.5 | -3,882 |
| Older people (65+ years) | Children | 568 | Primary sector | 0.2 | 323 | 0.0 | 30 |
| | | 568 | Hospital care | 1.3 | 2,358 | 0.2 | 433 |
| | | 568 | Filled prescriptions | 0.2 | 272 | 0.0 | 74 |
| | | 539 | Lost productivity (income loss) | . | . | 0.7 | 1,314 |
| Older people (65+ years) | Partners | 938 | Primary sector | 0.5 | 533 | 0.0 | 35 |
| | | 938 | Hospital care | 3.6 | 3,904 | -0.1 | -126 |
| | | 938 | Filled prescriptions | 0.5 | 514 | 0.1 | 55 |
| | | 48 | Lost productivity (income loss) | . | . | -0.7 | -14,234 |
| Older people (65+ years) | Siblings | 21 | Primary sector | 0.0 | 417 | 0.0 | 47 |
| | | 21 | Hospital care | 0.2 | 8,740 | 0.1 | 6,472 |
| | | 21 | Filled prescriptions | 0.0 | 506 | 0.0 | 360 |
| | | 9 | Lost productivity (income loss) | . | . | 0.2 | 19,727 |

### Supplementary Table 210
#### Parkinson's disease - Incident cohort 2016-2021
#### Identification of populations of relatives

| *Agegroup of index patients in matched population* | | *Closest relative type identified* | | *Available control with same type of closest relative* |
|---|---|---|---|---|
| Children and young people (0-24 years) | n = 113 | Fathers | n/a | n/a |
| | | Mothers | n/a | n/a |
| | | No available closest relative | n/a | n/a |
| Young adults and adults (25-64 years) | n = 3,071 | Children | n = 375 (12%) | 91% |
| | | Parents | n = 291 (9%) | 88% |
| | | Partners | n = 2,044 (67%) | 96% |
| | | Siblings | n = 281 (9%) | 62% |
| | | Pooled | n = 2,991 (97%) | 92% |
| | | No available closest relative | n = 80 (3%) | n/a |
| Older people (65+ years) | n = 10,502 | Children | n = 2,998 (29%) | 95% |
| | | Partners | n = 6,612 (63%) | 96% |
| | | Siblings | n = 99 (1%) | 32% |
| | | Pooled | n = 9,709 (92%) | 95% |
| | | No available closest relative | n = 793 (8%) | n/a |

### Supplementary Table 211
### Parkinson's disease - Incident cohort 2016-2021
### Patient characteristics - cases

| *Age strata of index patients* | *Characteristic* | |
|---|---|---|
| Children and young people (0-24 years) | Number of persons, n | 104 (100.0) |
| | Male, n (%) | 51 (49.0) |
| | Age, median (Q1;Q3) | 17.8 (11.1;21.2) |
| | CCI, n (%) | |
| | 0 | 83 (79.8) |
| | 1-2 | * |
| | +3 | * |
| | Education level, n (%) | |
| | No/Unknown education | 31 (29.8) |
| | Primary | 56 (53.8) |
| | Secondary | * |
| | Tertiary | * |
| Young adults and adults (25-64 years) | Number of persons, n | 2,752 (100.0) |
| | Male, n (%) | 1,404 (51.0) |
| | Age, median (Q1;Q3) | 56.7 (48.8;61.4) |
| | CCI, n (%) | |
| | 0 | 1,967 (71.5) |
| | 1-2 | 650 (23.6) |
| | +3 | 135 (4.9) |
| | Education level, n (%) | |
| | No/Unknown education | 86 (3.1) |
| | Primary | 597 (21.7) |
| | Secondary | 1,356 (49.3) |
| | Tertiary | 713 (25.9) |
| Older people (65+ years) | Number of persons, n | 9,297 (100.0) |
| | Male, n (%) | 5,586 (60.1) |
| | Age, median (Q1;Q3) | 76.2 (71.9;80.9) |
| | CCI, n (%) | |
| | 0 | 4,542 (48.9) |
| | 1-2 | 3,458 (37.2) |
| | +3 | 1,297 (14.0) |
| | Education level, n (%) | |
| | No/Unknown education | 775 (8.3) |
| | Primary | 2,574 (27.7) |
| | Secondary | 4,147 (44.6) |
| | Tertiary | 1,801 (19.4) |

**Supplementary Table 212**
**Parkinson's disease - Incident cohort 2016-2021**
**Patient characteristics - relatives - pooled**

| *Age strata of index patients* | *Closest relative type* | *Characteristic of family relatives by agestrata of index patients* | *Closest family relatives of brain disorder patients* | *Comparison relative* |
|---|---|---|---|---|
| Children and young people (0-24 years) | Fathers | Number of persons, n | 95 (100.0) | 95 (100.0) |
| | | Male, n (%) | 95 (100.0) | 95 (100.0) |
| | | Age, median (Q1;Q3) | 48.4 (42.0;52.4) | 48.9 (42.8;52.4) |
| | | CCI, n (%) | | |
| | | 0 | 83 (87.4) | 83 (87.4) |
| | | 1-2 | 12 (12.6) | * |
| | | +3 | 0 (0.0) | * |
| | | Education level, n (%) | | |
| | | No/Unknown education | * | * |
| | | Primary | 21 (22.1) | 21 (22.1) |
| | | Secondary | 53 (55.8) | 53 (55.8) |
| | | Tertiary | * | * |
| | | Relation to patient, n (%) | | |
| | | Father | 95 (100.0) | 95 (100.0) |
| Children and young people (0-24 years) | Mothers | Number of persons, n | 98 (100.0) | 98 (100.0) |
| | | Male, n (%) | 0 (0.0) | 0 (0.0) |
| | | Age, median (Q1;Q3) | 46.7 (40.6;51.4) | 47.3 (41.4;50.6) |
| | | CCI, n (%) | | |
| | | 0 | 85 (86.7) | 85 (86.7) |
| | | 1-2 | * | 13 (13.3) |
| | | +3 | * | 0 (0.0) |
| | | Education level, n (%) | | |
| | | No/Unknown education | * | * |
| | | Primary | * | * |
| | | Secondary | 49 (50.0) | 49 (50.0) |
| | | Tertiary | 32 (32.7) | 32 (32.7) |
| | | Relation to patient, n (%) | | |
| | | Mother | 98 (100.0) | 98 (100.0) |
| Young adults and adults (25-64 years) | Pooled | Number of persons, n | 2,752 (100.0) | 2,752 (100.0) |
| | | Male, n (%) | 1,287 (46.8) | 1,287 (46.8) |
| | | Age, median (Q1;Q3) | 55.5 (45.5;61.7) | 55.7 (45.7;61.7) |
| | | CCI, n (%) | | |
| | | 0 | 2,234 (81.2) | 2,212 (80.4) |
| | | 1-2 | 434 (15.8) | 459 (16.7) |
| | | +3 | 84 (3.1) | 81 (2.9) |
| | | Education level, n (%) | | |
| | | No/Unknown education | 96 (3.5) | 96 (3.5) |
| | | Primary | 660 (24.0) | 660 (24.0) |
| | | Secondary | 1,331 (48.4) | 1,331 (48.4) |
| | | Tertiary | 665 (24.2) | 665 (24.2) |
| | | Relation to patient, n (%) | | |
| | | Mother | 210 (7.6) | 210 (7.6) |
| | | Father | 47 (1.7) | 47 (1.7) |
| | | Partner | 1,974 (71.7) | 1,974 (71.7) |
| | | Child | 344 (12.5) | 344 (12.5) |

### Supplementary Table 212
### Parkinson's disease - Incident cohort 2016-2021
### Patient characteristics - relatives - pooled

| *Age strata of index patients* | *Closest relative type* | *Characteristic of family relatives by agestrata of index patients* | *Closest family relatives of brain disorder patients* | *Comparison relative* |
|---|---|---|---|---|
| | | Sibling | 177 (6.4) | 177 (6.4) |
| Older people (65+ years) | Pooled | Number of persons, n | 9,297 (100.0) | 9,297 (100.0) |
| | | Male, n (%) | 3,486 (37.5) | 3,486 (37.5) |
| | | Age, median (Q1;Q3) | 69.9 (58.0;76.2) | 70.0 (58.1;76.1) |
| | | CCI, n (%) | | |
| | | 0 | 6,421 (69.1) | 6,429 (69.2) |
| | | 1-2 | 2,216 (23.8) | 2,233 (24.0) |
| | | +3 | 660 (7.1) | 635 (6.8) |
| | | Education level, n (%) | | |
| | | No/Unknown education | 387 (4.2) | 387 (4.2) |
| | | Primary | 2,071 (22.3) | 2,071 (22.3) |
| | | Secondary | 4,413 (47.5) | 4,413 (47.5) |
| | | Tertiary | 2,426 (26.1) | 2,426 (26.1) |
| | | Relation to patient, n (%) | | |
| | | Partner | 6,400 (68.8) | 6,400 (68.8) |
| | | Child | 2,865 (30.8) | 2,865 (30.8) |
| | | Sibling | 32 (0.3) | 32 (0.3) |

**Supplementary Table 213**
**Parkinson's disease - Incident cohort 2016-2021**
**Patient characteristics - relatives - not pooled**

| *Age strata of index patients* | *Closest relative type* | *Characteristic of family relatives by agestrata of index patients* | *Closest family relatives of brain disorder patients* | *Comparison relative* |
|---|---|---|---|---|
| Children and young people (0-24 years) | Fathers | Number of persons, n | 95 (100.0) | 95 (100.0) |
| | | Male, n (%) | 95 (100.0) | 95 (100.0) |
| | | Age, median (Q1;Q3) | 48.4 (42.0;52.4) | 48.9 (42.8;52.4) |
| | | CCI, n (%) | | |
| | | 0 | 83 (87.4) | 83 (87.4) |
| | | 1-2 | 12 (12.6) | * |
| | | +3 | 0 (0.0) | * |
| | | Education level, n (%) | | |
| | | No/Unknown education | * | * |
| | | Primary | 21 (22.1) | 21 (22.1) |
| | | Secondary | 53 (55.8) | 53 (55.8) |
| | | Tertiary | * | * |
| | | Relation to patient, n (%) | | |
| | | Father | 95 (100.0) | 95 (100.0) |
| Children and young people (0-24 years) | Mothers | Number of persons, n | 98 (100.0) | 98 (100.0) |
| | | Male, n (%) | 0 (0.0) | 0 (0.0) |
| | | Age, median (Q1;Q3) | 46.7 (40.6;51.4) | 47.3 (41.4;50.6) |
| | | CCI, n (%) | | |
| | | 0 | 85 (86.7) | 85 (86.7) |
| | | 1-2 | * | 13 (13.3) |
| | | +3 | * | 0 (0.0) |
| | | Education level, n (%) | | |
| | | No/Unknown education | * | * |
| | | Primary | * | * |
| | | Secondary | 49 (50.0) | 49 (50.0) |
| | | Tertiary | 32 (32.7) | 32 (32.7) |
| | | Relation to patient, n (%) | | |
| | | Mother | 98 (100.0) | 98 (100.0) |
| Young adults and adults (25-64 years) | Children | Number of persons, n | 344 (100.0) | 344 (100.0) |
| | | Male, n (%) | 179 (52.0) | 179 (52.0) |
| | | Age, median (Q1;Q3) | 27.9 (19.6;34.7) | 28.1 (19.2;34.7) |
| | | CCI, n (%) | | |
| | | 0 | 317 (92.2) | 322 (93.6) |
| | | 1-2 | * | * |
| | | +3 | * | * |
| | | Education level, n (%) | | |
| | | No/Unknown education | 37 (10.8) | 37 (10.8) |
| | | Primary | 120 (34.9) | 120 (34.9) |
| | | Secondary | 109 (31.7) | 109 (31.7) |
| | | Tertiary | 78 (22.7) | 78 (22.7) |
| | | Relation to patient, n (%) | | |
| | | Child | 344 (100.0) | 344 (100.0) |
| Young adults and adults (25-64 years) | Parents | Number of persons, n | 257 (100.0) | 257 (100.0) |
| | | Male, n (%) | 47 (18.3) | 47 (18.3) |
| | | Age, median (Q1;Q3) | 71.7 (63.8;80.5) | 71.9 (65.1;80.5) |

**Supplementary Table 213**
**Parkinson's disease - Incident cohort 2016-2021**
**Patient characteristics - relatives - not pooled**

| *Age strata of index patients* | *Closest relative type* | *Characteristic of family relatives by agestrata of index patients* | *Closest family relatives of brain disorder patients* | *Comparison relative* |
|---|---|---|---|---|
| | | CCI, n (%) | | |
| | | 0 | 161 (62.6) | 138 (53.7) |
| | | 1-2 | 76 (29.6) | 90 (35.0) |
| | | +3 | 20 (7.8) | 29 (11.3) |
| | | Education level, n (%) | | |
| | | No/Unknown education | * | * |
| | | Primary | 118 (45.9) | 118 (45.9) |
| | | Secondary | 101 (39.3) | 101 (39.3) |
| | | Tertiary | * | * |
| | | Relation to patient, n (%) | | |
| | | Mother | 210 (81.7) | 210 (81.7) |
| | | Father | 47 (18.3) | 47 (18.3) |
| Young adults and adults (25-64 years) | Partners | Number of persons, n | 1,974 (100.0) | 1,974 (100.0) |
| | | Male, n (%) | 954 (48.3) | 954 (48.3) |
| | | Age, median (Q1;Q3) | 56.0 (49.2;61.2) | 56.2 (49.0;61.2) |
| | | CCI, n (%) | | |
| | | 0 | 1,611 (81.6) | 1,612 (81.7) |
| | | 1-2 | 310 (15.7) | 316 (16.0) |
| | | +3 | 53 (2.7) | 46 (2.3) |
| | | Education level, n (%) | | |
| | | No/Unknown education | 55 (2.8) | 55 (2.8) |
| | | Primary | 391 (19.8) | 391 (19.8) |
| | | Secondary | 1,015 (51.4) | 1,015 (51.4) |
| | | Tertiary | 513 (26.0) | 513 (26.0) |
| | | Relation to patient, n (%) | | |
| | | Partner | 1,974 (100.0) | 1,974 (100.0) |
| Young adults and adults (25-64 years) | Siblings | Number of persons, n | 177 (100.0) | 177 (100.0) |
| | | Male, n (%) | 107 (60.5) | 107 (60.5) |
| | | Age, median (Q1;Q3) | 57.3 (52.2;61.7) | 57.7 (52.1;61.4) |
| | | CCI, n (%) | | |
| | | 0 | 145 (81.9) | 140 (79.1) |
| | | 1-2 | 23 (13.0) | * |
| | | +3 | 9 (5.1) | * |
| | | Education level, n (%) | | |
| | | No/Unknown education | 0 (0.0) | 0 (0.0) |
| | | Primary | 31 (17.5) | 31 (17.5) |
| | | Secondary | 106 (59.9) | 106 (59.9) |
| | | Tertiary | 40 (22.6) | 40 (22.6) |
| | | Relation to patient, n (%) | | |
| | | Sibling | 177 (100.0) | 177 (100.0) |
| Older people (65+ years) | Children | Number of persons, n | 2,865 (100.0) | 2,865 (100.0) |
| | | Male, n (%) | 1,445 (50.4) | 1,445 (50.4) |
| | | Age, median (Q1;Q3) | 52.8 (47.6;57.4) | 52.9 (47.6;57.2) |
| | | CCI, n (%) | | |
| | | 0 | 2,436 (85.0) | 2,461 (85.9) |

**Supplementary Table 213**
**Parkinson's disease - Incident cohort 2016-2021**
**Patient characteristics - relatives - not pooled**

| *Age strata of index patients* | *Closest relative type* | *Characteristic of family relatives by agestrata of index patients* | *Closest family relatives of brain disorder patients* | *Comparison relative* |
|---|---|---|---|---|
| | | 1-2 | 377 (13.2) | 362 (12.6) |
| | | +3 | 52 (1.8) | 42 (1.5) |
| | | Education level, n (%) | | |
| | | No/Unknown education | 8 (0.3) | 8 (0.3) |
| | | Primary | 378 (13.2) | 378 (13.2) |
| | | Secondary | 1,460 (51.0) | 1,460 (51.0) |
| | | Tertiary | 1,019 (35.6) | 1,019 (35.6) |
| | | Relation to patient, n (%) | | |
| | | Child | 2,865 (100.0) | 2,865 (100.0) |
| Older people (65+ years) | Partners | Number of persons, n | 6,400 (100.0) | 6,400 (100.0) |
| | | Male, n (%) | 2,022 (31.6) | 2,022 (31.6) |
| | | Age, median (Q1;Q3) | 73.8 (69.4;78.3) | 73.8 (69.5;78.3) |
| | | CCI, n (%) | | |
| | | 0 | 3,966 (62.0) | 3,947 (61.7) |
| | | 1-2 | 1,830 (28.6) | 1,862 (29.1) |
| | | +3 | 604 (9.4) | 591 (9.2) |
| | | Education level, n (%) | | |
| | | No/Unknown education | 379 (5.9) | 379 (5.9) |
| | | Primary | 1,684 (26.3) | 1,684 (26.3) |
| | | Secondary | 2,938 (45.9) | 2,938 (45.9) |
| | | Tertiary | 1,399 (21.9) | 1,399 (21.9) |
| | | Relation to patient, n (%) | | |
| | | Partner | 6,400 (100.0) | 6,400 (100.0) |
| Older people (65+ years) | Siblings | Number of persons, n | 32 (100.0) | 32 (100.0) |
| | | Male, n (%) | 19 (59.4) | 19 (59.4) |
| | | Age, median (Q1;Q3) | 62.8 (60.9;66.9) | 64.6 (61.4;66.6) |
| | | CCI, n (%) | | |
| | | 0 | 19 (59.4) | 21 (65.6) |
| | | 1-2 | * | * |
| | | +3 | * | * |
| | | Education level, n (%) | | |
| | | No/Unknown education | 0 (0.0) | 0 (0.0) |
| | | Primary | 9 (28.1) | 9 (28.1) |
| | | Secondary | 15 (46.9) | 15 (46.9) |
| | | Tertiary | 8 (25.0) | 8 (25.0) |
| | | Relation to patient, n (%) | | |
| | | Sibling | 32 (100.0) | 32 (100.0) |

#### Supplementary Table 214
#### Parkinson's disease - Incident cohort 2016-2021
#### Cost analysis - pooled

| *Age strata of index patients* | *Closest relative type* | *Number of index patients included in cost component calculations* | *Cost component* | *Actual costs* | | *Attributable costs* | |
|---|---|---|---|---|---|---|---|
| | | | | *Total (million EUR)* | *Per person (EUR)* | *Total (million EUR)* | *Per person (EUR)* |
| Children and young people (0-24 years) | Fathers | 95 | Primary sector | 0.0 | 222 | 0.0 | 16 |
| | | 95 | Hospital care | 0.2 | 2,151 | 0.1 | 1,490 |
| | | 95 | Filled prescriptions | 0.0 | 204 | 0.0 | 45 |
| | | 94 | Lost productivity (income loss) | . | . | 0.0 | 116 |
| Children and young people (0-24 years) | Mothers | 98 | Primary sector | 0.0 | 362 | 0.0 | 15 |
| | | 98 | Hospital care | 0.1 | 1,459 | 0.1 | 563 |
| | | 98 | Filled prescriptions | 0.0 | 207 | 0.0 | 21 |
| | | 98 | Lost productivity (income loss) | . | . | -0.5 | -5,522 |
| Young adults and adults (25-64 years) | Pooled | 2,752 | Primary sector | 0.9 | 344 | 0.1 | 22 |
| | | 2,752 | Hospital care | 6.0 | 2,197 | 1.2 | 448 |
| | | 2,752 | Filled prescriptions | 0.8 | 283 | 0.0 | 16 |
| | | 2,056 | Lost productivity (income loss) | . | . | 6.1 | 2,962 |
| Older people (65+ years) | Pooled | 9,297 | Primary sector | 4.3 | 461 | 0.2 | 23 |
| | | 9,297 | Hospital care | 29.4 | 3,190 | 0.8 | 86 |
| | | 9,297 | Filled prescriptions | 4.0 | 435 | 0.2 | 26 |
| | | 2,919 | Lost productivity (income loss) | . | . | 4.8 | 1,654 |

**Supplementary Table 215**
**Parkinson's disease - Incident cohort 2016-2021**
**Cost analysis - not pooled**

| *Age strata of index patients* | *Closest relative type* | *Number of index patients included in cost component calculations* | *Cost component* | *Actual costs* Total (million EUR) | *Actual costs* Per person (EUR) | *Attributable costs* Total (million EUR) | *Attributable costs* Per person (EUR) |
|---|---|---|---|---|---|---|---|
| Children and young people (0-24 years) | Fathers | 95 | Primary sector | 0.0 | 222 | 0.0 | 16 |
| | | 95 | Hospital care | 0.2 | 2,151 | 0.1 | 1,490 |
| | | 95 | Filled prescriptions | 0.0 | 204 | 0.0 | 45 |
| | | 94 | Lost productivity (income loss) | . | . | 0.0 | 116 |
| Children and young people (0-24 years) | Mothers | 98 | Primary sector | 0.0 | 362 | 0.0 | 15 |
| | | 98 | Hospital care | 0.1 | 1,459 | 0.1 | 563 |
| | | 98 | Filled prescriptions | 0.0 | 207 | 0.0 | 21 |
| | | 98 | Lost productivity (income loss) | . | . | -0.5 | -5,522 |
| Young adults and adults (25-64 years) | Children | 344 | Primary sector | 0.1 | 261 | 0.0 | 28 |
| | | 344 | Hospital care | 0.5 | 1,368 | 0.2 | 719 |
| | | 344 | Filled prescriptions | 0.0 | 129 | 0.0 | 31 |
| | | 267 | Lost productivity (income loss) | . | . | 1.0 | 3,584 |
| Young adults and adults (25-64 years) | Parents | 257 | Primary sector | 0.1 | 469 | 0.0 | 4 |
| | | 257 | Hospital care | 1.0 | 3,855 | 0.2 | 866 |
| | | 257 | Filled prescriptions | 0.1 | 577 | 0.0 | 87 |
| | | 54 | Lost productivity (income loss) | . | . | -0.0 | -822 |
| Young adults and adults (25-64 years) | Partners | 1,974 | Primary sector | 0.7 | 344 | 0.1 | 29 |
| | | 1,974 | Hospital care | 4.2 | 2,128 | 0.8 | 417 |
| | | 1,974 | Filled prescriptions | 0.5 | 271 | 0.0 | 4 |
| | | 1,596 | Lost productivity (income loss) | . | . | 5.3 | 3,315 |
| Young adults and adults (25-64 years) | Siblings | 177 | Primary sector | 0.1 | 323 | -0.0 | -39 |
| | | 177 | Hospital care | 0.4 | 2,191 | -0.1 | -327 |
| | | 177 | Filled prescriptions | 0.1 | 299 | 0.0 | 9 |
| | | 139 | Lost productivity (income loss) | . | . | -0.1 | -822 |
| Older people (65+ years) | Children | 2,865 | Primary sector | 0.9 | 302 | 0.0 | 16 |
| | | 2,865 | Hospital care | 5.1 | 1,781 | 0.7 | 229 |
| | | 2,865 | Filled prescriptions | 0.7 | 249 | 0.1 | 35 |
| | | 2,673 | Lost productivity (income loss) | . | . | 6.1 | 2,278 |
| Older people (65+ years) | Partners | 6,400 | Primary sector | 3.4 | 534 | 0.2 | 26 |
| | | 6,400 | Hospital care | 24.1 | 3,817 | 0.2 | 26 |
| | | 6,400 | Filled prescriptions | 3.3 | 520 | 0.1 | 22 |
| | | 235 | Lost productivity (income loss) | . | . | -1.2 | -5,146 |
| Older people (65+ years) | Siblings | 32 | Primary sector | 0.0 | 319 | -0.0 | -47 |
| | | 32 | Hospital care | 0.2 | 5,110 | -0.0 | -646 |
| | | 32 | Filled prescriptions | 0.0 | 218 | -0.0 | -98 |
| | | 11 | Lost productivity (income loss) | . | . | -0.1 | -4,754 |

**Supplementary Table 216**
**Parkinson's disease - Prevalent cohort 2021**
**Identification of populations of relatives**

| ***Agegroup of index patients in matched population*** | | ***Closest relative type identified*** | | ***Available control with same type of closest relative*** |
|---|---|---|---|---|
| Children and young people (0-24 years) | n = 201 | Fathers | n = 174 (87%) | 93% |
| | | Mothers | n = 180 (90%) | 93% |
| | | No available closest relative | n = 8 (4%) | n/a |
| Young adults and adults (25-64 years) | n = 5,356 | Children | n = 791 (15%) | 92% |
| | | Parents | n = 598 (11%) | 88% |
| | | Partners | n = 3,314 (62%) | 96% |
| | | Siblings | n = 557 (10%) | 56% |
| | | Pooled | n = 5,260 (98%) | 90% |
| | | No available closest relative | n = 96 (2%) | n/a |
| Older people (65+ years) | n = 16,472 | Children | n = 5,920 (36%) | 95% |
| | | Partners | n = 8,950 (54%) | 96% |
| | | Siblings | n = 264 (2%) | 41% |
| | | Pooled | n = 15,134 (92%) | 95% |
| | | No available closest relative | n = 1,338 (8%) | n/a |

**Supplementary Table 217**
**Parkinson's disease - Prevalent cohort 2021**
**Patient characteristics - cases**

| *Age strata of index patients* | *Characteristic* | |
|---|---|---|
| Children and young people (0-24 years) | Number of persons, n | 181 (100.0) |
| | Male, n (%) | 90 (49.7) |
| | Age, median (Q1;Q3) | 19.9 (15.5;22.8) |
| | CCI, n (%) | |
| | 0 | 146 (80.7) |
| | 1-2 | 29 (16.0) |
| | +3 | 6 (3.3) |
| | Education level, n (%) | |
| | No/Unknown education | 36 (19.9) |
| | Primary | 117 (64.6) |
| | Secondary | * |
| | Tertiary | * |
| Young adults and adults (25-64 years) | Number of persons, n | 4,767 (100.0) |
| | Male, n (%) | 2,131 (44.7) |
| | Age, median (Q1;Q3) | 56.0 (48.3;61.1) |
| | CCI, n (%) | |
| | 0 | 3,369 (70.7) |
| | 1-2 | 1,158 (24.3) |
| | +3 | 240 (5.0) |
| | Education level, n (%) | |
| | No/Unknown education | 149 (3.1) |
| | Primary | 1,205 (25.3) |
| | Secondary | 2,318 (48.6) |
| | Tertiary | 1,095 (23.0) |
| Older people (65+ years) | Number of persons, n | 14,391 (100.0) |
| | Male, n (%) | 7,811 (54.3) |
| | Age, median (Q1;Q3) | 77.4 (72.9;82.4) |
| | CCI, n (%) | |
| | 0 | 6,616 (46.0) |
| | 1-2 | 5,517 (38.3) |
| | +3 | 2,258 (15.7) |
| | Education level, n (%) | |
| | No/Unknown education | 1,037 (7.2) |
| | Primary | 4,358 (30.3) |
| | Secondary | 6,308 (43.8) |
| | Tertiary | 2,688 (18.7) |

**Supplementary Table 218**
**Parkinson's disease - Prevalent cohort 2021**
**Patient characteristics - relatives - pooled**

| *Age strata of index patients* | *Closest relative type* | *Characteristic of family relatives by agestrata of index patients* | *Closest family relatives of brain disorder patients* | *Comparison relative* |
|---|---|---|---|---|
| Children and young people (0-24 years) | Fathers | Number of persons, n | 163 (100.0) | 163 (100.0) |
| | | Male, n (%) | 163 (100.0) | 163 (100.0) |
| | | Age, median (Q1;Q3) | 51.7 (47.2;55.2) | 52.0 (46.8;55.0) |
| | | CCI, n (%) | | |
| | | 0 | 142 (87.1) | 150 (92.0) |
| | | 1-2 | * | 13 (8.0) |
| | | +3 | * | 0 (0.0) |
| | | Education level, n (%) | | |
| | | No/Unknown education | * | * |
| | | Primary | * | * |
| | | Secondary | 90 (55.2) | 90 (55.2) |
| | | Tertiary | 37 (22.7) | 37 (22.7) |
| | | Relation to patient, n (%) | | |
| | | Father | 163 (100.0) | 163 (100.0) |
| Children and young people (0-24 years) | Mothers | Number of persons, n | 168 (100.0) | 168 (100.0) |
| | | Male, n (%) | 0 (0.0) | 0 (0.0) |
| | | Age, median (Q1;Q3) | 49.2 (44.9;53.9) | 50.0 (44.9;52.9) |
| | | CCI, n (%) | | |
| | | 0 | 152 (90.5) | 146 (86.9) |
| | | 1-2 | * | * |
| | | +3 | * | * |
| | | Education level, n (%) | | |
| | | No/Unknown education | * | * |
| | | Primary | * | * |
| | | Secondary | 75 (44.6) | 75 (44.6) |
| | | Tertiary | 63 (37.5) | 63 (37.5) |
| | | Relation to patient, n (%) | | |
| | | Mother | 168 (100.0) | 168 (100.0) |
| Young adults and adults (25-64 years) | Pooled | Number of persons, n | 4,767 (100.0) | 4,767 (100.0) |
| | | Male, n (%) | 2,468 (51.8) | 2,468 (51.8) |
| | | Age, median (Q1;Q3) | 55.3 (43.8;62.0) | 55.3 (43.9;62.0) |
| | | CCI, n (%) | | |
| | | 0 | 3,809 (79.9) | 3,934 (82.5) |
| | | 1-2 | 797 (16.7) | 704 (14.8) |
| | | +3 | 161 (3.4) | 129 (2.7) |
| | | Education level, n (%) | | |
| | | No/Unknown education | 143 (3.0) | 143 (3.0) |
| | | Primary | 1,268 (26.6) | 1,268 (26.6) |
| | | Secondary | 2,317 (48.6) | 2,317 (48.6) |
| | | Tertiary | 1,039 (21.8) | 1,039 (21.8) |
| | | Relation to patient, n (%) | | |
| | | Mother | 416 (8.7) | 416 (8.7) |
| | | Father | 112 (2.3) | 112 (2.3) |
| | | Partner | 3,196 (67.0) | 3,196 (67.0) |
| | | Child | 731 (15.3) | 731 (15.3) |

**Supplementary Table 218**
**Parkinson's disease - Prevalent cohort 2021**
**Patient characteristics - relatives - pooled**

| *Age strata of index patients* | *Closest relative type* | *Characteristic of family relatives by agestrata of index patients* | *Closest family relatives of brain disorder patients* | *Comparison relative* |
|---|---|---|---|---|
| | | Sibling | 312 (6.5) | 312 (6.5) |
| Older people (65+ years) | Pooled | Number of persons, n | 14,391 (100.0) | 14,391 (100.0) |
| | | Male, n (%) | 5,972 (41.5) | 5,972 (41.5) |
| | | Age, median (Q1;Q3) | 68.4 (56.9;76.2) | 68.5 (56.9;76.1) |
| | | CCI, n (%) | | |
| | | 0 | 9,962 (69.2) | 10,079 (70.0) |
| | | 1-2 | 3,402 (23.6) | 3,336 (23.2) |
| | | +3 | 1,027 (7.1) | 976 (6.8) |
| | | Education level, n (%) | | |
| | | No/Unknown education | 479 (3.3) | 479 (3.3) |
| | | Primary | 3,129 (21.7) | 3,129 (21.7) |
| | | Secondary | 6,968 (48.4) | 6,968 (48.4) |
| | | Tertiary | 3,815 (26.5) | 3,815 (26.5) |
| | | Relation to patient, n (%) | | |
| | | Partner | 8,643 (60.1) | 8,643 (60.1) |
| | | Child | 5,638 (39.2) | 5,638 (39.2) |
| | | Sibling | 110 (0.8) | 110 (0.8) |

**Supplementary Table 219**
**Parkinson's disease - Prevalent cohort 2021**
**Patient characteristics - relatives - not pooled**

| *Age strata of index patients* | *Closest relative type* | *Characteristic of family relatives by agestrata of index patients* | *Closest family relatives of brain disorder patients* | *Comparison relative* |
|---|---|---|---|---|
| Children and young people (0-24 years) | Fathers | Number of persons, n | 163 (100.0) | 163 (100.0) |
| | | Male, n (%) | 163 (100.0) | 163 (100.0) |
| | | Age, median (Q1;Q3) | 51.7 (47.2;55.2) | 52.0 (46.8;55.0) |
| | | CCI, n (%) | | |
| | | 0 | 142 (87.1) | 150 (92.0) |
| | | 1-2 | * | 13 (8.0) |
| | | +3 | * | 0 (0.0) |
| | | Education level, n (%) | | |
| | | No/Unknown education | * | * |
| | | Primary | * | * |
| | | Secondary | 90 (55.2) | 90 (55.2) |
| | | Tertiary | 37 (22.7) | 37 (22.7) |
| | | Relation to patient, n (%) | | |
| | | Father | 163 (100.0) | 163 (100.0) |
| Children and young people (0-24 years) | Mothers | Number of persons, n | 168 (100.0) | 168 (100.0) |
| | | Male, n (%) | 0 (0.0) | 0 (0.0) |
| | | Age, median (Q1;Q3) | 49.2 (44.9;53.9) | 50.0 (44.9;52.9) |
| | | CCI, n (%) | | |
| | | 0 | 152 (90.5) | 146 (86.9) |
| | | 1-2 | * | * |
| | | +3 | * | * |
| | | Education level, n (%) | | |
| | | No/Unknown education | * | * |
| | | Primary | * | * |
| | | Secondary | 75 (44.6) | 75 (44.6) |
| | | Tertiary | 63 (37.5) | 63 (37.5) |
| | | Relation to patient, n (%) | | |
| | | Mother | 168 (100.0) | 168 (100.0) |
| Young adults and adults (25-64 years) | Children | Number of persons, n | 731 (100.0) | 731 (100.0) |
| | | Male, n (%) | 381 (52.1) | 381 (52.1) |
| | | Age, median (Q1;Q3) | 27.8 (19.6;34.5) | 27.7 (19.3;34.3) |
| | | CCI, n (%) | | |
| | | 0 | 673 (92.1) | 692 (94.7) |
| | | 1-2 | * | * |
| | | +3 | * | * |
| | | Education level, n (%) | | |
| | | No/Unknown education | 58 (7.9) | 58 (7.9) |
| | | Primary | 318 (43.5) | 318 (43.5) |
| | | Secondary | 223 (30.5) | 223 (30.5) |
| | | Tertiary | 132 (18.1) | 132 (18.1) |
| | | Relation to patient, n (%) | | |
| | | Child | 731 (100.0) | 731 (100.0) |
| Young adults and adults (25-64 years) | Parents | Number of persons, n | 528 (100.0) | 528 (100.0) |
| | | Male, n (%) | 112 (21.2) | 112 (21.2) |
| | | Age, median (Q1;Q3) | 73.5 (64.0;80.1) | 73.0 (64.3;79.8) |

**Supplementary Table 219**
**Parkinson's disease - Prevalent cohort 2021**
**Patient characteristics - relatives - not pooled**

| *Age strata of index patients* | *Closest relative type* | *Characteristic of family relatives by agestrata of index patients* | *Closest family relatives of brain disorder patients* | *Comparison relative* |
|---|---|---|---|---|
| | | CCI, n (%) | | |
| | | 0 | 316 (59.8) | 328 (62.1) |
| | | 1-2 | 159 (30.1) | 152 (28.8) |
| | | +3 | 53 (10.0) | 48 (9.1) |
| | | Education level, n (%) | | |
| | | No/Unknown education | 6 (1.1) | 6 (1.1) |
| | | Primary | 211 (40.0) | 211 (40.0) |
| | | Secondary | 232 (43.9) | 232 (43.9) |
| | | Tertiary | 79 (15.0) | 79 (15.0) |
| | | Relation to patient, n (%) | | |
| | | Mother | 416 (78.8) | 416 (78.8) |
| | | Father | 112 (21.2) | 112 (21.2) |
| Young adults and adults (25-64 years) | Partners | Number of persons, n | 3,196 (100.0) | 3,196 (100.0) |
| | | Male, n (%) | 1,782 (55.8) | 1,782 (55.8) |
| | | Age, median (Q1;Q3) | 56.1 (48.9;61.2) | 55.9 (49.1;61.2) |
| | | CCI, n (%) | | |
| | | 0 | 2,582 (80.8) | 2,674 (83.7) |
| | | 1-2 | 524 (16.4) | 459 (14.4) |
| | | +3 | 90 (2.8) | 63 (2.0) |
| | | Education level, n (%) | | |
| | | No/Unknown education | 76 (2.4) | 76 (2.4) |
| | | Primary | 674 (21.1) | 674 (21.1) |
| | | Secondary | 1,682 (52.6) | 1,682 (52.6) |
| | | Tertiary | 764 (23.9) | 764 (23.9) |
| | | Relation to patient, n (%) | | |
| | | Partner | 3,196 (100.0) | 3,196 (100.0) |
| Young adults and adults (25-64 years) | Siblings | Number of persons, n | 312 (100.0) | 312 (100.0) |
| | | Male, n (%) | 193 (61.9) | 193 (61.9) |
| | | Age, median (Q1;Q3) | 58.3 (53.3;62.2) | 57.8 (53.1;62.4) |
| | | CCI, n (%) | | |
| | | 0 | 238 (76.3) | 240 (76.9) |
| | | 1-2 | 59 (18.9) | 56 (17.9) |
| | | +3 | 15 (4.8) | 16 (5.1) |
| | | Education level, n (%) | | |
| | | No/Unknown education | * | * |
| | | Primary | 65 (20.8) | 65 (20.8) |
| | | Secondary | 180 (57.7) | 180 (57.7) |
| | | Tertiary | * | * |
| | | Relation to patient, n (%) | | |
| | | Sibling | 312 (100.0) | 312 (100.0) |
| Older people (65+ years) | Children | Number of persons, n | 5,638 (100.0) | 5,638 (100.0) |
| | | Male, n (%) | 2,822 (50.1) | 2,822 (50.1) |
| | | Age, median (Q1;Q3) | 54.5 (48.8;59.5) | 54.6 (48.8;59.2) |
| | | CCI, n (%) | | |
| | | 0 | 4,680 (83.0) | 4,753 (84.3) |

**Supplementary Table 219**
**Parkinson's disease - Prevalent cohort 2021**
**Patient characteristics - relatives - not pooled**

| *Age strata of index patients* | *Closest relative type* | *Characteristic of family relatives by agestrata of index patients* | *Closest family relatives of brain disorder patients* | *Comparison relative* |
|---|---|---|---|---|
| | | 1-2 | 828 (14.7) | 753 (13.4) |
| | | +3 | 130 (2.3) | 132 (2.3) |
| | | Education level, n (%) | | |
| | | No/Unknown education | 19 (0.3) | 19 (0.3) |
| | | Primary | 831 (14.7) | 831 (14.7) |
| | | Secondary | 2,876 (51.0) | 2,876 (51.0) |
| | | Tertiary | 1,912 (33.9) | 1,912 (33.9) |
| | | Relation to patient, n (%) | | |
| | | Child | 5,638 (100.0) | 5,638 (100.0) |
| Older people (65+ years) | Partners | Number of persons, n | 8,643 (100.0) | 8,643 (100.0) |
| | | Male, n (%) | 3,088 (35.7) | 3,088 (35.7) |
| | | Age, median (Q1;Q3) | 74.8 (70.3;79.2) | 74.8 (70.4;79.0) |
| | | CCI, n (%) | | |
| | | 0 | 5,194 (60.1) | 5,248 (60.7) |
| | | 1-2 | 2,554 (29.5) | 2,556 (29.6) |
| | | +3 | 895 (10.4) | 839 (9.7) |
| | | Education level, n (%) | | |
| | | No/Unknown education | 459 (5.3) | 459 (5.3) |
| | | Primary | 2,282 (26.4) | 2,282 (26.4) |
| | | Secondary | 4,020 (46.5) | 4,020 (46.5) |
| | | Tertiary | 1,882 (21.8) | 1,882 (21.8) |
| | | Relation to patient, n (%) | | |
| | | Partner | 8,643 (100.0) | 8,643 (100.0) |
| Older people (65+ years) | Siblings | Number of persons, n | 110 (100.0) | 110 (100.0) |
| | | Male, n (%) | 62 (56.4) | 62 (56.4) |
| | | Age, median (Q1;Q3) | 63.4 (59.6;65.6) | 63.2 (60.9;65.8) |
| | | CCI, n (%) | | |
| | | 0 | 88 (80.0) | 78 (70.9) |
| | | 1-2 | * | 27 (24.5) |
| | | +3 | * | 5 (4.5) |
| | | Education level, n (%) | | |
| | | No/Unknown education | * | * |
| | | Primary | * | * |
| | | Secondary | 72 (65.5) | 72 (65.5) |
| | | Tertiary | 21 (19.1) | 21 (19.1) |
| | | Relation to patient, n (%) | | |
| | | Sibling | 110 (100.0) | 110 (100.0) |

#### Supplementary Table 220
#### Parkinson's disease - Prevalent cohort 2021
#### Cost analysis - pooled

| *Age strata of index patients* | *Closest relative type* | *Number of index patients included in cost component calculations* | *Cost component* | *Actual costs* | | *Attributable costs* | |
|---|---|---|---|---|---|---|---|
| | | | | *Total (million EUR)* | *Per person (EUR)* | *Total (million EUR)* | *Per person (EUR)* |
| Children and young people (0-24 years) | Fathers | 163 | Primary sector | 0.0 | 244 | 0.0 | 2 |
| | | 163 | Hospital care | 0.4 | 2,342 | 0.3 | 1,582 |
| | | 163 | Filled prescriptions | 0.0 | 165 | 0.0 | 7 |
| | | 161 | Lost productivity (income loss) | . | . | 1.3 | 8,327 |
| Children and young people (0-24 years) | Mothers | 168 | Primary sector | 0.1 | 354 | 0.0 | 8 |
| | | 168 | Hospital care | 0.4 | 2,493 | 0.2 | 1,044 |
| | | 168 | Filled prescriptions | 0.0 | 248 | 0.0 | 45 |
| | | 167 | Lost productivity (income loss) | . | . | 0.1 | 643 |
| Young adults and adults (25-64 years) | Pooled | 4,767 | Primary sector | 1.6 | 345 | 0.1 | 31 |
| | | 4,767 | Hospital care | 11.3 | 2,381 | 1.5 | 325 |
| | | 4,767 | Filled prescriptions | 1.5 | 326 | 0.3 | 55 |
| | | 3,531 | Lost productivity (income loss) | . | . | 16.4 | 4,646 |
| Older people (65+ years) | Pooled | 14,391 | Primary sector | 6.4 | 450 | 0.3 | 20 |
| | | 14,391 | Hospital care | 43.9 | 3,078 | 0.3 | 23 |
| | | 14,391 | Filled prescriptions | 6.1 | 429 | 0.3 | 20 |
| | | 5,413 | Lost productivity (income loss) | . | . | 14.4 | 2,666 |

### Supplementary Table 221
### Parkinson's disease - Prevalent cohort 2021
### Cost analysis - not pooled

| *Age strata of index patients* | *Closest relative type* | *Number of index patients included in cost component calculations* | *Cost component* | *Actual costs* | | *Attributable costs* | |
|---|---|---|---|---|---|---|---|
| | | | | *Total (million EUR)* | *Per person (EUR)* | *Total (million EUR)* | *Per person (EUR)* |
| Children and young people (0-24 years) | Fathers | 163 | Primary sector | 0.0 | 244 | 0.0 | 2 |
| | | 163 | Hospital care | 0.4 | 2,342 | 0.3 | 1,582 |
| | | 163 | Filled prescriptions | 0.0 | 165 | 0.0 | 7 |
| | | 161 | Lost productivity (income loss) | . | . | 1.3 | 8,327 |
| Children and young people (0-24 years) | Mothers | 168 | Primary sector | 0.1 | 354 | 0.0 | 8 |
| | | 168 | Hospital care | 0.4 | 2,493 | 0.2 | 1,044 |
| | | 168 | Filled prescriptions | 0.0 | 248 | 0.0 | 45 |
| | | 167 | Lost productivity (income loss) | . | . | 0.1 | 643 |
| Young adults and adults (25-64 years) | Children | 731 | Primary sector | 0.2 | 277 | 0.0 | 44 |
| | | 731 | Hospital care | 1.5 | 2,114 | 0.4 | 591 |
| | | 731 | Filled prescriptions | 0.1 | 172 | 0.0 | 45 |
| | | 545 | Lost productivity (income loss) | . | . | 1.5 | 2,716 |
| Young adults and adults (25-64 years) | Parents | 528 | Primary sector | 0.3 | 531 | 0.0 | 37 |
| | | 528 | Hospital care | 1.6 | 3,044 | 0.1 | 254 |
| | | 528 | Filled prescriptions | 0.3 | 585 | 0.0 | 71 |
| | | 110 | Lost productivity (income loss) | . | . | 1.1 | 9,820 |
| Young adults and adults (25-64 years) | Partners | 3,196 | Primary sector | 1.1 | 332 | 0.1 | 32 |
| | | 3,196 | Hospital care | 7.6 | 2,379 | 1.1 | 342 |
| | | 3,196 | Filled prescriptions | 1.0 | 315 | 0.2 | 58 |
| | | 2,631 | Lost productivity (income loss) | . | . | 12.3 | 4,663 |
| Young adults and adults (25-64 years) | Siblings | 312 | Primary sector | 0.1 | 324 | -0.0 | -24 |
| | | 312 | Hospital care | 0.6 | 1,927 | -0.1 | -356 |
| | | 312 | Filled prescriptions | 0.1 | 364 | 0.0 | 27 |
| | | 245 | Lost productivity (income loss) | . | . | 1.6 | 6,434 |
| Older people (65+ years) | Children | 5,638 | Primary sector | 1.8 | 320 | 0.1 | 13 |
| | | 5,638 | Hospital care | 11.0 | 1,958 | 1.1 | 203 |
| | | 5,638 | Filled prescriptions | 1.5 | 272 | 0.1 | 16 |
| | | 5,037 | Lost productivity (income loss) | . | . | 14.2 | 2,819 |
| Older people (65+ years) | Partners | 8,643 | Primary sector | 4.6 | 538 | 0.2 | 25 |
| | | 8,643 | Hospital care | 32.6 | 3,822 | -0.9 | -111 |
| | | 8,643 | Filled prescriptions | 4.5 | 532 | 0.2 | 23 |
| | | 329 | Lost productivity (income loss) | . | . | 0.4 | 1,109 |
| Older people (65+ years) | Siblings | 110 | Primary sector | 0.0 | 355 | 0.0 | 25 |
| | | 110 | Hospital care | 0.3 | 2,741 | 0.1 | 1,167 |
| | | 110 | Filled prescriptions | 0.0 | 425 | -0.0 | -9 |
| | | 47 | Lost productivity (income loss) | . | . | -0.1 | -2,853 |

**Supplementary Table 222**
**Personality disorders - Incident cohort 2016-2021**
**Identification of populations of relatives**

| ***Agegroup of index patients in matched population*** | | ***Closest relative type identified*** | | ***Available control with same type of closest relative*** |
|---|---|---|---|---|
| Children and young people (0-24 years) | n = 8,094 | Fathers | n = 6,631 (82%) | 96% |
| | | Mothers | n = 7,010 (87%) | 96% |
| | | No available closest relative | n = 399 (5%) | n/a |
| Young adults and adults (25-64 years) | n = 9,712 | Children | n = 1,391 (14%) | 95% |
| | | Parents | n = 3,015 (31%) | 92% |
| | | Partners | n = 3,369 (35%) | 95% |
| | | Siblings | n = 1,691 (17%) | 33% |
| | | Pooled | n = 9,466 (97%) | 83% |
| | | No available closest relative | n = 246 (3%) | n/a |
| Older people (65+ years) | n = 282 | Children | n = 131 (46%) | 93% |
| | | Partners | n = 86 (30%) | 93% |
| | | Siblings | n = 17 (6%) | 29% |
| | | Pooled | n = 234 (83%) | 88% |
| | | No available closest relative | n = 48 (17%) | n/a |

#### Supplementary Table 223
#### Personality disorders - Incident cohort 2016-2021
#### Patient characteristics - cases

| *Age strata of index patients* | *Characteristic* | |
|---|---|---|
| Children and young people (0-24 years) | Number of persons, n | 7,432 (100.0) |
| | Male, n (%) | 1,328 (17.9) |
| | Age, median (Q1;Q3) | 20.7 (18.4;22.7) |
| | CCI, n (%) | |
| | 0 | 6,884 (92.6) |
| | 1-2 | 531 (7.1) |
| | +3 | 17 (0.2) |
| | Education level, n (%) | |
| | No/Unknown education | 33 (0.4) |
| | Primary | 5,523 (74.3) |
| | Secondary | 1,783 (24.0) |
| | Tertiary | 93 (1.3) |
| Young adults and adults (25-64 years) | Number of persons, n | 7,916 (100.0) |
| | Male, n (%) | 2,556 (32.3) |
| | Age, median (Q1;Q3) | 34.8 (28.8;44.0) |
| | CCI, n (%) | |
| | 0 | 6,859 (86.6) |
| | 1-2 | 949 (12.0) |
| | +3 | 108 (1.4) |
| | Education level, n (%) | |
| | No/Unknown education | 147 (1.9) |
| | Primary | 2,717 (34.3) |
| | Secondary | 3,325 (42.0) |
| | Tertiary | 1,727 (21.8) |
| Older people (65+ years) | Number of persons, n | 207 (100.0) |
| | Male, n (%) | 65 (31.4) |
| | Age, median (Q1;Q3) | 73.2 (68.0;78.5) |
| | CCI, n (%) | |
| | 0 | 91 (44.0) |
| | 1-2 | 78 (37.7) |
| | +3 | 38 (18.4) |
| | Education level, n (%) | |
| | No/Unknown education | 11 (5.3) |
| | Primary | 75 (36.2) |
| | Secondary | 84 (40.6) |
| | Tertiary | 37 (17.9) |

#### Supplementary Table 224
#### Personality disorders - Incident cohort 2016-2021
#### Patient characteristics - relatives - pooled

| *Age strata of index patients* | *Closest relative type* | *Characteristic of family relatives by agestrata of index patients* | *Closest family relatives of brain disorder patients* | *Comparison relative* |
|---|---|---|---|---|
| Children and young people (0-24 years) | Fathers | Number of persons, n | 6,397 (100.0) | 6,397 (100.0) |
| | | Male, n (%) | 6,397 (100.0) | 6,397 (100.0) |
| | | Age, median (Q1;Q3) | 51.9 (47.8;55.9) | 52.0 (48.4;55.6) |
| | | CCI, n (%) | | |
| | | 0 | 5,266 (82.3) | 5,465 (85.4) |
| | | 1-2 | 967 (15.1) | 812 (12.7) |
| | | +3 | 164 (2.6) | 120 (1.9) |
| | | Education level, n (%) | | |
| | | No/Unknown education | 78 (1.2) | 78 (1.2) |
| | | Primary | 1,736 (27.1) | 1,736 (27.1) |
| | | Secondary | 3,303 (51.6) | 3,303 (51.6) |
| | | Tertiary | 1,280 (20.0) | 1,280 (20.0) |
| | | Relation to patient, n (%) | | |
| | | Father | 6,397 (100.0) | 6,397 (100.0) |
| Children and young people (0-24 years) | Mothers | Number of persons, n | 6,770 (100.0) | 6,770 (100.0) |
| | | Male, n (%) | 0 (0.0) | 0 (0.0) |
| | | Age, median (Q1;Q3) | 49.6 (45.5;53.5) | 49.7 (46.3;53.2) |
| | | CCI, n (%) | | |
| | | 0 | 5,576 (82.4) | 5,851 (86.4) |
| | | 1-2 | 1,084 (16.0) | 842 (12.4) |
| | | +3 | 110 (1.6) | 77 (1.1) |
| | | Education level, n (%) | | |
| | | No/Unknown education | 50 (0.7) | 50 (0.7) |
| | | Primary | 1,538 (22.7) | 1,538 (22.7) |
| | | Secondary | 3,185 (47.0) | 3,185 (47.0) |
| | | Tertiary | 1,997 (29.5) | 1,997 (29.5) |
| | | Relation to patient, n (%) | | |
| | | Mother | 6,770 (100.0) | 6,770 (100.0) |
| Young adults and adults (25-64 years) | Pooled | Number of persons, n | 7,916 (100.0) | 7,916 (100.0) |
| | | Male, n (%) | 4,114 (52.0) | 4,114 (52.0) |
| | | Age, median (Q1;Q3) | 43.5 (29.1;57.6) | 43.5 (29.3;57.8) |
| | | CCI, n (%) | | |
| | | 0 | 6,682 (84.4) | 6,818 (86.1) |
| | | 1-2 | 1,036 (13.1) | 935 (11.8) |
| | | +3 | 198 (2.5) | 163 (2.1) |
| | | Education level, n (%) | | |
| | | No/Unknown education | 708 (8.9) | 708 (8.9) |
| | | Primary | 2,236 (28.2) | 2,236 (28.2) |
| | | Secondary | 3,369 (42.6) | 3,369 (42.6) |
| | | Tertiary | 1,603 (20.3) | 1,603 (20.3) |
| | | Relation to patient, n (%) | | |
| | | Mother | 2,140 (27.0) | 2,140 (27.0) |
| | | Father | 663 (8.4) | 663 (8.4) |
| | | Partner | 3,217 (40.6) | 3,217 (40.6) |
| | | Child | 1,327 (16.8) | 1,327 (16.8) |

**Supplementary Table 224**
**Personality disorders - Incident cohort 2016-2021**
**Patient characteristics - relatives - pooled**

| *Age strata of index patients* | *Closest relative type* | *Characteristic of family relatives by agestrata of index patients* | *Closest family relatives of brain disorder patients* | *Comparison relative* |
|---|---|---|---|---|
| | | Sibling | 569 (7.2) | 569 (7.2) |
| Older people (65+ years) | Pooled | Number of persons, n | 207 (100.0) | 207 (100.0) |
| | | Male, n (%) | 104 (50.2) | 104 (50.2) |
| | | Age, median (Q1;Q3) | 57.1 (46.4;71.7) | 58.0 (46.7;71.5) |
| | | CCI, n (%) | | |
| | | 0 | 156 (75.4) | 155 (74.9) |
| | | 1-2 | 42 (20.3) | 37 (17.9) |
| | | +3 | 9 (4.3) | 15 (7.2) |
| | | Education level, n (%) | | |
| | | No/Unknown education | * | * |
| | | Primary | * | * |
| | | Secondary | 97 (46.9) | 97 (46.9) |
| | | Tertiary | 63 (30.4) | 63 (30.4) |
| | | Relation to patient, n (%) | | |
| | | Partner | 80 (38.6) | 80 (38.6) |
| | | Child | 122 (58.9) | 122 (58.9) |
| | | Sibling | 5 (2.4) | 5 (2.4) |

#### Supplementary Table 225
#### Personality disorders - Incident cohort 2016-2021
#### Patient characteristics - relatives - not pooled

| *Age strata of index patients* | *Closest relative type* | *Characteristic of family relatives by agestrata of index patients* | *Closest family relatives of brain disorder patients* | *Comparison relative* |
|---|---|---|---|---|
| Children and young people (0-24 years) | Fathers | Number of persons, n | 6,397 (100.0) | 6,397 (100.0) |
| | | Male, n (%) | 6,397 (100.0) | 6,397 (100.0) |
| | | Age, median (Q1;Q3) | 51.9 (47.8;55.9) | 52.0 (48.4;55.6) |
| | | CCI, n (%) | | |
| | | 0 | 5,266 (82.3) | 5,465 (85.4) |
| | | 1-2 | 967 (15.1) | 812 (12.7) |
| | | +3 | 164 (2.6) | 120 (1.9) |
| | | Education level, n (%) | | |
| | | No/Unknown education | 78 (1.2) | 78 (1.2) |
| | | Primary | 1,736 (27.1) | 1,736 (27.1) |
| | | Secondary | 3,303 (51.6) | 3,303 (51.6) |
| | | Tertiary | 1,280 (20.0) | 1,280 (20.0) |
| | | Relation to patient, n (%) | | |
| | | Father | 6,397 (100.0) | 6,397 (100.0) |
| Children and young people (0-24 years) | Mothers | Number of persons, n | 6,770 (100.0) | 6,770 (100.0) |
| | | Male, n (%) | 0 (0.0) | 0 (0.0) |
| | | Age, median (Q1;Q3) | 49.6 (45.5;53.5) | 49.7 (46.3;53.2) |
| | | CCI, n (%) | | |
| | | 0 | 5,576 (82.4) | 5,851 (86.4) |
| | | 1-2 | 1,084 (16.0) | 842 (12.4) |
| | | +3 | 110 (1.6) | 77 (1.1) |
| | | Education level, n (%) | | |
| | | No/Unknown education | 50 (0.7) | 50 (0.7) |
| | | Primary | 1,538 (22.7) | 1,538 (22.7) |
| | | Secondary | 3,185 (47.0) | 3,185 (47.0) |
| | | Tertiary | 1,997 (29.5) | 1,997 (29.5) |
| | | Relation to patient, n (%) | | |
| | | Mother | 6,770 (100.0) | 6,770 (100.0) |
| Young adults and adults (25-64 years) | Children | Number of persons, n | 1,327 (100.0) | 1,327 (100.0) |
| | | Male, n (%) | 682 (51.4) | 682 (51.4) |
| | | Age, median (Q1;Q3) | 14.6 (8.6;20.9) | 14.3 (7.7;20.2) |
| | | CCI, n (%) | | |
| | | 0 | 1,236 (93.1) | 1,247 (94.0) |
| | | 1-2 | * | 80 (6.0) |
| | | +3 | * | 0 (0.0) |
| | | Education level, n (%) | | |
| | | No/Unknown education | 602 (45.4) | 602 (45.4) |
| | | Primary | 524 (39.5) | 524 (39.5) |
| | | Secondary | 158 (11.9) | 158 (11.9) |
| | | Tertiary | 43 (3.2) | 43 (3.2) |
| | | Relation to patient, n (%) | | |
| | | Child | 1,327 (100.0) | 1,327 (100.0) |
| Young adults and adults (25-64 years) | Parents | Number of persons, n | 2,803 (100.0) | 2,803 (100.0) |
| | | Male, n (%) | 663 (23.7) | 663 (23.7) |
| | | Age, median (Q1;Q3) | 60.1 (55.2;66.4) | 60.2 (55.5;66.5) |

#### Supplementary Table 225
#### Personality disorders - Incident cohort 2016-2021
#### Patient characteristics - relatives - not pooled

| *Age strata of index patients* | *Closest relative type* | *Characteristic of family relatives by agestrata of index patients* | *Closest family relatives of brain disorder patients* | *Comparison relative* |
|---|---|---|---|---|
| | | CCI, n (%) | | |
| | | 0 | 2,015 (71.9) | 2,100 (74.9) |
| | | 1-2 | 627 (22.4) | 569 (20.3) |
| | | +3 | 161 (5.7) | 134 (4.8) |
| | | Education level, n (%) | | |
| | | No/Unknown education | 52 (1.9) | 52 (1.9) |
| | | Primary | 757 (27.0) | 757 (27.0) |
| | | Secondary | 1,378 (49.2) | 1,378 (49.2) |
| | | Tertiary | 616 (22.0) | 616 (22.0) |
| | | Relation to patient, n (%) | | |
| | | Mother | 2,140 (76.3) | 2,140 (76.3) |
| | | Father | 663 (23.7) | 663 (23.7) |
| Young adults and adults (25-64 years) | Partners | Number of persons, n | 3,217 (100.0) | 3,217 (100.0) |
| | | Male, n (%) | 2,417 (75.1) | 2,417 (75.1) |
| | | Age, median (Q1;Q3) | 37.2 (30.7;45.5) | 37.2 (30.8;45.1) |
| | | CCI, n (%) | | |
| | | 0 | 2,928 (91.0) | 2,964 (92.1) |
| | | 1-2 | 259 (8.1) | 228 (7.1) |
| | | +3 | 30 (0.9) | 25 (0.8) |
| | | Education level, n (%) | | |
| | | No/Unknown education | 53 (1.6) | 53 (1.6) |
| | | Primary | 799 (24.8) | 799 (24.8) |
| | | Secondary | 1,530 (47.6) | 1,530 (47.6) |
| | | Tertiary | 835 (26.0) | 835 (26.0) |
| | | Relation to patient, n (%) | | |
| | | Partner | 3,217 (100.0) | 3,217 (100.0) |
| Young adults and adults (25-64 years) | Siblings | Number of persons, n | 569 (100.0) | 569 (100.0) |
| | | Male, n (%) | 352 (61.9) | 352 (61.9) |
| | | Age, median (Q1;Q3) | 36.6 (28.5;51.6) | 36.6 (28.2;51.8) |
| | | CCI, n (%) | | |
| | | 0 | 503 (88.4) | 507 (89.1) |
| | | 1-2 | * | * |
| | | +3 | * | * |
| | | Education level, n (%) | | |
| | | No/Unknown education | * | * |
| | | Primary | 156 (27.4) | 156 (27.4) |
| | | Secondary | 303 (53.3) | 303 (53.3) |
| | | Tertiary | * | * |
| | | Relation to patient, n (%) | | |
| | | Sibling | 569 (100.0) | 569 (100.0) |
| Older people (65+ years) | Children | Number of persons, n | 122 (100.0) | 122 (100.0) |
| | | Male, n (%) | 55 (45.1) | 55 (45.1) |
| | | Age, median (Q1;Q3) | 48.3 (43.4;54.4) | 48.2 (44.5;55.5) |
| | | CCI, n (%) | | |
| | | 0 | 108 (88.5) | 110 (90.2) |

**Supplementary Table 225**
**Personality disorders - Incident cohort 2016-2021**
**Patient characteristics - relatives - not pooled**

| *Age strata of index patients* | *Closest relative type* | *Characteristic of family relatives by agestrata of index patients* | *Closest family relatives of brain disorder patients* | *Comparison relative* |
|---|---|---|---|---|
| | | 1-2 | * | * |
| | | +3 | * | * |
| | | Education level, n (%) | | |
| | | No/Unknown education | 0 (0.0) | 0 (0.0) |
| | | Primary | 20 (16.4) | 20 (16.4) |
| | | Secondary | 58 (47.5) | 58 (47.5) |
| | | Tertiary | 44 (36.1) | 44 (36.1) |
| | | Relation to patient, n (%) | | |
| | | Child | 122 (100.0) | 122 (100.0) |
| Older people (65+ years) | Partners | Number of persons, n | 80 (100.0) | 80 (100.0) |
| | | Male, n (%) | 47 (58.8) | 47 (58.8) |
| | | Age, median (Q1;Q3) | 73.1 (69.9;76.7) | 73.9 (68.6;78.2) |
| | | CCI, n (%) | | |
| | | 0 | 45 (56.3) | 42 (52.5) |
| | | 1-2 | 27 (33.8) | 26 (32.5) |
| | | +3 | 8 (10.0) | 12 (15.0) |
| | | Education level, n (%) | | |
| | | No/Unknown education | * | * |
| | | Primary | 22 (27.5) | 22 (27.5) |
| | | Secondary | 37 (46.3) | 37 (46.3) |
| | | Tertiary | * | * |
| | | Relation to patient, n (%) | | |
| | | Partner | 80 (100.0) | 80 (100.0) |
| Older people (65+ years) | Siblings | Number of persons, n | 5 (100.0) | 5 (100.0) |
| | | Male, n (%) | * | * |
| | | Age, median (Q1;Q3) | 63.7 (60.5;65.1) | 60.7 (59.8;65.6) |
| | | CCI, n (%) | | |
| | | 0 | * | * |
| | | 1-2 | * | * |
| | | +3 | 0 (0.0) | * |
| | | Education level, n (%) | | |
| | | No/Unknown education | 0 (0.0) | 0 (0.0) |
| | | Primary | * | * |
| | | Secondary | * | * |
| | | Tertiary | * | * |
| | | Relation to patient, n (%) | | |
| | | Sibling | 5 (100.0) | 5 (100.0) |

#### Supplementary Table 226
#### Personality disorders - Incident cohort 2016-2021
#### Cost analysis - pooled

| | | | | *Actual costs* | | *Attributable costs* | |
|---|---|---|---|---|---|---|---|
| *Age strata of index patients* | *Closest relative type* | *Number of index patients included in cost component calculations* | *Cost component* | *Total (million EUR)* | *Per person (EUR)* | *Total (million EUR)* | *Per person (EUR)* |
| Children and young people (0-24 years) | Fathers | 6,397 | Primary sector | 1.7 | 262 | 0.2 | 35 |
| | | 6,397 | Hospital care | 13.1 | 2,049 | 2.2 | 347 |
| | | 6,397 | Filled prescriptions | 1.7 | 264 | 0.5 | 71 |
| | | 6,059 | Lost productivity (income loss) | . | . | 62.4 | 10,301 |
| Children and young people (0-24 years) | Mothers | 6,770 | Primary sector | 2.7 | 397 | 0.4 | 55 |
| | | 6,770 | Hospital care | 16.9 | 2,494 | 5.4 | 803 |
| | | 6,770 | Filled prescriptions | 2.0 | 290 | 0.6 | 95 |
| | | 6,690 | Lost productivity (income loss) | . | . | 57.3 | 8,561 |
| Young adults and adults (25-64 years) | Pooled | 7,916 | Primary sector | 2.5 | 312 | 0.3 | 38 |
| | | 7,916 | Hospital care | 16.0 | 2,023 | 2.1 | 262 |
| | | 7,916 | Filled prescriptions | 2.1 | 262 | 0.5 | 62 |
| | | 5,764 | Lost productivity (income loss) | . | . | 44.7 | 7,759 |
| Older people (65+ years) | Pooled | 207 | Primary sector | 0.1 | 398 | 0.0 | 60 |
| | | 207 | Hospital care | 0.6 | 2,787 | -0.0 | -118 |
| | | 207 | Filled prescriptions | 0.1 | 319 | 0.0 | 2 |
| | | 120 | Lost productivity (income loss) | . | . | 0.2 | 1,298 |

#### Supplementary Table 227
#### Personality disorders - Incident cohort 2016-2021
#### Cost analysis - not pooled

| *Age strata of index patients* | *Closest relative type* | *Number of index patients included in cost component calculations* | *Cost component* | *Actual costs* | | *Attributable costs* | |
|---|---|---|---|---|---|---|---|
| | | | | *Total (million EUR)* | *Per person (EUR)* | *Total (million EUR)* | *Per person (EUR)* |
| Children and young people (0-24 years) | Fathers | 6,397 | Primary sector | 1.7 | 262 | 0.2 | 35 |
| | | 6,397 | Hospital care | 13.1 | 2,049 | 2.2 | 347 |
| | | 6,397 | Filled prescriptions | 1.7 | 264 | 0.5 | 71 |
| | | 6,059 | Lost productivity (income loss) | . | . | 62.4 | 10,301 |
| Children and young people (0-24 years) | Mothers | 6,770 | Primary sector | 2.7 | 397 | 0.4 | 55 |
| | | 6,770 | Hospital care | 16.9 | 2,494 | 5.4 | 803 |
| | | 6,770 | Filled prescriptions | 2.0 | 290 | 0.6 | 95 |
| | | 6,690 | Lost productivity (income loss) | . | . | 57.3 | 8,561 |
| Young adults and adults (25-64 years) | Children | 1,327 | Primary sector | 0.3 | 195 | 0.0 | 24 |
| | | 1,327 | Hospital care | 1.6 | 1,233 | 0.5 | 367 |
| | | 1,327 | Filled prescriptions | 0.1 | 101 | 0.0 | 34 |
| | | 416 | Lost productivity (income loss) | . | . | 1.2 | 2,872 |
| Young adults and adults (25-64 years) | Parents | 2,803 | Primary sector | 1.2 | 433 | 0.1 | 41 |
| | | 2,803 | Hospital care | 7.9 | 2,840 | 0.3 | 99 |
| | | 2,803 | Filled prescriptions | 1.3 | 448 | 0.2 | 88 |
| | | 1,676 | Lost productivity (income loss) | . | . | 8.2 | 4,881 |
| Young adults and adults (25-64 years) | Partners | 3,217 | Primary sector | 0.8 | 257 | 0.1 | 38 |
| | | 3,217 | Hospital care | 5.1 | 1,602 | 1.3 | 391 |
| | | 3,217 | Filled prescriptions | 0.6 | 180 | 0.2 | 60 |
| | | 3,152 | Lost productivity (income loss) | . | . | 34.3 | 10,894 |
| Young adults and adults (25-64 years) | Siblings | 569 | Primary sector | 0.2 | 303 | 0.0 | 55 |
| | | 569 | Hospital care | 1.3 | 2,239 | 0.0 | 88 |
| | | 569 | Filled prescriptions | 0.1 | 183 | 0.0 | 18 |
| | | 520 | Lost productivity (income loss) | . | . | 1.0 | 1,941 |
| Older people (65+ years) | Children | 122 | Primary sector | 0.0 | 312 | 0.0 | 45 |
| | | 122 | Hospital care | 0.3 | 2,056 | 0.1 | 806 |
| | | 122 | Filled prescriptions | 0.0 | 185 | -0.0 | -42 |
| | | 116 | Lost productivity (income loss) | . | . | 0.1 | 479 |
| Older people (65+ years) | Partners | 80 | Primary sector | 0.0 | 528 | 0.0 | 88 |
| | | 80 | Hospital care | 0.3 | 3,772 | 0.0 | 219 |
| | | 80 | Filled prescriptions | 0.0 | 524 | 0.0 | 58 |
| | | . | Lost productivity (income loss) | . | . | . | . |
| Older people (65+ years) | Siblings | . | Primary sector | . | . | . | . |
| | | . | Hospital care | . | . | . | . |
| | | . | Filled prescriptions | . | . | . | . |
| | | . | Lost productivity (income loss) | . | . | . | . |

**Supplementary Table 228**
**Personality disorders - Prevalent cohort 2021**
**Identification of populations of relatives**

| ***Agegroup of index patients in matched population*** | | ***Closest relative type identified*** | | ***Available control with same type of closest relative*** |
|---|---|---|---|---|
| Children and young people (0-24 years) | n = 6,574 | Fathers | n = 5,350 (81%) | 96% |
| | | Mothers | n = 5,583 (85%) | 96% |
| | | No available closest relative | n = 338 (5%) | n/a |
| Young adults and adults (25-64 years) | n = 58,363 | Children | n = 9,534 (16%) | 94% |
| | | Parents | n = 16,126 (28%) | 90% |
| | | Partners | n = 20,971 (36%) | 94% |
| | | Siblings | n = 10,536 (18%) | 38% |
| | | Pooled | n = 57,167 (98%) | 83% |
| | | No available closest relative | n = 1,196 (2%) | n/a |
| Older people (65+ years) | n = 4,456 | Children | n = 1,939 (44%) | 92% |
| | | Partners | n = 1,309 (29%) | 95% |
| | | Siblings | n = 408 (9%) | 50% |
| | | Pooled | n = 3,656 (82%) | 89% |
| | | No available closest relative | n = 800 (18%) | n/a |

#### Supplementary Table 229
#### Personality disorders - Prevalent cohort 2021
#### Patient characteristics - cases

| ***Age strata of index patients*** | ***Characteristic*** | |
|---|---|---|
| Children and young people (0-24 years) | Number of persons, n | 6,008 (100.0) |
| | Male, n (%) | 1,025 (17.1) |
| | Age, median (Q1;Q3) | 22.5 (20.6;23.8) |
| | CCI, n (%) | |
| | 0 | 5,475 (91.1) |
| | 1-2 | 512 (8.5) |
| | +3 | 21 (0.3) |
| | Education level, n (%) | |
| | No/Unknown education | 18 (0.3) |
| | Primary | 4,298 (71.5) |
| | Secondary | 1,585 (26.4) |
| | Tertiary | 107 (1.8) |
| Young adults and adults (25-64 years) | Number of persons, n | 47,515 (100.0) |
| | Male, n (%) | 14,594 (30.7) |
| | Age, median (Q1;Q3) | 40.0 (32.4;49.4) |
| | CCI, n (%) | |
| | 0 | 38,984 (82.0) |
| | 1-2 | 7,450 (15.7) |
| | +3 | 1,081 (2.3) |
| | Education level, n (%) | |
| | No/Unknown education | 670 (1.4) |
| | Primary | 19,312 (40.6) |
| | Secondary | 18,046 (38.0) |
| | Tertiary | 9,487 (20.0) |
| Older people (65+ years) | Number of persons, n | 3,260 (100.0) |
| | Male, n (%) | 950 (29.1) |
| | Age, median (Q1;Q3) | 70.9 (67.4;75.5) |
| | CCI, n (%) | |
| | 0 | 1,538 (47.2) |
| | 1-2 | 1,221 (37.5) |
| | +3 | 501 (15.4) |
| | Education level, n (%) | |
| | No/Unknown education | 118 (3.6) |
| | Primary | 1,131 (34.7) |
| | Secondary | 1,354 (41.5) |
| | Tertiary | 657 (20.2) |

**Supplementary Table 230**
**Personality disorders - Prevalent cohort 2021**
**Patient characteristics - relatives - pooled**

| *Age strata of index patients* | *Closest relative type* | *Characteristic of family relatives by agestrata of index patients* | *Closest family relatives of brain disorder patients* | *Comparison relative* |
|---|---|---|---|---|
| Children and young people (0-24 years) | Fathers | Number of persons, n | 5,152 (100.0) | 5,152 (100.0) |
| | | Male, n (%) | 5,152 (100.0) | 5,152 (100.0) |
| | | Age, median (Q1;Q3) | 53.5 (49.6;57.4) | 53.7 (50.1;57.0) |
| | | CCI, n (%) | | |
| | | 0 | 4,170 (80.9) | 4,393 (85.3) |
| | | 1-2 | 838 (16.3) | 670 (13.0) |
| | | +3 | 144 (2.8) | 89 (1.7) |
| | | Education level, n (%) | | |
| | | No/Unknown education | 59 (1.1) | 59 (1.1) |
| | | Primary | 1,414 (27.4) | 1,414 (27.4) |
| | | Secondary | 2,695 (52.3) | 2,695 (52.3) |
| | | Tertiary | 984 (19.1) | 984 (19.1) |
| | | Relation to patient, n (%) | | |
| | | Father | 5,152 (100.0) | 5,152 (100.0) |
| Children and young people (0-24 years) | Mothers | Number of persons, n | 5,371 (100.0) | 5,371 (100.0) |
| | | Male, n (%) | 0 (0.0) | 0 (0.0) |
| | | Age, median (Q1;Q3) | 51.0 (47.2;55.0) | 51.2 (47.9;54.6) |
| | | CCI, n (%) | | |
| | | 0 | 4,320 (80.4) | 4,591 (85.5) |
| | | 1-2 | 939 (17.5) | 699 (13.0) |
| | | +3 | 112 (2.1) | 81 (1.5) |
| | | Education level, n (%) | | |
| | | No/Unknown education | 30 (0.6) | 30 (0.6) |
| | | Primary | 1,206 (22.5) | 1,206 (22.5) |
| | | Secondary | 2,535 (47.2) | 2,535 (47.2) |
| | | Tertiary | 1,600 (29.8) | 1,600 (29.8) |
| | | Relation to patient, n (%) | | |
| | | Mother | 5,371 (100.0) | 5,371 (100.0) |
| Young adults and adults (25-64 years) | Pooled | Number of persons, n | 47,515 (100.0) | 47,515 (100.0) |
| | | Male, n (%) | 25,682 (54.1) | 25,682 (54.1) |
| | | Age, median (Q1;Q3) | 45.9 (30.9;60.0) | 45.9 (31.0;60.1) |
| | | CCI, n (%) | | |
| | | 0 | 39,040 (82.2) | 40,273 (84.8) |
| | | 1-2 | 7,066 (14.9) | 6,088 (12.8) |
| | | +3 | 1,409 (3.0) | 1,154 (2.4) |
| | | Education level, n (%) | | |
| | | No/Unknown education | 3,723 (7.8) | 3,723 (7.8) |
| | | Primary | 14,344 (30.2) | 14,344 (30.2) |
| | | Secondary | 20,452 (43.0) | 20,452 (43.0) |
| | | Tertiary | 8,996 (18.9) | 8,996 (18.9) |
| | | Relation to patient, n (%) | | |
| | | Mother | 11,211 (23.6) | 11,211 (23.6) |
| | | Father | 3,399 (7.2) | 3,399 (7.2) |
| | | Partner | 19,899 (41.9) | 19,899 (41.9) |
| | | Child | 8,993 (18.9) | 8,993 (18.9) |

#### Supplementary Table 230
#### Personality disorders - Prevalent cohort 2021
#### Patient characteristics - relatives - pooled

| *Age strata of index patients* | *Closest relative type* | *Characteristic of family relatives by agestrata of index patients* | *Closest family relatives of brain disorder patients* | *Comparison relative* |
|---|---|---|---|---|
| | | Sibling | 4,013 (8.4) | 4,013 (8.4) |
| Older people (65+ years) | Pooled | Number of persons, n | 3,260 (100.0) | 3,260 (100.0) |
| | | Male, n (%) | 1,843 (56.5) | 1,843 (56.5) |
| | | Age, median (Q1;Q3) | 57.3 (46.4;69.3) | 57.2 (45.9;69.4) |
| | | CCI, n (%) | | |
| | | 0 | 2,416 (74.1) | 2,472 (75.8) |
| | | 1-2 | 667 (20.5) | 631 (19.4) |
| | | +3 | 177 (5.4) | 157 (4.8) |
| | | Education level, n (%) | | |
| | | No/Unknown education | 79 (2.4) | 79 (2.4) |
| | | Primary | 682 (20.9) | 682 (20.9) |
| | | Secondary | 1,540 (47.2) | 1,540 (47.2) |
| | | Tertiary | 959 (29.4) | 959 (29.4) |
| | | Relation to patient, n (%) | | |
| | | Partner | 1,253 (38.4) | 1,253 (38.4) |
| | | Child | 1,799 (55.2) | 1,799 (55.2) |
| | | Sibling | 208 (6.4) | 208 (6.4) |

#### Supplementary Table 231
#### Personality disorders - Prevalent cohort 2021
#### Patient characteristics - relatives - not pooled

| *Age strata of index patients* | *Closest relative type* | *Characteristic of family relatives by agestrata of index patients* | *Closest family relatives of brain disorder patients* | *Comparison relative* |
|---|---|---|---|---|
| Children and young people (0-24 years) | Fathers | Number of persons, n | 5,152 (100.0) | 5,152 (100.0) |
| | | Male, n (%) | 5,152 (100.0) | 5,152 (100.0) |
| | | Age, median (Q1;Q3) | 53.5 (49.6;57.4) | 53.7 (50.1;57.0) |
| | | CCI, n (%) | | |
| | | 0 | 4,170 (80.9) | 4,393 (85.3) |
| | | 1-2 | 838 (16.3) | 670 (13.0) |
| | | +3 | 144 (2.8) | 89 (1.7) |
| | | Education level, n (%) | | |
| | | No/Unknown education | 59 (1.1) | 59 (1.1) |
| | | Primary | 1,414 (27.4) | 1,414 (27.4) |
| | | Secondary | 2,695 (52.3) | 2,695 (52.3) |
| | | Tertiary | 984 (19.1) | 984 (19.1) |
| | | Relation to patient, n (%) | | |
| | | Father | 5,152 (100.0) | 5,152 (100.0) |
| Children and young people (0-24 years) | Mothers | Number of persons, n | 5,371 (100.0) | 5,371 (100.0) |
| | | Male, n (%) | 0 (0.0) | 0 (0.0) |
| | | Age, median (Q1;Q3) | 51.0 (47.2;55.0) | 51.2 (47.9;54.6) |
| | | CCI, n (%) | | |
| | | 0 | 4,320 (80.4) | 4,591 (85.5) |
| | | 1-2 | 939 (17.5) | 699 (13.0) |
| | | +3 | 112 (2.1) | 81 (1.5) |
| | | Education level, n (%) | | |
| | | No/Unknown education | 30 (0.6) | 30 (0.6) |
| | | Primary | 1,206 (22.5) | 1,206 (22.5) |
| | | Secondary | 2,535 (47.2) | 2,535 (47.2) |
| | | Tertiary | 1,600 (29.8) | 1,600 (29.8) |
| | | Relation to patient, n (%) | | |
| | | Mother | 5,371 (100.0) | 5,371 (100.0) |
| Young adults and adults (25-64 years) | Children | Number of persons, n | 8,993 (100.0) | 8,993 (100.0) |
| | | Male, n (%) | 4,722 (52.5) | 4,722 (52.5) |
| | | Age, median (Q1;Q3) | 17.3 (10.5;25.7) | 17.0 (9.7;24.7) |
| | | CCI, n (%) | | |
| | | 0 | 8,364 (93.0) | 8,499 (94.5) |
| | | 1-2 | 615 (6.8) | 481 (5.3) |
| | | +3 | 14 (0.2) | 13 (0.1) |
| | | Education level, n (%) | | |
| | | No/Unknown education | 3,141 (34.9) | 3,141 (34.9) |
| | | Primary | 3,736 (41.5) | 3,736 (41.5) |
| | | Secondary | 1,570 (17.5) | 1,570 (17.5) |
| | | Tertiary | 546 (6.1) | 546 (6.1) |
| | | Relation to patient, n (%) | | |
| | | Child | 8,993 (100.0) | 8,993 (100.0) |
| Young adults and adults (25-64 years) | Parents | Number of persons, n | 14,610 (100.0) | 14,610 (100.0) |
| | | Male, n (%) | 3,399 (23.3) | 3,399 (23.3) |
| | | Age, median (Q1;Q3) | 64.3 (58.2;71.4) | 64.4 (58.5;71.6) |

**Supplementary Table 231**
**Personality disorders - Prevalent cohort 2021**
**Patient characteristics - relatives - not pooled**

| *Age strata of index patients* | *Closest relative type* | *Characteristic of family relatives by agestrata of index patients* | *Closest family relatives of brain disorder patients* | *Comparison relative* |
|---|---|---|---|---|
| | | CCI, n (%) | | |
| | | 0 | 9,713 (66.5) | 10,378 (71.0) |
| | | 1-2 | 3,858 (26.4) | 3,391 (23.2) |
| | | +3 | 1,039 (7.1) | 841 (5.8) |
| | | Education level, n (%) | | |
| | | No/Unknown education | 247 (1.7) | 247 (1.7) |
| | | Primary | 4,392 (30.1) | 4,392 (30.1) |
| | | Secondary | 7,029 (48.1) | 7,029 (48.1) |
| | | Tertiary | 2,942 (20.1) | 2,942 (20.1) |
| | | Relation to patient, n (%) | | |
| | | Mother | 11,211 (76.7) | 11,211 (76.7) |
| | | Father | 3,399 (23.3) | 3,399 (23.3) |
| Young adults and adults (25-64 years) | Partners | Number of persons, n | 19,899 (100.0) | 19,899 (100.0) |
| | | Male, n (%) | 15,240 (76.6) | 15,240 (76.6) |
| | | Age, median (Q1;Q3) | 40.7 (33.1;49.7) | 40.7 (33.3;49.6) |
| | | CCI, n (%) | | |
| | | 0 | 17,612 (88.5) | 17,959 (90.3) |
| | | 1-2 | 2,020 (10.2) | 1,716 (8.6) |
| | | +3 | 267 (1.3) | 224 (1.1) |
| | | Education level, n (%) | | |
| | | No/Unknown education | 325 (1.6) | 325 (1.6) |
| | | Primary | 5,115 (25.7) | 5,115 (25.7) |
| | | Secondary | 9,795 (49.2) | 9,795 (49.2) |
| | | Tertiary | 4,664 (23.4) | 4,664 (23.4) |
| | | Relation to patient, n (%) | | |
| | | Partner | 19,899 (100.0) | 19,899 (100.0) |
| Young adults and adults (25-64 years) | Siblings | Number of persons, n | 4,013 (100.0) | 4,013 (100.0) |
| | | Male, n (%) | 2,321 (57.8) | 2,321 (57.8) |
| | | Age, median (Q1;Q3) | 50.0 (35.2;57.7) | 50.1 (34.8;57.9) |
| | | CCI, n (%) | | |
| | | 0 | 3,351 (83.5) | 3,437 (85.6) |
| | | 1-2 | 573 (14.3) | 500 (12.5) |
| | | +3 | 89 (2.2) | 76 (1.9) |
| | | Education level, n (%) | | |
| | | No/Unknown education | 10 (0.2) | 10 (0.2) |
| | | Primary | 1,101 (27.4) | 1,101 (27.4) |
| | | Secondary | 2,058 (51.3) | 2,058 (51.3) |
| | | Tertiary | 844 (21.0) | 844 (21.0) |
| | | Relation to patient, n (%) | | |
| | | Sibling | 4,013 (100.0) | 4,013 (100.0) |
| Older people (65+ years) | Children | Number of persons, n | 1,799 (100.0) | 1,799 (100.0) |
| | | Male, n (%) | 899 (50.0) | 899 (50.0) |
| | | Age, median (Q1;Q3) | 47.2 (41.7;52.9) | 47.1 (41.8;52.8) |
| | | CCI, n (%) | | |
| | | 0 | 1,565 (87.0) | 1,563 (86.9) |

#### Supplementary Table 231
#### Personality disorders - Prevalent cohort 2021
#### Patient characteristics - relatives - not pooled

| *Age strata of index patients* | *Closest relative type* | *Characteristic of family relatives by agestrata of index patients* | *Closest family relatives of brain disorder patients* | *Comparison relative* |
|---|---|---|---|---|
| | | 1-2 | 217 (12.1) | 209 (11.6) |
| | | +3 | 17 (0.9) | 27 (1.5) |
| | | Education level, n (%) | | |
| | | No/Unknown education | 0 (0.0) | 0 (0.0) |
| | | Primary | 344 (19.1) | 344 (19.1) |
| | | Secondary | 815 (45.3) | 815 (45.3) |
| | | Tertiary | 640 (35.6) | 640 (35.6) |
| | | Relation to patient, n (%) | | |
| | | Child | 1,799 (100.0) | 1,799 (100.0) |
| Older people (65+ years) | Partners | Number of persons, n | 1,253 (100.0) | 1,253 (100.0) |
| | | Male, n (%) | 840 (67.0) | 840 (67.0) |
| | | Age, median (Q1;Q3) | 71.7 (67.4;75.9) | 71.4 (67.5;75.8) |
| | | CCI, n (%) | | |
| | | 0 | 696 (55.5) | 756 (60.3) |
| | | 1-2 | 409 (32.6) | 377 (30.1) |
| | | +3 | 148 (11.8) | 120 (9.6) |
| | | Education level, n (%) | | |
| | | No/Unknown education | 79 (6.3) | 79 (6.3) |
| | | Primary | 292 (23.3) | 292 (23.3) |
| | | Secondary | 607 (48.4) | 607 (48.4) |
| | | Tertiary | 275 (21.9) | 275 (21.9) |
| | | Relation to patient, n (%) | | |
| | | Partner | 1,253 (100.0) | 1,253 (100.0) |
| Older people (65+ years) | Siblings | Number of persons, n | 208 (100.0) | 208 (100.0) |
| | | Male, n (%) | 104 (50.0) | 104 (50.0) |
| | | Age, median (Q1;Q3) | 62.6 (59.0;64.9) | 62.6 (59.8;65.1) |
| | | CCI, n (%) | | |
| | | 0 | 155 (74.5) | 153 (73.6) |
| | | 1-2 | 41 (19.7) | 45 (21.6) |
| | | +3 | 12 (5.8) | 10 (4.8) |
| | | Education level, n (%) | | |
| | | No/Unknown education | 0 (0.0) | 0 (0.0) |
| | | Primary | 46 (22.1) | 46 (22.1) |
| | | Secondary | 118 (56.7) | 118 (56.7) |
| | | Tertiary | 44 (21.2) | 44 (21.2) |
| | | Relation to patient, n (%) | | |
| | | Sibling | 208 (100.0) | 208 (100.0) |

#### Supplementary Table 232
#### Personality disorders - Prevalent cohort 2021
#### Cost analysis - pooled

| *Age strata of index patients* | *Closest relative type* | *Number of index patients included in cost component calculations* | *Cost component* | *Actual costs* | | *Attributable costs* | |
|---|---|---|---|---|---|---|---|
| | | | | *Total (million EUR)* | *Per person (EUR)* | *Total (million EUR)* | *Per person (EUR)* |
| Children and young people (0-24 years) | Fathers | 5,152 | Primary sector | 1.4 | 273 | 0.2 | 32 |
| | | 5,152 | Hospital care | 11.9 | 2,325 | 3.1 | 601 |
| | | 5,152 | Filled prescriptions | 1.5 | 287 | 0.4 | 84 |
| | | 4,834 | Lost productivity (income loss) | . | . | 55.4 | 11,460 |
| Children and young people (0-24 years) | Mothers | 5,371 | Primary sector | 2.1 | 401 | 0.3 | 53 |
| | | 5,371 | Hospital care | 14.2 | 2,656 | 5.6 | 1,039 |
| | | 5,371 | Filled prescriptions | 1.8 | 327 | 0.6 | 105 |
| | | 5,290 | Lost productivity (income loss) | . | . | 50.6 | 9,557 |
| Young adults and adults (25-64 years) | Pooled | 47,515 | Primary sector | 15.1 | 319 | 1.7 | 35 |
| | | 47,515 | Hospital care | 108.4 | 2,291 | 23.7 | 501 |
| | | 47,515 | Filled prescriptions | 14.3 | 303 | 3.6 | 75 |
| | | 32,752 | Lost productivity (income loss) | . | . | 298.2 | 9,105 |
| Older people (65+ years) | Pooled | 3,260 | Primary sector | 1.3 | 398 | 0.1 | 44 |
| | | 3,260 | Hospital care | 10.4 | 3,223 | 1.6 | 500 |
| | | 3,260 | Filled prescriptions | 1.3 | 400 | 0.3 | 80 |
| | | 1,945 | Lost productivity (income loss) | . | . | 12.3 | 6,334 |

#### Supplementary Table 233
#### Personality disorders - Prevalent cohort 2021
#### Cost analysis - not pooled

| *Age strata of index patients* | *Closest relative type* | *Number of index patients included in cost component calculations* | *Cost component* | *Actual costs* | | *Attributable costs* | |
|---|---|---|---|---|---|---|---|
| | | | | *Total (million EUR)* | *Per person (EUR)* | *Total (million EUR)* | *Per person (EUR)* |
| Children and young people (0-24 years) | Fathers | 5,152 | Primary sector | 1.4 | 273 | 0.2 | 32 |
| | | 5,152 | Hospital care | 11.9 | 2,325 | 3.1 | 601 |
| | | 5,152 | Filled prescriptions | 1.5 | 287 | 0.4 | 84 |
| | | 4,834 | Lost productivity (income loss) | . | . | 55.4 | 11,460 |
| Children and young people (0-24 years) | Mothers | 5,371 | Primary sector | 2.1 | 401 | 0.3 | 53 |
| | | 5,371 | Hospital care | 14.2 | 2,656 | 5.6 | 1,039 |
| | | 5,371 | Filled prescriptions | 1.8 | 327 | 0.6 | 105 |
| | | 5,290 | Lost productivity (income loss) | . | . | 50.6 | 9,557 |
| Young adults and adults (25-64 years) | Children | 8,993 | Primary sector | 2.0 | 218 | 0.3 | 29 |
| | | 8,993 | Hospital care | 14.8 | 1,647 | 5.5 | 615 |
| | | 8,993 | Filled prescriptions | 1.1 | 120 | 0.3 | 29 |
| | | 3,745 | Lost productivity (income loss) | . | . | 15.4 | 4,108 |
| Young adults and adults (25-64 years) | Parents | 14,610 | Primary sector | 6.8 | 469 | 0.6 | 44 |
| | | 14,610 | Hospital care | 53.8 | 3,718 | 10.7 | 740 |
| | | 14,610 | Filled prescriptions | 7.8 | 537 | 1.7 | 117 |
| | | 6,318 | Lost productivity (income loss) | . | . | 45.0 | 7,127 |
| Young adults and adults (25-64 years) | Partners | 19,899 | Primary sector | 5.1 | 258 | 0.7 | 34 |
| | | 19,899 | Hospital care | 31.3 | 1,577 | 6.6 | 333 |
| | | 19,899 | Filled prescriptions | 4.4 | 221 | 1.4 | 71 |
| | | 19,116 | Lost productivity (income loss) | . | . | 211.7 | 11,072 |
| Young adults and adults (25-64 years) | Siblings | 4,013 | Primary sector | 1.2 | 307 | 0.1 | 23 |
| | | 4,013 | Hospital care | 8.5 | 2,129 | 0.9 | 216 |
| | | 4,013 | Filled prescriptions | 1.1 | 270 | 0.2 | 44 |
| | | 3,573 | Lost productivity (income loss) | . | . | 26.1 | 7,316 |
| Older people (65+ years) | Children | 1,799 | Primary sector | 0.6 | 323 | 0.1 | 48 |
| | | 1,799 | Hospital care | 4.1 | 2,305 | 1.0 | 531 |
| | | 1,799 | Filled prescriptions | 0.4 | 237 | 0.0 | 21 |
| | | 1,739 | Lost productivity (income loss) | . | . | 10.7 | 6,168 |
| Older people (65+ years) | Partners | 1,253 | Primary sector | 0.6 | 502 | 0.0 | 34 |
| | | 1,253 | Hospital care | 5.3 | 4,285 | 0.3 | 239 |
| | | 1,253 | Filled prescriptions | 0.8 | 614 | 0.2 | 133 |
| | | 94 | Lost productivity (income loss) | . | . | 0.9 | 9,107 |
| Older people (65+ years) | Siblings | 208 | Primary sector | 0.1 | 438 | 0.0 | 60 |
| | | 208 | Hospital care | 1.0 | 4,845 | 0.4 | 1,783 |
| | | 208 | Filled prescriptions | 0.1 | 542 | 0.1 | 280 |
| | | 112 | Lost productivity (income loss) | . | . | 0.7 | 6,587 |

**Supplementary Table 234**
**Polyneuropathy - Incident cohort 2016-2021**
**Identification of populations of relatives**

| ***Agegroup of index patients in matched population*** | | ***Closest relative type identified*** | | ***Available control with same type of closest relative*** |
|---|---|---|---|---|
| Children and young people (0-24 years) | n = 514 | Fathers | n = 408 (79%) | 96% |
| | | Mothers | n = 422 (82%) | 96% |
| | | No available closest relative | n = 42 (8%) | n/a |
| Young adults and adults (25-64 years) | n = 9,549 | Children | n = 1,465 (15%) | 91% |
| | | Parents | n = 1,010 (11%) | 86% |
| | | Partners | n = 5,674 (59%) | 95% |
| | | Siblings | n = 1,122 (12%) | 60% |
| | | Pooled | n = 9,271 (97%) | 89% |
| | | No available closest relative | n = 278 (3%) | n/a |
| Older people (65+ years) | n = 13,721 | Children | n = 4,422 (32%) | 94% |
| | | Partners | n = 7,912 (58%) | 96% |
| | | Siblings | n = 247 (2%) | 39% |
| | | Pooled | n = 12,581 (92%) | 94% |
| | | No available closest relative | n = 1,140 (8%) | n/a |

**Supplementary Table 235**
**Polyneuropathy - Incident cohort 2016-2021**
**Patient characteristics - cases**

| ***Age strata of index patients*** | ***Characteristic*** | |
|---|---|---|
| Children and young people (0-24 years) | Number of persons, n | 457 (100.0) |
| | Male, n (%) | 227 (49.7) |
| | Age, median (Q1;Q3) | 18.3 (12.7;22.0) |
| | CCI, n (%) | |
| | 0 | 361 (79.0) |
| | 1-2 | 88 (19.3) |
| | +3 | 8 (1.8) |
| | Education level, n (%) | |
| | No/Unknown education | 125 (27.4) |
| | Primary | 227 (49.7) |
| | Secondary | 97 (21.2) |
| | Tertiary | 8 (1.8) |
| Young adults and adults (25-64 years) | Number of persons, n | 8,306 (100.0) |
| | Male, n (%) | 4,875 (58.7) |
| | Age, median (Q1;Q3) | 55.3 (48.1;60.4) |
| | CCI, n (%) | |
| | 0 | 3,838 (46.2) |
| | 1-2 | 3,083 (37.1) |
| | +3 | 1,385 (16.7) |
| | Education level, n (%) | |
| | No/Unknown education | 255 (3.1) |
| | Primary | 2,306 (27.8) |
| | Secondary | 4,023 (48.4) |
| | Tertiary | 1,722 (20.7) |
| Older people (65+ years) | Number of persons, n | 11,925 (100.0) |
| | Male, n (%) | 7,190 (60.3) |
| | Age, median (Q1;Q3) | 74.8 (70.4;79.9) |
| | CCI, n (%) | |
| | 0 | 3,893 (32.6) |
| | 1-2 | 4,503 (37.8) |
| | +3 | 3,529 (29.6) |
| | Education level, n (%) | |
| | No/Unknown education | 737 (6.2) |
| | Primary | 3,736 (31.3) |
| | Secondary | 5,442 (45.6) |
| | Tertiary | 2,010 (16.9) |

**Supplementary Table 236**
**Polyneuropathy - Incident cohort 2016-2021**
**Patient characteristics - relatives - pooled**

| *Age strata of index patients* | *Closest relative type* | *Characteristic of family relatives by agestrata of index patients* | *Closest family relatives of brain disorder patients* | *Comparison relative* |
|---|---|---|---|---|
| Children and young people (0-24 years) | Fathers | Number of persons, n | 394 (100.0) | 394 (100.0) |
| | | Male, n (%) | 394 (100.0) | 394 (100.0) |
| | | Age, median (Q1;Q3) | 49.3 (44.3;53.8) | 49.8 (44.2;54.3) |
| | | CCI, n (%) | | |
| | | 0 | 345 (87.6) | 351 (89.1) |
| | | 1-2 | 40 (10.2) | 37 (9.4) |
| | | +3 | 9 (2.3) | 6 (1.5) |
| | | Education level, n (%) | | |
| | | No/Unknown education | 8 (2.0) | 8 (2.0) |
| | | Primary | 83 (21.1) | 83 (21.1) |
| | | Secondary | 210 (53.3) | 210 (53.3) |
| | | Tertiary | 93 (23.6) | 93 (23.6) |
| | | Relation to patient, n (%) | | |
| | | Father | 394 (100.0) | 394 (100.0) |
| Children and young people (0-24 years) | Mothers | Number of persons, n | 409 (100.0) | 409 (100.0) |
| | | Male, n (%) | 0 (0.0) | 0 (0.0) |
| | | Age, median (Q1;Q3) | 47.2 (41.7;50.9) | 47.4 (42.6;51.2) |
| | | CCI, n (%) | | |
| | | 0 | 357 (87.3) | 365 (89.2) |
| | | 1-2 | 45 (11.0) | 39 (9.5) |
| | | +3 | 7 (1.7) | 5 (1.2) |
| | | Education level, n (%) | | |
| | | No/Unknown education | 7 (1.7) | 7 (1.7) |
| | | Primary | 79 (19.3) | 79 (19.3) |
| | | Secondary | 175 (42.8) | 175 (42.8) |
| | | Tertiary | 148 (36.2) | 148 (36.2) |
| | | Relation to patient, n (%) | | |
| | | Mother | 409 (100.0) | 409 (100.0) |
| Young adults and adults (25-64 years) | Pooled | Number of persons, n | 8,306 (100.0) | 8,306 (100.0) |
| | | Male, n (%) | 3,596 (43.3) | 3,596 (43.3) |
| | | Age, median (Q1;Q3) | 54.1 (41.8;61.3) | 54.1 (41.9;61.0) |
| | | CCI, n (%) | | |
| | | 0 | 6,576 (79.2) | 6,877 (82.8) |
| | | 1-2 | 1,414 (17.0) | 1,192 (14.4) |
| | | +3 | 316 (3.8) | 237 (2.9) |
| | | Education level, n (%) | | |
| | | No/Unknown education | 302 (3.6) | 302 (3.6) |
| | | Primary | 2,296 (27.6) | 2,296 (27.6) |
| | | Secondary | 3,910 (47.1) | 3,910 (47.1) |
| | | Tertiary | 1,798 (21.6) | 1,798 (21.6) |
| | | Relation to patient, n (%) | | |
| | | Mother | 709 (8.5) | 709 (8.5) |
| | | Father | 165 (2.0) | 165 (2.0) |
| | | Partner | 5,405 (65.1) | 5,405 (65.1) |
| | | Child | 1,347 (16.2) | 1,347 (16.2) |

#### Supplementary Table 236
#### Polyneuropathy - Incident cohort 2016-2021
#### Patient characteristics - relatives - pooled

| *Age strata of index patients* | *Closest relative type* | *Characteristic of family relatives by agestrata of index patients* | *Closest family relatives of brain disorder patients* | *Comparison relative* |
|---|---|---|---|---|
| | | Sibling | 680 (8.2) | 680 (8.2) |
| Older people (65+ years) | Pooled | Number of persons, n | 11,925 (100.0) | 11,925 (100.0) |
| | | Male, n (%) | 4,536 (38.0) | 4,536 (38.0) |
| | | Age, median (Q1;Q3) | 67.7 (55.9;74.7) | 67.8 (55.8;74.6) |
| | | CCI, n (%) | | |
| | | 0 | 8,238 (69.1) | 8,521 (71.5) |
| | | 1-2 | 2,835 (23.8) | 2,693 (22.6) |
| | | +3 | 852 (7.1) | 711 (6.0) |
| | | Education level, n (%) | | |
| | | No/Unknown education | 347 (2.9) | 347 (2.9) |
| | | Primary | 2,922 (24.5) | 2,922 (24.5) |
| | | Secondary | 5,753 (48.2) | 5,753 (48.2) |
| | | Tertiary | 2,903 (24.3) | 2,903 (24.3) |
| | | Relation to patient, n (%) | | |
| | | Partner | 7,660 (64.2) | 7,660 (64.2) |
| | | Child | 4,167 (34.9) | 4,167 (34.9) |
| | | Sibling | 98 (0.8) | 98 (0.8) |

**Supplementary Table 237**
**Polyneuropathy - Incident cohort 2016-2021**
**Patient characteristics - relatives - not pooled**

| *Age strata of index patients* | *Closest relative type* | *Characteristic of family relatives by agestrata of index patients* | *Closest family relatives of brain disorder patients* | *Comparison relative* |
|---|---|---|---|---|
| Children and young people (0-24 years) | Fathers | Number of persons, n | 394 (100.0) | 394 (100.0) |
| | | Male, n (%) | 394 (100.0) | 394 (100.0) |
| | | Age, median (Q1;Q3) | 49.3 (44.3;53.8) | 49.8 (44.2;54.3) |
| | | CCI, n (%) | | |
| | | 0 | 345 (87.6) | 351 (89.1) |
| | | 1-2 | 40 (10.2) | 37 (9.4) |
| | | +3 | 9 (2.3) | 6 (1.5) |
| | | Education level, n (%) | | |
| | | No/Unknown education | 8 (2.0) | 8 (2.0) |
| | | Primary | 83 (21.1) | 83 (21.1) |
| | | Secondary | 210 (53.3) | 210 (53.3) |
| | | Tertiary | 93 (23.6) | 93 (23.6) |
| | | Relation to patient, n (%) | | |
| | | Father | 394 (100.0) | 394 (100.0) |
| Children and young people (0-24 years) | Mothers | Number of persons, n | 409 (100.0) | 409 (100.0) |
| | | Male, n (%) | 0 (0.0) | 0 (0.0) |
| | | Age, median (Q1;Q3) | 47.2 (41.7;50.9) | 47.4 (42.6;51.2) |
| | | CCI, n (%) | | |
| | | 0 | 357 (87.3) | 365 (89.2) |
| | | 1-2 | 45 (11.0) | 39 (9.5) |
| | | +3 | 7 (1.7) | 5 (1.2) |
| | | Education level, n (%) | | |
| | | No/Unknown education | 7 (1.7) | 7 (1.7) |
| | | Primary | 79 (19.3) | 79 (19.3) |
| | | Secondary | 175 (42.8) | 175 (42.8) |
| | | Tertiary | 148 (36.2) | 148 (36.2) |
| | | Relation to patient, n (%) | | |
| | | Mother | 409 (100.0) | 409 (100.0) |
| Young adults and adults (25-64 years) | Children | Number of persons, n | 1,347 (100.0) | 1,347 (100.0) |
| | | Male, n (%) | 731 (54.3) | 731 (54.3) |
| | | Age, median (Q1;Q3) | 26.7 (18.8;34.2) | 26.4 (18.7;33.9) |
| | | CCI, n (%) | | |
| | | 0 | 1,236 (91.8) | 1,261 (93.6) |
| | | 1-2 | 103 (7.6) | * |
| | | +3 | 8 (0.6) | * |
| | | Education level, n (%) | | |
| | | No/Unknown education | 147 (10.9) | 147 (10.9) |
| | | Primary | 531 (39.4) | 531 (39.4) |
| | | Secondary | 464 (34.4) | 464 (34.4) |
| | | Tertiary | 205 (15.2) | 205 (15.2) |
| | | Relation to patient, n (%) | | |
| | | Child | 1,347 (100.0) | 1,347 (100.0) |
| Young adults and adults (25-64 years) | Parents | Number of persons, n | 874 (100.0) | 874 (100.0) |
| | | Male, n (%) | 165 (18.9) | 165 (18.9) |
| | | Age, median (Q1;Q3) | 73.6 (66.0;79.7) | 73.8 (66.0;79.9) |

### Supplementary Table 237
### Polyneuropathy - Incident cohort 2016-2021
### Patient characteristics - relatives - not pooled

| *Age strata of index patients* | *Closest relative type* | *Characteristic of family relatives by agestrata of index patients* | *Closest family relatives of brain disorder patients* | *Comparison relative* |
|---|---|---|---|---|
| | | CCI, n (%) | | |
| | | 0 | 503 (57.6) | 525 (60.1) |
| | | 1-2 | 258 (29.5) | 264 (30.2) |
| | | +3 | 113 (12.9) | 85 (9.7) |
| | | Education level, n (%) | | |
| | | No/Unknown education | 10 (1.1) | 10 (1.1) |
| | | Primary | 375 (42.9) | 375 (42.9) |
| | | Secondary | 378 (43.2) | 378 (43.2) |
| | | Tertiary | 111 (12.7) | 111 (12.7) |
| | | Relation to patient, n (%) | | |
| | | Mother | 709 (81.1) | 709 (81.1) |
| | | Father | 165 (18.9) | 165 (18.9) |
| Young adults and adults (25-64 years) | Partners | Number of persons, n | 5,405 (100.0) | 5,405 (100.0) |
| | | Male, n (%) | 2,282 (42.2) | 2,282 (42.2) |
| | | Age, median (Q1;Q3) | 54.8 (47.7;60.5) | 54.8 (47.7;60.1) |
| | | CCI, n (%) | | |
| | | 0 | 4,320 (79.9) | 4,526 (83.7) |
| | | 1-2 | 925 (17.1) | 745 (13.8) |
| | | +3 | 160 (3.0) | 134 (2.5) |
| | | Education level, n (%) | | |
| | | No/Unknown education | 140 (2.6) | 140 (2.6) |
| | | Primary | 1,212 (22.4) | 1,212 (22.4) |
| | | Secondary | 2,687 (49.7) | 2,687 (49.7) |
| | | Tertiary | 1,366 (25.3) | 1,366 (25.3) |
| | | Relation to patient, n (%) | | |
| | | Partner | 5,405 (100.0) | 5,405 (100.0) |
| Young adults and adults (25-64 years) | Siblings | Number of persons, n | 680 (100.0) | 680 (100.0) |
| | | Male, n (%) | 418 (61.5) | 418 (61.5) |
| | | Age, median (Q1;Q3) | 57.0 (52.7;60.5) | 57.0 (52.3;60.9) |
| | | CCI, n (%) | | |
| | | 0 | 517 (76.0) | 565 (83.1) |
| | | 1-2 | 128 (18.8) | 100 (14.7) |
| | | +3 | 35 (5.1) | 15 (2.2) |
| | | Education level, n (%) | | |
| | | No/Unknown education | 5 (0.7) | 5 (0.7) |
| | | Primary | 178 (26.2) | 178 (26.2) |
| | | Secondary | 381 (56.0) | 381 (56.0) |
| | | Tertiary | 116 (17.1) | 116 (17.1) |
| | | Relation to patient, n (%) | | |
| | | Sibling | 680 (100.0) | 680 (100.0) |
| Older people (65+ years) | Children | Number of persons, n | 4,167 (100.0) | 4,167 (100.0) |
| | | Male, n (%) | 2,136 (51.3) | 2,136 (51.3) |
| | | Age, median (Q1;Q3) | 51.8 (45.7;57.1) | 51.6 (45.7;56.9) |
| | | CCI, n (%) | | |
| | | 0 | 3,501 (84.0) | 3,595 (86.3) |

### Supplementary Table 237
### Polyneuropathy - Incident cohort 2016-2021
### Patient characteristics - relatives - not pooled

| *Age strata of index patients* | *Closest relative type* | *Characteristic of family relatives by agestrata of index patients* | *Closest family relatives of brain disorder patients* | *Comparison relative* |
|---|---|---|---|---|
| | | 1-2 | 573 (13.8) | 510 (12.2) |
| | | +3 | 93 (2.2) | 62 (1.5) |
| | | Education level, n (%) | | |
| | | No/Unknown education | 12 (0.3) | 12 (0.3) |
| | | Primary | 712 (17.1) | 712 (17.1) |
| | | Secondary | 2,128 (51.1) | 2,128 (51.1) |
| | | Tertiary | 1,315 (31.6) | 1,315 (31.6) |
| | | Relation to patient, n (%) | | |
| | | Child | 4,167 (100.0) | 4,167 (100.0) |
| Older people (65+ years) | Partners | Number of persons, n | 7,660 (100.0) | 7,660 (100.0) |
| | | Male, n (%) | 2,340 (30.5) | 2,340 (30.5) |
| | | Age, median (Q1;Q3) | 72.7 (68.2;77.4) | 72.7 (68.3;77.2) |
| | | CCI, n (%) | | |
| | | 0 | 4,672 (61.0) | 4,850 (63.3) |
| | | 1-2 | 2,236 (29.2) | 2,162 (28.2) |
| | | +3 | 752 (9.8) | 648 (8.5) |
| | | Education level, n (%) | | |
| | | No/Unknown education | 333 (4.3) | 333 (4.3) |
| | | Primary | 2,194 (28.6) | 2,194 (28.6) |
| | | Secondary | 3,561 (46.5) | 3,561 (46.5) |
| | | Tertiary | 1,572 (20.5) | 1,572 (20.5) |
| | | Relation to patient, n (%) | | |
| | | Partner | 7,660 (100.0) | 7,660 (100.0) |
| Older people (65+ years) | Siblings | Number of persons, n | 98 (100.0) | 98 (100.0) |
| | | Male, n (%) | 60 (61.2) | 60 (61.2) |
| | | Age, median (Q1;Q3) | 61.2 (59.8;64.9) | 61.7 (59.1;64.5) |
| | | CCI, n (%) | | |
| | | 0 | 65 (66.3) | 76 (77.6) |
| | | 1-2 | 26 (26.5) | * |
| | | +3 | 7 (7.1) | * |
| | | Education level, n (%) | | |
| | | No/Unknown education | * | * |
| | | Primary | 16 (16.3) | 16 (16.3) |
| | | Secondary | 64 (65.3) | 64 (65.3) |
| | | Tertiary | * | * |
| | | Relation to patient, n (%) | | |
| | | Sibling | 98 (100.0) | 98 (100.0) |

#### Supplementary Table 238
#### Polyneuropathy - Incident cohort 2016-2021
#### Cost analysis - pooled

| | | | | *Actual costs* | | *Attributable costs* | |
|---|---|---|---|---|---|---|---|
| *Age strata of index patients* | *Closest relative type* | *Number of index patients included in cost component calculations* | *Cost component* | *Total (million EUR)* | *Per person (EUR)* | *Total (million EUR)* | *Per person (EUR)* |
| Children and young people (0-24 years) | Fathers | 394 | Primary sector | 0.1 | 226 | 0.0 | 6 |
| | | 394 | Hospital care | 0.7 | 1,897 | 0.3 | 721 |
| | | 394 | Filled prescriptions | 0.1 | 228 | 0.0 | 77 |
| | | 381 | Lost productivity (income loss) | . | . | 1.2 | 3,159 |
| Children and young people (0-24 years) | Mothers | 409 | Primary sector | 0.1 | 333 | 0.0 | 2 |
| | | 409 | Hospital care | 0.6 | 1,550 | 0.0 | 95 |
| | | 409 | Filled prescriptions | 0.1 | 172 | -0.0 | -2 |
| | | 408 | Lost productivity (income loss) | . | . | -0.1 | -181 |
| Young adults and adults (25-64 years) | Pooled | 8,306 | Primary sector | 2.8 | 337 | 0.2 | 25 |
| | | 8,306 | Hospital care | 20.2 | 2,436 | 2.7 | 331 |
| | | 8,306 | Filled prescriptions | 2.8 | 339 | 0.7 | 90 |
| | | 6,227 | Lost productivity (income loss) | . | . | 35.0 | 5,627 |
| Older people (65+ years) | Pooled | 11,925 | Primary sector | 5.3 | 445 | 0.3 | 27 |
| | | 11,925 | Hospital care | 38.5 | 3,253 | 1.4 | 114 |
| | | 11,925 | Filled prescriptions | 5.6 | 478 | 0.8 | 72 |
| | | 4,361 | Lost productivity (income loss) | . | . | 22.1 | 5,075 |

**Supplementary Table 239**
**Polyneuropathy - Incident cohort 2016-2021**
**Cost analysis - not pooled**

| *Age strata of index patients* | *Closest relative type* | *Number of index patients included in cost component calculations* | *Cost component* | *Actual costs* | | *Attributable costs* | |
|---|---|---|---|---|---|---|---|
| | | | | *Total (million EUR)* | *Per person (EUR)* | *Total (million EUR)* | *Per person (EUR)* |
| Children and young people (0-24 years) | Fathers | 394 | Primary sector | 0.1 | 226 | 0.0 | 6 |
| | | 394 | Hospital care | 0.7 | 1,897 | 0.3 | 721 |
| | | 394 | Filled prescriptions | 0.1 | 228 | 0.0 | 77 |
| | | 381 | Lost productivity (income loss) | . | . | 1.2 | 3,159 |
| Children and young people (0-24 years) | Mothers | 409 | Primary sector | 0.1 | 333 | 0.0 | 2 |
| | | 409 | Hospital care | 0.6 | 1,550 | 0.0 | 95 |
| | | 409 | Filled prescriptions | 0.1 | 172 | -0.0 | -2 |
| | | 408 | Lost productivity (income loss) | . | . | -0.1 | -181 |
| Young adults and adults (25-64 years) | Children | 1,347 | Primary sector | 0.3 | 242 | 0.0 | 23 |
| | | 1,347 | Hospital care | 2.4 | 1,754 | 0.8 | 617 |
| | | 1,347 | Filled prescriptions | 0.2 | 161 | 0.1 | 38 |
| | | 1,017 | Lost productivity (income loss) | . | . | 1.0 | 940 |
| Young adults and adults (25-64 years) | Parents | 874 | Primary sector | 0.4 | 505 | 0.0 | 34 |
| | | 874 | Hospital care | 3.6 | 4,209 | 0.4 | 454 |
| | | 874 | Filled prescriptions | 0.6 | 691 | 0.1 | 145 |
| | | 142 | Lost productivity (income loss) | . | . | 0.2 | 1,477 |
| Young adults and adults (25-64 years) | Partners | 5,405 | Primary sector | 1.8 | 335 | 0.1 | 27 |
| | | 5,405 | Hospital care | 12.3 | 2,281 | 1.0 | 185 |
| | | 5,405 | Filled prescriptions | 1.7 | 320 | 0.5 | 88 |
| | | 4,504 | Lost productivity (income loss) | . | . | 31.5 | 6,986 |
| Young adults and adults (25-64 years) | Siblings | 680 | Primary sector | 0.2 | 320 | 0.0 | 5 |
| | | 680 | Hospital care | 1.9 | 2,773 | 0.5 | 776 |
| | | 680 | Filled prescriptions | 0.3 | 395 | 0.1 | 139 |
| | | 564 | Lost productivity (income loss) | . | . | 2.4 | 4,275 |
| Older people (65+ years) | Children | 4,167 | Primary sector | 1.3 | 306 | 0.1 | 12 |
| | | 4,167 | Hospital care | 8.1 | 1,946 | -1.0 | -237 |
| | | 4,167 | Filled prescriptions | 1.1 | 273 | 0.1 | 27 |
| | | 3,892 | Lost productivity (income loss) | . | . | 18.6 | 4,781 |
| Older people (65+ years) | Partners | 7,660 | Primary sector | 4.0 | 523 | 0.3 | 36 |
| | | 7,660 | Hospital care | 30.1 | 3,977 | 2.3 | 301 |
| | | 7,660 | Filled prescriptions | 4.5 | 591 | 0.7 | 99 |
| | | 417 | Lost productivity (income loss) | . | . | 3.0 | 7,161 |
| Older people (65+ years) | Siblings | 98 | Primary sector | 0.0 | 354 | -0.0 | -13 |
| | | 98 | Hospital care | 0.3 | 2,702 | 0.1 | 555 |
| | | 98 | Filled prescriptions | 0.0 | 413 | -0.0 | -117 |
| | | 52 | Lost productivity (income loss) | . | . | 0.5 | 10,295 |

**Supplementary Table 240**
**Polyneuropathy - Prevalent cohort 2021**
**Identification of populations of relatives**

| *Agegroup of index patients in matched population* | | *Closest relative type identified* | | *Available control with same type of closest relative* |
|---|---|---|---|---|
| Children and young people (0-24 years) | n = 746 | Fathers | n = 609 (82%) | 96% |
| | | Mothers | n = 612 (82%) | 96% |
| | | No available closest relative | n = 38 (5%) | n/a |
| Young adults and adults (25-64 years) | n = 16,232 | Children | n = 2,577 (16%) | 92% |
| | | Parents | n = 1,889 (12%) | 87% |
| | | Partners | n = 9,346 (58%) | 95% |
| | | Siblings | n = 2,023 (12%) | 58% |
| | | Pooled | n = 15,835 (98%) | 89% |
| | | No available closest relative | n = 397 (2%) | n/a |
| Older people (65+ years) | n = 26,094 | Children | n = 9,281 (36%) | 94% |
| | | Partners | n = 13,847 (53%) | 96% |
| | | Siblings | n = 607 (2%) | 49% |
| | | Pooled | n = 23,735 (91%) | 94% |
| | | No available closest relative | n = 2,359 (9%) | n/a |

**Supplementary Table 241**
**Polyneuropathy - Prevalent cohort 2021**
**Patient characteristics - cases**

| *Age strata of index patients* | *Characteristic* | |
|---|---|---|
| Children and young people (0-24 years) | Number of persons, n | 683 (100.0) |
| | Male, n (%) | 362 (53.0) |
| | Age, median (Q1;Q3) | 19.2 (13.9;22.4) |
| | CCI, n (%) | |
| | 0 | 538 (78.8) |
| | 1-2 | 122 (17.9) |
| | +3 | 23 (3.4) |
| | Education level, n (%) | |
| | No/Unknown education | 155 (22.7) |
| | Primary | 385 (56.4) |
| | Secondary | 127 (18.6) |
| | Tertiary | 16 (2.3) |
| Young adults and adults (25-64 years) | Number of persons, n | 14,167 (100.0) |
| | Male, n (%) | 7,776 (54.9) |
| | Age, median (Q1;Q3) | 55.4 (47.7;60.6) |
| | CCI, n (%) | |
| | 0 | 7,412 (52.3) |
| | 1-2 | 4,599 (32.5) |
| | +3 | 2,156 (15.2) |
| | Education level, n (%) | |
| | No/Unknown education | 395 (2.8) |
| | Primary | 3,872 (27.3) |
| | Secondary | 6,743 (47.6) |
| | Tertiary | 3,157 (22.3) |
| Older people (65+ years) | Number of persons, n | 22,476 (100.0) |
| | Male, n (%) | 13,217 (58.8) |
| | Age, median (Q1;Q3) | 75.9 (71.0;81.5) |
| | CCI, n (%) | |
| | 0 | 7,322 (32.6) |
| | 1-2 | 8,640 (38.4) |
| | +3 | 6,514 (29.0) |
| | Education level, n (%) | |
| | No/Unknown education | 1,371 (6.1) |
| | Primary | 6,717 (29.9) |
| | Secondary | 10,383 (46.2) |
| | Tertiary | 4,005 (17.8) |

#### Supplementary Table 242
#### Polyneuropathy - Prevalent cohort 2021
#### Patient characteristics - relatives - pooled

| *Age strata of index patients* | *Closest relative type* | *Characteristic of family relatives by agestrata of index patients* | *Closest family relatives of brain disorder patients* | *Comparison relative* |
|---|---|---|---|---|
| Children and young people (0-24 years) | Fathers | Number of persons, n | 588 (100.0) | 588 (100.0) |
| | | Male, n (%) | 588 (100.0) | 588 (100.0) |
| | | Age, median (Q1;Q3) | 50.4 (45.6;54.7) | 50.5 (45.5;54.4) |
| | | CCI, n (%) | | |
| | | 0 | 522 (88.8) | 520 (88.4) |
| | | 1-2 | 59 (10.0) | 60 (10.2) |
| | | +3 | 7 (1.2) | 8 (1.4) |
| | | Education level, n (%) | | |
| | | No/Unknown education | * | * |
| | | Primary | * | * |
| | | Secondary | 289 (49.1) | 289 (49.1) |
| | | Tertiary | 166 (28.2) | 166 (28.2) |
| | | Relation to patient, n (%) | | |
| | | Father | 588 (100.0) | 588 (100.0) |
| Children and young people (0-24 years) | Mothers | Number of persons, n | 590 (100.0) | 590 (100.0) |
| | | Male, n (%) | 0 (0.0) | 0 (0.0) |
| | | Age, median (Q1;Q3) | 48.4 (43.7;52.8) | 48.5 (44.1;52.6) |
| | | CCI, n (%) | | |
| | | 0 | 518 (87.8) | 512 (86.8) |
| | | 1-2 | 63 (10.7) | 64 (10.8) |
| | | +3 | 9 (1.5) | 14 (2.4) |
| | | Education level, n (%) | | |
| | | No/Unknown education | * | * |
| | | Primary | * | * |
| | | Secondary | 250 (42.4) | 250 (42.4) |
| | | Tertiary | 230 (39.0) | 230 (39.0) |
| | | Relation to patient, n (%) | | |
| | | Mother | 590 (100.0) | 590 (100.0) |
| Young adults and adults (25-64 years) | Pooled | Number of persons, n | 14,167 (100.0) | 14,167 (100.0) |
| | | Male, n (%) | 6,337 (44.7) | 6,337 (44.7) |
| | | Age, median (Q1;Q3) | 54.6 (41.4;61.8) | 54.6 (41.4;61.6) |
| | | CCI, n (%) | | |
| | | 0 | 11,298 (79.7) | 11,669 (82.4) |
| | | 1-2 | 2,418 (17.1) | 2,143 (15.1) |
| | | +3 | 451 (3.2) | 355 (2.5) |
| | | Education level, n (%) | | |
| | | No/Unknown education | 450 (3.2) | 450 (3.2) |
| | | Primary | 3,774 (26.6) | 3,774 (26.6) |
| | | Secondary | 6,821 (48.1) | 6,821 (48.1) |
| | | Tertiary | 3,122 (22.0) | 3,122 (22.0) |
| | | Relation to patient, n (%) | | |
| | | Mother | 1,326 (9.4) | 1,326 (9.4) |
| | | Father | 332 (2.3) | 332 (2.3) |
| | | Partner | 8,931 (63.0) | 8,931 (63.0) |
| | | Child | 2,388 (16.9) | 2,388 (16.9) |

### Supplementary Table 242
### Polyneuropathy - Prevalent cohort 2021
### Patient characteristics - relatives - pooled

| *Age strata of index patients* | *Closest relative type* | *Characteristic of family relatives by agestrata of index patients* | *Closest family relatives of brain disorder patients* | *Comparison relative* |
|---|---|---|---|---|
| | | Sibling | 1,190 (8.4) | 1,190 (8.4) |
| Older people (65+ years) | Pooled | Number of persons, n | 22,476 (100.0) | 22,476 (100.0) |
| | | Male, n (%) | 8,722 (38.8) | 8,722 (38.8) |
| | | Age, median (Q1;Q3) | 66.9 (55.9;75.0) | 66.9 (55.8;74.9) |
| | | CCI, n (%) | | |
| | | 0 | 15,539 (69.1) | 15,987 (71.1) |
| | | 1-2 | 5,374 (23.9) | 5,173 (23.0) |
| | | +3 | 1,563 (7.0) | 1,316 (5.9) |
| | | Education level, n (%) | | |
| | | No/Unknown education | 645 (2.9) | 645 (2.9) |
| | | Primary | 4,991 (22.2) | 4,991 (22.2) |
| | | Secondary | 10,866 (48.3) | 10,866 (48.3) |
| | | Tertiary | 5,974 (26.6) | 5,974 (26.6) |
| | | Relation to patient, n (%) | | |
| | | Partner | 13,371 (59.5) | 13,371 (59.5) |
| | | Child | 8,806 (39.2) | 8,806 (39.2) |
| | | Sibling | 299 (1.3) | 299 (1.3) |

**Supplementary Table 243**
**Polyneuropathy - Prevalent cohort 2021**
**Patient characteristics - relatives - not pooled**

| *Age strata of index patients* | *Closest relative type* | *Characteristic of family relatives by agestrata of index patients* | *Closest family relatives of brain disorder patients* | *Comparison relative* |
|---|---|---|---|---|
| Children and young people (0-24 years) | Fathers | Number of persons, n | 588 (100.0) | 588 (100.0) |
| | | Male, n (%) | 588 (100.0) | 588 (100.0) |
| | | Age, median (Q1;Q3) | 50.4 (45.6;54.7) | 50.5 (45.5;54.4) |
| | | CCI, n (%) | | |
| | | 0 | 522 (88.8) | 520 (88.4) |
| | | 1-2 | 59 (10.0) | 60 (10.2) |
| | | +3 | 7 (1.2) | 8 (1.4) |
| | | Education level, n (%) | | |
| | | No/Unknown education | * | * |
| | | Primary | * | * |
| | | Secondary | 289 (49.1) | 289 (49.1) |
| | | Tertiary | 166 (28.2) | 166 (28.2) |
| | | Relation to patient, n (%) | | |
| | | Father | 588 (100.0) | 588 (100.0) |
| Children and young people (0-24 years) | Mothers | Number of persons, n | 590 (100.0) | 590 (100.0) |
| | | Male, n (%) | 0 (0.0) | 0 (0.0) |
| | | Age, median (Q1;Q3) | 48.4 (43.7;52.8) | 48.5 (44.1;52.6) |
| | | CCI, n (%) | | |
| | | 0 | 518 (87.8) | 512 (86.8) |
| | | 1-2 | 63 (10.7) | 64 (10.8) |
| | | +3 | 9 (1.5) | 14 (2.4) |
| | | Education level, n (%) | | |
| | | No/Unknown education | * | * |
| | | Primary | * | * |
| | | Secondary | 250 (42.4) | 250 (42.4) |
| | | Tertiary | 230 (39.0) | 230 (39.0) |
| | | Relation to patient, n (%) | | |
| | | Mother | 590 (100.0) | 590 (100.0) |
| Young adults and adults (25-64 years) | Children | Number of persons, n | 2,388 (100.0) | 2,388 (100.0) |
| | | Male, n (%) | 1,235 (51.7) | 1,235 (51.7) |
| | | Age, median (Q1;Q3) | 27.0 (19.2;33.7) | 26.7 (19.0;33.5) |
| | | CCI, n (%) | | |
| | | 0 | 2,205 (92.3) | 2,241 (93.8) |
| | | 1-2 | 173 (7.2) | 141 (5.9) |
| | | +3 | 10 (0.4) | 6 (0.3) |
| | | Education level, n (%) | | |
| | | No/Unknown education | 230 (9.6) | 230 (9.6) |
| | | Primary | 983 (41.2) | 983 (41.2) |
| | | Secondary | 798 (33.4) | 798 (33.4) |
| | | Tertiary | 377 (15.8) | 377 (15.8) |
| | | Relation to patient, n (%) | | |
| | | Child | 2,388 (100.0) | 2,388 (100.0) |
| Young adults and adults (25-64 years) | Parents | Number of persons, n | 1,658 (100.0) | 1,658 (100.0) |
| | | Male, n (%) | 332 (20.0) | 332 (20.0) |
| | | Age, median (Q1;Q3) | 73.6 (65.2;79.7) | 73.7 (65.7;79.9) |

### Supplementary Table 243
### Polyneuropathy - Prevalent cohort 2021
### Patient characteristics - relatives - not pooled

| *Age strata of index patients* | *Closest relative type* | *Characteristic of family relatives by agestrata of index patients* | *Closest family relatives of brain disorder patients* | *Comparison relative* |
|---|---|---|---|---|
| | | CCI, n (%) | | |
| | | 0 | 990 (59.7) | 1,041 (62.8) |
| | | 1-2 | 495 (29.9) | 482 (29.1) |
| | | +3 | 173 (10.4) | 135 (8.1) |
| | | Education level, n (%) | | |
| | | No/Unknown education | 23 (1.4) | 23 (1.4) |
| | | Primary | 656 (39.6) | 656 (39.6) |
| | | Secondary | 747 (45.1) | 747 (45.1) |
| | | Tertiary | 232 (14.0) | 232 (14.0) |
| | | Relation to patient, n (%) | | |
| | | Mother | 1,326 (80.0) | 1,326 (80.0) |
| | | Father | 332 (20.0) | 332 (20.0) |
| Young adults and adults (25-64 years) | Partners | Number of persons, n | 8,931 (100.0) | 8,931 (100.0) |
| | | Male, n (%) | 4,080 (45.7) | 4,080 (45.7) |
| | | Age, median (Q1;Q3) | 55.1 (47.4;60.8) | 55.2 (47.5;60.5) |
| | | CCI, n (%) | | |
| | | 0 | 7,164 (80.2) | 7,447 (83.4) |
| | | 1-2 | 1,540 (17.2) | 1,305 (14.6) |
| | | +3 | 227 (2.5) | 179 (2.0) |
| | | Education level, n (%) | | |
| | | No/Unknown education | 192 (2.1) | 192 (2.1) |
| | | Primary | 1,849 (20.7) | 1,849 (20.7) |
| | | Secondary | 4,570 (51.2) | 4,570 (51.2) |
| | | Tertiary | 2,320 (26.0) | 2,320 (26.0) |
| | | Relation to patient, n (%) | | |
| | | Partner | 8,931 (100.0) | 8,931 (100.0) |
| Young adults and adults (25-64 years) | Siblings | Number of persons, n | 1,190 (100.0) | 1,190 (100.0) |
| | | Male, n (%) | 690 (58.0) | 690 (58.0) |
| | | Age, median (Q1;Q3) | 58.2 (53.7;62.2) | 58.2 (53.2;62.3) |
| | | CCI, n (%) | | |
| | | 0 | 939 (78.9) | 940 (79.0) |
| | | 1-2 | 210 (17.6) | 215 (18.1) |
| | | +3 | 41 (3.4) | 35 (2.9) |
| | | Education level, n (%) | | |
| | | No/Unknown education | 5 (0.4) | 5 (0.4) |
| | | Primary | 286 (24.0) | 286 (24.0) |
| | | Secondary | 706 (59.3) | 706 (59.3) |
| | | Tertiary | 193 (16.2) | 193 (16.2) |
| | | Relation to patient, n (%) | | |
| | | Sibling | 1,190 (100.0) | 1,190 (100.0) |
| Older people (65+ years) | Children | Number of persons, n | 8,806 (100.0) | 8,806 (100.0) |
| | | Male, n (%) | 4,427 (50.3) | 4,427 (50.3) |
| | | Age, median (Q1;Q3) | 53.3 (46.7;58.9) | 53.2 (46.7;58.6) |
| | | CCI, n (%) | | |
| | | 0 | 7,274 (82.6) | 7,459 (84.7) |

**Supplementary Table 243**
**Polyneuropathy - Prevalent cohort 2021**
**Patient characteristics - relatives - not pooled**

| *Age strata of index patients* | *Closest relative type* | *Characteristic of family relatives by agestrata of index patients* | *Closest family relatives of brain disorder patients* | *Comparison relative* |
|---|---|---|---|---|
| | | 1-2 | 1,328 (15.1) | 1,183 (13.4) |
| | | +3 | 204 (2.3) | 164 (1.9) |
| | | Education level, n (%) | | |
| | | No/Unknown education | 31 (0.4) | 31 (0.4) |
| | | Primary | 1,437 (16.3) | 1,437 (16.3) |
| | | Secondary | 4,355 (49.5) | 4,355 (49.5) |
| | | Tertiary | 2,983 (33.9) | 2,983 (33.9) |
| | | Relation to patient, n (%) | | |
| | | Child | 8,806 (100.0) | 8,806 (100.0) |
| Older people (65+ years) | Partners | Number of persons, n | 13,371 (100.0) | 13,371 (100.0) |
| | | Male, n (%) | 4,131 (30.9) | 4,131 (30.9) |
| | | Age, median (Q1;Q3) | 73.7 (68.6;78.2) | 73.6 (68.7;78.2) |
| | | CCI, n (%) | | |
| | | 0 | 8,059 (60.3) | 8,296 (62.0) |
| | | 1-2 | 3,977 (29.7) | 3,934 (29.4) |
| | | +3 | 1,335 (10.0) | 1,141 (8.5) |
| | | Education level, n (%) | | |
| | | No/Unknown education | 613 (4.6) | 613 (4.6) |
| | | Primary | 3,491 (26.1) | 3,491 (26.1) |
| | | Secondary | 6,334 (47.4) | 6,334 (47.4) |
| | | Tertiary | 2,933 (21.9) | 2,933 (21.9) |
| | | Relation to patient, n (%) | | |
| | | Partner | 13,371 (100.0) | 13,371 (100.0) |
| Older people (65+ years) | Siblings | Number of persons, n | 299 (100.0) | 299 (100.0) |
| | | Male, n (%) | 164 (54.8) | 164 (54.8) |
| | | Age, median (Q1;Q3) | 61.8 (59.4;65.5) | 63.0 (59.5;65.4) |
| | | CCI, n (%) | | |
| | | 0 | 206 (68.9) | 232 (77.6) |
| | | 1-2 | 69 (23.1) | 56 (18.7) |
| | | +3 | 24 (8.0) | 11 (3.7) |
| | | Education level, n (%) | | |
| | | No/Unknown education | * | * |
| | | Primary | 63 (21.1) | 63 (21.1) |
| | | Secondary | 177 (59.2) | 177 (59.2) |
| | | Tertiary | * | * |
| | | Relation to patient, n (%) | | |
| | | Sibling | 299 (100.0) | 299 (100.0) |

#### Supplementary Table 244
#### Polyneuropathy - Prevalent cohort 2021
#### Cost analysis - pooled

| *Age strata of index patients* | *Closest relative type* | *Number of index patients included in cost component calculations* | *Cost component* | *Actual costs* | | *Attributable costs* | |
|---|---|---|---|---|---|---|---|
| | | | | *Total (million EUR)* | *Per person (EUR)* | *Total (million EUR)* | *Per person (EUR)* |
| Children and young people (0-24 years) | Fathers | 588 | Primary sector | 0.1 | 229 | 0.0 | 9 |
| | | 588 | Hospital care | 0.8 | 1,283 | 0.1 | 110 |
| | | 588 | Filled prescriptions | 0.1 | 195 | 0.0 | 49 |
| | | 565 | Lost productivity (income loss) | . | . | 4.8 | 8,515 |
| Children and young people (0-24 years) | Mothers | 590 | Primary sector | 0.2 | 338 | -0.0 | -7 |
| | | 590 | Hospital care | 0.7 | 1,168 | -0.5 | -930 |
| | | 590 | Filled prescriptions | 0.1 | 190 | -0.0 | -68 |
| | | 583 | Lost productivity (income loss) | . | . | 0.3 | 472 |
| Young adults and adults (25-64 years) | Pooled | 14,167 | Primary sector | 4.9 | 345 | 0.4 | 32 |
| | | 14,167 | Hospital care | 35.2 | 2,495 | 7.3 | 517 |
| | | 14,167 | Filled prescriptions | 4.5 | 321 | 0.7 | 52 |
| | | 10,537 | Lost productivity (income loss) | . | . | 58.3 | 5,533 |
| Older people (65+ years) | Pooled | 22,476 | Primary sector | 10.1 | 454 | 0.7 | 33 |
| | | 22,476 | Hospital care | 72.6 | 3,256 | 6.8 | 304 |
| | | 22,476 | Filled prescriptions | 10.2 | 459 | 1.3 | 58 |
| | | 8,731 | Lost productivity (income loss) | . | . | 40.3 | 4,620 |

### Supplementary Table 245
### Polyneuropathy - Prevalent cohort 2021
### Cost analysis - not pooled

| Age strata of index patients | Closest relative type | Number of index patients included in cost component calculations | Cost component | Actual costs | | Attributable costs | |
|---|---|---|---|---|---|---|---|
| | | | | Total (million EUR) | Per person (EUR) | Total (million EUR) | Per person (EUR) |
| Children and young people (0-24 years) | Fathers | 588 | Primary sector | 0.1 | 229 | 0.0 | 9 |
| | | 588 | Hospital care | 0.8 | 1,283 | 0.1 | 110 |
| | | 588 | Filled prescriptions | 0.1 | 195 | 0.0 | 49 |
| | | 565 | Lost productivity (income loss) | . | . | 4.8 | 8,515 |
| Children and young people (0-24 years) | Mothers | 590 | Primary sector | 0.2 | 338 | -0.0 | -7 |
| | | 590 | Hospital care | 0.7 | 1,168 | -0.5 | -930 |
| | | 590 | Filled prescriptions | 0.1 | 190 | -0.0 | -68 |
| | | 583 | Lost productivity (income loss) | . | . | 0.3 | 472 |
| Young adults and adults (25-64 years) | Children | 2,388 | Primary sector | 0.6 | 256 | 0.1 | 28 |
| | | 2,388 | Hospital care | 5.7 | 2,376 | 1.7 | 715 |
| | | 2,388 | Filled prescriptions | 0.4 | 154 | 0.0 | 17 |
| | | 1,796 | Lost productivity (income loss) | . | . | 4.6 | 2,574 |
| Young adults and adults (25-64 years) | Parents | 1,658 | Primary sector | 0.8 | 515 | 0.1 | 37 |
| | | 1,658 | Hospital care | 6.7 | 4,134 | 0.7 | 431 |
| | | 1,658 | Filled prescriptions | 1.0 | 631 | 0.1 | 76 |
| | | 318 | Lost productivity (income loss) | . | . | 3.0 | 9,444 |
| Young adults and adults (25-64 years) | Partners | 8,931 | Primary sector | 3.0 | 340 | 0.3 | 33 |
| | | 8,931 | Hospital care | 19.5 | 2,184 | 4.1 | 463 |
| | | 8,931 | Filled prescriptions | 2.7 | 305 | 0.5 | 59 |
| | | 7,519 | Lost productivity (income loss) | . | . | 42.3 | 5,624 |
| Young adults and adults (25-64 years) | Siblings | 1,190 | Primary sector | 0.4 | 330 | 0.0 | 18 |
| | | 1,190 | Hospital care | 3.4 | 2,827 | 0.8 | 637 |
| | | 1,190 | Filled prescriptions | 0.4 | 355 | 0.0 | 41 |
| | | 904 | Lost productivity (income loss) | . | . | 8.4 | 9,275 |
| Older people (65+ years) | Children | 8,806 | Primary sector | 2.9 | 329 | 0.2 | 28 |
| | | 8,806 | Hospital care | 17.7 | 2,012 | -1.4 | -160 |
| | | 8,806 | Filled prescriptions | 2.5 | 283 | 0.3 | 37 |
| | | 7,866 | Lost productivity (income loss) | . | . | 37.4 | 4,755 |
| Older people (65+ years) | Partners | 13,371 | Primary sector | 7.1 | 540 | 0.5 | 36 |
| | | 13,371 | Hospital care | 54.2 | 4,101 | 8.1 | 612 |
| | | 13,371 | Filled prescriptions | 7.6 | 575 | 0.9 | 69 |
| | | 703 | Lost productivity (income loss) | . | . | 1.6 | 2,267 |
| Older people (65+ years) | Siblings | 299 | Primary sector | 0.1 | 376 | 0.0 | 45 |
| | | 299 | Hospital care | 0.7 | 2,478 | 0.1 | 287 |
| | | 299 | Filled prescriptions | 0.1 | 466 | 0.0 | 151 |
| | | 162 | Lost productivity (income loss) | . | . | 1.3 | 8,280 |

### Supplementary Table 246
### Schizophrenia spectrum disorders - Incident cohort 2016-2021
### Identification of populations of relatives

| *Agegroup of index patients in matched population* | | *Closest relative type identified* | | *Available control with same type of closest relative* |
|---|---|---|---|---|
| Children and young people (0-24 years) | n = 9,758 | Fathers | n = 7,945 (81%) | 95% |
| | | Mothers | n = 8,627 (88%) | 95% |
| | | No available closest relative | n = 579 (6%) | n/a |
| Young adults and adults (25-64 years) | n = 9,792 | Children | n = 1,354 (14%) | 92% |
| | | Parents | n = 3,036 (31%) | 91% |
| | | Partners | n = 2,467 (25%) | 93% |
| | | Siblings | n = 2,148 (22%) | 38% |
| | | Pooled | n = 9,005 (92%) | 79% |
| | | No available closest relative | n = 787 (8%) | n/a |
| Older people (65+ years) | n = 2,162 | Children | n = 988 (46%) | 93% |
| | | Partners | n = 513 (24%) | 94% |
| | | Siblings | n = 95 (4%) | 33% |
| | | Pooled | n = 1,596 (74%) | 90% |
| | | No available closest relative | n = 566 (26%) | n/a |

**Supplementary Table 247**
**Schizophrenia spectrum disorders - Incident cohort 2016-2021**
**Patient characteristics - cases**

| *Age strata of index patients* | *Characteristic* | |
|---|---|---|
| Children and young people (0-24 years) | Number of persons, n | 8,774 (100.0) |
| | Male, n (%) | 4,024 (45.9) |
| | Age, median (Q1;Q3) | 19.5 (16.8;21.9) |
| | CCI, n (%) | |
| | 0 | 8,125 (92.6) |
| | 1-2 | 634 (7.2) |
| | +3 | 15 (0.2) |
| | Education level, n (%) | |
| | No/Unknown education | 442 (5.0) |
| | Primary | 6,864 (78.2) |
| | Secondary | 1,404 (16.0) |
| | Tertiary | 64 (0.7) |
| Young adults and adults (25-64 years) | Number of persons, n | 7,162 (100.0) |
| | Male, n (%) | 3,921 (54.7) |
| | Age, median (Q1;Q3) | 37.9 (29.3;49.5) |
| | CCI, n (%) | |
| | 0 | 5,949 (83.1) |
| | 1-2 | 1,036 (14.5) |
| | +3 | 177 (2.5) |
| | Education level, n (%) | |
| | No/Unknown education | 269 (3.8) |
| | Primary | 2,956 (41.3) |
| | Secondary | 2,722 (38.0) |
| | Tertiary | 1,215 (17.0) |
| Older people (65+ years) | Number of persons, n | 1,447 (100.0) |
| | Male, n (%) | 480 (33.2) |
| | Age, median (Q1;Q3) | 75.9 (70.4;82.6) |
| | CCI, n (%) | |
| | 0 | 631 (43.6) |
| | 1-2 | 538 (37.2) |
| | +3 | 278 (19.2) |
| | Education level, n (%) | |
| | No/Unknown education | 96 (6.6) |
| | Primary | 593 (41.0) |
| | Secondary | 556 (38.4) |
| | Tertiary | 202 (14.0) |

### Supplementary Table 248
### Schizophrenia spectrum disorders - Incident cohort 2016-2021
### Patient characteristics - relatives - pooled

| *Age strata of index patients* | *Closest relative type* | *Characteristic of family relatives by agestrata of index patients* | *Closest family relatives of brain disorder patients* | *Comparison relative* |
|---|---|---|---|---|
| Children and young people (0-24 years) | Fathers | Number of persons, n | 7,604 (100.0) | 7,604 (100.0) |
| | | Male, n (%) | 7,604 (100.0) | 7,604 (100.0) |
| | | Age, median (Q1;Q3) | 51.3 (47.1;55.6) | 51.4 (47.5;55.1) |
| | | CCI, n (%) | | |
| | | 0 | 6,437 (84.7) | 6,615 (87.0) |
| | | 1-2 | 1,000 (13.2) | 843 (11.1) |
| | | +3 | 167 (2.2) | 146 (1.9) |
| | | Education level, n (%) | | |
| | | No/Unknown education | 141 (1.9) | 141 (1.9) |
| | | Primary | 1,868 (24.6) | 1,868 (24.6) |
| | | Secondary | 3,708 (48.8) | 3,708 (48.8) |
| | | Tertiary | 1,887 (24.8) | 1,887 (24.8) |
| | | Relation to patient, n (%) | | |
| | | Father | 7,604 (100.0) | 7,604 (100.0) |
| Children and young people (0-24 years) | Mothers | Number of persons, n | 8,249 (100.0) | 8,249 (100.0) |
| | | Male, n (%) | 0 (0.0) | 0 (0.0) |
| | | Age, median (Q1;Q3) | 48.8 (44.9;52.9) | 48.9 (45.2;52.5) |
| | | CCI, n (%) | | |
| | | 0 | 6,902 (83.7) | 7,206 (87.4) |
| | | 1-2 | 1,227 (14.9) | 948 (11.5) |
| | | +3 | 120 (1.5) | 95 (1.2) |
| | | Education level, n (%) | | |
| | | No/Unknown education | 85 (1.0) | 85 (1.0) |
| | | Primary | 1,754 (21.3) | 1,754 (21.3) |
| | | Secondary | 3,653 (44.3) | 3,653 (44.3) |
| | | Tertiary | 2,757 (33.4) | 2,757 (33.4) |
| | | Relation to patient, n (%) | | |
| | | Mother | 8,249 (100.0) | 8,249 (100.0) |
| Young adults and adults (25-64 years) | Pooled | Number of persons, n | 7,162 (100.0) | 7,162 (100.0) |
| | | Male, n (%) | 3,185 (44.5) | 3,185 (44.5) |
| | | Age, median (Q1;Q3) | 50.4 (31.0;59.9) | 50.7 (31.1;59.9) |
| | | CCI, n (%) | | |
| | | 0 | 5,847 (81.6) | 6,039 (84.3) |
| | | 1-2 | 1,073 (15.0) | 932 (13.0) |
| | | +3 | 242 (3.4) | 191 (2.7) |
| | | Education level, n (%) | | |
| | | No/Unknown education | 576 (8.0) | 576 (8.0) |
| | | Primary | 2,206 (30.8) | 2,206 (30.8) |
| | | Secondary | 2,889 (40.3) | 2,889 (40.3) |
| | | Tertiary | 1,491 (20.8) | 1,491 (20.8) |
| | | Relation to patient, n (%) | | |
| | | Mother | 2,097 (29.3) | 2,097 (29.3) |
| | | Father | 679 (9.5) | 679 (9.5) |
| | | Partner | 2,308 (32.2) | 2,308 (32.2) |
| | | Child | 1,248 (17.4) | 1,248 (17.4) |

#### Supplementary Table 248
#### Schizophrenia spectrum disorders - Incident cohort 2016-2021
#### Patient characteristics - relatives - pooled

| *Age strata of index patients* | *Closest relative type* | *Characteristic of family relatives by agestrata of index patients* | *Closest family relatives of brain disorder patients* | *Comparison relative* |
|---|---|---|---|---|
| | | Sibling | 830 (11.6) | 830 (11.6) |
| Older people (65+ years) | Pooled | Number of persons, n | 1,447 (100.0) | 1,447 (100.0) |
| | | Male, n (%) | 753 (52.0) | 753 (52.0) |
| | | Age, median (Q1;Q3) | 58.4 (49.4;69.4) | 58.5 (49.7;69.9) |
| | | CCI, n (%) | | |
| | | 0 | 1,074 (74.2) | 1,075 (74.3) |
| | | 1-2 | 288 (19.9) | 292 (20.2) |
| | | +3 | 85 (5.9) | 80 (5.5) |
| | | Education level, n (%) | | |
| | | No/Unknown education | 36 (2.5) | 36 (2.5) |
| | | Primary | 330 (22.8) | 330 (22.8) |
| | | Secondary | 704 (48.7) | 704 (48.7) |
| | | Tertiary | 377 (26.1) | 377 (26.1) |
| | | Relation to patient, n (%) | | |
| | | Partner | 487 (33.7) | 487 (33.7) |
| | | Child | 928 (64.1) | 928 (64.1) |
| | | Sibling | 32 (2.2) | 32 (2.2) |

#### Supplementary Table 249
#### Schizophrenia spectrum disorders - Incident cohort 2016-2021
#### Patient characteristics - relatives - not pooled

| *Age strata of index patients* | *Closest relative type* | *Characteristic of family relatives by agestrata of index patients* | *Closest family relatives of brain disorder patients* | *Comparison relative* |
|---|---|---|---|---|
| Children and young people (0-24 years) | Fathers | Number of persons, n | 7,604 (100.0) | 7,604 (100.0) |
| | | Male, n (%) | 7,604 (100.0) | 7,604 (100.0) |
| | | Age, median (Q1;Q3) | 51.3 (47.1;55.6) | 51.4 (47.5;55.1) |
| | | CCI, n (%) | | |
| | | 0 | 6,437 (84.7) | 6,615 (87.0) |
| | | 1-2 | 1,000 (13.2) | 843 (11.1) |
| | | +3 | 167 (2.2) | 146 (1.9) |
| | | Education level, n (%) | | |
| | | No/Unknown education | 141 (1.9) | 141 (1.9) |
| | | Primary | 1,868 (24.6) | 1,868 (24.6) |
| | | Secondary | 3,708 (48.8) | 3,708 (48.8) |
| | | Tertiary | 1,887 (24.8) | 1,887 (24.8) |
| | | Relation to patient, n (%) | | |
| | | Father | 7,604 (100.0) | 7,604 (100.0) |
| Children and young people (0-24 years) | Mothers | Number of persons, n | 8,249 (100.0) | 8,249 (100.0) |
| | | Male, n (%) | 0 (0.0) | 0 (0.0) |
| | | Age, median (Q1;Q3) | 48.8 (44.9;52.9) | 48.9 (45.2;52.5) |
| | | CCI, n (%) | | |
| | | 0 | 6,902 (83.7) | 7,206 (87.4) |
| | | 1-2 | 1,227 (14.9) | 948 (11.5) |
| | | +3 | 120 (1.5) | 95 (1.2) |
| | | Education level, n (%) | | |
| | | No/Unknown education | 85 (1.0) | 85 (1.0) |
| | | Primary | 1,754 (21.3) | 1,754 (21.3) |
| | | Secondary | 3,653 (44.3) | 3,653 (44.3) |
| | | Tertiary | 2,757 (33.4) | 2,757 (33.4) |
| | | Relation to patient, n (%) | | |
| | | Mother | 8,249 (100.0) | 8,249 (100.0) |
| Young adults and adults (25-64 years) | Children | Number of persons, n | 1,248 (100.0) | 1,248 (100.0) |
| | | Male, n (%) | 648 (51.9) | 648 (51.9) |
| | | Age, median (Q1;Q3) | 18.6 (10.6;26.8) | 18.1 (10.0;26.7) |
| | | CCI, n (%) | | |
| | | 0 | 1,167 (93.5) | 1,173 (94.0) |
| | | 1-2 | * | 69 (5.5) |
| | | +3 | * | 6 (0.5) |
| | | Education level, n (%) | | |
| | | No/Unknown education | 394 (31.6) | 394 (31.6) |
| | | Primary | 531 (42.5) | 531 (42.5) |
| | | Secondary | 210 (16.8) | 210 (16.8) |
| | | Tertiary | 113 (9.1) | 113 (9.1) |
| | | Relation to patient, n (%) | | |
| | | Child | 1,248 (100.0) | 1,248 (100.0) |
| Young adults and adults (25-64 years) | Parents | Number of persons, n | 2,776 (100.0) | 2,776 (100.0) |
| | | Male, n (%) | 679 (24.5) | 679 (24.5) |
| | | Age, median (Q1;Q3) | 60.6 (55.4;68.2) | 60.6 (55.7;68.2) |

### Supplementary Table 249
### Schizophrenia spectrum disorders - Incident cohort 2016-2021
### Patient characteristics - relatives - not pooled

| *Age strata of index patients* | *Closest relative type* | *Characteristic of family relatives by agestrata of index patients* | *Closest family relatives of brain disorder patients* | *Comparison relative* |
|---|---|---|---|---|
| | | CCI, n (%) | | |
| | | 0 | 1,962 (70.7) | 2,041 (73.5) |
| | | 1-2 | 634 (22.8) | 590 (21.3) |
| | | +3 | 180 (6.5) | 145 (5.2) |
| | | Education level, n (%) | | |
| | | No/Unknown education | 64 (2.3) | 64 (2.3) |
| | | Primary | 772 (27.8) | 772 (27.8) |
| | | Secondary | 1,289 (46.4) | 1,289 (46.4) |
| | | Tertiary | 651 (23.5) | 651 (23.5) |
| | | Relation to patient, n (%) | | |
| | | Mother | 2,097 (75.5) | 2,097 (75.5) |
| | | Father | 679 (24.5) | 679 (24.5) |
| Young adults and adults (25-64 years) | Partners | Number of persons, n | 2,308 (100.0) | 2,308 (100.0) |
| | | Male, n (%) | 1,303 (56.5) | 1,303 (56.5) |
| | | Age, median (Q1;Q3) | 41.0 (31.9;51.5) | 41.0 (31.9;51.5) |
| | | CCI, n (%) | | |
| | | 0 | 1,988 (86.1) | 2,106 (91.2) |
| | | 1-2 | 274 (11.9) | 175 (7.6) |
| | | +3 | 46 (2.0) | 27 (1.2) |
| | | Education level, n (%) | | |
| | | No/Unknown education | 115 (5.0) | 115 (5.0) |
| | | Primary | 640 (27.7) | 640 (27.7) |
| | | Secondary | 984 (42.6) | 984 (42.6) |
| | | Tertiary | 569 (24.7) | 569 (24.7) |
| | | Relation to patient, n (%) | | |
| | | Partner | 2,308 (100.0) | 2,308 (100.0) |
| Young adults and adults (25-64 years) | Siblings | Number of persons, n | 830 (100.0) | 830 (100.0) |
| | | Male, n (%) | 555 (66.9) | 555 (66.9) |
| | | Age, median (Q1;Q3) | 45.3 (31.1;55.9) | 45.3 (31.2;55.9) |
| | | CCI, n (%) | | |
| | | 0 | 730 (88.0) | 719 (86.6) |
| | | 1-2 | 86 (10.4) | 98 (11.8) |
| | | +3 | 14 (1.7) | 13 (1.6) |
| | | Education level, n (%) | | |
| | | No/Unknown education | * | * |
| | | Primary | 263 (31.7) | 263 (31.7) |
| | | Secondary | 406 (48.9) | 406 (48.9) |
| | | Tertiary | * | * |
| | | Relation to patient, n (%) | | |
| | | Sibling | 830 (100.0) | 830 (100.0) |
| Older people (65+ years) | Children | Number of persons, n | 928 (100.0) | 928 (100.0) |
| | | Male, n (%) | 460 (49.6) | 460 (49.6) |
| | | Age, median (Q1;Q3) | 52.2 (45.7;58.1) | 52.2 (45.9;58.3) |
| | | CCI, n (%) | | |
| | | 0 | 781 (84.2) | 770 (83.0) |

**Supplementary Table 249**
**Schizophrenia spectrum disorders - Incident cohort 2016-2021**
**Patient characteristics - relatives - not pooled**

| *Age strata of index patients* | *Closest relative type* | *Characteristic of family relatives by agestrata of index patients* | *Closest family relatives of brain disorder patients* | *Comparison relative* |
|---|---|---|---|---|
| | | 1-2 | 130 (14.0) | 138 (14.9) |
| | | +3 | 17 (1.8) | 20 (2.2) |
| | | Education level, n (%) | | |
| | | No/Unknown education | * | * |
| | | Primary | * | * |
| | | Secondary | 460 (49.6) | 460 (49.6) |
| | | Tertiary | 283 (30.5) | 283 (30.5) |
| | | Relation to patient, n (%) | | |
| | | Child | 928 (100.0) | 928 (100.0) |
| Older people (65+ years) | Partners | Number of persons, n | 487 (100.0) | 487 (100.0) |
| | | Male, n (%) | 273 (56.1) | 273 (56.1) |
| | | Age, median (Q1;Q3) | 73.8 (68.6;79.6) | 73.7 (69.1;79.3) |
| | | CCI, n (%) | | |
| | | 0 | 267 (54.8) | 277 (56.9) |
| | | 1-2 | 152 (31.2) | 151 (31.0) |
| | | +3 | 68 (14.0) | 59 (12.1) |
| | | Education level, n (%) | | |
| | | No/Unknown education | 34 (7.0) | 34 (7.0) |
| | | Primary | 141 (29.0) | 141 (29.0) |
| | | Secondary | 223 (45.8) | 223 (45.8) |
| | | Tertiary | 89 (18.3) | 89 (18.3) |
| | | Relation to patient, n (%) | | |
| | | Partner | 487 (100.0) | 487 (100.0) |
| Older people (65+ years) | Siblings | Number of persons, n | 32 (100.0) | 32 (100.0) |
| | | Male, n (%) | 20 (62.5) | 20 (62.5) |
| | | Age, median (Q1;Q3) | 61.0 (58.5;64.1) | 60.3 (57.4;63.6) |
| | | CCI, n (%) | | |
| | | 0 | 26 (81.3) | * |
| | | 1-2 | 6 (18.8) | * |
| | | +3 | 0 (0.0) | * |
| | | Education level, n (%) | | |
| | | No/Unknown education | 0 (0.0) | 0 (0.0) |
| | | Primary | 6 (18.8) | 6 (18.8) |
| | | Secondary | 21 (65.6) | 21 (65.6) |
| | | Tertiary | 5 (15.6) | 5 (15.6) |
| | | Relation to patient, n (%) | | |
| | | Sibling | 32 (100.0) | 32 (100.0) |

#### Supplementary Table 250
#### Schizophrenia spectrum disorders - Incident cohort 2016-2021
#### Cost analysis - pooled

| | | | | *Actual costs* | | *Attributable costs* | |
|---|---|---|---|---|---|---|---|
| *Age strata of index patients* | *Closest relative type* | *Number of index patients included in cost component calculations* | *Cost component* | *Total (million EUR)* | *Per person (EUR)* | *Total (million EUR)* | *Per person (EUR)* |
| Children and young people (0-24 years) | Fathers | 7,604 | Primary sector | 1.9 | 254 | 0.2 | 22 |
| | | 7,604 | Hospital care | 14.4 | 1,896 | 3.3 | 432 |
| | | 7,604 | Filled prescriptions | 1.9 | 252 | 0.5 | 67 |
| | | 7,254 | Lost productivity (income loss) | . | . | 98.0 | 13,515 |
| Children and young people (0-24 years) | Mothers | 8,249 | Primary sector | 3.2 | 391 | 0.5 | 58 |
| | | 8,249 | Hospital care | 17.1 | 2,072 | 4.4 | 539 |
| | | 8,249 | Filled prescriptions | 2.2 | 271 | 0.6 | 70 |
| | | 8,151 | Lost productivity (income loss) | . | . | 56.3 | 6,906 |
| Young adults and adults (25-64 years) | Pooled | 7,162 | Primary sector | 2.4 | 337 | 0.3 | 42 |
| | | 7,162 | Hospital care | 15.6 | 2,198 | 3.0 | 419 |
| | | 7,162 | Filled prescriptions | 2.1 | 289 | 0.4 | 55 |
| | | 5,085 | Lost productivity (income loss) | . | . | 52.3 | 10,281 |
| Older people (65+ years) | Pooled | 1,447 | Primary sector | 0.6 | 395 | 0.1 | 40 |
| | | 1,447 | Hospital care | 4.0 | 2,819 | -0.5 | -361 |
| | | 1,447 | Filled prescriptions | 0.5 | 359 | -0.0 | -2 |
| | | 878 | Lost productivity (income loss) | . | . | 9.8 | 11,156 |

#### Supplementary Table 251
#### Schizophrenia spectrum disorders - Incident cohort 2016-2021
#### Cost analysis - not pooled

| | | | | *Actual costs* | | *Attributable costs* | |
|---|---|---|---|---|---|---|---|
| *Age strata of index patients* | *Closest relative type* | *Number of index patients included in cost component calculations* | *Cost component* | *Total (million EUR)* | *Per person (EUR)* | *Total (million EUR)* | *Per person (EUR)* |
| Children and young people (0-24 years) | Fathers | 7,604 | Primary sector | 1.9 | 254 | 0.2 | 22 |
| | | 7,604 | Hospital care | 14.4 | 1,896 | 3.3 | 432 |
| | | 7,604 | Filled prescriptions | 1.9 | 252 | 0.5 | 67 |
| | | 7,254 | Lost productivity (income loss) | . | . | 98.0 | 13,515 |
| Children and young people (0-24 years) | Mothers | 8,249 | Primary sector | 3.2 | 391 | 0.5 | 58 |
| | | 8,249 | Hospital care | 17.1 | 2,072 | 4.4 | 539 |
| | | 8,249 | Filled prescriptions | 2.2 | 271 | 0.6 | 70 |
| | | 8,151 | Lost productivity (income loss) | . | . | 56.3 | 6,906 |
| Young adults and adults (25-64 years) | Children | 1,248 | Primary sector | 0.3 | 216 | 0.0 | 22 |
| | | 1,248 | Hospital care | 2.2 | 1,741 | 1.0 | 776 |
| | | 1,248 | Filled prescriptions | 0.1 | 104 | -0.0 | -16 |
| | | 598 | Lost productivity (income loss) | . | . | 2.4 | 3,998 |
| Young adults and adults (25-64 years) | Parents | 2,776 | Primary sector | 1.2 | 431 | 0.1 | 42 |
| | | 2,776 | Hospital care | 8.3 | 3,014 | 1.6 | 573 |
| | | 2,776 | Filled prescriptions | 1.3 | 457 | 0.2 | 89 |
| | | 1,583 | Lost productivity (income loss) | . | . | 11.7 | 7,370 |
| Young adults and adults (25-64 years) | Partners | 2,308 | Primary sector | 0.7 | 316 | 0.2 | 68 |
| | | 2,308 | Hospital care | 4.0 | 1,749 | 0.6 | 244 |
| | | 2,308 | Filled prescriptions | 0.5 | 216 | 0.1 | 63 |
| | | 2,160 | Lost productivity (income loss) | . | . | 35.2 | 16,274 |
| Young adults and adults (25-64 years) | Siblings | 830 | Primary sector | 0.2 | 260 | 0.0 | 1 |
| | | 830 | Hospital care | 1.2 | 1,420 | -0.1 | -142 |
| | | 830 | Filled prescriptions | 0.2 | 210 | 0.0 | 25 |
| | | 744 | Lost productivity (income loss) | . | . | 3.1 | 4,127 |
| Older people (65+ years) | Children | 928 | Primary sector | 0.3 | 325 | 0.0 | 39 |
| | | 928 | Hospital care | 1.8 | 1,907 | -0.3 | -352 |
| | | 928 | Filled prescriptions | 0.2 | 246 | 0.0 | 21 |
| | | 831 | Lost productivity (income loss) | . | . | 8.7 | 10,518 |
| Older people (65+ years) | Partners | 487 | Primary sector | 0.3 | 533 | 0.0 | 42 |
| | | 487 | Hospital care | 2.2 | 4,603 | -0.2 | -344 |
| | | 487 | Filled prescriptions | 0.3 | 584 | -0.0 | -55 |
| | | 25 | Lost productivity (income loss) | . | . | 0.6 | 24,742 |
| Older people (65+ years) | Siblings | 32 | Primary sector | 0.0 | 372 | 0.0 | 27 |
| | | 32 | Hospital care | 0.1 | 2,749 | -0.0 | -873 |
| | | 32 | Filled prescriptions | 0.0 | 272 | 0.0 | 96 |
| | | 22 | Lost productivity (income loss) | . | . | 0.4 | 19,820 |

**Supplementary Table 252**
**Schizophrenia spectrum disorders - Prevalent cohort 2021**
**Identification of populations of relatives**

| *Agegroup of index patients in matched population* | | *Closest relative type identified* | | *Available control with same type of closest relative* |
|---|---|---|---|---|
| Children and young people (0-24 years) | n = 8,528 | Fathers | n = 6,963 (82%) | 95% |
| | | Mothers | n = 7,571 (89%) | 95% |
| | | No available closest relative | n = 410 (5%) | n/a |
| Young adults and adults (25-64 years) | n = 47,279 | Children | n = 5,726 (12%) | 91% |
| | | Parents | n = 15,814 (33%) | 89% |
| | | Partners | n = 11,174 (24%) | 93% |
| | | Siblings | n = 11,700 (25%) | 43% |
| | | Pooled | n = 44,414 (94%) | 78% |
| | | No available closest relative | n = 2,865 (6%) | n/a |
| Older people (65+ years) | n = 9,090 | Children | n = 3,488 (38%) | 94% |
| | | Partners | n = 1,786 (20%) | 94% |
| | | Siblings | n = 1,023 (11%) | 51% |
| | | Pooled | n = 6,297 (69%) | 87% |
| | | No available closest relative | n = 2,793 (31%) | n/a |

#### Supplementary Table 253
#### Schizophrenia spectrum disorders - Prevalent cohort 2021
#### Patient characteristics - cases

| *Age strata of index patients* | *Characteristic* | |
|---|---|---|
| Children and young people (0-24 years) | Number of persons, n | 7,789 (100.0) |
| | Male, n (%) | 3,426 (44.0) |
| | Age, median (Q1;Q3) | 21.5 (19.1;23.3) |
| | CCI, n (%) | |
| | 0 | 7,133 (91.6) |
| | 1-2 | 635 (8.2) |
| | +3 | 21 (0.3) |
| | Education level, n (%) | |
| | No/Unknown education | 189 (2.4) |
| | Primary | 6,129 (78.7) |
| | Secondary | 1,415 (18.2) |
| | Tertiary | 56 (0.7) |
| Young adults and adults (25-64 years) | Number of persons, n | 34,931 (100.0) |
| | Male, n (%) | 18,935 (54.2) |
| | Age, median (Q1;Q3) | 42.2 (32.4;53.0) |
| | CCI, n (%) | |
| | 0 | 27,996 (80.1) |
| | 1-2 | 5,905 (16.9) |
| | +3 | 1,030 (2.9) |
| | Education level, n (%) | |
| | No/Unknown education | 1,085 (3.1) |
| | Primary | 16,911 (48.4) |
| | Secondary | 11,968 (34.3) |
| | Tertiary | 4,967 (14.2) |
| Older people (65+ years) | Number of persons, n | 5,506 (100.0) |
| | Male, n (%) | 1,785 (32.4) |
| | Age, median (Q1;Q3) | 72.2 (68.0;77.6) |
| | CCI, n (%) | |
| | 0 | 2,554 (46.4) |
| | 1-2 | 2,065 (37.5) |
| | +3 | 887 (16.1) |
| | Education level, n (%) | |
| | No/Unknown education | 318 (5.8) |
| | Primary | 2,067 (37.5) |
| | Secondary | 2,247 (40.8) |
| | Tertiary | 874 (15.9) |

**Supplementary Table 254**
**Schizophrenia spectrum disorders - Prevalent cohort 2021**
**Patient characteristics - relatives - pooled**

| *Age strata of index patients* | *Closest relative type* | *Characteristic of family relatives by agestrata of index patients* | *Closest family relatives of brain disorder patients* | *Comparison relative* |
|---|---|---|---|---|
| Children and young people (0-24 years) | Fathers | Number of persons, n | 6,678 (100.0) | 6,678 (100.0) |
| | | Male, n (%) | 6,678 (100.0) | 6,678 (100.0) |
| | | Age, median (Q1;Q3) | 53.1 (48.8;57.0) | 53.1 (49.3;56.7) |
| | | CCI, n (%) | | |
| | | 0 | 5,506 (82.4) | 5,697 (85.3) |
| | | 1-2 | 989 (14.8) | 859 (12.9) |
| | | +3 | 183 (2.7) | 122 (1.8) |
| | | Education level, n (%) | | |
| | | No/Unknown education | 107 (1.6) | 107 (1.6) |
| | | Primary | 1,592 (23.8) | 1,592 (23.8) |
| | | Secondary | 3,338 (50.0) | 3,338 (50.0) |
| | | Tertiary | 1,641 (24.6) | 1,641 (24.6) |
| | | Relation to patient, n (%) | | |
| | | Father | 6,678 (100.0) | 6,678 (100.0) |
| Children and young people (0-24 years) | Mothers | Number of persons, n | 7,268 (100.0) | 7,268 (100.0) |
| | | Male, n (%) | 0 (0.0) | 0 (0.0) |
| | | Age, median (Q1;Q3) | 50.4 (46.6;54.5) | 50.7 (47.1;54.2) |
| | | CCI, n (%) | | |
| | | 0 | 6,007 (82.6) | 6,261 (86.1) |
| | | 1-2 | 1,144 (15.7) | 897 (12.3) |
| | | +3 | 117 (1.6) | 110 (1.5) |
| | | Education level, n (%) | | |
| | | No/Unknown education | 74 (1.0) | 74 (1.0) |
| | | Primary | 1,490 (20.5) | 1,490 (20.5) |
| | | Secondary | 3,257 (44.8) | 3,257 (44.8) |
| | | Tertiary | 2,447 (33.7) | 2,447 (33.7) |
| | | Relation to patient, n (%) | | |
| | | Mother | 7,268 (100.0) | 7,268 (100.0) |
| Young adults and adults (25-64 years) | Pooled | Number of persons, n | 34,931 (100.0) | 34,931 (100.0) |
| | | Male, n (%) | 15,353 (44.0) | 15,353 (44.0) |
| | | Age, median (Q1;Q3) | 54.1 (34.6;63.8) | 54.4 (34.5;63.7) |
| | | CCI, n (%) | | |
| | | 0 | 27,762 (79.5) | 28,282 (81.0) |
| | | 1-2 | 5,791 (16.6) | 5,506 (15.8) |
| | | +3 | 1,378 (3.9) | 1,143 (3.3) |
| | | Education level, n (%) | | |
| | | No/Unknown education | 2,028 (5.8) | 2,028 (5.8) |
| | | Primary | 10,475 (30.0) | 10,475 (30.0) |
| | | Secondary | 15,106 (43.2) | 15,106 (43.2) |
| | | Tertiary | 7,322 (21.0) | 7,322 (21.0) |
| | | Relation to patient, n (%) | | |
| | | Mother | 10,761 (30.8) | 10,761 (30.8) |
| | | Father | 3,355 (9.6) | 3,355 (9.6) |
| | | Partner | 10,436 (29.9) | 10,436 (29.9) |
| | | Child | 5,262 (15.1) | 5,262 (15.1) |

#### Supplementary Table 254
#### Schizophrenia spectrum disorders - Prevalent cohort 2021
#### Patient characteristics - relatives - pooled

| *Age strata of index patients* | *Closest relative type* | *Characteristic of family relatives by agestrata of index patients* | *Closest family relatives of brain disorder patients* | *Comparison relative* |
|---|---|---|---|---|
| | | Sibling | 5,117 (14.6) | 5,117 (14.6) |
| Older people (65+ years) | Pooled | Number of persons, n | 5,506 (100.0) | 5,506 (100.0) |
| | | Male, n (%) | 3,028 (55.0) | 3,028 (55.0) |
| | | Age, median (Q1;Q3) | 57.0 (47.4;67.6) | 57.0 (47.0;67.7) |
| | | CCI, n (%) | | |
| | | 0 | 4,226 (76.8) | 4,261 (77.4) |
| | | 1-2 | 994 (18.1) | 1,008 (18.3) |
| | | +3 | 286 (5.2) | 237 (4.3) |
| | | Education level, n (%) | | |
| | | No/Unknown education | 154 (2.8) | 154 (2.8) |
| | | Primary | 1,193 (21.7) | 1,193 (21.7) |
| | | Secondary | 2,577 (46.8) | 2,577 (46.8) |
| | | Tertiary | 1,582 (28.7) | 1,582 (28.7) |
| | | Relation to patient, n (%) | | |
| | | Partner | 1,689 (30.7) | 1,689 (30.7) |
| | | Child | 3,286 (59.7) | 3,286 (59.7) |
| | | Sibling | 531 (9.6) | 531 (9.6) |

**Supplementary Table 255**
**Schizophrenia spectrum disorders - Prevalent cohort 2021**
**Patient characteristics - relatives - not pooled**

| *Age strata of index patients* | *Closest relative type* | *Characteristic of family relatives by agestrata of index patients* | *Closest family relatives of brain disorder patients* | *Comparison relative* |
|---|---|---|---|---|
| Children and young people (0-24 years) | Fathers | Number of persons, n | 6,678 (100.0) | 6,678 (100.0) |
| | | Male, n (%) | 6,678 (100.0) | 6,678 (100.0) |
| | | Age, median (Q1;Q3) | 53.1 (48.8;57.0) | 53.1 (49.3;56.7) |
| | | CCI, n (%) | | |
| | | 0 | 5,506 (82.4) | 5,697 (85.3) |
| | | 1-2 | 989 (14.8) | 859 (12.9) |
| | | +3 | 183 (2.7) | 122 (1.8) |
| | | Education level, n (%) | | |
| | | No/Unknown education | 107 (1.6) | 107 (1.6) |
| | | Primary | 1,592 (23.8) | 1,592 (23.8) |
| | | Secondary | 3,338 (50.0) | 3,338 (50.0) |
| | | Tertiary | 1,641 (24.6) | 1,641 (24.6) |
| | | Relation to patient, n (%) | | |
| | | Father | 6,678 (100.0) | 6,678 (100.0) |
| Children and young people (0-24 years) | Mothers | Number of persons, n | 7,268 (100.0) | 7,268 (100.0) |
| | | Male, n (%) | 0 (0.0) | 0 (0.0) |
| | | Age, median (Q1;Q3) | 50.4 (46.6;54.5) | 50.7 (47.1;54.2) |
| | | CCI, n (%) | | |
| | | 0 | 6,007 (82.6) | 6,261 (86.1) |
| | | 1-2 | 1,144 (15.7) | 897 (12.3) |
| | | +3 | 117 (1.6) | 110 (1.5) |
| | | Education level, n (%) | | |
| | | No/Unknown education | 74 (1.0) | 74 (1.0) |
| | | Primary | 1,490 (20.5) | 1,490 (20.5) |
| | | Secondary | 3,257 (44.8) | 3,257 (44.8) |
| | | Tertiary | 2,447 (33.7) | 2,447 (33.7) |
| | | Relation to patient, n (%) | | |
| | | Mother | 7,268 (100.0) | 7,268 (100.0) |
| Young adults and adults (25-64 years) | Children | Number of persons, n | 5,262 (100.0) | 5,262 (100.0) |
| | | Male, n (%) | 2,825 (53.7) | 2,825 (53.7) |
| | | Age, median (Q1;Q3) | 21.1 (12.9;29.6) | 20.5 (12.8;29.1) |
| | | CCI, n (%) | | |
| | | 0 | 4,947 (94.0) | 4,934 (93.8) |
| | | 1-2 | 303 (5.8) | 318 (6.0) |
| | | +3 | 12 (0.2) | 10 (0.2) |
| | | Education level, n (%) | | |
| | | No/Unknown education | 1,343 (25.5) | 1,343 (25.5) |
| | | Primary | 2,150 (40.9) | 2,150 (40.9) |
| | | Secondary | 1,225 (23.3) | 1,225 (23.3) |
| | | Tertiary | 544 (10.3) | 544 (10.3) |
| | | Relation to patient, n (%) | | |
| | | Child | 5,262 (100.0) | 5,262 (100.0) |
| Young adults and adults (25-64 years) | Parents | Number of persons, n | 14,116 (100.0) | 14,116 (100.0) |
| | | Male, n (%) | 3,355 (23.8) | 3,355 (23.8) |
| | | Age, median (Q1;Q3) | 64.9 (58.2;73.3) | 64.8 (58.4;73.2) |

#### Supplementary Table 255
#### Schizophrenia spectrum disorders - Prevalent cohort 2021
#### Patient characteristics - relatives - not pooled

| *Age strata of index patients* | *Closest relative type* | *Characteristic of family relatives by agestrata of index patients* | *Closest family relatives of brain disorder patients* | *Comparison relative* |
|---|---|---|---|---|
| | | CCI, n (%) | | |
| | | 0 | 9,517 (67.4) | 9,769 (69.2) |
| | | 1-2 | 3,557 (25.2) | 3,463 (24.5) |
| | | +3 | 1,042 (7.4) | 884 (6.3) |
| | | Education level, n (%) | | |
| | | No/Unknown education | 306 (2.2) | 306 (2.2) |
| | | Primary | 3,982 (28.2) | 3,982 (28.2) |
| | | Secondary | 6,637 (47.0) | 6,637 (47.0) |
| | | Tertiary | 3,191 (22.6) | 3,191 (22.6) |
| | | Relation to patient, n (%) | | |
| | | Mother | 10,761 (76.2) | 10,761 (76.2) |
| | | Father | 3,355 (23.8) | 3,355 (23.8) |
| Young adults and adults (25-64 years) | Partners | Number of persons, n | 10,436 (100.0) | 10,436 (100.0) |
| | | Male, n (%) | 6,035 (57.8) | 6,035 (57.8) |
| | | Age, median (Q1;Q3) | 41.8 (32.8;53.0) | 41.8 (32.9;52.9) |
| | | CCI, n (%) | | |
| | | 0 | 9,055 (86.8) | 9,318 (89.3) |
| | | 1-2 | 1,192 (11.4) | 994 (9.5) |
| | | +3 | 189 (1.8) | 124 (1.2) |
| | | Education level, n (%) | | |
| | | No/Unknown education | 366 (3.5) | 366 (3.5) |
| | | Primary | 3,011 (28.9) | 3,011 (28.9) |
| | | Secondary | 4,580 (43.9) | 4,580 (43.9) |
| | | Tertiary | 2,479 (23.8) | 2,479 (23.8) |
| | | Relation to patient, n (%) | | |
| | | Partner | 10,436 (100.0) | 10,436 (100.0) |
| Young adults and adults (25-64 years) | Siblings | Number of persons, n | 5,117 (100.0) | 5,117 (100.0) |
| | | Male, n (%) | 3,138 (61.3) | 3,138 (61.3) |
| | | Age, median (Q1;Q3) | 53.2 (40.0;59.7) | 53.4 (40.1;59.7) |
| | | CCI, n (%) | | |
| | | 0 | 4,243 (82.9) | 4,261 (83.3) |
| | | 1-2 | 739 (14.4) | 731 (14.3) |
| | | +3 | 135 (2.6) | 125 (2.4) |
| | | Education level, n (%) | | |
| | | No/Unknown education | 13 (0.3) | 13 (0.3) |
| | | Primary | 1,332 (26.0) | 1,332 (26.0) |
| | | Secondary | 2,664 (52.1) | 2,664 (52.1) |
| | | Tertiary | 1,108 (21.7) | 1,108 (21.7) |
| | | Relation to patient, n (%) | | |
| | | Sibling | 5,117 (100.0) | 5,117 (100.0) |
| Older people (65+ years) | Children | Number of persons, n | 3,286 (100.0) | 3,286 (100.0) |
| | | Male, n (%) | 1,688 (51.4) | 1,688 (51.4) |
| | | Age, median (Q1;Q3) | 49.0 (43.2;55.3) | 48.9 (43.2;55.2) |
| | | CCI, n (%) | | |
| | | 0 | 2,853 (86.8) | 2,854 (86.9) |

**Supplementary Table 255**
**Schizophrenia spectrum disorders - Prevalent cohort 2021**
**Patient characteristics - relatives - not pooled**

| *Age strata of index patients* | *Closest relative type* | *Characteristic of family relatives by agestrata of index patients* | *Closest family relatives of brain disorder patients* | *Comparison relative* |
|---|---|---|---|---|
| | | 1-2 | 364 (11.1) | 379 (11.5) |
| | | +3 | 69 (2.1) | 53 (1.6) |
| | | Education level, n (%) | | |
| | | No/Unknown education | 5 (0.2) | 5 (0.2) |
| | | Primary | 629 (19.1) | 629 (19.1) |
| | | Secondary | 1,478 (45.0) | 1,478 (45.0) |
| | | Tertiary | 1,174 (35.7) | 1,174 (35.7) |
| | | Relation to patient, n (%) | | |
| | | Child | 3,286 (100.0) | 3,286 (100.0) |
| Older people (65+ years) | Partners | Number of persons, n | 1,689 (100.0) | 1,689 (100.0) |
| | | Male, n (%) | 1,048 (62.0) | 1,048 (62.0) |
| | | Age, median (Q1;Q3) | 72.5 (67.6;77.1) | 72.4 (68.0;76.8) |
| | | CCI, n (%) | | |
| | | 0 | 981 (58.1) | 1,004 (59.4) |
| | | 1-2 | 519 (30.7) | 518 (30.7) |
| | | +3 | 189 (11.2) | 167 (9.9) |
| | | Education level, n (%) | | |
| | | No/Unknown education | 143 (8.5) | 143 (8.5) |
| | | Primary | 480 (28.4) | 480 (28.4) |
| | | Secondary | 791 (46.8) | 791 (46.8) |
| | | Tertiary | 275 (16.3) | 275 (16.3) |
| | | Relation to patient, n (%) | | |
| | | Partner | 1,689 (100.0) | 1,689 (100.0) |
| Older people (65+ years) | Siblings | Number of persons, n | 531 (100.0) | 531 (100.0) |
| | | Male, n (%) | 292 (55.0) | 292 (55.0) |
| | | Age, median (Q1;Q3) | 62.4 (59.4;65.1) | 62.5 (60.0;64.9) |
| | | CCI, n (%) | | |
| | | 0 | 392 (73.8) | 403 (75.9) |
| | | 1-2 | 111 (20.9) | 111 (20.9) |
| | | +3 | 28 (5.3) | 17 (3.2) |
| | | Education level, n (%) | | |
| | | No/Unknown education | 6 (1.1) | 6 (1.1) |
| | | Primary | 84 (15.8) | 84 (15.8) |
| | | Secondary | 308 (58.0) | 308 (58.0) |
| | | Tertiary | 133 (25.0) | 133 (25.0) |
| | | Relation to patient, n (%) | | |
| | | Sibling | 531 (100.0) | 531 (100.0) |

### Supplementary Table 256
### Schizophrenia spectrum disorders - Prevalent cohort 2021
### Cost analysis - pooled

| Age strata of index patients | Closest relative type | Number of index patients included in cost component calculations | Cost component | Actual costs: Total (million EUR) | Actual costs: Per person (EUR) | Attributable costs: Total (million EUR) | Attributable costs: Per person (EUR) |
|---|---|---|---|---|---|---|---|
| Children and young people (0-24 years) | Fathers | 6,678 | Primary sector | 1.8 | 269 | 0.2 | 27 |
| | | 6,678 | Hospital care | 14.2 | 2,139 | 2.8 | 415 |
| | | 6,678 | Filled prescriptions | 2.0 | 297 | 0.6 | 84 |
| | | 6,272 | Lost productivity (income loss) | . | . | 70.1 | 11,184 |
| Children and young people (0-24 years) | Mothers | 7,268 | Primary sector | 3.0 | 409 | 0.5 | 68 |
| | | 7,268 | Hospital care | 15.1 | 2,084 | 3.7 | 508 |
| | | 7,268 | Filled prescriptions | 2.1 | 290 | 0.6 | 84 |
| | | 7,179 | Lost productivity (income loss) | . | . | 60.8 | 8,471 |
| Young adults and adults (25-64 years) | Pooled | 34,931 | Primary sector | 12.3 | 353 | 1.0 | 28 |
| | | 34,931 | Hospital care | 86.9 | 2,502 | 12.6 | 362 |
| | | 34,931 | Filled prescriptions | 11.5 | 331 | 1.6 | 46 |
| | | 22,960 | Lost productivity (income loss) | . | . | 251.7 | 10,963 |
| Older people (65+ years) | Pooled | 5,506 | Primary sector | 2.0 | 370 | 0.0 | 7 |
| | | 5,506 | Hospital care | 14.8 | 2,708 | 1.9 | 339 |
| | | 5,506 | Filled prescriptions | 2.0 | 361 | 0.1 | 13 |
| | | 3,486 | Lost productivity (income loss) | . | . | 21.2 | 6,093 |

#### Supplementary Table 257
#### Schizophrenia spectrum disorders - Prevalent cohort 2021
#### Cost analysis - not pooled

| *Age strata of index patients* | *Closest relative type* | *Number of index patients included in cost component calculations* | *Cost component* | *Actual costs* | | *Attributable costs* | |
|---|---|---|---|---|---|---|---|
| | | | | *Total (million EUR)* | *Per person (EUR)* | *Total (million EUR)* | *Per person (EUR)* |
| Children and young people (0-24 years) | Fathers | 6,678 | Primary sector | 1.8 | 269 | 0.2 | 27 |
| | | 6,678 | Hospital care | 14.2 | 2,139 | 2.8 | 415 |
| | | 6,678 | Filled prescriptions | 2.0 | 297 | 0.6 | 84 |
| | | 6,272 | Lost productivity (income loss) | . | . | 70.1 | 11,184 |
| Children and young people (0-24 years) | Mothers | 7,268 | Primary sector | 3.0 | 409 | 0.5 | 68 |
| | | 7,268 | Hospital care | 15.1 | 2,084 | 3.7 | 508 |
| | | 7,268 | Filled prescriptions | 2.1 | 290 | 0.6 | 84 |
| | | 7,179 | Lost productivity (income loss) | . | . | 60.8 | 8,471 |
| Young adults and adults (25-64 years) | Children | 5,262 | Primary sector | 1.1 | 217 | 0.1 | 21 |
| | | 5,262 | Hospital care | 7.3 | 1,389 | 2.1 | 403 |
| | | 5,262 | Filled prescriptions | 0.5 | 103 | 0.0 | 1 |
| | | 2,897 | Lost productivity (income loss) | . | . | 15.7 | 5,417 |
| Young adults and adults (25-64 years) | Parents | 14,116 | Primary sector | 6.5 | 463 | 0.5 | 36 |
| | | 14,116 | Hospital care | 48.2 | 3,452 | 5.3 | 377 |
| | | 14,116 | Filled prescriptions | 7.1 | 506 | 0.9 | 66 |
| | | 5,977 | Lost productivity (income loss) | . | . | 44.0 | 7,365 |
| Young adults and adults (25-64 years) | Partners | 10,436 | Primary sector | 3.1 | 299 | 0.3 | 28 |
| | | 10,436 | Hospital care | 20.4 | 1,956 | 4.3 | 408 |
| | | 10,436 | Filled prescriptions | 2.6 | 254 | 0.7 | 68 |
| | | 9,728 | Lost productivity (income loss) | . | . | 158.8 | 16,328 |
| Young adults and adults (25-64 years) | Siblings | 5,117 | Primary sector | 1.6 | 306 | 0.1 | 15 |
| | | 5,117 | Hospital care | 11.0 | 2,158 | 0.9 | 183 |
| | | 5,117 | Filled prescriptions | 1.2 | 244 | -0.0 | -8 |
| | | 4,358 | Lost productivity (income loss) | . | . | 33.2 | 7,609 |
| Older people (65+ years) | Children | 3,286 | Primary sector | 1.0 | 303 | 0.0 | 8 |
| | | 3,286 | Hospital care | 6.2 | 1,902 | 1.3 | 382 |
| | | 3,286 | Filled prescriptions | 0.8 | 231 | -0.1 | -28 |
| | | 3,088 | Lost productivity (income loss) | . | . | 20.8 | 6,743 |
| Older people (65+ years) | Partners | 1,689 | Primary sector | 0.8 | 503 | 0.0 | 7 |
| | | 1,689 | Hospital care | 7.6 | 4,570 | 1.0 | 625 |
| | | 1,689 | Filled prescriptions | 1.0 | 599 | 0.1 | 76 |
| | | 120 | Lost productivity (income loss) | . | . | 2.2 | 18,230 |
| Older people (65+ years) | Siblings | 531 | Primary sector | 0.2 | 370 | -0.0 | -4 |
| | | 531 | Hospital care | 1.0 | 1,849 | -0.4 | -825 |
| | | 531 | Filled prescriptions | 0.2 | 416 | 0.0 | 66 |
| | | 278 | Lost productivity (income loss) | . | . | -1.8 | -6,369 |

### Supplementary Table 258
### Sleep disorders - Incident cohort 2016-2021
### Identification of populations of relatives

| *Agegroup of index patients in matched population* | | *Closest relative type identified* | | *Available control with same type of closest relative* |
|---|---|---|---|---|
| Children and young people (0-24 years) | n = 54,475 | Fathers | n = 37,378 (69%) | 96% |
| | | Mothers | n = 37,852 (69%) | 96% |
| | | No available closest relative | n = 7,097 (13%) | n/a |
| Young adults and adults (25-64 years) | n = 160,148 | Children | n = 31,363 (20%) | 95% |
| | | Parents | n = 23,300 (15%) | 88% |
| | | Partners | n = 78,000 (49%) | 95% |
| | | Siblings | n = 20,229 (13%) | 60% |
| | | Pooled | n = 152,892 (95%) | 89% |
| | | No available closest relative | n = 7,256 (5%) | n/a |
| Older people (65+ years) | n = 87,813 | Children | n = 39,997 (46%) | 95% |
| | | Partners | n = 33,685 (38%) | 95% |
| | | Siblings | n = 1,478 (2%) | 36% |
| | | Pooled | n = 75,160 (86%) | 94% |
| | | No available closest relative | n = 12,653 (14%) | n/a |

#### Supplementary Table 259
#### Sleep disorders - Incident cohort 2016-2021
#### Patient characteristics - cases

| *Age strata of index patients* | *Characteristic* | |
|---|---|---|
| Children and young people (0-24 years) | Number of persons, n | 45,794 (100.0) |
| | Male, n (%) | 21,516 (47.0) |
| | Age, median (Q1;Q3) | 17.7 (13.2;21.4) |
| | CCI, n (%) | |
| | 0 | 41,702 (91.1) |
| | 1-2 | 3,914 (8.5) |
| | +3 | 178 (0.4) |
| | Education level, n (%) | |
| | No/Unknown education | 11,484 (25.1) |
| | Primary | 26,092 (57.0) |
| | Secondary | 7,529 (16.4) |
| | Tertiary | 689 (1.5) |
| Young adults and adults (25-64 years) | Number of persons, n | 137,414 (100.0) |
| | Male, n (%) | 64,032 (46.6) |
| | Age, median (Q1;Q3) | 47.9 (37.2;55.8) |
| | CCI, n (%) | |
| | 0 | 108,669 (79.1) |
| | 1-2 | 23,467 (17.1) |
| | +3 | 5,278 (3.8) |
| | Education level, n (%) | |
| | No/Unknown education | 4,369 (3.2) |
| | Primary | 29,321 (21.3) |
| | Secondary | 62,318 (45.4) |
| | Tertiary | 41,406 (30.1) |
| Older people (65+ years) | Number of persons, n | 70,773 (100.0) |
| | Male, n (%) | 34,272 (48.4) |
| | Age, median (Q1;Q3) | 75.1 (70.4;80.9) |
| | CCI, n (%) | |
| | 0 | 28,612 (40.4) |
| | 1-2 | 27,000 (38.2) |
| | +3 | 15,161 (21.4) |
| | Education level, n (%) | |
| | No/Unknown education | 4,625 (6.5) |
| | Primary | 21,961 (31.0) |
| | Secondary | 31,635 (44.7) |
| | Tertiary | 12,552 (17.7) |

**Supplementary Table 260**
**Sleep disorders - Incident cohort 2016-2021**
**Patient characteristics - relatives - pooled**

| *Age strata of index patients* | *Closest relative type* | *Characteristic of family relatives by agestrata of index patients* | *Closest family relatives of brain disorder patients* | *Comparison relative* |
|---|---|---|---|---|
| Children and young people (0-24 years) | Fathers | Number of persons, n | 36,023 (100.0) | 36,023 (100.0) |
| | | Male, n (%) | 36,023 (100.0) | 36,023 (100.0) |
| | | Age, median (Q1;Q3) | 49.2 (43.8;53.9) | 49.3 (44.2;53.7) |
| | | CCI, n (%) | | |
| | | 0 | 31,780 (88.2) | 32,263 (89.6) |
| | | 1-2 | 3,788 (10.5) | 3,348 (9.3) |
| | | +3 | 455 (1.3) | 412 (1.1) |
| | | Education level, n (%) | | |
| | | No/Unknown education | 443 (1.2) | 443 (1.2) |
| | | Primary | 7,821 (21.7) | 7,821 (21.7) |
| | | Secondary | 18,464 (51.3) | 18,464 (51.3) |
| | | Tertiary | 9,295 (25.8) | 9,295 (25.8) |
| | | Relation to patient, n (%) | | |
| | | Father | 36,023 (100.0) | 36,023 (100.0) |
| Children and young people (0-24 years) | Mothers | Number of persons, n | 36,550 (100.0) | 36,550 (100.0) |
| | | Male, n (%) | 0 (0.0) | 0 (0.0) |
| | | Age, median (Q1;Q3) | 46.9 (41.7;51.5) | 47.1 (42.2;51.3) |
| | | CCI, n (%) | | |
| | | 0 | 31,755 (86.9) | 32,779 (89.7) |
| | | 1-2 | 4,429 (12.1) | 3,458 (9.5) |
| | | +3 | 366 (1.0) | 313 (0.9) |
| | | Education level, n (%) | | |
| | | No/Unknown education | 365 (1.0) | 365 (1.0) |
| | | Primary | 6,644 (18.2) | 6,644 (18.2) |
| | | Secondary | 16,034 (43.9) | 16,034 (43.9) |
| | | Tertiary | 13,507 (37.0) | 13,507 (37.0) |
| | | Relation to patient, n (%) | | |
| | | Mother | 36,550 (100.0) | 36,550 (100.0) |
| Young adults and adults (25-64 years) | Pooled | Number of persons, n | 137,414 (100.0) | 137,414 (100.0) |
| | | Male, n (%) | 68,719 (50.0) | 68,719 (50.0) |
| | | Age, median (Q1;Q3) | 46.6 (30.9;57.6) | 46.7 (30.9;57.6) |
| | | CCI, n (%) | | |
| | | 0 | 118,246 (86.1) | 119,941 (87.3) |
| | | 1-2 | 16,388 (11.9) | 15,141 (11.0) |
| | | +3 | 2,780 (2.0) | 2,332 (1.7) |
| | | Education level, n (%) | | |
| | | No/Unknown education | 10,089 (7.3) | 10,089 (7.3) |
| | | Primary | 32,868 (23.9) | 32,868 (23.9) |
| | | Secondary | 60,057 (43.7) | 60,057 (43.7) |
| | | Tertiary | 34,400 (25.0) | 34,400 (25.0) |
| | | Relation to patient, n (%) | | |
| | | Mother | 14,764 (10.7) | 14,764 (10.7) |
| | | Father | 5,794 (4.2) | 5,794 (4.2) |
| | | Partner | 74,728 (54.4) | 74,728 (54.4) |
| | | Child | 29,928 (21.8) | 29,928 (21.8) |

#### Supplementary Table 260
#### Sleep disorders - Incident cohort 2016-2021
#### Patient characteristics - relatives - pooled

| *Age strata of index patients* | *Closest relative type* | *Characteristic of family relatives by agestrata of index patients* | *Closest family relatives of brain disorder patients* | *Comparison relative* |
|---|---|---|---|---|
| | | Sibling | 12,200 (8.9) | 12,200 (8.9) |
| Older people (65+ years) | Pooled | Number of persons, n | 70,773 (100.0) | 70,773 (100.0) |
| | | Male, n (%) | 34,577 (48.9) | 34,577 (48.9) |
| | | Age, median (Q1;Q3) | 60.8 (50.3;72.1) | 60.7 (50.1;72.1) |
| | | CCI, n (%) | | |
| | | 0 | 53,215 (75.2) | 54,657 (77.2) |
| | | 1-2 | 13,898 (19.6) | 13,011 (18.4) |
| | | +3 | 3,660 (5.2) | 3,105 (4.4) |
| | | Education level, n (%) | | |
| | | No/Unknown education | 2,027 (2.9) | 2,027 (2.9) |
| | | Primary | 14,143 (20.0) | 14,143 (20.0) |
| | | Secondary | 34,675 (49.0) | 34,675 (49.0) |
| | | Tertiary | 19,928 (28.2) | 19,928 (28.2) |
| | | Relation to patient, n (%) | | |
| | | Partner | 32,114 (45.4) | 32,114 (45.4) |
| | | Child | 38,126 (53.9) | 38,126 (53.9) |
| | | Sibling | 533 (0.8) | 533 (0.8) |

**Supplementary Table 261**
**Sleep disorders - Incident cohort 2016-2021**
**Patient characteristics - relatives - not pooled**

| *Age strata of index patients* | *Closest relative type* | *Characteristic of family relatives by agestrata of index patients* | *Closest family relatives of brain disorder patients* | *Comparison relative* |
|---|---|---|---|---|
| Children and young people (0-24 years) | Fathers | Number of persons, n | 36,023 (100.0) | 36,023 (100.0) |
| | | Male, n (%) | 36,023 (100.0) | 36,023 (100.0) |
| | | Age, median (Q1;Q3) | 49.2 (43.8;53.9) | 49.3 (44.2;53.7) |
| | | CCI, n (%) | | |
| | | 0 | 31,780 (88.2) | 32,263 (89.6) |
| | | 1-2 | 3,788 (10.5) | 3,348 (9.3) |
| | | +3 | 455 (1.3) | 412 (1.1) |
| | | Education level, n (%) | | |
| | | No/Unknown education | 443 (1.2) | 443 (1.2) |
| | | Primary | 7,821 (21.7) | 7,821 (21.7) |
| | | Secondary | 18,464 (51.3) | 18,464 (51.3) |
| | | Tertiary | 9,295 (25.8) | 9,295 (25.8) |
| | | Relation to patient, n (%) | | |
| | | Father | 36,023 (100.0) | 36,023 (100.0) |
| Children and young people (0-24 years) | Mothers | Number of persons, n | 36,550 (100.0) | 36,550 (100.0) |
| | | Male, n (%) | 0 (0.0) | 0 (0.0) |
| | | Age, median (Q1;Q3) | 46.9 (41.7;51.5) | 47.1 (42.2;51.3) |
| | | CCI, n (%) | | |
| | | 0 | 31,755 (86.9) | 32,779 (89.7) |
| | | 1-2 | 4,429 (12.1) | 3,458 (9.5) |
| | | +3 | 366 (1.0) | 313 (0.9) |
| | | Education level, n (%) | | |
| | | No/Unknown education | 365 (1.0) | 365 (1.0) |
| | | Primary | 6,644 (18.2) | 6,644 (18.2) |
| | | Secondary | 16,034 (43.9) | 16,034 (43.9) |
| | | Tertiary | 13,507 (37.0) | 13,507 (37.0) |
| | | Relation to patient, n (%) | | |
| | | Mother | 36,550 (100.0) | 36,550 (100.0) |
| Young adults and adults (25-64 years) | Children | Number of persons, n | 29,928 (100.0) | 29,928 (100.0) |
| | | Male, n (%) | 15,631 (52.2) | 15,631 (52.2) |
| | | Age, median (Q1;Q3) | 20.4 (13.3;28.9) | 20.3 (13.2;28.6) |
| | | CCI, n (%) | | |
| | | 0 | 28,082 (93.8) | 28,303 (94.6) |
| | | 1-2 | 1,773 (5.9) | 1,562 (5.2) |
| | | +3 | 73 (0.2) | 63 (0.2) |
| | | Education level, n (%) | | |
| | | No/Unknown education | 7,376 (24.6) | 7,376 (24.6) |
| | | Primary | 11,233 (37.5) | 11,233 (37.5) |
| | | Secondary | 7,318 (24.5) | 7,318 (24.5) |
| | | Tertiary | 4,001 (13.4) | 4,001 (13.4) |
| | | Relation to patient, n (%) | | |
| | | Child | 29,928 (100.0) | 29,928 (100.0) |
| Young adults and adults (25-64 years) | Parents | Number of persons, n | 20,558 (100.0) | 20,558 (100.0) |
| | | Male, n (%) | 5,794 (28.2) | 5,794 (28.2) |
| | | Age, median (Q1;Q3) | 65.0 (57.9;73.5) | 64.9 (57.9;73.5) |

**Supplementary Table 261**
**Sleep disorders - Incident cohort 2016-2021**
**Patient characteristics - relatives - not pooled**

| *Age strata of index patients* | *Closest relative type* | *Characteristic of family relatives by agestrata of index patients* | *Closest family relatives of brain disorder patients* | *Comparison relative* |
|---|---|---|---|---|
| | | CCI, n (%) | | |
| | | 0 | 14,413 (70.1) | 14,816 (72.1) |
| | | 1-2 | 4,841 (23.5) | 4,623 (22.5) |
| | | +3 | 1,304 (6.3) | 1,119 (5.4) |
| | | Education level, n (%) | | |
| | | No/Unknown education | 325 (1.6) | 325 (1.6) |
| | | Primary | 6,046 (29.4) | 6,046 (29.4) |
| | | Secondary | 10,042 (48.8) | 10,042 (48.8) |
| | | Tertiary | 4,145 (20.2) | 4,145 (20.2) |
| | | Relation to patient, n (%) | | |
| | | Mother | 14,764 (71.8) | 14,764 (71.8) |
| | | Father | 5,794 (28.2) | 5,794 (28.2) |
| Young adults and adults (25-64 years) | Partners | Number of persons, n | 74,728 (100.0) | 74,728 (100.0) |
| | | Male, n (%) | 40,596 (54.3) | 40,596 (54.3) |
| | | Age, median (Q1;Q3) | 48.7 (39.1;56.5) | 48.8 (39.1;56.4) |
| | | CCI, n (%) | | |
| | | 0 | 65,070 (87.1) | 66,089 (88.4) |
| | | 1-2 | 8,437 (11.3) | 7,655 (10.2) |
| | | +3 | 1,221 (1.6) | 984 (1.3) |
| | | Education level, n (%) | | |
| | | No/Unknown education | 2,356 (3.2) | 2,356 (3.2) |
| | | Primary | 12,671 (17.0) | 12,671 (17.0) |
| | | Secondary | 36,428 (48.7) | 36,428 (48.7) |
| | | Tertiary | 23,273 (31.1) | 23,273 (31.1) |
| | | Relation to patient, n (%) | | |
| | | Partner | 74,728 (100.0) | 74,728 (100.0) |
| Young adults and adults (25-64 years) | Siblings | Number of persons, n | 12,200 (100.0) | 12,200 (100.0) |
| | | Male, n (%) | 6,698 (54.9) | 6,698 (54.9) |
| | | Age, median (Q1;Q3) | 47.2 (31.1;56.7) | 47.3 (30.9;56.8) |
| | | CCI, n (%) | | |
| | | 0 | 10,681 (87.5) | 10,733 (88.0) |
| | | 1-2 | 1,337 (11.0) | 1,301 (10.7) |
| | | +3 | 182 (1.5) | 166 (1.4) |
| | | Education level, n (%) | | |
| | | No/Unknown education | 32 (0.3) | 32 (0.3) |
| | | Primary | 2,918 (23.9) | 2,918 (23.9) |
| | | Secondary | 6,269 (51.4) | 6,269 (51.4) |
| | | Tertiary | 2,981 (24.4) | 2,981 (24.4) |
| | | Relation to patient, n (%) | | |
| | | Sibling | 12,200 (100.0) | 12,200 (100.0) |
| Older people (65+ years) | Children | Number of persons, n | 38,126 (100.0) | 38,126 (100.0) |
| | | Male, n (%) | 19,837 (52.0) | 19,837 (52.0) |
| | | Age, median (Q1;Q3) | 51.1 (45.1;56.7) | 51.0 (45.1;56.5) |
| | | CCI, n (%) | | |
| | | 0 | 33,090 (86.8) | 33,550 (88.0) |

#### Supplementary Table 261
#### Sleep disorders - Incident cohort 2016-2021
#### Patient characteristics - relatives - not pooled

| *Age strata of index patients* | *Closest relative type* | *Characteristic of family relatives by agestrata of index patients* | *Closest family relatives of brain disorder patients* | *Comparison relative* |
|---|---|---|---|---|
| | | 1-2 | 4,439 (11.6) | 4,082 (10.7) |
| | | +3 | 597 (1.6) | 494 (1.3) |
| | | Education level, n (%) | | |
| | | No/Unknown education | 115 (0.3) | 115 (0.3) |
| | | Primary | 5,594 (14.7) | 5,594 (14.7) |
| | | Secondary | 19,118 (50.1) | 19,118 (50.1) |
| | | Tertiary | 13,299 (34.9) | 13,299 (34.9) |
| | | Relation to patient, n (%) | | |
| | | Child | 38,126 (100.0) | 38,126 (100.0) |
| Older people (65+ years) | Partners | Number of persons, n | 32,114 (100.0) | 32,114 (100.0) |
| | | Male, n (%) | 14,424 (44.9) | 14,424 (44.9) |
| | | Age, median (Q1;Q3) | 73.0 (68.4;78.0) | 72.9 (68.5;77.8) |
| | | CCI, n (%) | | |
| | | 0 | 19,730 (61.4) | 20,677 (64.4) |
| | | 1-2 | 9,341 (29.1) | 8,840 (27.5) |
| | | +3 | 3,043 (9.5) | 2,597 (8.1) |
| | | Education level, n (%) | | |
| | | No/Unknown education | 1,911 (6.0) | 1,911 (6.0) |
| | | Primary | 8,470 (26.4) | 8,470 (26.4) |
| | | Secondary | 15,223 (47.4) | 15,223 (47.4) |
| | | Tertiary | 6,510 (20.3) | 6,510 (20.3) |
| | | Relation to patient, n (%) | | |
| | | Partner | 32,114 (100.0) | 32,114 (100.0) |
| Older people (65+ years) | Siblings | Number of persons, n | 533 (100.0) | 533 (100.0) |
| | | Male, n (%) | 316 (59.3) | 316 (59.3) |
| | | Age, median (Q1;Q3) | 62.1 (59.1;64.5) | 62.3 (59.5;64.3) |
| | | CCI, n (%) | | |
| | | 0 | 395 (74.1) | 430 (80.7) |
| | | 1-2 | 118 (22.1) | 89 (16.7) |
| | | +3 | 20 (3.8) | 14 (2.6) |
| | | Education level, n (%) | | |
| | | No/Unknown education | * | * |
| | | Primary | * | * |
| | | Secondary | 334 (62.7) | 334 (62.7) |
| | | Tertiary | 119 (22.3) | 119 (22.3) |
| | | Relation to patient, n (%) | | |
| | | Sibling | 533 (100.0) | 533 (100.0) |

#### Supplementary Table 262
#### Sleep disorders - Incident cohort 2016-2021
#### Cost analysis - pooled

| Age strata of index patients | Closest relative type | Number of index patients included in cost component calculations | Cost component | Actual costs | | Attributable costs | |
|---|---|---|---|---|---|---|---|
| | | | | Total (million EUR) | Per person (EUR) | Total (million EUR) | Per person (EUR) |
| Children and young people (0-24 years) | Fathers | 36,023 | Primary sector | 7.9 | 220 | 0.7 | 19 |
| | | 36,023 | Hospital care | 53.6 | 1,492 | 10.5 | 291 |
| | | 36,023 | Filled prescriptions | 6.5 | 181 | 1.6 | 44 |
| | | 34,901 | Lost productivity (income loss) | . | . | 135.4 | 3,879 |
| Children and young people (0-24 years) | Mothers | 36,550 | Primary sector | 13.5 | 369 | 2.2 | 61 |
| | | 36,550 | Hospital care | 64.2 | 1,758 | 16.0 | 438 |
| | | 36,550 | Filled prescriptions | 8.0 | 221 | 2.7 | 73 |
| | | 36,301 | Lost productivity (income loss) | . | . | 175.6 | 4,836 |
| Young adults and adults (25-64 years) | Pooled | 137,414 | Primary sector | 40.3 | 295 | 4.4 | 32 |
| | | 137,414 | Hospital care | 264.0 | 1,931 | 54.3 | 397 |
| | | 137,414 | Filled prescriptions | 28.9 | 212 | 4.4 | 32 |
| | | 103,871 | Lost productivity (income loss) | . | . | 224.8 | 2,164 |
| Older people (65+ years) | Pooled | 70,773 | Primary sector | 26.7 | 382 | 2.1 | 31 |
| | | 70,773 | Hospital care | 200.2 | 2,867 | 33.4 | 479 |
| | | 70,773 | Filled prescriptions | 23.2 | 332 | 2.0 | 29 |
| | | 37,192 | Lost productivity (income loss) | . | . | -3.9 | -104 |

#### Supplementary Table 263
#### Sleep disorders - Incident cohort 2016-2021
#### Cost analysis - not pooled

| | | | | *Actual costs* | | *Attributable costs* | |
|---|---|---|---|---|---|---|---|
| *Age strata of index patients* | *Closest relative type* | *Number of index patients included in cost component calculations* | *Cost component* | *Total (million EUR)* | *Per person (EUR)* | *Total (million EUR)* | *Per person (EUR)* |
| Children and young people (0-24 years) | Fathers | 36,023 | Primary sector | 7.9 | 220 | 0.7 | 19 |
| | | 36,023 | Hospital care | 53.6 | 1,492 | 10.5 | 291 |
| | | 36,023 | Filled prescriptions | 6.5 | 181 | 1.6 | 44 |
| | | 34,901 | Lost productivity (income loss) | . | . | 135.4 | 3,879 |
| Children and young people (0-24 years) | Mothers | 36,550 | Primary sector | 13.5 | 369 | 2.2 | 61 |
| | | 36,550 | Hospital care | 64.2 | 1,758 | 16.0 | 438 |
| | | 36,550 | Filled prescriptions | 8.0 | 221 | 2.7 | 73 |
| | | 36,301 | Lost productivity (income loss) | . | . | 175.6 | 4,836 |
| Young adults and adults (25-64 years) | Children | 29,928 | Primary sector | 6.3 | 211 | 0.7 | 25 |
| | | 29,928 | Hospital care | 36.2 | 1,216 | 8.2 | 275 |
| | | 29,928 | Filled prescriptions | 3.0 | 101 | 0.3 | 10 |
| | | 16,830 | Lost productivity (income loss) | . | . | 16.2 | 961 |
| Young adults and adults (25-64 years) | Parents | 20,558 | Primary sector | 8.7 | 427 | 0.7 | 34 |
| | | 20,558 | Hospital care | 62.0 | 3,044 | 6.5 | 321 |
| | | 20,558 | Filled prescriptions | 9.0 | 441 | 1.4 | 67 |
| | | 8,474 | Lost productivity (income loss) | . | . | 37.9 | 4,471 |
| Young adults and adults (25-64 years) | Partners | 74,728 | Primary sector | 21.9 | 295 | 2.7 | 36 |
| | | 74,728 | Hospital care | 144.2 | 1,937 | 37.1 | 499 |
| | | 74,728 | Filled prescriptions | 14.5 | 195 | 2.5 | 33 |
| | | 67,527 | Lost productivity (income loss) | . | . | 155.5 | 2,303 |
| Young adults and adults (25-64 years) | Siblings | 12,200 | Primary sector | 3.4 | 277 | 0.2 | 19 |
| | | 12,200 | Hospital care | 21.6 | 1,775 | 2.5 | 205 |
| | | 12,200 | Filled prescriptions | 2.4 | 200 | 0.3 | 23 |
| | | 11,040 | Lost productivity (income loss) | . | . | 15.2 | 1,380 |
| Older people (65+ years) | Children | 38,126 | Primary sector | 10.8 | 285 | 0.9 | 24 |
| | | 38,126 | Hospital care | 62.5 | 1,642 | 6.4 | 169 |
| | | 38,126 | Filled prescriptions | 7.4 | 194 | 0.6 | 16 |
| | | 35,329 | Lost productivity (income loss) | . | . | -1.1 | -32 |
| Older people (65+ years) | Partners | 32,114 | Primary sector | 15.7 | 502 | 1.2 | 38 |
| | | 32,114 | Hospital care | 135.8 | 4,345 | 26.1 | 834 |
| | | 32,114 | Filled prescriptions | 15.6 | 500 | 1.4 | 44 |
| | | 1,566 | Lost productivity (income loss) | . | . | -1.3 | -819 |
| Older people (65+ years) | Siblings | 533 | Primary sector | 0.2 | 362 | 0.0 | 62 |
| | | 533 | Hospital care | 1.9 | 3,576 | 0.9 | 1,733 |
| | | 533 | Filled prescriptions | 0.2 | 323 | 0.0 | 45 |
| | | 297 | Lost productivity (income loss) | . | . | -1.5 | -4,943 |

**Supplementary Table 264**
**Sleep disorders - Prevalent cohort 2021**
**Identification of populations of relatives**

| *Agegroup of index patients in matched population* | | *Closest relative type identified* | | *Available control with same type of closest relative* |
|---|---|---|---|---|
| Children and young people (0-24 years) | n = 55,115 | Fathers | n = 36,998 (67%) | 96% |
| | | Mothers | n = 36,566 (66%) | 96% |
| | | No available closest relative | n = 7,644 (14%) | n/a |
| Young adults and adults (25-64 years) | n = 402,783 | Children | n = 94,352 (23%) | 95% |
| | | Parents | n = 58,212 (14%) | 85% |
| | | Partners | n = 177,542 (44%) | 95% |
| | | Siblings | n = 56,136 (14%) | 60% |
| | | Pooled | n = 386,242 (96%) | 88% |
| | | No available closest relative | n = 16,541 (4%) | n/a |
| Older people (65+ years) | n = 285,693 | Children | n = 133,360 (47%) | 95% |
| | | Partners | n = 100,189 (35%) | 95% |
| | | Siblings | n = 7,976 (3%) | 46% |
| | | Pooled | n = 241,525 (85%) | 94% |
| | | No available closest relative | n = 44,168 (15%) | n/a |

#### Supplementary Table 265
#### Sleep disorders - Prevalent cohort 2021
#### Patient characteristics - cases

| *Age strata of index patients* | *Characteristic* | |
|---|---|---|
| Children and young people (0-24 years) | Number of persons, n | 46,038 (100.0) |
| | Male, n (%) | 24,225 (52.6) |
| | Age, median (Q1;Q3) | 19.0 (14.5;22.1) |
| | CCI, n (%) | |
| | 0 | 41,470 (90.1) |
| | 1-2 | 4,315 (9.4) |
| | +3 | 253 (0.5) |
| | Education level, n (%) | |
| | No/Unknown education | 9,187 (20.0) |
| | Primary | 28,772 (62.5) |
| | Secondary | 7,512 (16.3) |
| | Tertiary | 567 (1.2) |
| Young adults and adults (25-64 years) | Number of persons, n | 343,196 (100.0) |
| | Male, n (%) | 152,829 (44.5) |
| | Age, median (Q1;Q3) | 51.4 (41.6;58.1) |
| | CCI, n (%) | |
| | 0 | 268,666 (78.3) |
| | 1-2 | 62,212 (18.1) |
| | +3 | 12,318 (3.6) |
| | Education level, n (%) | |
| | No/Unknown education | 8,215 (2.4) |
| | Primary | 79,726 (23.2) |
| | Secondary | 155,914 (45.4) |
| | Tertiary | 99,341 (28.9) |
| Older people (65+ years) | Number of persons, n | 227,088 (100.0) |
| | Male, n (%) | 90,282 (39.8) |
| | Age, median (Q1;Q3) | 74.9 (70.0;80.6) |
| | CCI, n (%) | |
| | 0 | 110,758 (48.8) |
| | 1-2 | 81,160 (35.7) |
| | +3 | 35,170 (15.5) |
| | Education level, n (%) | |
| | No/Unknown education | 13,944 (6.1) |
| | Primary | 63,964 (28.2) |
| | Secondary | 102,110 (45.0) |
| | Tertiary | 47,070 (20.7) |

**Supplementary Table 266**
**Sleep disorders - Prevalent cohort 2021**
**Patient characteristics - relatives - pooled**

| *Age strata of index patients* | *Closest relative type* | *Characteristic of family relatives by agestrata of index patients* | *Closest family relatives of brain disorder patients* | *Comparison relative* |
|---|---|---|---|---|
| Children and young people (0-24 years) | Fathers | Number of persons, n | 35,778 (100.0) | 35,778 (100.0) |
| | | Male, n (%) | 35,778 (100.0) | 35,778 (100.0) |
| | | Age, median (Q1;Q3) | 50.2 (45.2;54.8) | 50.3 (45.5;54.6) |
| | | CCI, n (%) | | |
| | | 0 | 31,339 (87.6) | 31,908 (89.2) |
| | | 1-2 | 3,977 (11.1) | 3,464 (9.7) |
| | | +3 | 462 (1.3) | 406 (1.1) |
| | | Education level, n (%) | | |
| | | No/Unknown education | 388 (1.1) | 388 (1.1) |
| | | Primary | 7,934 (22.2) | 7,934 (22.2) |
| | | Secondary | 18,405 (51.4) | 18,405 (51.4) |
| | | Tertiary | 9,051 (25.3) | 9,051 (25.3) |
| | | Relation to patient, n (%) | | |
| | | Father | 35,778 (100.0) | 35,778 (100.0) |
| Children and young people (0-24 years) | Mothers | Number of persons, n | 35,423 (100.0) | 35,423 (100.0) |
| | | Male, n (%) | 0 (0.0) | 0 (0.0) |
| | | Age, median (Q1;Q3) | 47.9 (43.1;52.5) | 48.2 (43.5;52.3) |
| | | CCI, n (%) | | |
| | | 0 | 30,470 (86.0) | 31,537 (89.0) |
| | | 1-2 | 4,564 (12.9) | 3,579 (10.1) |
| | | +3 | 389 (1.1) | 307 (0.9) |
| | | Education level, n (%) | | |
| | | No/Unknown education | 263 (0.7) | 263 (0.7) |
| | | Primary | 6,385 (18.0) | 6,385 (18.0) |
| | | Secondary | 15,615 (44.1) | 15,615 (44.1) |
| | | Tertiary | 13,160 (37.2) | 13,160 (37.2) |
| | | Relation to patient, n (%) | | |
| | | Mother | 35,423 (100.0) | 35,423 (100.0) |
| Young adults and adults (25-64 years) | Pooled | Number of persons, n | 343,196 (100.0) | 343,196 (100.0) |
| | | Male, n (%) | 175,780 (51.2) | 175,780 (51.2) |
| | | Age, median (Q1;Q3) | 48.4 (31.2;59.5) | 48.5 (31.2;59.4) |
| | | CCI, n (%) | | |
| | | 0 | 292,123 (85.1) | 295,917 (86.2) |
| | | 1-2 | 43,770 (12.8) | 40,580 (11.8) |
| | | +3 | 7,303 (2.1) | 6,699 (2.0) |
| | | Education level, n (%) | | |
| | | No/Unknown education | 20,941 (6.1) | 20,941 (6.1) |
| | | Primary | 86,084 (25.1) | 86,084 (25.1) |
| | | Secondary | 152,639 (44.5) | 152,639 (44.5) |
| | | Tertiary | 83,532 (24.3) | 83,532 (24.3) |
| | | Relation to patient, n (%) | | |
| | | Mother | 35,597 (10.4) | 35,597 (10.4) |
| | | Father | 14,461 (4.2) | 14,461 (4.2) |
| | | Partner | 169,632 (49.4) | 169,632 (49.4) |
| | | Child | 89,679 (26.1) | 89,679 (26.1) |

**Supplementary Table 266**
**Sleep disorders - Prevalent cohort 2021**
**Patient characteristics - relatives - pooled**

| *Age strata of index patients* | *Closest relative type* | *Characteristic of family relatives by agestrata of index patients* | *Closest family relatives of brain disorder patients* | *Comparison relative* |
|---|---|---|---|---|
| | | Sibling | 33,827 (9.9) | 33,827 (9.9) |
| Older people (65+ years) | Pooled | Number of persons, n | 227,088 (100.0) | 227,088 (100.0) |
| | | Male, n (%) | 118,599 (52.2) | 118,599 (52.2) |
| | | Age, median (Q1;Q3) | 60.2 (49.7;71.5) | 60.2 (49.7;71.6) |
| | | CCI, n (%) | | |
| | | 0 | 173,626 (76.5) | 175,939 (77.5) |
| | | 1-2 | 43,131 (19.0) | 41,397 (18.2) |
| | | +3 | 10,331 (4.5) | 9,752 (4.3) |
| | | Education level, n (%) | | |
| | | No/Unknown education | 6,242 (2.7) | 6,242 (2.7) |
| | | Primary | 39,448 (17.4) | 39,448 (17.4) |
| | | Secondary | 110,560 (48.7) | 110,560 (48.7) |
| | | Tertiary | 70,838 (31.2) | 70,838 (31.2) |
| | | Relation to patient, n (%) | | |
| | | Partner | 95,623 (42.1) | 95,623 (42.1) |
| | | Child | 127,786 (56.3) | 127,786 (56.3) |
| | | Sibling | 3,679 (1.6) | 3,679 (1.6) |

**Supplementary Table 267**
**Sleep disorders - Prevalent cohort 2021**
**Patient characteristics - relatives - not pooled**

| *Age strata of index patients* | *Closest relative type* | *Characteristic of family relatives by agestrata of index patients* | *Closest family relatives of brain disorder patients* | *Comparison relative* |
|---|---|---|---|---|
| Children and young people (0-24 years) | Fathers | Number of persons, n | 35,778 (100.0) | 35,778 (100.0) |
| | | Male, n (%) | 35,778 (100.0) | 35,778 (100.0) |
| | | Age, median (Q1;Q3) | 50.2 (45.2;54.8) | 50.3 (45.5;54.6) |
| | | CCI, n (%) | | |
| | | 0 | 31,339 (87.6) | 31,908 (89.2) |
| | | 1-2 | 3,977 (11.1) | 3,464 (9.7) |
| | | +3 | 462 (1.3) | 406 (1.1) |
| | | Education level, n (%) | | |
| | | No/Unknown education | 388 (1.1) | 388 (1.1) |
| | | Primary | 7,934 (22.2) | 7,934 (22.2) |
| | | Secondary | 18,405 (51.4) | 18,405 (51.4) |
| | | Tertiary | 9,051 (25.3) | 9,051 (25.3) |
| | | Relation to patient, n (%) | | |
| | | Father | 35,778 (100.0) | 35,778 (100.0) |
| Children and young people (0-24 years) | Mothers | Number of persons, n | 35,423 (100.0) | 35,423 (100.0) |
| | | Male, n (%) | 0 (0.0) | 0 (0.0) |
| | | Age, median (Q1;Q3) | 47.9 (43.1;52.5) | 48.2 (43.5;52.3) |
| | | CCI, n (%) | | |
| | | 0 | 30,470 (86.0) | 31,537 (89.0) |
| | | 1-2 | 4,564 (12.9) | 3,579 (10.1) |
| | | +3 | 389 (1.1) | 307 (0.9) |
| | | Education level, n (%) | | |
| | | No/Unknown education | 263 (0.7) | 263 (0.7) |
| | | Primary | 6,385 (18.0) | 6,385 (18.0) |
| | | Secondary | 15,615 (44.1) | 15,615 (44.1) |
| | | Tertiary | 13,160 (37.2) | 13,160 (37.2) |
| | | Relation to patient, n (%) | | |
| | | Mother | 35,423 (100.0) | 35,423 (100.0) |
| Young adults and adults (25-64 years) | Children | Number of persons, n | 89,679 (100.0) | 89,679 (100.0) |
| | | Male, n (%) | 47,150 (52.6) | 47,150 (52.6) |
| | | Age, median (Q1;Q3) | 23.3 (15.8;30.8) | 23.0 (15.8;30.5) |
| | | CCI, n (%) | | |
| | | 0 | 84,181 (93.9) | 85,014 (94.8) |
| | | 1-2 | 5,311 (5.9) | 4,508 (5.0) |
| | | +3 | 187 (0.2) | 157 (0.2) |
| | | Education level, n (%) | | |
| | | No/Unknown education | 16,010 (17.9) | 16,010 (17.9) |
| | | Primary | 33,803 (37.7) | 33,803 (37.7) |
| | | Secondary | 25,277 (28.2) | 25,277 (28.2) |
| | | Tertiary | 14,589 (16.3) | 14,589 (16.3) |
| | | Relation to patient, n (%) | | |
| | | Child | 89,679 (100.0) | 89,679 (100.0) |
| Young adults and adults (25-64 years) | Parents | Number of persons, n | 50,058 (100.0) | 50,058 (100.0) |
| | | Male, n (%) | 14,461 (28.9) | 14,461 (28.9) |
| | | Age, median (Q1;Q3) | 69.6 (61.1;76.9) | 69.6 (61.2;76.9) |

### Supplementary Table 267
### Sleep disorders - Prevalent cohort 2021
### Patient characteristics - relatives - not pooled

| *Age strata of index patients* | *Closest relative type* | *Characteristic of family relatives by agestrata of index patients* | *Closest family relatives of brain disorder patients* | *Comparison relative* |
|---|---|---|---|---|
| | | CCI, n (%) | | |
| | | 0 | 32,956 (65.8) | 34,082 (68.1) |
| | | 1-2 | 13,379 (26.7) | 12,603 (25.2) |
| | | +3 | 3,723 (7.4) | 3,373 (6.7) |
| | | Education level, n (%) | | |
| | | No/Unknown education | 737 (1.5) | 737 (1.5) |
| | | Primary | 15,786 (31.5) | 15,786 (31.5) |
| | | Secondary | 24,470 (48.9) | 24,470 (48.9) |
| | | Tertiary | 9,065 (18.1) | 9,065 (18.1) |
| | | Relation to patient, n (%) | | |
| | | Mother | 35,597 (71.1) | 35,597 (71.1) |
| | | Father | 14,461 (28.9) | 14,461 (28.9) |
| Young adults and adults (25-64 years) | Partners | Number of persons, n | 169,632 (100.0) | 169,632 (100.0) |
| | | Male, n (%) | 95,615 (56.4) | 95,615 (56.4) |
| | | Age, median (Q1;Q3) | 51.4 (41.9;58.5) | 51.6 (42.0;58.5) |
| | | CCI, n (%) | | |
| | | 0 | 146,326 (86.3) | 148,099 (87.3) |
| | | 1-2 | 20,616 (12.2) | 19,055 (11.2) |
| | | +3 | 2,690 (1.6) | 2,478 (1.5) |
| | | Education level, n (%) | | |
| | | No/Unknown education | 4,100 (2.4) | 4,100 (2.4) |
| | | Primary | 29,180 (17.2) | 29,180 (17.2) |
| | | Secondary | 84,573 (49.9) | 84,573 (49.9) |
| | | Tertiary | 51,779 (30.5) | 51,779 (30.5) |
| | | Relation to patient, n (%) | | |
| | | Partner | 169,632 (100.0) | 169,632 (100.0) |
| Young adults and adults (25-64 years) | Siblings | Number of persons, n | 33,827 (100.0) | 33,827 (100.0) |
| | | Male, n (%) | 18,554 (54.8) | 18,554 (54.8) |
| | | Age, median (Q1;Q3) | 53.9 (40.1;59.8) | 53.9 (40.0;59.8) |
| | | CCI, n (%) | | |
| | | 0 | 28,660 (84.7) | 28,722 (84.9) |
| | | 1-2 | 4,464 (13.2) | 4,414 (13.0) |
| | | +3 | 703 (2.1) | 691 (2.0) |
| | | Education level, n (%) | | |
| | | No/Unknown education | 94 (0.3) | 94 (0.3) |
| | | Primary | 7,315 (21.6) | 7,315 (21.6) |
| | | Secondary | 18,319 (54.2) | 18,319 (54.2) |
| | | Tertiary | 8,099 (23.9) | 8,099 (23.9) |
| | | Relation to patient, n (%) | | |
| | | Sibling | 33,827 (100.0) | 33,827 (100.0) |
| Older people (65+ years) | Children | Number of persons, n | 127,786 (100.0) | 127,786 (100.0) |
| | | Male, n (%) | 66,485 (52.0) | 66,485 (52.0) |
| | | Age, median (Q1;Q3) | 51.2 (44.9;57.1) | 51.1 (44.9;56.9) |
| | | CCI, n (%) | | |
| | | 0 | 110,974 (86.8) | 112,201 (87.8) |

**Supplementary Table 267**
**Sleep disorders - Prevalent cohort 2021**
**Patient characteristics - relatives - not pooled**

| *Age strata of index patients* | *Closest relative type* | *Characteristic of family relatives by agestrata of index patients* | *Closest family relatives of brain disorder patients* | *Comparison relative* |
|---|---|---|---|---|
| | | 1-2 | 14,910 (11.7) | 13,893 (10.9) |
| | | +3 | 1,902 (1.5) | 1,692 (1.3) |
| | | Education level, n (%) | | |
| | | No/Unknown education | 296 (0.2) | 296 (0.2) |
| | | Primary | 17,652 (13.8) | 17,652 (13.8) |
| | | Secondary | 61,317 (48.0) | 61,317 (48.0) |
| | | Tertiary | 48,521 (38.0) | 48,521 (38.0) |
| | | Relation to patient, n (%) | | |
| | | Child | 127,786 (100.0) | 127,786 (100.0) |
| Older people (65+ years) | Partners | Number of persons, n | 95,623 (100.0) | 95,623 (100.0) |
| | | Male, n (%) | 50,138 (52.4) | 50,138 (52.4) |
| | | Age, median (Q1;Q3) | 73.2 (68.3;77.7) | 73.1 (68.5;77.6) |
| | | CCI, n (%) | | |
| | | 0 | 59,809 (62.5) | 60,889 (63.7) |
| | | 1-2 | 27,512 (28.8) | 26,810 (28.0) |
| | | +3 | 8,302 (8.7) | 7,924 (8.3) |
| | | Education level, n (%) | | |
| | | No/Unknown education | 5,926 (6.2) | 5,926 (6.2) |
| | | Primary | 21,250 (22.2) | 21,250 (22.2) |
| | | Secondary | 46,959 (49.1) | 46,959 (49.1) |
| | | Tertiary | 21,488 (22.5) | 21,488 (22.5) |
| | | Relation to patient, n (%) | | |
| | | Partner | 95,623 (100.0) | 95,623 (100.0) |
| Older people (65+ years) | Siblings | Number of persons, n | 3,679 (100.0) | 3,679 (100.0) |
| | | Male, n (%) | 1,976 (53.7) | 1,976 (53.7) |
| | | Age, median (Q1;Q3) | 62.5 (59.6;65.1) | 62.7 (59.8;65.1) |
| | | CCI, n (%) | | |
| | | 0 | 2,843 (77.3) | 2,849 (77.4) |
| | | 1-2 | 709 (19.3) | 694 (18.9) |
| | | +3 | 127 (3.5) | 136 (3.7) |
| | | Education level, n (%) | | |
| | | No/Unknown education | 20 (0.5) | 20 (0.5) |
| | | Primary | 546 (14.8) | 546 (14.8) |
| | | Secondary | 2,284 (62.1) | 2,284 (62.1) |
| | | Tertiary | 829 (22.5) | 829 (22.5) |
| | | Relation to patient, n (%) | | |
| | | Sibling | 3,679 (100.0) | 3,679 (100.0) |

### Supplementary Table 268
### Sleep disorders - Prevalent cohort 2021
### Cost analysis - pooled

| *Age strata of index patients* | *Closest relative type* | *Number of index patients included in cost component calculations* | *Cost component* | *Actual costs* | | *Attributable costs* | |
|---|---|---|---|---|---|---|---|
| | | | | *Total (million EUR)* | *Per person (EUR)* | *Total (million EUR)* | *Per person (EUR)* |
| Children and young people (0-24 years) | Fathers | 35,778 | Primary sector | 8.1 | 227 | 0.8 | 22 |
| | | 35,778 | Hospital care | 50.2 | 1,405 | 8.7 | 243 |
| | | 35,778 | Filled prescriptions | 6.8 | 190 | 1.6 | 43 |
| | | 34,725 | Lost productivity (income loss) | . | . | 208.1 | 5,992 |
| Children and young people (0-24 years) | Mothers | 35,423 | Primary sector | 13.3 | 376 | 2.3 | 66 |
| | | 35,423 | Hospital care | 63.7 | 1,799 | 14.5 | 410 |
| | | 35,423 | Filled prescriptions | 8.4 | 238 | 3.0 | 84 |
| | | 35,221 | Lost productivity (income loss) | . | . | 219.5 | 6,231 |
| Young adults and adults (25-64 years) | Pooled | 343,196 | Primary sector | 102.3 | 299 | 8.5 | 25 |
| | | 343,196 | Hospital care | 621.0 | 1,816 | 59.9 | 175 |
| | | 343,196 | Filled prescriptions | 77.7 | 227 | 10.8 | 31 |
| | | 250,679 | Lost productivity (income loss) | . | . | 589.1 | 2,350 |
| Older people (65+ years) | Pooled | 227,088 | Primary sector | 85.6 | 379 | 6.5 | 29 |
| | | 227,088 | Hospital care | 552.7 | 2,448 | 20.2 | 90 |
| | | 227,088 | Filled prescriptions | 72.2 | 320 | 6.2 | 28 |
| | | 125,557 | Lost productivity (income loss) | . | . | -82.4 | -656 |

### Supplementary Table 269
### Sleep disorders - Prevalent cohort 2021
### Cost analysis - not pooled

| *Age strata of index patients* | *Closest relative type* | *Number of index patients included in cost component calculations* | *Cost component* | *Actual costs* | | *Attributable costs* | |
|---|---|---|---|---|---|---|---|
| | | | | *Total (million EUR)* | *Per person (EUR)* | *Total (million EUR)* | *Per person (EUR)* |
| Children and young people (0-24 years) | Fathers | 35,778 | Primary sector | 8.1 | 227 | 0.8 | 22 |
| | | 35,778 | Hospital care | 50.2 | 1,405 | 8.7 | 243 |
| | | 35,778 | Filled prescriptions | 6.8 | 190 | 1.6 | 43 |
| | | 34,725 | Lost productivity (income loss) | . | . | 208.1 | 5,992 |
| Children and young people (0-24 years) | Mothers | 35,423 | Primary sector | 13.3 | 376 | 2.3 | 66 |
| | | 35,423 | Hospital care | 63.7 | 1,799 | 14.5 | 410 |
| | | 35,423 | Filled prescriptions | 8.4 | 238 | 3.0 | 84 |
| | | 35,221 | Lost productivity (income loss) | . | . | 219.5 | 6,231 |
| Young adults and adults (25-64 years) | Children | 89,679 | Primary sector | 20.4 | 228 | 2.5 | 28 |
| | | 89,679 | Hospital care | 104.0 | 1,163 | 14.3 | 160 |
| | | 89,679 | Filled prescriptions | 9.7 | 108 | 1.6 | 18 |
| | | 56,675 | Lost productivity (income loss) | . | . | 67.4 | 1,189 |
| Young adults and adults (25-64 years) | Parents | 50,058 | Primary sector | 22.7 | 459 | 1.6 | 32 |
| | | 50,058 | Hospital care | 168.5 | 3,407 | 13.8 | 278 |
| | | 50,058 | Filled prescriptions | 24.3 | 492 | 3.2 | 65 |
| | | 14,821 | Lost productivity (income loss) | . | . | 73.6 | 4,963 |
| Young adults and adults (25-64 years) | Partners | 169,632 | Primary sector | 49.1 | 290 | 3.7 | 22 |
| | | 169,632 | Hospital care | 281.5 | 1,662 | 25.0 | 148 |
| | | 169,632 | Filled prescriptions | 35.7 | 211 | 4.9 | 29 |
| | | 150,255 | Lost productivity (income loss) | . | . | 398.4 | 2,652 |
| Young adults and adults (25-64 years) | Siblings | 33,827 | Primary sector | 10.1 | 300 | 0.7 | 20 |
| | | 33,827 | Hospital care | 67.0 | 1,987 | 6.9 | 204 |
| | | 33,827 | Filled prescriptions | 8.0 | 238 | 1.0 | 29 |
| | | 28,928 | Lost productivity (income loss) | . | . | 49.7 | 1,717 |
| Older people (65+ years) | Children | 127,786 | Primary sector | 37.3 | 292 | 3.2 | 25 |
| | | 127,786 | Hospital care | 196.5 | 1,540 | 9.4 | 73 |
| | | 127,786 | Filled prescriptions | 25.4 | 199 | 2.5 | 19 |
| | | 118,153 | Lost productivity (income loss) | . | . | -82.2 | -696 |
| Older people (65+ years) | Partners | 95,623 | Primary sector | 47.0 | 496 | 3.1 | 33 |
| | | 95,623 | Hospital care | 348.3 | 3,683 | 12.4 | 131 |
| | | 95,623 | Filled prescriptions | 45.7 | 483 | 3.7 | 39 |
| | | 5,445 | Lost productivity (income loss) | . | . | -0.7 | -131 |
| Older people (65+ years) | Siblings | 3,679 | Primary sector | 1.4 | 371 | 0.1 | 25 |
| | | 3,679 | Hospital care | 7.9 | 2,159 | -1.5 | -409 |
| | | 3,679 | Filled prescriptions | 1.1 | 306 | 0.1 | 18 |
| | | 1,959 | Lost productivity (income loss) | . | . | 0.5 | 255 |

#### Supplementary Table 270
#### Stress related disorders - Incident cohort 2016-2021
#### Identification of populations of relatives

| *Agegroup of index patients in matched population* | | *Closest relative type identified* | | *Available control with same type of closest relative* |
|---|---|---|---|---|
| Children and young people (0-24 years) | n = 34,694 | Fathers | n = 26,418 (76%) | 96% |
| | | Mothers | n = 27,174 (78%) | 96% |
| | | No available closest relative | n = 3,198 (9%) | n/a |
| Young adults and adults (25-64 years) | n = 48,869 | Children | n = 9,051 (19%) | 93% |
| | | Parents | n = 8,481 (17%) | 91% |
| | | Partners | n = 23,392 (48%) | 95% |
| | | Siblings | n = 5,447 (11%) | 45% |
| | | Pooled | n = 46,371 (95%) | 88% |
| | | No available closest relative | n = 2,498 (5%) | n/a |
| Older people (65+ years) | n = 6,100 | Children | n = 2,538 (42%) | 93% |
| | | Partners | n = 2,628 (43%) | 95% |
| | | Siblings | n = 133 (2%) | 31% |
| | | Pooled | n = 5,299 (87%) | 93% |
| | | No available closest relative | n = 801 (13%) | n/a |

**Supplementary Table 271**
**Stress related disorders - Incident cohort 2016-2021**
**Patient characteristics - cases**

| *Age strata of index patients* | *Characteristic* | |
|---|---|---|
| Children and young people (0-24 years) | Number of persons, n | 30,330 (100.0) |
| | Male, n (%) | 11,569 (38.1) |
| | Age, median (Q1;Q3) | 16.8 (14.3;20.1) |
| | CCI, n (%) | |
| | 0 | 27,880 (91.9) |
| | 1-2 | 2,377 (7.8) |
| | +3 | 73 (0.2) |
| | Education level, n (%) | |
| | No/Unknown education | 5,369 (17.7) |
| | Primary | 21,187 (69.9) |
| | Secondary | 3,545 (11.7) |
| | Tertiary | 229 (0.8) |
| Young adults and adults (25-64 years) | Number of persons, n | 40,982 (100.0) |
| | Male, n (%) | 16,312 (39.8) |
| | Age, median (Q1;Q3) | 43.3 (33.7;52.2) |
| | CCI, n (%) | |
| | 0 | 33,956 (82.9) |
| | 1-2 | 6,043 (14.7) |
| | +3 | 983 (2.4) |
| | Education level, n (%) | |
| | No/Unknown education | 1,434 (3.5) |
| | Primary | 11,596 (28.3) |
| | Secondary | 16,415 (40.1) |
| | Tertiary | 11,537 (28.2) |
| Older people (65+ years) | Number of persons, n | 4,931 (100.0) |
| | Male, n (%) | 1,976 (40.1) |
| | Age, median (Q1;Q3) | 74.3 (69.6;80.2) |
| | CCI, n (%) | |
| | 0 | 2,100 (42.6) |
| | 1-2 | 1,838 (37.3) |
| | +3 | 993 (20.1) |
| | Education level, n (%) | |
| | No/Unknown education | 292 (5.9) |
| | Primary | 1,546 (31.4) |
| | Secondary | 2,223 (45.1) |
| | Tertiary | 870 (17.6) |

#### Supplementary Table 272
#### Stress related disorders - Incident cohort 2016-2021
#### Patient characteristics - relatives - pooled

| *Age strata of index patients* | *Closest relative type* | *Characteristic of family relatives by agestrata of index patients* | *Closest family relatives of brain disorder patients* | *Comparison relative* |
|---|---|---|---|---|
| Children and young people (0-24 years) | Fathers | Number of persons, n | 25,411 (100.0) | 25,411 (100.0) |
| | | Male, n (%) | 25,411 (100.0) | 25,411 (100.0) |
| | | Age, median (Q1;Q3) | 49.0 (44.6;53.6) | 49.1 (45.0;53.3) |
| | | CCI, n (%) | | |
| | | 0 | 21,996 (86.6) | 22,603 (88.9) |
| | | 1-2 | 2,977 (11.7) | 2,495 (9.8) |
| | | +3 | 438 (1.7) | 313 (1.2) |
| | | Education level, n (%) | | |
| | | No/Unknown education | 341 (1.3) | 341 (1.3) |
| | | Primary | 5,915 (23.3) | 5,915 (23.3) |
| | | Secondary | 13,233 (52.1) | 13,233 (52.1) |
| | | Tertiary | 5,922 (23.3) | 5,922 (23.3) |
| | | Relation to patient, n (%) | | |
| | | Father | 25,411 (100.0) | 25,411 (100.0) |
| Children and young people (0-24 years) | Mothers | Number of persons, n | 26,194 (100.0) | 26,194 (100.0) |
| | | Male, n (%) | 0 (0.0) | 0 (0.0) |
| | | Age, median (Q1;Q3) | 46.7 (42.4;51.0) | 46.9 (43.0;50.8) |
| | | CCI, n (%) | | |
| | | 0 | 22,410 (85.6) | 23,274 (88.9) |
| | | 1-2 | 3,413 (13.0) | 2,662 (10.2) |
| | | +3 | 371 (1.4) | 258 (1.0) |
| | | Education level, n (%) | | |
| | | No/Unknown education | 267 (1.0) | 267 (1.0) |
| | | Primary | 5,198 (19.8) | 5,198 (19.8) |
| | | Secondary | 11,964 (45.7) | 11,964 (45.7) |
| | | Tertiary | 8,765 (33.5) | 8,765 (33.5) |
| | | Relation to patient, n (%) | | |
| | | Mother | 26,194 (100.0) | 26,194 (100.0) |
| Young adults and adults (25-64 years) | Pooled | Number of persons, n | 40,982 (100.0) | 40,982 (100.0) |
| | | Male, n (%) | 21,888 (53.4) | 21,888 (53.4) |
| | | Age, median (Q1;Q3) | 44.3 (29.8;56.6) | 44.3 (29.8;56.7) |
| | | CCI, n (%) | | |
| | | 0 | 34,797 (84.9) | 35,606 (86.9) |
| | | 1-2 | 5,202 (12.7) | 4,608 (11.2) |
| | | +3 | 983 (2.4) | 768 (1.9) |
| | | Education level, n (%) | | |
| | | No/Unknown education | 3,838 (9.4) | 3,838 (9.4) |
| | | Primary | 11,506 (28.1) | 11,506 (28.1) |
| | | Secondary | 16,929 (41.3) | 16,929 (41.3) |
| | | Tertiary | 8,709 (21.3) | 8,709 (21.3) |
| | | Relation to patient, n (%) | | |
| | | Mother | 5,874 (14.3) | 5,874 (14.3) |
| | | Father | 1,894 (4.6) | 1,894 (4.6) |
| | | Partner | 22,265 (54.3) | 22,265 (54.3) |
| | | Child | 8,493 (20.7) | 8,493 (20.7) |

#### Supplementary Table 272
#### Stress related disorders - Incident cohort 2016-2021
#### Patient characteristics - relatives - pooled

| *Age strata of index patients* | *Closest relative type* | *Characteristic of family relatives by agestrata of index patients* | *Closest family relatives of brain disorder patients* | *Comparison relative* |
|---|---|---|---|---|
| | | Sibling | 2,456 (6.0) | 2,456 (6.0) |
| Older people (65+ years) | Pooled | Number of persons, n | 4,931 (100.0) | 4,931 (100.0) |
| | | Male, n (%) | 2,638 (53.5) | 2,638 (53.5) |
| | | Age, median (Q1;Q3) | 63.1 (51.5;73.4) | 63.2 (51.5;73.1) |
| | | CCI, n (%) | | |
| | | 0 | 3,383 (68.6) | 3,619 (73.4) |
| | | 1-2 | 1,106 (22.4) | 1,030 (20.9) |
| | | +3 | 442 (9.0) | 282 (5.7) |
| | | Education level, n (%) | | |
| | | No/Unknown education | 155 (3.1) | 155 (3.1) |
| | | Primary | 1,107 (22.4) | 1,107 (22.4) |
| | | Secondary | 2,360 (47.9) | 2,360 (47.9) |
| | | Tertiary | 1,309 (26.5) | 1,309 (26.5) |
| | | Relation to patient, n (%) | | |
| | | Partner | 2,508 (50.9) | 2,508 (50.9) |
| | | Child | 2,381 (48.3) | 2,381 (48.3) |
| | | Sibling | 42 (0.9) | 42 (0.9) |

#### Supplementary Table 273
#### Stress related disorders - Incident cohort 2016-2021
#### Patient characteristics - relatives - not pooled

| *Age strata of index patients* | *Closest relative type* | *Characteristic of family relatives by agestrata of index patients* | *Closest family relatives of brain disorder patients* | *Comparison relative* |
|---|---|---|---|---|
| Children and young people (0-24 years) | Fathers | Number of persons, n | 25,411 (100.0) | 25,411 (100.0) |
| | | Male, n (%) | 25,411 (100.0) | 25,411 (100.0) |
| | | Age, median (Q1;Q3) | 49.0 (44.6;53.6) | 49.1 (45.0;53.3) |
| | | CCI, n (%) | | |
| | | 0 | 21,996 (86.6) | 22,603 (88.9) |
| | | 1-2 | 2,977 (11.7) | 2,495 (9.8) |
| | | +3 | 438 (1.7) | 313 (1.2) |
| | | Education level, n (%) | | |
| | | No/Unknown education | 341 (1.3) | 341 (1.3) |
| | | Primary | 5,915 (23.3) | 5,915 (23.3) |
| | | Secondary | 13,233 (52.1) | 13,233 (52.1) |
| | | Tertiary | 5,922 (23.3) | 5,922 (23.3) |
| | | Relation to patient, n (%) | | |
| | | Father | 25,411 (100.0) | 25,411 (100.0) |
| Children and young people (0-24 years) | Mothers | Number of persons, n | 26,194 (100.0) | 26,194 (100.0) |
| | | Male, n (%) | 0 (0.0) | 0 (0.0) |
| | | Age, median (Q1;Q3) | 46.7 (42.4;51.0) | 46.9 (43.0;50.8) |
| | | CCI, n (%) | | |
| | | 0 | 22,410 (85.6) | 23,274 (88.9) |
| | | 1-2 | 3,413 (13.0) | 2,662 (10.2) |
| | | +3 | 371 (1.4) | 258 (1.0) |
| | | Education level, n (%) | | |
| | | No/Unknown education | 267 (1.0) | 267 (1.0) |
| | | Primary | 5,198 (19.8) | 5,198 (19.8) |
| | | Secondary | 11,964 (45.7) | 11,964 (45.7) |
| | | Tertiary | 8,765 (33.5) | 8,765 (33.5) |
| | | Relation to patient, n (%) | | |
| | | Mother | 26,194 (100.0) | 26,194 (100.0) |
| Young adults and adults (25-64 years) | Children | Number of persons, n | 8,493 (100.0) | 8,493 (100.0) |
| | | Male, n (%) | 4,538 (53.4) | 4,538 (53.4) |
| | | Age, median (Q1;Q3) | 17.3 (10.3;24.4) | 16.9 (9.6;24.0) |
| | | CCI, n (%) | | |
| | | 0 | 7,935 (93.4) | 8,026 (94.5) |
| | | 1-2 | 538 (6.3) | 448 (5.3) |
| | | +3 | 20 (0.2) | 19 (0.2) |
| | | Education level, n (%) | | |
| | | No/Unknown education | 2,991 (35.2) | 2,991 (35.2) |
| | | Primary | 3,415 (40.2) | 3,415 (40.2) |
| | | Secondary | 1,446 (17.0) | 1,446 (17.0) |
| | | Tertiary | 641 (7.5) | 641 (7.5) |
| | | Relation to patient, n (%) | | |
| | | Child | 8,493 (100.0) | 8,493 (100.0) |
| Young adults and adults (25-64 years) | Parents | Number of persons, n | 7,768 (100.0) | 7,768 (100.0) |
| | | Male, n (%) | 1,894 (24.4) | 1,894 (24.4) |
| | | Age, median (Q1;Q3) | 63.1 (57.0;71.3) | 63.1 (57.1;71.3) |

#### Supplementary Table 273
#### Stress related disorders - Incident cohort 2016-2021
#### Patient characteristics - relatives - not pooled

| *Age strata of index patients* | *Closest relative type* | *Characteristic of family relatives by agestrata of index patients* | *Closest family relatives of brain disorder patients* | *Comparison relative* |
|---|---|---|---|---|
| | | CCI, n (%) | | |
| | | 0 | 5,266 (67.8) | 5,558 (71.5) |
| | | 1-2 | 1,961 (25.2) | 1,778 (22.9) |
| | | +3 | 541 (7.0) | 432 (5.6) |
| | | Education level, n (%) | | |
| | | No/Unknown education | 138 (1.8) | 138 (1.8) |
| | | Primary | 2,422 (31.2) | 2,422 (31.2) |
| | | Secondary | 3,693 (47.5) | 3,693 (47.5) |
| | | Tertiary | 1,515 (19.5) | 1,515 (19.5) |
| | | Relation to patient, n (%) | | |
| | | Mother | 5,874 (75.6) | 5,874 (75.6) |
| | | Father | 1,894 (24.4) | 1,894 (24.4) |
| Young adults and adults (25-64 years) | Partners | Number of persons, n | 22,265 (100.0) | 22,265 (100.0) |
| | | Male, n (%) | 14,012 (62.9) | 14,012 (62.9) |
| | | Age, median (Q1;Q3) | 44.7 (35.7;53.5) | 44.8 (35.7;53.4) |
| | | CCI, n (%) | | |
| | | 0 | 19,475 (87.5) | 19,867 (89.2) |
| | | 1-2 | 2,419 (10.9) | 2,118 (9.5) |
| | | +3 | 371 (1.7) | 280 (1.3) |
| | | Education level, n (%) | | |
| | | No/Unknown education | 701 (3.1) | 701 (3.1) |
| | | Primary | 5,001 (22.5) | 5,001 (22.5) |
| | | Secondary | 10,486 (47.1) | 10,486 (47.1) |
| | | Tertiary | 6,077 (27.3) | 6,077 (27.3) |
| | | Relation to patient, n (%) | | |
| | | Partner | 22,265 (100.0) | 22,265 (100.0) |
| Young adults and adults (25-64 years) | Siblings | Number of persons, n | 2,456 (100.0) | 2,456 (100.0) |
| | | Male, n (%) | 1,444 (58.8) | 1,444 (58.8) |
| | | Age, median (Q1;Q3) | 46.7 (30.5;56.2) | 46.7 (30.5;56.3) |
| | | CCI, n (%) | | |
| | | 0 | 2,121 (86.4) | 2,155 (87.7) |
| | | 1-2 | 284 (11.6) | 264 (10.7) |
| | | +3 | 51 (2.1) | 37 (1.5) |
| | | Education level, n (%) | | |
| | | No/Unknown education | 8 (0.3) | 8 (0.3) |
| | | Primary | 668 (27.2) | 668 (27.2) |
| | | Secondary | 1,304 (53.1) | 1,304 (53.1) |
| | | Tertiary | 476 (19.4) | 476 (19.4) |
| | | Relation to patient, n (%) | | |
| | | Sibling | 2,456 (100.0) | 2,456 (100.0) |
| Older people (65+ years) | Children | Number of persons, n | 2,381 (100.0) | 2,381 (100.0) |
| | | Male, n (%) | 1,274 (53.5) | 1,274 (53.5) |
| | | Age, median (Q1;Q3) | 51.2 (44.8;57.2) | 51.1 (44.8;56.9) |
| | | CCI, n (%) | | |
| | | 0 | 2,027 (85.1) | 2,069 (86.9) |

**Supplementary Table 273**
**Stress related disorders - Incident cohort 2016-2021**
**Patient characteristics - relatives - not pooled**

| *Age strata of index patients* | *Closest relative type* | *Characteristic of family relatives by agestrata of index patients* | *Closest family relatives of brain disorder patients* | *Comparison relative* |
|---|---|---|---|---|
| | | 1-2 | 305 (12.8) | 274 (11.5) |
| | | +3 | 49 (2.1) | 38 (1.6) |
| | | Education level, n (%) | | |
| | | No/Unknown education | 6 (0.3) | 6 (0.3) |
| | | Primary | 404 (17.0) | 404 (17.0) |
| | | Secondary | 1,166 (49.0) | 1,166 (49.0) |
| | | Tertiary | 805 (33.8) | 805 (33.8) |
| | | Relation to patient, n (%) | | |
| | | Child | 2,381 (100.0) | 2,381 (100.0) |
| Older people (65+ years) | Partners | Number of persons, n | 2,508 (100.0) | 2,508 (100.0) |
| | | Male, n (%) | 1,342 (53.5) | 1,342 (53.5) |
| | | Age, median (Q1;Q3) | 73.3 (68.3;78.5) | 73.0 (68.5;78.5) |
| | | CCI, n (%) | | |
| | | 0 | 1,322 (52.7) | 1,516 (60.4) |
| | | 1-2 | 796 (31.7) | 748 (29.8) |
| | | +3 | 390 (15.6) | 244 (9.7) |
| | | Education level, n (%) | | |
| | | No/Unknown education | 149 (5.9) | 149 (5.9) |
| | | Primary | 693 (27.6) | 693 (27.6) |
| | | Secondary | 1,173 (46.8) | 1,173 (46.8) |
| | | Tertiary | 493 (19.7) | 493 (19.7) |
| | | Relation to patient, n (%) | | |
| | | Partner | 2,508 (100.0) | 2,508 (100.0) |
| Older people (65+ years) | Siblings | Number of persons, n | 42 (100.0) | 42 (100.0) |
| | | Male, n (%) | 22 (52.4) | 22 (52.4) |
| | | Age, median (Q1;Q3) | 62.6 (60.7;64.6) | 62.4 (59.7;65.3) |
| | | CCI, n (%) | | |
| | | 0 | * | 34 (81.0) |
| | | 1-2 | * | 8 (19.0) |
| | | +3 | * | 0 (0.0) |
| | | Education level, n (%) | | |
| | | No/Unknown education | 0 (0.0) | 0 (0.0) |
| | | Primary | 10 (23.8) | 10 (23.8) |
| | | Secondary | 21 (50.0) | 21 (50.0) |
| | | Tertiary | 11 (26.2) | 11 (26.2) |
| | | Relation to patient, n (%) | | |
| | | Sibling | 42 (100.0) | 42 (100.0) |

### Supplementary Table 274
### Stress related disorders - Incident cohort 2016-2021
### Cost analysis - pooled

| | | | | *Actual costs* | | *Attributable costs* | |
|---|---|---|---|---|---|---|---|
| *Age strata of index patients* | *Closest relative type* | *Number of index patients included in cost component calculations* | *Cost component* | *Total (million EUR)* | *Per person (EUR)* | *Total (million EUR)* | *Per person (EUR)* |
| Children and young people (0-24 years) | Fathers | 25,411 | Primary sector | 5.9 | 233 | 0.5 | 22 |
| | | 25,411 | Hospital care | 44.3 | 1,747 | 11.3 | 445 |
| | | 25,411 | Filled prescriptions | 5.4 | 214 | 1.5 | 59 |
| | | 24,627 | Lost productivity (income loss) | . | . | 244.7 | 9,936 |
| Children and young people (0-24 years) | Mothers | 26,194 | Primary sector | 9.9 | 378 | 1.5 | 57 |
| | | 26,194 | Hospital care | 52.4 | 2,005 | 15.4 | 588 |
| | | 26,194 | Filled prescriptions | 6.5 | 250 | 2.0 | 77 |
| | | 25,998 | Lost productivity (income loss) | . | . | 201.2 | 7,740 |
| Young adults and adults (25-64 years) | Pooled | 40,982 | Primary sector | 12.5 | 307 | 1.7 | 42 |
| | | 40,982 | Hospital care | 83.6 | 2,053 | 16.6 | 407 |
| | | 40,982 | Filled prescriptions | 9.5 | 233 | 1.5 | 37 |
| | | 30,318 | Lost productivity (income loss) | . | . | 270.8 | 8,931 |
| Older people (65+ years) | Pooled | 4,931 | Primary sector | 2.1 | 442 | 0.3 | 64 |
| | | 4,931 | Hospital care | 19.6 | 4,065 | 6.3 | 1,314 |
| | | 4,931 | Filled prescriptions | 2.1 | 430 | 0.4 | 88 |
| | | 2,369 | Lost productivity (income loss) | . | . | 17.0 | 7,184 |

#### Supplementary Table 275
#### Stress related disorders - Incident cohort 2016-2021
#### Cost analysis - not pooled

| *Age strata of index patients* | *Closest relative type* | *Number of index patients included in cost component calculations* | *Cost component* | *Actual costs* Total (million EUR) | *Actual costs* Per person (EUR) | *Attributable costs* Total (million EUR) | *Attributable costs* Per person (EUR) |
|---|---|---|---|---|---|---|---|
| Children and young people (0-24 years) | Fathers | 25,411 | Primary sector | 5.9 | 233 | 0.5 | 22 |
| | | 25,411 | Hospital care | 44.3 | 1,747 | 11.3 | 445 |
| | | 25,411 | Filled prescriptions | 5.4 | 214 | 1.5 | 59 |
| | | 24,627 | Lost productivity (income loss) | . | . | 244.7 | 9,936 |
| Children and young people (0-24 years) | Mothers | 26,194 | Primary sector | 9.9 | 378 | 1.5 | 57 |
| | | 26,194 | Hospital care | 52.4 | 2,005 | 15.4 | 588 |
| | | 26,194 | Filled prescriptions | 6.5 | 250 | 2.0 | 77 |
| | | 25,998 | Lost productivity (income loss) | . | . | 201.2 | 7,740 |
| Young adults and adults (25-64 years) | Children | 8,493 | Primary sector | 1.7 | 201 | 0.2 | 27 |
| | | 8,493 | Hospital care | 10.0 | 1,189 | 2.6 | 302 |
| | | 8,493 | Filled prescriptions | 0.8 | 94 | 0.1 | 9 |
| | | 3,641 | Lost productivity (income loss) | . | . | 5.2 | 1,440 |
| Young adults and adults (25-64 years) | Parents | 7,768 | Primary sector | 3.4 | 447 | 0.3 | 41 |
| | | 7,768 | Hospital care | 27.5 | 3,581 | 4.2 | 543 |
| | | 7,768 | Filled prescriptions | 3.8 | 490 | 0.7 | 93 |
| | | 3,576 | Lost productivity (income loss) | . | . | 27.9 | 7,806 |
| Young adults and adults (25-64 years) | Partners | 22,265 | Primary sector | 6.7 | 303 | 1.1 | 51 |
| | | 22,265 | Hospital care | 41.7 | 1,881 | 9.8 | 442 |
| | | 22,265 | Filled prescriptions | 4.4 | 199 | 0.7 | 31 |
| | | 20,865 | Lost productivity (income loss) | . | . | 232.7 | 11,155 |
| Young adults and adults (25-64 years) | Siblings | 2,456 | Primary sector | 0.7 | 275 | 0.0 | 10 |
| | | 2,456 | Hospital care | 4.4 | 1,791 | 0.1 | 31 |
| | | 2,456 | Filled prescriptions | 0.5 | 220 | 0.1 | 21 |
| | | 2,236 | Lost productivity (income loss) | . | . | 4.9 | 2,183 |
| Older people (65+ years) | Children | 2,381 | Primary sector | 0.7 | 303 | 0.1 | 27 |
| | | 2,381 | Hospital care | 4.5 | 1,885 | 0.9 | 385 |
| | | 2,381 | Filled prescriptions | 0.6 | 232 | 0.1 | 29 |
| | | 2,216 | Lost productivity (income loss) | . | . | 18.3 | 8,280 |
| Older people (65+ years) | Partners | 2,508 | Primary sector | 1.4 | 582 | 0.3 | 105 |
| | | 2,508 | Hospital care | 14.9 | 6,218 | 5.4 | 2,241 |
| | | 2,508 | Filled prescriptions | 1.5 | 625 | 0.3 | 142 |
| | | 133 | Lost productivity (income loss) | . | . | -0.0 | -46 |
| Older people (65+ years) | Siblings | 42 | Primary sector | 0.0 | 358 | -0.0 | -156 |
| | | 42 | Hospital care | 0.2 | 4,348 | 0.0 | 840 |
| | | 42 | Filled prescriptions | 0.0 | 525 | 0.0 | 300 |
| | | 20 | Lost productivity (income loss) | . | . | -1.3 | -66,184 |

### Supplementary Table 276
### Stress related disorders - Prevalent cohort 2021
### Identification of populations of relatives

| *Agegroup of index patients in matched population* | | *Closest relative type identified* | | *Available control with same type of closest relative* |
|---|---|---|---|---|
| Children and young people (0-24 years) | n = 41,901 | Fathers | n = 31,811 (76%) | 96% |
| | | Mothers | n = 31,871 (76%) | 96% |
| | | No available closest relative | n = 3,768 (9%) | n/a |
| Young adults and adults (25-64 years) | n = 168,671 | Children | n = 35,057 (21%) | 93% |
| | | Parents | n = 33,582 (20%) | 90% |
| | | Partners | n = 70,863 (42%) | 94% |
| | | Siblings | n = 23,404 (14%) | 45% |
| | | Pooled | n = 162,906 (97%) | 86% |
| | | No available closest relative | n = 5,765 (3%) | n/a |
| Older people (65+ years) | n = 25,445 | Children | n = 11,025 (43%) | 93% |
| | | Partners | n = 9,692 (38%) | 95% |
| | | Siblings | n = 1,379 (5%) | 55% |
| | | Pooled | n = 22,096 (87%) | 91% |
| | | No available closest relative | n = 3,349 (13%) | n/a |

**Supplementary Table 277**
**Stress related disorders - Prevalent cohort 2021**
**Patient characteristics - cases**

| *Age strata of index patients* | *Characteristic* | |
|---|---|---|
| Children and young people (0-24 years) | Number of persons, n | 36,832 (100.0) |
| | Male, n (%) | 14,506 (39.4) |
| | Age, median (Q1;Q3) | 19.8 (16.6;22.5) |
| | CCI, n (%) | |
| | 0 | 33,132 (90.0) |
| | 1-2 | 3,502 (9.5) |
| | +3 | 198 (0.5) |
| | Education level, n (%) | |
| | No/Unknown education | 3,476 (9.4) |
| | Primary | 25,976 (70.5) |
| | Secondary | 6,938 (18.8) |
| | Tertiary | 442 (1.2) |
| Young adults and adults (25-64 years) | Number of persons, n | 140,902 (100.0) |
| | Male, n (%) | 54,178 (38.5) |
| | Age, median (Q1;Q3) | 44.9 (34.5;54.1) |
| | CCI, n (%) | |
| | 0 | 113,927 (80.9) |
| | 1-2 | 23,150 (16.4) |
| | +3 | 3,825 (2.7) |
| | Education level, n (%) | |
| | No/Unknown education | 3,457 (2.5) |
| | Primary | 44,429 (31.5) |
| | Secondary | 57,845 (41.1) |
| | Tertiary | 35,171 (25.0) |
| Older people (65+ years) | Number of persons, n | 20,249 (100.0) |
| | Male, n (%) | 7,341 (36.3) |
| | Age, median (Q1;Q3) | 71.3 (67.6;76.6) |
| | CCI, n (%) | |
| | 0 | 9,616 (47.5) |
| | 1-2 | 7,415 (36.6) |
| | +3 | 3,218 (15.9) |
| | Education level, n (%) | |
| | No/Unknown education | 1,000 (4.9) |
| | Primary | 5,812 (28.7) |
| | Secondary | 8,708 (43.0) |
| | Tertiary | 4,729 (23.4) |

**Supplementary Table 278**
**Stress related disorders - Prevalent cohort 2021**
**Patient characteristics - relatives - pooled**

| *Age strata of index patients* | *Closest relative type* | *Characteristic of family relatives by agestrata of index patients* | *Closest family relatives of brain disorder patients* | *Comparison relative* |
|---|---|---|---|---|
| Children and young people (0-24 years) | Fathers | Number of persons, n | 30,691 (100.0) | 30,691 (100.0) |
| | | Male, n (%) | 30,691 (100.0) | 30,691 (100.0) |
| | | Age, median (Q1;Q3) | 51.2 (46.8;55.6) | 51.3 (47.3;55.3) |
| | | CCI, n (%) | | |
| | | 0 | 26,030 (84.8) | 26,872 (87.6) |
| | | 1-2 | 4,058 (13.2) | 3,372 (11.0) |
| | | +3 | 603 (2.0) | 447 (1.5) |
| | | Education level, n (%) | | |
| | | No/Unknown education | 337 (1.1) | 337 (1.1) |
| | | Primary | 6,921 (22.6) | 6,921 (22.6) |
| | | Secondary | 16,288 (53.1) | 16,288 (53.1) |
| | | Tertiary | 7,145 (23.3) | 7,145 (23.3) |
| | | Relation to patient, n (%) | | |
| | | Father | 30,691 (100.0) | 30,691 (100.0) |
| Children and young people (0-24 years) | Mothers | Number of persons, n | 30,785 (100.0) | 30,785 (100.0) |
| | | Male, n (%) | 0 (0.0) | 0 (0.0) |
| | | Age, median (Q1;Q3) | 48.9 (44.9;53.2) | 49.1 (45.3;53.0) |
| | | CCI, n (%) | | |
| | | 0 | 25,911 (84.2) | 26,917 (87.4) |
| | | 1-2 | 4,383 (14.2) | 3,530 (11.5) |
| | | +3 | 491 (1.6) | 338 (1.1) |
| | | Education level, n (%) | | |
| | | No/Unknown education | 209 (0.7) | 209 (0.7) |
| | | Primary | 5,797 (18.8) | 5,797 (18.8) |
| | | Secondary | 14,237 (46.2) | 14,237 (46.2) |
| | | Tertiary | 10,542 (34.2) | 10,542 (34.2) |
| | | Relation to patient, n (%) | | |
| | | Mother | 30,785 (100.0) | 30,785 (100.0) |
| Young adults and adults (25-64 years) | Pooled | Number of persons, n | 140,902 (100.0) | 140,902 (100.0) |
| | | Male, n (%) | 75,575 (53.6) | 75,575 (53.6) |
| | | Age, median (Q1;Q3) | 45.6 (29.8;58.7) | 45.6 (29.8;58.8) |
| | | CCI, n (%) | | |
| | | 0 | 117,631 (83.5) | 120,758 (85.7) |
| | | 1-2 | 19,507 (13.8) | 17,010 (12.1) |
| | | +3 | 3,764 (2.7) | 3,134 (2.2) |
| | | Education level, n (%) | | |
| | | No/Unknown education | 11,608 (8.2) | 11,608 (8.2) |
| | | Primary | 40,347 (28.6) | 40,347 (28.6) |
| | | Secondary | 59,921 (42.5) | 59,921 (42.5) |
| | | Tertiary | 29,026 (20.6) | 29,026 (20.6) |
| | | Relation to patient, n (%) | | |
| | | Mother | 22,852 (16.2) | 22,852 (16.2) |
| | | Father | 7,484 (5.3) | 7,484 (5.3) |
| | | Partner | 67,225 (47.7) | 67,225 (47.7) |
| | | Child | 32,743 (23.2) | 32,743 (23.2) |

**Supplementary Table 278**
**Stress related disorders - Prevalent cohort 2021**
**Patient characteristics - relatives - pooled**

| *Age strata of index patients* | *Closest relative type* | *Characteristic of family relatives by agestrata of index patients* | *Closest family relatives of brain disorder patients* | *Comparison relative* |
|---|---|---|---|---|
| | | Sibling | 10,598 (7.5) | 10,598 (7.5) |
| Older people (65+ years) | Pooled | Number of persons, n | 20,249 (100.0) | 20,249 (100.0) |
| | | Male, n (%) | 11,009 (54.4) | 11,009 (54.4) |
| | | Age, median (Q1;Q3) | 60.7 (47.8;70.3) | 60.7 (47.8;70.3) |
| | | CCI, n (%) | | |
| | | 0 | 14,919 (73.7) | 15,386 (76.0) |
| | | 1-2 | 4,149 (20.5) | 3,847 (19.0) |
| | | +3 | 1,181 (5.8) | 1,016 (5.0) |
| | | Education level, n (%) | | |
| | | No/Unknown education | 585 (2.9) | 585 (2.9) |
| | | Primary | 4,039 (19.9) | 4,039 (19.9) |
| | | Secondary | 9,701 (47.9) | 9,701 (47.9) |
| | | Tertiary | 5,924 (29.3) | 5,924 (29.3) |
| | | Relation to patient, n (%) | | |
| | | Partner | 9,223 (45.5) | 9,223 (45.5) |
| | | Child | 10,254 (50.6) | 10,254 (50.6) |
| | | Sibling | 772 (3.8) | 772 (3.8) |

**Supplementary Table 279**
**Stress related disorders - Prevalent cohort 2021**
**Patient characteristics - relatives - not pooled**

| *Age strata of index patients* | *Closest relative type* | *Characteristic of family relatives by agestrata of index patients* | *Closest family relatives of brain disorder patients* | *Comparison relative* |
|---|---|---|---|---|
| Children and young people (0-24 years) | Fathers | Number of persons, n | 30,691 (100.0) | 30,691 (100.0) |
| | | Male, n (%) | 30,691 (100.0) | 30,691 (100.0) |
| | | Age, median (Q1;Q3) | 51.2 (46.8;55.6) | 51.3 (47.3;55.3) |
| | | CCI, n (%) | | |
| | | 0 | 26,030 (84.8) | 26,872 (87.6) |
| | | 1-2 | 4,058 (13.2) | 3,372 (11.0) |
| | | +3 | 603 (2.0) | 447 (1.5) |
| | | Education level, n (%) | | |
| | | No/Unknown education | 337 (1.1) | 337 (1.1) |
| | | Primary | 6,921 (22.6) | 6,921 (22.6) |
| | | Secondary | 16,288 (53.1) | 16,288 (53.1) |
| | | Tertiary | 7,145 (23.3) | 7,145 (23.3) |
| | | Relation to patient, n (%) | | |
| | | Father | 30,691 (100.0) | 30,691 (100.0) |
| Children and young people (0-24 years) | Mothers | Number of persons, n | 30,785 (100.0) | 30,785 (100.0) |
| | | Male, n (%) | 0 (0.0) | 0 (0.0) |
| | | Age, median (Q1;Q3) | 48.9 (44.9;53.2) | 49.1 (45.3;53.0) |
| | | CCI, n (%) | | |
| | | 0 | 25,911 (84.2) | 26,917 (87.4) |
| | | 1-2 | 4,383 (14.2) | 3,530 (11.5) |
| | | +3 | 491 (1.6) | 338 (1.1) |
| | | Education level, n (%) | | |
| | | No/Unknown education | 209 (0.7) | 209 (0.7) |
| | | Primary | 5,797 (18.8) | 5,797 (18.8) |
| | | Secondary | 14,237 (46.2) | 14,237 (46.2) |
| | | Tertiary | 10,542 (34.2) | 10,542 (34.2) |
| | | Relation to patient, n (%) | | |
| | | Mother | 30,785 (100.0) | 30,785 (100.0) |
| Young adults and adults (25-64 years) | Children | Number of persons, n | 32,743 (100.0) | 32,743 (100.0) |
| | | Male, n (%) | 17,621 (53.8) | 17,621 (53.8) |
| | | Age, median (Q1;Q3) | 19.3 (12.2;27.6) | 18.8 (11.5;27.2) |
| | | CCI, n (%) | | |
| | | 0 | 30,600 (93.5) | 30,969 (94.6) |
| | | 1-2 | 2,075 (6.3) | 1,709 (5.2) |
| | | +3 | 68 (0.2) | 65 (0.2) |
| | | Education level, n (%) | | |
| | | No/Unknown education | 9,302 (28.4) | 9,302 (28.4) |
| | | Primary | 13,406 (40.9) | 13,406 (40.9) |
| | | Secondary | 6,922 (21.1) | 6,922 (21.1) |
| | | Tertiary | 3,113 (9.5) | 3,113 (9.5) |
| | | Relation to patient, n (%) | | |
| | | Child | 32,743 (100.0) | 32,743 (100.0) |
| Young adults and adults (25-64 years) | Parents | Number of persons, n | 30,336 (100.0) | 30,336 (100.0) |
| | | Male, n (%) | 7,484 (24.7) | 7,484 (24.7) |
| | | Age, median (Q1;Q3) | 65.0 (58.3;73.2) | 65.1 (58.6;73.4) |

#### Supplementary Table 279
#### Stress related disorders - Prevalent cohort 2021
#### Patient characteristics - relatives - not pooled

| *Age strata of index patients* | *Closest relative type* | *Characteristic of family relatives by agestrata of index patients* | *Closest family relatives of brain disorder patients* | *Comparison relative* |
|---|---|---|---|---|
| | | CCI, n (%) | | |
| | | 0 | 19,933 (65.7) | 21,147 (69.7) |
| | | 1-2 | 8,061 (26.6) | 7,269 (24.0) |
| | | +3 | 2,342 (7.7) | 1,920 (6.3) |
| | | Education level, n (%) | | |
| | | No/Unknown education | 531 (1.8) | 531 (1.8) |
| | | Primary | 9,396 (31.0) | 9,396 (31.0) |
| | | Secondary | 14,573 (48.0) | 14,573 (48.0) |
| | | Tertiary | 5,836 (19.2) | 5,836 (19.2) |
| | | Relation to patient, n (%) | | |
| | | Mother | 22,852 (75.3) | 22,852 (75.3) |
| | | Father | 7,484 (24.7) | 7,484 (24.7) |
| Young adults and adults (25-64 years) | Partners | Number of persons, n | 67,225 (100.0) | 67,225 (100.0) |
| | | Male, n (%) | 44,480 (66.2) | 44,480 (66.2) |
| | | Age, median (Q1;Q3) | 45.7 (35.6;54.9) | 45.7 (35.7;54.9) |
| | | CCI, n (%) | | |
| | | 0 | 58,231 (86.6) | 59,585 (88.6) |
| | | 1-2 | 7,882 (11.7) | 6,724 (10.0) |
| | | +3 | 1,112 (1.7) | 916 (1.4) |
| | | Education level, n (%) | | |
| | | No/Unknown education | 1,744 (2.6) | 1,744 (2.6) |
| | | Primary | 14,686 (21.8) | 14,686 (21.8) |
| | | Secondary | 32,795 (48.8) | 32,795 (48.8) |
| | | Tertiary | 18,000 (26.8) | 18,000 (26.8) |
| | | Relation to patient, n (%) | | |
| | | Partner | 67,225 (100.0) | 67,225 (100.0) |
| Young adults and adults (25-64 years) | Siblings | Number of persons, n | 10,598 (100.0) | 10,598 (100.0) |
| | | Male, n (%) | 5,990 (56.5) | 5,990 (56.5) |
| | | Age, median (Q1;Q3) | 50.9 (33.7;58.7) | 51.0 (33.5;58.8) |
| | | CCI, n (%) | | |
| | | 0 | 8,867 (83.7) | 9,057 (85.5) |
| | | 1-2 | 1,489 (14.0) | 1,308 (12.3) |
| | | +3 | 242 (2.3) | 233 (2.2) |
| | | Education level, n (%) | | |
| | | No/Unknown education | 31 (0.3) | 31 (0.3) |
| | | Primary | 2,859 (27.0) | 2,859 (27.0) |
| | | Secondary | 5,631 (53.1) | 5,631 (53.1) |
| | | Tertiary | 2,077 (19.6) | 2,077 (19.6) |
| | | Relation to patient, n (%) | | |
| | | Sibling | 10,598 (100.0) | 10,598 (100.0) |
| Older people (65+ years) | Children | Number of persons, n | 10,254 (100.0) | 10,254 (100.0) |
| | | Male, n (%) | 5,330 (52.0) | 5,330 (52.0) |
| | | Age, median (Q1;Q3) | 48.1 (42.1;54.7) | 47.9 (42.1;54.5) |
| | | CCI, n (%) | | |
| | | 0 | 8,904 (86.8) | 8,962 (87.4) |

#### Supplementary Table 279
#### Stress related disorders - Prevalent cohort 2021
#### Patient characteristics - relatives - not pooled

| *Age strata of index patients* | *Closest relative type* | *Characteristic of family relatives by agestrata of index patients* | *Closest family relatives of brain disorder patients* | *Comparison relative* |
|---|---|---|---|---|
| | | 1-2 | 1,185 (11.6) | 1,127 (11.0) |
| | | +3 | 165 (1.6) | 165 (1.6) |
| | | Education level, n (%) | | |
| | | No/Unknown education | 13 (0.1) | 13 (0.1) |
| | | Primary | 1,859 (18.1) | 1,859 (18.1) |
| | | Secondary | 4,671 (45.6) | 4,671 (45.6) |
| | | Tertiary | 3,711 (36.2) | 3,711 (36.2) |
| | | Relation to patient, n (%) | | |
| | | Child | 10,254 (100.0) | 10,254 (100.0) |
| Older people (65+ years) | Partners | Number of persons, n | 9,223 (100.0) | 9,223 (100.0) |
| | | Male, n (%) | 5,286 (57.3) | 5,286 (57.3) |
| | | Age, median (Q1;Q3) | 71.0 (66.7;75.6) | 71.0 (67.0;75.3) |
| | | CCI, n (%) | | |
| | | 0 | 5,454 (59.1) | 5,832 (63.2) |
| | | 1-2 | 2,783 (30.2) | 2,567 (27.8) |
| | | +3 | 986 (10.7) | 824 (8.9) |
| | | Education level, n (%) | | |
| | | No/Unknown education | 566 (6.1) | 566 (6.1) |
| | | Primary | 2,046 (22.2) | 2,046 (22.2) |
| | | Secondary | 4,541 (49.2) | 4,541 (49.2) |
| | | Tertiary | 2,070 (22.4) | 2,070 (22.4) |
| | | Relation to patient, n (%) | | |
| | | Partner | 9,223 (100.0) | 9,223 (100.0) |
| Older people (65+ years) | Siblings | Number of persons, n | 772 (100.0) | 772 (100.0) |
| | | Male, n (%) | 393 (50.9) | 393 (50.9) |
| | | Age, median (Q1;Q3) | 62.4 (59.5;64.9) | 62.5 (59.9;64.7) |
| | | CCI, n (%) | | |
| | | 0 | 561 (72.7) | 592 (76.7) |
| | | 1-2 | 181 (23.4) | 153 (19.8) |
| | | +3 | 30 (3.9) | 27 (3.5) |
| | | Education level, n (%) | | |
| | | No/Unknown education | 6 (0.8) | 6 (0.8) |
| | | Primary | 134 (17.4) | 134 (17.4) |
| | | Secondary | 489 (63.3) | 489 (63.3) |
| | | Tertiary | 143 (18.5) | 143 (18.5) |
| | | Relation to patient, n (%) | | |
| | | Sibling | 772 (100.0) | 772 (100.0) |

#### Supplementary Table 280
#### Stress related disorders - Prevalent cohort 2021
#### Cost analysis - pooled

| Age strata of index patients | Closest relative type | Number of index patients included in cost component calculations | Cost component | Actual costs: Total (million EUR) | Actual costs: Per person (EUR) | Attributable costs: Total (million EUR) | Attributable costs: Per person (EUR) |
|---|---|---|---|---|---|---|---|
| Children and young people (0-24 years) | Fathers | 30,691 | Primary sector | 7.6 | 247 | 0.7 | 22 |
| | | 30,691 | Hospital care | 52.8 | 1,724 | 7.7 | 253 |
| | | 30,691 | Filled prescriptions | 7.2 | 236 | 1.8 | 60 |
| | | 29,497 | Lost productivity (income loss) | . | . | 330.2 | 11,194 |
| Children and young people (0-24 years) | Mothers | 30,785 | Primary sector | 12.1 | 394 | 1.8 | 60 |
| | | 30,785 | Hospital care | 64.4 | 2,094 | 16.8 | 548 |
| | | 30,785 | Filled prescriptions | 8.5 | 277 | 2.6 | 85 |
| | | 30,543 | Lost productivity (income loss) | . | . | 268.3 | 8,785 |
| Young adults and adults (25-64 years) | Pooled | 140,902 | Primary sector | 42.9 | 306 | 3.9 | 28 |
| | | 140,902 | Hospital care | 278.9 | 1,987 | 34.0 | 242 |
| | | 140,902 | Filled prescriptions | 36.8 | 262 | 5.7 | 41 |
| | | 100,266 | Lost productivity (income loss) | . | . | 785.2 | 7,831 |
| Older people (65+ years) | Pooled | 20,249 | Primary sector | 8.1 | 401 | 0.6 | 29 |
| | | 20,249 | Hospital care | 61.9 | 3,079 | 6.9 | 341 |
| | | 20,249 | Filled prescriptions | 7.8 | 389 | 0.8 | 39 |
| | | 10,900 | Lost productivity (income loss) | . | . | 60.7 | 5,568 |

**Supplementary Table 281**
**Stress related disorders - Prevalent cohort 2021**
**Cost analysis - not pooled**

| *Age strata of index patients* | *Closest relative type* | *Number of index patients included in cost component calculations* | *Cost component* | *Actual costs* | | *Attributable costs* | |
|---|---|---|---|---|---|---|---|
| | | | | *Total (million EUR)* | *Per person (EUR)* | *Total (million EUR)* | *Per person (EUR)* |
| Children and young people (0-24 years) | Fathers | 30,691 | Primary sector | 7.6 | 247 | 0.7 | 22 |
| | | 30,691 | Hospital care | 52.8 | 1,724 | 7.7 | 253 |
| | | 30,691 | Filled prescriptions | 7.2 | 236 | 1.8 | 60 |
| | | 29,497 | Lost productivity (income loss) | . | . | 330.2 | 11,194 |
| Children and young people (0-24 years) | Mothers | 30,785 | Primary sector | 12.1 | 394 | 1.8 | 60 |
| | | 30,785 | Hospital care | 64.4 | 2,094 | 16.8 | 548 |
| | | 30,785 | Filled prescriptions | 8.5 | 277 | 2.6 | 85 |
| | | 30,543 | Lost productivity (income loss) | . | . | 268.3 | 8,785 |
| Young adults and adults (25-64 years) | Children | 32,743 | Primary sector | 6.8 | 210 | 0.7 | 21 |
| | | 32,743 | Hospital care | 36.5 | 1,117 | 6.4 | 195 |
| | | 32,743 | Filled prescriptions | 3.3 | 101 | 0.4 | 12 |
| | | 16,152 | Lost productivity (income loss) | . | . | 31.3 | 1,936 |
| Young adults and adults (25-64 years) | Parents | 30,336 | Primary sector | 13.9 | 464 | 1.1 | 37 |
| | | 30,336 | Hospital care | 105.4 | 3,509 | 11.2 | 373 |
| | | 30,336 | Filled prescriptions | 16.0 | 533 | 2.4 | 81 |
| | | 12,593 | Lost productivity (income loss) | . | . | 90.2 | 7,165 |
| Young adults and adults (25-64 years) | Partners | 67,225 | Primary sector | 18.9 | 282 | 1.9 | 29 |
| | | 67,225 | Hospital care | 115.4 | 1,720 | 15.1 | 225 |
| | | 67,225 | Filled prescriptions | 14.7 | 218 | 2.5 | 37 |
| | | 62,310 | Lost productivity (income loss) | . | . | 633.1 | 10,160 |
| Young adults and adults (25-64 years) | Siblings | 10,598 | Primary sector | 3.2 | 303 | 0.2 | 19 |
| | | 10,598 | Hospital care | 21.7 | 2,049 | 1.3 | 125 |
| | | 10,598 | Filled prescriptions | 2.9 | 270 | 0.4 | 38 |
| | | 9,211 | Lost productivity (income loss) | . | . | 30.7 | 3,330 |
| Older people (65+ years) | Children | 10,254 | Primary sector | 3.1 | 307 | 0.3 | 27 |
| | | 10,254 | Hospital care | 20.4 | 1,994 | 2.9 | 288 |
| | | 10,254 | Filled prescriptions | 2.4 | 232 | 0.3 | 28 |
| | | 9,717 | Lost productivity (income loss) | . | . | 57.6 | 5,931 |
| Older people (65+ years) | Partners | 9,223 | Primary sector | 4.6 | 506 | 0.3 | 32 |
| | | 9,223 | Hospital care | 39.1 | 4,294 | 3.6 | 395 |
| | | 9,223 | Filled prescriptions | 5.1 | 565 | 0.5 | 53 |
| | | 763 | Lost productivity (income loss) | . | . | 4.0 | 5,281 |
| Older people (65+ years) | Siblings | 772 | Primary sector | 0.3 | 411 | 0.0 | 38 |
| | | 772 | Hospital care | 2.4 | 3,139 | 0.3 | 411 |
| | | 772 | Filled prescriptions | 0.3 | 397 | 0.0 | 27 |
| | | 420 | Lost productivity (income loss) | . | . | -1.0 | -2,293 |

**Supplementary Table 282**
**Stroke - Incident cohort 2016-2021**
**Identification of populations of relatives**

| ***Agegroup of index patients in matched population*** | | ***Closest relative type identified*** | | ***Available control with same type of closest relative*** |
|---|---|---|---|---|
| Children and young people (0-24 years) | n = 696 | Fathers | n = 467 (67%) | 96% |
| | | Mothers | n = 500 (72%) | 95% |
| | | No available closest relative | n = 169 (24%) | n/a |
| Young adults and adults (25-64 years) | n = 23,203 | Children | n = 4,456 (19%) | 90% |
| | | Parents | n = 2,029 (9%) | 86% |
| | | Partners | n = 12,868 (55%) | 95% |
| | | Siblings | n = 2,996 (13%) | 60% |
| | | Pooled | n = 22,349 (96%) | 89% |
| | | No available closest relative | n = 854 (4%) | n/a |
| Older people (65+ years) | n = 58,980 | Children | n = 26,460 (45%) | 95% |
| | | Partners | n = 24,393 (41%) | 95% |
| | | Siblings | n = 805 (1%) | 33% |
| | | Pooled | n = 51,658 (88%) | 94% |
| | | No available closest relative | n = 7,322 (12%) | n/a |

### Supplementary Table 283
### Stroke - Incident cohort 2016-2021
### Patient characteristics - cases

| *Age strata of index patients* | *Characteristic* | |
|---|---|---|
| Children and young people (0-24 years) | Number of persons, n | 504 (100.0) |
| | Male, n (%) | 290 (57.5) |
| | Age, median (Q1;Q3) | 18.4 (10.0;22.1) |
| | CCI, n (%) | |
| | 0 | 424 (84.1) |
| | 1-2 | 70 (13.9) |
| | +3 | 10 (2.0) |
| | Education level, n (%) | |
| | No/Unknown education | 153 (30.4) |
| | Primary | 255 (50.6) |
| | Secondary | 90 (17.9) |
| | Tertiary | 6 (1.2) |
| Young adults and adults (25-64 years) | Number of persons, n | 19,963 (100.0) |
| | Male, n (%) | 12,170 (61.0) |
| | Age, median (Q1;Q3) | 56.6 (50.5;61.1) |
| | CCI, n (%) | |
| | 0 | 13,497 (67.6) |
| | 1-2 | 4,949 (24.8) |
| | +3 | 1,517 (7.6) |
| | Education level, n (%) | |
| | No/Unknown education | 651 (3.3) |
| | Primary | 5,345 (26.8) |
| | Secondary | 9,993 (50.1) |
| | Tertiary | 3,974 (19.9) |
| Older people (65+ years) | Number of persons, n | 48,874 (100.0) |
| | Male, n (%) | 25,165 (51.5) |
| | Age, median (Q1;Q3) | 77.4 (72.1;83.5) |
| | CCI, n (%) | |
| | 0 | 22,874 (46.8) |
| | 1-2 | 17,490 (35.8) |
| | +3 | 8,510 (17.4) |
| | Education level, n (%) | |
| | No/Unknown education | 2,931 (6.0) |
| | Primary | 17,140 (35.1) |
| | Secondary | 21,468 (43.9) |
| | Tertiary | 7,335 (15.0) |

**Supplementary Table 284**
**Stroke - Incident cohort 2016-2021**
**Patient characteristics - relatives - pooled**

| *Age strata of index patients* | *Closest relative type* | *Characteristic of family relatives by agestrata of index patients* | *Closest family relatives of brain disorder patients* | *Comparison relative* |
|---|---|---|---|---|
| Children and young people (0-24 years) | Fathers | Number of persons, n | 449 (100.0) | 449 (100.0) |
| | | Male, n (%) | 449 (100.0) | 449 (100.0) |
| | | Age, median (Q1;Q3) | 48.4 (42.2;53.7) | 48.7 (42.1;53.8) |
| | | CCI, n (%) | | |
| | | 0 | 400 (89.1) | 409 (91.1) |
| | | 1-2 | 39 (8.7) | * |
| | | +3 | 10 (2.2) | * |
| | | Education level, n (%) | | |
| | | No/Unknown education | 7 (1.6) | 7 (1.6) |
| | | Primary | 91 (20.3) | 91 (20.3) |
| | | Secondary | 233 (51.9) | 233 (51.9) |
| | | Tertiary | 118 (26.3) | 118 (26.3) |
| | | Relation to patient, n (%) | | |
| | | Father | 449 (100.0) | 449 (100.0) |
| Children and young people (0-24 years) | Mothers | Number of persons, n | 477 (100.0) | 477 (100.0) |
| | | Male, n (%) | 0 (0.0) | 0 (0.0) |
| | | Age, median (Q1;Q3) | 46.9 (39.9;51.4) | 47.6 (39.7;51.8) |
| | | CCI, n (%) | | |
| | | 0 | 408 (85.5) | 423 (88.7) |
| | | 1-2 | 64 (13.4) | 47 (9.9) |
| | | +3 | 5 (1.0) | 7 (1.5) |
| | | Education level, n (%) | | |
| | | No/Unknown education | 16 (3.4) | 16 (3.4) |
| | | Primary | 89 (18.7) | 89 (18.7) |
| | | Secondary | 220 (46.1) | 220 (46.1) |
| | | Tertiary | 152 (31.9) | 152 (31.9) |
| | | Relation to patient, n (%) | | |
| | | Mother | 477 (100.0) | 477 (100.0) |
| Young adults and adults (25-64 years) | Pooled | Number of persons, n | 19,963 (100.0) | 19,963 (100.0) |
| | | Male, n (%) | 8,316 (41.7) | 8,316 (41.7) |
| | | Age, median (Q1;Q3) | 54.0 (41.3;60.9) | 54.2 (41.4;60.8) |
| | | CCI, n (%) | | |
| | | 0 | 16,473 (82.5) | 16,769 (84.0) |
| | | 1-2 | 2,918 (14.6) | 2,698 (13.5) |
| | | +3 | 572 (2.9) | 496 (2.5) |
| | | Education level, n (%) | | |
| | | No/Unknown education | 666 (3.3) | 666 (3.3) |
| | | Primary | 5,539 (27.7) | 5,539 (27.7) |
| | | Secondary | 9,436 (47.3) | 9,436 (47.3) |
| | | Tertiary | 4,322 (21.7) | 4,322 (21.7) |
| | | Relation to patient, n (%) | | |
| | | Mother | 1,409 (7.1) | 1,409 (7.1) |
| | | Father | 343 (1.7) | 343 (1.7) |
| | | Partner | 12,352 (61.9) | 12,352 (61.9) |
| | | Child | 4,039 (20.2) | 4,039 (20.2) |

**Supplementary Table 284**
**Stroke - Incident cohort 2016-2021**
**Patient characteristics - relatives - pooled**

| *Age strata of index patients* | *Closest relative type* | *Characteristic of family relatives by agestrata of index patients* | *Closest family relatives of brain disorder patients* | *Comparison relative* |
|---|---|---|---|---|
| | | Sibling | 1,820 (9.1) | 1,820 (9.1) |
| Older people (65+ years) | Pooled | Number of persons, n | 48,874 (100.0) | 48,874 (100.0) |
| | | Male, n (%) | 21,027 (43.0) | 21,027 (43.0) |
| | | Age, median (Q1;Q3) | 63.0 (53.9;73.4) | 63.0 (53.8;73.3) |
| | | CCI, n (%) | | |
| | | 0 | 36,242 (74.2) | 36,867 (75.4) |
| | | 1-2 | 10,034 (20.5) | 9,547 (19.5) |
| | | +3 | 2,598 (5.3) | 2,460 (5.0) |
| | | Education level, n (%) | | |
| | | No/Unknown education | 1,284 (2.6) | 1,284 (2.6) |
| | | Primary | 10,998 (22.5) | 10,998 (22.5) |
| | | Secondary | 24,242 (49.6) | 24,242 (49.6) |
| | | Tertiary | 12,350 (25.3) | 12,350 (25.3) |
| | | Relation to patient, n (%) | | |
| | | Partner | 23,401 (47.9) | 23,401 (47.9) |
| | | Child | 25,204 (51.6) | 25,204 (51.6) |
| | | Sibling | 269 (0.6) | 269 (0.6) |

**Supplementary Table 285**
**Stroke - Incident cohort 2016-2021**
**Patient characteristics - relatives - not pooled**

| *Age strata of index patients* | *Closest relative type* | *Characteristic of family relatives by agestrata of index patients* | *Closest family relatives of brain disorder patients* | *Comparison relative* |
|---|---|---|---|---|
| Children and young people (0-24 years) | Fathers | Number of persons, n | 449 (100.0) | 449 (100.0) |
| | | Male, n (%) | 449 (100.0) | 449 (100.0) |
| | | Age, median (Q1;Q3) | 48.4 (42.2;53.7) | 48.7 (42.1;53.8) |
| | | CCI, n (%) | | |
| | | 0 | 400 (89.1) | 409 (91.1) |
| | | 1-2 | 39 (8.7) | * |
| | | +3 | 10 (2.2) | * |
| | | Education level, n (%) | | |
| | | No/Unknown education | 7 (1.6) | 7 (1.6) |
| | | Primary | 91 (20.3) | 91 (20.3) |
| | | Secondary | 233 (51.9) | 233 (51.9) |
| | | Tertiary | 118 (26.3) | 118 (26.3) |
| | | Relation to patient, n (%) | | |
| | | Father | 449 (100.0) | 449 (100.0) |
| Children and young people (0-24 years) | Mothers | Number of persons, n | 477 (100.0) | 477 (100.0) |
| | | Male, n (%) | 0 (0.0) | 0 (0.0) |
| | | Age, median (Q1;Q3) | 46.9 (39.9;51.4) | 47.6 (39.7;51.8) |
| | | CCI, n (%) | | |
| | | 0 | 408 (85.5) | 423 (88.7) |
| | | 1-2 | 64 (13.4) | 47 (9.9) |
| | | +3 | 5 (1.0) | 7 (1.5) |
| | | Education level, n (%) | | |
| | | No/Unknown education | 16 (3.4) | 16 (3.4) |
| | | Primary | 89 (18.7) | 89 (18.7) |
| | | Secondary | 220 (46.1) | 220 (46.1) |
| | | Tertiary | 152 (31.9) | 152 (31.9) |
| | | Relation to patient, n (%) | | |
| | | Mother | 477 (100.0) | 477 (100.0) |
| Young adults and adults (25-64 years) | Children | Number of persons, n | 4,039 (100.0) | 4,039 (100.0) |
| | | Male, n (%) | 2,187 (54.1) | 2,187 (54.1) |
| | | Age, median (Q1;Q3) | 28.3 (21.1;34.6) | 28.2 (20.6;34.5) |
| | | CCI, n (%) | | |
| | | 0 | 3,758 (93.0) | 3,783 (93.7) |
| | | 1-2 | 268 (6.6) | 242 (6.0) |
| | | +3 | 13 (0.3) | 14 (0.3) |
| | | Education level, n (%) | | |
| | | No/Unknown education | 317 (7.8) | 317 (7.8) |
| | | Primary | 1,568 (38.8) | 1,568 (38.8) |
| | | Secondary | 1,454 (36.0) | 1,454 (36.0) |
| | | Tertiary | 700 (17.3) | 700 (17.3) |
| | | Relation to patient, n (%) | | |
| | | Child | 4,039 (100.0) | 4,039 (100.0) |
| Young adults and adults (25-64 years) | Parents | Number of persons, n | 1,752 (100.0) | 1,752 (100.0) |
| | | Male, n (%) | 343 (19.6) | 343 (19.6) |
| | | Age, median (Q1;Q3) | 74.8 (68.1;80.4) | 74.9 (68.4;80.6) |

### Supplementary Table 285
### Stroke - Incident cohort 2016-2021
### Patient characteristics - relatives - not pooled

| *Age strata of index patients* | *Closest relative type* | *Characteristic of family relatives by agestrata of index patients* | *Closest family relatives of brain disorder patients* | *Comparison relative* |
|---|---|---|---|---|
| | | CCI, n (%) | | |
| | | 0 | 1,064 (60.7) | 1,082 (61.8) |
| | | 1-2 | 508 (29.0) | 502 (28.7) |
| | | +3 | 180 (10.3) | 168 (9.6) |
| | | Education level, n (%) | | |
| | | No/Unknown education | 20 (1.1) | 20 (1.1) |
| | | Primary | 827 (47.2) | 827 (47.2) |
| | | Secondary | 713 (40.7) | 713 (40.7) |
| | | Tertiary | 192 (11.0) | 192 (11.0) |
| | | Relation to patient, n (%) | | |
| | | Mother | 1,409 (80.4) | 1,409 (80.4) |
| | | Father | 343 (19.6) | 343 (19.6) |
| Young adults and adults (25-64 years) | Partners | Number of persons, n | 12,352 (100.0) | 12,352 (100.0) |
| | | Male, n (%) | 4,714 (38.2) | 4,714 (38.2) |
| | | Age, median (Q1;Q3) | 55.4 (49.1;60.6) | 55.6 (49.4;60.5) |
| | | CCI, n (%) | | |
| | | 0 | 10,211 (82.7) | 10,442 (84.5) |
| | | 1-2 | 1,827 (14.8) | 1,655 (13.4) |
| | | +3 | 314 (2.5) | 255 (2.1) |
| | | Education level, n (%) | | |
| | | No/Unknown education | 318 (2.6) | 318 (2.6) |
| | | Primary | 2,637 (21.3) | 2,637 (21.3) |
| | | Secondary | 6,270 (50.8) | 6,270 (50.8) |
| | | Tertiary | 3,127 (25.3) | 3,127 (25.3) |
| | | Relation to patient, n (%) | | |
| | | Partner | 12,352 (100.0) | 12,352 (100.0) |
| Young adults and adults (25-64 years) | Siblings | Number of persons, n | 1,820 (100.0) | 1,820 (100.0) |
| | | Male, n (%) | 1,072 (58.9) | 1,072 (58.9) |
| | | Age, median (Q1;Q3) | 57.4 (52.8;61.1) | 57.6 (53.1;61.0) |
| | | CCI, n (%) | | |
| | | 0 | 1,440 (79.1) | 1,462 (80.3) |
| | | 1-2 | 315 (17.3) | 299 (16.4) |
| | | +3 | 65 (3.6) | 59 (3.2) |
| | | Education level, n (%) | | |
| | | No/Unknown education | 11 (0.6) | 11 (0.6) |
| | | Primary | 507 (27.9) | 507 (27.9) |
| | | Secondary | 999 (54.9) | 999 (54.9) |
| | | Tertiary | 303 (16.6) | 303 (16.6) |
| | | Relation to patient, n (%) | | |
| | | Sibling | 1,820 (100.0) | 1,820 (100.0) |
| Older people (65+ years) | Children | Number of persons, n | 25,204 (100.0) | 25,204 (100.0) |
| | | Male, n (%) | 13,024 (51.7) | 13,024 (51.7) |
| | | Age, median (Q1;Q3) | 54.3 (48.3;59.4) | 54.2 (48.3;59.1) |
| | | CCI, n (%) | | |
| | | 0 | 21,321 (84.6) | 21,607 (85.7) |

### Supplementary Table 285
### Stroke - Incident cohort 2016-2021
### Patient characteristics - relatives - not pooled

| *Age strata of index patients* | *Closest relative type* | *Characteristic of family relatives by agestrata of index patients* | *Closest family relatives of brain disorder patients* | *Comparison relative* |
|---|---|---|---|---|
| | | 1-2 | 3,375 (13.4) | 3,113 (12.4) |
| | | +3 | 508 (2.0) | 484 (1.9) |
| | | Education level, n (%) | | |
| | | No/Unknown education | 127 (0.5) | 127 (0.5) |
| | | Primary | 4,142 (16.4) | 4,142 (16.4) |
| | | Secondary | 13,139 (52.1) | 13,139 (52.1) |
| | | Tertiary | 7,796 (30.9) | 7,796 (30.9) |
| | | Relation to patient, n (%) | | |
| | | Child | 25,204 (100.0) | 25,204 (100.0) |
| Older people (65+ years) | Partners | Number of persons, n | 23,401 (100.0) | 23,401 (100.0) |
| | | Male, n (%) | 7,856 (33.6) | 7,856 (33.6) |
| | | Age, median (Q1;Q3) | 73.7 (69.0;78.7) | 73.7 (69.1;78.6) |
| | | CCI, n (%) | | |
| | | 0 | 14,725 (62.9) | 15,048 (64.3) |
| | | 1-2 | 6,598 (28.2) | 6,385 (27.3) |
| | | +3 | 2,078 (8.9) | 1,968 (8.4) |
| | | Education level, n (%) | | |
| | | No/Unknown education | 1,155 (4.9) | 1,155 (4.9) |
| | | Primary | 6,798 (29.1) | 6,798 (29.1) |
| | | Secondary | 10,941 (46.8) | 10,941 (46.8) |
| | | Tertiary | 4,507 (19.3) | 4,507 (19.3) |
| | | Relation to patient, n (%) | | |
| | | Partner | 23,401 (100.0) | 23,401 (100.0) |
| Older people (65+ years) | Siblings | Number of persons, n | 269 (100.0) | 269 (100.0) |
| | | Male, n (%) | 147 (54.6) | 147 (54.6) |
| | | Age, median (Q1;Q3) | 61.7 (58.7;64.2) | 61.8 (58.9;64.2) |
| | | CCI, n (%) | | |
| | | 0 | 196 (72.9) | 212 (78.8) |
| | | 1-2 | 61 (22.7) | 49 (18.2) |
| | | +3 | 12 (4.5) | 8 (3.0) |
| | | Education level, n (%) | | |
| | | No/Unknown education | * | * |
| | | Primary | 58 (21.6) | 58 (21.6) |
| | | Secondary | 162 (60.2) | 162 (60.2) |
| | | Tertiary | * | * |
| | | Relation to patient, n (%) | | |
| | | Sibling | 269 (100.0) | 269 (100.0) |

### Supplementary Table 286
### Stroke - Incident cohort 2016-2021
### Cost analysis - pooled

| *Age strata of index patients* | *Closest relative type* | *Number of index patients included in cost component calculations* | *Cost component* | *Actual costs* | | *Attributable costs* | |
|---|---|---|---|---|---|---|---|
| | | | | *Total (million EUR)* | *Per person (EUR)* | *Total (million EUR)* | *Per person (EUR)* |
| Children and young people (0-24 years) | Fathers | 449 | Primary sector | 0.1 | 234 | 0.0 | 18 |
| | | 449 | Hospital care | 0.8 | 1,757 | -0.0 | -14 |
| | | 449 | Filled prescriptions | 0.1 | 194 | -0.0 | -23 |
| | | 432 | Lost productivity (income loss) | . | . | 1.8 | 4,279 |
| Children and young people (0-24 years) | Mothers | 477 | Primary sector | 0.2 | 348 | 0.0 | 10 |
| | | 477 | Hospital care | 1.0 | 1,992 | 0.2 | 503 |
| | | 477 | Filled prescriptions | 0.1 | 204 | -0.0 | -0 |
| | | 471 | Lost productivity (income loss) | . | . | -0.2 | -523 |
| Young adults and adults (25-64 years) | Pooled | 19,963 | Primary sector | 6.5 | 328 | 0.3 | 16 |
| | | 19,963 | Hospital care | 46.5 | 2,341 | 3.7 | 184 |
| | | 19,963 | Filled prescriptions | 5.6 | 282 | 0.4 | 20 |
| | | 15,211 | Lost productivity (income loss) | . | . | 55.4 | 3,643 |
| Older people (65+ years) | Pooled | 48,874 | Primary sector | 19.5 | 402 | 0.4 | 9 |
| | | 48,874 | Hospital care | 142.2 | 2,933 | 9.8 | 201 |
| | | 48,874 | Filled prescriptions | 18.6 | 383 | 1.0 | 21 |
| | | 23,376 | Lost productivity (income loss) | . | . | 51.4 | 2,201 |

### Supplementary Table 287
### Stroke - Incident cohort 2016-2021
### Cost analysis - not pooled

| Age strata of index patients | Closest relative type | Number of index patients included in cost component calculations | Cost component | Actual costs: Total (million EUR) | Actual costs: Per person (EUR) | Attributable costs: Total (million EUR) | Attributable costs: Per person (EUR) |
|---|---|---|---|---|---|---|---|
| Children and young people (0-24 years) | Fathers | 449 | Primary sector | 0.1 | 234 | 0.0 | 18 |
| | | 449 | Hospital care | 0.8 | 1,757 | -0.0 | -14 |
| | | 449 | Filled prescriptions | 0.1 | 194 | -0.0 | -23 |
| | | 432 | Lost productivity (income loss) | . | . | 1.8 | 4,279 |
| Children and young people (0-24 years) | Mothers | 477 | Primary sector | 0.2 | 348 | 0.0 | 10 |
| | | 477 | Hospital care | 1.0 | 1,992 | 0.2 | 503 |
| | | 477 | Filled prescriptions | 0.1 | 204 | -0.0 | -0 |
| | | 471 | Lost productivity (income loss) | . | . | -0.2 | -523 |
| Young adults and adults (25-64 years) | Children | 4,039 | Primary sector | 1.0 | 236 | 0.1 | 17 |
| | | 4,039 | Hospital care | 7.0 | 1,736 | 0.8 | 195 |
| | | 4,039 | Filled prescriptions | 0.5 | 129 | 0.0 | 6 |
| | | 3,296 | Lost productivity (income loss) | . | . | 6.1 | 1,862 |
| Young adults and adults (25-64 years) | Parents | 1,752 | Primary sector | 0.9 | 500 | 0.0 | 4 |
| | | 1,752 | Hospital care | 7.9 | 4,568 | 0.4 | 237 |
| | | 1,752 | Filled prescriptions | 1.1 | 628 | 0.1 | 72 |
| | | 231 | Lost productivity (income loss) | . | . | 0.2 | 795 |
| Young adults and adults (25-64 years) | Partners | 12,352 | Primary sector | 4.1 | 336 | 0.2 | 19 |
| | | 12,352 | Hospital care | 26.9 | 2,182 | 2.9 | 233 |
| | | 12,352 | Filled prescriptions | 3.4 | 279 | 0.3 | 25 |
| | | 10,217 | Lost productivity (income loss) | . | . | 44.5 | 4,360 |
| Young adults and adults (25-64 years) | Siblings | 1,820 | Primary sector | 0.6 | 313 | 0.0 | 6 |
| | | 1,820 | Hospital care | 4.8 | 2,653 | -0.4 | -220 |
| | | 1,820 | Filled prescriptions | 0.6 | 309 | -0.1 | -29 |
| | | 1,467 | Lost productivity (income loss) | . | . | 4.5 | 3,099 |
| Older people (65+ years) | Children | 25,204 | Primary sector | 7.8 | 309 | 0.3 | 11 |
| | | 25,204 | Hospital care | 50.4 | 2,003 | 4.1 | 161 |
| | | 25,204 | Filled prescriptions | 6.4 | 256 | 0.6 | 25 |
| | | 22,110 | Lost productivity (income loss) | . | . | 50.1 | 2,267 |
| Older people (65+ years) | Partners | 23,401 | Primary sector | 11.6 | 505 | 0.2 | 7 |
| | | 23,401 | Hospital care | 91.3 | 3,958 | 5.7 | 246 |
| | | 23,401 | Filled prescriptions | 12.0 | 521 | 0.4 | 17 |
| | | 1,107 | Lost productivity (income loss) | . | . | -1.2 | -1,106 |
| Older people (65+ years) | Siblings | 269 | Primary sector | 0.1 | 341 | 0.0 | 8 |
| | | 269 | Hospital care | 0.5 | 1,918 | 0.0 | 83 |
| | | 269 | Filled prescriptions | 0.1 | 381 | 0.0 | 58 |
| | | 159 | Lost productivity (income loss) | . | . | 2.6 | 16,046 |

**Supplementary Table 288**
**Stroke - Prevalent cohort 2021**
**Identification of populations of relatives**

| ***Agegroup of index patients in matched population*** | | ***Closest relative type identified*** | | ***Available control with same type of closest relative*** |
|---|---|---|---|---|
| Children and young people (0-24 years) | n = 1,084 | Fathers | n = 937 (86%) | 96% |
| | | Mothers | n = 966 (89%) | 97% |
| | | No available closest relative | n = 48 (4%) | n/a |
| Young adults and adults (25-64 years) | n = 36,931 | Children | n = 6,899 (19%) | 91% |
| | | Parents | n = 3,575 (10%) | 85% |
| | | Partners | n = 20,708 (56%) | 95% |
| | | Siblings | n = 4,774 (13%) | 61% |
| | | Pooled | n = 35,956 (97%) | 89% |
| | | No available closest relative | n = 975 (3%) | n/a |
| Older people (65+ years) | n = 89,392 | Children | n = 37,213 (42%) | 95% |
| | | Partners | n = 41,302 (46%) | 96% |
| | | Siblings | n = 1,795 (2%) | 44% |
| | | Pooled | n = 80,310 (90%) | 94% |
| | | No available closest relative | n = 9,082 (10%) | n/a |

**Supplementary Table 289**
**Stroke - Prevalent cohort 2021**
**Patient characteristics - cases**

| *Age strata of index patients* | *Characteristic* | |
|---|---|---|
| Children and young people (0-24 years) | Number of persons, n | 1,004 (100.0) |
| | Male, n (%) | 579 (57.7) |
| | Age, median (Q1;Q3) | 15.9 (9.4;20.9) |
| | CCI, n (%) | |
| | 0 | 250 (24.9) |
| | 1-2 | 643 (64.0) |
| | +3 | 111 (11.1) |
| | Education level, n (%) | |
| | No/Unknown education | 400 (39.8) |
| | Primary | 471 (46.9) |
| | Secondary | 118 (11.8) |
| | Tertiary | 15 (1.5) |
| Young adults and adults (25-64 years) | Number of persons, n | 32,204 (100.0) |
| | Male, n (%) | 17,773 (55.2) |
| | Age, median (Q1;Q3) | 57.0 (50.8;61.4) |
| | CCI, n (%) | |
| | 0 | 6,530 (20.3) |
| | 1-2 | 20,568 (63.9) |
| | +3 | 5,106 (15.9) |
| | Education level, n (%) | |
| | No/Unknown education | 891 (2.8) |
| | Primary | 8,620 (26.8) |
| | Secondary | 15,887 (49.3) |
| | Tertiary | 6,806 (21.1) |
| Older people (65+ years) | Number of persons, n | 75,968 (100.0) |
| | Male, n (%) | 40,776 (53.7) |
| | Age, median (Q1;Q3) | 77.1 (72.1;82.9) |
| | CCI, n (%) | |
| | 0 | 9,094 (12.0) |
| | 1-2 | 43,545 (57.3) |
| | +3 | 23,329 (30.7) |
| | Education level, n (%) | |
| | No/Unknown education | 4,338 (5.7) |
| | Primary | 25,147 (33.1) |
| | Secondary | 34,535 (45.5) |
| | Tertiary | 11,948 (15.7) |

**Supplementary Table 290**
**Stroke - Prevalent cohort 2021**
**Patient characteristics - relatives - pooled**

| *Age strata of index patients* | *Closest relative type* | *Characteristic of family relatives by agestrata of index patients* | *Closest family relatives of brain disorder patients* | *Comparison relative* |
|---|---|---|---|---|
| Children and young people (0-24 years) | Fathers | Number of persons, n | 907 (100.0) | 907 (100.0) |
| | | Male, n (%) | 907 (100.0) | 907 (100.0) |
| | | Age, median (Q1;Q3) | 47.9 (41.4;53.4) | 47.8 (41.7;53.1) |
| | | CCI, n (%) | | |
| | | 0 | 814 (89.7) | 829 (91.4) |
| | | 1-2 | 79 (8.7) | 72 (7.9) |
| | | +3 | 14 (1.5) | 6 (0.7) |
| | | Education level, n (%) | | |
| | | No/Unknown education | 19 (2.1) | 19 (2.1) |
| | | Primary | 162 (17.9) | 162 (17.9) |
| | | Secondary | 477 (52.6) | 477 (52.6) |
| | | Tertiary | 249 (27.5) | 249 (27.5) |
| | | Relation to patient, n (%) | | |
| | | Father | 907 (100.0) | 907 (100.0) |
| Children and young people (0-24 years) | Mothers | Number of persons, n | 938 (100.0) | 938 (100.0) |
| | | Male, n (%) | 0 (0.0) | 0 (0.0) |
| | | Age, median (Q1;Q3) | 45.5 (38.8;50.9) | 45.9 (39.2;51.3) |
| | | CCI, n (%) | | |
| | | 0 | 833 (88.8) | 834 (88.9) |
| | | 1-2 | 98 (10.4) | 98 (10.4) |
| | | +3 | 7 (0.7) | 6 (0.6) |
| | | Education level, n (%) | | |
| | | No/Unknown education | 21 (2.2) | 21 (2.2) |
| | | Primary | 155 (16.5) | 155 (16.5) |
| | | Secondary | 386 (41.2) | 386 (41.2) |
| | | Tertiary | 376 (40.1) | 376 (40.1) |
| | | Relation to patient, n (%) | | |
| | | Mother | 938 (100.0) | 938 (100.0) |
| Young adults and adults (25-64 years) | Pooled | Number of persons, n | 32,204 (100.0) | 32,204 (100.0) |
| | | Male, n (%) | 14,605 (45.4) | 14,605 (45.4) |
| | | Age, median (Q1;Q3) | 55.0 (41.7;61.8) | 55.2 (41.7;61.6) |
| | | CCI, n (%) | | |
| | | 0 | 26,302 (81.7) | 26,885 (83.5) |
| | | 1-2 | 4,908 (15.2) | 4,524 (14.0) |
| | | +3 | 994 (3.1) | 795 (2.5) |
| | | Education level, n (%) | | |
| | | No/Unknown education | 925 (2.9) | 925 (2.9) |
| | | Primary | 8,691 (27.0) | 8,691 (27.0) |
| | | Secondary | 15,648 (48.6) | 15,648 (48.6) |
| | | Tertiary | 6,940 (21.6) | 6,940 (21.6) |
| | | Relation to patient, n (%) | | |
| | | Mother | 2,488 (7.7) | 2,488 (7.7) |
| | | Father | 580 (1.8) | 580 (1.8) |
| | | Partner | 19,839 (61.6) | 19,839 (61.6) |
| | | Child | 6,338 (19.7) | 6,338 (19.7) |

**Supplementary Table 290**
**Stroke - Prevalent cohort 2021**
**Patient characteristics - relatives - pooled**

| *Age strata of index patients* | *Closest relative type* | *Characteristic of family relatives by agestrata of index patients* | *Closest family relatives of brain disorder patients* | *Comparison relative* |
|---|---|---|---|---|
| | | Sibling | 2,959 (9.2) | 2,959 (9.2) |
| Older people (65+ years) | Pooled | Number of persons, n | 75,968 (100.0) | 75,968 (100.0) |
| | | Male, n (%) | 31,504 (41.5) | 31,504 (41.5) |
| | | Age, median (Q1;Q3) | 65.0 (55.1;74.5) | 65.1 (55.0;74.5) |
| | | CCI, n (%) | | |
| | | 0 | 55,362 (72.9) | 56,407 (74.3) |
| | | 1-2 | 16,401 (21.6) | 15,784 (20.8) |
| | | +3 | 4,205 (5.5) | 3,777 (5.0) |
| | | Education level, n (%) | | |
| | | No/Unknown education | 1,939 (2.6) | 1,939 (2.6) |
| | | Primary | 17,178 (22.6) | 17,178 (22.6) |
| | | Secondary | 37,709 (49.6) | 37,709 (49.6) |
| | | Tertiary | 19,142 (25.2) | 19,142 (25.2) |
| | | Relation to patient, n (%) | | |
| | | Partner | 39,706 (52.3) | 39,706 (52.3) |
| | | Child | 35,459 (46.7) | 35,459 (46.7) |
| | | Sibling | 803 (1.1) | 803 (1.1) |

#### Supplementary Table 291
#### Stroke - Prevalent cohort 2021
#### Patient characteristics - relatives - not pooled

| *Age strata of index patients* | *Closest relative type* | *Characteristic of family relatives by agestrata of index patients* | *Closest family relatives of brain disorder patients* | *Comparison relative* |
|---|---|---|---|---|
| Children and young people (0-24 years) | Fathers | Number of persons, n | 907 (100.0) | 907 (100.0) |
| | | Male, n (%) | 907 (100.0) | 907 (100.0) |
| | | Age, median (Q1;Q3) | 47.9 (41.4;53.4) | 47.8 (41.7;53.1) |
| | | CCI, n (%) | | |
| | | 0 | 814 (89.7) | 829 (91.4) |
| | | 1-2 | 79 (8.7) | 72 (7.9) |
| | | +3 | 14 (1.5) | 6 (0.7) |
| | | Education level, n (%) | | |
| | | No/Unknown education | 19 (2.1) | 19 (2.1) |
| | | Primary | 162 (17.9) | 162 (17.9) |
| | | Secondary | 477 (52.6) | 477 (52.6) |
| | | Tertiary | 249 (27.5) | 249 (27.5) |
| | | Relation to patient, n (%) | | |
| | | Father | 907 (100.0) | 907 (100.0) |
| Children and young people (0-24 years) | Mothers | Number of persons, n | 938 (100.0) | 938 (100.0) |
| | | Male, n (%) | 0 (0.0) | 0 (0.0) |
| | | Age, median (Q1;Q3) | 45.5 (38.8;50.9) | 45.9 (39.2;51.3) |
| | | CCI, n (%) | | |
| | | 0 | 833 (88.8) | 834 (88.9) |
| | | 1-2 | 98 (10.4) | 98 (10.4) |
| | | +3 | 7 (0.7) | 6 (0.6) |
| | | Education level, n (%) | | |
| | | No/Unknown education | 21 (2.2) | 21 (2.2) |
| | | Primary | 155 (16.5) | 155 (16.5) |
| | | Secondary | 386 (41.2) | 386 (41.2) |
| | | Tertiary | 376 (40.1) | 376 (40.1) |
| | | Relation to patient, n (%) | | |
| | | Mother | 938 (100.0) | 938 (100.0) |
| Young adults and adults (25-64 years) | Children | Number of persons, n | 6,338 (100.0) | 6,338 (100.0) |
| | | Male, n (%) | 3,443 (54.3) | 3,443 (54.3) |
| | | Age, median (Q1;Q3) | 28.8 (21.2;35.1) | 28.5 (21.1;34.7) |
| | | CCI, n (%) | | |
| | | 0 | 5,901 (93.1) | 5,949 (93.9) |
| | | 1-2 | 416 (6.6) | 372 (5.9) |
| | | +3 | 21 (0.3) | 17 (0.3) |
| | | Education level, n (%) | | |
| | | No/Unknown education | 433 (6.8) | 433 (6.8) |
| | | Primary | 2,558 (40.4) | 2,558 (40.4) |
| | | Secondary | 2,280 (36.0) | 2,280 (36.0) |
| | | Tertiary | 1,067 (16.8) | 1,067 (16.8) |
| | | Relation to patient, n (%) | | |
| | | Child | 6,338 (100.0) | 6,338 (100.0) |
| Young adults and adults (25-64 years) | Parents | Number of persons, n | 3,068 (100.0) | 3,068 (100.0) |
| | | Male, n (%) | 580 (18.9) | 580 (18.9) |
| | | Age, median (Q1;Q3) | 75.5 (68.5;81.2) | 75.8 (68.9;81.1) |

**Supplementary Table 291**
**Stroke - Prevalent cohort 2021**
**Patient characteristics - relatives - not pooled**

| *Age strata of index patients* | *Closest relative type* | *Characteristic of family relatives by agestrata of index patients* | *Closest family relatives of brain disorder patients* | *Comparison relative* |
|---|---|---|---|---|
| | | CCI, n (%) | | |
| | | 0 | 1,836 (59.8) | 1,882 (61.3) |
| | | 1-2 | 907 (29.6) | 912 (29.7) |
| | | +3 | 325 (10.6) | 274 (8.9) |
| | | Education level, n (%) | | |
| | | No/Unknown education | 43 (1.4) | 43 (1.4) |
| | | Primary | 1,327 (43.3) | 1,327 (43.3) |
| | | Secondary | 1,341 (43.7) | 1,341 (43.7) |
| | | Tertiary | 357 (11.6) | 357 (11.6) |
| | | Relation to patient, n (%) | | |
| | | Mother | 2,488 (81.1) | 2,488 (81.1) |
| | | Father | 580 (18.9) | 580 (18.9) |
| Young adults and adults (25-64 years) | Partners | Number of persons, n | 19,839 (100.0) | 19,839 (100.0) |
| | | Male, n (%) | 8,875 (44.7) | 8,875 (44.7) |
| | | Age, median (Q1;Q3) | 56.2 (49.8;61.2) | 56.3 (50.0;61.0) |
| | | CCI, n (%) | | |
| | | 0 | 16,268 (82.0) | 16,688 (84.1) |
| | | 1-2 | 3,038 (15.3) | 2,748 (13.9) |
| | | +3 | 533 (2.7) | 403 (2.0) |
| | | Education level, n (%) | | |
| | | No/Unknown education | 434 (2.2) | 434 (2.2) |
| | | Primary | 4,066 (20.5) | 4,066 (20.5) |
| | | Secondary | 10,288 (51.9) | 10,288 (51.9) |
| | | Tertiary | 5,051 (25.5) | 5,051 (25.5) |
| | | Relation to patient, n (%) | | |
| | | Partner | 19,839 (100.0) | 19,839 (100.0) |
| Young adults and adults (25-64 years) | Siblings | Number of persons, n | 2,959 (100.0) | 2,959 (100.0) |
| | | Male, n (%) | 1,707 (57.7) | 1,707 (57.7) |
| | | Age, median (Q1;Q3) | 58.6 (54.2;62.6) | 58.7 (54.3;62.3) |
| | | CCI, n (%) | | |
| | | 0 | 2,297 (77.6) | 2,366 (80.0) |
| | | 1-2 | 547 (18.5) | 492 (16.6) |
| | | +3 | 115 (3.9) | 101 (3.4) |
| | | Education level, n (%) | | |
| | | No/Unknown education | 15 (0.5) | 15 (0.5) |
| | | Primary | 740 (25.0) | 740 (25.0) |
| | | Secondary | 1,739 (58.8) | 1,739 (58.8) |
| | | Tertiary | 465 (15.7) | 465 (15.7) |
| | | Relation to patient, n (%) | | |
| | | Sibling | 2,959 (100.0) | 2,959 (100.0) |
| Older people (65+ years) | Children | Number of persons, n | 35,459 (100.0) | 35,459 (100.0) |
| | | Male, n (%) | 17,965 (50.7) | 17,965 (50.7) |
| | | Age, median (Q1;Q3) | 54.5 (48.2;59.8) | 54.4 (48.2;59.4) |
| | | CCI, n (%) | | |
| | | 0 | 29,868 (84.2) | 30,310 (85.5) |

**Supplementary Table 291**
**Stroke - Prevalent cohort 2021**
**Patient characteristics - relatives - not pooled**

| *Age strata of index patients* | *Closest relative type* | *Characteristic of family relatives by agestrata of index patients* | *Closest family relatives of brain disorder patients* | *Comparison relative* |
|---|---|---|---|---|
| | | 1-2 | 4,866 (13.7) | 4,513 (12.7) |
| | | +3 | 725 (2.0) | 636 (1.8) |
| | | Education level, n (%) | | |
| | | No/Unknown education | 106 (0.3) | 106 (0.3) |
| | | Primary | 5,996 (16.9) | 5,996 (16.9) |
| | | Secondary | 18,187 (51.3) | 18,187 (51.3) |
| | | Tertiary | 11,170 (31.5) | 11,170 (31.5) |
| | | Relation to patient, n (%) | | |
| | | Child | 35,459 (100.0) | 35,459 (100.0) |
| Older people (65+ years) | Partners | Number of persons, n | 39,706 (100.0) | 39,706 (100.0) |
| | | Male, n (%) | 13,115 (33.0) | 13,115 (33.0) |
| | | Age, median (Q1;Q3) | 74.1 (69.2;78.8) | 74.2 (69.3;78.6) |
| | | CCI, n (%) | | |
| | | 0 | 24,895 (62.7) | 25,481 (64.2) |
| | | 1-2 | 11,364 (28.6) | 11,126 (28.0) |
| | | +3 | 3,447 (8.7) | 3,099 (7.8) |
| | | Education level, n (%) | | |
| | | No/Unknown education | 1,827 (4.6) | 1,827 (4.6) |
| | | Primary | 11,012 (27.7) | 11,012 (27.7) |
| | | Secondary | 19,044 (48.0) | 19,044 (48.0) |
| | | Tertiary | 7,823 (19.7) | 7,823 (19.7) |
| | | Relation to patient, n (%) | | |
| | | Partner | 39,706 (100.0) | 39,706 (100.0) |
| Older people (65+ years) | Siblings | Number of persons, n | 803 (100.0) | 803 (100.0) |
| | | Male, n (%) | 424 (52.8) | 424 (52.8) |
| | | Age, median (Q1;Q3) | 62.4 (59.1;64.8) | 62.6 (59.5;64.8) |
| | | CCI, n (%) | | |
| | | 0 | 599 (74.6) | 616 (76.7) |
| | | 1-2 | 171 (21.3) | 145 (18.1) |
| | | +3 | 33 (4.1) | 42 (5.2) |
| | | Education level, n (%) | | |
| | | No/Unknown education | 6 (0.7) | 6 (0.7) |
| | | Primary | 170 (21.2) | 170 (21.2) |
| | | Secondary | 478 (59.5) | 478 (59.5) |
| | | Tertiary | 149 (18.6) | 149 (18.6) |
| | | Relation to patient, n (%) | | |
| | | Sibling | 803 (100.0) | 803 (100.0) |

### Supplementary Table 292
#### Stroke - Prevalent cohort 2021
#### Cost analysis - pooled

| Age strata of index patients | Closest relative type | Number of index patients included in cost component calculations | Cost component | Actual costs: Total (million EUR) | Actual costs: Per person (EUR) | Attributable costs: Total (million EUR) | Attributable costs: Per person (EUR) |
|---|---|---|---|---|---|---|---|
| Children and young people (0-24 years) | Fathers | 907 | Primary sector | 0.2 | 226 | 0.0 | 21 |
| | | 907 | Hospital care | 1.1 | 1,234 | 0.3 | 282 |
| | | 907 | Filled prescriptions | 0.2 | 177 | 0.0 | 17 |
| | | 893 | Lost productivity (income loss) | . | . | -2.0 | -2,195 |
| Children and young people (0-24 years) | Mothers | 938 | Primary sector | 0.3 | 319 | -0.0 | -16 |
| | | 938 | Hospital care | 1.8 | 1,961 | 0.3 | 295 |
| | | 938 | Filled prescriptions | 0.2 | 203 | 0.0 | 37 |
| | | 932 | Lost productivity (income loss) | . | . | -0.0 | -12 |
| Young adults and adults (25-64 years) | Pooled | 32,204 | Primary sector | 10.8 | 335 | 0.6 | 18 |
| | | 32,204 | Hospital care | 76.5 | 2,387 | 11.6 | 362 |
| | | 32,204 | Filled prescriptions | 9.9 | 308 | 1.3 | 41 |
| | | 24,255 | Lost productivity (income loss) | . | . | 125.2 | 5,162 |
| Older people (65+ years) | Pooled | 75,968 | Primary sector | 32.0 | 425 | 1.2 | 16 |
| | | 75,968 | Hospital care | 234.0 | 3,105 | 20.0 | 265 |
| | | 75,968 | Filled prescriptions | 31.2 | 415 | 3.0 | 40 |
| | | 33,280 | Lost productivity (income loss) | . | . | 86.6 | 2,603 |

### Supplementary Table 293
### Stroke - Prevalent cohort 2021
### Cost analysis - not pooled

| Age strata of index patients | Closest relative type | Number of index patients included in cost component calculations | Cost component | Actual costs: Total (million EUR) | Actual costs: Per person (EUR) | Attributable costs: Total (million EUR) | Attributable costs: Per person (EUR) |
|---|---|---|---|---|---|---|---|
| Children and young people (0-24 years) | Fathers | 907 | Primary sector | 0.2 | 226 | 0.0 | 21 |
| | | 907 | Hospital care | 1.1 | 1,234 | 0.3 | 282 |
| | | 907 | Filled prescriptions | 0.2 | 177 | 0.0 | 17 |
| | | 893 | Lost productivity (income loss) | . | . | -2.0 | -2,195 |
| Children and young people (0-24 years) | Mothers | 938 | Primary sector | 0.3 | 319 | -0.0 | -16 |
| | | 938 | Hospital care | 1.8 | 1,961 | 0.3 | 295 |
| | | 938 | Filled prescriptions | 0.2 | 203 | 0.0 | 37 |
| | | 932 | Lost productivity (income loss) | . | . | -0.0 | -12 |
| Young adults and adults (25-64 years) | Children | 6,338 | Primary sector | 1.6 | 250 | 0.1 | 18 |
| | | 6,338 | Hospital care | 9.8 | 1,556 | 0.5 | 74 |
| | | 6,338 | Filled prescriptions | 0.9 | 144 | -0.0 | -1 |
| | | 5,134 | Lost productivity (income loss) | . | . | 9.1 | 1,774 |
| Young adults and adults (25-64 years) | Parents | 3,068 | Primary sector | 1.6 | 525 | 0.1 | 31 |
| | | 3,068 | Hospital care | 14.2 | 4,735 | 2.4 | 796 |
| | | 3,068 | Filled prescriptions | 2.0 | 651 | 0.3 | 91 |
| | | 402 | Lost productivity (income loss) | . | . | -2.5 | -6,127 |
| Young adults and adults (25-64 years) | Partners | 19,839 | Primary sector | 6.6 | 333 | 0.3 | 16 |
| | | 19,839 | Hospital care | 43.9 | 2,217 | 7.2 | 363 |
| | | 19,839 | Filled prescriptions | 5.9 | 297 | 0.9 | 45 |
| | | 16,448 | Lost productivity (income loss) | . | . | 102.6 | 6,236 |
| Young adults and adults (25-64 years) | Siblings | 2,959 | Primary sector | 1.0 | 340 | 0.1 | 18 |
| | | 2,959 | Hospital care | 8.6 | 2,923 | 1.6 | 535 |
| | | 2,959 | Filled prescriptions | 1.1 | 382 | 0.2 | 58 |
| | | 2,271 | Lost productivity (income loss) | . | . | 16.0 | 7,039 |
| Older people (65+ years) | Children | 35,459 | Primary sector | 11.4 | 323 | 0.7 | 19 |
| | | 35,459 | Hospital care | 72.8 | 2,059 | 6.5 | 183 |
| | | 35,459 | Filled prescriptions | 9.8 | 277 | 1.5 | 41 |
| | | 30,872 | Lost productivity (income loss) | . | . | 87.4 | 2,831 |
| Older people (65+ years) | Partners | 39,706 | Primary sector | 20.3 | 518 | 0.5 | 14 |
| | | 39,706 | Hospital care | 158.9 | 4,057 | 13.1 | 335 |
| | | 39,706 | Filled prescriptions | 21.1 | 539 | 1.5 | 39 |
| | | 1,972 | Lost productivity (income loss) | . | . | -2.2 | -1,119 |
| Older people (65+ years) | Siblings | 803 | Primary sector | 0.3 | 372 | 0.0 | 7 |
| | | 803 | Hospital care | 2.2 | 2,757 | 0.4 | 488 |
| | | 803 | Filled prescriptions | 0.3 | 388 | 0.0 | 36 |
| | | 436 | Lost productivity (income loss) | . | . | 1.4 | 3,302 |

**Supplementary Table 294**
**Traumatic brain injury - Incident cohort 2016-2021**
**Identification of populations of relatives**

| ***Agegroup of index patients in matched population*** | | ***Closest relative type identified*** | | ***Available control with same type of closest relative*** |
|---|---|---|---|---|
| Children and young people (0-24 years) | n = 28,031 | Fathers | n = 21,823 (78%) | 96% |
| | | Mothers | n = 23,152 (83%) | 96% |
| | | No available closest relative | n = 2,612 (9%) | n/a |
| Young adults and adults (25-64 years) | n = 24,110 | Children | n = 3,506 (15%) | 93% |
| | | Parents | n = 4,223 (18%) | 91% |
| | | Partners | n = 12,072 (50%) | 95% |
| | | Siblings | n = 3,199 (13%) | 47% |
| | | Pooled | n = 23,000 (95%) | 88% |
| | | No available closest relative | n = 1,110 (5%) | n/a |
| Older people (65+ years) | n = 22,986 | Children | n = 10,463 (46%) | 94% |
| | | Partners | n = 8,847 (38%) | 95% |
| | | Siblings | n = 374 (2%) | 29% |
| | | Pooled | n = 19,684 (86%) | 94% |
| | | No available closest relative | n = 3,302 (14%) | n/a |

**Supplementary Table 295**
**Traumatic brain injury - Incident cohort 2016-2021**
**Patient characteristics - cases**

| *Age strata of index patients* | *Characteristic* | |
|---|---|---|
| Children and young people (0-24 years) | Number of persons, n | 24,615 (100.0) |
| | Male, n (%) | 13,498 (54.8) |
| | Age, median (Q1;Q3) | 12.8 (4.8;19.1) |
| | CCI, n (%) | |
| | 0 | 23,027 (93.5) |
| | 1-2 | 1,551 (6.3) |
| | +3 | 37 (0.2) |
| | Education level, n (%) | |
| | No/Unknown education | 12,540 (50.9) |
| | Primary | 9,102 (37.0) |
| | Secondary | 2,737 (11.1) |
| | Tertiary | 236 (1.0) |
| Young adults and adults (25-64 years) | Number of persons, n | 20,249 (100.0) |
| | Male, n (%) | 10,654 (52.6) |
| | Age, median (Q1;Q3) | 46.4 (34.4;55.8) |
| | CCI, n (%) | |
| | 0 | 16,319 (80.6) |
| | 1-2 | 3,254 (16.1) |
| | +3 | 676 (3.3) |
| | Education level, n (%) | |
| | No/Unknown education | 669 (3.3) |
| | Primary | 4,779 (23.6) |
| | Secondary | 8,884 (43.9) |
| | Tertiary | 5,917 (29.2) |
| Older people (65+ years) | Number of persons, n | 18,518 (100.0) |
| | Male, n (%) | 9,199 (49.7) |
| | Age, median (Q1;Q3) | 78.8 (72.8;85.0) |
| | CCI, n (%) | |
| | 0 | 7,010 (37.9) |
| | 1-2 | 7,510 (40.6) |
| | +3 | 3,998 (21.6) |
| | Education level, n (%) | |
| | No/Unknown education | 1,252 (6.8) |
| | Primary | 6,326 (34.2) |
| | Secondary | 7,857 (42.4) |
| | Tertiary | 3,083 (16.6) |

### Supplementary Table 296
### Traumatic brain injury - Incident cohort 2016-2021
### Patient characteristics - relatives - pooled

| *Age strata of index patients* | *Closest relative type* | *Characteristic of family relatives by agestrata of index patients* | *Closest family relatives of brain disorder patients* | *Comparison relative* |
|---|---|---|---|---|
| Children and young people (0-24 years) | Fathers | Number of persons, n | 21,096 (100.0) | 21,096 (100.0) |
| | | Male, n (%) | 21,096 (100.0) | 21,096 (100.0) |
| | | Age, median (Q1;Q3) | 45.2 (38.1;51.5) | 45.4 (38.2;51.3) |
| | | CCI, n (%) | | |
| | | 0 | 18,917 (89.7) | 19,159 (90.8) |
| | | 1-2 | 1,923 (9.1) | 1,741 (8.3) |
| | | +3 | 256 (1.2) | 196 (0.9) |
| | | Education level, n (%) | | |
| | | No/Unknown education | 426 (2.0) | 426 (2.0) |
| | | Primary | 4,125 (19.6) | 4,125 (19.6) |
| | | Secondary | 10,438 (49.5) | 10,438 (49.5) |
| | | Tertiary | 6,107 (28.9) | 6,107 (28.9) |
| | | Relation to patient, n (%) | | |
| | | Father | 21,096 (100.0) | 21,096 (100.0) |
| Children and young people (0-24 years) | Mothers | Number of persons, n | 22,411 (100.0) | 22,411 (100.0) |
| | | Male, n (%) | 0 (0.0) | 0 (0.0) |
| | | Age, median (Q1;Q3) | 42.9 (35.8;49.0) | 43.1 (35.9;49.0) |
| | | CCI, n (%) | | |
| | | 0 | 19,789 (88.3) | 20,267 (90.4) |
| | | 1-2 | 2,435 (10.9) | 1,976 (8.8) |
| | | +3 | 187 (0.8) | 168 (0.7) |
| | | Education level, n (%) | | |
| | | No/Unknown education | 452 (2.0) | 452 (2.0) |
| | | Primary | 3,802 (17.0) | 3,802 (17.0) |
| | | Secondary | 9,259 (41.3) | 9,259 (41.3) |
| | | Tertiary | 8,898 (39.7) | 8,898 (39.7) |
| | | Relation to patient, n (%) | | |
| | | Mother | 22,411 (100.0) | 22,411 (100.0) |
| Young adults and adults (25-64 years) | Pooled | Number of persons, n | 20,249 (100.0) | 20,249 (100.0) |
| | | Male, n (%) | 9,147 (45.2) | 9,147 (45.2) |
| | | Age, median (Q1;Q3) | 48.5 (32.9;58.5) | 48.7 (33.0;58.7) |
| | | CCI, n (%) | | |
| | | 0 | 17,073 (84.3) | 17,297 (85.4) |
| | | 1-2 | 2,622 (12.9) | 2,522 (12.5) |
| | | +3 | 554 (2.7) | 430 (2.1) |
| | | Education level, n (%) | | |
| | | No/Unknown education | 1,356 (6.7) | 1,356 (6.7) |
| | | Primary | 4,811 (23.8) | 4,811 (23.8) |
| | | Secondary | 8,775 (43.3) | 8,775 (43.3) |
| | | Tertiary | 5,307 (26.2) | 5,307 (26.2) |
| | | Relation to patient, n (%) | | |
| | | Mother | 2,967 (14.7) | 2,967 (14.7) |
| | | Father | 897 (4.4) | 897 (4.4) |
| | | Partner | 11,579 (57.2) | 11,579 (57.2) |
| | | Child | 3,271 (16.2) | 3,271 (16.2) |

**Supplementary Table 296**
**Traumatic brain injury - Incident cohort 2016-2021**
**Patient characteristics - relatives - pooled**

| *Age strata of index patients* | *Closest relative type* | *Characteristic of family relatives by agestrata of index patients* | *Closest family relatives of brain disorder patients* | *Comparison relative* |
|---|---|---|---|---|
| | | Sibling | 1,535 (7.6) | 1,535 (7.6) |
| Older people (65+ years) | Pooled | Number of persons, n | 18,518 (100.0) | 18,518 (100.0) |
| | | Male, n (%) | 8,377 (45.2) | 8,377 (45.2) |
| | | Age, median (Q1;Q3) | 63.0 (54.4;73.8) | 63.0 (54.3;73.9) |
| | | CCI, n (%) | | |
| | | 0 | 13,151 (71.0) | 13,632 (73.6) |
| | | 1-2 | 4,118 (22.2) | 3,844 (20.8) |
| | | +3 | 1,249 (6.7) | 1,042 (5.6) |
| | | Education level, n (%) | | |
| | | No/Unknown education | 601 (3.2) | 601 (3.2) |
| | | Primary | 3,931 (21.2) | 3,931 (21.2) |
| | | Secondary | 9,093 (49.1) | 9,093 (49.1) |
| | | Tertiary | 4,893 (26.4) | 4,893 (26.4) |
| | | Relation to patient, n (%) | | |
| | | Partner | 8,474 (45.8) | 8,474 (45.8) |
| | | Child | 9,934 (53.6) | 9,934 (53.6) |
| | | Sibling | 110 (0.6) | 110 (0.6) |

#### Supplementary Table 297
#### Traumatic brain injury - Incident cohort 2016-2021
#### Patient characteristics - relatives - not pooled

| *Age strata of index patients* | *Closest relative type* | *Characteristic of family relatives by agestrata of index patients* | *Closest family relatives of brain disorder patients* | *Comparison relative* |
|---|---|---|---|---|
| Children and young people (0-24 years) | Fathers | Number of persons, n | 21,096 (100.0) | 21,096 (100.0) |
| | | Male, n (%) | 21,096 (100.0) | 21,096 (100.0) |
| | | Age, median (Q1;Q3) | 45.2 (38.1;51.5) | 45.4 (38.2;51.3) |
| | | CCI, n (%) | | |
| | | 0 | 18,917 (89.7) | 19,159 (90.8) |
| | | 1-2 | 1,923 (9.1) | 1,741 (8.3) |
| | | +3 | 256 (1.2) | 196 (0.9) |
| | | Education level, n (%) | | |
| | | No/Unknown education | 426 (2.0) | 426 (2.0) |
| | | Primary | 4,125 (19.6) | 4,125 (19.6) |
| | | Secondary | 10,438 (49.5) | 10,438 (49.5) |
| | | Tertiary | 6,107 (28.9) | 6,107 (28.9) |
| | | Relation to patient, n (%) | | |
| | | Father | 21,096 (100.0) | 21,096 (100.0) |
| Children and young people (0-24 years) | Mothers | Number of persons, n | 22,411 (100.0) | 22,411 (100.0) |
| | | Male, n (%) | 0 (0.0) | 0 (0.0) |
| | | Age, median (Q1;Q3) | 42.9 (35.8;49.0) | 43.1 (35.9;49.0) |
| | | CCI, n (%) | | |
| | | 0 | 19,789 (88.3) | 20,267 (90.4) |
| | | 1-2 | 2,435 (10.9) | 1,976 (8.8) |
| | | +3 | 187 (0.8) | 168 (0.7) |
| | | Education level, n (%) | | |
| | | No/Unknown education | 452 (2.0) | 452 (2.0) |
| | | Primary | 3,802 (17.0) | 3,802 (17.0) |
| | | Secondary | 9,259 (41.3) | 9,259 (41.3) |
| | | Tertiary | 8,898 (39.7) | 8,898 (39.7) |
| | | Relation to patient, n (%) | | |
| | | Mother | 22,411 (100.0) | 22,411 (100.0) |
| Young adults and adults (25-64 years) | Children | Number of persons, n | 3,271 (100.0) | 3,271 (100.0) |
| | | Male, n (%) | 1,678 (51.3) | 1,678 (51.3) |
| | | Age, median (Q1;Q3) | 20.0 (12.2;29.5) | 19.9 (11.9;29.3) |
| | | CCI, n (%) | | |
| | | 0 | 3,044 (93.1) | 3,074 (94.0) |
| | | 1-2 | 218 (6.7) | 187 (5.7) |
| | | +3 | 9 (0.3) | 10 (0.3) |
| | | Education level, n (%) | | |
| | | No/Unknown education | 903 (27.6) | 903 (27.6) |
| | | Primary | 1,235 (37.8) | 1,235 (37.8) |
| | | Secondary | 729 (22.3) | 729 (22.3) |
| | | Tertiary | 404 (12.4) | 404 (12.4) |
| | | Relation to patient, n (%) | | |
| | | Child | 3,271 (100.0) | 3,271 (100.0) |
| Young adults and adults (25-64 years) | Parents | Number of persons, n | 3,864 (100.0) | 3,864 (100.0) |
| | | Male, n (%) | 897 (23.2) | 897 (23.2) |
| | | Age, median (Q1;Q3) | 62.8 (56.4;72.0) | 62.8 (57.0;72.3) |

### Supplementary Table 297
### Traumatic brain injury - Incident cohort 2016-2021
### Patient characteristics - relatives - not pooled

| *Age strata of index patients* | *Closest relative type* | *Characteristic of family relatives by agestrata of index patients* | *Closest family relatives of brain disorder patients* | *Comparison relative* |
|---|---|---|---|---|
| | | CCI, n (%) | | |
| | | 0 | 2,645 (68.5) | 2,767 (71.6) |
| | | 1-2 | 904 (23.4) | 886 (22.9) |
| | | +3 | 315 (8.2) | 211 (5.5) |
| | | Education level, n (%) | | |
| | | No/Unknown education | 70 (1.8) | 70 (1.8) |
| | | Primary | 1,143 (29.6) | 1,143 (29.6) |
| | | Secondary | 1,826 (47.3) | 1,826 (47.3) |
| | | Tertiary | 825 (21.4) | 825 (21.4) |
| | | Relation to patient, n (%) | | |
| | | Mother | 2,967 (76.8) | 2,967 (76.8) |
| | | Father | 897 (23.2) | 897 (23.2) |
| Young adults and adults (25-64 years) | Partners | Number of persons, n | 11,579 (100.0) | 11,579 (100.0) |
| | | Male, n (%) | 5,641 (48.7) | 5,641 (48.7) |
| | | Age, median (Q1;Q3) | 47.5 (37.1;56.2) | 47.5 (37.0;56.2) |
| | | CCI, n (%) | | |
| | | 0 | 10,080 (87.1) | 10,150 (87.7) |
| | | 1-2 | 1,321 (11.4) | 1,255 (10.8) |
| | | +3 | 178 (1.5) | 174 (1.5) |
| | | Education level, n (%) | | |
| | | No/Unknown education | 378 (3.3) | 378 (3.3) |
| | | Primary | 2,020 (17.4) | 2,020 (17.4) |
| | | Secondary | 5,412 (46.7) | 5,412 (46.7) |
| | | Tertiary | 3,769 (32.6) | 3,769 (32.6) |
| | | Relation to patient, n (%) | | |
| | | Partner | 11,579 (100.0) | 11,579 (100.0) |
| Young adults and adults (25-64 years) | Siblings | Number of persons, n | 1,535 (100.0) | 1,535 (100.0) |
| | | Male, n (%) | 931 (60.7) | 931 (60.7) |
| | | Age, median (Q1;Q3) | 51.9 (33.4;58.8) | 51.9 (33.7;58.7) |
| | | CCI, n (%) | | |
| | | 0 | 1,304 (85.0) | 1,306 (85.1) |
| | | 1-2 | 179 (11.7) | 194 (12.6) |
| | | +3 | 52 (3.4) | 35 (2.3) |
| | | Education level, n (%) | | |
| | | No/Unknown education | 5 (0.3) | 5 (0.3) |
| | | Primary | 413 (26.9) | 413 (26.9) |
| | | Secondary | 808 (52.6) | 808 (52.6) |
| | | Tertiary | 309 (20.1) | 309 (20.1) |
| | | Relation to patient, n (%) | | |
| | | Sibling | 1,535 (100.0) | 1,535 (100.0) |
| Older people (65+ years) | Children | Number of persons, n | 9,934 (100.0) | 9,934 (100.0) |
| | | Male, n (%) | 5,052 (50.9) | 5,052 (50.9) |
| | | Age, median (Q1;Q3) | 55.1 (49.2;60.2) | 55.0 (49.3;59.8) |
| | | CCI, n (%) | | |
| | | 0 | 8,244 (83.0) | 8,342 (84.0) |

### Supplementary Table 297
### Traumatic brain injury - Incident cohort 2016-2021
### Patient characteristics - relatives - not pooled

| *Age strata of index patients* | *Closest relative type* | *Characteristic of family relatives by agestrata of index patients* | *Closest family relatives of brain disorder patients* | *Comparison relative* |
|---|---|---|---|---|
| | | 1-2 | 1,454 (14.6) | 1,381 (13.9) |
| | | +3 | 236 (2.4) | 211 (2.1) |
| | | Education level, n (%) | | |
| | | No/Unknown education | 68 (0.7) | 68 (0.7) |
| | | Primary | 1,536 (15.5) | 1,536 (15.5) |
| | | Secondary | 5,137 (51.7) | 5,137 (51.7) |
| | | Tertiary | 3,193 (32.1) | 3,193 (32.1) |
| | | Relation to patient, n (%) | | |
| | | Child | 9,934 (100.0) | 9,934 (100.0) |
| Older people (65+ years) | Partners | Number of persons, n | 8,474 (100.0) | 8,474 (100.0) |
| | | Male, n (%) | 3,260 (38.5) | 3,260 (38.5) |
| | | Age, median (Q1;Q3) | 74.7 (69.8;80.1) | 74.7 (69.8;80.1) |
| | | CCI, n (%) | | |
| | | 0 | 4,833 (57.0) | 5,206 (61.4) |
| | | 1-2 | 2,634 (31.1) | 2,439 (28.8) |
| | | +3 | 1,007 (11.9) | 829 (9.8) |
| | | Education level, n (%) | | |
| | | No/Unknown education | 532 (6.3) | 532 (6.3) |
| | | Primary | 2,379 (28.1) | 2,379 (28.1) |
| | | Secondary | 3,891 (45.9) | 3,891 (45.9) |
| | | Tertiary | 1,672 (19.7) | 1,672 (19.7) |
| | | Relation to patient, n (%) | | |
| | | Partner | 8,474 (100.0) | 8,474 (100.0) |
| Older people (65+ years) | Siblings | Number of persons, n | 110 (100.0) | 110 (100.0) |
| | | Male, n (%) | 65 (59.1) | 65 (59.1) |
| | | Age, median (Q1;Q3) | 62.1 (59.8;64.3) | 62.8 (59.4;64.8) |
| | | CCI, n (%) | | |
| | | 0 | 74 (67.3) | 84 (76.4) |
| | | 1-2 | 30 (27.3) | * |
| | | +3 | 6 (5.5) | * |
| | | Education level, n (%) | | |
| | | No/Unknown education | * | * |
| | | Primary | * | * |
| | | Secondary | 65 (59.1) | 65 (59.1) |
| | | Tertiary | 28 (25.5) | 28 (25.5) |
| | | Relation to patient, n (%) | | |
| | | Sibling | 110 (100.0) | 110 (100.0) |

#### Supplementary Table 298
#### Traumatic brain injury - Incident cohort 2016-2021
#### Cost analysis - pooled

| | | | | Actual costs | | Attributable costs | |
|---|---|---|---|---|---|---|---|
| *Age strata of index patients* | *Closest relative type* | *Number of index patients included in cost component calculations* | *Cost component* | *Total (million EUR)* | *Per person (EUR)* | *Total (million EUR)* | *Per person (EUR)* |
| Children and young people (0-24 years) | Fathers | 21,096 | Primary sector | 4.4 | 210 | 0.3 | 13 |
| | | 21,096 | Hospital care | 28.7 | 1,365 | 4.7 | 225 |
| | | 21,096 | Filled prescriptions | 3.1 | 147 | 0.3 | 13 |
| | | 20,655 | Lost productivity (income loss) | . | . | 35.1 | 1,698 |
| Children and young people (0-24 years) | Mothers | 22,411 | Primary sector | 7.9 | 354 | 0.7 | 33 |
| | | 22,411 | Hospital care | 42.0 | 1,877 | 7.7 | 345 |
| | | 22,411 | Filled prescriptions | 3.9 | 176 | 0.5 | 23 |
| | | 22,301 | Lost productivity (income loss) | . | . | 31.9 | 1,431 |
| Young adults and adults (25-64 years) | Pooled | 20,249 | Primary sector | 6.2 | 308 | 0.3 | 15 |
| | | 20,249 | Hospital care | 43.8 | 2,173 | 4.6 | 227 |
| | | 20,249 | Filled prescriptions | 4.8 | 238 | 0.1 | 6 |
| | | 15,493 | Lost productivity (income loss) | . | . | 35.3 | 2,278 |
| Older people (65+ years) | Pooled | 18,518 | Primary sector | 7.8 | 427 | 0.5 | 27 |
| | | 18,518 | Hospital care | 60.9 | 3,323 | 10.1 | 551 |
| | | 18,518 | Filled prescriptions | 7.6 | 416 | 1.1 | 59 |
| | | 8,837 | Lost productivity (income loss) | . | . | 18.3 | 2,065 |

#### Supplementary Table 299
#### Traumatic brain injury - Incident cohort 2016-2021
#### Cost analysis - not pooled

| *Age strata of index patients* | *Closest relative type* | *Number of index patients included in cost component calculations* | *Cost component* | *Actual costs* | | *Attributable costs* | |
|---|---|---|---|---|---|---|---|
| | | | | *Total (million EUR)* | *Per person (EUR)* | *Total (million EUR)* | *Per person (EUR)* |
| Children and young people (0-24 years) | Fathers | 21,096 | Primary sector | 4.4 | 210 | 0.3 | 13 |
| | | 21,096 | Hospital care | 28.7 | 1,365 | 4.7 | 225 |
| | | 21,096 | Filled prescriptions | 3.1 | 147 | 0.3 | 13 |
| | | 20,655 | Lost productivity (income loss) | . | . | 35.1 | 1,698 |
| Children and young people (0-24 years) | Mothers | 22,411 | Primary sector | 7.9 | 354 | 0.7 | 33 |
| | | 22,411 | Hospital care | 42.0 | 1,877 | 7.7 | 345 |
| | | 22,411 | Filled prescriptions | 3.9 | 176 | 0.5 | 23 |
| | | 22,301 | Lost productivity (income loss) | . | . | 31.9 | 1,431 |
| Young adults and adults (25-64 years) | Children | 3,271 | Primary sector | 0.7 | 214 | 0.0 | 14 |
| | | 3,271 | Hospital care | 5.0 | 1,550 | 0.9 | 284 |
| | | 3,271 | Filled prescriptions | 0.4 | 110 | 0.0 | 5 |
| | | 1,765 | Lost productivity (income loss) | . | . | 1.9 | 1,071 |
| Young adults and adults (25-64 years) | Parents | 3,864 | Primary sector | 1.7 | 438 | 0.1 | 20 |
| | | 3,864 | Hospital care | 13.2 | 3,453 | 2.6 | 671 |
| | | 3,864 | Filled prescriptions | 1.7 | 447 | 0.1 | 22 |
| | | 1,872 | Lost productivity (income loss) | . | . | 8.9 | 4,746 |
| Young adults and adults (25-64 years) | Partners | 11,579 | Primary sector | 3.4 | 295 | 0.1 | 13 |
| | | 11,579 | Hospital care | 22.0 | 1,905 | 1.0 | 85 |
| | | 11,579 | Filled prescriptions | 2.4 | 204 | 0.0 | 2 |
| | | 10,527 | Lost productivity (income loss) | . | . | 20.6 | 1,955 |
| Young adults and adults (25-64 years) | Siblings | 1,535 | Primary sector | 0.4 | 285 | 0.0 | 13 |
| | | 1,535 | Hospital care | 3.5 | 2,322 | 0.1 | 57 |
| | | 1,535 | Filled prescriptions | 0.4 | 248 | 0.0 | 6 |
| | | 1,329 | Lost productivity (income loss) | . | . | 3.9 | 2,960 |
| Older people (65+ years) | Children | 9,934 | Primary sector | 3.2 | 325 | 0.3 | 26 |
| | | 9,934 | Hospital care | 22.7 | 2,287 | 4.4 | 443 |
| | | 9,934 | Filled prescriptions | 2.7 | 270 | 0.5 | 47 |
| | | 8,436 | Lost productivity (income loss) | . | . | 20.3 | 2,402 |
| Older people (65+ years) | Partners | 8,474 | Primary sector | 4.6 | 550 | 0.3 | 31 |
| | | 8,474 | Hospital care | 37.8 | 4,554 | 5.5 | 662 |
| | | 8,474 | Filled prescriptions | 4.9 | 591 | 0.6 | 74 |
| | | 346 | Lost productivity (income loss) | . | . | -1.9 | -5,631 |
| Older people (65+ years) | Siblings | 110 | Primary sector | 0.0 | 321 | -0.0 | -118 |
| | | 110 | Hospital care | 0.4 | 3,767 | 0.2 | 1,886 |
| | | 110 | Filled prescriptions | 0.0 | 305 | -0.0 | -67 |
| | | 55 | Lost productivity (income loss) | . | . | -0.1 | -1,115 |

**Supplementary Table 300**
**Traumatic brain injury - Prevalent cohort 2021**
**Identification of populations of relatives**

| ***Agegroup of index patients in matched population*** | | ***Closest relative type identified*** | | ***Available control with same type of closest relative*** |
|---|---|---|---|---|
| Children and young people (0-24 years) | n = 63,208 | Fathers | n = 52,188 (83%) | 97% |
| | | Mothers | n = 54,908 (87%) | 97% |
| | | No available closest relative | n = 2,926 (5%) | n/a |
| Young adults and adults (25-64 years) | n = 115,506 | Children | n = 14,849 (13%) | 93% |
| | | Parents | n = 23,373 (20%) | 92% |
| | | Partners | n = 59,505 (52%) | 96% |
| | | Siblings | n = 15,660 (14%) | 46% |
| | | Pooled | n = 113,387 (98%) | 88% |
| | | No available closest relative | n = 2,119 (2%) | n/a |
| Older people (65+ years) | n = 40,446 | Children | n = 16,209 (40%) | 94% |
| | | Partners | n = 18,081 (45%) | 95% |
| | | Siblings | n = 1,225 (3%) | 48% |
| | | Pooled | n = 35,515 (88%) | 93% |
| | | No available closest relative | n = 4,931 (12%) | n/a |

**Supplementary Table 301**
**Traumatic brain injury - Prevalent cohort 2021**
**Patient characteristics - cases**

| ***Age strata of index patients*** | ***Characteristic*** | |
|---|---|---|
| Children and young people (0-24 years) | Number of persons, n | 58,596 (100.0) |
| | Male, n (%) | 32,938 (56.2) |
| | Age, median (Q1;Q3) | 17.3 (11.9;21.3) |
| | CCI, n (%) | |
| | 0 | 54,398 (92.8) |
| | 1-2 | 4,064 (6.9) |
| | +3 | 134 (0.2) |
| | Education level, n (%) | |
| | No/Unknown education | 17,605 (30.0) |
| | Primary | 29,543 (50.4) |
| | Secondary | 10,539 (18.0) |
| | Tertiary | 909 (1.6) |
| Young adults and adults (25-64 years) | Number of persons, n | 99,982 (100.0) |
| | Male, n (%) | 56,552 (56.6) |
| | Age, median (Q1;Q3) | 40.4 (31.7;52.4) |
| | CCI, n (%) | |
| | 0 | 84,225 (84.2) |
| | 1-2 | 13,582 (13.6) |
| | +3 | 2,175 (2.2) |
| | Education level, n (%) | |
| | No/Unknown education | 1,636 (1.6) |
| | Primary | 24,836 (24.8) |
| | Secondary | 44,262 (44.3) |
| | Tertiary | 29,248 (29.3) |
| Older people (65+ years) | Number of persons, n | 33,260 (100.0) |
| | Male, n (%) | 16,311 (49.0) |
| | Age, median (Q1;Q3) | 75.7 (70.5;81.9) |
| | CCI, n (%) | |
| | 0 | 14,801 (44.5) |
| | 1-2 | 12,773 (38.4) |
| | +3 | 5,686 (17.1) |
| | Education level, n (%) | |
| | No/Unknown education | 2,032 (6.1) |
| | Primary | 10,057 (30.2) |
| | Secondary | 14,771 (44.4) |
| | Tertiary | 6,400 (19.2) |

**Supplementary Table 302**
**Traumatic brain injury - Prevalent cohort 2021**
**Patient characteristics - relatives - pooled**

| *Age strata of index patients* | *Closest relative type* | *Characteristic of family relatives by agestrata of index patients* | *Closest family relatives of brain disorder patients* | *Comparison relative* |
|---|---|---|---|---|
| Children and young people (0-24 years) | Fathers | Number of persons, n | 50,657 (100.0) | 50,657 (100.0) |
| | | Male, n (%) | 50,657 (100.0) | 50,657 (100.0) |
| | | Age, median (Q1;Q3) | 49.1 (43.6;54.0) | 49.2 (43.9;53.9) |
| | | CCI, n (%) | | |
| | | 0 | 44,450 (87.7) | 45,026 (88.9) |
| | | 1-2 | 5,476 (10.8) | 5,035 (9.9) |
| | | +3 | 731 (1.4) | 596 (1.2) |
| | | Education level, n (%) | | |
| | | No/Unknown education | 716 (1.4) | 716 (1.4) |
| | | Primary | 9,472 (18.7) | 9,472 (18.7) |
| | | Secondary | 25,924 (51.2) | 25,924 (51.2) |
| | | Tertiary | 14,545 (28.7) | 14,545 (28.7) |
| | | Relation to patient, n (%) | | |
| | | Father | 50,657 (100.0) | 50,657 (100.0) |
| Children and young people (0-24 years) | Mothers | Number of persons, n | 53,396 (100.0) | 53,396 (100.0) |
| | | Male, n (%) | 0 (0.0) | 0 (0.0) |
| | | Age, median (Q1;Q3) | 46.9 (41.4;51.6) | 47.1 (41.7;51.5) |
| | | CCI, n (%) | | |
| | | 0 | 46,308 (86.7) | 47,260 (88.5) |
| | | 1-2 | 6,492 (12.2) | 5,660 (10.6) |
| | | +3 | 596 (1.1) | 476 (0.9) |
| | | Education level, n (%) | | |
| | | No/Unknown education | 606 (1.1) | 606 (1.1) |
| | | Primary | 8,505 (15.9) | 8,505 (15.9) |
| | | Secondary | 22,957 (43.0) | 22,957 (43.0) |
| | | Tertiary | 21,328 (39.9) | 21,328 (39.9) |
| | | Relation to patient, n (%) | | |
| | | Mother | 53,396 (100.0) | 53,396 (100.0) |
| Young adults and adults (25-64 years) | Pooled | Number of persons, n | 99,982 (100.0) | 99,982 (100.0) |
| | | Male, n (%) | 42,559 (42.6) | 42,559 (42.6) |
| | | Age, median (Q1;Q3) | 44.2 (30.8;57.8) | 44.1 (30.9;57.9) |
| | | CCI, n (%) | | |
| | | 0 | 85,062 (85.1) | 86,115 (86.1) |
| | | 1-2 | 12,655 (12.7) | 11,923 (11.9) |
| | | +3 | 2,265 (2.3) | 1,944 (1.9) |
| | | Education level, n (%) | | |
| | | No/Unknown education | 6,385 (6.4) | 6,385 (6.4) |
| | | Primary | 22,710 (22.7) | 22,710 (22.7) |
| | | Secondary | 43,567 (43.6) | 43,567 (43.6) |
| | | Tertiary | 27,320 (27.3) | 27,320 (27.3) |
| | | Relation to patient, n (%) | | |
| | | Mother | 16,478 (16.5) | 16,478 (16.5) |
| | | Father | 5,080 (5.1) | 5,080 (5.1) |
| | | Partner | 57,288 (57.3) | 57,288 (57.3) |
| | | Child | 13,906 (13.9) | 13,906 (13.9) |

**Supplementary Table 302**
**Traumatic brain injury - Prevalent cohort 2021**
**Patient characteristics - relatives - pooled**

| *Age strata of index patients* | *Closest relative type* | *Characteristic of family relatives by agestrata of index patients* | *Closest family relatives of brain disorder patients* | *Comparison relative* |
|---|---|---|---|---|
| | | Sibling | 7,230 (7.2) | 7,230 (7.2) |
| Older people (65+ years) | Pooled | Number of persons, n | 33,260 (100.0) | 33,260 (100.0) |
| | | Male, n (%) | 14,999 (45.1) | 14,999 (45.1) |
| | | Age, median (Q1;Q3) | 64.7 (54.1;73.7) | 64.8 (54.0;73.7) |
| | | CCI, n (%) | | |
| | | 0 | 23,693 (71.2) | 24,169 (72.7) |
| | | 1-2 | 7,460 (22.4) | 7,164 (21.5) |
| | | +3 | 2,107 (6.3) | 1,927 (5.8) |
| | | Education level, n (%) | | |
| | | No/Unknown education | 1,081 (3.3) | 1,081 (3.3) |
| | | Primary | 6,818 (20.5) | 6,818 (20.5) |
| | | Secondary | 15,912 (47.8) | 15,912 (47.8) |
| | | Tertiary | 9,449 (28.4) | 9,449 (28.4) |
| | | Relation to patient, n (%) | | |
| | | Partner | 17,357 (52.2) | 17,357 (52.2) |
| | | Child | 15,305 (46.0) | 15,305 (46.0) |
| | | Sibling | 598 (1.8) | 598 (1.8) |

#### Supplementary Table 303
#### Traumatic brain injury - Prevalent cohort 2021
#### Patient characteristics - relatives - not pooled

| *Age strata of index patients* | *Closest relative type* | *Characteristic of family relatives by agestrata of index patients* | *Closest family relatives of brain disorder patients* | *Comparison relative* |
|---|---|---|---|---|
| Children and young people (0-24 years) | Fathers | Number of persons, n | 50,657 (100.0) | 50,657 (100.0) |
| | | Male, n (%) | 50,657 (100.0) | 50,657 (100.0) |
| | | Age, median (Q1;Q3) | 49.1 (43.6;54.0) | 49.2 (43.9;53.9) |
| | | CCI, n (%) | | |
| | | 0 | 44,450 (87.7) | 45,026 (88.9) |
| | | 1-2 | 5,476 (10.8) | 5,035 (9.9) |
| | | +3 | 731 (1.4) | 596 (1.2) |
| | | Education level, n (%) | | |
| | | No/Unknown education | 716 (1.4) | 716 (1.4) |
| | | Primary | 9,472 (18.7) | 9,472 (18.7) |
| | | Secondary | 25,924 (51.2) | 25,924 (51.2) |
| | | Tertiary | 14,545 (28.7) | 14,545 (28.7) |
| | | Relation to patient, n (%) | | |
| | | Father | 50,657 (100.0) | 50,657 (100.0) |
| Children and young people (0-24 years) | Mothers | Number of persons, n | 53,396 (100.0) | 53,396 (100.0) |
| | | Male, n (%) | 0 (0.0) | 0 (0.0) |
| | | Age, median (Q1;Q3) | 46.9 (41.4;51.6) | 47.1 (41.7;51.5) |
| | | CCI, n (%) | | |
| | | 0 | 46,308 (86.7) | 47,260 (88.5) |
| | | 1-2 | 6,492 (12.2) | 5,660 (10.6) |
| | | +3 | 596 (1.1) | 476 (0.9) |
| | | Education level, n (%) | | |
| | | No/Unknown education | 606 (1.1) | 606 (1.1) |
| | | Primary | 8,505 (15.9) | 8,505 (15.9) |
| | | Secondary | 22,957 (43.0) | 22,957 (43.0) |
| | | Tertiary | 21,328 (39.9) | 21,328 (39.9) |
| | | Relation to patient, n (%) | | |
| | | Mother | 53,396 (100.0) | 53,396 (100.0) |
| Young adults and adults (25-64 years) | Children | Number of persons, n | 13,906 (100.0) | 13,906 (100.0) |
| | | Male, n (%) | 7,258 (52.2) | 7,258 (52.2) |
| | | Age, median (Q1;Q3) | 17.8 (9.7;27.3) | 17.5 (9.1;27.1) |
| | | CCI, n (%) | | |
| | | 0 | 13,041 (93.8) | 13,165 (94.7) |
| | | 1-2 | 831 (6.0) | 723 (5.2) |
| | | +3 | 34 (0.2) | 18 (0.1) |
| | | Education level, n (%) | | |
| | | No/Unknown education | 4,967 (35.7) | 4,967 (35.7) |
| | | Primary | 5,033 (36.2) | 5,033 (36.2) |
| | | Secondary | 2,659 (19.1) | 2,659 (19.1) |
| | | Tertiary | 1,247 (9.0) | 1,247 (9.0) |
| | | Relation to patient, n (%) | | |
| | | Child | 13,906 (100.0) | 13,906 (100.0) |
| Young adults and adults (25-64 years) | Parents | Number of persons, n | 21,558 (100.0) | 21,558 (100.0) |
| | | Male, n (%) | 5,080 (23.6) | 5,080 (23.6) |
| | | Age, median (Q1;Q3) | 62.2 (56.8;70.2) | 62.3 (57.1;70.4) |

#### Supplementary Table 303
#### Traumatic brain injury - Prevalent cohort 2021
#### Patient characteristics - relatives - not pooled

| *Age strata of index patients* | *Closest relative type* | *Characteristic of family relatives by agestrata of index patients* | *Closest family relatives of brain disorder patients* | *Comparison relative* |
|---|---|---|---|---|
| | | CCI, n (%) | | |
| | | 0 | 15,059 (69.9) | 15,490 (71.9) |
| | | 1-2 | 5,149 (23.9) | 4,879 (22.6) |
| | | +3 | 1,350 (6.3) | 1,189 (5.5) |
| | | Education level, n (%) | | |
| | | No/Unknown education | 313 (1.5) | 313 (1.5) |
| | | Primary | 6,009 (27.9) | 6,009 (27.9) |
| | | Secondary | 10,567 (49.0) | 10,567 (49.0) |
| | | Tertiary | 4,669 (21.7) | 4,669 (21.7) |
| | | Relation to patient, n (%) | | |
| | | Mother | 16,478 (76.4) | 16,478 (76.4) |
| | | Father | 5,080 (23.6) | 5,080 (23.6) |
| Young adults and adults (25-64 years) | Partners | Number of persons, n | 57,288 (100.0) | 57,288 (100.0) |
| | | Male, n (%) | 25,733 (44.9) | 25,733 (44.9) |
| | | Age, median (Q1;Q3) | 40.7 (32.0;52.5) | 40.7 (32.1;52.4) |
| | | CCI, n (%) | | |
| | | 0 | 50,840 (88.7) | 51,292 (89.5) |
| | | 1-2 | 5,736 (10.0) | 5,397 (9.4) |
| | | +3 | 712 (1.2) | 599 (1.0) |
| | | Education level, n (%) | | |
| | | No/Unknown education | 1,077 (1.9) | 1,077 (1.9) |
| | | Primary | 9,697 (16.9) | 9,697 (16.9) |
| | | Secondary | 26,593 (46.4) | 26,593 (46.4) |
| | | Tertiary | 19,921 (34.8) | 19,921 (34.8) |
| | | Relation to patient, n (%) | | |
| | | Partner | 57,288 (100.0) | 57,288 (100.0) |
| Young adults and adults (25-64 years) | Siblings | Number of persons, n | 7,230 (100.0) | 7,230 (100.0) |
| | | Male, n (%) | 4,488 (62.1) | 4,488 (62.1) |
| | | Age, median (Q1;Q3) | 49.3 (32.4;58.7) | 49.5 (32.4;58.7) |
| | | CCI, n (%) | | |
| | | 0 | 6,122 (84.7) | 6,168 (85.3) |
| | | 1-2 | 939 (13.0) | 924 (12.8) |
| | | +3 | 169 (2.3) | 138 (1.9) |
| | | Education level, n (%) | | |
| | | No/Unknown education | 28 (0.4) | 28 (0.4) |
| | | Primary | 1,971 (27.3) | 1,971 (27.3) |
| | | Secondary | 3,748 (51.8) | 3,748 (51.8) |
| | | Tertiary | 1,483 (20.5) | 1,483 (20.5) |
| | | Relation to patient, n (%) | | |
| | | Sibling | 7,230 (100.0) | 7,230 (100.0) |
| Older people (65+ years) | Children | Number of persons, n | 15,305 (100.0) | 15,305 (100.0) |
| | | Male, n (%) | 7,637 (49.9) | 7,637 (49.9) |
| | | Age, median (Q1;Q3) | 53.3 (46.3;59.2) | 53.2 (46.3;59.0) |
| | | CCI, n (%) | | |
| | | 0 | 12,841 (83.9) | 13,042 (85.2) |

#### Supplementary Table 303
#### Traumatic brain injury - Prevalent cohort 2021
#### Patient characteristics - relatives - not pooled

| *Age strata of index patients* | *Closest relative type* | *Characteristic of family relatives by agestrata of index patients* | *Closest family relatives of brain disorder patients* | *Comparison relative* |
|---|---|---|---|---|
| | | 1-2 | 2,101 (13.7) | 1,974 (12.9) |
| | | +3 | 363 (2.4) | 289 (1.9) |
| | | Education level, n (%) | | |
| | | No/Unknown education | 56 (0.4) | 56 (0.4) |
| | | Primary | 2,458 (16.1) | 2,458 (16.1) |
| | | Secondary | 7,413 (48.4) | 7,413 (48.4) |
| | | Tertiary | 5,378 (35.1) | 5,378 (35.1) |
| | | Relation to patient, n (%) | | |
| | | Child | 15,305 (100.0) | 15,305 (100.0) |
| Older people (65+ years) | Partners | Number of persons, n | 17,357 (100.0) | 17,357 (100.0) |
| | | Male, n (%) | 7,032 (40.5) | 7,032 (40.5) |
| | | Age, median (Q1;Q3) | 73.3 (68.2;78.1) | 73.3 (68.5;77.9) |
| | | CCI, n (%) | | |
| | | 0 | 10,410 (60.0) | 10,697 (61.6) |
| | | 1-2 | 5,234 (30.2) | 5,046 (29.1) |
| | | +3 | 1,713 (9.9) | 1,614 (9.3) |
| | | Education level, n (%) | | |
| | | No/Unknown education | 1,022 (5.9) | 1,022 (5.9) |
| | | Primary | 4,239 (24.4) | 4,239 (24.4) |
| | | Secondary | 8,137 (46.9) | 8,137 (46.9) |
| | | Tertiary | 3,959 (22.8) | 3,959 (22.8) |
| | | Relation to patient, n (%) | | |
| | | Partner | 17,357 (100.0) | 17,357 (100.0) |
| Older people (65+ years) | Siblings | Number of persons, n | 598 (100.0) | 598 (100.0) |
| | | Male, n (%) | 330 (55.2) | 330 (55.2) |
| | | Age, median (Q1;Q3) | 62.5 (59.6;65.0) | 62.7 (59.7;64.9) |
| | | CCI, n (%) | | |
| | | 0 | 442 (73.9) | 430 (71.9) |
| | | 1-2 | 125 (20.9) | 144 (24.1) |
| | | +3 | 31 (5.2) | 24 (4.0) |
| | | Education level, n (%) | | |
| | | No/Unknown education | * | * |
| | | Primary | 121 (20.2) | 121 (20.2) |
| | | Secondary | 362 (60.5) | 362 (60.5) |
| | | Tertiary | * | * |
| | | Relation to patient, n (%) | | |
| | | Sibling | 598 (100.0) | 598 (100.0) |

#### Supplementary Table 304
#### Traumatic brain injury - Prevalent cohort 2021
#### Cost analysis - pooled

| | | | | *Actual costs* | | *Attributable costs* | |
|---|---|---|---|---|---|---|---|
| *Age strata of index patients* | *Closest relative type* | *Number of index patients included in cost component calculations* | *Cost component* | *Total (million EUR)* | *Per person (EUR)* | *Total (million EUR)* | *Per person (EUR)* |
| Children and young people (0-24 years) | Fathers | 50,657 | Primary sector | 11.5 | 227 | 0.8 | 16 |
| | | 50,657 | Hospital care | 74.6 | 1,475 | 11.0 | 218 |
| | | 50,657 | Filled prescriptions | 8.8 | 174 | 0.6 | 12 |
| | | 49,360 | Lost productivity (income loss) | . | . | 48.9 | 991 |
| Children and young people (0-24 years) | Mothers | 53,396 | Primary sector | 19.4 | 365 | 1.7 | 32 |
| | | 53,396 | Hospital care | 100.1 | 1,876 | 17.6 | 331 |
| | | 53,396 | Filled prescriptions | 11.4 | 214 | 1.0 | 19 |
| | | 53,105 | Lost productivity (income loss) | . | . | 55.1 | 1,038 |
| Young adults and adults (25-64 years) | Pooled | 99,982 | Primary sector | 30.9 | 310 | 1.4 | 14 |
| | | 99,982 | Hospital care | 204.7 | 2,054 | 23.8 | 238 |
| | | 99,982 | Filled prescriptions | 24.6 | 246 | 2.7 | 28 |
| | | 77,847 | Lost productivity (income loss) | . | . | 44.3 | 569 |
| Older people (65+ years) | Pooled | 33,260 | Primary sector | 14.2 | 429 | 0.7 | 21 |
| | | 33,260 | Hospital care | 101.6 | 3,080 | 5.5 | 168 |
| | | 33,260 | Filled prescriptions | 13.9 | 421 | 1.4 | 42 |
| | | 14,828 | Lost productivity (income loss) | . | . | 34.4 | 2,321 |

### Supplementary Table 305
### Traumatic brain injury - Prevalent cohort 2021
### Cost analysis - not pooled

| Age strata of index patients | Closest relative type | Number of index patients included in cost component calculations | Cost component | Actual costs | | Attributable costs | |
|---|---|---|---|---|---|---|---|
| | | | | Total (million EUR) | Per person (EUR) | Total (million EUR) | Per person (EUR) |
| Children and young people (0-24 years) | Fathers | 50,657 | Primary sector | 11.5 | 227 | 0.8 | 16 |
| | | 50,657 | Hospital care | 74.6 | 1,475 | 11.0 | 218 |
| | | 50,657 | Filled prescriptions | 8.8 | 174 | 0.6 | 12 |
| | | 49,360 | Lost productivity (income loss) | . | . | 48.9 | 991 |
| Children and young people (0-24 years) | Mothers | 53,396 | Primary sector | 19.4 | 365 | 1.7 | 32 |
| | | 53,396 | Hospital care | 100.1 | 1,876 | 17.6 | 331 |
| | | 53,396 | Filled prescriptions | 11.4 | 214 | 1.0 | 19 |
| | | 53,105 | Lost productivity (income loss) | . | . | 55.1 | 1,038 |
| Young adults and adults (25-64 years) | Children | 13,906 | Primary sector | 2.9 | 212 | 0.3 | 18 |
| | | 13,906 | Hospital care | 18.1 | 1,303 | -1.0 | -70 |
| | | 13,906 | Filled prescriptions | 1.6 | 113 | 0.3 | 24 |
| | | 6,255 | Lost productivity (income loss) | . | . | 14.2 | 2,267 |
| Young adults and adults (25-64 years) | Parents | 21,558 | Primary sector | 9.4 | 439 | 0.6 | 27 |
| | | 21,558 | Hospital care | 68.7 | 3,215 | 10.5 | 493 |
| | | 21,558 | Filled prescriptions | 10.0 | 467 | 1.0 | 44 |
| | | 11,131 | Lost productivity (income loss) | . | . | 4.7 | 418 |
| Young adults and adults (25-64 years) | Partners | 57,288 | Primary sector | 16.5 | 289 | 0.5 | 9 |
| | | 57,288 | Hospital care | 101.9 | 1,781 | 11.7 | 204 |
| | | 57,288 | Filled prescriptions | 11.2 | 196 | 1.5 | 26 |
| | | 54,155 | Lost productivity (income loss) | . | . | 18.9 | 349 |
| Young adults and adults (25-64 years) | Siblings | 7,230 | Primary sector | 2.1 | 291 | 0.1 | 7 |
| | | 7,230 | Hospital care | 16.1 | 2,230 | 2.5 | 348 |
| | | 7,230 | Filled prescriptions | 1.8 | 253 | -0.0 | -4 |
| | | 6,306 | Lost productivity (income loss) | . | . | 6.5 | 1,037 |
| Older people (65+ years) | Children | 15,305 | Primary sector | 5.0 | 326 | 0.2 | 16 |
| | | 15,305 | Hospital care | 31.8 | 2,082 | 1.4 | 95 |
| | | 15,305 | Filled prescriptions | 4.0 | 261 | 0.3 | 19 |
| | | 13,439 | Lost productivity (income loss) | . | . | 33.0 | 2,459 |
| Older people (65+ years) | Partners | 17,357 | Primary sector | 9.0 | 524 | 0.4 | 24 |
| | | 17,357 | Hospital care | 68.0 | 3,974 | 3.3 | 194 |
| | | 17,357 | Filled prescriptions | 9.6 | 563 | 1.0 | 61 |
| | | 1,068 | Lost productivity (income loss) | . | . | 0.2 | 172 |
| Older people (65+ years) | Siblings | 598 | Primary sector | 0.2 | 388 | 0.0 | 29 |
| | | 598 | Hospital care | 1.8 | 2,990 | 0.8 | 1,296 |
| | | 598 | Filled prescriptions | 0.2 | 409 | 0.1 | 95 |
| | | 321 | Lost productivity (income loss) | . | . | 1.2 | 3,684 |
